# Biomedical consequences of elevated cholesterol-containing lipoproteins and apolipoproteins

**DOI:** 10.1101/2022.03.11.22272251

**Authors:** AF Schmidt, R Joshi, M Gordillo-Marañón, F Drenos, P Charoen, C Giambartolomei, JC Bis, TR Gaunt, AD Hughes, DA Lawlor, A Wong, JF Price, N Chaturvedi, G Wannamethee, N Franceschini, M Kivimaki, AD Hingorani, C Finan

## Abstract

**Aims:** To provide a comprehensive evaluation of the biomedical effects of circulating concentrations of cholesterol-containing lipoproteins and apolipoproteins.

**Methods and Results:** Nuclear magnetic resonance (NMR) spectroscopy was used to measure the cholesterol content of high density (HDL-C), very low-density (VLDL-C), intermediate-density (IDL-C), and low-density (LDL-C) lipoprotein fractions; apolipoproteins Apo-A1 and Apo-B; as well as total triglycerides (TG), remnant-cholesterol (Rem-chol) and total cholesterol (TC). The causal effects of these exposures were assessed against 33 cardiovascular as well as non-cardiovascular outcomes using two-sample univariable and multivariable Mendelian randomization. We observed that most cholesterol containing lipoproteins and apolipoproteins affected coronary heart disease (CHD), cIMT, carotid plaque, CRP and blood pressure. Through MVMR we showed that many of these exposures acted independently of the more commonly measured blood lipids: HDL-C, LDL-C and TG. We furthermore found that HF risk was increased by higher concentrations of TG, VLDL-C, Rem-Chol and Apo-B, often independently of LDL-C, HDL-C or TG. Finally, a smaller subset of these exposures could be robustly mapped to non-CVD traits such as Alzheimer’s disease (HDL-C, LDL-C, IDL-C, Apo-B), type 2 diabetes (VLDL-C, IDL-C, LDL-C), and inflammatory bowel disease (LDL-C, IDL-C).

**Conclusion:** The cholesterol content of a wide range of lipoprotein and apolipoproteins affected measures of atherosclerosis and CHD, implicating subfractions beyond LDL-C. Many of the observed effects acted independently of LDL-C, HDL-C, and TG, supporting the potential for additional, non-LDL-C, avenues to disease prevention.

## Introduction

Circulating concentrations of cholesterol-containing lipoproteins have been linked to risk of atherosclerotic cardiovascular disease (CVD)^1^, in particular coronary heart disease (CHD). Certain circulating lipids have also been implicated in other late life disorders such as dementia^2^, type 2 diabetes (T2DM)^3^, and Crohn’s disease (CD)^4^.

The major blood lipid components, free cholesterol, cholesteryl-esters, and triglycerides are transported in the core of membrane bound lipoprotein particles. Large lipoprotein particles are triglyceride-rich and encompass chylomicrons derived from dietary fat, and very-low density lipoproteins synthesised in the liver. These particles carry a single apolipoprotein B (Apo-B) on the surface (Apo-B 48 for chylomicrons and Apo-B 100 otherwise), and are progressively depleted of triglycerides, through the action of lipoprotein lipase, becoming smaller, denser, and proportionately richer in cholesterol. Lipoproteins, are involved in the process of transporting cholesterol to peripheral tissues (endogenous transport), and are classified according to density gradient centrifugation as (VLDL) very-low-density-, (IDL) intermediate-density- and (LDL) low-density-lipoproteins. Reverse cholesterol transport, from tissues to liver, is mediated by high-density lipoprotein (HDL) particles that are synthesised and released from the liver in nascent form, and which possess membrane-bound apolipoprotein A1 (Apo-A1).

Evidence from non-randomized (i.e., observations) studies, monogenic disorders (FH)^5^, and drug trials of LDL-C lowering drugs^6,7^ have convincingly shown that higher concentrations of LDL-C increase CHD risk. While non-randomized studies have provide similar evidence^8,9^ of a CHD association by HDL-C and total triglycerides (TG, the aggregated across all lipoprotein particles) concentrations, the lack of successful drugs targeting these blood lipids casts doubt on their potential causal role in CHD. For example, the protective CHD effect of the recently marketed ANGPTL3-inhibitor evinacumab was attributed to its LDL-C reducing ability, despite evinacumab showing strong decreasing TG and HDL-C increasing effects^10^.

In fact most lipid lower drugs, including statins and PCSK9 inhibitors, affect multiple lipid fractions^11–13^. This highlights an inferential challenge, where an exposure may affect disease through multiple independent pathways its (marginal) effect reflects the sum of all pathways and is referred to as the *“total effect”*. To consider the potentially distinct causal effect of each pathway, mediation analyses can be used to decompose a total effect in multiple, pathway specific, effects; for example into CHD effects attributable to LDL-C, HDL-C and TG.

Recently, low-cost, high-throughput nuclear magnetic resonance (NMR) spectroscopy allows for accurate, and detailed quantification of cholesterol containing lipoprotein and apolipoprotein concentrations^14^. This makes it feasible to investigate the associations of VLDL-C and IDL-C^14^, as well as remnant cholesterol (Rem-Chol = TC minus HDL-C and LDL-C). Cholesterol-containing lipoproteins and apolipoproteins are however involved in the same lipid-metabolism. As such it becomes relevant to consider to what extend the various cholesterol containing lipoproteins and apolipoproteins *“directly effect”* disease, or whether some of their effects are mediated by the more routinely measured blood lipids: LDL-C, HDL-C, and TG. Identification of direct effects (independent of LDL-C, HDL-C, and TG) is important because these provide evidence for additional avenues for disease prevention; see figure 1 for an illustrative example.

**Figure 1.**
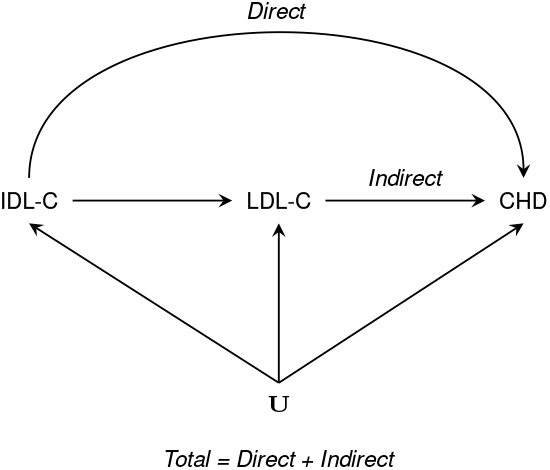
Illustrating the difference between total, direct, and indirect effects, using a hypothetical diagram of intermediate-density lipoprotein cholesterol, low-density lipoprotein cholesterol and coronary heart disease. N.b. IDL-C: intermediate-density lipoprotein cholesterol, LDL-C: low-density lipoprotein cholesterol, CHD: coronary heart disease, and common causes (confounders) represented by **U**.

Genome-wide association studies (GWAS)^15^ of NMR quantified lipoprotein subfractions have identified genetic variants that can be used to undertake Mendelian randomisation (MR) analyses to help ascertain their causal relevance for common disorders. By leveraging genetic variants associated with the exposure(s) of interest, and in the absence of (residual) horizontal pleiotropy, MR protects against bias due to confounding^16^ and reverse causation, biases which may befall non-randomized studies. Multivariable MR (MVMR) incorporates additional information on variants effect on multiple exposures increasing the plausibility of the no-horizontal pleiotropy assumption, and identifies direct effects of the considered exposures^17–19^.

Here we use genetic associations on NMR-measured metabolites and apply two-sample MR to determine the causal relevance of the cholesterol content on different lipoprotein subfractions (including Rem-Chol), as well as Apo-A1 and Apo-B, on a range of cardiovascular (CVD) outcomes, disease biomarkers, measures of organ or systems function as well as late-in-life non-CVD conditions. MVMR is subsequently performed to ascertain whether causal effects might be independent of the routinely measured blood lipids LDL-C, HDL-C, and TG. While NMR spectroscopy provides a detailed sub-classification by particle size, clinical practice, and most interventions, are guided by effects of the total cholesterol content of distinct lipoprotein, hence in the following we will focus on identifying the likely causal effects of these exposures.

## Methods

### Available NMR data

To evaluate the consequences of elevated concentration of circulating cholesterol-containing lipoproteins and apolipoproteins, we sourced genetic associations from meta-analyses of Kettunen^15^ *et al*., and UCLEB^20^ (n=33,029) utilizing NMR-based measurements made using the Nightingale platform on VLDL-C, IDL-C, LDL-C, HDL-C, Rem-Chol, TC, TG, Apo-A1, and Apo-B. Independent replication data on LDL-C, HDL-C, and TG, were available from the Global Lipids genetics Consortium(GLGC^21^, n=188,577) based on clinical chemistry measures. While the UK biobank (UKB) has NMR measurement available for a large sample of participants, it is also a major contributor to the outcome data described in the data availability section. In the presence of sample overlap, weak-instruments may result anti-conservative behaviour (due to an inflated type 1 error rate). We therefore used the relatively smaller UCLEB-Kettunen data, which more closely follows the two-sample paradigm, guarding against the aforementioned type 1 error inflation^22^.

### Selection of genetic instruments for lipoproteins and apolipoproteins

Genetic instruments were selected from throughout the genome using a p-value ≤ 10^−6^ threshold (i.e., F-statistic > 24) and a minor allele frequency (MAF) of at least 0.01. Variants were clumped to a linkage disequilibrium (LD) R-squared threshold of 0.10 based on a random sample of 5,000 unrelated UKB participants of European ancestry.

Following Schmidt *et al*. 2020^41^, we repeated the Apo-B and Apo-A1 genome-wide MR analyses, additionally applying a *cis-*MR approach, which is often more robust to possible horizontal pleiotropy. For *cis-*MR analysis, variants were selected from within a 50kbp window surrounding *APOB* (ENSG00000084674) and *APOA1* (ENSG00000118137). Given the lower number of candidate instruments in a *cis* region (compared to a genome-wide MR), we increased our p-value threshold to 10^−4^ (F-statistic > 15).

### Statistical analyses

To determine the interdependence between the various exposures, pairwise Spearman’s correlation coefficients were calculated between the genetic effect estimates of cholesterol-containing lipoproteins and apolipoproteins and compared to the correlation coefficient of their actual blood concentrations using an 14,834 individual participant sample contributed by UCLEB.

MR analyses were conducted following the two-sample paradigm, ensuring that any bias due to (conditional) weak-instruments is attenuated towards the null. Residual LD was modelled through generalised least squares (GLS)^42,43^ implementations of the inverse variance weighted (IVW) and MR-Egger estimators. Here the univariable MR methods provide *total effect* estimates, and multivariable MR (MVMR) implementations of IVW and MR-Egger (both implemented as GLS) were used to estimate *direct effects*, independent from combinations of LDL-C, HDL-C and TG. Additionally, addressing the growing interest in Apo-B as a fundamental cause of atherosclerosis, we explored a MVMR model with Apo-B conditioned on HDL-C and TG, excluding LDL-C due to its high correlation (0.90) with Apo-B (Figure S1).

To minimize the potential influence of horizontal pleiotropy we excluded variants with large leverage or outlier statistics^41,44^ and used the Q-statistic to identify possible remaining violations^44,45^. A model selection framework^45^ was applied to select the most appropriate estimator (IVW or MR-Egger) for each specific exposure – outcome relationship, where the Egger correction is unbiased even in the extreme setting where 100% of the selected variants affect disease through horizontal pleiotropy but has markedly less power.

Multivariable methods, such as MVMR, may falter when considering (conditionally) multicollinear variables – whose inclusion leads to numerically unstable models with noticeably lower precision^46^, and can result in conditionally weak-instrument settings^47^. For example, the strong correlation between LDL-C and Apo-B (Figure S1) is anticipated to destabilize a model that includes both. While there are MR methods specifically designed to address such highly correlated data they assume a complete absence of horizontal pleiotropy, which is unlikely to hold^47,48^ and computationally prohibitive^47^. We therefore decided to identify and downweigh evidence likely affected by multicollinearity. First, we included estimates of precision (the multiplicative inverse of the standard error) as well as conditional F-statistics, where any substantial change is indicative of model instability and weak-instrument settings. Second, erroneous results were identified by gradually extending the MVMR models to first consider the influence of each single covariate (genetic instruments with LDL-C, HDL-C, or TG only), before fitting a “*fully conditional*” MVMR model including all three blood lipids. When LDL-C, HDL-C, or TG was the exposure of interest, we only corrected for the remaining two covariates; for example, LDL-C was only considered conditional on HDL-C and TG. After filtering on significance (at an alpha of 0.05), MR estimates potentially affected by instability were removed by focussing on exposure-outcome relationships with 60% or higher directional concordance (i.e., significant, and directionally concordant in 3 out of 5 models). The five models constituted estimates of i) the total effect (from the univariable MR models), and direct effects adjusting for ii) LDL-C, iii) HDL-C, or iv) TG, and v) all three exposures jointly. When LDL-C, HDL-C, or TG was the exposure of interest, adjustments were made for the two remaining exposures only.

### Prioritization analyses

Under the null-hypothesis the p-values of a group of tests follow an uniform distribution between zero and one^49^. Hence to explore the influence of multiplicity, we performed “overall” null-hypothesis tests. Here, p-values were grouped by exposure or outcome, and the empirical p-value distribution was tested against the uniform distribution expected under the null-hypothesis using Kolmogorov-Smirnov “KS”-tests^49^. Significant KS-tests indicate that results do not follow the uniform distribution expected under the null, and therefore were unlikely driven by multiple testing. KS-tests were compared against an alpha of 0.05 divided by 9 when considering the exposure-specific tests, or by 33 when conducting outcome-specific testing, and ranked on their deviation from the uniform distribution.

Furthermore, we independently replicated total effect estimates for LDL-C, HDL-C and TG using GLGC data where lipid concentration was measured using clinical chemistry. Apo-B and Apo-A1 from a genome-wide analysis were analytically replicated using a *cis-*MR approach based on the UCLEB-Kettunen data.

### Software

Analyses were conducted using Python v3.7.4 (for GNU Linux), Pandas v0.25, Numpy v1.15^50^, Seaborn v0.11.5, R v4.0.3^51^ (for GNU Linux), ggforesplot^52^, and Cytoscape v3.8.2 (for GNU Linux).

Results were presented as mean difference (MD, for continuous traits) or odds ratio (OR, for binary traits) with 95% confidence interval (95%CI) for increasing blood lipid or lipoprotein concentration, scaled to one standard deviation (Table S1).

## Results

### Phenotypic and genetic correlation

Aside from an inverse correlation of HDL-C and ApoA1 with TG and VLDL-C concentration, the remaining exposures were positively correlated (Figure S1). Particularly strong correlations were noted between HDL-C and Apo-A1 (correlation coefficient: 0.9); among TC, LDL-C, IDL-C and Apo-B (0.8 to 0.9); between TG and VLDL-C (0.8), and TG with Apo-B, and Rem-Chol (both 0.70). The correlation between the genetic effect estimates followed a similar pattern, albeit with a slightly weaker magnitude, and with VLDL-C and Rem-Chol predominantly correlated among themselves (Figure S1). Please see the supplementary results for a description of the participant characteristics contributing to the sourced NMR data.

### Univariable MR effects of cholesterol-containing lipoproteins on disease incidence and biomarkers

We used univariable IVW MR or MR-Egger (in the presence of directional horizontal pleiotropy) to estimate the total effect of individual cholesterol-containing lipoproteins with risk factors and disease endpoints (Figure 2, and Tables S2-14).

**Figure 2.**
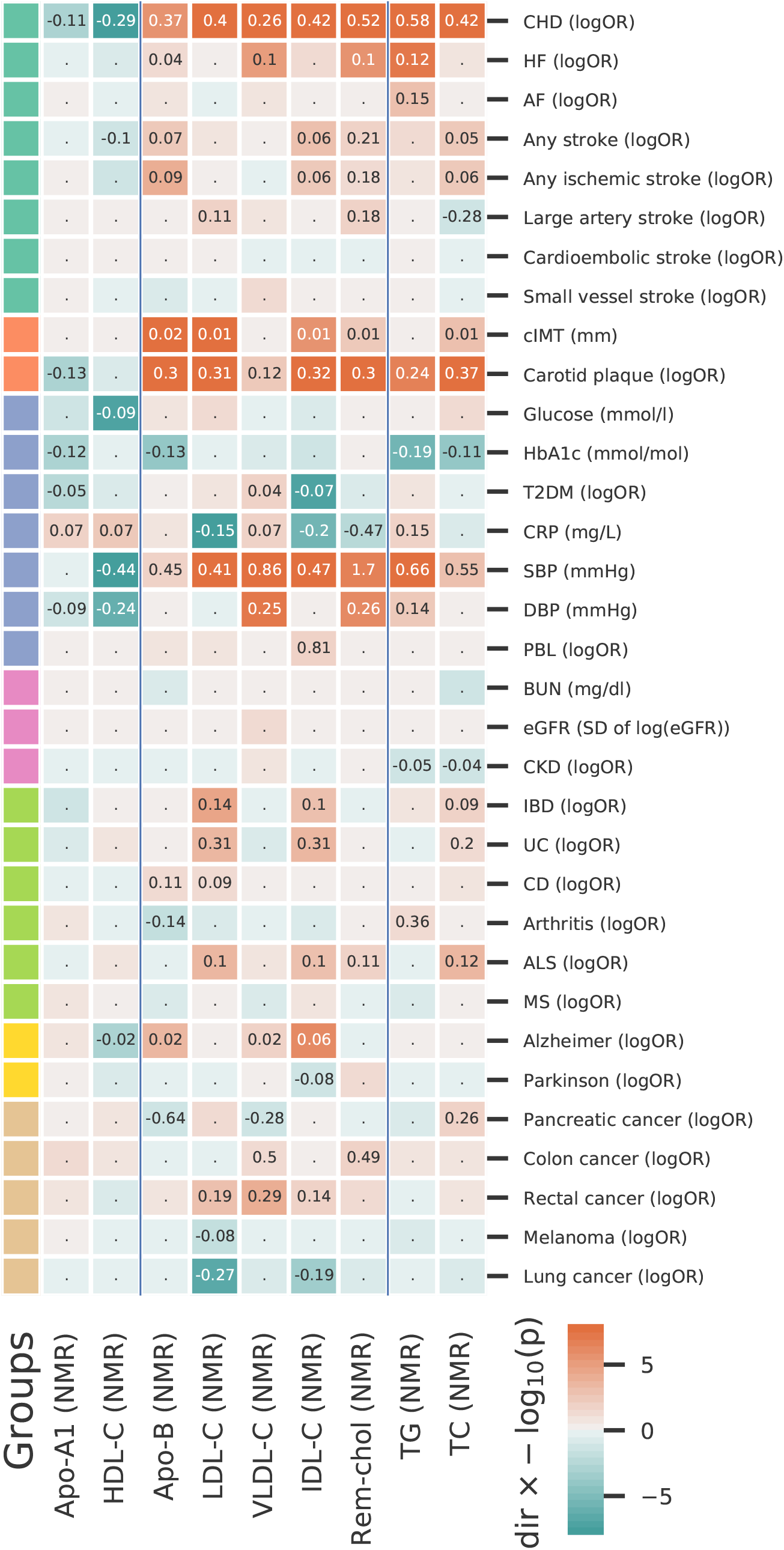
Mendelian randomization estimates of the total effects of a one SD increase in cholesterol-containing lipoprotein and apolipoprotein concentrations. N.b. Cells are coloured by effect direction multiplied by -log_10_(p-value), with the point estimate (the mean difference or log odds ratio) provided for results with p-values smaller than 0.05. The p-values were truncated at 10^−16^ for display purposes. NMR analyses are based on a 33,029 subject meta-analysis of Kettunen^15^ and UCLEB^20^. LDL-C: low-density lipoprotein cholesterol, HDL-C: high-density lipoprotein cholesterol, TG: triglycerides, VLDL-C: very-low-density lipoprotein cholesterol, IDL-C: intermediate-density lipoprotein cholesterol, Rem-chol: remnant-cholesterol, TC: total cholesterol, Apo-B: apolipoprotein-B, Apo-A1: apoliprotein-A1. CHD: coronary heart disease, HF: heart failure, AF: atrial fibrillation, T2DM: type 2 diabetes mellitus, CKD: chronic kidney disease, IBD: inflammatory bowel disease, CD: Crohn’s disease, UC: ulcerative colitis, ALS: Amyotrofe Laterale Sclerose, MS: multiple sclerosis, PBL: primary biliary liver cirrhosis, DBP and SBP: diastolic and systolic blood pressure, CRP: c-reactive protein, HbA1c: glycated haemoglobin; BUN: blood urea nitrogen, eGFR: estimated glomerular filtration rate, cIMT: carotid artery intima media thickness.

Higher concentrations of LDL-C, TC, TG, VLDL-C, IDL-C, and Rem-Chol, were associated with higher CHD risk (OR range: 1.29 to 1.79 per SD), while higher HDL-C cocentration decreased CHD risk; OR 0.75 (95%CI 0.70; 0.80). Heat failure (HF) risk increased with higher concentrations of TG, OR 1.12 (95%CI 1.08; 1.17), VLDL-C, 1.10 (95%CI 1.06; 1.15) and Rem-Chol, OR 1.11 (95%CI 1.06; 1.16), additionally atrial fibrillation (AF) risk increased with TG OR 1.16 (95%CI 1.06; 1.27). The risk of any stroke and ischemic stroke increased with higher concentrations of TC, IDL-C, and Rem-Chol. Higher HDL-C was associated with lower risk of any stroke (Figure 2). Elevated cholesterol-containing lipoproteins were associated with imaging measures of carotid artery atherosclerosis: LDL-C, TC, IDL-C, and Rem-Chol increased cIMT as well as carotid plaque, with the latter trait also affected by higher TG and VLDL-C (Figure 2).

Higher concentration of VLDL-C was associated with increased T2DM risk (OR 1.04 95%CI 1.01; 1.08), while higher IDL-C decreased the risk of T2DM (Figure 2). A one SD higher LDL-C, IDL-C, and Rem-Chol concentration was associated with lower c-reactive protein (CRP) concentration, while higher HDL-C, TG, and VLDL-C were associated with higher CRP concentration. Systolic and/or diastolic blood pressure (SBP, DBP) increased with higher concentrations of LDL-C, TG, TC, VLDL-C, IDL-C, and Rem-Chol, while higher concentrations of HDL-C decreased SBP and DBP (MD range in mmHg: -0.57 to 1.70; Figure 2).

Higher LDL-C concentration was associated with increased the risk of inflammatory bowel disease (IBD, OR 1.15 95%CI 1.07; 1.22), ulcerative colitis (UC, OR 1.37 95%CI 1.15; 1.63), and CD (OR 1.10 95%CI 1.00; 1.20). Higher IDL-C and TC had a similar risk increasing effect on IBD and UC. An SD higher HDL-C decreased Alzheimer’s disease risk (AD, OR 0.98, 95%CI 0.97; 0.99), while AD risk increased with higher concentrations of VLDL-C (OR 1.02, 95%CI 1.00; 1.03) and ILD-C (OR 1.06, 95%CI 1.04; 1.08). Higher LDL-C concentrations decreased risk of lung cancer (OR 0.76, 95%CI 0.69; 0.84) and melanoma (OR 0.92, 95%CI 0.86; 0.99), with the latter also decreased through higher IDL-C. The risk of incident rectal cancer and/or colon cancer were increased by higher concentrations of LDL-C, VLDL-C, IDL-C and Rem-Chol (Figure 2). Finally, we did not observe strong lipoprotein associations with kidney traits, or the smaller stroke sub-types such as cardioembolic stroke. Please see the supplementary results for independent replication of the univariable “total effects” for LDL-C, HDL-C, and TG concentration.

### Univariable MR of Apo-B and Apo-A1 concentration

Higher Apo-B concentration was positively associated with the risk of CHD, (ischemic) stroke, CD, AD, and furthermore increased cIMT, carotid plaque and SBP. Conversely, increased Apo-B concentration was associated with lower HbA1c concentration as well as with pancreatic cancer and arthritis risk (Figure 2). Higher ApoA-1 concentration decreases the risk of CHD, T2DM, carotid plaque, and DBP, while increasing CRP concentrations (Figure 2). Please see the supplementary results for a technical replication using *cis* instruments for Apo-A1 and Apo-B.

### Multivariable MR to identify effects independent of LDL-C, HDL-C and TG

Leveraging NMR data, we applied multivariable MR (MVMR) to investigate whether the above-described causal effect acted independent of the more commonly measured lipids LDL-C, HDL-C, and TG. To do so, for each cholesterol-containing lipoprotein subfraction considered, we derived three MVMR models conditioning on LDL-C, HDL-C or TG, and a fourth model jointly conditioning on all three (Figure S2-S6).

Prioritization on significance and directional concordance (see Methods, Figure 3) resulted in the following *frequency-ranked* list of outcomes with strong support for an independent causal role of cholesterol-containing lipoproteins or/and apolipoproteins: CHD, CRP, SBP, carotid plaque, cIMT, HF, AD, T2DM, HbA1c, IBD, lung cancer, estimated glomerular filtration rate (eGFR), DBP, rectal cancer, CD, UC, glucose concentration, pancreatic cancer, melanoma, primary biliary cirrhosis (PBL) and large artery stroke. Specifically, we observed that higher LDL-C, HDL-C, and TG increased CHD risk independently of one another (Figure 4): OR 1.43 (95%CI 1.34; 1.52) per SD LDL-C, OR 0.76 (95%CI 0.70; 0.80) per HDL-C, and OR 1.15 (95%CI 1.04; 1.28) per SD TG. We also observed effects independent of LDL-C, HDL-C and TG for TC, VLDL-C, IDL-C, Rem-Chol, Apo-B, and Apo-A1. Similarly, CRP was independently affected by LDL-C -0.24 mg/L (95%CI -0.29; -0.19), HDL-C 0.20 mg/L (95%CI 0.14; 0.26) and TG 0.32 mg/L (95%CI 0.25; 0.38), with VLDL-C and Apo-B being associated with higher, and IDL-C and Apo-A1 with lower CRP concentration (Figure 4, Tables S2-10). We also found independent effects for LDL-C, IDL-C, VLDL-C, Rem-Chol, TC, and Apo-B on carotid plaque and cIMT. Furthermore, we found evidence to support independent HF risk increasing effects of VLDL-C OR 1.10 (95%CI 1.02; 1.19), Rem-Chol, Apo-B and TG OR 1.06 (95%CI 1.00; 1.12) (Figure 3 and 5). SBP decreased with higher concentrations of LDL-C, HDL-C, TG, VLDL-C, IDL-C and Rem-Chol (Figures 3 and 5), and AD risk was associated with higher concentration of LDL-C, IDL-C, and Apo-B, while higher HDL-C decreased AD risk: OR 0.97 (95%CI 0.96; 0.98). We also found evidence to support an independent role for VLDL-C increasing T2DM risk OR 1.11 (95%CI 1.04; 1.20), while higher LDL-C (OR 0.90 95%CI 0.88; 0.93) and IDL-C (OR 0.85 95%CI 0.74; 0.97) decreased T2DM risk; Figure 3 and Tables S2-10.

**Figure 3.**
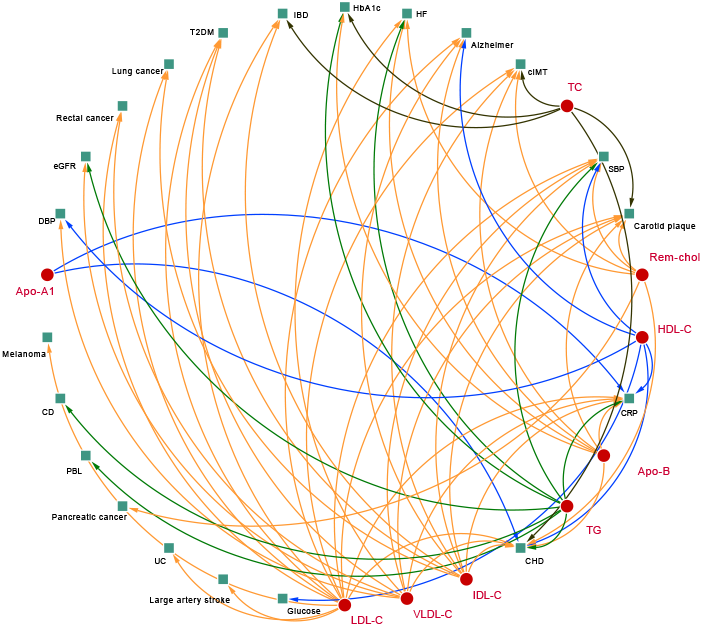
A causal network of phenotypic consequences of higher cholesterol-containing lipoprotein and apolipoprotein blood concentration. N.b. Arcs belonging to the endogenous pathway (VLDL-C, IDL-C, LDL-C, and Apo-B) were coloured *yellow*, arcs for HDL-C and Apo-A1, belonging to the reverse cholesterol transport pathway were depicted in *blue*, TC and TG arcs were represented as black and green, respectively. The network represents highly supported pathways, where the MR effect was significant at an alpha of 0.05 and showed directionally concordant results in at least three of out five potential models (four for LDL-C, HDL-C, and TG): total effects, the direct effects conditional on LDL-C, on HDL-C, on TG, or all three blood lipids.

**Figure 4.**
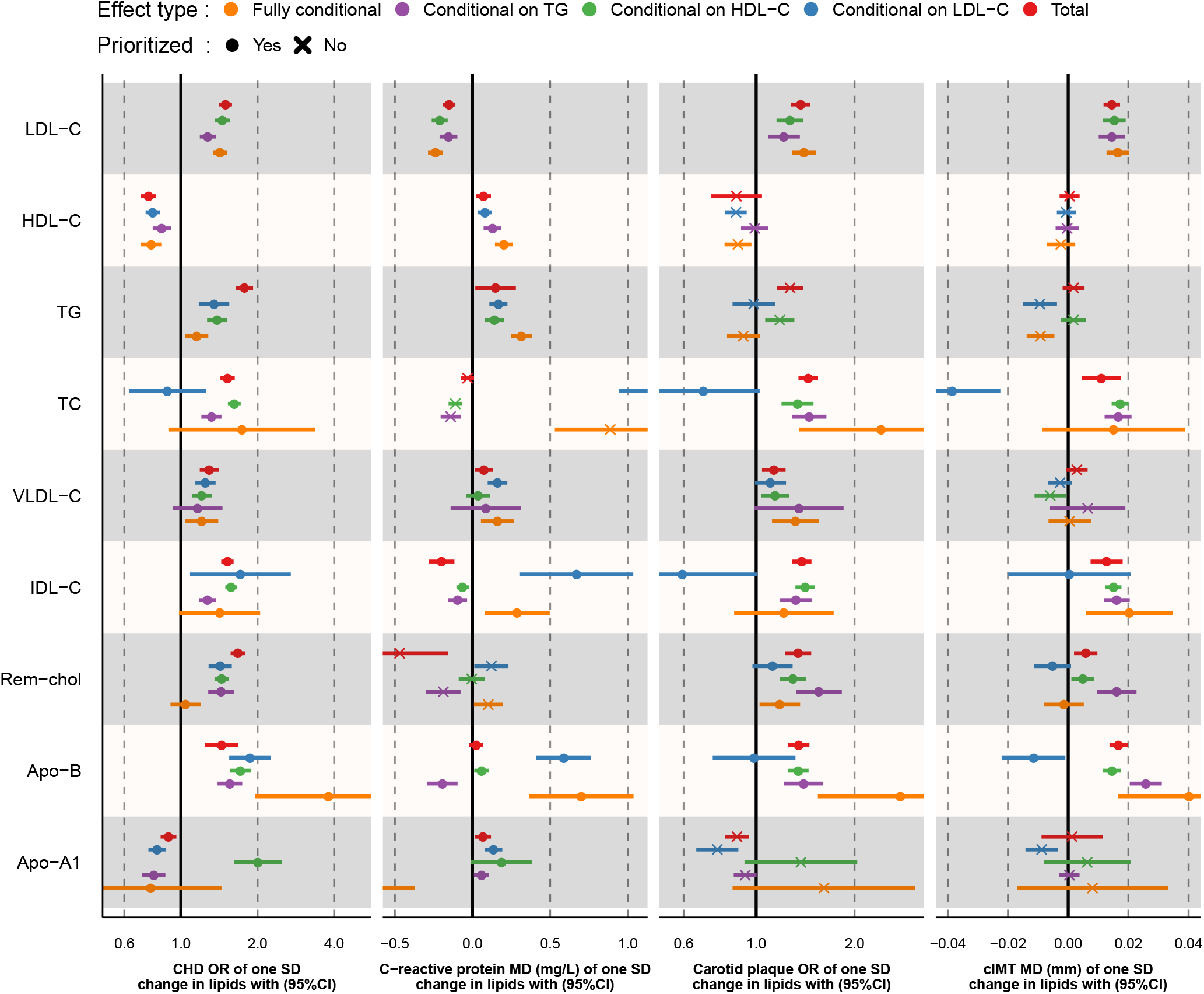
Mendelian randomization effect estimates of a standard deviation change in cholesterol-containing lipoprotein and apolipoprotein concentration on coronary heart disease (CHD), c-reactive protein (CRP), carotid intima media thickness (cIMT), and carotid plaque. N.b. Prioritized results reflect associations depicted in the causal network of Figure 3, where 3 out of 5 (or 4 for LDL-C, HDL-C, and TG) estimates were significant at an alpha of 0.05 and directionally concordant. Total: the total lipid effect, Conditional effects either, represent the blood lipid effect of LDL-C, HDL-C or TG singularly, or off all three blood lipids in a single multivariable MR (fully adjusted) model. Fully adjusted models for LDL-C, HDL-C, or TG exposures only conditioned on two of the three blood lipids (e.g., the fully conditional model for LDL-C exposure only conditioned on HDL-C and TG). Estimates are provided as odds ratio (OR) or mean difference (MD) with 95% confidence intervals (95%CI).

**Figure 5.**
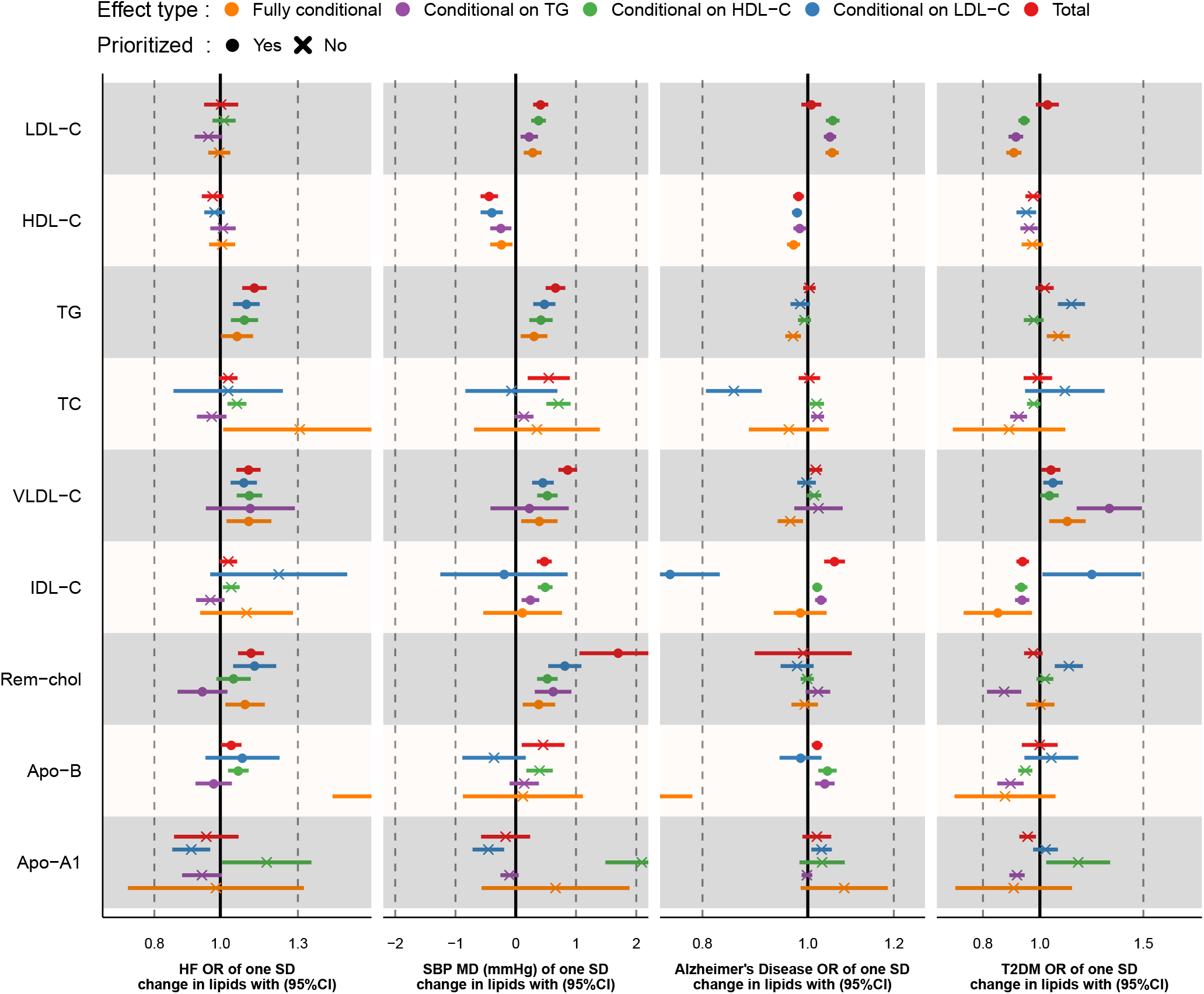
Mendelian randomization effect estimates of a standard deviation change in cholesterol-containing lipoprotein and apolipoprotein concentration on heart failure (HF), systolic blood pressure (SBP), Alzheimer’s disease (AD), and type 2 diabetes (T2DM). N.b. Prioritized results reflect associations depicted in the causal network of Figure 3, where 3 out of 5 (or 4 for LDL-C, HDL-C, and TG) estimates were significant at an alpha of 0.05 and directionally concordant. Total: the total lipid effect, Conditional effects either, represent the blood lipid effect of LDL-C, HDL-C or TG singularly, or off all three blood lipids in a single multivariable MR (fully adjusted) model. Fully adjusted models for LDL-C, HDL-C, or TG exposures only conditioned on two of the three blood lipids (e.g., the fully conditional model for LDL-C exposure only conditioned on HDL-C and TG). Estimates are provided as odds ratio (OR) or mean difference (MD) with 95% confidence intervals (95%CI).

### Further prioritization: assessing the overall null-hypothesis

Under the null-hypothesis the p-values of a group of tests follow a standard continuous uniform distribution between zero and one^49^. Hence to assess to what extend the described results were driven by multiple testing we use Kolmogorov-Smirnov tests (KS-tests) to compare empirical p-values distributions against a uniform distribution (Figure 6). Additionally, KS-tests naturally rank results by their joint evidence against a null-hypothesis of no-effect, providing a further prioritization metric.

**Figure 6.**
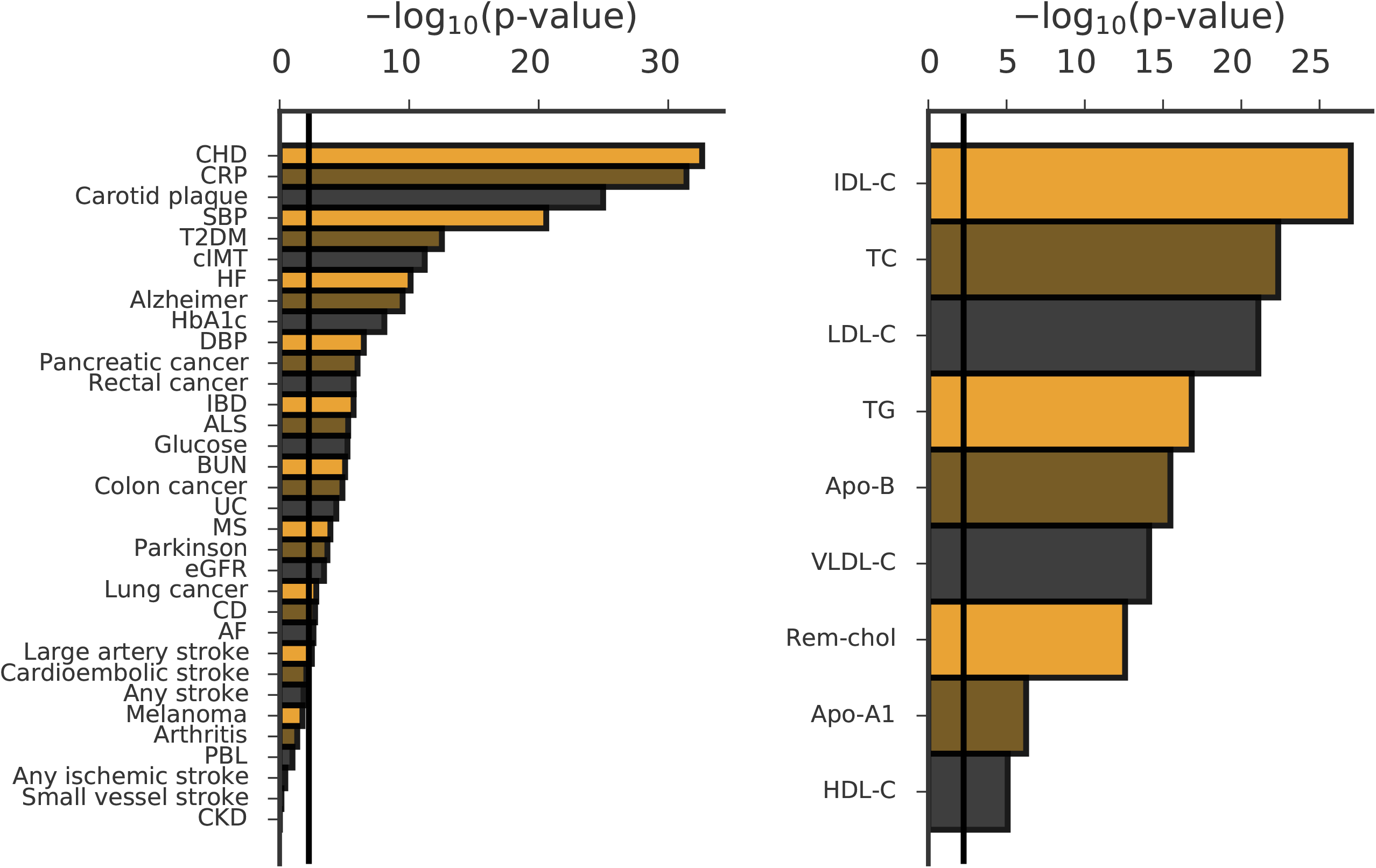
Kolmogorov-Smirnov overall-null hypothesis tests. N.b. Kolmogorov-Smirnov goodness-off-fit tests were used to compare empirical p-value distribution against the continuous uniform p-value distribution expected when the strict null-hypothesis holds. The **top** panel, grouped empirical p-values by exposure and explored whether their distribution agreed with the expected p-value distribution when all test would be false-positive. The **bottom** panel, grouped empirical p-values by outcome and explored whether their distribution agreed with the expected p-value distribution when all test would be false-positive. The horizontal lines represent the multiplicity corrected p-value threshold.

Grouping p-values by exposure indicated that none of our results could readily be explained by multiple testing alone and ranked IDL-C as the relatively most influential cholesterol-containing lipoprotein, with HDL-C providing relatively less evidence against the overall null-hypothesis. Aggregating results by outcome resulted in similar ranking obtained by simply counting the number of exposures affecting a single trait (Figure 3; i.e., the indegree of a digraph), indicating cholesterol-containing lipoprotein and apolipoproteins particularly affect CHD, CRP, Carotid plaque, cIMT, SBP, HF, T2DM, and AD (Figure 6). Additionally, this analysis revealed there was little evidence to support involvement of cholesterol-containing lipoprotein and apolipoprotein concentrations with CKD, ischemic and small vessel stroke, PBL, and Arthritis.

## Discussion

We used Mendelian randomization (MR) to catalogue, and prioritize, the biomedical consequences of elevated concentrations of cholesterol-containing lipoproteins beyond LDL-C, HDL-C, and total triglycerides (TG), including remnant cholesterol, IDL-C and VLDL-C, as well as apolipoproteins A1 and B. Major findings include that CHD is affected by all of the major cholesterol-rich lipoproteins including HDL-C, IDL-C, VLDL-C, Rem-Chol as well as apolipoproteins A1 and B, and TG. Similar ubiquitous effects were observed for cIMT, and carotid plaque, suggesting the observed CHD effects are partially mediated through atherosclerosis. Additionally, we found strong evidence linking higher concentrations of TG, VLDL-C, Apo-B, and Rem-Chol to increased HF risk. Cholesterol-containing lipoproteins, apolipoproteins, as well triglycerides also affected non-CVD traits such as T2DM, CRP, IBD, and AD. Multivariable MR was used to confirm many of these associations act independently of the three widely measured lipid subfractions: LDL-C, HDL-C, and TG.

There has been considerable debate on higher HDL-C potentially reducing CHD risk. With an imprecise OR estimate of 0.93 per SD (95% CI 0·68;1·26) by Voight *et al*^53^ often cited as definitively proving that HDL-C does not affect CHD risk. We note however that our estimate OR 0.75 per SD (95%CI 0.70; 0.80) falls almost completely within the 95%CI provided by Voight *et al*. Hence our results, suggesting a protective CHD effect of higher HDL-C concentration, are fully supported by previous findings. The major difference here is the added precision offered by the available larger sample size data (12K CHD cases by Voight *et al*. vs 60K in the current paper). More recently MVMR conditioning on HDL-C, Apo-A1 and Apo-B, failed to show an independent HDL-C effect on CHD, with suggestions that the univariable protective CHD effect of HDL-C (OR 0.80, 95%CI 0.77; 0.89) was attributable to Apo-B^54^. Based on the understanding of lipid metabolism however, it seems unlikely that HDL-C asserts its CHD effect through Apo-B. Similarly, the absence of a strong correlation between Apo-B and HDL-C does not suggest there is any clear potential for horizontal pleiotropy that might be explained by Apo-B (Figure S1). Instead, the lack of significant HDL-C after conditioning on Apo-B and Apo-A1, likely reflects model instability due to including both HDL-C and Apo-A1. Using independent data and a stepwise analysis, we confirmed the presence of such model instability, observing a decrease in precision when including HDL-C and Apo-A1, and a flipped effect direction erroneously suggesting that higher Apo-A1 (conditional on HDL-C) increases CHD risk. Through the previously described heuristic approach, we discounted such clearly problematic models, and looked for consistency in a majority vote of at least 3 significant and directionally concordant models (Figure 4), resulting in the above mentioned protective effect of HDL-C on CHD.

Our analysis supports the proposition by Ference, Kastelijn and Catapano^55^ that Apo-B might play a central role in lipid lowering therapies intended to decrease CHD risk. Apo-B (specifically Apo-B 100) is carried by VLDL, IDL and LDL particles and as such provides a summary measure of these lipoproteins. Our finding that Apo-B, VLDL-C, IDL-C, LDL-C and Rem-Chol (a summary of IDL-C and VLDL-C) all increase CHD risk suggests that possibly the entire endogenous pathway is relevant for CHD. This is further supported by our observation that higher concentrations of these same subfractions increase carotid plaque risk and cIMT. Given that Apo-B is carried on top of the major endogenous lipoproteins and does not naturally occur in isolation off these lipoporteins^56^, the effects of Apo-B conditional on LDL-C may be less relevant. We therefore repeated our MVMR analyses conditioning exclusively on Apo-B, TG and HDL-C (Table S13), confirming independent CHD effects of both Apo-B and HDL-C: OR 1.81 (95%CI 1.64; 1.99), OR 0.80 (95%CI 0.74; 0.86).

Rem-Chol provides a summary measure of the cholesterol content on IDL and VLDL particles which have been proposed as additional atherogenic lipid fractions^57^. We now provide empirical proof showing that conditional on LDL-C (as well as HDL-C and TG) Rem-Chol, and it’s constituents VLDL-C and IDL-C independently increases the risk of CHD, HF, ischemic strokes and cIMT and carotid plaque. These findings suggest that that IDL-C and VLDL-C may provide additional, LDL-C independent, therapeutic avenues.

In general, we showed that many of the cholesterol-containing lipoproteins acted independently of TG, suggesting that these effects are at least partially cholesterol mediated. By specifically conditioning on TG, that is the total triglyceride content aggregated across all lipid particles, we attempted to account for all sources of triglyceride at the cost of implicitly assuming a shared effect direction irrespective of the lipoprotein triglyceride carrier. This may not always hold, and further studies could consider TG effects per subfraction. Nevertheless, the joint consideration of TG provides valuable insights, finding that conditional on LDL-C and HDL-C higher concentrations of TG increase the risk of CHD, HF, PBL, CD, as well as increasing CRP concentration, SBP, carotid plaque and cIMT (Figure 3). Importantly, the observation that higher TG concentration increased CHD risk independently of LDL-C suggests that the ANGPTL3-inhibitor evinacumab may decrease CHD risk jointly through lowering TG and LDL-C, instead of exclusively through LDL-C^58^.

While the considered cholesterol-containing lipoprotein and apolipoproteins had a predominant cardiac and atherosclerotic fingerprint, we found that specific subfractions affected non-CVD diseases. For example, higher LDL-C concentrations decreased T2DM risk, and CRP concentrations, while increasing risk of AD, UC, IBD, and SBP. IDL-C showed comparable effect, also affecting SBP, CRPAD, T2DM, and IBD in the same direction. Higher VLDL-C concentration increased CRP, and T2DM risk. HDL-C (and to a lesser extent Apo-A1) typically acted in opposite direction of the endogenous pathway blood lipids, with higher concentrations protecting against CHD, AD, decreasing SBP and DBP, and increasing CRP concentrations as well as T2DM risk. We observed some suggestion that LDL-C, IDL-C and VLDL-C might be related to cancer incidence, with for example both LDL-C and VLDL-C increasing rectal cancer risk (OR 1.21 95%CI 1.08; 1.35 for LDL-C and OR 1.34, 95%CI 1.15; 1.56 for VLDL-C); Figures 2-3.

Randomized controlled trials of statins, which primarily affect LDL-C, are known to reduce cIMT^59^. Our results now suggest this favourable effect might be (partially) mediated through lipid fraction perturbation, and possibly shared by other lipid related drug targets. Recently we showed that lower plasma concentrations of CETP and PCSK9, both lipid related drug targets, are associated with decreases of cIMT, supporting the lipid mediation proposition^11^. The observed CRP decreasing effect of higher LDL-C seems at odds with previous finding that statins (targeting HMGCR) and ezetimibe (targeting NCP1L1) reduce LDL-C as well as CRP^60,61^. If instead we consider the entire endogenous pathway, we find that higher concentration of Apo-B and VLDL-C increase CRP levels (Figure 4) matching the reported drug compound effect direction, where statins and to a lesser extent, ezetimibe are known to decrease Apo-B and VLDL-C. We and others^11,62–65^ have previously reported that the lipid drug targets HMGCR (statins), NCP1L1 (for ezetimibe) and PCSK9 increase T2DM risk. In the current study we found robust evidence that is likely mediated by changes in LDL-C, and IDL-C, where IDL-C acts independently of LDL-C.

This study has employed MR to determine two types of effects 1) the total effect which consists of a direct and indirect effect (where both, or either could be zero), and 2) the direct effect accounting for any potential mediation by LDL-C, HDL-C, and TG (Figure 1). Both the total and direct effects are valid causal effects, and the absence of a direct effect should not be interpreted as disqualifying any observed total effect, or vice versa. We specifically utilized MVMR to explore to what extent the observed effect acted independently from the thoroughly studied exposures LDL-C, TG, or HDL-C. It is important to highlight that our genetics instruments were selected on F-statistic > 24 which protects against weak instrument bias. Because MVMR performs a conditional analysis it becomes relevant to also consider conditional F-statistics (Supplementary Table 15), which suggest that MVMR models jointly accounting for LDL-C, HDL-C, and TG, were especially vulnerable conditional weak-instruments. Our analysis was designed to ensure that any weak-instrument bias would act towards a null effect, resulting in conservative findings. Specifically, analyses were conducted in a two-sample setting, and MVMR-Egger was employed to protect against any potential horizontal pleiotropy not captured by MVMR. While this minimizes the false-positive rate, this also implies (even more than usual) that we should not overinterpret non-significant findings as proof of a null-effect^66^.

Given the high phenotypic and genetic correlation (Figure S1) among the considered exposures, and the multiple *nested* models (one total effect and four multivariable direct effect models), straightforward control of the family-wise error or of the false discovery rate^49^ would be overly conservative and likely greatly inflate type 2 error rates. Instead, we addressed multiple testing in several complementary ways. First, we leveraged independent validation data for LDL-C, HDL-C, and TG (from GLGC), showing general concordance. Furthermore, the apolipoprotein results were validated using *cis* acting instruments. While the Apo-A1 *cis-*MR results were limited by the availability of only 2 instruments, the Apo-B *cis-*MR generally agreed with the genome-wide MR results for Apo-B, supporting the robustness of our analytical approach - combining model selection techniques with removal of variant with relatively larger outlier or leverage statics. KS-test were used to assess overall null-hypothesis that results were false-positive, which we could strongly reject (Figure 6). Furthermore, when considering multivariable results, we discounted directionally discordance results, more likely to be false positives.

Due to the considerable correlation between lipid subfractions, we focussed on total cholesterol-content of each lipoprotein fraction instead of subdivisions by particle sizes. Nevertheless, considerations of particle size are important when interpreting the results. For example, it is likely that the atherogenic VLDL-C affects are predominantly driven by the smaller VLDL-C particles that may penetrate endothelial tissue^55^. The CHD effect of HDL-C may similarly be size specific, with studies suggesting the large HDL-C particles are particularly protective against CHD occurrence^67,68^.

In conclusion, we have catalogued and prioritized the phenotypic consequences of cholesterol-containing lipoprotein and apolipoprotein blood concentrations, finding that many of these exposures act independently of the commonly measured blood lipids: LDL-C, HDL-C and TG. We found evidence that CHD and related traits, such as cIMT, carotid plaque, CRP, and blood pressure, are causally affected by most lipid fractions typically including LDL-C, HDL-C, VLDL-C, IDL-C, TG, and apolipoproteins B and A1. Furthermore, we found that higher concentrations of TG, VLDL-C, Rem-Chol and Apo-B increased HF risk. Our analyses additionally identified certain non-CVD traits that are more exclusively affected by smaller subset of exposures, such as Alzheimer’s disease (HDL-C, LDL-C, IDL-C, Apo-B), IBD (LDL-C, IDL-C), and T2DM (VLDL-C, IDL-C and LDL-C).

## Data Availability

Summary genetic effect estimates for outcomes were extracted from publicly accessible GWAS on glucose and HbA1c, and C-reactive protein (all from the UKB (nealelab.is/uk-biobank; removing low confidence variants), as well as blood pressure (systolic and diastolic), available from Evangelou et al.23. The CKDGen consortium provided GWAS associations on blood urea nitrogen, estimated glomerular filtration rate, and chronic kidney disease24. Genetic associations with primary biliary cirrhosis were available from Jostins et al25. A meta-analysis of CHARGE26 and UCLEB20 provided genetic associations with carotid artery intima media thickness and plaque. CHD data were available for 42,335 cases from CardiogramplusC4D27; 40,585 stroke cases (including four subtypes) from MEGASTROKE28; 47,309 heart failure cases from HERMES29, 60620 atrial fibrillation cases from Nielson et al30, 74,124 type 2 diabetes31 cases from DIAGRAM, 32,637 cases of inflammatory bowel disease32, 5,956 cases of Crohn's disease33 and 6,687 cases of ulcerative colitis34 from IIBDGC, 29880 rheumatoid arthritis cases from Okada et al35, 14,498 cases of multiple sclerosis36 from the IMSG consortium, 15156 amyotrophic lateral sclerosis cases from Rheenen et al37, 71,880 cases of Alzheimer's disease from Jansen et al38, and 56,306 cases of Parkinson's disease from Nalls et al.39. Finally, we sourced data on pancreatic cancer, colon cancer, rectal cancer, lung cancer and melanoma from Rashkin et al40. Please contact AFS for access to analysis scripts, processed analyses tables are presented in the supplemental, and crude pickled results file have been deposited at XXX.

## Author’s contributions

AFS and ADH, CF contributed to the idea and design of the study. AFS performed the analyses. AFS drafted the manuscript. All authors provided critical input on the analyses and the drafted manuscript.

## Guarantor

Amand F Schmidt performed the here presented analyses, had full access to all the data in the study and takes responsibility for the integrity of the data and the accuracy of the data analysis.

## Acknowledgement

This research has been conducted using the UK Biobank Resource under Application Number 12113. The authors are grateful to UK Biobank participants. UK Biobank was established by the Wellcome Trust medical charity, Medical Research Council, Department of Health, Scottish Government, and the Northwest Regional Development Agency. It has also had funding from the Welsh Assembly Government and the British Heart Foundation.

## Prior postings and presentations

A pre-print version of this manuscript has been deposited at XXX.

## Data availability

Summary genetic effect estimates for outcomes were extracted from publicly accessible GWAS on glucose and HbA1c, and C-reactive protein (all from the UKB (nealelab.is/uk-biobank; removing low confidence variants), as well as blood pressure (systolic and diastolic), available from Evangelou *et al*.^23^. The CKDGen consortium provided GWAS associations on blood urea nitrogen, estimated glomerular filtration rate, and chronic kidney disease^24^. Genetic associations with primary biliary cirrhosis were available from Jostins *et al*^25^. A meta-analysis of CHARGE^26^ and UCLEB^20^ provided genetic associations with carotid artery intima media thickness and plaque. CHD data were available for 42,335 cases from CardiogramplusC4D^27^; 40,585 stroke cases (including four subtypes) from MEGASTROKE^28^; 47,309 heart failure cases from HERMES^29^, 60620 atrial fibrillation cases from Nielson *et al*^30^, 74,124 type 2 diabetes^31^ cases from DIAGRAM, 32,637 cases of inflammatory bowel disease^32^, 5,956 cases of Crohn’s disease^33^ and 6,687 cases of ulcerative colitis^34^ from IIBDGC, 29880 rheumatoid arthritis cases from Okada *et al*^35^, 14,498 cases of multiple sclerosis^36^ from the IMSG consortium, 15156 amyotrophic lateral sclerosis cases from Rheenen *et al*^37^, 71,880 cases of Alzheimer’s disease from Jansen *et al*^38^, and 56,306 cases of Parkinson’s disease from Nalls *et al*.^39^. Finally, we sourced data on pancreatic cancer, colon cancer, rectal cancer, lung cancer and melanoma from Rashkin *et al*^40^. Please contact AFS for access to analysis scripts, processed analyses tables are presented in the supplemental, and crude pickled results file have been deposited at XXX.

