## Supplementary for "Biomedical consequences of elevated cholesterol-containing lipoproteins and apolipoproteins"

### Biomedical consequences of elevated blood lipoproteins and apolipoprotein concentration

*Blood lipoproteins and apolipoprotein*

***A F Schmidt et al***

### *Contents*

*Figures* 2

*Tables* 8

*Supplementary results* 275

*Acknowledgements* 277

*Bibliography* 278

### *Figures*

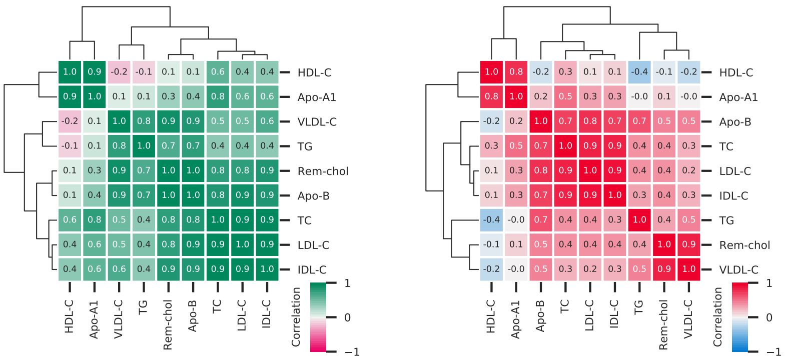


**Figure S1:** Spearman's pair-wise correlation between the phenotypic NMR measured blood lipids (*left*: based on a n=14,834 UCLEB[1] sample) and between the genetic association with these blood lipids (*right*: based on a n=33,029 meta-analysis of UCLEB and Kettunen[2]); the margin order was based on hierarchical clustering of the Euclidean distance.

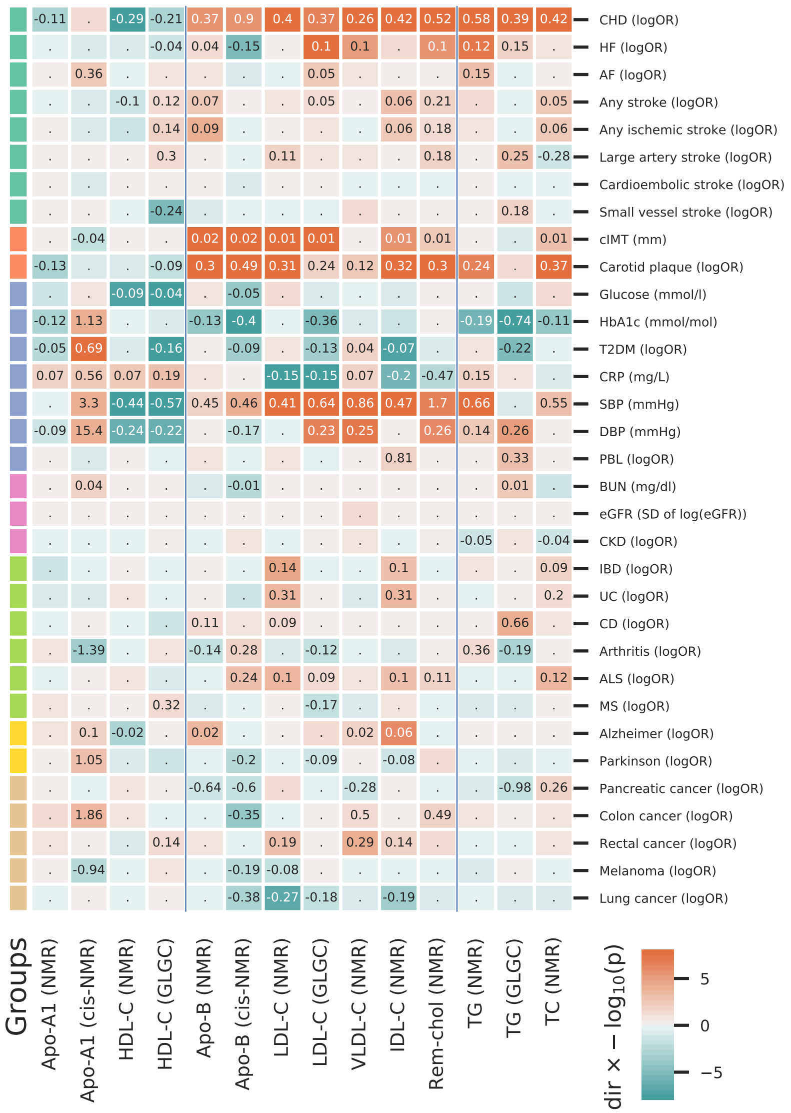


**Figure S2:** Mendelian randomization estimates of the total effects of a one SD increase in cholesterol-containing lipoprotein and apolipoprotein concentration. With independent replication data from the GLGC GWAS [3], and technical replication using *cis* MR analysis of Apo-A1 and Apo-B concentrations.

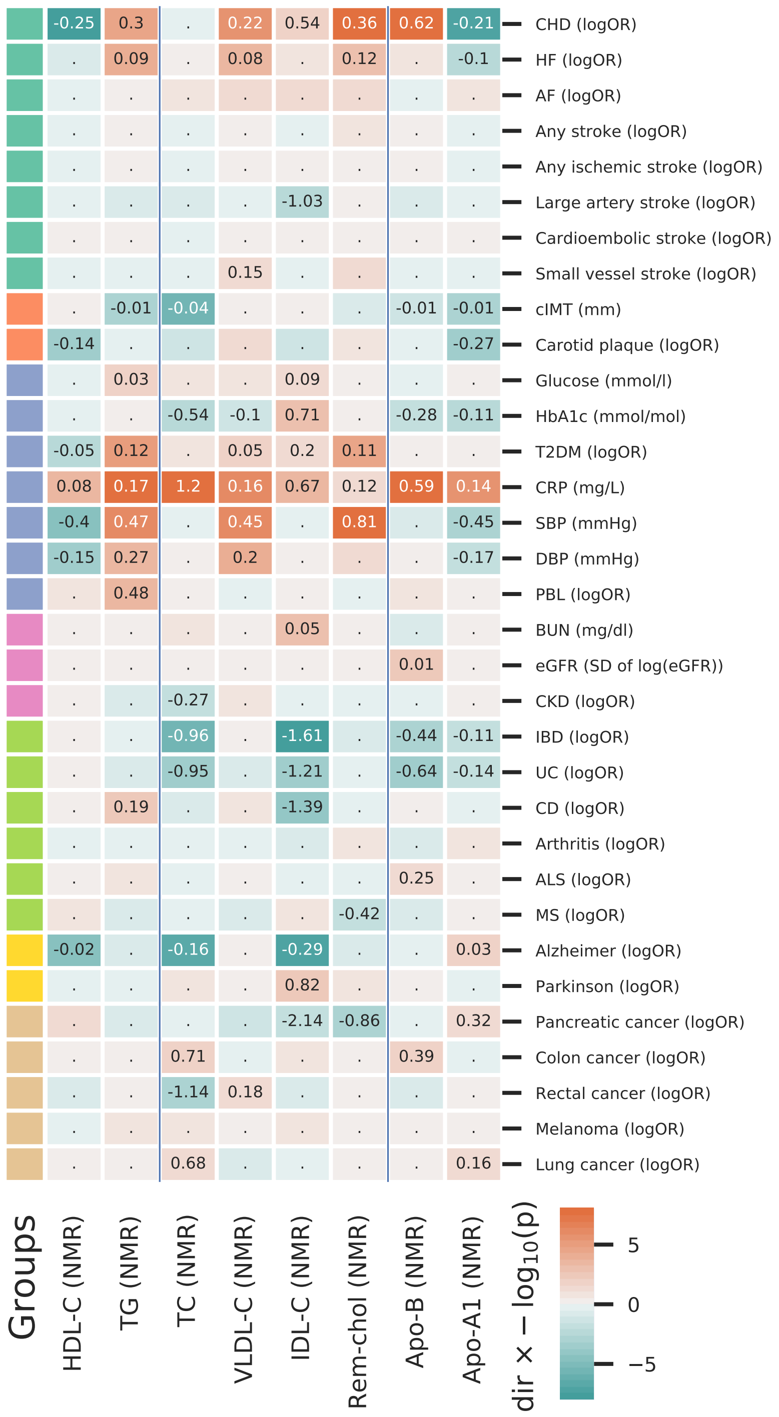


**Figure S3:** Multivariable Mendelian randomization estimates of the direct pathway effect of one SD change in blood lipid, conditional on LDL-C.

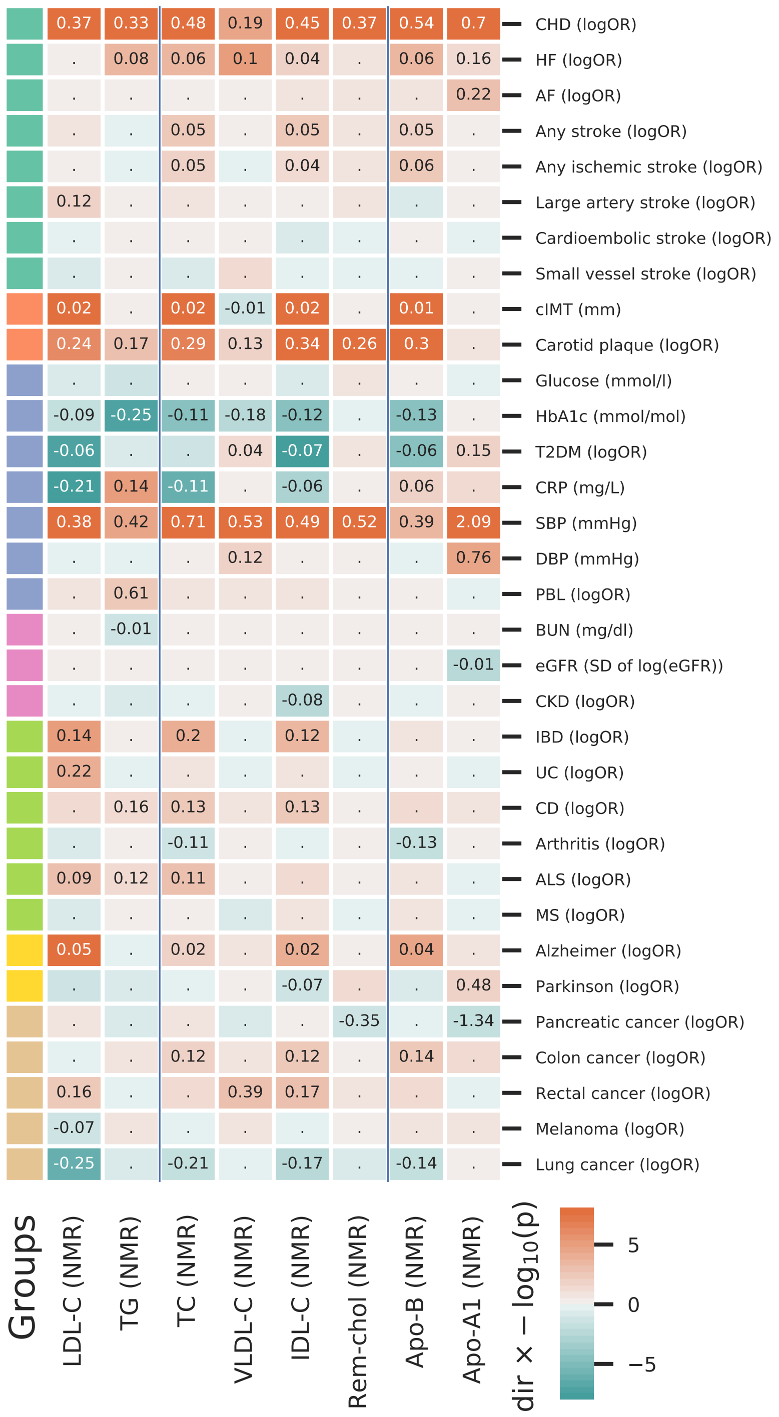


**Figure S4:** Multivariable Mendelian randomization estimates of the direct pathway effect of one SD change in blood lipid, conditional on HDL-C.

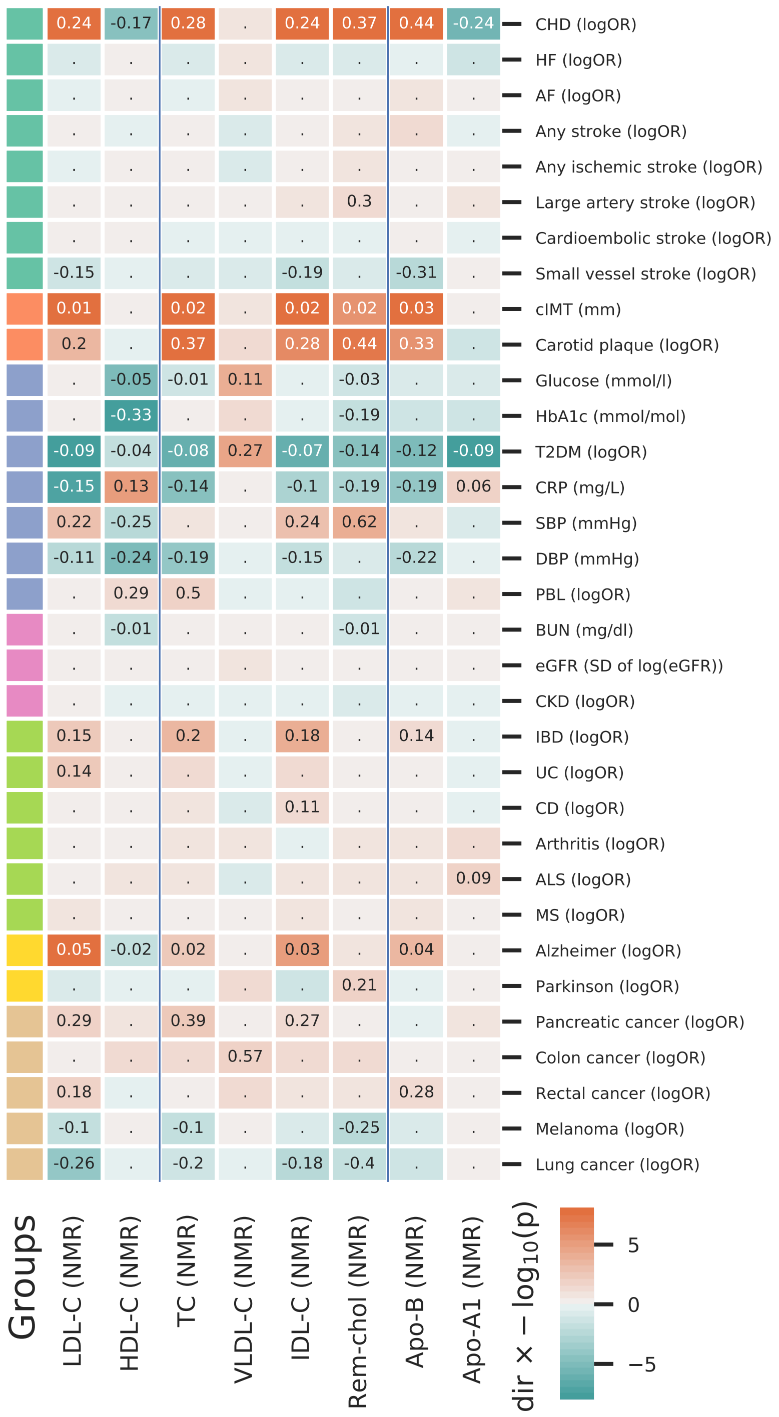


**Figure S5:** Multivariable Mendelian randomization estimates of the direct pathway effect of one SD change in blood lipid, conditional on Triglycerides.

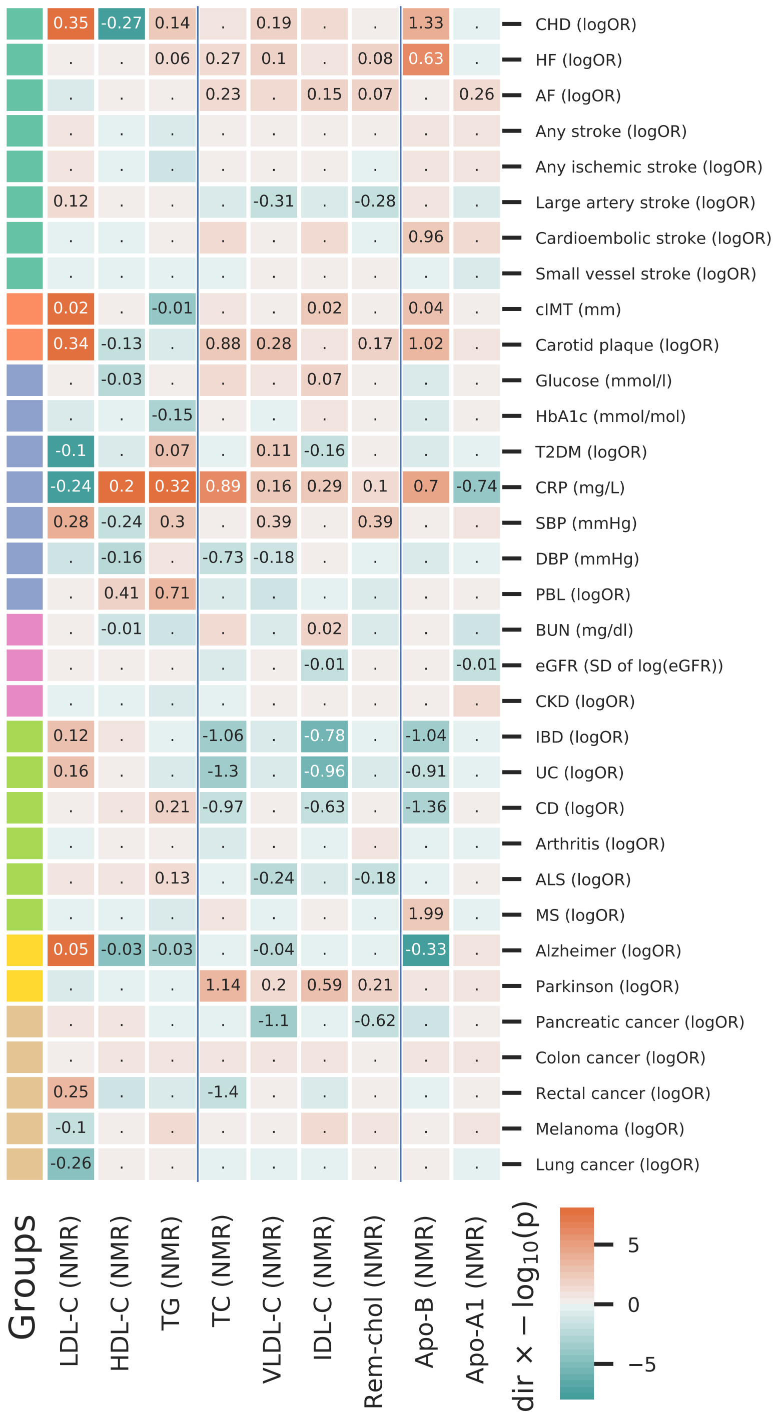


**Figure S6:** Multivariable Mendelian randomization estimates of the direct pathway effect of one SD change in blood lipid, (fully) conditional on LDL-C, HDL-C, and Triglycerides.

### Tables

**Table S1:** The standard deviation of NMR measured blood lipids in mmol/L (total cholesterol, triglycerides, lipoproteins) or g/L (apolipoproteins)

| **Blood lipid** | **Standard deviation** |
|---|---|
| LDL-C | 0.646 |
| HDL-C | 0.416 |
| TG | 0.613 |
| VLDL-C | 0.282 |
| IDL-C | 0.232 |
| Rem-chol | 0.461 |
| Chol | 1.238 |
| Apo-B | 0.238 |
| Apo-A1 | 0.276 |

**Table S2:** (Multivariable) Mendelian randomization effect estimates for one standard deviation change in LDL-C

| Outcome | Effect type | Point estimate (95%CI) | P-value | Estimator | Heterogeneity P-value | Precision | No. variants |
|---|---|---|---|---|---|---|---|
| CHD (OR) | Total | 1.50 (1.41; 1.59) | < 0.001 | IVW | < 0.001 | $1.1\times10^3$ | 25 |
| CHD (OR) | HDL-C independent | 1.45 (1.36; 1.56) | < 0.001 | MR Egger | < 0.001 | $8.1\times10^2$ | 50 |
| CHD (OR) | TG independent | 1.28 (1.19; 1.37) | < 0.001 | MR Egger | < 0.001 | $7.4\times10^2$ | 45 |
| CHD (OR) | HDL-C & TG independent | 1.43 (1.34; 1.52) | < 0.001 | IVW | < 0.001 | $9.5\times10^2$ | 57 |
| HF (OR) | Total | 1.00 (0.95; 1.06) | 0.926 | MR Egger | < 0.001 | $1.2\times10^3$ | 111 |
| HF (OR) | HDL-C independent | 1.01 (0.97; 1.05) | 0.530 | MR Egger | < 0.001 | $2.5\times10^3$ | 185 |
| HF (OR) | TG independent | 0.96 (0.92; 1.01) | 0.090 | MR Egger | < 0.001 | $1.8\times10^3$ | 162 |
| HF (OR) | HDL-C & TG independent | 1.00 (0.96; 1.03) | 0.848 | IVW | < 0.001 | $2.8\times10^3$ | 216 |

**Table S2:** (Multivariable) Mendelian randomization effect estimates for one standard deviation change in LDL-C *(continued)*

| Outcome | Effect type | Point estimate (95%CI) | P-value | Estimator | Heterogeneity P-value | Precision | No. variants |
|---|---|---|---|---|---|---|---|
| AF (OR) | Total | 0.99 (0.96; 1.02) | 0.476 | IVW | 0.080 | $5.0\times10^3$ | 121 |
| AF (OR) | HDL-C independent | 1.00 (0.97; 1.02) | 0.875 | IVW | 0.067 | $5.5\times10^3$ | 202 |
| AF (OR) | TG independent | 0.98 (0.95; 1.01) | 0.285 | IVW | < 0.001 | $4.2\times10^3$ | 181 |
| AF (OR) | HDL-C & TG independent | 0.98 (0.95; 1.01) | 0.178 | IVW | < 0.001 | $4.0\times10^3$ | 238 |
| Any stroke (OR) | Total | 1.04 (0.97; 1.12) | 0.261 | MR Egger | 0.004 | $7.5\times10^2$ | 112 |
| Any stroke (OR) | HDL-C independent | 1.03 (0.99; 1.07) | 0.106 | IVW | < 0.001 | $3.1\times10^3$ | 192 |
| Any stroke (OR) | TG independent | 1.01 (0.95; 1.07) | 0.722 | MR Egger | < 0.001 | $1.2\times10^3$ | 167 |
| Any stroke (OR) | HDL-C & TG independent | 1.04 (0.99; 1.09) | 0.089 | IVW | < 0.001 | $1.9\times10^3$ | 228 |
| Any ischemic stroke (OR) | Total | 1.02 (0.94; 1.11) | 0.646 | MR Egger | 0.067 | $5.3\times10^2$ | 111 |
| Any ischemic stroke (OR) | HDL-C independent | 1.02 (0.98; 1.06) | 0.387 | IVW | < 0.001 | $2.5\times10^3$ | 189 |
| Any ischemic stroke (OR) | TG independent | 0.99 (0.93; 1.06) | 0.843 | MR Egger | 0.096 | $8.7\times10^2$ | 165 |
| Any ischemic stroke (OR) | HDL-C & TG independent | 1.03 (0.98; 1.08) | 0.206 | IVW | 0.010 | $1.6\times10^3$ | 226 |
| Large artery stroke (OR) | Total | 1.12 (1.02; 1.23) | 0.018 | IVW | < 0.001 | $4.2\times10^2$ | 113 |
| Large artery stroke (OR) | HDL-C independent | 1.12 (1.02; 1.23) | 0.015 | IVW | < 0.001 | $4.4\times10^2$ | 192 |
| Large artery stroke (OR) | TG independent | 1.02 (0.88; 1.19) | 0.768 | MR Egger | < 0.001 | $1.6\times10^2$ | 168 |
| Large artery stroke (OR) | HDL-C & TG independent | 1.13 (1.00; 1.27) | 0.046 | IVW | < 0.001 | $2.7\times10^2$ | 229 |
| Cardioembolic stroke (OR) | Total | 1.00 (0.93; 1.08) | 0.994 | IVW | 0.026 | $6.9\times10^2$ | 114 |
| Cardioembolic stroke (OR) | HDL-C independent | 0.98 (0.91; 1.06) | 0.650 | IVW | 0.039 | $6.8\times10^2$ | 191 |
| Cardioembolic stroke (OR) | TG independent | 1.02 (0.89; 1.17) | 0.775 | MR Egger | 0.054 | $2.2\times10^2$ | 167 |
| Cardioembolic stroke (OR) | HDL-C & TG independent | 0.99 (0.90; 1.08) | 0.769 | IVW | 0.041 | $4.4\times10^2$ | 228 |
| Small vessel stroke (OR) | Total | 0.99 (0.90; 1.08) | 0.758 | IVW | 0.070 | $4.2\times10^2$ | 116 |
| Small vessel stroke (OR) | HDL-C independent | 0.95 (0.87; 1.04) | 0.254 | IVW | 0.008 | $5.1\times10^2$ | 195 |
| Small vessel stroke (OR) | TG independent | 0.86 (0.75; 0.99) | 0.042 | MR Egger | 0.006 | $1.9\times10^2$ | 170 |
| Small vessel stroke (OR) | HDL-C & TG independent | 0.97 (0.87; 1.08) | 0.572 | IVW | 0.007 | $3.2\times10^2$ | 232 |
| cIMT (mm) | Total | 0.01 (0.01; 0.02) | < 0.001 | IVW | < 0.001 | $5.1\times10^5$ | 116 |
| cIMT (mm) | HDL-C independent | 0.02 (0.01; 0.02) | < 0.001 | MR Egger | < 0.001 | $2.7\times10^5$ | 193 |
| cIMT (mm) | TG independent | 0.01 (0.01; 0.02) | < 0.001 | MR Egger | < 0.001 | $2.0\times10^5$ | 172 |
| cIMT (mm) | HDL-C & TG independent | 0.02 (0.01; 0.02) | < 0.001 | MR Egger | < 0.001 | $2.7\times10^5$ | 234 |
| Carotid plaque (OR) | Total | 1.37 (1.28; 1.46) | < 0.001 | IVW | 0.009 | $8.7\times10^2$ | 111 |
| Carotid plaque (OR) | HDL-C independent | 1.27 (1.15; 1.40) | < 0.001 | MR Egger | < 0.001 | $4.3\times10^2$ | 187 |
| Carotid plaque (OR) | TG independent | 1.22 (1.09; 1.36) | < 0.001 | MR Egger | 0.013 | $3.1\times10^2$ | 162 |
| Carotid plaque (OR) | HDL-C & TG independent | 1.40 (1.29; 1.52) | < 0.001 | IVW | 0.004 | $5.6\times10^2$ | 222 |
| Glucose (mmol/l) | Total | 0.02 (-0.00; 0.04) | 0.075 | MR Egger | 0.049 | $9.2\times10^3$ | 115 |
| Glucose (mmol/l) | HDL-C independent | -0.01 (-0.02; 0.00) | 0.097 | IVW | < 0.001 | $4.1\times10^4$ | 191 |
| Glucose (mmol/l) | TG independent | 0.01 (-0.01; 0.02) | 0.361 | MR Egger | < 0.001 | $1.7\times10^4$ | 172 |
| Glucose (mmol/l) | HDL-C & TG independent | 0.01 (-0.01; 0.02) | 0.449 | MR Egger | < 0.001 | $2.0\times10^4$ | 226 |
| HbA1c (mmol/mol) | Total | -0.03 (-0.14; 0.07) | 0.520 | MR Egger | < 0.001 | $3.4\times10^2$ | 113 |
| HbA1c (mmol/mol) | HDL-C independent | -0.09 (-0.16; -0.02) | 0.013 | MR Egger | < 0.001 | $7.7\times10^2$ | 187 |
| HbA1c (mmol/mol) | TG independent | 0.02 (-0.06; 0.10) | 0.587 | MR Egger | < 0.001 | $6.2\times10^2$ | 169 |

**Table S2:** (Multivariable) Mendelian randomization effect estimates for one standard deviation change in LDL-C *(continued)*

| Outcome | Effect type | Point estimate (95%CI) | P-value | Estimator | Heterogeneity P-value | Precision | No. variants |
|---|---|---|---|---|---|---|---|
| HbA1c (mmol/mol) | HDL-C & TG independent | -0.04 (-0.11; 0.03) | 0.248 | MR Egger | < 0.001 | $7.8\times10^{2}$ | 223 |
| T2DM (OR) | Total | 1.03 (0.98; 1.08) | 0.205 | MR Egger | < 0.001 | $1.9\times10^{3}$ | 105 |
| T2DM (OR) | HDL-C independent | 0.94 (0.92; 0.96) | < 0.001 | IVW | < 0.001 | $7.7\times10^{3}$ | 171 |
| T2DM (OR) | TG independent | 0.91 (0.88; 0.94) | < 0.001 | IVW | < 0.001 | $4.6\times10^{3}$ | 149 |
| T2DM (OR) | HDL-C & TG independent | 0.90 (0.88; 0.93) | < 0.001 | IVW | < 0.001 | $4.4\times10^{3}$ | 202 |
| CRP (mg/L) | Total | -0.15 (-0.19; -0.11) | < 0.001 | IVW | < 0.001 | $2.3\times10^{3}$ | 93 |
| CRP (mg/L) | HDL-C independent | -0.21 (-0.26; -0.16) | < 0.001 | MR Egger | < 0.001 | $1.5\times10^{3}$ | 173 |
| CRP (mg/L) | TG independent | -0.15 (-0.21; -0.10) | < 0.001 | MR Egger | < 0.001 | $1.2\times10^{3}$ | 155 |
| CRP (mg/L) | HDL-C & TG independent | -0.24 (-0.29; -0.19) | < 0.001 | IVW | < 0.001 | $1.7\times10^{3}$ | 210 |
| SBP (mmHg) | Total | 0.41 (0.29; 0.54) | < 0.001 | IVW | < 0.001 | $2.4\times10^{2}$ | 90 |
| SBP (mmHg) | HDL-C independent | 0.38 (0.26; 0.50) | < 0.001 | IVW | < 0.001 | $2.6\times10^{2}$ | 156 |
| SBP (mmHg) | TG independent | 0.22 (0.08; 0.37) | 0.002 | IVW | < 0.001 | $1.9\times10^{2}$ | 140 |
| SBP (mmHg) | HDL-C & TG independent | 0.28 (0.13; 0.43) | < 0.001 | IVW | < 0.001 | $1.8\times10^{2}$ | 185 |
| DBP (mmHg) | Total | -0.03 (-0.11; 0.04) | 0.356 | IVW | < 0.001 | $7.5\times10^{2}$ | 91 |
| DBP (mmHg) | HDL-C independent | -0.04 (-0.10; 0.03) | 0.306 | IVW | < 0.001 | $8.1\times10^{2}$ | 156 |
| DBP (mmHg) | TG independent | -0.11 (-0.19; -0.03) | 0.007 | IVW | < 0.001 | $5.8\times10^{2}$ | 139 |
| DBP (mmHg) | HDL-C & TG independent | -0.07 (-0.16; 0.01) | 0.080 | IVW | < 0.001 | $5.6\times10^{2}$ | 187 |
| PBL (OR) | Total | 1.18 (0.96; 1.44) | 0.108 | IVW | 0.084 | $9.5\times10^{1}$ | 51 |
| PBL (OR) | HDL-C independent | 1.18 (0.97; 1.43) | 0.089 | IVW | 0.157 | $1.0\times10^{2}$ | 91 |
| PBL (OR) | TG independent | 1.13 (0.81; 1.58) | 0.467 | MR Egger | 0.023 | $3.4\times10^{1}$ | 81 |
| PBL (OR) | HDL-C & TG independent | 1.03 (0.78; 1.36) | 0.814 | MR Egger | 0.041 | $5.0\times10^{1}$ | 113 |
| BUN (mg/dl) | Total | -0.00 (-0.01; 0.00) | 0.481 | MR Egger | < 0.001 | $7.7\times10^{4}$ | 110 |
| BUN (mg/dl) | HDL-C independent | 0.00 (-0.00; 0.01) | 0.130 | IVW | < 0.001 | $3.5\times10^{5}$ | 181 |
| BUN (mg/dl) | TG independent | 0.00 (-0.00; 0.01) | 0.340 | MR Egger | < 0.001 | $1.4\times10^{5}$ | 165 |
| BUN (mg/dl) | HDL-C & TG independent | 0.00 (-0.00; 0.01) | 0.071 | IVW | < 0.001 | $2.1\times10^{5}$ | 219 |
| eGFR (SD of log(eGFR)) | Total | 0.00 (0.00; 0.01) | 0.026 | MR Egger | < 0.001 | $5.6\times10^{5}$ | 107 |
| eGFR (SD of log(eGFR)) | HDL-C independent | 0.00 (-0.00; 0.00) | 0.649 | MR Egger | < 0.001 | $1.3\times10^{6}$ | 183 |
| eGFR (SD of log(eGFR)) | TG independent | -0.00 (-0.00; 0.00) | 0.151 | IVW | < 0.001 | $1.6\times10^{6}$ | 161 |
| eGFR (SD of log(eGFR)) | HDL-C & TG independent | -0.00 (-0.00; 0.00) | 0.239 | IVW | < 0.001 | $1.5\times10^{6}$ | 214 |
| CKD (OR) | Total | 0.99 (0.95; 1.03) | 0.614 | IVW | 0.058 | $2.8\times10^{3}$ | 116 |
| CKD (OR) | HDL-C independent | 0.99 (0.95; 1.02) | 0.434 | IVW | 0.027 | $3.5\times10^{3}$ | 190 |
| CKD (OR) | TG independent | 1.00 (0.94; 1.05) | 0.942 | MR Egger | 0.005 | $1.3\times10^{3}$ | 171 |
| CKD (OR) | HDL-C & TG independent | 0.99 (0.95; 1.04) | 0.720 | IVW | < 0.001 | $2.1\times10^{3}$ | 227 |
| IBD (OR) | Total | 1.15 (1.07; 1.22) | < 0.001 | IVW | 0.008 | $9.4\times10^{2}$ | 117 |
| IBD (OR) | HDL-C independent | 1.15 (1.08; 1.22) | < 0.001 | IVW | < 0.001 | $9.9\times10^{2}$ | 196 |
| IBD (OR) | TG independent | 1.16 (1.05; 1.29) | 0.003 | MR Egger | < 0.001 | $3.7\times10^{2}$ | 171 |
| IBD (OR) | HDL-C & TG independent | 1.13 (1.05; 1.23) | 0.002 | IVW | < 0.001 | $6.1\times10^{2}$ | 232 |
| UC (OR) | Total | 1.37 (1.15; 1.63) | < 0.001 | MR Egger | 0.196 | $1.3\times10^{2}$ | 117 |

**Table S2:** (Multivariable) Mendelian randomization effect estimates for one standard deviation change in LDL-C *(continued)*

| Outcome | Effect type | Point estimate (95%CI) | P-value | Estimator | Heterogeneity P-value | Precision | No. variants |
|---|---|---|---|---|---|---|---|
| UC (OR) | HDL-C independent | 1.24 (1.11; 1.38) | < 0.001 | MR Egger | 0.008 | 3.3×10² | 196 |
| UC (OR) | TG independent | 1.15 (1.04; 1.27) | 0.006 | IVW | 0.002 | 4.0×10² | 176 |
| UC (OR) | HDL-C & TG independent | 1.17 (1.06; 1.30) | 0.002 | IVW | < 0.001 | 3.8×10² | 233 |
| CD (OR) | Total | 1.10 (1.00; 1.20) | 0.042 | IVW | 0.497 | 5.0×10² | 117 |
| CD (OR) | HDL-C independent | 1.09 (1.00; 1.19) | 0.054 | IVW | 0.395 | 5.2×10² | 197 |
| CD (OR) | TG independent | 1.01 (0.87; 1.17) | 0.928 | MR Egger | 0.051 | 1.8×10² | 173 |
| CD (OR) | HDL-C & TG independent | 1.00 (0.89; 1.13) | 0.967 | IVW | 0.056 | 2.8×10² | 233 |
| Arthritis (OR) | Total | 0.93 (0.84; 1.03) | 0.158 | IVW | 0.060 | 4.0×10² | 91 |
| Arthritis (OR) | HDL-C independent | 0.95 (0.87; 1.04) | 0.262 | IVW | 0.212 | 5.3×10² | 157 |
| Arthritis (OR) | TG independent | 1.05 (0.90; 1.23) | 0.520 | MR Egger | 0.181 | 1.6×10² | 141 |
| Arthritis (OR) | HDL-C & TG independent | 0.97 (0.85; 1.11) | 0.693 | MR Egger | 0.186 | 2.2×10² | 185 |
| ALS (OR) | Total | 1.11 (1.04; 1.17) | < 0.001 | IVW | 0.612 | 1.1×10³ | 119 |
| ALS (OR) | HDL-C independent | 1.09 (1.03; 1.15) | 0.002 | IVW | 0.487 | 1.3×10³ | 200 |
| ALS (OR) | TG independent | 1.03 (0.94; 1.13) | 0.547 | MR Egger | 0.292 | 4.5×10² | 175 |
| ALS (OR) | HDL-C & TG independent | 1.05 (0.97; 1.13) | 0.196 | IVW | 0.210 | 6.8×10² | 231 |
| MS (OR) | Total | 1.07 (0.92; 1.24) | 0.381 | IVW | 0.106 | 1.8×10² | 19 |
| MS (OR) | HDL-C independent | 0.86 (0.71; 1.04) | 0.119 | MR Egger | 0.008 | 1.0×10² | 32 |
| MS (OR) | TG independent | 1.11 (0.94; 1.31) | 0.209 | IVW | 0.115 | 1.5×10² | 33 |
| MS (OR) | HDL-C & TG independent | 0.92 (0.77; 1.10) | 0.366 | MR Egger | 0.004 | 1.2×10² | 39 |
| Alzheimer (OR) | Total | 1.01 (0.99; 1.03) | 0.484 | MR Egger | < 0.001 | 8.6×10³ | 82 |
| Alzheimer (OR) | HDL-C independent | 1.05 (1.04; 1.07) | < 0.001 | MR Egger | < 0.001 | 1.8×10⁴ | 156 |
| Alzheimer (OR) | TG independent | 1.05 (1.03; 1.06) | < 0.001 | IVW | < 0.001 | 2.2×10⁴ | 135 |
| Alzheimer (OR) | HDL-C & TG independent | 1.05 (1.04; 1.07) | < 0.001 | MR Egger | < 0.001 | 1.9×10⁴ | 198 |
| Parkinson (OR) | Total | 0.93 (0.81; 1.07) | 0.315 | MR Egger | 0.216 | 1.9×10² | 117 |
| Parkinson (OR) | HDL-C independent | 0.93 (0.87; 1.00) | 0.050 | IVW | 0.082 | 7.9×10² | 189 |
| Parkinson (OR) | TG independent | 0.94 (0.86; 1.02) | 0.133 | IVW | 0.002 | 5.6×10² | 174 |
| Parkinson (OR) | HDL-C & TG independent | 0.93 (0.85; 1.01) | 0.096 | IVW | < 0.001 | 5.3×10² | 230 |
| Pancreatic cancer (OR) | Total | 1.20 (0.99; 1.46) | 0.066 | IVW | 0.002 | 1.0×10² | 117 |
| Pancreatic cancer (OR) | HDL-C independent | 1.11 (0.91; 1.34) | 0.305 | IVW | 0.003 | 1.0×10² | 192 |
| Pancreatic cancer (OR) | TG independent | 1.34 (1.06; 1.69) | 0.014 | IVW | 0.021 | 7.1×10¹ | 177 |
| Pancreatic cancer (OR) | HDL-C & TG independent | 1.14 (0.90; 1.45) | 0.269 | IVW | < 0.001 | 6.8×10¹ | 233 |
| Colon cancer (OR) | Total | 0.95 (0.79; 1.13) | 0.548 | MR Egger | 0.087 | 1.2×10² | 118 |
| Colon cancer (OR) | HDL-C independent | 0.99 (0.89; 1.11) | 0.890 | MR Egger | < 0.001 | 3.2×10² | 198 |
| Colon cancer (OR) | TG independent | 1.03 (0.93; 1.14) | 0.580 | IVW | < 0.001 | 3.9×10² | 177 |
| Colon cancer (OR) | HDL-C & TG independent | 1.02 (0.92; 1.12) | 0.754 | IVW | < 0.001 | 3.9×10² | 236 |
| Rectal cancer (OR) | Total | 1.21 (1.08; 1.35) | < 0.001 | IVW | 0.833 | 3.1×10² | 116 |
| Rectal cancer (OR) | HDL-C independent | 1.17 (1.05; 1.30) | 0.005 | IVW | 0.500 | 3.3×10² | 193 |
| Rectal cancer (OR) | TG independent | 1.20 (1.05; 1.37) | 0.009 | IVW | 0.840 | 2.1×10² | 176 |

**Table S2:** (Multivariable) Mendelian randomization effect estimates for one standard deviation change in LDL-C *(continued)*

| Outcome | Effect type | Point estimate (95%CI) | P-value | Estimator | Heterogeneity P-value | Precision | No. variants |
|---|---|---|---|---|---|---|---|
| Rectal cancer (OR) | HDL-C & TG independent | 1.28 (1.12; 1.47) | < 0.001 | IVW | 0.586 | $2.1\times10^2$ | 231 |
| Melanoma (OR) | Total | 0.92 (0.86; 0.99) | 0.018 | IVW | 0.171 | $8.6\times10^2$ | 117 |
| Melanoma (OR) | HDL-C independent | 0.93 (0.87; 0.99) | 0.024 | IVW | 0.014 | $9.9\times10^2$ | 191 |
| Melanoma (OR) | TG independent | 0.90 (0.84; 0.97) | 0.008 | IVW | 0.024 | $6.6\times10^2$ | 176 |
| Melanoma (OR) | HDL-C & TG independent | 0.90 (0.83; 0.97) | 0.008 | IVW | < 0.001 | $6.5\times10^2$ | 231 |
| Lung cancer (OR) | Total | 0.76 (0.69; 0.84) | < 0.001 | IVW | 0.045 | $3.6\times10^2$ | 116 |
| Lung cancer (OR) | HDL-C independent | 0.78 (0.70; 0.86) | < 0.001 | IVW | 0.017 | $3.8\times10^2$ | 194 |
| Lung cancer (OR) | TG independent | 0.77 (0.68; 0.88) | < 0.001 | IVW | 0.061 | $2.2\times10^2$ | 177 |
| Lung cancer (OR) | HDL-C & TG independent | 0.77 (0.68; 0.87) | < 0.001 | IVW | 0.022 | $2.5\times10^2$ | 235 |

**Table S3:** (Multivariable) Mendelian randomization effect estimates for one standard deviation change in HDL-C

| Outcome | Effect type | Point estimate (95%CI) | P-value | Estimator | Heterogeneity P-value | Precision | No. variants |
|---|---|---|---|---|---|---|---|
| CHD (OR) | Total | 0.75 (0.70; 0.80) | < 0.001 | IVW | < 0.001 | $8.1\times10^2$ | 26 |
| CHD (OR) | LDL-C independent | 0.78 (0.73; 0.83) | < 0.001 | IVW | < 0.001 | $9.4\times10^2$ | 50 |
| CHD (OR) | TG independent | 0.84 (0.78; 0.91) | < 0.001 | IVW | < 0.001 | $5.7\times10^2$ | 37 |
| CHD (OR) | TG & LDL-C independent | 0.76 (0.70; 0.84) | < 0.001 | IVW | < 0.001 | $4.4\times10^2$ | 57 |
| HF (OR) | Total | 0.97 (0.94; 1.01) | 0.165 | IVW | < 0.001 | $2.9\times10^3$ | 81 |
| HF (OR) | LDL-C independent | 0.98 (0.95; 1.02) | 0.272 | IVW | < 0.001 | $3.2\times10^3$ | 185 |
| HF (OR) | TG independent | 1.01 (0.97; 1.05) | 0.682 | IVW | < 0.001 | $2.1\times10^3$ | 129 |
| HF (OR) | TG & LDL-C independent | 1.01 (0.96; 1.05) | 0.786 | IVW | < 0.001 | $2.0\times10^3$ | 216 |
| AF (OR) | Total | 0.99 (0.96; 1.02) | 0.490 | IVW | 0.251 | $3.7\times10^3$ | 86 |
| AF (OR) | LDL-C independent | 0.99 (0.96; 1.02) | 0.509 | IVW | 0.067 | $3.8\times10^3$ | 202 |
| AF (OR) | TG independent | 1.01 (0.97; 1.05) | 0.579 | IVW | 0.008 | $2.8\times10^3$ | 139 |
| AF (OR) | TG & LDL-C independent | 1.01 (0.98; 1.05) | 0.440 | IVW | < 0.001 | $2.9\times10^3$ | 238 |
| Any stroke (OR) | Total | 0.91 (0.83; 0.99) | 0.036 | MR Egger | 0.002 | $4.5\times10^2$ | 85 |
| Any stroke (OR) | LDL-C independent | 0.98 (0.94; 1.02) | 0.429 | IVW | < 0.001 | $2.3\times10^3$ | 192 |
| Any stroke (OR) | TG independent | 0.98 (0.94; 1.03) | 0.535 | IVW | 0.029 | $1.6\times10^3$ | 138 |
| Any stroke (OR) | TG & LDL-C independent | 0.98 (0.93; 1.03) | 0.426 | IVW | < 0.001 | $1.5\times10^3$ | 228 |
| Any ischemic stroke (OR) | Total | 0.91 (0.83; 1.01) | 0.067 | MR Egger | < 0.001 | $3.9\times10^2$ | 85 |
| Any ischemic stroke (OR) | LDL-C independent | 0.99 (0.95; 1.04) | 0.753 | IVW | < 0.001 | $2.0\times10^3$ | 189 |
| Any ischemic stroke (OR) | TG independent | 1.00 (0.94; 1.06) | 0.967 | IVW | 0.058 | $1.1\times10^3$ | 137 |
| Any ischemic stroke (OR) | TG & LDL-C independent | 0.98 (0.93; 1.04) | 0.483 | IVW | 0.010 | $1.2\times10^3$ | 226 |
| Large artery stroke (OR) | Total | 1.01 (0.89; 1.15) | 0.856 | IVW | 0.052 | $2.2\times10^2$ | 85 |
| Large artery stroke (OR) | LDL-C independent | 0.98 (0.88; 1.10) | 0.752 | IVW | < 0.001 | $3.1\times10^2$ | 192 |
| Large artery stroke (OR) | TG independent | 1.04 (0.90; 1.21) | 0.580 | IVW | 0.068 | $1.8\times10^2$ | 140 |

**Table S3:** (Multivariable) Mendelian randomization effect estimates for one standard deviation change in HDL-C *(continued)*

| Outcome | Effect type | Point estimate (95%CI) | P-value | Estimator | Heterogeneity P-value | Precision | No. variants |
|---|---|---|---|---|---|---|---|
| Large artery stroke (OR) | TG & LDL-C independent | 1.03 (0.90; 1.18) | 0.679 | IVW | < 0.001 | $2.0\times10^{2}$ | 229 |
| Cardioembolic stroke (OR) | Total | 1.00 (0.91; 1.10) | 0.986 | IVW | 0.424 | $4.5\times10^{2}$ | 85 |
| Cardioembolic stroke (OR) | LDL-C independent | 1.00 (0.91; 1.09) | 0.930 | IVW | 0.039 | $5.1\times10^{2}$ | 191 |
| Cardioembolic stroke (OR) | TG independent | 1.00 (0.90; 1.11) | 0.961 | IVW | 0.509 | $3.5\times10^{2}$ | 139 |
| Cardioembolic stroke (OR) | TG & LDL-C independent | 0.98 (0.88; 1.09) | 0.722 | IVW | 0.041 | $3.5\times10^{2}$ | 228 |
| Small vessel stroke (OR) | Total | 0.93 (0.73; 1.18) | 0.533 | MR Egger | 0.302 | $6.5\times10^{1}$ | 84 |
| Small vessel stroke (OR) | LDL-C independent | 0.96 (0.83; 1.11) | 0.581 | MR Egger | 0.017 | $1.9\times10^{2}$ | 195 |
| Small vessel stroke (OR) | TG independent | 0.99 (0.88; 1.12) | 0.886 | IVW | 0.046 | $2.5\times10^{2}$ | 138 |
| Small vessel stroke (OR) | TG & LDL-C independent | 0.95 (0.81; 1.11) | 0.506 | MR Egger | 0.011 | $1.6\times10^{2}$ | 232 |
| cIMT (mm) | Total | 0.00 (-0.00; 0.00) | 0.781 | IVW | 0.006 | $3.5\times10^{5}$ | 84 |
| cIMT (mm) | LDL-C independent | -0.00 (-0.00; 0.00) | 0.696 | IVW | < 0.001 | $3.9\times10^{5}$ | 196 |
| cIMT (mm) | TG independent | -0.00 (-0.00; 0.00) | 0.859 | IVW | < 0.001 | $2.6\times10^{5}$ | 138 |
| cIMT (mm) | TG & LDL-C independent | -0.00 (-0.01; 0.00) | 0.319 | MR Egger | < 0.001 | $1.7\times10^{5}$ | 232 |
| Carotid plaque (OR) | Total | 0.87 (0.73; 1.04) | 0.130 | MR Egger | 0.039 | $1.2\times10^{2}$ | 83 |
| Carotid plaque (OR) | LDL-C independent | 0.87 (0.80; 0.94) | < 0.001 | IVW | < 0.001 | $6.8\times10^{2}$ | 188 |
| Carotid plaque (OR) | TG independent | 0.99 (0.90; 1.09) | 0.836 | IVW | 0.006 | $4.1\times10^{2}$ | 132 |
| Carotid plaque (OR) | TG & LDL-C independent | 0.88 (0.80; 0.97) | 0.008 | IVW | 0.004 | $4.3\times10^{2}$ | 222 |
| Glucose (mmol/l) | Total | -0.09 (-0.12; -0.05) | < 0.001 | MR Egger | 0.115 | $4.0\times10^{3}$ | 79 |
| Glucose (mmol/l) | LDL-C independent | -0.01 (-0.02; 0.01) | 0.291 | IVW | < 0.001 | $2.2\times10^{4}$ | 191 |
| Glucose (mmol/l) | TG independent | -0.05 (-0.07; -0.03) | < 0.001 | MR Egger | < 0.001 | $8.4\times10^{3}$ | 131 |
| Glucose (mmol/l) | TG & LDL-C independent | -0.03 (-0.05; -0.01) | 0.003 | MR Egger | < 0.001 | $1.2\times10^{4}$ | 229 |
| HbA1c (mmol/mol) | Total | -0.03 (-0.10; 0.04) | 0.352 | IVW | 0.008 | $7.8\times10^{2}$ | 76 |
| HbA1c (mmol/mol) | LDL-C independent | -0.03 (-0.09; 0.04) | 0.439 | IVW | < 0.001 | $8.6\times10^{2}$ | 187 |
| HbA1c (mmol/mol) | TG independent | -0.33 (-0.44; -0.22) | < 0.001 | MR Egger | < 0.001 | $3.3\times10^{2}$ | 127 |
| HbA1c (mmol/mol) | TG & LDL-C independent | -0.05 (-0.13; 0.04) | 0.279 | IVW | < 0.001 | $5.7\times10^{2}$ | 220 |
| T2DM (OR) | Total | 0.97 (0.94; 1.00) | 0.087 | IVW | < 0.001 | $4.2\times10^{3}$ | 71 |
| T2DM (OR) | LDL-C independent | 0.95 (0.91; 0.99) | 0.007 | MR Egger | < 0.001 | $2.6\times10^{3}$ | 171 |
| T2DM (OR) | TG independent | 0.96 (0.93; 0.99) | 0.016 | IVW | < 0.001 | $3.3\times10^{3}$ | 118 |
| T2DM (OR) | TG & LDL-C independent | 0.97 (0.93; 1.01) | 0.165 | MR Egger | < 0.001 | $2.2\times10^{3}$ | 204 |
| CRP (mg/L) | Total | 0.07 (0.02; 0.12) | 0.003 | IVW | 0.002 | $1.7\times10^{3}$ | 78 |
| CRP (mg/L) | LDL-C independent | 0.08 (0.03; 0.13) | < 0.001 | IVW | < 0.001 | $1.9\times10^{3}$ | 174 |
| CRP (mg/L) | TG independent | 0.13 (0.07; 0.19) | < 0.001 | IVW | < 0.001 | $1.2\times10^{3}$ | 124 |
| CRP (mg/L) | TG & LDL-C independent | 0.20 (0.14; 0.26) | < 0.001 | IVW | < 0.001 | $1.2\times10^{3}$ | 210 |
| SBP (mmHg) | Total | -0.44 (-0.58; -0.29) | < 0.001 | IVW | < 0.001 | $1.8\times10^{2}$ | 70 |
| SBP (mmHg) | LDL-C independent | -0.40 (-0.58; -0.21) | < 0.001 | MR Egger | < 0.001 | $1.1\times10^{2}$ | 159 |
| SBP (mmHg) | TG independent | -0.25 (-0.42; -0.07) | 0.006 | IVW | < 0.001 | $1.2\times10^{2}$ | 112 |
| SBP (mmHg) | TG & LDL-C independent | -0.24 (-0.42; -0.05) | 0.011 | MR Egger | < 0.001 | $1.1\times10^{2}$ | 190 |
| DBP (mmHg) | Total | -0.24 (-0.33; -0.15) | < 0.001 | IVW | 0.054 | $4.4\times10^{2}$ | 70 |

**Table S3:** (Multivariable) Mendelian randomization effect estimates for one standard deviation change in HDL-C *(continued)*

| Outcome | Effect type | Point estimate (95%CI) | P-value | Estimator | Heterogeneity P-value | Precision | No. variants |
|---|---|---|---|---|---|---|---|
| DBP (mmHg) | LDL-C independent | -0.15 (-0.23; -0.07) | < 0.001 | IVW | < 0.001 | $5.9\times10^2$ | 156 |
| DBP (mmHg) | TG independent | -0.24 (-0.34; -0.13) | < 0.001 | IVW | < 0.001 | $3.7\times10^2$ | 111 |
| DBP (mmHg) | TG & LDL-C independent | -0.16 (-0.26; -0.05) | 0.004 | MR Egger | < 0.001 | $3.4\times10^2$ | 189 |
| PBL (OR) | Total | 1.03 (0.59; 1.80) | 0.910 | MR Egger | 0.661 | $1.2\times10^1$ | 44 |
| PBL (OR) | LDL-C independent | 1.15 (0.92; 1.44) | 0.212 | IVW | 0.157 | $7.9\times10^1$ | 91 |
| PBL (OR) | TG independent | 1.34 (1.03; 1.75) | 0.029 | IVW | 0.007 | $5.5\times10^1$ | 70 |
| PBL (OR) | TG & LDL-C independent | 1.51 (1.10; 2.08) | 0.011 | MR Egger | 0.015 | $3.8\times10^1$ | 112 |
| BUN (mg/dl) | Total | -0.00 (-0.01; 0.00) | 0.182 | IVW | 0.293 | $1.9\times10^5$ | 76 |
| BUN (mg/dl) | LDL-C independent | -0.00 (-0.01; 0.00) | 0.150 | IVW | < 0.001 | $2.2\times10^5$ | 181 |
| BUN (mg/dl) | TG independent | -0.01 (-0.01; -0.00) | 0.009 | IVW | 0.003 | $1.9\times10^5$ | 131 |
| BUN (mg/dl) | TG & LDL-C independent | -0.01 (-0.01; -0.00) | 0.031 | IVW | < 0.001 | $1.6\times10^5$ | 219 |
| eGFR (SD of log(eGFR)) | Total | 0.00 (-0.00; 0.00) | 0.828 | IVW | < 0.001 | $1.4\times10^6$ | 78 |
| eGFR (SD of log(eGFR)) | LDL-C independent | 0.00 (-0.00; 0.00) | 0.542 | IVW | < 0.001 | $1.5\times10^6$ | 183 |
| eGFR (SD of log(eGFR)) | TG independent | 0.00 (-0.00; 0.00) | 0.909 | IVW | < 0.001 | $8.6\times10^5$ | 124 |
| eGFR (SD of log(eGFR)) | TG & LDL-C independent | -0.00 (-0.00; 0.00) | 0.521 | IVW | < 0.001 | $9.4\times10^5$ | 214 |
| CKD (OR) | Total | 0.99 (0.95; 1.04) | 0.816 | IVW | 0.118 | $1.6\times10^3$ | 80 |
| CKD (OR) | LDL-C independent | 1.00 (0.96; 1.04) | 0.984 | IVW | 0.027 | $2.2\times10^3$ | 190 |
| CKD (OR) | TG independent | 0.98 (0.93; 1.03) | 0.407 | IVW | < 0.001 | $1.4\times10^3$ | 134 |
| CKD (OR) | TG & LDL-C independent | 0.98 (0.93; 1.03) | 0.397 | IVW | < 0.001 | $1.4\times10^3$ | 227 |
| IBD (OR) | Total | 1.03 (0.94; 1.12) | 0.561 | IVW | 0.095 | $5.2\times10^2$ | 84 |
| IBD (OR) | LDL-C independent | 1.01 (0.94; 1.09) | 0.742 | IVW | < 0.001 | $7.0\times10^2$ | 196 |
| IBD (OR) | TG independent | 1.03 (0.93; 1.13) | 0.601 | IVW | 0.004 | $4.4\times10^2$ | 136 |
| IBD (OR) | TG & LDL-C independent | 1.06 (0.95; 1.18) | 0.277 | MR Egger | < 0.001 | $3.3\times10^2$ | 233 |
| UC (OR) | Total | 1.06 (0.96; 1.16) | 0.273 | IVW | 0.023 | $4.0\times10^2$ | 85 |
| UC (OR) | LDL-C independent | 1.04 (0.94; 1.14) | 0.458 | IVW | 0.005 | $4.5\times10^2$ | 197 |
| UC (OR) | TG independent | 1.02 (0.91; 1.14) | 0.769 | IVW | 0.007 | $2.8\times10^2$ | 138 |
| UC (OR) | TG & LDL-C independent | 1.07 (0.93; 1.22) | 0.338 | MR Egger | 0.006 | $2.1\times10^2$ | 234 |
| CD (OR) | Total | 0.99 (0.89; 1.10) | 0.842 | IVW | 0.595 | $3.3\times10^2$ | 84 |
| CD (OR) | LDL-C independent | 1.00 (0.90; 1.10) | 0.957 | IVW | 0.395 | $3.6\times10^2$ | 197 |
| CD (OR) | TG independent | 1.06 (0.92; 1.21) | 0.443 | IVW | 0.096 | $2.0\times10^2$ | 137 |
| CD (OR) | TG & LDL-C independent | 1.07 (0.94; 1.23) | 0.311 | IVW | 0.056 | $2.1\times10^2$ | 233 |
| Arthritis (OR) | Total | 0.98 (0.88; 1.10) | 0.733 | IVW | 0.666 | $3.2\times10^2$ | 66 |
| Arthritis (OR) | LDL-C independent | 0.98 (0.88; 1.09) | 0.682 | IVW | 0.212 | $3.3\times10^2$ | 157 |
| Arthritis (OR) | TG independent | 1.05 (0.92; 1.19) | 0.449 | IVW | 0.395 | $2.3\times10^2$ | 108 |
| Arthritis (OR) | TG & LDL-C independent | 1.06 (0.92; 1.22) | 0.397 | IVW | 0.143 | $2.0\times10^2$ | 187 |
| ALS (OR) | Total | 1.05 (0.97; 1.14) | 0.192 | IVW | 0.465 | $6.1\times10^2$ | 84 |
| ALS (OR) | LDL-C independent | 1.04 (0.96; 1.12) | 0.355 | IVW | 0.487 | $6.8\times10^2$ | 200 |
| ALS (OR) | TG independent | 1.08 (0.97; 1.19) | 0.156 | IVW | 0.053 | $3.7\times10^2$ | 135 |

**Table S3:** (Multivariable) Mendelian randomization effect estimates for one standard deviation change in HDL-C *(continued)*

| Outcome | Effect type | Point estimate (95%CI) | P-value | Estimator | Heterogeneity P-value | Precision | No. variants |
|---|---|---|---|---|---|---|---|
| ALS (OR) | TG & LDL-C independent | 1.07 (0.98; 1.18) | 0.143 | IVW | 0.210 | $4.2\times10^{2}$ | 231 |
| MS (OR) | Total | 1.06 (0.93; 1.21) | 0.346 | IVW | 0.013 | $2.2\times10^{2}$ | 15 |
| MS (OR) | LDL-C independent | 1.07 (0.94; 1.22) | 0.292 | IVW | 0.006 | $2.4\times10^{2}$ | 33 |
| MS (OR) | TG independent | 1.08 (0.91; 1.28) | 0.351 | IVW | 0.020 | $1.3\times10^{2}$ | 24 |
| MS (OR) | TG & LDL-C independent | 0.97 (0.77; 1.21) | 0.762 | MR Egger | 0.002 | $7.6\times10^{1}$ | 40 |
| Alzheimer (OR) | Total | 0.98 (0.97; 0.99) | < 0.001 | IVW | 0.151 | $2.9\times10^{4}$ | 82 |
| Alzheimer (OR) | LDL-C independent | 0.98 (0.97; 0.99) | < 0.001 | IVW | < 0.001 | $3.4\times10^{4}$ | 159 |
| Alzheimer (OR) | TG independent | 0.98 (0.97; 1.00) | 0.013 | IVW | 0.003 | $2.1\times10^{4}$ | 130 |
| Alzheimer (OR) | TG & LDL-C independent | 0.97 (0.96; 0.98) | < 0.001 | IVW | < 0.001 | $2.0\times10^{4}$ | 195 |
| Parkinson (OR) | Total | 0.94 (0.86; 1.04) | 0.225 | IVW | 0.351 | $4.6\times10^{2}$ | 79 |
| Parkinson (OR) | LDL-C independent | 0.96 (0.87; 1.05) | 0.345 | IVW | 0.082 | $4.5\times10^{2}$ | 189 |
| Parkinson (OR) | TG independent | 0.95 (0.85; 1.05) | 0.286 | IVW | 0.008 | $3.7\times10^{2}$ | 136 |
| Parkinson (OR) | TG & LDL-C independent | 0.95 (0.86; 1.05) | 0.314 | IVW | < 0.001 | $3.8\times10^{2}$ | 230 |
| Pancreatic cancer (OR) | Total | 1.19 (0.92; 1.55) | 0.178 | IVW | 0.030 | $5.7\times10^{1}$ | 82 |
| Pancreatic cancer (OR) | LDL-C independent | 1.26 (0.98; 1.61) | 0.069 | IVW | 0.003 | $6.3\times10^{1}$ | 192 |
| Pancreatic cancer (OR) | TG independent | 1.18 (0.87; 1.60) | 0.296 | IVW | 0.017 | $4.1\times10^{1}$ | 136 |
| Pancreatic cancer (OR) | TG & LDL-C independent | 1.25 (0.93; 1.69) | 0.138 | IVW | < 0.001 | $4.3\times10^{1}$ | 233 |
| Colon cancer (OR) | Total | 1.07 (0.96; 1.19) | 0.205 | IVW | 0.047 | $3.3\times10^{2}$ | 83 |
| Colon cancer (OR) | LDL-C independent | 1.05 (0.95; 1.16) | 0.342 | IVW | < 0.001 | $3.7\times10^{2}$ | 195 |
| Colon cancer (OR) | TG independent | 1.13 (0.99; 1.28) | 0.061 | IVW | < 0.001 | $2.4\times10^{2}$ | 137 |
| Colon cancer (OR) | TG & LDL-C independent | 1.10 (0.98; 1.25) | 0.118 | IVW | < 0.001 | $2.6\times10^{2}$ | 236 |
| Rectal cancer (OR) | Total | 0.92 (0.79; 1.07) | 0.262 | IVW | 0.353 | $1.7\times10^{2}$ | 80 |
| Rectal cancer (OR) | LDL-C independent | 0.91 (0.79; 1.04) | 0.177 | IVW | 0.500 | $2.0\times10^{2}$ | 193 |
| Rectal cancer (OR) | TG independent | 0.96 (0.81; 1.15) | 0.679 | IVW | 0.356 | $1.2\times10^{2}$ | 134 |
| Rectal cancer (OR) | TG & LDL-C independent | 0.85 (0.72; 1.01) | 0.070 | IVW | 0.586 | $1.3\times10^{2}$ | 231 |
| Melanoma (OR) | Total | 0.98 (0.90; 1.07) | 0.678 | IVW | 0.008 | $5.5\times10^{2}$ | 79 |
| Melanoma (OR) | LDL-C independent | 0.99 (0.92; 1.07) | 0.866 | IVW | 0.014 | $6.2\times10^{2}$ | 191 |
| Melanoma (OR) | TG independent | 1.03 (0.93; 1.13) | 0.600 | IVW | < 0.001 | $4.1\times10^{2}$ | 134 |
| Melanoma (OR) | TG & LDL-C independent | 1.03 (0.93; 1.13) | 0.590 | IVW | < 0.001 | $4.2\times10^{2}$ | 231 |
| Lung cancer (OR) | Total | 0.99 (0.85; 1.14) | 0.861 | IVW | 0.126 | $1.9\times10^{2}$ | 83 |
| Lung cancer (OR) | LDL-C independent | 1.01 (0.89; 1.15) | 0.857 | IVW | 0.017 | $2.4\times10^{2}$ | 194 |
| Lung cancer (OR) | TG independent | 0.92 (0.78; 1.09) | 0.331 | IVW | 0.173 | $1.4\times10^{2}$ | 137 |
| Lung cancer (OR) | TG & LDL-C independent | 1.00 (0.86; 1.16) | 0.972 | IVW | 0.022 | $1.7\times10^{2}$ | 235 |

**Table S4:** (Multivariable) Mendelian randomization effect estimates for one standard deviation change in TG

| Outcome | Effect type | Point estimate (95%CI) | P-value | Estimator | Heterogeneity P-value | Precision | No. variants |
|---|---|---|---|---|---|---|---|
| CHD (OR) | Total | 1.78 (1.65; 1.92) | < 0.001 | IVW | < 0.001 | $6.5\times10^2$ | 23 |
| CHD (OR) | LDL-C independent | 1.35 (1.18; 1.55) | < 0.001 | MR Egger | < 0.001 | $2.0\times10^2$ | 46 |
| CHD (OR) | HDL-C independent | 1.39 (1.27; 1.52) | < 0.001 | IVW | < 0.001 | $4.6\times10^2$ | 37 |
| CHD (OR) | LDL-C & HDL-C & TG independent | 1.15 (1.04; 1.28) | 0.007 | IVW | < 0.001 | $3.5\times10^2$ | 57 |
| HF (OR) | Total | 1.12 (1.08; 1.17) | < 0.001 | IVW | < 0.001 | $2.2\times10^3$ | 71 |
| HF (OR) | LDL-C independent | 1.09 (1.04; 1.14) | < 0.001 | IVW | < 0.001 | $1.9\times10^3$ | 166 |
| HF (OR) | HDL-C independent | 1.09 (1.04; 1.14) | < 0.001 | IVW | < 0.001 | $1.8\times10^3$ | 129 |
| HF (OR) | LDL-C & HDL-C & TG independent | 1.06 (1.00; 1.12) | 0.040 | IVW | < 0.001 | $1.3\times10^3$ | 216 |
| AF (OR) | Total | 1.16 (1.06; 1.27) | < 0.001 | MR Egger | 0.009 | $4.8\times10^2$ | 76 |
| AF (OR) | LDL-C independent | 1.01 (0.98; 1.05) | 0.466 | IVW | < 0.001 | $2.8\times10^3$ | 181 |
| AF (OR) | HDL-C independent | 1.02 (0.98; 1.06) | 0.353 | IVW | 0.008 | $2.7\times10^3$ | 139 |
| AF (OR) | LDL-C & HDL-C & TG independent | 1.02 (0.97; 1.06) | 0.443 | IVW | < 0.001 | $2.0\times10^3$ | 238 |
| Any stroke (OR) | Total | 1.05 (1.00; 1.10) | 0.067 | IVW | 0.431 | $1.6\times10^3$ | 77 |
| Any stroke (OR) | LDL-C independent | 1.00 (0.95; 1.06) | 0.891 | IVW | < 0.001 | $1.4\times10^3$ | 172 |
| Any stroke (OR) | HDL-C independent | 0.98 (0.93; 1.03) | 0.415 | IVW | 0.029 | $1.3\times10^3$ | 138 |
| Any stroke (OR) | LDL-C & HDL-C & TG independent | 0.95 (0.90; 1.02) | 0.144 | IVW | < 0.001 | $9.9\times10^2$ | 228 |
| Any ischemic stroke (OR) | Total | 1.04 (0.99; 1.10) | 0.117 | IVW | 0.636 | $1.4\times10^3$ | 77 |
| Any ischemic stroke (OR) | LDL-C independent | 1.00 (0.94; 1.07) | 0.912 | IVW | 0.075 | $1.0\times10^3$ | 170 |
| Any ischemic stroke (OR) | HDL-C independent | 0.98 (0.92; 1.05) | 0.545 | IVW | 0.058 | $8.9\times10^2$ | 137 |
| Any ischemic stroke (OR) | LDL-C & HDL-C & TG independent | 0.94 (0.88; 1.01) | 0.080 | IVW | 0.010 | $8.4\times10^2$ | 226 |
| Large artery stroke (OR) | Total | 1.06 (0.92; 1.23) | 0.411 | IVW | 0.054 | $1.8\times10^2$ | 78 |
| Large artery stroke (OR) | LDL-C independent | 0.86 (0.70; 1.06) | 0.153 | MR Egger | < 0.001 | $9.2\times10^1$ | 170 |
| Large artery stroke (OR) | HDL-C independent | 1.04 (0.88; 1.21) | 0.668 | IVW | 0.068 | $1.5\times10^2$ | 140 |
| Large artery stroke (OR) | LDL-C & HDL-C & TG independent | 1.04 (0.88; 1.23) | 0.623 | IVW | < 0.001 | $1.4\times10^2$ | 229 |
| Cardioembolic stroke (OR) | Total | 1.04 (0.94; 1.15) | 0.477 | IVW | 0.701 | $3.7\times10^2$ | 77 |
| Cardioembolic stroke (OR) | LDL-C independent | 1.06 (0.94; 1.19) | 0.371 | IVW | 0.062 | $2.7\times10^2$ | 172 |
| Cardioembolic stroke (OR) | HDL-C independent | 1.03 (0.92; 1.15) | 0.656 | IVW | 0.509 | $3.0\times10^2$ | 139 |
| Cardioembolic stroke (OR) | LDL-C & HDL-C & TG independent | 1.00 (0.88; 1.13) | 0.960 | IVW | 0.041 | $2.3\times10^2$ | 228 |
| Small vessel stroke (OR) | Total | 1.09 (0.77; 1.54) | 0.640 | MR Egger | 0.108 | $3.1\times10^1$ | 75 |
| Small vessel stroke (OR) | LDL-C independent | 0.96 (0.79; 1.16) | 0.651 | MR Egger | 0.007 | $1.1\times10^2$ | 173 |
| Small vessel stroke (OR) | HDL-C independent | 1.00 (0.87; 1.14) | 0.985 | IVW | 0.046 | $2.1\times10^2$ | 138 |
| Small vessel stroke (OR) | LDL-C & HDL-C & TG independent | 0.98 (0.84; 1.14) | 0.789 | IVW | 0.007 | $1.7\times10^2$ | 232 |
| cIMT (mm) | Total | 0.00 (-0.00; 0.01) | 0.336 | IVW | 0.025 | $2.9\times10^5$ | 77 |
| cIMT (mm) | LDL-C independent | -0.01 (-0.02; -0.00) | < 0.001 | MR Egger | < 0.001 | $1.2\times10^5$ | 174 |
| cIMT (mm) | HDL-C independent | 0.00 (-0.00; 0.01) | 0.400 | IVW | < 0.001 | $2.3\times10^5$ | 138 |
| cIMT (mm) | LDL-C & HDL-C & TG independent | -0.01 (-0.01; -0.00) | < 0.001 | IVW | < 0.001 | $1.8\times10^5$ | 233 |
| Carotid plaque (OR) | Total | 1.27 (1.16; 1.39) | < 0.001 | IVW | 0.043 | $4.6\times10^2$ | 71 |
| Carotid plaque (OR) | LDL-C independent | 0.98 (0.85; 1.14) | 0.827 | MR Egger | 0.012 | $1.7\times10^2$ | 163 |

**Table S4:** (Multivariable) Mendelian randomization effect estimates for one standard deviation change in TG *(continued)*

| Outcome | Effect type | Point estimate (95%CI) | P-value | Estimator | Heterogeneity P-value | Precision | No. variants |
|---|---|---|---|---|---|---|---|
| Carotid plaque (OR) | HDL-C independent | 1.18 (1.07; 1.31) | < 0.001 | IVW | 0.006 | $3.6\times10^{2}$ | 132 |
| Carotid plaque (OR) | LDL-C & HDL-C & TG independent | 0.91 (0.81; 1.03) | 0.125 | IVW | 0.004 | $2.9\times10^{2}$ | 222 |
| Glucose (mmol/l) | Total | 0.01 (-0.03; 0.04) | 0.702 | MR Egger | < 0.001 | $2.8\times10^{3}$ | 73 |
| Glucose (mmol/l) | LDL-C independent | 0.03 (0.01; 0.05) | 0.009 | MR Egger | < 0.001 | $8.7\times10^{3}$ | 174 |
| Glucose (mmol/l) | HDL-C independent | -0.02 (-0.03; 0.00) | 0.077 | IVW | < 0.001 | $1.3\times10^{4}$ | 128 |
| Glucose (mmol/l) | LDL-C & HDL-C & TG independent | 0.00 (-0.02; 0.02) | 0.961 | IVW | < 0.001 | $1.1\times10^{4}$ | 225 |
| HbA1c (mmol/mol) | Total | -0.19 (-0.27; -0.11) | < 0.001 | IVW | < 0.001 | $6.3\times10^{2}$ | 71 |
| HbA1c (mmol/mol) | LDL-C independent | 0.01 (-0.10; 0.11) | 0.915 | MR Egger | < 0.001 | $3.2\times10^{2}$ | 170 |
| HbA1c (mmol/mol) | HDL-C independent | -0.25 (-0.34; -0.16) | < 0.001 | IVW | < 0.001 | $4.8\times10^{2}$ | 125 |
| HbA1c (mmol/mol) | LDL-C & HDL-C & TG independent | -0.15 (-0.25; -0.06) | 0.002 | IVW | < 0.001 | $4.1\times10^{2}$ | 220 |
| T2DM (OR) | Total | 1.02 (0.98; 1.06) | 0.313 | IVW | < 0.001 | $3.0\times10^{3}$ | 61 |
| T2DM (OR) | LDL-C independent | 1.13 (1.07; 1.19) | < 0.001 | MR Egger | < 0.001 | $1.4\times10^{3}$ | 145 |
| T2DM (OR) | HDL-C independent | 0.98 (0.94; 1.01) | 0.221 | IVW | < 0.001 | $2.5\times10^{3}$ | 118 |
| T2DM (OR) | LDL-C & HDL-C & TG independent | 1.07 (1.03; 1.12) | 0.002 | IVW | < 0.001 | $1.9\times10^{3}$ | 202 |
| CRP (mg/L) | Total | 0.15 (0.02; 0.28) | 0.025 | MR Egger | 0.002 | $2.3\times10^{2}$ | 69 |
| CRP (mg/L) | LDL-C independent | 0.17 (0.11; 0.23) | < 0.001 | IVW | < 0.001 | $1.1\times10^{3}$ | 154 |
| CRP (mg/L) | HDL-C independent | 0.14 (0.08; 0.20) | < 0.001 | IVW | < 0.001 | $1.0\times10^{3}$ | 124 |
| CRP (mg/L) | LDL-C & HDL-C & TG independent | 0.32 (0.25; 0.38) | < 0.001 | IVW | < 0.001 | $8.3\times10^{2}$ | 210 |
| SBP (mmHg) | Total | 0.66 (0.50; 0.82) | < 0.001 | IVW | 0.006 | $1.4\times10^{2}$ | 63 |
| SBP (mmHg) | LDL-C independent | 0.47 (0.29; 0.66) | < 0.001 | IVW | < 0.001 | $1.1\times10^{2}$ | 140 |
| SBP (mmHg) | HDL-C independent | 0.42 (0.23; 0.61) | < 0.001 | IVW | < 0.001 | $1.0\times10^{2}$ | 112 |
| SBP (mmHg) | LDL-C & HDL-C & TG independent | 0.30 (0.08; 0.53) | 0.007 | IVW | < 0.001 | $7.8\times10^{1}$ | 185 |
| DBP (mmHg) | Total | 0.14 (0.05; 0.24) | 0.003 | IVW | < 0.001 | $4.3\times10^{2}$ | 62 |
| DBP (mmHg) | LDL-C independent | 0.27 (0.13; 0.42) | < 0.001 | MR Egger | < 0.001 | $1.8\times10^{2}$ | 138 |
| DBP (mmHg) | HDL-C independent | -0.02 (-0.13; 0.09) | 0.729 | IVW | < 0.001 | $3.0\times10^{2}$ | 111 |
| DBP (mmHg) | LDL-C & HDL-C & TG independent | 0.10 (-0.03; 0.22) | 0.139 | IVW | < 0.001 | $2.4\times10^{2}$ | 187 |
| PBL (OR) | Total | 1.34 (0.37; 4.80) | 0.653 | MR Egger | 0.011 | 2.4 | 37 |
| PBL (OR) | LDL-C independent | 1.62 (1.25; 2.09) | < 0.001 | IVW | 0.003 | $5.9\times10^{1}$ | 84 |
| PBL (OR) | HDL-C independent | 1.84 (1.21; 2.80) | 0.004 | MR Egger | 0.041 | $2.2\times10^{1}$ | 71 |
| PBL (OR) | LDL-C & HDL-C & TG independent | 2.04 (1.40; 2.99) | < 0.001 | MR Egger | 0.016 | $2.6\times10^{1}$ | 112 |
| BUN (mg/dl) | Total | -0.00 (-0.01; 0.00) | 0.333 | IVW | 0.003 | $1.7\times10^{5}$ | 73 |
| BUN (mg/dl) | LDL-C independent | -0.00 (-0.01; 0.00) | 0.311 | IVW | < 0.001 | $1.5\times10^{5}$ | 167 |
| BUN (mg/dl) | HDL-C independent | -0.01 (-0.01; -0.00) | 0.035 | IVW | 0.003 | $1.3\times10^{5}$ | 131 |
| BUN (mg/dl) | LDL-C & HDL-C & TG independent | -0.01 (-0.01; 0.00) | 0.056 | IVW | < 0.001 | $1.0\times10^{5}$ | 219 |
| eGFR (SD of log(eGFR)) | Total | 0.00 (0.00; 0.00) | 0.011 | IVW | < 0.001 | $1.1\times10^{6}$ | 69 |
| eGFR (SD of log(eGFR)) | LDL-C independent | 0.00 (0.00; 0.01) | 0.004 | MR Egger | < 0.001 | $5.1\times10^{5}$ | 159 |
| eGFR (SD of log(eGFR)) | HDL-C independent | 0.00 (0.00; 0.00) | 0.029 | IVW | < 0.001 | $7.9\times10^{5}$ | 124 |
| eGFR (SD of log(eGFR)) | LDL-C & HDL-C & TG independent | 0.00 (-0.00; 0.00) | 0.067 | IVW | < 0.001 | $6.4\times10^{5}$ | 214 |

**Table S4:** (Multivariable) Mendelian randomization effect estimates for one standard deviation change in TG *(continued)*

| Outcome | Effect type | Point estimate (95%CI) | P-value | Estimator | Heterogeneity P-value | Precision | No. variants |
|---|---|---|---|---|---|---|---|
| CKD (OR) | Total | 0.95 (0.91; 1.00) | 0.032 | IVW | 0.002 | $1.7\times10^3$ | 77 |
| CKD (OR) | LDL-C independent | 0.95 (0.88; 1.02) | 0.187 | MR Egger | 0.004 | $7.0\times10^2$ | 172 |
| CKD (OR) | HDL-C independent | 0.95 (0.90; 1.01) | 0.106 | IVW | < 0.001 | $1.2\times10^3$ | 134 |
| CKD (OR) | LDL-C & HDL-C & TG independent | 0.96 (0.90; 1.02) | 0.216 | IVW | < 0.001 | $9.5\times10^2$ | 227 |
| IBD (OR) | Total | 1.14 (0.91; 1.43) | 0.240 | MR Egger | 0.036 | $7.8\times10^1$ | 75 |
| IBD (OR) | LDL-C independent | 0.99 (0.90; 1.08) | 0.773 | IVW | < 0.001 | $4.5\times10^2$ | 175 |
| IBD (OR) | HDL-C independent | 1.00 (0.91; 1.11) | 0.961 | IVW | 0.004 | $4.0\times10^2$ | 136 |
| IBD (OR) | LDL-C & HDL-C & TG independent | 0.96 (0.86; 1.07) | 0.476 | IVW | < 0.001 | $3.2\times10^2$ | 232 |
| UC (OR) | Total | 0.96 (0.86; 1.07) | 0.481 | IVW | 0.024 | $3.3\times10^2$ | 78 |
| UC (OR) | LDL-C independent | 0.93 (0.83; 1.04) | 0.193 | IVW | 0.002 | $2.9\times10^2$ | 176 |
| UC (OR) | HDL-C independent | 0.95 (0.84; 1.07) | 0.383 | IVW | 0.007 | $2.6\times10^2$ | 138 |
| UC (OR) | LDL-C & HDL-C & TG independent | 0.89 (0.78; 1.02) | 0.099 | IVW | < 0.001 | $2.0\times10^2$ | 233 |
| CD (OR) | Total | 1.15 (0.82; 1.61) | 0.426 | MR Egger | 0.055 | $3.3\times10^1$ | 76 |
| CD (OR) | LDL-C independent | 1.21 (1.07; 1.37) | 0.003 | IVW | 0.043 | $2.4\times10^2$ | 176 |
| CD (OR) | HDL-C independent | 1.17 (1.01; 1.35) | 0.034 | IVW | 0.096 | $1.8\times10^2$ | 137 |
| CD (OR) | LDL-C & HDL-C & TG independent | 1.24 (1.05; 1.45) | 0.009 | IVW | 0.056 | $1.5\times10^2$ | 233 |
| Arthritis (OR) | Total | 1.43 (1.04; 1.96) | 0.029 | MR Egger | 0.299 | $3.8\times10^1$ | 64 |
| Arthritis (OR) | LDL-C independent | 0.96 (0.85; 1.09) | 0.563 | IVW | 0.086 | $2.6\times10^2$ | 144 |
| Arthritis (OR) | HDL-C independent | 1.02 (0.90; 1.16) | 0.716 | IVW | 0.395 | $2.4\times10^2$ | 108 |
| Arthritis (OR) | LDL-C & HDL-C & TG independent | 1.06 (0.90; 1.23) | 0.490 | IVW | 0.143 | $1.6\times10^2$ | 187 |
| ALS (OR) | Total | 0.91 (0.72; 1.17) | 0.473 | MR Egger | 0.182 | $6.5\times10^1$ | 75 |
| ALS (OR) | LDL-C independent | 1.08 (0.98; 1.19) | 0.114 | IVW | 0.171 | $4.0\times10^2$ | 175 |
| ALS (OR) | HDL-C independent | 1.13 (1.01; 1.26) | 0.030 | IVW | 0.053 | $3.2\times10^2$ | 135 |
| ALS (OR) | LDL-C & HDL-C & TG independent | 1.14 (1.01; 1.28) | 0.029 | IVW | 0.210 | $2.9\times10^2$ | 231 |
| MS (OR) | Total | 0.90 (0.79; 1.03) | 0.126 | IVW | 0.941 | $2.2\times10^2$ | 17 |
| MS (OR) | LDL-C independent | 0.87 (0.73; 1.04) | 0.136 | IVW | 0.115 | $1.2\times10^2$ | 33 |
| MS (OR) | HDL-C independent | 1.15 (0.86; 1.54) | 0.342 | MR Egger | 0.089 | $4.5\times10^1$ | 24 |
| MS (OR) | LDL-C & HDL-C & TG independent | 0.90 (0.75; 1.07) | 0.227 | IVW | < 0.001 | $1.2\times10^2$ | 40 |
| Alzheimer (OR) | Total | 1.00 (0.99; 1.02) | 0.611 | IVW | 0.053 | $2.2\times10^4$ | 71 |
| Alzheimer (OR) | LDL-C independent | 0.98 (0.96; 1.00) | 0.127 | MR Egger | < 0.001 | $9.0\times10^3$ | 129 |
| Alzheimer (OR) | HDL-C independent | 0.99 (0.98; 1.01) | 0.312 | IVW | 0.003 | $2.0\times10^4$ | 130 |
| Alzheimer (OR) | LDL-C & HDL-C & TG independent | 0.97 (0.95; 0.99) | < 0.001 | IVW | < 0.001 | $1.4\times10^4$ | 195 |
| Parkinson (OR) | Total | 0.88 (0.67; 1.15) | 0.347 | MR Egger | 0.076 | $5.1\times10^1$ | 75 |
| Parkinson (OR) | LDL-C independent | 0.95 (0.86; 1.06) | 0.388 | IVW | 0.002 | $3.3\times10^2$ | 174 |
| Parkinson (OR) | HDL-C independent | 0.93 (0.83; 1.04) | 0.215 | IVW | 0.008 | $3.1\times10^2$ | 136 |
| Parkinson (OR) | LDL-C & HDL-C & TG independent | 0.99 (0.87; 1.12) | 0.843 | IVW | < 0.001 | $2.4\times10^2$ | 230 |
| Pancreatic cancer (OR) | Total | 0.81 (0.61; 1.07) | 0.143 | IVW | 0.378 | $4.7\times10^1$ | 79 |
| Pancreatic cancer (OR) | LDL-C independent | 0.77 (0.56; 1.04) | 0.088 | IVW | 0.021 | $4.1\times10^1$ | 177 |
| Pancreatic cancer (OR) | HDL-C independent | 0.76 (0.55; 1.05) | 0.092 | IVW | 0.017 | $3.8\times10^1$ | 136 |

**Table S4:** (Multivariable) Mendelian randomization effect estimates for one standard deviation change in TG *(continued)*

| Outcome | Effect type | Point estimate (95%CI) | P-value | Estimator | Heterogeneity P-value | Precision | No. variants |
|---|---|---|---|---|---|---|---|
| Pancreatic cancer (OR) | LDL-C & HDL-C & TG independent | 0.84 (0.59; 1.19) | 0.322 | IVW | < 0.001 | $3.2\times10^{1}$ | 233 |
| Colon cancer (OR) | Total | 1.30 (0.95; 1.80) | 0.106 | MR Egger | 0.060 | $3.7\times10^{1}$ | 77 |
| Colon cancer (OR) | LDL-C independent | 1.03 (0.87; 1.22) | 0.712 | MR Egger | 0.006 | $1.4\times10^{2}$ | 181 |
| Colon cancer (OR) | HDL-C independent | 1.12 (0.98; 1.27) | 0.102 | IVW | < 0.001 | $2.2\times10^{2}$ | 137 |
| Colon cancer (OR) | LDL-C & HDL-C & TG independent | 1.10 (0.95; 1.27) | 0.207 | IVW | < 0.001 | $1.8\times10^{2}$ | 236 |
| Rectal cancer (OR) | Total | 0.98 (0.67; 1.44) | 0.936 | MR Egger | 0.663 | $2.7\times10^{1}$ | 78 |
| Rectal cancer (OR) | LDL-C independent | 1.06 (0.89; 1.26) | 0.517 | IVW | 0.840 | $1.3\times10^{2}$ | 176 |
| Rectal cancer (OR) | HDL-C independent | 0.97 (0.80; 1.16) | 0.706 | IVW | 0.356 | $1.2\times10^{2}$ | 134 |
| Rectal cancer (OR) | LDL-C & HDL-C & TG independent | 0.85 (0.70; 1.04) | 0.109 | IVW | 0.586 | $9.8\times10^{1}$ | 231 |
| Melanoma (OR) | Total | 0.82 (0.65; 1.04) | 0.103 | MR Egger | 0.196 | $6.7\times10^{1}$ | 77 |
| Melanoma (OR) | LDL-C independent | 1.07 (0.97; 1.18) | 0.183 | IVW | 0.024 | $3.9\times10^{2}$ | 176 |
| Melanoma (OR) | HDL-C independent | 1.08 (0.97; 1.19) | 0.143 | IVW | < 0.001 | $3.8\times10^{2}$ | 134 |
| Melanoma (OR) | LDL-C & HDL-C & TG independent | 1.11 (0.99; 1.24) | 0.075 | IVW | < 0.001 | $3.0\times10^{2}$ | 231 |
| Lung cancer (OR) | Total | 0.88 (0.62; 1.24) | 0.459 | MR Egger | 0.854 | $3.3\times10^{1}$ | 78 |
| Lung cancer (OR) | LDL-C independent | 1.04 (0.88; 1.23) | 0.651 | IVW | 0.061 | $1.3\times10^{2}$ | 177 |
| Lung cancer (OR) | HDL-C independent | 0.87 (0.73; 1.03) | 0.111 | IVW | 0.173 | $1.3\times10^{2}$ | 137 |
| Lung cancer (OR) | LDL-C & HDL-C & TG independent | 1.06 (0.88; 1.26) | 0.548 | IVW | 0.022 | $1.2\times10^{2}$ | 235 |

**Table S5:** (Multivariable) Mendelian randomization effect estimates for one standard deviation change in VLDL-C

| Outcome | Effect type | Point estimate (95%CI) | P-value | Estimator | Heterogeneity P-value | Precision | No. variants |
|---|---|---|---|---|---|---|---|
| CHD (OR) | Total | 1.29 (1.19; 1.41) | < 0.001 | IVW | 0.120 | $5.2\times10^{2}$ | 11 |
| CHD (OR) | LDL-C independent | 1.25 (1.14; 1.37) | < 0.001 | IVW | < 0.001 | $4.6\times10^{2}$ | 33 |
| CHD (OR) | HDL-C independent | 1.21 (1.10; 1.32) | < 0.001 | IVW | 0.138 | $4.7\times10^{2}$ | 29 |
| CHD (OR) | TG independent | 1.16 (0.93; 1.46) | 0.191 | IVW | < 0.001 | $7.6\times10^{1}$ | 21 |
| CHD (OR) | LDL-C & HDL-C & TG independent | 1.21 (1.04; 1.40) | 0.014 | IVW | < 0.001 | $1.7\times10^{2}$ | 58 |
| HF (OR) | Total | 1.10 (1.06; 1.15) | < 0.001 | IVW | < 0.001 | $2.3\times10^{3}$ | 27 |
| HF (OR) | LDL-C independent | 1.08 (1.04; 1.13) | < 0.001 | IVW | < 0.001 | $1.9\times10^{3}$ | 103 |
| HF (OR) | HDL-C independent | 1.10 (1.06; 1.15) | < 0.001 | IVW | < 0.001 | $2.1\times10^{3}$ | 89 |
| HF (OR) | TG independent | 1.11 (0.95; 1.29) | 0.186 | IVW | 0.006 | $1.7\times10^{2}$ | 67 |
| HF (OR) | LDL-C & HDL-C & TG independent | 1.10 (1.02; 1.19) | 0.012 | IVW | < 0.001 | $6.7\times10^{2}$ | 188 |
| AF (OR) | Total | 1.03 (0.99; 1.06) | 0.186 | IVW | 0.198 | $2.7\times10^{3}$ | 28 |
| AF (OR) | LDL-C independent | 1.04 (1.00; 1.08) | 0.064 | IVW | 0.017 | $2.6\times10^{3}$ | 107 |
| AF (OR) | HDL-C independent | 1.01 (0.97; 1.05) | 0.628 | IVW | 0.443 | $3.0\times10^{3}$ | 93 |
| AF (OR) | TG independent | 1.08 (0.95; 1.22) | 0.239 | IVW | 0.030 | $2.4\times10^{2}$ | 69 |
| AF (OR) | LDL-C & HDL-C & TG independent | 1.06 (0.99; 1.13) | 0.083 | IVW | 0.002 | $9.2\times10^{2}$ | 195 |

**Table S5:** (Multivariable) Mendelian randomization effect estimates for one standard deviation change in VLDL-C *(continued)*

| Outcome | Effect type | Point estimate (95%CI) | P-value | Estimator | Heterogeneity P-value | Precision | No. variants |
|---|---|---|---|---|---|---|---|
| Any stroke (OR) | Total | 1.02 (0.97; 1.07) | 0.403 | IVW | 0.501 | $1.8\times10^{3}$ | 27 |
| Any stroke (OR) | LDL-C independent | 1.02 (0.97; 1.08) | 0.400 | IVW | 0.002 | $1.4\times10^{3}$ | 103 |
| Any stroke (OR) | HDL-C independent | 1.03 (0.96; 1.11) | 0.366 | MR Egger | 0.008 | $7.9\times10^{2}$ | 92 |
| Any stroke (OR) | TG independent | 0.90 (0.75; 1.08) | 0.258 | IVW | 0.335 | $1.1\times10^{2}$ | 70 |
| Any stroke (OR) | LDL-C & HDL-C & TG independent | 1.02 (0.93; 1.12) | 0.612 | IVW | < 0.001 | $4.5\times10^{2}$ | 193 |
| Any ischemic stroke (OR) | Total | 0.90 (0.74; 1.10) | 0.309 | MR Egger | 0.756 | $9.4\times10^{1}$ | 26 |
| Any ischemic stroke (OR) | LDL-C independent | 1.01 (0.95; 1.07) | 0.769 | IVW | 0.010 | $1.2\times10^{3}$ | 102 |
| Any ischemic stroke (OR) | HDL-C independent | 0.99 (0.94; 1.05) | 0.807 | IVW | 0.013 | $1.3\times10^{3}$ | 91 |
| Any ischemic stroke (OR) | TG independent | 0.89 (0.73; 1.08) | 0.229 | IVW | 0.503 | $1.0\times10^{2}$ | 70 |
| Any ischemic stroke (OR) | LDL-C & HDL-C & TG independent | 1.00 (0.91; 1.11) | 0.926 | IVW | 0.006 | $3.8\times10^{2}$ | 192 |
| Large artery stroke (OR) | Total | 1.10 (0.97; 1.25) | 0.140 | IVW | 0.339 | $2.3\times10^{2}$ | 28 |
| Large artery stroke (OR) | LDL-C independent | 0.97 (0.85; 1.12) | 0.726 | IVW | < 0.001 | $1.9\times10^{2}$ | 105 |
| Large artery stroke (OR) | HDL-C independent | 1.02 (0.89; 1.17) | 0.766 | IVW | 0.006 | $2.2\times10^{2}$ | 92 |
| Large artery stroke (OR) | TG independent | 1.28 (0.78; 2.09) | 0.324 | IVW | 0.320 | $1.6\times10^{1}$ | 72 |
| Large artery stroke (OR) | LDL-C & HDL-C & TG independent | 0.74 (0.57; 0.94) | 0.015 | IVW | 0.006 | $6.3\times10^{1}$ | 194 |
| Cardioembolic stroke (OR) | Total | 0.98 (0.89; 1.08) | 0.698 | IVW | 0.621 | $4.1\times10^{2}$ | 28 |
| Cardioembolic stroke (OR) | LDL-C independent | 1.02 (0.91; 1.14) | 0.724 | IVW | 0.028 | $3.1\times10^{2}$ | 104 |
| Cardioembolic stroke (OR) | HDL-C independent | 1.01 (0.91; 1.12) | 0.845 | IVW | 0.732 | $3.6\times10^{2}$ | 92 |
| Cardioembolic stroke (OR) | TG independent | 0.82 (0.57; 1.19) | 0.297 | IVW | 0.714 | $2.9\times10^{1}$ | 72 |
| Cardioembolic stroke (OR) | LDL-C & HDL-C & TG independent | 1.00 (0.82; 1.21) | 0.981 | IVW | 0.009 | $1.0\times10^{2}$ | 195 |
| Small vessel stroke (OR) | Total | 1.11 (0.99; 1.25) | 0.068 | IVW | 0.737 | $3.0\times10^{2}$ | 28 |
| Small vessel stroke (OR) | LDL-C independent | 1.16 (1.01; 1.34) | 0.035 | IVW | 0.156 | $2.0\times10^{2}$ | 106 |
| Small vessel stroke (OR) | HDL-C independent | 1.17 (0.98; 1.39) | 0.086 | MR Egger | 0.270 | $1.3\times10^{2}$ | 94 |
| Small vessel stroke (OR) | TG independent | 0.73 (0.50; 1.07) | 0.109 | MR Egger | 0.009 | $2.7\times10^{1}$ | 73 |
| Small vessel stroke (OR) | LDL-C & HDL-C & TG independent | 1.03 (0.82; 1.29) | 0.803 | IVW | 0.002 | $7.5\times10^{1}$ | 196 |
| cIMT (mm) | Total | 0.00 (-0.00; 0.01) | 0.111 | IVW | 0.005 | $3.0\times10^{5}$ | 26 |
| cIMT (mm) | LDL-C independent | -0.00 (-0.01; 0.00) | 0.183 | IVW | < 0.001 | $2.4\times10^{5}$ | 105 |
| cIMT (mm) | HDL-C independent | -0.01 (-0.01; -0.00) | 0.025 | MR Egger | < 0.001 | $1.4\times10^{5}$ | 92 |
| cIMT (mm) | TG independent | 0.01 (-0.01; 0.02) | 0.310 | IVW | < 0.001 | $2.5\times10^{4}$ | 72 |
| cIMT (mm) | LDL-C & HDL-C & TG independent | 0.00 (-0.01; 0.01) | 0.891 | IVW | < 0.001 | $7.8\times10^{4}$ | 194 |
| Carotid plaque (OR) | Total | 1.13 (1.04; 1.23) | 0.003 | IVW | 0.014 | $5.6\times10^{2}$ | 26 |
| Carotid plaque (OR) | LDL-C independent | 1.11 (0.99; 1.23) | 0.069 | IVW | 0.083 | $3.2\times10^{2}$ | 103 |
| Carotid plaque (OR) | HDL-C independent | 1.14 (1.04; 1.26) | 0.008 | IVW | 0.094 | $4.0\times10^{2}$ | 89 |
| Carotid plaque (OR) | TG independent | 1.35 (0.99; 1.85) | 0.062 | IVW | 0.031 | $3.9\times10^{1}$ | 70 |
| Carotid plaque (OR) | LDL-C & HDL-C & TG independent | 1.32 (1.12; 1.56) | < 0.001 | IVW | 0.019 | $1.4\times10^{2}$ | 192 |
| Glucose (mmol/l) | Total | -0.01 (-0.08; 0.05) | 0.652 | MR Egger | < 0.001 | $9.3\times10^{2}$ | 23 |
| Glucose (mmol/l) | LDL-C independent | 0.01 (-0.01; 0.03) | 0.192 | IVW | 0.002 | $1.2\times10^{4}$ | 98 |
| Glucose (mmol/l) | HDL-C independent | 0.01 (-0.01; 0.02) | 0.460 | IVW | < 0.001 | $1.4\times10^{4}$ | 83 |
| Glucose (mmol/l) | TG independent | 0.11 (0.06; 0.17) | < 0.001 | IVW | < 0.001 | $1.2\times10^{3}$ | 66 |
| Glucose (mmol/l) | LDL-C & HDL-C & TG independent | 0.02 (-0.01; 0.06) | 0.144 | MR Egger | < 0.001 | $3.7\times10^{3}$ | 180 |

**Table S5:** (Multivariable) Mendelian randomization effect estimates for one standard deviation change in VLDL-C *(continued)*

| Outcome | Effect type | Point estimate (95%CI) | P-value | Estimator | Heterogeneity P-value | Precision | No. variants |
|---|---|---|---|---|---|---|---|
| HbA1c (mmol/mol) | Total | -0.07 (-0.15; 0.01) | 0.103 | IVW | < 0.001 | $5.7\times10^{2}$ | 22 |
| HbA1c (mmol/mol) | LDL-C independent | -0.10 (-0.19; -0.01) | 0.036 | IVW | < 0.001 | $4.6\times10^{2}$ | 97 |
| HbA1c (mmol/mol) | HDL-C independent | -0.18 (-0.29; -0.06) | 0.002 | MR Egger | < 0.001 | $2.9\times10^{2}$ | 82 |
| HbA1c (mmol/mol) | TG independent | 0.28 (-0.02; 0.58) | 0.069 | IVW | < 0.001 | $4.2\times10^{1}$ | 59 |
| HbA1c (mmol/mol) | LDL-C & HDL-C & TG independent | -0.02 (-0.18; 0.13) | 0.746 | IVW | < 0.001 | $1.7\times10^{2}$ | 178 |
| T2DM (OR) | Total | 1.04 (1.01; 1.08) | 0.024 | IVW | < 0.001 | $2.8\times10^{3}$ | 19 |
| T2DM (OR) | LDL-C independent | 1.05 (1.01; 1.09) | 0.008 | IVW | < 0.001 | $2.6\times10^{3}$ | 87 |
| T2DM (OR) | HDL-C independent | 1.04 (1.00; 1.08) | 0.043 | IVW | < 0.001 | $2.9\times10^{3}$ | 79 |
| T2DM (OR) | TG independent | 1.31 (1.16; 1.49) | < 0.001 | IVW | < 0.001 | $2.4\times10^{2}$ | 58 |
| T2DM (OR) | LDL-C & HDL-C & TG independent | 1.11 (1.04; 1.20) | 0.003 | MR Egger | < 0.001 | $7.5\times10^{2}$ | 170 |
| CRP (mg/L) | Total | 0.07 (0.01; 0.13) | 0.014 | IVW | 0.014 | $1.1\times10^{3}$ | 21 |
| CRP (mg/L) | LDL-C independent | 0.16 (0.10; 0.22) | < 0.001 | IVW | < 0.001 | $9.6\times10^{2}$ | 87 |
| CRP (mg/L) | HDL-C independent | 0.04 (-0.04; 0.11) | 0.369 | MR Egger | 0.014 | $6.3\times10^{2}$ | 85 |
| CRP (mg/L) | TG independent | 0.09 (-0.14; 0.31) | 0.458 | IVW | < 0.001 | $7.5\times10^{1}$ | 58 |
| CRP (mg/L) | LDL-C & HDL-C & TG independent | 0.16 (0.05; 0.27) | 0.003 | IVW | < 0.001 | $3.4\times10^{2}$ | 168 |
| SBP (mmHg) | Total | 0.86 (0.71; 1.02) | < 0.001 | IVW | < 0.001 | $1.6\times10^{2}$ | 24 |
| SBP (mmHg) | LDL-C independent | 0.45 (0.27; 0.63) | < 0.001 | IVW | < 0.001 | $1.2\times10^{2}$ | 91 |
| SBP (mmHg) | HDL-C independent | 0.53 (0.36; 0.69) | < 0.001 | IVW | < 0.001 | $1.3\times10^{2}$ | 81 |
| SBP (mmHg) | TG independent | 0.23 (-0.42; 0.88) | 0.488 | IVW | 0.004 | 9.1 | 60 |
| SBP (mmHg) | LDL-C & HDL-C & TG independent | 0.39 (0.09; 0.69) | 0.011 | IVW | < 0.001 | $4.2\times10^{1}$ | 172 |
| DBP (mmHg) | Total | 0.25 (0.16; 0.34) | < 0.001 | IVW | < 0.001 | $4.6\times10^{2}$ | 23 |
| DBP (mmHg) | LDL-C independent | 0.20 (0.10; 0.30) | < 0.001 | IVW | < 0.001 | $3.6\times10^{2}$ | 89 |
| DBP (mmHg) | HDL-C independent | 0.12 (0.03; 0.22) | 0.013 | IVW | 0.034 | $4.2\times10^{2}$ | 80 |
| DBP (mmHg) | TG independent | -0.04 (-0.41; 0.33) | 0.825 | IVW | < 0.001 | $2.8\times10^{1}$ | 58 |
| DBP (mmHg) | LDL-C & HDL-C & TG independent | -0.18 (-0.36; -0.00) | 0.044 | IVW | < 0.001 | $1.2\times10^{2}$ | 166 |
| PBL (OR) | Total | 1.76 (0.41; 7.55) | 0.446 | MR Egger | 0.103 | 1.8 | 17 |
| PBL (OR) | LDL-C independent | 0.97 (0.77; 1.23) | 0.825 | IVW | 0.023 | $7.2\times10^{1}$ | 62 |
| PBL (OR) | HDL-C independent | 1.14 (0.89; 1.46) | 0.294 | IVW | 0.170 | $6.3\times10^{1}$ | 53 |
| PBL (OR) | TG independent | 0.81 (0.41; 1.62) | 0.559 | IVW | 0.003 | 8.0 | 41 |
| PBL (OR) | LDL-C & HDL-C & TG independent | 0.73 (0.51; 1.04) | 0.084 | IVW | < 0.001 | $3.1\times10^{1}$ | 115 |
| BUN (mg/dl) | Total | -0.00 (-0.01; 0.00) | 0.301 | IVW | < 0.001 | $1.7\times10^{5}$ | 23 |
| BUN (mg/dl) | LDL-C independent | -0.00 (-0.01; 0.00) | 0.212 | IVW | < 0.001 | $1.3\times10^{5}$ | 98 |
| BUN (mg/dl) | HDL-C independent | -0.00 (-0.01; 0.00) | 0.664 | IVW | 0.002 | $1.6\times10^{5}$ | 87 |
| BUN (mg/dl) | TG independent | -0.00 (-0.02; 0.01) | 0.647 | IVW | 0.023 | $1.3\times10^{4}$ | 68 |
| BUN (mg/dl) | LDL-C & HDL-C & TG independent | -0.01 (-0.01; 0.00) | 0.223 | IVW | < 0.001 | $4.9\times10^{4}$ | 186 |
| eGFR (SD of log(eGFR)) | Total | 0.01 (-0.00; 0.01) | 0.068 | MR Egger | < 0.001 | $7.6\times10^{4}$ | 24 |
| eGFR (SD of log(eGFR)) | LDL-C independent | 0.00 (0.00; 0.01) | 0.004 | MR Egger | < 0.001 | $5.6\times10^{5}$ | 98 |
| eGFR (SD of log(eGFR)) | HDL-C independent | 0.00 (0.00; 0.01) | 0.012 | MR Egger | 0.005 | $5.5\times10^{5}$ | 85 |
| eGFR (SD of log(eGFR)) | TG independent | 0.01 (-0.00; 0.01) | 0.155 | IVW | < 0.001 | $6.5\times10^{4}$ | 63 |
| eGFR (SD of log(eGFR)) | LDL-C & HDL-C & TG independent | 0.00 (0.00; 0.01) | 0.021 | IVW | < 0.001 | $3.0\times10^{5}$ | 179 |

**Table S5:** (Multivariable) Mendelian randomization effect estimates for one standard deviation change in VLDL-C *(continued)*

| Outcome | Effect type | Point estimate (95%CI) | P-value | Estimator | Heterogeneity P-value | Precision | No. variants |
|---|---|---|---|---|---|---|---|
| CKD (OR) | Total | 1.03 (0.98; 1.07) | 0.296 | IVW | 0.028 | $1.7\times10^3$ | 26 |
| CKD (OR) | LDL-C independent | 1.03 (0.97; 1.09) | 0.299 | IVW | 0.004 | $1.3\times10^3$ | 103 |
| CKD (OR) | HDL-C independent | 1.02 (0.96; 1.08) | 0.567 | IVW | 0.051 | $1.2\times10^3$ | 87 |
| CKD (OR) | TG independent | 0.98 (0.82; 1.17) | 0.803 | IVW | 0.002 | $1.2\times10^2$ | 70 |
| CKD (OR) | LDL-C & HDL-C & TG independent | 1.01 (0.92; 1.11) | 0.797 | IVW | < 0.001 | $4.5\times10^2$ | 191 |
| IBD (OR) | Total | 0.86 (0.63; 1.18) | 0.358 | MR Egger | 0.031 | $3.8\times10^1$ | 26 |
| IBD (OR) | LDL-C independent | 1.00 (0.90; 1.10) | 0.977 | IVW | 0.006 | $3.8\times10^2$ | 102 |
| IBD (OR) | HDL-C independent | 0.99 (0.91; 1.08) | 0.859 | IVW | 0.009 | $4.8\times10^2$ | 90 |
| IBD (OR) | TG independent | 0.95 (0.70; 1.29) | 0.722 | IVW | < 0.001 | $4.1\times10^1$ | 71 |
| IBD (OR) | LDL-C & HDL-C & TG independent | 0.89 (0.76; 1.06) | 0.193 | IVW | < 0.001 | $1.4\times10^2$ | 193 |
| UC (OR) | Total | 0.77 (0.52; 1.15) | 0.201 | MR Egger | 0.032 | $2.4\times10^1$ | 27 |
| UC (OR) | LDL-C independent | 0.92 (0.81; 1.05) | 0.209 | IVW | 0.008 | $2.4\times10^2$ | 104 |
| UC (OR) | HDL-C independent | 0.98 (0.84; 1.15) | 0.831 | MR Egger | 0.010 | $1.6\times10^2$ | 92 |
| UC (OR) | TG independent | 0.90 (0.62; 1.32) | 0.600 | IVW | 0.021 | $2.6\times10^1$ | 72 |
| UC (OR) | LDL-C & HDL-C & TG independent | 0.86 (0.70; 1.06) | 0.161 | IVW | < 0.001 | $8.7\times10^1$ | 195 |
| CD (OR) | Total | 1.02 (0.66; 1.56) | 0.938 | MR Egger | 0.048 | $2.1\times10^1$ | 26 |
| CD (OR) | LDL-C independent | 1.11 (0.96; 1.27) | 0.158 | IVW | 0.376 | $2.0\times10^2$ | 102 |
| CD (OR) | HDL-C independent | 1.09 (0.95; 1.24) | 0.235 | IVW | 0.077 | $2.1\times10^2$ | 90 |
| CD (OR) | TG independent | 0.72 (0.48; 1.09) | 0.122 | IVW | 0.039 | $2.2\times10^1$ | 72 |
| CD (OR) | LDL-C & HDL-C & TG independent | 1.00 (0.78; 1.28) | 0.978 | IVW | 0.054 | $6.3\times10^1$ | 194 |
| Arthritis (OR) | Total | 0.97 (0.87; 1.09) | 0.604 | IVW | 0.155 | $3.1\times10^2$ | 26 |
| Arthritis (OR) | LDL-C independent | 0.99 (0.88; 1.10) | 0.814 | IVW | < 0.001 | $3.0\times10^2$ | 95 |
| Arthritis (OR) | HDL-C independent | 1.02 (0.86; 1.22) | 0.781 | MR Egger | 0.148 | $1.3\times10^2$ | 79 |
| Arthritis (OR) | TG independent | 1.30 (0.84; 1.99) | 0.235 | IVW | 0.234 | $2.1\times10^1$ | 62 |
| Arthritis (OR) | LDL-C & HDL-C & TG independent | 1.11 (0.89; 1.38) | 0.344 | IVW | 0.022 | $8.3\times10^1$ | 166 |
| ALS (OR) | Total | 1.07 (0.98; 1.17) | 0.131 | IVW | 0.510 | $5.1\times10^2$ | 28 |
| ALS (OR) | LDL-C independent | 1.00 (0.90; 1.10) | 0.970 | IVW | 0.491 | $3.9\times10^2$ | 106 |
| ALS (OR) | HDL-C independent | 1.00 (0.91; 1.09) | 0.924 | IVW | 0.259 | $4.3\times10^2$ | 92 |
| ALS (OR) | TG independent | 0.81 (0.58; 1.15) | 0.237 | IVW | 0.242 | $3.3\times10^1$ | 69 |
| ALS (OR) | LDL-C & HDL-C & TG independent | 0.78 (0.66; 0.93) | 0.004 | IVW | 0.428 | $1.4\times10^2$ | 191 |
| MS (OR) | Total | 0.92 (0.81; 1.03) | 0.154 | IVW | 0.758 | $2.7\times10^2$ | 11 |
| MS (OR) | LDL-C independent | 0.87 (0.73; 1.04) | 0.129 | IVW | 0.155 | $1.2\times10^2$ | 27 |
| MS (OR) | HDL-C independent | 0.89 (0.74; 1.06) | 0.198 | IVW | 0.187 | $1.2\times10^2$ | 19 |
| MS (OR) | TG independent | 1.23 (0.60; 2.53) | 0.576 | IVW | 0.902 | 7.4 | 17 |
| MS (OR) | LDL-C & HDL-C & TG independent | 0.98 (0.65; 1.46) | 0.910 | IVW | 0.007 | $2.4\times10^1$ | 38 |
| Alzheimer (OR) | Total | 1.02 (1.00; 1.03) | 0.014 | IVW | 0.839 | $2.1\times10^4$ | 22 |
| Alzheimer (OR) | LDL-C independent | 1.00 (0.98; 1.02) | 0.792 | MR Egger | < 0.001 | $9.9\times10^3$ | 67 |
| Alzheimer (OR) | HDL-C independent | 1.01 (1.00; 1.03) | 0.087 | IVW | 0.094 | $1.6\times10^4$ | 86 |
| Alzheimer (OR) | TG independent | 1.02 (0.97; 1.08) | 0.387 | IVW | 0.072 | $1.5\times10^3$ | 63 |
| Alzheimer (OR) | LDL-C & HDL-C & TG independent | 0.96 (0.94; 0.99) | 0.007 | IVW | < 0.001 | $5.3\times10^3$ | 150 |

**Table S5:** (Multivariable) Mendelian randomization effect estimates for one standard deviation change in VLDL-C *(continued)*

| Outcome | Effect type | Point estimate (95%CI) | P-value | Estimator | Heterogeneity P-value | Precision | No. variants |
|---|---|---|---|---|---|---|---|
| Parkinson (OR) | Total | 1.17 (0.80; 1.71) | 0.424 | MR Egger | 0.781 | $2.6\times10^{1}$ | 24 |
| Parkinson (OR) | LDL-C independent | 1.05 (0.89; 1.23) | 0.570 | MR Egger | 0.070 | $1.5\times10^{2}$ | 102 |
| Parkinson (OR) | HDL-C independent | 1.02 (0.93; 1.13) | 0.631 | IVW | 0.434 | $3.9\times10^{2}$ | 89 |
| Parkinson (OR) | TG independent | 1.39 (0.99; 1.95) | 0.059 | IVW | 0.021 | $3.3\times10^{1}$ | 71 |
| Parkinson (OR) | LDL-C & HDL-C & TG independent | 1.22 (1.02; 1.46) | 0.029 | IVW | 0.005 | $1.2\times10^{2}$ | 192 |
| Pancreatic cancer (OR) | Total | 0.76 (0.57; 1.00) | 0.047 | IVW | 0.951 | $5.0\times10^{1}$ | 27 |
| Pancreatic cancer (OR) | LDL-C independent | 0.73 (0.53; 1.00) | 0.052 | IVW | 0.003 | $3.7\times10^{1}$ | 104 |
| Pancreatic cancer (OR) | HDL-C independent | 0.76 (0.55; 1.06) | 0.103 | IVW | 0.248 | $3.7\times10^{1}$ | 87 |
| Pancreatic cancer (OR) | TG independent | 1.30 (0.41; 4.10) | 0.650 | IVW | 0.234 | 2.9 | 70 |
| Pancreatic cancer (OR) | LDL-C & HDL-C & TG independent | 0.33 (0.18; 0.62) | < 0.001 | MR Egger | < 0.001 | $1.0\times10^{1}$ | 192 |
| Colon cancer (OR) | Total | 1.64 (1.02; 2.64) | 0.041 | MR Egger | 0.823 | $1.7\times10^{1}$ | 25 |
| Colon cancer (OR) | LDL-C independent | 0.94 (0.82; 1.07) | 0.350 | IVW | 0.020 | $2.1\times10^{2}$ | 101 |
| Colon cancer (OR) | HDL-C independent | 1.13 (0.94; 1.37) | 0.192 | MR Egger | 0.261 | $1.1\times10^{2}$ | 90 |
| Colon cancer (OR) | TG independent | 1.77 (1.14; 2.75) | 0.011 | IVW | 0.017 | $2.0\times10^{1}$ | 70 |
| Colon cancer (OR) | LDL-C & HDL-C & TG independent | 1.13 (0.90; 1.43) | 0.303 | IVW | 0.011 | $7.0\times10^{1}$ | 188 |
| Rectal cancer (OR) | Total | 1.34 (1.15; 1.56) | < 0.001 | IVW | 0.485 | $1.6\times10^{2}$ | 27 |
| Rectal cancer (OR) | LDL-C independent | 1.20 (1.00; 1.43) | 0.049 | IVW | 0.870 | $1.2\times10^{2}$ | 102 |
| Rectal cancer (OR) | HDL-C independent | 1.48 (1.15; 1.91) | 0.002 | MR Egger | 0.265 | $6.0\times10^{1}$ | 89 |
| Rectal cancer (OR) | TG independent | 1.74 (0.95; 3.16) | 0.071 | IVW | 0.463 | $1.1\times10^{1}$ | 70 |
| Rectal cancer (OR) | LDL-C & HDL-C & TG independent | 1.16 (0.84; 1.58) | 0.372 | IVW | 0.358 | $3.8\times10^{1}$ | 188 |
| Melanoma (OR) | Total | 0.99 (0.68; 1.43) | 0.937 | MR Egger | 0.339 | $2.8\times10^{1}$ | 26 |
| Melanoma (OR) | LDL-C independent | 1.00 (0.90; 1.10) | 0.945 | IVW | 0.038 | $3.7\times10^{2}$ | 103 |
| Melanoma (OR) | HDL-C independent | 1.05 (0.95; 1.16) | 0.314 | IVW | < 0.001 | $4.1\times10^{2}$ | 88 |
| Melanoma (OR) | TG independent | 1.10 (0.79; 1.55) | 0.573 | IVW | 0.002 | $3.4\times10^{1}$ | 70 |
| Melanoma (OR) | LDL-C & HDL-C & TG independent | 1.00 (0.84; 1.19) | 0.971 | IVW | < 0.001 | $1.2\times10^{2}$ | 191 |
| Lung cancer (OR) | Total | 0.89 (0.77; 1.02) | 0.104 | IVW | 0.505 | $2.0\times10^{2}$ | 27 |
| Lung cancer (OR) | LDL-C independent | 0.90 (0.76; 1.08) | 0.251 | IVW | 0.262 | $1.2\times10^{2}$ | 102 |
| Lung cancer (OR) | HDL-C independent | 0.96 (0.82; 1.13) | 0.642 | IVW | 0.016 | $1.5\times10^{2}$ | 88 |
| Lung cancer (OR) | TG independent | 0.98 (0.60; 1.59) | 0.938 | IVW | 0.906 | $1.6\times10^{1}$ | 71 |
| Lung cancer (OR) | LDL-C & HDL-C & TG independent | 0.98 (0.73; 1.32) | 0.889 | IVW | 0.233 | $4.3\times10^{1}$ | 191 |

**Table S6:** (Multivariable) Mendelian randomization effect estimates for one standard deviation change in IDL-C

| Outcome | Effect type | Point estimate (95%CI) | P-value | Estimator | Heterogeneity P-value | Precision | No. variants |
|---|---|---|---|---|---|---|---|
| CHD (OR) | Total | 1.52 (1.44; 1.61) | < 0.001 | IVW | < 0.001 | $1.2\times10^{3}$ | 29 |
| CHD (OR) | LDL-C independent | 1.71 (1.09; 2.70) | 0.020 | IVW | < 0.001 | $1.9\times10^{1}$ | 32 |
| CHD (OR) | HDL-C independent | 1.57 (1.49; 1.66) | < 0.001 | IVW | < 0.001 | $1.4\times10^{3}$ | 51 |
| CHD (OR) | TG independent | 1.27 (1.18; 1.37) | < 0.001 | MR Egger | < 0.001 | $6.4\times10^{2}$ | 48 |

**Table S6:** (Multivariable) Mendelian randomization effect estimates for one standard deviation change in IDL-C *(continued)*

| Outcome | Effect type | Point estimate (95%CI) | P-value | Estimator | Heterogeneity P-value | Precision | No. variants |
|---|---|---|---|---|---|---|---|
| CHD (OR) | LDL-C & HDL-C & TG independent | 1.42 (0.98; 2.05) | 0.062 | IVW | < 0.001 | $2.9\times10^{1}$ | 62 |
| HF (OR) | Total | 1.03 (1.00; 1.06) | 0.077 | IVW | < 0.001 | $4.3\times10^{3}$ | 107 |
| HF (OR) | LDL-C independent | 1.22 (0.97; 1.54) | 0.095 | MR Egger | < 0.001 | $7.2\times10^{1}$ | 130 |
| HF (OR) | HDL-C independent | 1.04 (1.01; 1.07) | 0.011 | IVW | < 0.001 | $4.7\times10^{3}$ | 179 |
| HF (OR) | TG independent | 0.97 (0.92; 1.01) | 0.168 | MR Egger | < 0.001 | $1.7\times10^{3}$ | 158 |
| HF (OR) | LDL-C & HDL-C & TG independent | 1.09 (0.93; 1.28) | 0.267 | IVW | < 0.001 | $1.6\times10^{2}$ | 236 |
| AF (OR) | Total | 1.01 (0.98; 1.04) | 0.495 | IVW | 0.100 | $4.6\times10^{3}$ | 115 |
| AF (OR) | LDL-C independent | 1.20 (0.98; 1.47) | 0.082 | MR Egger | 0.016 | $9.4\times10^{1}$ | 142 |
| AF (OR) | HDL-C independent | 1.02 (1.00; 1.05) | 0.107 | IVW | 0.135 | $5.5\times10^{3}$ | 191 |
| AF (OR) | TG independent | 1.01 (0.98; 1.04) | 0.415 | IVW | < 0.001 | $4.7\times10^{3}$ | 176 |
| AF (OR) | LDL-C & HDL-C & TG independent | 1.16 (1.02; 1.32) | 0.021 | IVW | < 0.001 | $2.3\times10^{2}$ | 256 |
| Any stroke (OR) | Total | 1.06 (1.02; 1.10) | < 0.001 | IVW | < 0.001 | $3.1\times10^{3}$ | 111 |
| Any stroke (OR) | LDL-C independent | 0.96 (0.73; 1.25) | 0.764 | IVW | < 0.001 | $5.4\times10^{1}$ | 132 |
| Any stroke (OR) | HDL-C independent | 1.05 (1.01; 1.09) | 0.006 | IVW | < 0.001 | $3.2\times10^{3}$ | 185 |
| Any stroke (OR) | TG independent | 1.00 (0.95; 1.06) | 0.897 | MR Egger | < 0.001 | $1.4\times10^{3}$ | 168 |
| Any stroke (OR) | LDL-C & HDL-C & TG independent | 1.04 (0.86; 1.26) | 0.685 | IVW | < 0.001 | $1.1\times10^{2}$ | 242 |
| Any ischemic stroke (OR) | Total | 1.06 (1.02; 1.10) | 0.003 | IVW | < 0.001 | $2.7\times10^{3}$ | 112 |
| Any ischemic stroke (OR) | LDL-C independent | 1.09 (0.82; 1.46) | 0.544 | IVW | < 0.001 | $4.6\times10^{1}$ | 133 |
| Any ischemic stroke (OR) | HDL-C independent | 1.04 (1.00; 1.08) | 0.033 | IVW | < 0.001 | $2.8\times10^{3}$ | 187 |
| Any ischemic stroke (OR) | TG independent | 1.00 (0.95; 1.06) | 0.866 | MR Egger | < 0.001 | $1.2\times10^{3}$ | 168 |
| Any ischemic stroke (OR) | LDL-C & HDL-C & TG independent | 1.00 (0.82; 1.23) | 0.985 | IVW | < 0.001 | $9.3\times10^{1}$ | 243 |
| Large artery stroke (OR) | Total | 1.05 (0.95; 1.16) | 0.346 | IVW | < 0.001 | $4.0\times10^{2}$ | 108 |
| Large artery stroke (OR) | LDL-C independent | 0.36 (0.17; 0.74) | 0.006 | IVW | < 0.001 | 7.2 | 133 |
| Large artery stroke (OR) | HDL-C independent | 1.04 (0.94; 1.14) | 0.437 | IVW | < 0.001 | $4.2\times10^{2}$ | 185 |
| Large artery stroke (OR) | TG independent | 1.08 (0.97; 1.21) | 0.182 | IVW | < 0.001 | $3.1\times10^{2}$ | 166 |
| Large artery stroke (OR) | LDL-C & HDL-C & TG independent | 0.70 (0.42; 1.17) | 0.177 | IVW | < 0.001 | $1.5\times10^{1}$ | 244 |
| Cardioembolic stroke (OR) | Total | 0.96 (0.89; 1.04) | 0.332 | IVW | 0.033 | $6.8\times10^{2}$ | 111 |
| Cardioembolic stroke (OR) | LDL-C independent | 1.24 (0.71; 2.17) | 0.458 | IVW | 0.006 | $1.2\times10^{1}$ | 135 |
| Cardioembolic stroke (OR) | HDL-C independent | 0.95 (0.89; 1.02) | 0.189 | IVW | 0.006 | $7.2\times10^{2}$ | 187 |
| Cardioembolic stroke (OR) | TG independent | 0.99 (0.90; 1.08) | 0.753 | IVW | 0.092 | $4.6\times10^{2}$ | 168 |
| Cardioembolic stroke (OR) | LDL-C & HDL-C & TG independent | 1.43 (0.95; 2.13) | 0.083 | IVW | 0.004 | $2.4\times10^{1}$ | 244 |
| Small vessel stroke (OR) | Total | 1.00 (0.91; 1.10) | 0.956 | IVW | 0.291 | $4.6\times10^{2}$ | 112 |
| Small vessel stroke (OR) | LDL-C independent | 0.83 (0.40; 1.71) | 0.608 | IVW | 0.054 | 7.3 | 137 |
| Small vessel stroke (OR) | HDL-C independent | 0.96 (0.84; 1.09) | 0.543 | MR Egger | 0.099 | $2.2\times10^{2}$ | 186 |
| Small vessel stroke (OR) | TG independent | 0.83 (0.72; 0.95) | 0.008 | MR Egger | 0.024 | $2.0\times10^{2}$ | 167 |
| Small vessel stroke (OR) | LDL-C & HDL-C & TG independent | 1.10 (0.68; 1.76) | 0.702 | IVW | 0.002 | $1.7\times10^{1}$ | 245 |
| cIMT (mm) | Total | 0.01 (0.01; 0.02) | < 0.001 | MR Egger | < 0.001 | $1.4\times10^{5}$ | 117 |
| cIMT (mm) | LDL-C independent | 0.00 (-0.02; 0.02) | 0.969 | IVW | < 0.001 | $9.3\times10^{3}$ | 137 |
| cIMT (mm) | HDL-C independent | 0.02 (0.01; 0.02) | < 0.001 | IVW | < 0.001 | $5.4\times10^{5}$ | 186 |

**Table S6:** (Multivariable) Mendelian randomization effect estimates for one standard deviation change in IDL-C *(continued)*

| Outcome | Effect type | Point estimate (95%CI) | P-value | Estimator | Heterogeneity P-value | Precision | No. variants |
|---|---|---|---|---|---|---|---|
| cIMT (mm) | TG independent | 0.02 (0.01; 0.02) | < 0.001 | MR Egger | < 0.001 | $2.1\times10^5$ | 169 |
| cIMT (mm) | LDL-C & HDL-C & TG independent | 0.02 (0.01; 0.03) | 0.006 | IVW | < 0.001 | $1.8\times10^4$ | 247 |
| Carotid plaque (OR) | Total | 1.38 (1.29; 1.48) | < 0.001 | IVW | 0.008 | $8.4\times10^2$ | 105 |
| Carotid plaque (OR) | LDL-C independent | 0.59 (0.35; 1.01) | 0.052 | MR Egger | 0.023 | $1.4\times10^1$ | 129 |
| Carotid plaque (OR) | HDL-C independent | 1.41 (1.32; 1.51) | < 0.001 | IVW | 0.011 | $8.5\times10^2$ | 178 |
| Carotid plaque (OR) | TG independent | 1.32 (1.18; 1.48) | < 0.001 | MR Egger | 0.002 | $3.1\times10^2$ | 156 |
| Carotid plaque (OR) | LDL-C & HDL-C & TG independent | 1.22 (0.86; 1.73) | 0.275 | MR Egger | 0.013 | $3.1\times10^1$ | 239 |
| Glucose (mmol/l) | Total | -0.01 (-0.02; 0.00) | 0.194 | IVW | 0.010 | $3.7\times10^4$ | 110 |
| Glucose (mmol/l) | LDL-C independent | 0.09 (0.01; 0.17) | 0.033 | IVW | 0.039 | $5.8\times10^2$ | 135 |
| Glucose (mmol/l) | HDL-C independent | -0.01 (-0.02; 0.00) | 0.115 | IVW | < 0.001 | $3.6\times10^4$ | 179 |
| Glucose (mmol/l) | TG independent | -0.01 (-0.02; 0.01) | 0.487 | MR Egger | < 0.001 | $1.5\times10^4$ | 164 |
| Glucose (mmol/l) | LDL-C & HDL-C & TG independent | 0.07 (0.01; 0.12) | 0.020 | IVW | < 0.001 | $1.3\times10^3$ | 241 |
| HbA1c (mmol/mol) | Total | -0.10 (-0.21; 0.01) | 0.083 | MR Egger | < 0.001 | $3.2\times10^2$ | 106 |
| HbA1c (mmol/mol) | LDL-C independent | 0.71 (0.30; 1.12) | < 0.001 | MR Egger | < 0.001 | $2.3\times10^1$ | 132 |
| HbA1c (mmol/mol) | HDL-C independent | -0.12 (-0.17; -0.06) | < 0.001 | IVW | < 0.001 | $1.4\times10^3$ | 174 |
| HbA1c (mmol/mol) | TG independent | -0.02 (-0.08; 0.04) | 0.535 | IVW | < 0.001 | $1.0\times10^3$ | 161 |
| HbA1c (mmol/mol) | LDL-C & HDL-C & TG independent | 0.18 (-0.10; 0.46) | 0.215 | IVW | < 0.001 | $4.9\times10^1$ | 237 |
| T2DM (OR) | Total | 0.93 (0.91; 0.96) | < 0.001 | IVW | < 0.001 | $6.4\times10^3$ | 96 |
| T2DM (OR) | LDL-C independent | 1.23 (1.01; 1.49) | 0.039 | MR Egger | < 0.001 | $1.0\times10^2$ | 122 |
| T2DM (OR) | HDL-C independent | 0.93 (0.91; 0.95) | < 0.001 | IVW | < 0.001 | $6.7\times10^3$ | 159 |
| T2DM (OR) | TG independent | 0.93 (0.91; 0.96) | < 0.001 | IVW | < 0.001 | $5.0\times10^3$ | 142 |
| T2DM (OR) | LDL-C & HDL-C & TG independent | 0.85 (0.74; 0.97) | 0.016 | MR Egger | < 0.001 | $2.1\times10^2$ | 215 |
| CRP (mg/L) | Total | -0.20 (-0.28; -0.12) | < 0.001 | MR Egger | < 0.001 | $5.7\times10^2$ | 100 |
| CRP (mg/L) | LDL-C independent | 0.67 (0.31; 1.03) | < 0.001 | MR Egger | < 0.001 | $2.9\times10^1$ | 114 |
| CRP (mg/L) | HDL-C independent | -0.06 (-0.10; -0.02) | 0.002 | IVW | < 0.001 | $2.4\times10^3$ | 164 |
| CRP (mg/L) | TG independent | -0.10 (-0.16; -0.04) | 0.002 | MR Egger | < 0.001 | $1.0\times10^3$ | 149 |
| CRP (mg/L) | LDL-C & HDL-C & TG independent | 0.29 (0.08; 0.50) | 0.007 | IVW | < 0.001 | $8.7\times10^1$ | 226 |
| SBP (mmHg) | Total | 0.47 (0.35; 0.60) | < 0.001 | IVW | < 0.001 | $2.5\times10^2$ | 85 |
| SBP (mmHg) | LDL-C independent | -0.20 (-1.25; 0.86) | 0.717 | MR Egger | < 0.001 | 3.4 | 99 |
| SBP (mmHg) | HDL-C independent | 0.49 (0.37; 0.61) | < 0.001 | IVW | < 0.001 | $2.6\times10^2$ | 145 |
| SBP (mmHg) | TG independent | 0.24 (0.10; 0.39) | < 0.001 | IVW | < 0.001 | $1.8\times10^2$ | 130 |
| SBP (mmHg) | LDL-C & HDL-C & TG independent | 0.11 (-0.54; 0.77) | 0.733 | IVW | < 0.001 | 9.0 | 191 |
| DBP (mmHg) | Total | 0.03 (-0.04; 0.10) | 0.436 | IVW | < 0.001 | $7.4\times10^2$ | 83 |
| DBP (mmHg) | LDL-C independent | 0.16 (-0.45; 0.77) | 0.611 | MR Egger | < 0.001 | $1.0\times10^1$ | 104 |
| DBP (mmHg) | HDL-C independent | 0.05 (-0.06; 0.15) | 0.367 | MR Egger | < 0.001 | $3.5\times10^2$ | 146 |
| DBP (mmHg) | TG independent | -0.15 (-0.27; -0.03) | 0.012 | MR Egger | < 0.001 | $2.7\times10^2$ | 127 |
| DBP (mmHg) | LDL-C & HDL-C & TG independent | 0.06 (-0.32; 0.44) | 0.761 | IVW | < 0.001 | $2.7\times10^1$ | 191 |
| PBL (OR) | Total | 2.25 (1.23; 4.14) | 0.009 | MR Egger | 0.519 | $1.0\times10^1$ | 59 |
| PBL (OR) | LDL-C independent | 1.20 (0.32; 4.48) | 0.784 | IVW | 0.029 | 2.2 | 63 |

**Table S6:** (Multivariable) Mendelian randomization effect estimates for one standard deviation change in IDL-C *(continued)*

| Outcome | Effect type | Point estimate (95%CI) | P-value | Estimator | Heterogeneity P-value | Precision | No. variants |
|---|---|---|---|---|---|---|---|
| PBL (OR) | HDL-C independent | 1.15 (0.95; 1.39) | 0.150 | IVW | 0.247 | $1.1\times10^{2}$ | 92 |
| PBL (OR) | TG independent | 0.97 (0.78; 1.21) | 0.794 | IVW | 0.027 | $8.2\times10^{1}$ | 86 |
| PBL (OR) | LDL-C & HDL-C & TG independent | 0.60 (0.22; 1.64) | 0.316 | MR Egger | 0.057 | 3.8 | 119 |
| BUN (mg/dl) | Total | 0.00 (-0.00; 0.01) | 0.230 | IVW | < 0.001 | $3.2\times10^{5}$ | 108 |
| BUN (mg/dl) | LDL-C independent | 0.05 (0.02; 0.07) | < 0.001 | IVW | < 0.001 | $5.4\times10^{3}$ | 130 |
| BUN (mg/dl) | HDL-C independent | 0.00 (-0.00; 0.01) | 0.071 | IVW | < 0.001 | $3.3\times10^{5}$ | 174 |
| BUN (mg/dl) | TG independent | 0.00 (-0.01; 0.01) | 0.895 | MR Egger | < 0.001 | $1.3\times10^{5}$ | 164 |
| BUN (mg/dl) | LDL-C & HDL-C & TG independent | 0.02 (0.00; 0.04) | 0.019 | IVW | < 0.001 | $1.1\times10^{4}$ | 230 |
| eGFR (SD of log(eGFR)) | Total | -0.00 (-0.00; 0.00) | 0.479 | IVW | < 0.001 | $2.0\times10^{6}$ | 97 |
| eGFR (SD of log(eGFR)) | LDL-C independent | -0.00 (-0.01; 0.01) | 0.729 | IVW | < 0.001 | $3.7\times10^{4}$ | 125 |
| eGFR (SD of log(eGFR)) | HDL-C independent | 0.00 (0.00; 0.00) | 0.032 | MR Egger | < 0.001 | $1.1\times10^{6}$ | 168 |
| eGFR (SD of log(eGFR)) | TG independent | 0.00 (-0.00; 0.00) | 0.868 | IVW | < 0.001 | $1.6\times10^{6}$ | 155 |
| eGFR (SD of log(eGFR)) | LDL-C & HDL-C & TG independent | -0.01 (-0.02; -0.00) | 0.022 | IVW | < 0.001 | $7.5\times10^{4}$ | 226 |
| CKD (OR) | Total | 0.99 (0.95; 1.02) | 0.500 | IVW | 0.006 | $3.0\times10^{3}$ | 111 |
| CKD (OR) | LDL-C independent | 0.92 (0.70; 1.19) | 0.510 | IVW | 0.005 | $5.5\times10^{1}$ | 134 |
| CKD (OR) | HDL-C independent | 0.92 (0.88; 0.97) | 0.003 | MR Egger | 0.091 | $1.4\times10^{3}$ | 182 |
| CKD (OR) | TG independent | 0.98 (0.94; 1.02) | 0.315 | IVW | < 0.001 | $2.4\times10^{3}$ | 169 |
| CKD (OR) | LDL-C & HDL-C & TG independent | 1.09 (0.91; 1.32) | 0.348 | IVW | < 0.001 | $1.1\times10^{2}$ | 242 |
| IBD (OR) | Total | 1.11 (1.04; 1.18) | 0.002 | IVW | < 0.001 | $9.0\times10^{2}$ | 109 |
| IBD (OR) | LDL-C independent | 0.20 (0.12; 0.34) | < 0.001 | MR Egger | 0.036 | $1.4\times10^{1}$ | 133 |
| IBD (OR) | HDL-C independent | 1.12 (1.05; 1.20) | < 0.001 | IVW | < 0.001 | $9.0\times10^{2}$ | 182 |
| IBD (OR) | TG independent | 1.20 (1.09; 1.32) | < 0.001 | MR Egger | < 0.001 | $4.2\times10^{2}$ | 168 |
| IBD (OR) | LDL-C & HDL-C & TG independent | 0.46 (0.33; 0.64) | < 0.001 | IVW | < 0.001 | $3.6\times10^{1}$ | 245 |
| UC (OR) | Total | 1.37 (1.15; 1.63) | < 0.001 | MR Egger | 0.170 | $1.3\times10^{2}$ | 117 |
| UC (OR) | LDL-C independent | 0.30 (0.15; 0.59) | < 0.001 | MR Egger | 0.323 | 8.4 | 134 |
| UC (OR) | HDL-C independent | 1.07 (0.98; 1.15) | 0.121 | IVW | < 0.001 | $6.0\times10^{2}$ | 188 |
| UC (OR) | TG independent | 1.09 (1.00; 1.19) | 0.051 | IVW | < 0.001 | $4.9\times10^{2}$ | 173 |
| UC (OR) | LDL-C & HDL-C & TG independent | 0.38 (0.25; 0.57) | < 0.001 | IVW | 0.011 | $2.3\times10^{1}$ | 248 |
| CD (OR) | Total | 1.04 (0.86; 1.25) | 0.698 | MR Egger | 0.002 | $1.1\times10^{2}$ | 111 |
| CD (OR) | LDL-C independent | 0.25 (0.12; 0.51) | < 0.001 | IVW | 0.160 | 7.6 | 136 |
| CD (OR) | HDL-C independent | 1.14 (1.04; 1.24) | 0.005 | IVW | 0.033 | $4.9\times10^{2}$ | 184 |
| CD (OR) | TG independent | 1.11 (1.01; 1.23) | 0.032 | IVW | < 0.001 | $4.0\times10^{2}$ | 171 |
| CD (OR) | LDL-C & HDL-C & TG independent | 0.53 (0.33; 0.86) | 0.010 | IVW | 0.067 | $1.7\times10^{1}$ | 248 |
| Arthritis (OR) | Total | 0.93 (0.86; 1.02) | 0.123 | IVW | 0.008 | $5.1\times10^{2}$ | 91 |
| Arthritis (OR) | LDL-C independent | 0.67 (0.34; 1.33) | 0.249 | IVW | 0.014 | 8.1 | 107 |
| Arthritis (OR) | HDL-C independent | 0.96 (0.89; 1.04) | 0.351 | IVW | 0.042 | $5.8\times10^{2}$ | 151 |
| Arthritis (OR) | TG independent | 0.96 (0.87; 1.07) | 0.458 | IVW | 0.040 | $3.6\times10^{2}$ | 137 |
| Arthritis (OR) | LDL-C & HDL-C & TG independent | 0.97 (0.61; 1.55) | 0.915 | IVW | 0.084 | $1.8\times10^{1}$ | 195 |
| ALS (OR) | Total | 1.10 (1.04; 1.17) | 0.002 | IVW | 0.368 | $9.8\times10^{2}$ | 114 |

**Table S6:** (Multivariable) Mendelian randomization effect estimates for one standard deviation change in IDL-C *(continued)*

| Outcome | Effect type | Point estimate (95%CI) | P-value | Estimator | Heterogeneity P-value | Precision | No. variants |
|---|---|---|---|---|---|---|---|
| ALS (OR) | LDL-C independent | 0.87 (0.52; 1.44) | 0.578 | IVW | 0.367 | $1.5\times10^{1}$ | 139 |
| ALS (OR) | HDL-C independent | 1.06 (1.00; 1.13) | 0.053 | IVW | 0.543 | $1.1\times10^{3}$ | 187 |
| ALS (OR) | TG independent | 1.05 (0.97; 1.13) | 0.192 | IVW | 0.079 | $6.7\times10^{2}$ | 168 |
| ALS (OR) | LDL-C & HDL-C & TG independent | 0.76 (0.53; 1.08) | 0.122 | IVW | 0.083 | $3.1\times10^{1}$ | 250 |
| MS (OR) | Total | 1.08 (0.95; 1.22) | 0.264 | IVW | 0.165 | $2.3\times10^{2}$ | 25 |
| MS (OR) | LDL-C independent | 1.73 (0.84; 3.59) | 0.138 | IVW | 0.296 | 7.3 | 24 |
| MS (OR) | HDL-C independent | 1.09 (0.97; 1.23) | 0.135 | IVW | 0.016 | $2.9\times10^{2}$ | 33 |
| MS (OR) | TG independent | 1.09 (0.95; 1.26) | 0.229 | IVW | 0.095 | $1.9\times10^{2}$ | 37 |
| MS (OR) | LDL-C & HDL-C & TG independent | 1.09 (0.52; 2.28) | 0.814 | MR Egger | 0.005 | 7.1 | 40 |
| Alzheimer (OR) | Total | 1.06 (1.04; 1.08) | < 0.001 | MR Egger | < 0.001 | $7.9\times10^{3}$ | 90 |
| Alzheimer (OR) | LDL-C independent | 0.75 (0.67; 0.83) | < 0.001 | MR Egger | < 0.001 | $3.5\times10^{2}$ | 96 |
| Alzheimer (OR) | HDL-C independent | 1.02 (1.01; 1.03) | < 0.001 | IVW | < 0.001 | $3.5\times10^{4}$ | 153 |
| Alzheimer (OR) | TG independent | 1.03 (1.02; 1.04) | < 0.001 | IVW | < 0.001 | $2.5\times10^{4}$ | 133 |
| Alzheimer (OR) | LDL-C & HDL-C & TG independent | 0.98 (0.93; 1.04) | 0.573 | MR Egger | < 0.001 | $1.2\times10^{3}$ | 208 |
| Parkinson (OR) | Total | 0.92 (0.86; 0.99) | 0.025 | IVW | 0.012 | $8.1\times10^{2}$ | 112 |
| Parkinson (OR) | LDL-C independent | 2.28 (1.34; 3.88) | 0.002 | IVW | 0.209 | $1.4\times10^{1}$ | 134 |
| Parkinson (OR) | HDL-C independent | 0.93 (0.87; 1.00) | 0.046 | IVW | 0.018 | $8.4\times10^{2}$ | 181 |
| Parkinson (OR) | TG independent | 0.92 (0.85; 1.00) | 0.056 | IVW | < 0.001 | $5.9\times10^{2}$ | 168 |
| Parkinson (OR) | LDL-C & HDL-C & TG independent | 1.81 (1.25; 2.62) | 0.002 | IVW | 0.009 | $2.8\times10^{1}$ | 239 |
| Pancreatic cancer (OR) | Total | 1.04 (0.67; 1.61) | 0.859 | MR Egger | 0.080 | $2.0\times10^{1}$ | 115 |
| Pancreatic cancer (OR) | LDL-C independent | 0.12 (0.02; 0.58) | 0.009 | MR Egger | 0.004 | 1.5 | 140 |
| Pancreatic cancer (OR) | HDL-C independent | 1.01 (0.84; 1.23) | 0.884 | IVW | < 0.001 | $1.0\times10^{2}$ | 187 |
| Pancreatic cancer (OR) | TG independent | 1.30 (1.01; 1.68) | 0.041 | IVW | 0.056 | $5.9\times10^{1}$ | 172 |
| Pancreatic cancer (OR) | LDL-C & HDL-C & TG independent | 0.92 (0.32; 2.69) | 0.886 | IVW | < 0.001 | 3.4 | 247 |
| Colon cancer (OR) | Total | 1.04 (0.85; 1.26) | 0.722 | MR Egger | 0.068 | $1.0\times10^{2}$ | 115 |
| Colon cancer (OR) | LDL-C independent | 1.51 (0.76; 3.00) | 0.235 | MR Egger | 0.024 | 8.2 | 140 |
| Colon cancer (OR) | HDL-C independent | 1.12 (1.04; 1.22) | 0.005 | IVW | 0.002 | $5.9\times10^{2}$ | 188 |
| Colon cancer (OR) | TG independent | 1.10 (0.99; 1.21) | 0.063 | IVW | 0.002 | $4.0\times10^{2}$ | 172 |
| Colon cancer (OR) | LDL-C & HDL-C & TG independent | 1.33 (0.86; 2.06) | 0.202 | IVW | < 0.001 | $2.0\times10^{1}$ | 251 |
| Rectal cancer (OR) | Total | 1.15 (1.03; 1.29) | 0.016 | IVW | 0.823 | $2.9\times10^{2}$ | 114 |
| Rectal cancer (OR) | LDL-C independent | 0.58 (0.23; 1.44) | 0.237 | MR Egger | 0.792 | 4.6 | 139 |
| Rectal cancer (OR) | HDL-C independent | 1.19 (1.07; 1.33) | 0.002 | IVW | 0.477 | $3.3\times10^{2}$ | 186 |
| Rectal cancer (OR) | TG independent | 1.10 (0.97; 1.26) | 0.136 | IVW | 0.621 | $2.3\times10^{2}$ | 174 |
| Rectal cancer (OR) | LDL-C & HDL-C & TG independent | 0.72 (0.40; 1.30) | 0.271 | IVW | 0.481 | $1.1\times10^{1}$ | 249 |
| Melanoma (OR) | Total | 0.97 (0.90; 1.04) | 0.341 | IVW | 0.155 | $8.2\times10^{2}$ | 115 |
| Melanoma (OR) | LDL-C independent | 1.43 (0.85; 2.39) | 0.177 | IVW | 0.007 | $1.4\times10^{1}$ | 140 |
| Melanoma (OR) | HDL-C independent | 0.98 (0.92; 1.04) | 0.514 | IVW | 0.004 | $1.0\times10^{3}$ | 185 |
| Melanoma (OR) | TG independent | 0.96 (0.88; 1.03) | 0.265 | IVW | 0.074 | $6.1\times10^{2}$ | 172 |
| Melanoma (OR) | LDL-C & HDL-C & TG independent | 1.35 (0.96; 1.88) | 0.082 | IVW | < 0.001 | $3.4\times10^{1}$ | 249 |

**Table S6:** (Multivariable) Mendelian randomization effect estimates for one standard deviation change in IDL-C *(continued)*

| Outcome | Effect type | Point estimate (95%CI) | P-value | Estimator | Heterogeneity P-value | Precision | No. variants |
|---|---|---|---|---|---|---|---|
| Lung cancer (OR) | Total | 0.83 (0.75; 0.92) | < 0.001 | IVW | 0.338 | $3.4\times10^2$ | 115 |
| Lung cancer (OR) | LDL-C independent | 0.80 (0.34; 1.87) | 0.608 | IVW | 0.190 | 5.3 | 141 |
| Lung cancer (OR) | HDL-C independent | 0.85 (0.76; 0.94) | 0.002 | IVW | 0.100 | $3.3\times10^2$ | 188 |
| Lung cancer (OR) | TG independent | 0.84 (0.74; 0.95) | 0.006 | IVW | 0.185 | $2.4\times10^2$ | 172 |
| Lung cancer (OR) | LDL-C & HDL-C & TG independent | 0.97 (0.54; 1.73) | 0.908 | IVW | 0.062 | $1.1\times10^1$ | 251 |

**Table S7:** (Multivariable) Mendelian randomization effect estimates for one standard deviation change in Rem-chol

| Outcome | Effect type | Point estimate (95%CI) | P-value | Estimator | Heterogeneity P-value | Precision | No. variants |
|---|---|---|---|---|---|---|---|
| CHD (OR) | Total | 1.67 (1.57; 1.79) | < 0.001 | IVW | < 0.001 | $9.0\times10^2$ | 14 |
| CHD (OR) | LDL-C independent | 1.43 (1.28; 1.58) | < 0.001 | IVW | < 0.001 | $3.5\times10^2$ | 34 |
| CHD (OR) | HDL-C independent | 1.45 (1.36; 1.54) | < 0.001 | IVW | < 0.001 | $9.3\times10^2$ | 32 |
| CHD (OR) | TG independent | 1.44 (1.28; 1.62) | < 0.001 | IVW | < 0.001 | $2.8\times10^2$ | 25 |
| CHD (OR) | LDL-C & HDL-C & TG independent | 1.04 (0.91; 1.20) | 0.534 | IVW | < 0.001 | $2.0\times10^2$ | 55 |
| HF (OR) | Total | 1.11 (1.06; 1.16) | < 0.001 | IVW | < 0.001 | $2.0\times10^3$ | 24 |
| HF (OR) | LDL-C independent | 1.12 (1.04; 1.21) | 0.002 | IVW | < 0.001 | $7.2\times10^2$ | 90 |
| HF (OR) | HDL-C independent | 1.05 (0.99; 1.11) | 0.131 | MR Egger | < 0.001 | $1.1\times10^3$ | 92 |
| HF (OR) | TG independent | 0.94 (0.87; 1.02) | 0.162 | MR Egger | < 0.001 | $5.4\times10^2$ | 71 |
| HF (OR) | LDL-C & HDL-C & TG independent | 1.09 (1.02; 1.16) | 0.014 | IVW | < 0.001 | $8.6\times10^2$ | 184 |
| AF (OR) | Total | 1.01 (0.97; 1.05) | 0.588 | IVW | 0.287 | $2.2\times10^3$ | 24 |
| AF (OR) | LDL-C independent | 1.06 (1.00; 1.13) | 0.050 | IVW | 0.004 | $1.1\times10^3$ | 95 |
| AF (OR) | HDL-C independent | 1.02 (0.98; 1.06) | 0.265 | IVW | 0.102 | $2.7\times10^3$ | 94 |
| AF (OR) | TG independent | 1.03 (0.96; 1.10) | 0.410 | MR Egger | 0.006 | $8.4\times10^2$ | 77 |
| AF (OR) | LDL-C & HDL-C & TG independent | 1.07 (1.01; 1.13) | 0.015 | IVW | < 0.001 | $1.2\times10^3$ | 191 |
| Any stroke (OR) | Total | 1.23 (1.04; 1.45) | 0.014 | MR Egger | 0.254 | $1.4\times10^2$ | 25 |
| Any stroke (OR) | LDL-C independent | 1.08 (0.99; 1.18) | 0.089 | IVW | 0.004 | $4.8\times10^2$ | 89 |
| Any stroke (OR) | HDL-C independent | 1.03 (0.98; 1.08) | 0.241 | IVW | < 0.001 | $1.6\times10^3$ | 94 |
| Any stroke (OR) | TG independent | 1.07 (0.96; 1.19) | 0.200 | MR Egger | 0.130 | $3.4\times10^2$ | 77 |
| Any stroke (OR) | LDL-C & HDL-C & TG independent | 1.00 (0.93; 1.09) | 0.952 | IVW | < 0.001 | $6.1\times10^2$ | 190 |
| Any ischemic stroke (OR) | Total | 1.19 (1.00; 1.42) | 0.044 | MR Egger | 0.379 | $1.3\times10^2$ | 25 |
| Any ischemic stroke (OR) | LDL-C independent | 1.03 (0.94; 1.14) | 0.485 | IVW | 0.014 | $4.2\times10^2$ | 88 |
| Any ischemic stroke (OR) | HDL-C independent | 1.04 (0.98; 1.09) | 0.191 | IVW | < 0.001 | $1.4\times10^3$ | 95 |
| Any ischemic stroke (OR) | TG independent | 1.07 (0.96; 1.19) | 0.243 | MR Egger | 0.712 | $3.4\times10^2$ | 76 |
| Any ischemic stroke (OR) | LDL-C & HDL-C & TG independent | 0.98 (0.90; 1.07) | 0.719 | IVW | 0.005 | $5.2\times10^2$ | 189 |
| Large artery stroke (OR) | Total | 1.20 (1.03; 1.39) | 0.020 | IVW | 0.221 | $1.7\times10^2$ | 25 |
| Large artery stroke (OR) | LDL-C independent | 1.09 (0.86; 1.38) | 0.473 | IVW | < 0.001 | $6.9\times10^1$ | 90 |
| Large artery stroke (OR) | HDL-C independent | 1.09 (0.95; 1.24) | 0.218 | IVW | 0.003 | $2.2\times10^2$ | 94 |
| Large artery stroke (OR) | TG independent | 1.35 (1.02; 1.77) | 0.035 | IVW | 0.069 | $5.0\times10^1$ | 77 |

**Table S7:** (Multivariable) Mendelian randomization effect estimates for one standard deviation change in Rem-chol *(continued)*

| Outcome | Effect type | Point estimate (95%CI) | P-value | Estimator | Heterogeneity P-value | Precision | No. variants |
|---|---|---|---|---|---|---|---|
| Large artery stroke (OR) | LDL-C & HDL-C & TG independent | 0.76 (0.61; 0.93) | 0.010 | IVW | 0.012 | $8.5\times10^{1}$ | 191 |
| Cardioembolic stroke (OR) | Total | 0.99 (0.89; 1.10) | 0.876 | IVW | 0.652 | $3.3\times10^{2}$ | 25 |
| Cardioembolic stroke (OR) | LDL-C independent | 1.09 (0.91; 1.30) | 0.335 | IVW | 0.006 | $1.2\times10^{2}$ | 91 |
| Cardioembolic stroke (OR) | HDL-C independent | 0.99 (0.90; 1.10) | 0.920 | IVW | 0.735 | $3.7\times10^{2}$ | 95 |
| Cardioembolic stroke (OR) | TG independent | 0.94 (0.78; 1.14) | 0.544 | IVW | 0.730 | $1.1\times10^{2}$ | 78 |
| Cardioembolic stroke (OR) | LDL-C & HDL-C & TG independent | 0.98 (0.83; 1.16) | 0.782 | IVW | 0.008 | $1.4\times10^{2}$ | 191 |
| Small vessel stroke (OR) | Total | 1.05 (0.92; 1.19) | 0.460 | IVW | 0.842 | $2.4\times10^{2}$ | 25 |
| Small vessel stroke (OR) | LDL-C independent | 1.24 (0.98; 1.57) | 0.069 | IVW | 0.142 | $7.0\times10^{1}$ | 92 |
| Small vessel stroke (OR) | HDL-C independent | 0.96 (0.84; 1.09) | 0.501 | IVW | 0.186 | $2.3\times10^{2}$ | 95 |
| Small vessel stroke (OR) | TG independent | 0.85 (0.67; 1.08) | 0.189 | MR Egger | 0.026 | $6.8\times10^{1}$ | 78 |
| Small vessel stroke (OR) | LDL-C & HDL-C & TG independent | 1.10 (0.90; 1.34) | 0.357 | IVW | 0.002 | $9.9\times10^{1}$ | 193 |
| cIMT (mm) | Total | 0.01 (0.00; 0.01) | 0.003 | IVW | < 0.001 | $2.5\times10^{5}$ | 23 |
| cIMT (mm) | LDL-C independent | -0.01 (-0.01; 0.00) | 0.096 | IVW | 0.007 | $1.0\times10^{5}$ | 93 |
| cIMT (mm) | HDL-C independent | 0.00 (0.00; 0.01) | 0.012 | IVW | < 0.001 | $2.7\times10^{5}$ | 93 |
| cIMT (mm) | TG independent | 0.02 (0.01; 0.02) | < 0.001 | IVW | 0.002 | $8.8\times10^{4}$ | 79 |
| cIMT (mm) | LDL-C & HDL-C & TG independent | -0.00 (-0.01; 0.01) | 0.676 | MR Egger | < 0.001 | $8.9\times10^{4}$ | 194 |
| Carotid plaque (OR) | Total | 1.34 (1.23; 1.47) | < 0.001 | IVW | 0.016 | $4.5\times10^{2}$ | 24 |
| Carotid plaque (OR) | LDL-C independent | 1.12 (0.97; 1.29) | 0.110 | IVW | 0.031 | $1.9\times10^{2}$ | 93 |
| Carotid plaque (OR) | HDL-C independent | 1.30 (1.18; 1.42) | < 0.001 | IVW | 0.010 | $4.7\times10^{2}$ | 92 |
| Carotid plaque (OR) | TG independent | 1.56 (1.32; 1.83) | < 0.001 | IVW | 0.042 | $1.5\times10^{2}$ | 75 |
| Carotid plaque (OR) | LDL-C & HDL-C & TG independent | 1.18 (1.02; 1.36) | 0.022 | IVW | 0.012 | $1.9\times10^{2}$ | 189 |
| Glucose (mmol/l) | Total | -0.00 (-0.02; 0.01) | 0.781 | IVW | 0.040 | $1.5\times10^{4}$ | 25 |
| Glucose (mmol/l) | LDL-C independent | 0.00 (-0.03; 0.03) | 0.853 | IVW | 0.060 | $4.5\times10^{3}$ | 87 |
| Glucose (mmol/l) | HDL-C independent | 0.01 (-0.00; 0.03) | 0.180 | IVW | < 0.001 | $1.7\times10^{4}$ | 89 |
| Glucose (mmol/l) | TG independent | -0.03 (-0.06; -0.00) | 0.028 | IVW | < 0.001 | $5.2\times10^{3}$ | 73 |
| Glucose (mmol/l) | LDL-C & HDL-C & TG independent | 0.00 (-0.02; 0.03) | 0.736 | IVW | < 0.001 | $6.3\times10^{3}$ | 178 |
| HbA1c (mmol/mol) | Total | 0.08 (-0.19; 0.36) | 0.545 | MR Egger | < 0.001 | $5.2\times10^{1}$ | 21 |
| HbA1c (mmol/mol) | LDL-C independent | 0.02 (-0.11; 0.16) | 0.727 | IVW | < 0.001 | $2.1\times10^{2}$ | 85 |
| HbA1c (mmol/mol) | HDL-C independent | -0.03 (-0.11; 0.05) | 0.430 | IVW | < 0.001 | $6.0\times10^{2}$ | 85 |
| HbA1c (mmol/mol) | TG independent | -0.19 (-0.34; -0.05) | 0.008 | IVW | < 0.001 | $1.9\times10^{2}$ | 69 |
| HbA1c (mmol/mol) | LDL-C & HDL-C & TG independent | -0.00 (-0.13; 0.13) | 0.987 | IVW | < 0.001 | $2.3\times10^{2}$ | 175 |
| T2DM (OR) | Total | 0.97 (0.94; 1.01) | 0.166 | IVW | 0.007 | $2.9\times10^{3}$ | 22 |
| T2DM (OR) | LDL-C independent | 1.12 (1.06; 1.18) | < 0.001 | MR Egger | < 0.001 | $1.3\times10^{3}$ | 83 |
| T2DM (OR) | HDL-C independent | 1.02 (0.99; 1.05) | 0.243 | IVW | < 0.001 | $3.5\times10^{3}$ | 84 |
| T2DM (OR) | TG independent | 0.87 (0.81; 0.93) | < 0.001 | IVW | < 0.001 | $8.5\times10^{2}$ | 57 |
| T2DM (OR) | LDL-C & HDL-C & TG independent | 1.00 (0.95; 1.06) | 0.939 | IVW | < 0.001 | $1.3\times10^{3}$ | 165 |
| CRP (mg/L) | Total | -0.47 (-0.78; -0.16) | 0.003 | MR Egger | < 0.001 | $4.0\times10^{1}$ | 14 |
| CRP (mg/L) | LDL-C independent | 0.12 (0.01; 0.23) | 0.030 | IVW | < 0.001 | $3.2\times10^{2}$ | 65 |
| CRP (mg/L) | HDL-C independent | -0.00 (-0.09; 0.08) | 0.910 | MR Egger | 0.005 | $5.5\times10^{2}$ | 82 |

**Table S7:** (Multivariable) Mendelian randomization effect estimates for one standard deviation change in Rem-chol *(continued)*

| Outcome | Effect type | Point estimate (95%CI) | P-value | Estimator | Heterogeneity P-value | Precision | No. variants |
|---|---|---|---|---|---|---|---|
| CRP (mg/L) | TG independent | -0.19 (-0.30; -0.08) | < 0.001 | IVW | < 0.001 | $3.1\times10^{2}$ | 61 |
| CRP (mg/L) | LDL-C & HDL-C & TG independent | 0.10 (0.01; 0.19) | 0.031 | IVW | < 0.001 | $4.5\times10^{2}$ | 165 |
| SBP (mmHg) | Total | 1.70 (1.06; 2.34) | < 0.001 | MR Egger | 0.106 | 9.3 | 21 |
| SBP (mmHg) | LDL-C independent | 0.81 (0.54; 1.09) | < 0.001 | IVW | < 0.001 | $5.1\times10^{1}$ | 79 |
| SBP (mmHg) | HDL-C independent | 0.52 (0.36; 0.69) | < 0.001 | IVW | < 0.001 | $1.3\times10^{2}$ | 84 |
| SBP (mmHg) | TG independent | 0.62 (0.32; 0.92) | < 0.001 | IVW | 0.006 | $4.2\times10^{1}$ | 66 |
| SBP (mmHg) | LDL-C & HDL-C & TG independent | 0.39 (0.12; 0.65) | 0.005 | MR Egger | < 0.001 | $5.3\times10^{1}$ | 171 |
| DBP (mmHg) | Total | 0.26 (0.16; 0.37) | < 0.001 | IVW | < 0.001 | $3.5\times10^{2}$ | 20 |
| DBP (mmHg) | LDL-C independent | 0.15 (-0.02; 0.31) | 0.078 | IVW | < 0.001 | $1.4\times10^{2}$ | 76 |
| DBP (mmHg) | HDL-C independent | 0.04 (-0.06; 0.13) | 0.451 | IVW | < 0.001 | $4.2\times10^{2}$ | 81 |
| DBP (mmHg) | TG independent | -0.12 (-0.31; 0.06) | 0.198 | MR Egger | < 0.001 | $1.1\times10^{2}$ | 64 |
| DBP (mmHg) | LDL-C & HDL-C & TG independent | -0.08 (-0.22; 0.07) | 0.291 | IVW | < 0.001 | $1.9\times10^{2}$ | 163 |
| PBL (OR) | Total | 1.03 (0.36; 2.95) | 0.963 | MR Egger | 0.004 | 3.4 | 17 |
| PBL (OR) | LDL-C independent | 0.87 (0.63; 1.19) | 0.367 | IVW | 0.002 | $3.9\times10^{1}$ | 55 |
| PBL (OR) | HDL-C independent | 1.01 (0.83; 1.23) | 0.908 | IVW | 0.005 | $9.9\times10^{1}$ | 58 |
| PBL (OR) | TG independent | 0.69 (0.47; 1.02) | 0.064 | IVW | 0.019 | $2.6\times10^{1}$ | 46 |
| PBL (OR) | LDL-C & HDL-C & TG independent | 0.80 (0.56; 1.15) | 0.226 | IVW | 0.003 | $3.0\times10^{1}$ | 112 |
| BUN (mg/dl) | Total | -0.00 (-0.01; 0.00) | 0.168 | IVW | < 0.001 | $1.6\times10^{5}$ | 24 |
| BUN (mg/dl) | LDL-C independent | -0.00 (-0.01; 0.01) | 0.859 | IVW | < 0.001 | $7.0\times10^{4}$ | 89 |
| BUN (mg/dl) | HDL-C independent | -0.00 (-0.01; 0.00) | 0.555 | IVW | < 0.001 | $1.7\times10^{5}$ | 89 |
| BUN (mg/dl) | TG independent | -0.01 (-0.02; -0.00) | 0.025 | IVW | < 0.001 | $5.8\times10^{4}$ | 76 |
| BUN (mg/dl) | LDL-C & HDL-C & TG independent | -0.01 (-0.01; 0.00) | 0.159 | IVW | < 0.001 | $6.9\times10^{4}$ | 185 |
| eGFR (SD of log(eGFR)) | Total | 0.00 (-0.00; 0.00) | 0.503 | IVW | < 0.001 | $9.9\times10^{5}$ | 23 |
| eGFR (SD of log(eGFR)) | LDL-C independent | 0.00 (-0.00; 0.01) | 0.103 | MR Egger | < 0.001 | $4.0\times10^{5}$ | 87 |
| eGFR (SD of log(eGFR)) | HDL-C independent | -0.00 (-0.00; 0.00) | 0.244 | IVW | < 0.001 | $1.1\times10^{6}$ | 89 |
| eGFR (SD of log(eGFR)) | TG independent | 0.00 (0.00; 0.01) | 0.011 | IVW | < 0.001 | $2.9\times10^{5}$ | 68 |
| eGFR (SD of log(eGFR)) | LDL-C & HDL-C & TG independent | 0.00 (0.00; 0.01) | 0.013 | MR Egger | < 0.001 | $3.6\times10^{5}$ | 178 |
| CKD (OR) | Total | 1.00 (0.95; 1.05) | 0.994 | IVW | 0.040 | $1.5\times10^{3}$ | 25 |
| CKD (OR) | LDL-C independent | 0.98 (0.91; 1.07) | 0.700 | IVW | 0.009 | $5.9\times10^{2}$ | 92 |
| CKD (OR) | HDL-C independent | 1.01 (0.94; 1.08) | 0.838 | MR Egger | 0.032 | $8.4\times10^{2}$ | 92 |
| CKD (OR) | TG independent | 0.92 (0.84; 1.01) | 0.087 | IVW | < 0.001 | $4.6\times10^{2}$ | 76 |
| CKD (OR) | LDL-C & HDL-C & TG independent | 1.02 (0.94; 1.11) | 0.571 | IVW | < 0.001 | $6.0\times10^{2}$ | 188 |
| IBD (OR) | Total | 0.97 (0.88; 1.06) | 0.460 | IVW | 0.009 | $4.5\times10^{2}$ | 25 |
| IBD (OR) | LDL-C independent | 0.92 (0.79; 1.06) | 0.224 | IVW | 0.012 | $1.9\times10^{2}$ | 93 |
| IBD (OR) | HDL-C independent | 0.97 (0.89; 1.05) | 0.429 | IVW | 0.004 | $5.5\times10^{2}$ | 95 |
| IBD (OR) | TG independent | 1.07 (0.91; 1.27) | 0.419 | IVW | < 0.001 | $1.4\times10^{2}$ | 77 |
| IBD (OR) | LDL-C & HDL-C & TG independent | 0.93 (0.80; 1.07) | 0.292 | IVW | < 0.001 | $1.8\times10^{2}$ | 190 |
| UC (OR) | Total | 1.04 (0.92; 1.18) | 0.520 | IVW | 0.772 | $2.5\times10^{2}$ | 24 |
| UC (OR) | LDL-C independent | 0.98 (0.80; 1.21) | 0.863 | MR Egger | 0.095 | $9.1\times10^{1}$ | 94 |

**Table S7:** (Multivariable) Mendelian randomization effect estimates for one standard deviation change in Rem-chol *(continued)*

| Outcome | Effect type | Point estimate (95%CI) | P-value | Estimator | Heterogeneity P-value | Precision | No. variants |
|---|---|---|---|---|---|---|---|
| UC (OR) | HDL-C independent | 0.99 (0.89; 1.10) | 0.813 | IVW | 0.008 | $3.5{\times}10^2$ | 96 |
| UC (OR) | TG independent | 1.04 (0.84; 1.28) | 0.748 | IVW | 0.005 | $8.7{\times}10^1$ | 78 |
| UC (OR) | LDL-C & HDL-C & TG independent | 0.88 (0.73; 1.05) | 0.151 | IVW | < 0.001 | $1.2{\times}10^2$ | 192 |
| CD (OR) | Total | 1.00 (0.88; 1.14) | 0.967 | IVW | 0.009 | $2.4{\times}10^2$ | 25 |
| CD (OR) | LDL-C independent | 0.91 (0.74; 1.12) | 0.382 | IVW | 0.384 | $9.2{\times}10^1$ | 92 |
| CD (OR) | HDL-C independent | 1.02 (0.89; 1.16) | 0.796 | IVW | 0.135 | $2.3{\times}10^2$ | 94 |
| CD (OR) | TG independent | 1.04 (0.83; 1.30) | 0.762 | IVW | 0.003 | $7.5{\times}10^1$ | 78 |
| CD (OR) | LDL-C & HDL-C & TG independent | 1.06 (0.87; 1.29) | 0.582 | IVW | 0.049 | $9.5{\times}10^1$ | 189 |
| Arthritis (OR) | Total | 1.02 (0.89; 1.17) | 0.782 | IVW | 0.081 | $2.1{\times}10^2$ | 23 |
| Arthritis (OR) | LDL-C independent | 1.10 (0.92; 1.31) | 0.302 | MR Egger | 0.013 | $1.2{\times}10^2$ | 82 |
| Arthritis (OR) | HDL-C independent | 1.05 (0.94; 1.19) | 0.368 | IVW | 0.375 | $2.8{\times}10^2$ | 80 |
| Arthritis (OR) | TG independent | 1.14 (0.94; 1.39) | 0.184 | IVW | 0.267 | $9.9{\times}10^1$ | 71 |
| Arthritis (OR) | LDL-C & HDL-C & TG independent | 1.14 (0.95; 1.37) | 0.170 | MR Egger | 0.035 | $1.1{\times}10^2$ | 167 |
| ALS (OR) | Total | 1.12 (1.02; 1.23) | 0.022 | IVW | 0.683 | $4.3{\times}10^2$ | 25 |
| ALS (OR) | LDL-C independent | 0.94 (0.81; 1.09) | 0.431 | MR Egger | 0.517 | $1.7{\times}10^2$ | 94 |
| ALS (OR) | HDL-C independent | 1.02 (0.93; 1.11) | 0.666 | IVW | 0.662 | $4.9{\times}10^2$ | 95 |
| ALS (OR) | TG independent | 1.12 (0.93; 1.35) | 0.234 | IVW | 0.104 | $1.1{\times}10^2$ | 75 |
| ALS (OR) | LDL-C & HDL-C & TG independent | 0.83 (0.72; 0.96) | 0.013 | IVW | 0.471 | $1.8{\times}10^2$ | 188 |
| MS (OR) | Total | 0.93 (0.80; 1.07) | 0.320 | IVW | 0.669 | $1.8{\times}10^2$ | 9 |
| MS (OR) | LDL-C independent | 0.66 (0.49; 0.90) | 0.008 | MR Egger | 0.309 | $4.0{\times}10^1$ | 24 |
| MS (OR) | HDL-C independent | 0.92 (0.79; 1.08) | 0.320 | IVW | 0.133 | $1.5{\times}10^2$ | 21 |
| MS (OR) | TG independent | 1.12 (0.86; 1.48) | 0.397 | IVW | 0.941 | $5.2{\times}10^1$ | 19 |
| MS (OR) | LDL-C & HDL-C & TG independent | 0.91 (0.63; 1.30) | 0.592 | IVW | 0.007 | $3.0{\times}10^1$ | 38 |
| Alzheimer (OR) | Total | 0.99 (0.89; 1.10) | 0.854 | MR Egger | 0.220 | $3.6{\times}10^2$ | 15 |
| Alzheimer (OR) | LDL-C independent | 0.98 (0.94; 1.01) | 0.205 | IVW | < 0.001 | $3.1{\times}10^3$ | 52 |
| Alzheimer (OR) | HDL-C independent | 1.00 (0.98; 1.01) | 0.865 | IVW | 0.434 | $2.0{\times}10^4$ | 83 |
| Alzheimer (OR) | TG independent | 1.02 (1.00; 1.05) | 0.105 | IVW | 0.177 | $5.6{\times}10^3$ | 69 |
| Alzheimer (OR) | LDL-C & HDL-C & TG independent | 0.99 (0.97; 1.02) | 0.650 | MR Egger | < 0.001 | $4.9{\times}10^3$ | 146 |
| Parkinson (OR) | Total | 1.37 (1.00; 1.88) | 0.051 | MR Egger | 0.480 | $3.8{\times}10^1$ | 24 |
| Parkinson (OR) | LDL-C independent | 1.12 (0.95; 1.33) | 0.183 | IVW | 0.076 | $1.3{\times}10^2$ | 92 |
| Parkinson (OR) | HDL-C independent | 1.09 (0.99; 1.20) | 0.076 | IVW | 0.548 | $4.3{\times}10^2$ | 92 |
| Parkinson (OR) | TG independent | 1.24 (1.05; 1.47) | 0.013 | IVW | 0.044 | $1.3{\times}10^2$ | 78 |
| Parkinson (OR) | LDL-C & HDL-C & TG independent | 1.23 (1.05; 1.44) | 0.009 | IVW | 0.005 | $1.6{\times}10^2$ | 190 |
| Pancreatic cancer (OR) | Total | 0.89 (0.66; 1.20) | 0.447 | IVW | 0.565 | $4.2{\times}10^1$ | 25 |
| Pancreatic cancer (OR) | LDL-C independent | 0.42 (0.26; 0.70) | < 0.001 | IVW | 0.003 | $1.5{\times}10^1$ | 91 |
| Pancreatic cancer (OR) | HDL-C independent | 0.71 (0.52; 0.96) | 0.029 | IVW | 0.058 | $4.0{\times}10^1$ | 92 |
| Pancreatic cancer (OR) | TG independent | 1.17 (0.69; 1.99) | 0.564 | IVW | 0.611 | $1.4{\times}10^1$ | 76 |
| Pancreatic cancer (OR) | LDL-C & HDL-C & TG independent | 0.54 (0.33; 0.88) | 0.014 | IVW | < 0.001 | $1.6{\times}10^1$ | 186 |
| Colon cancer (OR) | Total | 1.63 (1.06; 2.51) | 0.026 | MR Egger | 0.301 | $2.1{\times}10^1$ | 24 |

**Table S7:** (Multivariable) Mendelian randomization effect estimates for one standard deviation change in Rem-chol *(continued)*

| Outcome | Effect type | Point estimate (95%CI) | P-value | Estimator | Heterogeneity P-value | Precision | No. variants |
|---|---|---|---|---|---|---|---|
| Colon cancer (OR) | LDL-C independent | 1.02 (0.83; 1.27) | 0.829 | MR Egger | 0.010 | $8.5\times10^1$ | 90 |
| Colon cancer (OR) | HDL-C independent | 1.03 (0.90; 1.18) | 0.617 | IVW | 0.115 | $2.1\times10^2$ | 92 |
| Colon cancer (OR) | TG independent | 1.26 (0.98; 1.62) | 0.069 | IVW | 0.072 | $6.2\times10^1$ | 76 |
| Colon cancer (OR) | LDL-C & HDL-C & TG independent | 1.09 (0.88; 1.34) | 0.424 | IVW | 0.010 | $9.0\times10^1$ | 185 |
| Rectal cancer (OR) | Total | 1.18 (1.00; 1.40) | 0.055 | IVW | 0.469 | $1.3\times10^2$ | 25 |
| Rectal cancer (OR) | LDL-C independent | 1.05 (0.80; 1.38) | 0.722 | IVW | 0.908 | $5.3\times10^1$ | 91 |
| Rectal cancer (OR) | HDL-C independent | 1.11 (0.93; 1.33) | 0.257 | IVW | 0.081 | $1.2\times10^2$ | 91 |
| Rectal cancer (OR) | TG independent | 1.22 (0.91; 1.64) | 0.190 | IVW | 0.591 | $4.3\times10^1$ | 76 |
| Rectal cancer (OR) | LDL-C & HDL-C & TG independent | 1.02 (0.77; 1.34) | 0.903 | IVW | 0.375 | $5.0\times10^1$ | 186 |
| Melanoma (OR) | Total | 0.97 (0.88; 1.07) | 0.504 | IVW | 0.007 | $4.1\times10^2$ | 25 |
| Melanoma (OR) | LDL-C independent | 1.05 (0.90; 1.22) | 0.547 | IVW | 0.048 | $1.7\times10^2$ | 92 |
| Melanoma (OR) | HDL-C independent | 1.05 (0.96; 1.14) | 0.329 | IVW | 0.002 | $4.8\times10^2$ | 90 |
| Melanoma (OR) | TG independent | 0.78 (0.65; 0.92) | 0.003 | IVW | 0.040 | $1.3\times10^2$ | 75 |
| Melanoma (OR) | LDL-C & HDL-C & TG independent | 1.12 (0.96; 1.31) | 0.152 | IVW | < 0.001 | $1.6\times10^2$ | 187 |
| Lung cancer (OR) | Total | 0.88 (0.76; 1.03) | 0.118 | IVW | 0.675 | $1.6\times10^2$ | 25 |
| Lung cancer (OR) | LDL-C independent | 1.01 (0.78; 1.30) | 0.954 | IVW | 0.317 | $5.9\times10^1$ | 92 |
| Lung cancer (OR) | HDL-C independent | 0.92 (0.79; 1.06) | 0.242 | IVW | 0.016 | $1.9\times10^2$ | 93 |
| Lung cancer (OR) | TG independent | 0.67 (0.50; 0.90) | 0.008 | MR Egger | 0.939 | $4.5\times10^1$ | 76 |
| Lung cancer (OR) | LDL-C & HDL-C & TG independent | 1.11 (0.85; 1.44) | 0.449 | IVW | 0.275 | $5.5\times10^1$ | 187 |

**Table S8:** (Multivariable) Mendelian randomization effect estimates for one standard deviation change in TC

| Outcome | Effect type | Point estimate (95%CI) | P-value | Estimator | Heterogeneity P-value | Precision | No. variants |
|---|---|---|---|---|---|---|---|
| CHD (OR) | Total | 1.53 (1.43; 1.63) | < 0.001 | IVW | < 0.001 | $9.2\times10^2$ | 24 |
| CHD (OR) | LDL-C independent | 0.88 (0.62; 1.25) | 0.491 | IVW | < 0.001 | $3.2\times10^1$ | 32 |
| CHD (OR) | HDL-C independent | 1.62 (1.53; 1.72) | < 0.001 | IVW | < 0.001 | $1.1\times10^3$ | 46 |
| CHD (OR) | TG independent | 1.32 (1.20; 1.44) | < 0.001 | MR Egger | < 0.001 | $4.7\times10^2$ | 37 |
| CHD (OR) | LDL-C & HDL-C & TG independent | 1.73 (0.89; 3.37) | 0.104 | MR Egger | < 0.001 | 8.7 | 57 |
| HF (OR) | Total | 1.03 (1.00; 1.06) | 0.090 | IVW | < 0.001 | $4.0\times10^3$ | 101 |
| HF (OR) | LDL-C independent | 1.03 (0.85; 1.24) | 0.779 | IVW | < 0.001 | $1.1\times10^2$ | 126 |
| HF (OR) | HDL-C independent | 1.06 (1.02; 1.09) | < 0.001 | IVW | < 0.001 | $3.7\times10^3$ | 167 |
| HF (OR) | TG independent | 0.97 (0.92; 1.02) | 0.253 | MR Egger | < 0.001 | $1.5\times10^3$ | 147 |
| HF (OR) | LDL-C & HDL-C & TG independent | 1.31 (1.01; 1.70) | 0.042 | IVW | < 0.001 | $5.7\times10^1$ | 226 |
| AF (OR) | Total | 1.01 (0.98; 1.04) | 0.650 | IVW | 0.068 | $4.4\times10^3$ | 111 |
| AF (OR) | LDL-C independent | 1.11 (0.95; 1.30) | 0.204 | IVW | 0.005 | $1.5\times10^2$ | 138 |
| AF (OR) | HDL-C independent | 1.01 (0.98; 1.04) | 0.448 | IVW | 0.133 | $4.7\times10^3$ | 181 |
| AF (OR) | TG independent | 0.98 (0.94; 1.03) | 0.459 | MR Egger | < 0.001 | $2.1\times10^3$ | 164 |

**Table S8:** (Multivariable) Mendelian randomization effect estimates for one standard deviation change in TC *(continued)*

| Outcome | Effect type | Point estimate (95%CI) | P-value | Estimator | Heterogeneity P-value | Precision | No. variants |
|---|---|---|---|---|---|---|---|
| AF (OR) | LDL-C & HDL-C & TG independent | 1.26 (1.02; 1.57) | 0.036 | IVW | < 0.001 | $8.1\times10^{1}$ | 248 |
| Any stroke (OR) | Total | 1.06 (1.02; 1.10) | 0.004 | IVW | 0.033 | $2.7\times10^{3}$ | 103 |
| Any stroke (OR) | LDL-C independent | 0.93 (0.75; 1.15) | 0.489 | IVW | < 0.001 | $8.5\times10^{1}$ | 130 |
| Any stroke (OR) | HDL-C independent | 1.06 (1.01; 1.10) | 0.007 | IVW | < 0.001 | $2.5\times10^{3}$ | 172 |
| Any stroke (OR) | TG independent | 1.02 (0.96; 1.08) | 0.477 | MR Egger | 0.027 | $1.1\times10^{3}$ | 155 |
| Any stroke (OR) | LDL-C & HDL-C & TG independent | 1.06 (0.78; 1.45) | 0.699 | MR Egger | < 0.001 | $4.0\times10^{1}$ | 235 |
| Any ischemic stroke (OR) | Total | 1.07 (1.02; 1.11) | 0.002 | IVW | 0.009 | $2.3\times10^{3}$ | 103 |
| Any ischemic stroke (OR) | LDL-C independent | 0.98 (0.78; 1.23) | 0.860 | IVW | 0.004 | $7.2\times10^{1}$ | 128 |
| Any ischemic stroke (OR) | HDL-C independent | 1.05 (1.01; 1.10) | 0.017 | IVW | < 0.001 | $2.1\times10^{3}$ | 172 |
| Any ischemic stroke (OR) | TG independent | 1.02 (0.95; 1.09) | 0.611 | MR Egger | 0.052 | $7.7\times10^{2}$ | 155 |
| Any ischemic stroke (OR) | LDL-C & HDL-C & TG independent | 1.06 (0.76; 1.48) | 0.744 | IVW | 0.004 | $3.4\times10^{1}$ | 234 |
| Large artery stroke (OR) | Total | 0.76 (0.59; 0.97) | 0.026 | MR Egger | 0.024 | $6.3\times10^{1}$ | 102 |
| Large artery stroke (OR) | LDL-C independent | 0.70 (0.38; 1.29) | 0.256 | MR Egger | < 0.001 | $1.0\times10^{1}$ | 127 |
| Large artery stroke (OR) | HDL-C independent | 1.06 (0.96; 1.18) | 0.251 | IVW | < 0.001 | $3.5\times10^{2}$ | 174 |
| Large artery stroke (OR) | TG independent | 1.02 (0.87; 1.21) | 0.772 | MR Egger | < 0.001 | $1.4\times10^{2}$ | 154 |
| Large artery stroke (OR) | LDL-C & HDL-C & TG independent | 0.50 (0.22; 1.16) | 0.106 | MR Egger | < 0.001 | 5.4 | 236 |
| Cardioembolic stroke (OR) | Total | 0.92 (0.75; 1.13) | 0.452 | MR Egger | 0.121 | $9.3\times10^{1}$ | 106 |
| Cardioembolic stroke (OR) | LDL-C independent | 0.91 (0.57; 1.44) | 0.677 | MR Egger | 0.040 | $1.8\times10^{1}$ | 129 |
| Cardioembolic stroke (OR) | HDL-C independent | 1.02 (0.94; 1.10) | 0.705 | IVW | 0.026 | $6.1\times10^{2}$ | 176 |
| Cardioembolic stroke (OR) | TG independent | 0.99 (0.90; 1.09) | 0.857 | IVW | 0.234 | $4.1\times10^{2}$ | 159 |
| Cardioembolic stroke (OR) | LDL-C & HDL-C & TG independent | 1.78 (0.92; 3.43) | 0.085 | IVW | 0.027 | 8.9 | 236 |
| Small vessel stroke (OR) | Total | 0.97 (0.88; 1.07) | 0.584 | IVW | 0.145 | $4.0\times10^{2}$ | 106 |
| Small vessel stroke (OR) | LDL-C independent | 0.94 (0.55; 1.59) | 0.804 | IVW | 0.022 | $1.4\times10^{1}$ | 133 |
| Small vessel stroke (OR) | HDL-C independent | 0.91 (0.78; 1.06) | 0.236 | MR Egger | 0.115 | $1.7\times10^{2}$ | 174 |
| Small vessel stroke (OR) | TG independent | 0.88 (0.75; 1.03) | 0.112 | MR Egger | 0.057 | $1.5\times10^{2}$ | 156 |
| Small vessel stroke (OR) | LDL-C & HDL-C & TG independent | 0.75 (0.35; 1.63) | 0.469 | IVW | < 0.001 | 6.4 | 239 |
| cIMT (mm) | Total | 0.01 (0.00; 0.02) | < 0.001 | MR Egger | < 0.001 | $9.2\times10^{4}$ | 105 |
| cIMT (mm) | LDL-C independent | -0.04 (-0.05; -0.02) | < 0.001 | IVW | 0.002 | $1.5\times10^{4}$ | 135 |
| cIMT (mm) | HDL-C independent | 0.02 (0.01; 0.02) | < 0.001 | IVW | < 0.001 | $4.8\times10^{5}$ | 176 |
| cIMT (mm) | TG independent | 0.02 (0.01; 0.02) | < 0.001 | MR Egger | < 0.001 | $1.9\times10^{5}$ | 158 |
| cIMT (mm) | LDL-C & HDL-C & TG independent | 0.02 (-0.01; 0.04) | 0.215 | IVW | < 0.001 | $6.8\times10^{3}$ | 239 |
| Carotid plaque (OR) | Total | 1.44 (1.35; 1.55) | < 0.001 | IVW | < 0.001 | $8.1\times10^{2}$ | 99 |
| Carotid plaque (OR) | LDL-C independent | 0.69 (0.46; 1.02) | 0.066 | MR Egger | 0.008 | $2.4\times10^{1}$ | 126 |
| Carotid plaque (OR) | HDL-C independent | 1.34 (1.20; 1.50) | < 0.001 | MR Egger | 0.002 | $3.0\times10^{2}$ | 167 |
| Carotid plaque (OR) | TG independent | 1.45 (1.29; 1.64) | < 0.001 | MR Egger | < 0.001 | $2.6\times10^{2}$ | 145 |
| Carotid plaque (OR) | LDL-C & HDL-C & TG independent | 2.41 (1.35; 4.30) | 0.003 | MR Egger | 0.017 | $1.1\times10^{1}$ | 228 |
| Glucose (mmol/l) | Total | 0.02 (-0.00; 0.05) | 0.062 | MR Egger | 0.364 | $6.2\times10^{3}$ | 102 |
| Glucose (mmol/l) | LDL-C independent | 0.04 (-0.02; 0.11) | 0.210 | MR Egger | < 0.001 | $9.0\times10^{2}$ | 133 |
| Glucose (mmol/l) | HDL-C independent | -0.00 (-0.01; 0.01) | 0.518 | IVW | < 0.001 | $3.8\times10^{4}$ | 168 |

**Table S8:** (Multivariable) Mendelian randomization effect estimates for one standard deviation change in TC *(continued)*

| Outcome | Effect type | Point estimate (95%CI) | P-value | Estimator | Heterogeneity P-value | Precision | No. variants |
|---|---|---|---|---|---|---|---|
| Glucose (mmol/l) | TG independent | -0.01 (-0.03; -0.00) | 0.038 | IVW | < 0.001 | $2.3\times10^4$ | 157 |
| Glucose (mmol/l) | LDL-C & HDL-C & TG independent | 0.09 (-0.00; 0.19) | 0.061 | IVW | < 0.001 | $4.3\times10^2$ | 229 |
| HbA1c (mmol/mol) | Total | -0.11 (-0.16; -0.05) | < 0.001 | IVW | < 0.001 | $1.3\times10^3$ | 99 |
| HbA1c (mmol/mol) | LDL-C independent | -0.54 (-0.90; -0.19) | 0.003 | IVW | < 0.001 | $3.0\times10^1$ | 127 |
| HbA1c (mmol/mol) | HDL-C independent | -0.11 (-0.16; -0.06) | < 0.001 | IVW | < 0.001 | $1.4\times10^3$ | 164 |
| HbA1c (mmol/mol) | TG independent | 0.02 (-0.07; 0.10) | 0.701 | MR Egger | < 0.001 | $5.0\times10^2$ | 153 |
| HbA1c (mmol/mol) | LDL-C & HDL-C & TG independent | 0.15 (-0.34; 0.64) | 0.553 | IVW | < 0.001 | $1.6\times10^1$ | 224 |
| T2DM (OR) | Total | 0.99 (0.94; 1.05) | 0.778 | MR Egger | < 0.001 | $1.2\times10^3$ | 91 |
| T2DM (OR) | LDL-C independent | 1.10 (0.94; 1.29) | 0.219 | MR Egger | < 0.001 | $1.6\times10^2$ | 118 |
| T2DM (OR) | HDL-C independent | 0.98 (0.95; 1.00) | 0.075 | IVW | < 0.001 | $5.5\times10^3$ | 153 |
| T2DM (OR) | TG independent | 0.92 (0.89; 0.95) | < 0.001 | IVW | < 0.001 | $3.5\times10^3$ | 134 |
| T2DM (OR) | LDL-C & HDL-C & TG independent | 0.89 (0.71; 1.10) | 0.283 | IVW | < 0.001 | $7.9\times10^1$ | 209 |
| CRP (mg/L) | Total | -0.03 (-0.07; 0.01) | 0.137 | IVW | < 0.001 | $2.2\times10^3$ | 88 |
| CRP (mg/L) | LDL-C independent | 1.20 (0.94; 1.46) | < 0.001 | MR Egger | < 0.001 | $5.7\times10^1$ | 118 |
| CRP (mg/L) | HDL-C independent | -0.11 (-0.15; -0.07) | < 0.001 | IVW | < 0.001 | $2.1\times10^3$ | 148 |
| CRP (mg/L) | TG independent | -0.14 (-0.20; -0.08) | < 0.001 | MR Egger | < 0.001 | $9.2\times10^2$ | 139 |
| CRP (mg/L) | LDL-C & HDL-C & TG independent | 0.89 (0.53; 1.24) | < 0.001 | IVW | < 0.001 | $3.0\times10^1$ | 219 |
| SBP (mmHg) | Total | 0.55 (0.20; 0.90) | 0.002 | MR Egger | < 0.001 | $3.1\times10^1$ | 80 |
| SBP (mmHg) | LDL-C independent | -0.07 (-0.84; 0.69) | 0.853 | IVW | < 0.001 | 6.6 | 106 |
| SBP (mmHg) | HDL-C independent | 0.71 (0.51; 0.91) | < 0.001 | MR Egger | < 0.001 | $9.5\times10^1$ | 143 |
| SBP (mmHg) | TG independent | 0.14 (-0.02; 0.30) | 0.093 | IVW | < 0.001 | $1.5\times10^2$ | 127 |
| SBP (mmHg) | LDL-C & HDL-C & TG independent | 0.35 (-0.69; 1.40) | 0.509 | IVW | < 0.001 | 3.5 | 193 |
| DBP (mmHg) | Total | -0.00 (-0.08; 0.07) | 0.948 | IVW | < 0.001 | $7.0\times10^2$ | 81 |
| DBP (mmHg) | LDL-C independent | 0.22 (-0.24; 0.67) | 0.354 | IVW | < 0.001 | $1.8\times10^1$ | 103 |
| DBP (mmHg) | HDL-C independent | 0.02 (-0.06; 0.10) | 0.652 | IVW | < 0.001 | $6.4\times10^2$ | 138 |
| DBP (mmHg) | TG independent | -0.19 (-0.28; -0.10) | < 0.001 | IVW | < 0.001 | $4.4\times10^2$ | 124 |
| DBP (mmHg) | LDL-C & HDL-C & TG independent | -0.73 (-1.36; -0.11) | 0.021 | IVW | < 0.001 | 9.9 | 191 |
| PBL (OR) | Total | 1.09 (0.87; 1.37) | 0.458 | IVW | 0.053 | $7.5\times10^1$ | 51 |
| PBL (OR) | LDL-C independent | 1.15 (0.59; 2.22) | 0.686 | IVW | 0.016 | 8.8 | 64 |
| PBL (OR) | HDL-C independent | 1.16 (0.94; 1.43) | 0.169 | IVW | 0.202 | $8.7\times10^1$ | 83 |
| PBL (OR) | TG independent | 1.64 (1.07; 2.53) | 0.023 | MR Egger | 0.114 | $2.1\times10^1$ | 74 |
| PBL (OR) | LDL-C & HDL-C & TG independent | 0.41 (0.11; 1.62) | 0.205 | IVW | 0.019 | 2.1 | 115 |
| BUN (mg/dl) | Total | -0.01 (-0.02; 0.00) | 0.055 | MR Egger | 0.007 | $5.1\times10^4$ | 102 |
| BUN (mg/dl) | LDL-C independent | 0.02 (-0.00; 0.04) | 0.119 | IVW | < 0.001 | $9.1\times10^3$ | 127 |
| BUN (mg/dl) | HDL-C independent | 0.00 (0.00; 0.01) | 0.014 | IVW | < 0.001 | $2.7\times10^5$ | 165 |
| BUN (mg/dl) | TG independent | 0.00 (-0.00; 0.01) | 0.293 | MR Egger | < 0.001 | $1.1\times10^5$ | 153 |
| BUN (mg/dl) | LDL-C & HDL-C & TG independent | 0.03 (-0.00; 0.06) | 0.077 | MR Egger | < 0.001 | $4.3\times10^3$ | 229 |
| eGFR (SD of log(eGFR)) | Total | 0.00 (-0.00; 0.01) | 0.186 | MR Egger | < 0.001 | $3.7\times10^5$ | 95 |
| eGFR (SD of log(eGFR)) | LDL-C independent | 0.00 (-0.01; 0.01) | 0.704 | IVW | < 0.001 | $5.4\times10^4$ | 124 |

**Table S8:** (Multivariable) Mendelian randomization effect estimates for one standard deviation change in TC *(continued)*

| Outcome | Effect type | Point estimate (95%CI) | P-value | Estimator | Heterogeneity P-value | Precision | No. variants |
|---|---|---|---|---|---|---|---|
| eGFR (SD of log(eGFR)) | HDL-C independent | -0.00 (-0.00; 0.00) | 0.800 | IVW | < 0.001 | $1.9\times10^6$ | 162 |
| eGFR (SD of log(eGFR)) | TG independent | -0.00 (-0.00; 0.00) | 0.694 | IVW | < 0.001 | $1.4\times10^6$ | 146 |
| eGFR (SD of log(eGFR)) | LDL-C & HDL-C & TG independent | -0.01 (-0.02; 0.01) | 0.263 | IVW | < 0.001 | $2.8\times10^4$ | 222 |
| CKD (OR) | Total | 0.96 (0.92; 1.00) | 0.030 | IVW | < 0.001 | $2.7\times10^3$ | 104 |
| CKD (OR) | LDL-C independent | 0.77 (0.61; 0.95) | 0.017 | MR Egger | 0.011 | $8.0\times10^1$ | 131 |
| CKD (OR) | HDL-C independent | 0.99 (0.95; 1.03) | 0.495 | IVW | 0.005 | $2.7\times10^3$ | 172 |
| CKD (OR) | TG independent | 0.99 (0.94; 1.06) | 0.850 | MR Egger | < 0.001 | $1.1\times10^3$ | 157 |
| CKD (OR) | LDL-C & HDL-C & TG independent | 0.93 (0.68; 1.26) | 0.624 | IVW | < 0.001 | $4.0\times10^1$ | 236 |
| IBD (OR) | Total | 1.09 (1.02; 1.16) | 0.010 | IVW | < 0.001 | $8.8\times10^2$ | 107 |
| IBD (OR) | LDL-C independent | 0.38 (0.26; 0.57) | < 0.001 | IVW | 0.011 | $2.5\times10^1$ | 134 |
| IBD (OR) | HDL-C independent | 1.22 (1.10; 1.35) | < 0.001 | MR Egger | < 0.001 | $3.6\times10^2$ | 175 |
| IBD (OR) | TG independent | 1.22 (1.10; 1.35) | < 0.001 | MR Egger | < 0.001 | $3.5\times10^2$ | 158 |
| IBD (OR) | LDL-C & HDL-C & TG independent | 0.35 (0.20; 0.60) | < 0.001 | MR Egger | < 0.001 | $1.2\times10^1$ | 241 |
| UC (OR) | Total | 1.22 (1.01; 1.48) | 0.036 | MR Egger | 0.010 | $1.1\times10^2$ | 109 |
| UC (OR) | LDL-C independent | 0.39 (0.23; 0.65) | < 0.001 | IVW | 0.303 | $1.5\times10^1$ | 133 |
| UC (OR) | HDL-C independent | 1.06 (0.98; 1.16) | 0.150 | IVW | < 0.001 | $5.3\times10^2$ | 178 |
| UC (OR) | TG independent | 1.09 (0.99; 1.20) | 0.077 | IVW | 0.005 | $4.1\times10^2$ | 163 |
| UC (OR) | LDL-C & HDL-C & TG independent | 0.27 (0.13; 0.55) | < 0.001 | IVW | 0.005 | 7.8 | 240 |
| CD (OR) | Total | 1.08 (0.99; 1.18) | 0.089 | IVW | 0.019 | $4.7\times10^2$ | 107 |
| CD (OR) | LDL-C independent | 0.69 (0.39; 1.21) | 0.192 | IVW | 0.060 | $1.2\times10^1$ | 135 |
| CD (OR) | HDL-C independent | 1.14 (1.04; 1.25) | 0.006 | IVW | 0.024 | $4.4\times10^2$ | 176 |
| CD (OR) | TG independent | 1.12 (0.97; 1.30) | 0.119 | MR Egger | 0.003 | $1.8\times10^2$ | 160 |
| CD (OR) | LDL-C & HDL-C & TG independent | 0.38 (0.17; 0.87) | 0.022 | IVW | 0.077 | 5.6 | 238 |
| Arthritis (OR) | Total | 1.05 (0.80; 1.38) | 0.740 | MR Egger | 0.070 | $5.1\times10^1$ | 85 |
| Arthritis (OR) | LDL-C independent | 0.90 (0.51; 1.60) | 0.719 | IVW | 0.061 | $1.2\times10^1$ | 108 |
| Arthritis (OR) | HDL-C independent | 0.90 (0.81; 0.99) | 0.031 | IVW | 0.069 | $3.9\times10^2$ | 143 |
| Arthritis (OR) | TG independent | 1.12 (0.96; 1.30) | 0.156 | MR Egger | 0.044 | $1.6\times10^2$ | 132 |
| Arthritis (OR) | LDL-C & HDL-C & TG independent | 0.61 (0.28; 1.35) | 0.226 | IVW | 0.041 | 6.2 | 191 |
| ALS (OR) | Total | 1.12 (1.05; 1.20) | < 0.001 | IVW | 0.228 | $9.3\times10^2$ | 107 |
| ALS (OR) | LDL-C independent | 0.98 (0.66; 1.46) | 0.926 | MR Egger | 0.282 | $2.4\times10^1$ | 136 |
| ALS (OR) | HDL-C independent | 1.11 (1.05; 1.18) | < 0.001 | IVW | 0.336 | $1.0\times10^3$ | 178 |
| ALS (OR) | TG independent | 1.07 (0.99; 1.17) | 0.087 | IVW | 0.074 | $5.7\times10^2$ | 159 |
| ALS (OR) | LDL-C & HDL-C & TG independent | 0.94 (0.53; 1.68) | 0.839 | IVW | 0.178 | $1.1\times10^1$ | 239 |
| MS (OR) | Total | 1.01 (0.89; 1.15) | 0.831 | IVW | 0.901 | $2.3\times10^2$ | 21 |
| MS (OR) | LDL-C independent | 0.64 (0.28; 1.46) | 0.288 | MR Egger | 0.250 | 5.6 | 23 |
| MS (OR) | HDL-C independent | 1.00 (0.88; 1.14) | 0.974 | IVW | 0.038 | $2.3\times10^2$ | 30 |
| MS (OR) | TG independent | 1.04 (0.90; 1.21) | 0.576 | IVW | 0.980 | $1.7\times10^2$ | 31 |
| MS (OR) | LDL-C & HDL-C & TG independent | 2.08 (0.65; 6.71) | 0.218 | MR Egger | 0.005 | 2.8 | 41 |
| Alzheimer (OR) | Total | 1.00 (0.98; 1.03) | 0.772 | MR Egger | 0.009 | $7.4\times10^3$ | 88 |

**Table S8:** (Multivariable) Mendelian randomization effect estimates for one standard deviation change in TC *(continued)*

| Outcome | Effect type | Point estimate (95%CI) | P-value | Estimator | Heterogeneity P-value | Precision | No. variants |
|---|---|---|---|---|---|---|---|
| Alzheimer (OR) | LDL-C independent | 0.86 (0.81; 0.91) | < 0.001 | IVW | < 0.001 | $1.1\times10^3$ | 101 |
| Alzheimer (OR) | HDL-C independent | 1.02 (1.00; 1.03) | 0.024 | MR Egger | < 0.001 | $1.5\times10^4$ | 144 |
| Alzheimer (OR) | TG independent | 1.02 (1.01; 1.04) | 0.003 | IVW | 0.003 | $2.0\times10^4$ | 129 |
| Alzheimer (OR) | LDL-C & HDL-C & TG independent | 0.96 (0.88; 1.05) | 0.352 | IVW | < 0.001 | $5.4\times10^2$ | 199 |
| Parkinson (OR) | Total | 0.97 (0.80; 1.19) | 0.787 | MR Egger | 0.081 | $9.6\times10^1$ | 103 |
| Parkinson (OR) | LDL-C independent | 1.45 (0.94; 2.24) | 0.093 | IVW | 0.084 | $2.0\times10^1$ | 132 |
| Parkinson (OR) | HDL-C independent | 0.95 (0.84; 1.07) | 0.404 | MR Egger | 0.034 | $2.7\times10^2$ | 168 |
| Parkinson (OR) | TG independent | 0.98 (0.90; 1.08) | 0.705 | IVW | < 0.001 | $4.5\times10^2$ | 158 |
| Parkinson (OR) | LDL-C & HDL-C & TG independent | 3.13 (1.64; 5.99) | < 0.001 | IVW | 0.004 | 9.1 | 233 |
| Pancreatic cancer (OR) | Total | 1.29 (1.04; 1.62) | 0.023 | IVW | 0.104 | $7.8\times10^1$ | 109 |
| Pancreatic cancer (OR) | LDL-C independent | 0.96 (0.27; 3.36) | 0.948 | IVW | < 0.001 | 2.4 | 137 |
| Pancreatic cancer (OR) | HDL-C independent | 1.18 (0.96; 1.45) | 0.119 | IVW | 0.002 | $8.9\times10^1$ | 176 |
| Pancreatic cancer (OR) | TG independent | 1.47 (1.12; 1.94) | 0.005 | IVW | 0.062 | $5.1\times10^1$ | 165 |
| Pancreatic cancer (OR) | LDL-C & HDL-C & TG independent | 0.63 (0.10; 4.07) | 0.629 | IVW | < 0.001 | 1.1 | 238 |
| Colon cancer (OR) | Total | 1.11 (0.92; 1.34) | 0.283 | MR Egger | 0.366 | $1.1\times10^2$ | 109 |
| Colon cancer (OR) | LDL-C independent | 2.04 (1.16; 3.57) | 0.013 | IVW | 0.010 | $1.2\times10^1$ | 136 |
| Colon cancer (OR) | HDL-C independent | 1.13 (1.03; 1.23) | 0.008 | IVW | 0.021 | $5.0\times10^2$ | 176 |
| Colon cancer (OR) | TG independent | 1.10 (0.99; 1.23) | 0.083 | IVW | 0.024 | $3.3\times10^2$ | 165 |
| Colon cancer (OR) | LDL-C & HDL-C & TG independent | 1.65 (0.76; 3.59) | 0.204 | IVW | < 0.001 | 6.4 | 240 |
| Rectal cancer (OR) | Total | 1.14 (0.89; 1.45) | 0.300 | MR Egger | 0.807 | $6.6\times10^1$ | 110 |
| Rectal cancer (OR) | LDL-C independent | 0.32 (0.16; 0.65) | 0.002 | IVW | 0.648 | 7.7 | 137 |
| Rectal cancer (OR) | HDL-C independent | 1.18 (0.99; 1.41) | 0.066 | MR Egger | 0.692 | $1.2\times10^2$ | 177 |
| Rectal cancer (OR) | TG independent | 1.04 (0.89; 1.20) | 0.644 | IVW | 0.476 | $1.8\times10^2$ | 165 |
| Rectal cancer (OR) | LDL-C & HDL-C & TG independent | 0.25 (0.09; 0.72) | 0.010 | IVW | 0.689 | 3.4 | 236 |
| Melanoma (OR) | Total | 0.95 (0.82; 1.09) | 0.452 | MR Egger | 0.037 | $1.9\times10^2$ | 107 |
| Melanoma (OR) | LDL-C independent | 1.37 (0.91; 2.04) | 0.129 | MR Egger | 0.206 | $2.4\times10^1$ | 139 |
| Melanoma (OR) | HDL-C independent | 0.98 (0.92; 1.05) | 0.633 | IVW | 0.003 | $8.6\times10^2$ | 173 |
| Melanoma (OR) | TG independent | 0.91 (0.84; 0.98) | 0.016 | IVW | < 0.001 | $5.9\times10^2$ | 164 |
| Melanoma (OR) | LDL-C & HDL-C & TG independent | 1.26 (0.70; 2.25) | 0.439 | IVW | < 0.001 | $1.1\times10^1$ | 238 |
| Lung cancer (OR) | Total | 0.84 (0.65; 1.07) | 0.158 | MR Egger | 0.190 | $6.1\times10^1$ | 108 |
| Lung cancer (OR) | LDL-C independent | 1.97 (1.02; 3.80) | 0.044 | MR Egger | 0.073 | 8.8 | 140 |
| Lung cancer (OR) | HDL-C independent | 0.81 (0.69; 0.95) | 0.012 | MR Egger | 0.169 | $1.4\times10^2$ | 180 |
| Lung cancer (OR) | TG independent | 0.82 (0.69; 0.98) | 0.033 | MR Egger | 0.104 | $1.2\times10^2$ | 165 |
| Lung cancer (OR) | LDL-C & HDL-C & TG independent | 0.93 (0.35; 2.43) | 0.878 | IVW | 0.018 | 4.2 | 241 |

**Table S9:** (Multivariable) Mendelian randomization effect estimates for one standard deviation change in Apo-B

| Outcome | Effect type | Point estimate (95%CI) | P-value | Estimator | Heterogeneity P-value | Precision | No. variants |
|---|---|---|---|---|---|---|---|
| CHD (OR) | Total | 1.45 (1.24; 1.68) | < 0.001 | MR Egger | < 0.001 | $1.7{\times}10^2$ | 29 |
| CHD (OR) | LDL-C independent | 1.87 (1.55; 2.25) | < 0.001 | IVW | < 0.001 | $1.1{\times}10^2$ | 36 |
| CHD (OR) | HDL-C independent | 1.71 (1.56; 1.88) | < 0.001 | MR Egger | < 0.001 | $4.3{\times}10^2$ | 49 |
| CHD (OR) | TG independent | 1.56 (1.39; 1.74) | < 0.001 | MR Egger | < 0.001 | $3.1{\times}10^2$ | 39 |
| CHD (OR) | LDL-C & HDL-C & TG independent | 3.80 (1.96; 7.38) | < 0.001 | IVW | < 0.001 | 8.7 | 59 |
| HF (OR) | Total | 1.04 (1.00; 1.07) | 0.036 | IVW | 0.050 | $3.2{\times}10^3$ | 92 |
| HF (OR) | LDL-C independent | 1.08 (0.95; 1.22) | 0.241 | IVW | < 0.001 | $2.4{\times}10^2$ | 132 |
| HF (OR) | HDL-C independent | 1.06 (1.03; 1.10) | < 0.001 | IVW | 0.002 | $3.2{\times}10^3$ | 162 |
| HF (OR) | TG independent | 0.98 (0.92; 1.04) | 0.472 | IVW | 0.003 | $1.0{\times}10^3$ | 127 |
| HF (OR) | LDL-C & HDL-C & TG independent | 1.89 (1.46; 2.43) | < 0.001 | IVW | < 0.001 | $5.9{\times}10^1$ | 230 |
| AF (OR) | Total | 1.02 (0.99; 1.05) | 0.288 | IVW | 0.148 | $4.0{\times}10^3$ | 99 |
| AF (OR) | LDL-C independent | 0.97 (0.87; 1.08) | 0.559 | IVW | 0.022 | $3.5{\times}10^2$ | 144 |
| AF (OR) | HDL-C independent | 1.02 (0.99; 1.06) | 0.137 | IVW | 0.088 | $4.1{\times}10^3$ | 175 |
| AF (OR) | TG independent | 1.03 (0.98; 1.08) | 0.304 | IVW | 0.003 | $1.4{\times}10^3$ | 140 |
| AF (OR) | LDL-C & HDL-C & TG independent | 1.00 (0.81; 1.25) | 0.978 | IVW | < 0.001 | $8.0{\times}10^1$ | 251 |
| Any stroke (OR) | Total | 1.07 (1.02; 1.12) | 0.003 | IVW | 0.127 | $2.0{\times}10^3$ | 93 |
| Any stroke (OR) | LDL-C independent | 1.02 (0.88; 1.18) | 0.799 | IVW | < 0.001 | $1.8{\times}10^2$ | 132 |
| Any stroke (OR) | HDL-C independent | 1.05 (1.01; 1.09) | 0.011 | IVW | 0.002 | $2.5{\times}10^3$ | 167 |
| Any stroke (OR) | TG independent | 1.07 (1.00; 1.16) | 0.052 | IVW | 0.017 | $7.3{\times}10^2$ | 133 |
| Any stroke (OR) | LDL-C & HDL-C & TG independent | 1.28 (0.94; 1.75) | 0.121 | IVW | < 0.001 | $4.0{\times}10^1$ | 239 |
| Any ischemic stroke (OR) | Total | 1.09 (1.04; 1.15) | < 0.001 | IVW | 0.157 | $1.7{\times}10^3$ | 92 |
| Any ischemic stroke (OR) | LDL-C independent | 1.00 (0.85; 1.17) | 0.972 | IVW | 0.002 | $1.5{\times}10^2$ | 131 |
| Any ischemic stroke (OR) | HDL-C independent | 1.06 (1.02; 1.11) | 0.004 | IVW | < 0.001 | $2.2{\times}10^3$ | 169 |
| Any ischemic stroke (OR) | TG independent | 1.05 (0.95; 1.15) | 0.347 | MR Egger | 0.090 | $4.2{\times}10^2$ | 135 |
| Any ischemic stroke (OR) | LDL-C & HDL-C & TG independent | 1.27 (0.90; 1.78) | 0.171 | IVW | < 0.001 | $3.3{\times}10^1$ | 237 |
| Large artery stroke (OR) | Total | 1.05 (0.94; 1.17) | 0.371 | IVW | 0.005 | $3.2{\times}10^2$ | 95 |
| Large artery stroke (OR) | LDL-C independent | 0.74 (0.51; 1.07) | 0.114 | MR Egger | < 0.001 | $2.8{\times}10^1$ | 138 |
| Large artery stroke (OR) | HDL-C independent | 0.92 (0.78; 1.07) | 0.272 | MR Egger | 0.009 | $1.5{\times}10^2$ | 170 |
| Large artery stroke (OR) | TG independent | 1.05 (0.86; 1.28) | 0.638 | IVW | 0.002 | $9.9{\times}10^1$ | 135 |
| Large artery stroke (OR) | LDL-C & HDL-C & TG independent | 1.62 (0.69; 3.79) | 0.269 | IVW | < 0.001 | 5.3 | 240 |
| Cardioembolic stroke (OR) | Total | 1.04 (0.96; 1.13) | 0.367 | IVW | 0.004 | $5.4{\times}10^2$ | 95 |
| Cardioembolic stroke (OR) | LDL-C independent | 1.09 (0.80; 1.47) | 0.601 | IVW | < 0.001 | $4.1{\times}10^1$ | 134 |
| Cardioembolic stroke (OR) | HDL-C independent | 1.01 (0.93; 1.10) | 0.772 | IVW | 0.006 | $5.8{\times}10^2$ | 171 |
| Cardioembolic stroke (OR) | TG independent | 1.07 (0.92; 1.25) | 0.366 | IVW | 0.009 | $1.6{\times}10^2$ | 134 |
| Cardioembolic stroke (OR) | LDL-C & HDL-C & TG independent | 2.61 (1.33; 5.11) | 0.005 | IVW | 0.008 | 8.5 | 237 |
| Small vessel stroke (OR) | Total | 0.84 (0.64; 1.10) | 0.201 | MR Egger | 0.108 | $5.3{\times}10^1$ | 96 |
| Small vessel stroke (OR) | LDL-C independent | 0.98 (0.69; 1.38) | 0.894 | IVW | 0.022 | $3.3{\times}10^1$ | 137 |
| Small vessel stroke (OR) | HDL-C independent | 0.99 (0.86; 1.14) | 0.923 | MR Egger | 0.044 | $2.0{\times}10^2$ | 173 |
| Small vessel stroke (OR) | TG independent | 0.73 (0.59; 0.90) | 0.003 | MR Egger | 0.014 | $9.1{\times}10^1$ | 137 |

**Table S9:** (Multivariable) Mendelian randomization effect estimates for one standard deviation change in Apo-B *(continued)*

| Outcome | Effect type | Point estimate (95%CI) | P-value | Estimator | Heterogeneity P-value | Precision | No. variants |
|---|---|---|---|---|---|---|---|
| Small vessel stroke (OR) | LDL-C & HDL-C & TG independent | 0.84 (0.39; 1.84) | 0.672 | IVW | 0.003 | 6.3 | 241 |
| cIMT (mm) | Total | 0.02 (0.01; 0.02) | < 0.001 | IVW | < 0.001 | $4.2\times10^5$ | 95 |
| cIMT (mm) | LDL-C independent | -0.01 (-0.02; -0.00) | 0.033 | IVW | < 0.001 | $3.4\times10^4$ | 138 |
| cIMT (mm) | HDL-C independent | 0.01 (0.01; 0.02) | < 0.001 | IVW | < 0.001 | $4.3\times10^5$ | 168 |
| cIMT (mm) | TG independent | 0.03 (0.02; 0.03) | < 0.001 | IVW | 0.008 | $1.4\times10^5$ | 138 |
| cIMT (mm) | LDL-C & HDL-C & TG independent | 0.04 (0.02; 0.06) | < 0.001 | IVW | < 0.001 | $6.9\times10^3$ | 241 |
| Carotid plaque (OR) | Total | 1.35 (1.25; 1.46) | < 0.001 | IVW | 0.025 | $6.7\times10^2$ | 89 |
| Carotid plaque (OR) | LDL-C independent | 0.99 (0.74; 1.32) | 0.922 | MR Egger | 0.050 | $4.5\times10^1$ | 133 |
| Carotid plaque (OR) | HDL-C independent | 1.35 (1.25; 1.45) | < 0.001 | IVW | 0.003 | $7.3\times10^2$ | 163 |
| Carotid plaque (OR) | TG independent | 1.40 (1.22; 1.60) | < 0.001 | IVW | 0.029 | $2.0\times10^2$ | 125 |
| Carotid plaque (OR) | LDL-C & HDL-C & TG independent | 2.76 (1.54; 4.93) | < 0.001 | IVW | 0.006 | $1.1\times10^1$ | 231 |
| Glucose (mmol/l) | Total | 0.02 (-0.01; 0.06) | 0.157 | MR Egger | 0.002 | $3.2\times10^3$ | 90 |
| Glucose (mmol/l) | LDL-C independent | -0.01 (-0.05; 0.04) | 0.765 | IVW | 0.008 | $2.0\times10^3$ | 134 |
| Glucose (mmol/l) | HDL-C independent | -0.00 (-0.02; 0.01) | 0.517 | IVW | < 0.001 | $2.6\times10^4$ | 159 |
| Glucose (mmol/l) | TG independent | -0.02 (-0.04; 0.01) | 0.163 | IVW | < 0.001 | $8.3\times10^3$ | 135 |
| Glucose (mmol/l) | LDL-C & HDL-C & TG independent | -0.06 (-0.15; 0.03) | 0.215 | MR Egger | < 0.001 | $4.3\times10^2$ | 236 |
| HbA1c (mmol/mol) | Total | -0.13 (-0.19; -0.07) | < 0.001 | IVW | < 0.001 | $9.4\times10^2$ | 86 |
| HbA1c (mmol/mol) | LDL-C independent | -0.28 (-0.50; -0.07) | 0.011 | IVW | < 0.001 | $8.1\times10^1$ | 135 |
| HbA1c (mmol/mol) | HDL-C independent | -0.13 (-0.20; -0.07) | < 0.001 | IVW | < 0.001 | $9.2\times10^2$ | 151 |
| HbA1c (mmol/mol) | TG independent | -0.10 (-0.21; 0.01) | 0.078 | IVW | < 0.001 | $3.1\times10^2$ | 129 |
| HbA1c (mmol/mol) | LDL-C & HDL-C & TG independent | -0.40 (-0.88; 0.09) | 0.107 | MR Egger | < 0.001 | $1.6\times10^1$ | 232 |
| T2DM (OR) | Total | 1.00 (0.93; 1.07) | 0.987 | MR Egger | < 0.001 | $7.8\times10^2$ | 82 |
| T2DM (OR) | LDL-C independent | 1.05 (0.94; 1.16) | 0.404 | MR Egger | < 0.001 | $3.4\times10^2$ | 122 |
| T2DM (OR) | HDL-C independent | 0.94 (0.92; 0.97) | < 0.001 | IVW | < 0.001 | $5.0\times10^3$ | 144 |
| T2DM (OR) | TG independent | 0.89 (0.85; 0.94) | < 0.001 | IVW | < 0.001 | $1.4\times10^3$ | 111 |
| T2DM (OR) | LDL-C & HDL-C & TG independent | 0.87 (0.72; 1.06) | 0.176 | IVW | < 0.001 | $9.8\times10^1$ | 212 |
| CRP (mg/L) | Total | 0.02 (-0.02; 0.07) | 0.289 | IVW | < 0.001 | $1.9\times10^3$ | 83 |
| CRP (mg/L) | LDL-C independent | 0.59 (0.41; 0.76) | < 0.001 | IVW | < 0.001 | $1.2\times10^2$ | 116 |
| CRP (mg/L) | HDL-C independent | 0.06 (0.01; 0.11) | 0.014 | IVW | < 0.001 | $1.8\times10^3$ | 144 |
| CRP (mg/L) | TG independent | -0.19 (-0.29; -0.10) | < 0.001 | MR Egger | < 0.001 | $4.0\times10^2$ | 121 |
| CRP (mg/L) | LDL-C & HDL-C & TG independent | 0.70 (0.36; 1.04) | < 0.001 | IVW | < 0.001 | $3.4\times10^1$ | 223 |
| SBP (mmHg) | Total | 0.45 (0.10; 0.81) | 0.012 | MR Egger | < 0.001 | $3.0\times10^1$ | 78 |
| SBP (mmHg) | LDL-C independent | -0.36 (-0.89; 0.17) | 0.180 | IVW | < 0.001 | $1.4\times10^1$ | 107 |
| SBP (mmHg) | HDL-C independent | 0.39 (0.18; 0.61) | < 0.001 | MR Egger | < 0.001 | $8.0\times10^1$ | 138 |
| SBP (mmHg) | TG independent | 0.14 (-0.10; 0.38) | 0.256 | IVW | < 0.001 | $6.5\times10^1$ | 113 |
| SBP (mmHg) | LDL-C & HDL-C & TG independent | 0.12 (-0.88; 1.11) | 0.815 | IVW | < 0.001 | 3.9 | 197 |
| DBP (mmHg) | Total | 0.03 (-0.06; 0.11) | 0.526 | IVW | < 0.001 | $5.7\times10^2$ | 76 |
| DBP (mmHg) | LDL-C independent | 0.09 (-0.18; 0.37) | 0.506 | IVW | < 0.001 | $4.9\times10^1$ | 111 |
| DBP (mmHg) | HDL-C independent | -0.03 (-0.11; 0.06) | 0.531 | IVW | < 0.001 | $5.8\times10^2$ | 135 |

**Table S9:** (Multivariable) Mendelian randomization effect estimates for one standard deviation change in Apo-B *(continued)*

| Outcome | Effect type | Point estimate (95%CI) | P-value | Estimator | Heterogeneity P-value | Precision | No. variants |
|---|---|---|---|---|---|---|---|
| DBP (mmHg) | TG independent | -0.22 (-0.37; -0.07) | 0.004 | IVW | < 0.001 | $1.7\times10^{2}$ | 109 |
| DBP (mmHg) | LDL-C & HDL-C & TG independent | -0.37 (-0.97; 0.24) | 0.232 | IVW | < 0.001 | $1.1\times10^{1}$ | 198 |
| PBL (OR) | Total | 1.12 (0.92; 1.35) | 0.262 | IVW | 0.002 | $1.1\times10^{2}$ | 48 |
| PBL (OR) | LDL-C independent | 1.36 (0.85; 2.18) | 0.197 | IVW | 0.015 | $1.7\times10^{1}$ | 66 |
| PBL (OR) | HDL-C independent | 1.08 (0.89; 1.32) | 0.411 | IVW | 0.014 | $1.0\times10^{2}$ | 85 |
| PBL (OR) | TG independent | 1.00 (0.72; 1.39) | 0.993 | IVW | 0.002 | $3.5\times10^{1}$ | 69 |
| PBL (OR) | LDL-C & HDL-C & TG independent | 1.03 (0.19; 5.45) | 0.975 | IVW | 0.007 | 1.4 | 114 |
| BUN (mg/dl) | Total | -0.01 (-0.02; 0.00) | 0.266 | MR Egger | < 0.001 | $3.3\times10^{4}$ | 90 |
| BUN (mg/dl) | LDL-C independent | -0.01 (-0.02; 0.00) | 0.149 | IVW | < 0.001 | $1.9\times10^{4}$ | 130 |
| BUN (mg/dl) | HDL-C independent | -0.00 (-0.01; 0.01) | 0.941 | MR Egger | < 0.001 | $9.1\times10^{4}$ | 155 |
| BUN (mg/dl) | TG independent | 0.00 (-0.00; 0.01) | 0.225 | IVW | < 0.001 | $8.1\times10^{4}$ | 133 |
| BUN (mg/dl) | LDL-C & HDL-C & TG independent | 0.00 (-0.02; 0.03) | 0.752 | IVW | < 0.001 | $4.5\times10^{3}$ | 230 |
| eGFR (SD of log(eGFR)) | Total | 0.00 (-0.00; 0.00) | 0.359 | IVW | < 0.001 | $1.7\times10^{6}$ | 88 |
| eGFR (SD of log(eGFR)) | LDL-C independent | 0.01 (0.00; 0.01) | 0.003 | IVW | < 0.001 | $1.2\times10^{5}$ | 127 |
| eGFR (SD of log(eGFR)) | HDL-C independent | 0.00 (-0.00; 0.00) | 0.580 | IVW | < 0.001 | $1.6\times10^{6}$ | 155 |
| eGFR (SD of log(eGFR)) | TG independent | 0.00 (-0.00; 0.00) | 0.830 | IVW | < 0.001 | $5.4\times10^{5}$ | 125 |
| eGFR (SD of log(eGFR)) | LDL-C & HDL-C & TG independent | 0.00 (-0.01; 0.01) | 0.603 | MR Egger | < 0.001 | $2.9\times10^{4}$ | 225 |
| CKD (OR) | Total | 0.98 (0.94; 1.02) | 0.419 | IVW | 0.018 | $2.3\times10^{3}$ | 91 |
| CKD (OR) | LDL-C independent | 0.94 (0.81; 1.07) | 0.344 | IVW | 0.013 | $2.0\times10^{2}$ | 135 |
| CKD (OR) | HDL-C independent | 0.98 (0.95; 1.02) | 0.439 | IVW | 0.006 | $2.4\times10^{3}$ | 162 |
| CKD (OR) | TG independent | 0.96 (0.90; 1.03) | 0.286 | IVW | 0.002 | $7.7\times10^{2}$ | 135 |
| CKD (OR) | LDL-C & HDL-C & TG independent | 1.00 (0.74; 1.35) | 0.999 | IVW | < 0.001 | $4.2\times10^{1}$ | 238 |
| IBD (OR) | Total | 1.04 (0.87; 1.24) | 0.659 | MR Egger | 0.006 | $1.2\times10^{2}$ | 95 |
| IBD (OR) | LDL-C independent | 0.64 (0.49; 0.84) | < 0.001 | IVW | 0.007 | $5.4\times10^{1}$ | 136 |
| IBD (OR) | HDL-C independent | 1.06 (0.98; 1.14) | 0.127 | IVW | 0.011 | $7.4\times10^{2}$ | 167 |
| IBD (OR) | TG independent | 1.15 (1.01; 1.30) | 0.035 | IVW | < 0.001 | $2.4\times10^{2}$ | 135 |
| IBD (OR) | LDL-C & HDL-C & TG independent | 0.35 (0.20; 0.62) | < 0.001 | IVW | < 0.001 | $1.2\times10^{1}$ | 241 |
| UC (OR) | Total | 1.00 (0.90; 1.10) | 0.971 | IVW | 0.051 | $3.7\times10^{2}$ | 96 |
| UC (OR) | LDL-C independent | 0.53 (0.37; 0.76) | < 0.001 | IVW | 0.075 | $2.9\times10^{1}$ | 136 |
| UC (OR) | HDL-C independent | 1.03 (0.94; 1.13) | 0.554 | IVW | 0.006 | $4.7\times10^{2}$ | 170 |
| UC (OR) | TG independent | 1.09 (0.92; 1.30) | 0.324 | IVW | 0.079 | $1.3\times10^{2}$ | 137 |
| UC (OR) | LDL-C & HDL-C & TG independent | 0.40 (0.20; 0.82) | 0.012 | IVW | 0.008 | 7.6 | 241 |
| CD (OR) | Total | 1.12 (1.00; 1.25) | 0.045 | IVW | 0.081 | $3.2\times10^{2}$ | 93 |
| CD (OR) | LDL-C independent | 1.15 (0.77; 1.70) | 0.501 | IVW | 0.097 | $2.4\times10^{1}$ | 134 |
| CD (OR) | HDL-C independent | 1.11 (1.00; 1.23) | 0.051 | IVW | 0.111 | $3.5\times10^{2}$ | 167 |
| CD (OR) | TG independent | 1.06 (0.89; 1.26) | 0.524 | IVW | 0.008 | $1.3\times10^{2}$ | 135 |
| CD (OR) | LDL-C & HDL-C & TG independent | 0.26 (0.12; 0.56) | < 0.001 | IVW | 0.040 | 6.2 | 240 |
| Arthritis (OR) | Total | 0.87 (0.78; 0.96) | 0.008 | IVW | 0.131 | $3.5\times10^{2}$ | 77 |
| Arthritis (OR) | LDL-C independent | 0.80 (0.56; 1.13) | 0.202 | IVW | 0.067 | $3.2\times10^{1}$ | 113 |

**Table S9:** (Multivariable) Mendelian randomization effect estimates for one standard deviation change in Apo-B *(continued)*

| Outcome | Effect type | Point estimate (95%CI) | P-value | Estimator | Heterogeneity P-value | Precision | No. variants |
|---|---|---|---|---|---|---|---|
| Arthritis (OR) | HDL-C independent | 0.88 (0.79; 0.97) | 0.008 | IVW | 0.191 | $4.0\times10^{2}$ | 135 |
| Arthritis (OR) | TG independent | 1.19 (0.96; 1.46) | 0.113 | MR Egger | 0.546 | $8.7\times10^{1}$ | 111 |
| Arthritis (OR) | LDL-C & HDL-C & TG independent | 0.67 (0.30; 1.50) | 0.330 | MR Egger | 0.158 | 6.0 | 198 |
| ALS (OR) | Total | 0.94 (0.78; 1.13) | 0.486 | MR Egger | 0.720 | $1.1\times10^{2}$ | 93 |
| ALS (OR) | LDL-C independent | 1.29 (1.00; 1.65) | 0.046 | IVW | 0.477 | $6.3\times10^{1}$ | 139 |
| ALS (OR) | HDL-C independent | 1.05 (0.99; 1.13) | 0.124 | IVW | 0.433 | $8.3\times10^{2}$ | 171 |
| ALS (OR) | TG independent | 1.10 (0.97; 1.24) | 0.128 | IVW | 0.030 | $2.6\times10^{2}$ | 135 |
| ALS (OR) | LDL-C & HDL-C & TG independent | 0.87 (0.50; 1.52) | 0.624 | IVW | 0.043 | $1.2\times10^{1}$ | 244 |
| MS (OR) | Total | 0.74 (0.52; 1.06) | 0.106 | MR Egger | 0.346 | $3.0\times10^{1}$ | 25 |
| MS (OR) | LDL-C independent | 0.72 (0.41; 1.26) | 0.248 | MR Egger | 0.136 | $1.2\times10^{1}$ | 27 |
| MS (OR) | HDL-C independent | 1.07 (0.94; 1.20) | 0.299 | IVW | 0.045 | $2.6\times10^{2}$ | 34 |
| MS (OR) | TG independent | 1.20 (0.97; 1.49) | 0.101 | IVW | 0.269 | $8.1\times10^{1}$ | 31 |
| MS (OR) | LDL-C & HDL-C & TG independent | 7.29 (1.92; 27.70) | 0.004 | IVW | 0.007 | 2.2 | 40 |
| Alzheimer (OR) | Total | 1.02 (1.01; 1.03) | < 0.001 | IVW | < 0.001 | $3.0\times10^{4}$ | 79 |
| Alzheimer (OR) | LDL-C independent | 0.98 (0.94; 1.03) | 0.500 | IVW | < 0.001 | $1.9\times10^{3}$ | 93 |
| Alzheimer (OR) | HDL-C independent | 1.04 (1.02; 1.06) | < 0.001 | MR Egger | 0.013 | $1.0\times10^{4}$ | 137 |
| Alzheimer (OR) | TG independent | 1.04 (1.02; 1.06) | < 0.001 | IVW | < 0.001 | $9.0\times10^{3}$ | 121 |
| Alzheimer (OR) | LDL-C & HDL-C & TG independent | 0.72 (0.66; 0.78) | < 0.001 | IVW | < 0.001 | $5.1\times10^{2}$ | 201 |
| Parkinson (OR) | Total | 0.98 (0.78; 1.24) | 0.887 | MR Egger | 0.264 | $6.9\times10^{1}$ | 92 |
| Parkinson (OR) | LDL-C independent | 1.11 (0.84; 1.47) | 0.475 | IVW | 0.392 | $4.8\times10^{1}$ | 135 |
| Parkinson (OR) | HDL-C independent | 0.89 (0.77; 1.02) | 0.098 | MR Egger | 0.061 | $2.0\times10^{2}$ | 163 |
| Parkinson (OR) | TG independent | 0.99 (0.82; 1.19) | 0.878 | MR Egger | 0.005 | $1.1\times10^{2}$ | 134 |
| Parkinson (OR) | LDL-C & HDL-C & TG independent | 1.67 (0.90; 3.13) | 0.105 | IVW | 0.005 | 9.9 | 238 |
| Pancreatic cancer (OR) | Total | 0.53 (0.30; 0.94) | 0.031 | MR Egger | 0.295 | $1.1\times10^{1}$ | 93 |
| Pancreatic cancer (OR) | LDL-C independent | 0.73 (0.32; 1.66) | 0.455 | MR Egger | 0.016 | 5.7 | 141 |
| Pancreatic cancer (OR) | HDL-C independent | 0.94 (0.74; 1.20) | 0.621 | IVW | 0.050 | $6.4\times10^{1}$ | 167 |
| Pancreatic cancer (OR) | TG independent | 0.86 (0.54; 1.36) | 0.510 | MR Egger | 0.432 | $1.8\times10^{1}$ | 140 |
| Pancreatic cancer (OR) | LDL-C & HDL-C & TG independent | 0.20 (0.03; 1.23) | 0.082 | IVW | < 0.001 | 1.2 | 244 |
| Colon cancer (OR) | Total | 0.97 (0.74; 1.26) | 0.796 | MR Egger | 0.255 | $5.4\times10^{1}$ | 93 |
| Colon cancer (OR) | LDL-C independent | 1.48 (1.06; 2.08) | 0.023 | IVW | 0.017 | $3.3\times10^{1}$ | 138 |
| Colon cancer (OR) | HDL-C independent | 1.15 (1.04; 1.26) | 0.005 | IVW | 0.020 | $4.3\times10^{2}$ | 168 |
| Colon cancer (OR) | TG independent | 1.08 (0.92; 1.28) | 0.358 | IVW | 0.036 | $1.4\times10^{2}$ | 140 |
| Colon cancer (OR) | LDL-C & HDL-C & TG independent | 1.49 (0.70; 3.19) | 0.299 | IVW | < 0.001 | 6.7 | 244 |
| Rectal cancer (OR) | Total | 1.31 (0.93; 1.85) | 0.124 | MR Egger | 0.156 | $3.2\times10^{1}$ | 93 |
| Rectal cancer (OR) | LDL-C independent | 0.69 (0.44; 1.09) | 0.111 | IVW | 0.472 | $1.9\times10^{1}$ | 140 |
| Rectal cancer (OR) | HDL-C independent | 1.21 (0.99; 1.49) | 0.067 | MR Egger | 0.203 | $8.9\times10^{1}$ | 166 |
| Rectal cancer (OR) | TG independent | 1.32 (1.01; 1.74) | 0.045 | MR Egger | 0.379 | $5.1\times10^{1}$ | 140 |
| Rectal cancer (OR) | LDL-C & HDL-C & TG independent | 0.76 (0.27; 2.11) | 0.592 | IVW | 0.313 | 3.6 | 245 |
| Melanoma (OR) | Total | 0.94 (0.78; 1.13) | 0.531 | MR Egger | 0.025 | $1.1\times10^{2}$ | 93 |

**Table S9:** (Multivariable) Mendelian randomization effect estimates for one standard deviation change in Apo-B *(continued)*

| Outcome | Effect type | Point estimate (95%CI) | P-value | Estimator | Heterogeneity P-value | Precision | No. variants |
|---|---|---|---|---|---|---|---|
| Melanoma (OR) | LDL-C independent | 1.13 (0.86; 1.48) | 0.368 | IVW | 0.138 | $5.3{\times}10^1$ | 140 |
| Melanoma (OR) | HDL-C independent | 1.05 (0.98; 1.13) | 0.140 | IVW | < 0.001 | $7.7{\times}10^2$ | 166 |
| Melanoma (OR) | TG independent | 0.91 (0.79; 1.06) | 0.235 | MR Egger | < 0.001 | $1.8{\times}10^2$ | 140 |
| Melanoma (OR) | LDL-C & HDL-C & TG independent | 1.22 (0.68; 2.18) | 0.511 | IVW | < 0.001 | $1.1{\times}10^1$ | 242 |
| Lung cancer (OR) | Total | 0.96 (0.70; 1.32) | 0.813 | MR Egger | 0.392 | $3.8{\times}10^1$ | 93 |
| Lung cancer (OR) | LDL-C independent | 1.24 (0.79; 1.93) | 0.351 | IVW | 0.080 | $1.9{\times}10^1$ | 140 |
| Lung cancer (OR) | HDL-C independent | 0.87 (0.77; 0.97) | 0.013 | IVW | 0.031 | $3.0{\times}10^2$ | 170 |
| Lung cancer (OR) | TG independent | 0.83 (0.68; 1.02) | 0.072 | IVW | 0.035 | $9.6{\times}10^1$ | 141 |
| Lung cancer (OR) | LDL-C & HDL-C & TG independent | 1.03 (0.40; 2.64) | 0.953 | IVW | 0.007 | 4.3 | 247 |

**Table S10:** (Multivariable) Mendelian randomization effect estimates for one standard deviation change in Apo-A1

| Outcome | Effect type | Point estimate (95%CI) | P-value | Estimator | Heterogeneity P-value | Precision | No. variants |
|---|---|---|---|---|---|---|---|
| CHD (OR) | Total | 0.89 (0.83; 0.96) | 0.002 | IVW | 0.008 | $7.6{\times}10^2$ | 18 |
| CHD (OR) | LDL-C independent | 0.81 (0.75; 0.87) | < 0.001 | IVW | < 0.001 | $6.2{\times}10^2$ | 43 |
| CHD (OR) | HDL-C independent | 2.01 (1.62; 2.49) | < 0.001 | MR Egger | 0.002 | $8.2{\times}10^1$ | 31 |
| CHD (OR) | TG independent | 0.78 (0.70; 0.87) | < 0.001 | MR Egger | < 0.001 | $3.5{\times}10^2$ | 38 |
| CHD (OR) | LDL-C & HDL-C & TG independent | 0.76 (0.40; 1.44) | 0.403 | IVW | < 0.001 | 9.4 | 63 |
| HF (OR) | Total | 0.95 (0.86; 1.06) | 0.394 | MR Egger | 0.039 | $3.2{\times}10^2$ | 66 |
| HF (OR) | LDL-C independent | 0.91 (0.85; 0.97) | 0.003 | MR Egger | < 0.001 | $9.3{\times}10^2$ | 166 |
| HF (OR) | HDL-C independent | 1.17 (1.01; 1.36) | 0.042 | IVW | < 0.001 | $1.7{\times}10^2$ | 95 |
| HF (OR) | TG independent | 0.94 (0.88; 1.00) | 0.069 | MR Egger | < 0.001 | $8.5{\times}10^2$ | 128 |
| HF (OR) | LDL-C & HDL-C & TG independent | 0.99 (0.73; 1.33) | 0.922 | IVW | < 0.001 | $4.3{\times}10^1$ | 227 |
| AF (OR) | Total | 1.00 (0.96; 1.03) | 0.857 | IVW | 0.012 | $3.4{\times}10^3$ | 71 |
| AF (OR) | LDL-C independent | 1.02 (0.99; 1.06) | 0.215 | IVW | 0.002 | $3.1{\times}10^3$ | 185 |
| AF (OR) | HDL-C independent | 1.25 (1.08; 1.44) | 0.002 | IVW | 0.017 | $1.9{\times}10^2$ | 100 |
| AF (OR) | TG independent | 1.01 (0.96; 1.06) | 0.775 | MR Egger | < 0.001 | $1.5{\times}10^3$ | 141 |
| AF (OR) | LDL-C & HDL-C & TG independent | 1.30 (1.03; 1.64) | 0.028 | IVW | < 0.001 | $7.0{\times}10^1$ | 251 |
| Any stroke (OR) | Total | 0.98 (0.94; 1.02) | 0.374 | IVW | 0.014 | $1.9{\times}10^3$ | 70 |
| Any stroke (OR) | LDL-C independent | 0.97 (0.93; 1.02) | 0.295 | IVW | < 0.001 | $1.6{\times}10^3$ | 173 |
| Any stroke (OR) | HDL-C independent | 1.04 (0.86; 1.25) | 0.704 | IVW | 0.023 | $1.1{\times}10^2$ | 98 |
| Any stroke (OR) | TG independent | 0.99 (0.95; 1.04) | 0.755 | IVW | 0.051 | $1.7{\times}10^3$ | 139 |
| Any stroke (OR) | LDL-C & HDL-C & TG independent | 1.34 (0.95; 1.89) | 0.092 | IVW | 0.003 | $3.3{\times}10^1$ | 237 |
| Any ischemic stroke (OR) | Total | 1.00 (0.95; 1.04) | 0.838 | IVW | 0.011 | $1.7{\times}10^3$ | 71 |
| Any ischemic stroke (OR) | LDL-C independent | 0.98 (0.93; 1.03) | 0.452 | IVW | 0.003 | $1.3{\times}10^3$ | 171 |
| Any ischemic stroke (OR) | HDL-C independent | 1.09 (0.89; 1.34) | 0.396 | IVW | 0.038 | $9.3{\times}10^1$ | 98 |
| Any ischemic stroke (OR) | TG independent | 1.01 (0.96; 1.06) | 0.808 | IVW | 0.056 | $1.5{\times}10^3$ | 139 |

**Table S10:** (Multivariable) Mendelian randomization effect estimates for one standard deviation change in Apo-A1 *(continued)*

| Outcome | Effect type | Point estimate (95%CI) | P-value | Estimator | Heterogeneity P-value | Precision | No. variants |
|---|---|---|---|---|---|---|---|
| Any ischemic stroke (OR) | LDL-C & HDL-C & TG independent | 1.31 (0.90; 1.90) | 0.152 | IVW | 0.012 | $2.8\times10^{1}$ | 236 |
| Large artery stroke (OR) | Total | 1.04 (0.92; 1.17) | 0.544 | IVW | 0.004 | $2.6\times10^{2}$ | 71 |
| Large artery stroke (OR) | LDL-C independent | 0.90 (0.74; 1.09) | 0.277 | MR Egger | < 0.001 | $1.1\times10^{2}$ | 173 |
| Large artery stroke (OR) | HDL-C independent | 1.07 (0.60; 1.88) | 0.825 | IVW | 0.050 | $1.2\times10^{1}$ | 98 |
| Large artery stroke (OR) | TG independent | 1.10 (0.98; 1.24) | 0.105 | IVW | 0.015 | $2.8\times10^{2}$ | 140 |
| Large artery stroke (OR) | LDL-C & HDL-C & TG independent | 0.59 (0.23; 1.51) | 0.268 | IVW | 0.002 | 4.3 | 237 |
| Cardioembolic stroke (OR) | Total | 1.01 (0.92; 1.12) | 0.780 | IVW | 0.343 | $4.1\times10^{2}$ | 71 |
| Cardioembolic stroke (OR) | LDL-C independent | 1.00 (0.85; 1.18) | 0.968 | MR Egger | 0.067 | $1.4\times10^{2}$ | 173 |
| Cardioembolic stroke (OR) | HDL-C independent | 0.91 (0.61; 1.35) | 0.630 | IVW | 0.725 | $2.4\times10^{1}$ | 98 |
| Cardioembolic stroke (OR) | TG independent | 0.98 (0.90; 1.08) | 0.744 | IVW | 0.403 | $4.5\times10^{2}$ | 139 |
| Cardioembolic stroke (OR) | LDL-C & HDL-C & TG independent | 2.03 (0.99; 4.15) | 0.052 | IVW | 0.043 | 7.5 | 237 |
| Small vessel stroke (OR) | Total | 1.06 (0.76; 1.48) | 0.742 | MR Egger | 0.221 | $3.5\times10^{1}$ | 69 |
| Small vessel stroke (OR) | LDL-C independent | 0.98 (0.87; 1.11) | 0.791 | IVW | 0.014 | $2.4\times10^{2}$ | 176 |
| Small vessel stroke (OR) | HDL-C independent | 1.04 (0.67; 1.63) | 0.852 | IVW | 0.024 | $2.0\times10^{1}$ | 99 |
| Small vessel stroke (OR) | TG independent | 1.00 (0.90; 1.11) | 0.967 | IVW | < 0.001 | $3.3\times10^{2}$ | 139 |
| Small vessel stroke (OR) | LDL-C & HDL-C & TG independent | 0.59 (0.25; 1.41) | 0.239 | IVW | 0.002 | 5.1 | 239 |
| cIMT (mm) | Total | 0.00 (-0.01; 0.01) | 0.806 | MR Egger | 0.031 | $3.7\times10^{4}$ | 67 |
| cIMT (mm) | LDL-C independent | -0.01 (-0.01; -0.00) | < 0.001 | MR Egger | < 0.001 | $1.3\times10^{5}$ | 178 |
| cIMT (mm) | HDL-C independent | 0.01 (-0.01; 0.02) | 0.390 | IVW | 0.007 | $1.9\times10^{4}$ | 98 |
| cIMT (mm) | TG independent | 0.00 (-0.00; 0.00) | 0.805 | IVW | < 0.001 | $3.4\times10^{5}$ | 139 |
| cIMT (mm) | LDL-C & HDL-C & TG independent | 0.01 (-0.02; 0.03) | 0.529 | IVW | < 0.001 | $6.1\times10^{3}$ | 243 |
| Carotid plaque (OR) | Total | 0.87 (0.80; 0.95) | 0.002 | IVW | 0.529 | $5.3\times10^{2}$ | 66 |
| Carotid plaque (OR) | LDL-C independent | 0.76 (0.66; 0.88) | < 0.001 | MR Egger | 0.066 | $1.8\times10^{2}$ | 168 |
| Carotid plaque (OR) | HDL-C independent | 1.37 (0.92; 2.04) | 0.120 | IVW | 0.181 | $2.4\times10^{1}$ | 93 |
| Carotid plaque (OR) | TG independent | 0.93 (0.85; 1.00) | 0.060 | IVW | 0.044 | $5.9\times10^{2}$ | 132 |
| Carotid plaque (OR) | LDL-C & HDL-C & TG independent | 1.61 (0.85; 3.07) | 0.146 | IVW | 0.009 | 9.3 | 231 |
| Glucose (mmol/l) | Total | -0.01 (-0.03; 0.00) | 0.065 | IVW | 0.343 | $1.7\times10^{4}$ | 70 |
| Glucose (mmol/l) | LDL-C independent | 0.01 (-0.02; 0.03) | 0.565 | MR Egger | 0.047 | $8.1\times10^{3}$ | 180 |
| Glucose (mmol/l) | HDL-C independent | -0.03 (-0.09; 0.03) | 0.325 | MR Egger | 0.275 | $1.1\times10^{3}$ | 95 |
| Glucose (mmol/l) | TG independent | -0.01 (-0.03; 0.00) | 0.086 | IVW | < 0.001 | $2.0\times10^{4}$ | 136 |
| Glucose (mmol/l) | LDL-C & HDL-C & TG independent | 0.01 (-0.10; 0.11) | 0.920 | IVW | < 0.001 | $3.8\times10^{2}$ | 237 |
| HbA1c (mmol/mol) | Total | -0.12 (-0.19; -0.05) | < 0.001 | IVW | < 0.001 | $7.7\times10^{2}$ | 72 |
| HbA1c (mmol/mol) | LDL-C independent | -0.11 (-0.18; -0.03) | 0.006 | IVW | < 0.001 | $6.8\times10^{2}$ | 177 |
| HbA1c (mmol/mol) | HDL-C independent | 0.03 (-0.26; 0.32) | 0.836 | IVW | < 0.001 | $4.5\times10^{1}$ | 93 |
| HbA1c (mmol/mol) | TG independent | -0.10 (-0.22; 0.01) | 0.073 | MR Egger | < 0.001 | $3.0\times10^{2}$ | 133 |
| HbA1c (mmol/mol) | LDL-C & HDL-C & TG independent | 0.06 (-0.43; 0.56) | 0.797 | MR Egger | < 0.001 | $1.6\times10^{1}$ | 236 |
| T2DM (OR) | Total | 0.95 (0.92; 0.98) | 0.004 | IVW | < 0.001 | $3.6\times10^{3}$ | 63 |
| T2DM (OR) | LDL-C independent | 1.02 (0.97; 1.07) | 0.373 | MR Egger | < 0.001 | $1.6\times10^{3}$ | 162 |
| T2DM (OR) | HDL-C independent | 1.16 (1.02; 1.32) | 0.019 | IVW | < 0.001 | $2.4\times10^{2}$ | 89 |

**Table S10:** (Multivariable) Mendelian randomization effect estimates for one standard deviation change in Apo-A1 *(continued)*

| Outcome | Effect type | Point estimate (95%CI) | P-value | Estimator | Heterogeneity P-value | Precision | No. variants |
|---|---|---|---|---|---|---|---|
| T2DM (OR) | TG independent | 0.91 (0.89; 0.94) | < 0.001 | IVW | < 0.001 | $4.3\times10^{3}$ | 122 |
| T2DM (OR) | LDL-C & HDL-C & TG independent | 0.90 (0.72; 1.13) | 0.379 | IVW | < 0.001 | $7.3\times10^{1}$ | 216 |
| CRP (mg/L) | Total | 0.07 (0.02; 0.12) | 0.009 | IVW | 0.002 | $1.5\times10^{3}$ | 67 |
| CRP (mg/L) | LDL-C independent | 0.14 (0.08; 0.19) | < 0.001 | IVW | < 0.001 | $1.2\times10^{3}$ | 153 |
| CRP (mg/L) | HDL-C independent | 0.19 (-0.01; 0.38) | 0.061 | IVW | 0.002 | $9.9\times10^{1}$ | 94 |
| CRP (mg/L) | TG independent | 0.06 (0.01; 0.11) | 0.020 | IVW | < 0.001 | $1.6\times10^{3}$ | 125 |
| CRP (mg/L) | LDL-C & HDL-C & TG independent | -0.74 (-1.10; -0.37) | < 0.001 | IVW | < 0.001 | $2.9\times10^{1}$ | 224 |
| SBP (mmHg) | Total | -0.17 (-0.58; 0.24) | 0.420 | MR Egger | < 0.001 | $2.3\times10^{1}$ | 60 |
| SBP (mmHg) | LDL-C independent | -0.45 (-0.72; -0.19) | < 0.001 | MR Egger | < 0.001 | $5.6\times10^{1}$ | 141 |
| SBP (mmHg) | HDL-C independent | 2.09 (1.49; 2.69) | < 0.001 | IVW | < 0.001 | $1.1\times10^{1}$ | 85 |
| SBP (mmHg) | TG independent | -0.10 (-0.25; 0.05) | 0.182 | IVW | < 0.001 | $1.7\times10^{2}$ | 116 |
| SBP (mmHg) | LDL-C & HDL-C & TG independent | 0.66 (-0.57; 1.89) | 0.292 | IVW | < 0.001 | 2.5 | 192 |
| DBP (mmHg) | Total | -0.09 (-0.18; -0.00) | 0.043 | IVW | 0.017 | $4.8\times10^{2}$ | 61 |
| DBP (mmHg) | LDL-C independent | -0.17 (-0.32; -0.03) | 0.020 | MR Egger | < 0.001 | $1.9\times10^{2}$ | 145 |
| DBP (mmHg) | HDL-C independent | 0.76 (0.40; 1.13) | < 0.001 | IVW | 0.181 | $2.9\times10^{1}$ | 85 |
| DBP (mmHg) | TG independent | -0.05 (-0.13; 0.04) | 0.288 | IVW | < 0.001 | $5.1\times10^{2}$ | 114 |
| DBP (mmHg) | LDL-C & HDL-C & TG independent | -0.04 (-0.70; 0.63) | 0.907 | MR Egger | < 0.001 | 8.7 | 195 |
| PBL (OR) | Total | 1.04 (0.82; 1.32) | 0.718 | IVW | 0.050 | $6.9\times10^{1}$ | 36 |
| PBL (OR) | LDL-C independent | 1.06 (0.83; 1.35) | 0.627 | IVW | 0.010 | $6.6\times10^{1}$ | 83 |
| PBL (OR) | HDL-C independent | 0.95 (0.40; 2.22) | 0.898 | IVW | 0.203 | 5.3 | 52 |
| PBL (OR) | TG independent | 1.13 (0.90; 1.42) | 0.291 | IVW | 0.002 | $7.4\times10^{1}$ | 69 |
| PBL (OR) | LDL-C & HDL-C & TG independent | 1.49 (0.27; 8.22) | 0.646 | IVW | 0.005 | 1.3 | 118 |
| BUN (mg/dl) | Total | -0.00 (-0.01; 0.01) | 0.711 | MR Egger | 0.003 | $2.6\times10^{4}$ | 68 |
| BUN (mg/dl) | LDL-C independent | -0.00 (-0.01; 0.01) | 0.925 | MR Egger | < 0.001 | $8.1\times10^{4}$ | 172 |
| BUN (mg/dl) | HDL-C independent | -0.00 (-0.02; 0.02) | 0.990 | IVW | 0.005 | $1.2\times10^{4}$ | 93 |
| BUN (mg/dl) | TG independent | 0.00 (-0.01; 0.01) | 0.932 | MR Egger | < 0.001 | $7.6\times10^{4}$ | 133 |
| BUN (mg/dl) | LDL-C & HDL-C & TG independent | -0.03 (-0.07; 0.00) | 0.056 | MR Egger | < 0.001 | $3.5\times10^{3}$ | 231 |
| eGFR (SD of log(eGFR)) | Total | 0.00 (-0.00; 0.00) | 0.790 | MR Egger | 0.044 | $2.0\times10^{5}$ | 68 |
| eGFR (SD of log(eGFR)) | LDL-C independent | -0.00 (-0.00; 0.00) | 0.711 | MR Egger | < 0.001 | $4.9\times10^{5}$ | 164 |
| eGFR (SD of log(eGFR)) | HDL-C independent | -0.01 (-0.02; -0.00) | 0.006 | MR Egger | 0.006 | $8.2\times10^{4}$ | 94 |
| eGFR (SD of log(eGFR)) | TG independent | -0.00 (-0.00; 0.00) | 0.620 | IVW | < 0.001 | $1.3\times10^{6}$ | 127 |
| eGFR (SD of log(eGFR)) | LDL-C & HDL-C & TG independent | -0.01 (-0.03; -0.00) | 0.022 | IVW | < 0.001 | $2.4\times10^{4}$ | 225 |
| CKD (OR) | Total | 0.98 (0.86; 1.12) | 0.793 | MR Egger | 0.200 | $2.2\times10^{2}$ | 70 |
| CKD (OR) | LDL-C independent | 1.00 (0.93; 1.08) | 0.979 | MR Egger | 0.007 | $7.0\times10^{2}$ | 174 |
| CKD (OR) | HDL-C independent | 1.03 (0.85; 1.26) | 0.741 | IVW | 0.372 | $9.7\times10^{1}$ | 97 |
| CKD (OR) | TG independent | 0.98 (0.94; 1.03) | 0.473 | IVW | < 0.001 | $2.0\times10^{3}$ | 138 |
| CKD (OR) | LDL-C & HDL-C & TG independent | 1.40 (1.00; 1.97) | 0.052 | IVW | 0.003 | $3.3\times10^{1}$ | 240 |
| IBD (OR) | Total | 0.92 (0.85; 1.00) | 0.057 | IVW | < 0.001 | $5.5\times10^{2}$ | 70 |
| IBD (OR) | LDL-C independent | 0.89 (0.82; 0.97) | 0.010 | IVW | < 0.001 | $5.1\times10^{2}$ | 180 |

**Table S10:** (Multivariable) Mendelian randomization effect estimates for one standard deviation change in Apo-A1 *(continued)*

| Outcome | Effect type | Point estimate (95%CI) | P-value | Estimator | Heterogeneity P-value | Precision | No. variants |
|---|---|---|---|---|---|---|---|
| IBD (OR) | HDL-C independent | 1.02 (0.72; 1.43) | 0.930 | IVW | 0.011 | $3.3\times10^{1}$ | 98 |
| IBD (OR) | TG independent | 0.98 (0.90; 1.06) | 0.568 | IVW | < 0.001 | $6.3\times10^{2}$ | 139 |
| IBD (OR) | LDL-C & HDL-C & TG independent | 0.83 (0.45; 1.54) | 0.552 | IVW | < 0.001 | $1.0\times10^{1}$ | 240 |
| UC (OR) | Total | 0.94 (0.84; 1.04) | 0.222 | IVW | < 0.001 | $3.5\times10^{2}$ | 70 |
| UC (OR) | LDL-C independent | 0.87 (0.78; 0.97) | 0.015 | IVW | < 0.001 | $3.2\times10^{2}$ | 179 |
| UC (OR) | HDL-C independent | 0.97 (0.63; 1.49) | 0.892 | IVW | 0.005 | $2.1\times10^{1}$ | 98 |
| UC (OR) | TG independent | 0.96 (0.87; 1.06) | 0.417 | IVW | < 0.001 | $3.8\times10^{2}$ | 139 |
| UC (OR) | LDL-C & HDL-C & TG independent | 0.90 (0.41; 1.95) | 0.781 | IVW | 0.002 | 6.3 | 240 |
| CD (OR) | Total | 0.85 (0.61; 1.18) | 0.338 | MR Egger | 0.205 | $3.5\times10^{1}$ | 68 |
| CD (OR) | LDL-C independent | 0.99 (0.87; 1.12) | 0.884 | IVW | 0.479 | $2.5\times10^{2}$ | 178 |
| CD (OR) | HDL-C independent | 1.37 (0.85; 2.19) | 0.194 | IVW | 0.844 | $1.7\times10^{1}$ | 96 |
| CD (OR) | TG independent | 0.94 (0.77; 1.14) | 0.520 | MR Egger | 0.066 | $1.0\times10^{2}$ | 138 |
| CD (OR) | LDL-C & HDL-C & TG independent | 1.02 (0.43; 2.42) | 0.966 | IVW | 0.133 | 5.1 | 241 |
| Arthritis (OR) | Total | 1.09 (0.95; 1.24) | 0.231 | IVW | 0.075 | $2.1\times10^{2}$ | 60 |
| Arthritis (OR) | LDL-C independent | 1.07 (0.94; 1.23) | 0.311 | IVW | 0.068 | $2.1\times10^{2}$ | 147 |
| Arthritis (OR) | HDL-C independent | 1.20 (0.75; 1.91) | 0.454 | MR Egger | 0.519 | $1.8\times10^{1}$ | 82 |
| Arthritis (OR) | TG independent | 1.12 (0.99; 1.25) | 0.064 | IVW | 0.107 | $2.8\times10^{2}$ | 114 |
| Arthritis (OR) | LDL-C & HDL-C & TG independent | 0.62 (0.26; 1.52) | 0.297 | IVW | 0.074 | 4.9 | 196 |
| ALS (OR) | Total | 0.96 (0.75; 1.23) | 0.751 | MR Egger | 0.292 | $6.4\times10^{1}$ | 72 |
| ALS (OR) | LDL-C independent | 1.04 (0.95; 1.14) | 0.382 | IVW | 0.376 | $4.8\times10^{2}$ | 186 |
| ALS (OR) | HDL-C independent | 0.94 (0.66; 1.35) | 0.745 | MR Egger | 0.324 | $2.9\times10^{1}$ | 98 |
| ALS (OR) | TG independent | 1.09 (1.01; 1.18) | 0.025 | IVW | 0.022 | $6.3\times10^{2}$ | 139 |
| ALS (OR) | LDL-C & HDL-C & TG independent | 1.21 (0.65; 2.25) | 0.552 | IVW | 0.113 | 9.9 | 242 |
| MS (OR) | Total | 1.10 (0.95; 1.27) | 0.213 | IVW | 0.040 | $1.8\times10^{2}$ | 13 |
| MS (OR) | LDL-C independent | 1.02 (0.86; 1.21) | 0.840 | IVW | 0.021 | $1.3\times10^{2}$ | 31 |
| MS (OR) | HDL-C independent | 0.97 (0.67; 1.41) | 0.885 | IVW | 0.041 | $2.8\times10^{1}$ | 20 |
| MS (OR) | TG independent | 1.07 (0.91; 1.26) | 0.406 | IVW | 0.198 | $1.5\times10^{2}$ | 26 |
| MS (OR) | LDL-C & HDL-C & TG independent | 0.73 (0.19; 2.74) | 0.640 | IVW | < 0.001 | 2.2 | 43 |
| Alzheimer (OR) | Total | 1.02 (0.99; 1.05) | 0.218 | MR Egger | 0.006 | $4.1\times10^{3}$ | 70 |
| Alzheimer (OR) | LDL-C independent | 1.03 (1.01; 1.05) | 0.008 | MR Egger | < 0.001 | $8.2\times10^{3}$ | 135 |
| Alzheimer (OR) | HDL-C independent | 1.03 (0.98; 1.08) | 0.215 | MR Egger | 0.155 | $1.7\times10^{3}$ | 93 |
| Alzheimer (OR) | TG independent | 1.00 (0.99; 1.01) | 0.752 | IVW | < 0.001 | $3.0\times10^{4}$ | 132 |
| Alzheimer (OR) | LDL-C & HDL-C & TG independent | 1.08 (0.98; 1.18) | 0.102 | MR Egger | < 0.001 | $4.5\times10^{2}$ | 199 |
| Parkinson (OR) | Total | 1.02 (0.92; 1.13) | 0.698 | IVW | 0.165 | $4.0\times10^{2}$ | 71 |
| Parkinson (OR) | LDL-C independent | 0.99 (0.86; 1.14) | 0.840 | MR Egger | 0.105 | $1.9\times10^{2}$ | 178 |
| Parkinson (OR) | HDL-C independent | 1.62 (1.11; 2.37) | 0.013 | MR Egger | 0.633 | $2.7\times10^{1}$ | 95 |
| Parkinson (OR) | TG independent | 1.02 (0.93; 1.11) | 0.684 | IVW | 0.008 | $5.0\times10^{2}$ | 140 |
| Parkinson (OR) | LDL-C & HDL-C & TG independent | 1.63 (0.83; 3.20) | 0.159 | IVW | < 0.001 | 8.4 | 241 |
| Pancreatic cancer (OR) | Total | 1.04 (0.54; 1.99) | 0.913 | MR Egger | 0.562 | 9.1 | 75 |

**Table S10:** (Multivariable) Mendelian randomization effect estimates for one standard deviation change in Apo-A1 *(continued)*

| Outcome | Effect type | Point estimate (95%CI) | P-value | Estimator | Heterogeneity P-value | Precision | No. variants |
|---|---|---|---|---|---|---|---|
| Pancreatic cancer (OR) | LDL-C independent | 1.38 (1.03; 1.84) | 0.030 | IVW | 0.010 | $4.6\times10^{1}$ | 182 |
| Pancreatic cancer (OR) | HDL-C independent | 0.26 (0.08; 0.81) | 0.020 | IVW | 0.214 | 3.0 | 101 |
| Pancreatic cancer (OR) | TG independent | 1.18 (0.89; 1.55) | 0.249 | IVW | 0.162 | $5.0\times10^{1}$ | 143 |
| Pancreatic cancer (OR) | LDL-C & HDL-C & TG independent | 1.66 (0.27; 10.27) | 0.583 | MR Egger | 0.017 | 1.2 | 250 |
| Colon cancer (OR) | Total | 1.13 (0.99; 1.28) | 0.061 | IVW | 0.131 | $2.4\times10^{2}$ | 74 |
| Colon cancer (OR) | LDL-C independent | 0.98 (0.82; 1.16) | 0.807 | MR Egger | 0.008 | $1.3\times10^{2}$ | 185 |
| Colon cancer (OR) | HDL-C independent | 1.59 (0.98; 2.57) | 0.060 | IVW | 0.022 | $1.7\times10^{1}$ | 100 |
| Colon cancer (OR) | TG independent | 1.05 (0.94; 1.17) | 0.371 | IVW | 0.004 | $3.2\times10^{2}$ | 142 |
| Colon cancer (OR) | LDL-C & HDL-C & TG independent | 1.79 (0.83; 3.88) | 0.137 | IVW | < 0.001 | 6.5 | 250 |
| Rectal cancer (OR) | Total | 1.30 (0.84; 2.01) | 0.231 | MR Egger | 0.051 | $2.0\times10^{1}$ | 72 |
| Rectal cancer (OR) | LDL-C independent | 1.00 (0.83; 1.19) | 0.984 | IVW | 0.142 | $1.2\times10^{2}$ | 179 |
| Rectal cancer (OR) | HDL-C independent | 0.94 (0.49; 1.80) | 0.858 | IVW | 0.026 | 9.2 | 100 |
| Rectal cancer (OR) | TG independent | 1.08 (0.92; 1.27) | 0.351 | IVW | 0.054 | $1.4\times10^{2}$ | 141 |
| Rectal cancer (OR) | LDL-C & HDL-C & TG independent | 1.19 (0.39; 3.68) | 0.760 | IVW | 0.401 | 3.0 | 245 |
| Melanoma (OR) | Total | 1.02 (0.94; 1.12) | 0.580 | IVW | 0.012 | $5.0\times10^{2}$ | 73 |
| Melanoma (OR) | LDL-C independent | 1.03 (0.94; 1.13) | 0.539 | IVW | 0.011 | $4.6\times10^{2}$ | 183 |
| Melanoma (OR) | HDL-C independent | 1.31 (0.90; 1.90) | 0.162 | IVW | 0.012 | $2.7\times10^{1}$ | 98 |
| Melanoma (OR) | TG independent | 1.01 (0.93; 1.10) | 0.745 | IVW | < 0.001 | $5.4\times10^{2}$ | 140 |
| Melanoma (OR) | LDL-C & HDL-C & TG independent | 1.37 (0.75; 2.51) | 0.300 | IVW | < 0.001 | $1.1\times10^{1}$ | 247 |
| Lung cancer (OR) | Total | 0.91 (0.64; 1.28) | 0.580 | MR Egger | 0.449 | $3.1\times10^{1}$ | 74 |
| Lung cancer (OR) | LDL-C independent | 1.17 (1.01; 1.36) | 0.036 | IVW | 0.015 | $1.7\times10^{2}$ | 184 |
| Lung cancer (OR) | HDL-C independent | 1.06 (0.56; 2.01) | 0.861 | IVW | 0.091 | 9.3 | 100 |
| Lung cancer (OR) | TG independent | 1.03 (0.90; 1.18) | 0.678 | IVW | 0.553 | $2.1\times10^{2}$ | 143 |
| Lung cancer (OR) | LDL-C & HDL-C & TG independent | 0.66 (0.23; 1.87) | 0.434 | IVW | 0.058 | 3.5 | 248 |

**Table S11:** *cis* Mendelian randomization effect estimates for one standard deviation change in Apo-B

| Outcome | Effect type | Point estimate (95%CI) | P-value | Estimator | Heterogeneity P-value | Precision | No. variants |
|---|---|---|---|---|---|---|---|
| CHD (OR) | Total | 2.46 (1.68; 3.61) | < 0.001 | MR Egger | 0.331 | $2.6\times10^{1}$ | 3 |
| HF (OR) | Total | 0.86 (0.81; 0.92) | < 0.001 | IVW | 0.379 | $9.4\times10^{2}$ | 8 |
| AF (OR) | Total | 0.97 (0.92; 1.02) | 0.211 | IVW | 0.493 | $1.3\times10^{3}$ | 8 |
| Any stroke (OR) | Total | 1.00 (0.77; 1.31) | 0.988 | MR Egger | 0.591 | $5.5\times10^{1}$ | 8 |
| Any ischemic stroke (OR) | Total | 0.93 (0.70; 1.24) | 0.642 | MR Egger | 0.496 | $4.7\times10^{1}$ | 8 |
| Large artery stroke (OR) | Total | 0.93 (0.76; 1.14) | 0.478 | IVW | 0.743 | $9.1\times10^{1}$ | 8 |
| Cardioembolic stroke (OR) | Total | 0.68 (0.39; 1.18) | 0.175 | MR Egger | 0.832 | $1.3\times10^{1}$ | 8 |
| Small vessel stroke (OR) | Total | 0.73 (0.33; 1.58) | 0.418 | MR Egger | 0.226 | 6.4 | 8 |
| cIMT (mm) | Total | 0.02 (0.02; 0.03) | < 0.001 | IVW | 0.316 | $9.7\times10^{4}$ | 8 |
| Carotid plaque (OR) | Total | 1.63 (1.42; 1.86) | < 0.001 | IVW | 0.545 | $2.0\times10^{2}$ | 8 |

**Table S11:** *cis* Mendelian randomization effect estimates for one standard deviation change in Apo-B *(continued)*

| Outcome | Effect type | Point estimate (95%CI) | P-value | Estimator | Heterogeneity P-value | Precision | No. variants |
|---|---|---|---|---|---|---|---|
| Glucose (mmol/l) | Total | -0.05 (-0.07; -0.02) | < 0.001 | IVW | 0.517 | $6.8\times10^3$ | 8 |
| HbA1c (mmol/mol) | Total | -0.40 (-0.53; -0.28) | < 0.001 | IVW | 0.998 | $2.6\times10^2$ | 8 |
| T2DM (OR) | Total | 0.91 (0.87; 0.96) | < 0.001 | IVW | 0.854 | $1.5\times10^3$ | 8 |
| CRP (mg/L) | Total | 0.02 (-0.06; 0.10) | 0.660 | IVW | 0.898 | $5.5\times10^2$ | 8 |
| SBP (mmHg) | Total | 0.46 (0.22; 0.70) | < 0.001 | IVW | 0.015 | $6.6\times10^1$ | 8 |
| DBP (mmHg) | Total | -0.17 (-0.31; -0.03) | 0.015 | IVW | 0.425 | $2.0\times10^2$ | 8 |
| PBL (OR) | Total | 0.87 (0.33; 2.25) | 0.771 | IVW | 0.971 | 4.2 | 4 |
| BUN (mg/dl) | Total | -0.01 (-0.02; -0.00) | 0.004 | IVW | 0.37 | $7.1\times10^4$ | 8 |
| eGFR (SD of log(eGFR)) | Total | -0.00 (-0.01; -0.00) | 0.047 | IVW | 0.828 | $5.2\times10^5$ | 8 |
| CKD (OR) | Total | 1.06 (0.96; 1.16) | 0.272 | IVW | 0.101 | $4.2\times10^2$ | 8 |
| IBD (OR) | Total | 0.97 (0.84; 1.11) | 0.662 | IVW | 0.585 | $2.0\times10^2$ | 8 |
| UC (OR) | Total | 0.84 (0.71; 1.00) | 0.052 | IVW | 0.547 | $1.3\times10^2$ | 8 |
| CD (OR) | Total | 1.14 (0.94; 1.37) | 0.188 | IVW | 0.767 | $1.1\times10^2$ | 8 |
| Arthritis (OR) | Total | 1.32 (1.06; 1.63) | 0.011 | IVW | 0.366 | $8.4\times10^1$ | 7 |
| ALS (OR) | Total | 1.27 (1.11; 1.46) | < 0.001 | IVW | 0.9 | $2.0\times10^2$ | 8 |
| MS (OR) | Total | 0.91 (0.66; 1.25) | 0.564 | IVW | 0.791 | $3.9\times10^1$ | 3 |
| Alzheimer (OR) | Total | 0.99 (0.98; 1.01) | 0.348 | IVW | 0.484 | $1.3\times10^4$ | 8 |
| Parkinson (OR) | Total | 0.82 (0.71; 0.94) | 0.004 | IVW | 0.755 | $2.0\times10^2$ | 8 |
| Pancreatic cancer (OR) | Total | 0.55 (0.34; 0.89) | 0.015 | IVW | 0.302 | $1.6\times10^1$ | 8 |
| Colon cancer (OR) | Total | 0.70 (0.58; 0.84) | < 0.001 | IVW | 0.721 | $1.1\times10^2$ | 8 |
| Rectal cancer (OR) | Total | 0.97 (0.72; 1.31) | 0.834 | IVW | 0.161 | $4.3\times10^1$ | 8 |
| Melanoma (OR) | Total | 0.82 (0.71; 0.95) | 0.007 | IVW | 0.664 | $1.9\times10^2$ | 8 |
| Lung cancer (OR) | Total | 0.68 (0.54; 0.86) | < 0.001 | IVW | 0.981 | $7.4\times10^1$ | 8 |

**Table S12:** *cis* Mendelian randomization effect estimates for one standard deviation change in Apo-A1

| Outcome | Effect type | Point estimate (95%CI) | P-value | Estimator | Heterogeneity P-value | Precision | No. variants |
|---|---|---|---|---|---|---|---|
| CHD (OR) | Total | 1.76 (0.96; 3.22) | 0.067 | Wald | NA | $1.1\times10^1$ | 1 |
| HF (OR) | Total | 0.79 (0.59; 1.05) | 0.104 | IVW | 0.389 | $4.6\times10^1$ | 2 |
| AF (OR) | Total | 1.44 (1.12; 1.84) | 0.004 | IVW | 0.833 | $6.2\times10^1$ | 2 |
| Any stroke (OR) | Total | 0.82 (0.58; 1.17) | 0.269 | IVW | 0.949 | $3.1\times10^1$ | 2 |
| Any ischemic stroke (OR) | Total | 0.73 (0.49; 1.09) | 0.126 | IVW | 0.932 | $2.4\times10^1$ | 2 |
| Large artery stroke (OR) | Total | 1.43 (0.57; 3.63) | 0.448 | IVW | 0.48 | 4.5 | 2 |
| Cardioembolic stroke (OR) | Total | 0.62 (0.30; 1.28) | 0.194 | IVW | 0.781 | 7.3 | 2 |
| Small vessel stroke (OR) | Total | 1.31 (0.55; 3.11) | 0.546 | IVW | 0.602 | 5.1 | 2 |
| cIMT (mm) | Total | -0.04 (-0.07; -0.01) | 0.012 | IVW | 0.231 | $3.9\times10^3$ | 2 |
| Carotid plaque (OR) | Total | 0.61 (0.33; 1.12) | 0.114 | IVW | 0.769 | $1.0\times10^1$ | 2 |
| Glucose (mmol/l) | Total | 0.10 (-0.02; 0.21) | 0.089 | IVW | 0.6 | $2.9\times10^2$ | 2 |
| HbA1c (mmol/mol) | Total | 1.13 (0.55; 1.71) | < 0.001 | IVW | 0.853 | $1.1\times10^1$ | 2 |
| T2DM (OR) | Total | 2.00 (1.57; 2.54) | < 0.001 | IVW | 0.308 | $6.6\times10^1$ | 2 |
| CRP (mg/L) | Total | 0.56 (0.16; 0.96) | 0.006 | IVW | 0.91 | $2.4\times10^1$ | 2 |
| SBP (mmHg) | Total | 3.30 (1.58; 5.03) | < 0.001 | IVW | 0.134 | 1.3 | 2 |

**Table S12:** *cis* Mendelian randomization effect estimates for one standard deviation change in Apo-A1 *(continued)*

| Outcome | Effect type | Point estimate (95%CI) | P-value | Estimator | Heterogeneity P-value | Precision | No. variants |
|---|---|---|---|---|---|---|---|
| DBP (mmHg) | Total | 15.45 (7.43; 23.47) | < 0.001 | MR Egger | NA | $6.0\times10^{-2}$ | 2 |
| PBL (OR) | Total | 0.42 (0.13; 1.37) | 0.149 | IVW | 0.306 | 2.8 | 2 |
| BUN (mg/dl) | Total | 0.04 (0.00; 0.08) | 0.026 | IVW | 0.469 | $3.1\times10^{3}$ | 2 |
| eGFR (SD of log(eGFR)) | Total | -0.00 (-0.02; 0.01) | 0.616 | IVW | 0.568 | $2.2\times10^{4}$ | 2 |
| CKD (OR) | Total | 0.90 (0.63; 1.28) | 0.559 | IVW | 0.703 | $3.0\times10^{1}$ | 2 |
| IBD (OR) | Total | 0.78 (0.42; 1.48) | 0.453 | IVW | 0.947 | 9.6 | 2 |
| UC (OR) | Total | 0.57 (0.26; 1.27) | 0.169 | IVW | 0.532 | 6.1 | 2 |
| CD (OR) | Total | 1.18 (0.45; 3.09) | 0.734 | IVW | 0.267 | 4.1 | 2 |
| Arthritis (OR) | Total | 0.25 (0.11; 0.54) | < 0.001 | IVW | 0.834 | 6.3 | 2 |
| ALS (OR) | Total | 1.02 (0.53; 1.99) | 0.943 | IVW | 0.384 | 8.7 | 2 |
| MS (OR) | Total | NA | NA | NA | NA | NA | NA |
| Alzheimer (OR) | Total | 1.11 (1.02; 1.20) | 0.011 | IVW | 0.428 | $6.3\times10^{2}$ | 2 |
| Parkinson (OR) | Total | 2.86 (1.50; 5.43) | < 0.001 | IVW | 0.596 | 9.3 | 2 |
| Pancreatic cancer (OR) | Total | 0.41 (0.05; 3.36) | 0.410 | IVW | 0.493 | $8.8\times10^{-1}$ | 2 |
| Colon cancer (OR) | Total | 6.45 (2.33; 17.87) | < 0.001 | IVW | 0.235 | 3.7 | 2 |
| Rectal cancer (OR) | Total | 1.51 (0.47; 4.86) | 0.494 | IVW | 0.78 | 2.8 | 2 |
| Melanoma (OR) | Total | 0.39 (0.20; 0.76) | 0.006 | IVW | 0.612 | 8.7 | 2 |
| Lung cancer (OR) | Total | 1.06 (0.36; 3.12) | 0.917 | IVW | 0.535 | 3.3 | 2 |

**Table S13:** Multivariable Mendelian randomization effect estimates for one standard deviation change in HDL-C, TG or Apo-B

| Exposure | Outcome | Effect type | Point estimate (95%CI) | P-value | Estimator | Heterogeneity P-value | Precision | No. variants |
|---|---|---|---|---|---|---|---|---|
| HDL-C | CHD (OR) | Conditional on Apo-B & TG | 0.80 (0.74; 0.86) | < 0.001 | IVW | < 0.001 | $6.9\times10^{2}$ | 53 |
| TG | CHD (OR) | Conditional on HDL-C & Apo-B | 0.88 (0.78; 1.01) | 0.065 | IVW | < 0.001 | $2.2\times10^{2}$ | 53 |
| Apo-B | CHD (OR) | Conditional on TG & HDL-C | 1.81 (1.64; 1.99) | < 0.001 | IVW | < 0.001 | $4.2\times10^{2}$ | 53 |
| HDL-C | HF (OR) | Conditional on Apo-B & TG | 1.00 (0.96; 1.05) | 0.860 | IVW | < 0.001 | $2.0\times10^{3}$ | 184 |
| TG | HF (OR) | Conditional on HDL-C & Apo-B | 1.08 (1.00; 1.17) | 0.065 | IVW | < 0.001 | $6.2\times10^{2}$ | 184 |
| Apo-B | HF (OR) | Conditional on TG & HDL-C | 1.01 (0.95; 1.08) | 0.642 | IVW | < 0.001 | $1.0\times10^{3}$ | 184 |
| HDL-C | AF (OR) | Conditional on Apo-B & TG | 1.00 (0.97; 1.04) | 0.809 | IVW | 0.008 | $2.7\times10^{3}$ | 200 |
| TG | AF (OR) | Conditional on HDL-C & Apo-B | 0.98 (0.92; 1.04) | 0.488 | IVW | 0.008 | $9.6\times10^{2}$ | 200 |
| Apo-B | AF (OR) | Conditional on TG & HDL-C | 1.05 (1.00; 1.10) | 0.068 | IVW | 0.008 | $1.5\times10^{3}$ | 200 |
| HDL-C | Any stroke (OR) | Conditional on Apo-B & TG | 0.98 (0.93; 1.02) | 0.323 | IVW | < 0.001 | $1.6\times10^{3}$ | 195 |
| TG | Any stroke (OR) | Conditional on HDL-C & Apo-B | 0.89 (0.81; 0.97) | 0.011 | IVW | < 0.001 | $4.7\times10^{2}$ | 195 |
| Apo-B | Any stroke (OR) | Conditional on TG & HDL-C | 1.13 (1.05; 1.22) | < 0.001 | IVW | < 0.001 | $7.5\times10^{2}$ | 195 |
| HDL-C | Any ischemic stroke (OR) | Conditional on Apo-B & TG | 0.98 (0.93; 1.04) | 0.526 | IVW | < 0.001 | $1.3\times10^{3}$ | 195 |
| TG | Any ischemic stroke (OR) | Conditional on HDL-C & Apo-B | 0.89 (0.80; 0.98) | 0.023 | IVW | < 0.001 | $3.8\times10^{2}$ | 195 |
| Apo-B | Any ischemic stroke (OR) | Conditional on TG & HDL-C | 1.15 (1.06; 1.24) | < 0.001 | IVW | < 0.001 | $6.0\times10^{2}$ | 195 |
| HDL-C | Large artery stroke (OR) | Conditional on Apo-B & TG | 1.02 (0.89; 1.17) | 0.774 | IVW | 0.002 | $2.1\times10^{2}$ | 198 |
| TG | Large artery stroke (OR) | Conditional on HDL-C & Apo-B | 0.96 (0.76; 1.22) | 0.744 | IVW | 0.002 | $6.8\times10^{1}$ | 198 |
| Apo-B | Large artery stroke (OR) | Conditional on TG & HDL-C | 1.08 (0.90; 1.31) | 0.401 | IVW | 0.002 | $1.1\times10^{2}$ | 198 |
| HDL-C | Cardioembolic stroke (OR) | Conditional on Apo-B & TG | 0.98 (0.88; 1.09) | 0.733 | IVW | 0.003 | $3.5\times10^{2}$ | 197 |
| TG | Cardioembolic stroke (OR) | Conditional on HDL-C & Apo-B | 0.93 (0.77; 1.12) | 0.433 | IVW | 0.003 | $1.1\times10^{2}$ | 197 |
| Apo-B | Cardioembolic stroke (OR) | Conditional on TG & HDL-C | 1.13 (0.97; 1.32) | 0.103 | IVW | 0.003 | $1.7\times10^{2}$ | 197 |
| HDL-C | Small vessel stroke (OR) | Conditional on Apo-B & TG | 0.97 (0.86; 1.10) | 0.645 | IVW | 0.025 | $2.5\times10^{2}$ | 195 |
| TG | Small vessel stroke (OR) | Conditional on HDL-C & Apo-B | 0.99 (0.79; 1.25) | 0.954 | IVW | 0.025 | $7.5\times10^{1}$ | 195 |
| Apo-B | Small vessel stroke (OR) | Conditional on TG & HDL-C | 0.98 (0.82; 1.17) | 0.804 | IVW | 0.025 | $1.2\times10^{2}$ | 195 |
| HDL-C | cIMT (mm) | Conditional on Apo-B & TG | -0.00 (-0.01; 0.00) | 0.143 | IVW | < 0.001 | $2.6\times10^{5}$ | 199 |
| TG | cIMT (mm) | Conditional on HDL-C & Apo-B | -0.02 (-0.03; -0.01) | < 0.001 | IVW | < 0.001 | $8.7\times10^{4}$ | 199 |
| Apo-B | cIMT (mm) | Conditional on TG & HDL-C | 0.03 (0.02; 0.03) | < 0.001 | IVW | < 0.001 | $1.4\times10^{5}$ | 199 |
| HDL-C | Carotid plaque (OR) | Conditional on Apo-B & TG | 0.91 (0.83; 1.00) | 0.060 | IVW | 0.017 | $4.2\times10^{2}$ | 187 |
| TG | Carotid plaque (OR) | Conditional on HDL-C & Apo-B | 0.74 (0.63; 0.88) | < 0.001 | IVW | 0.017 | $1.4\times10^{2}$ | 187 |
| Apo-B | Carotid plaque (OR) | Conditional on TG & HDL-C | 1.60 (1.40; 1.82) | < 0.001 | IVW | 0.017 | $2.2\times10^{2}$ | 187 |
| HDL-C | Glucose (mmol/l) | Conditional on Apo-B & TG | -0.00 (-0.02; 0.01) | 0.616 | IVW | < 0.001 | $1.4\times10^{4}$ | 187 |

**Table S13:** Multivariable Mendelian randomization effect estimates for one standa
*(continued)*

| Exposure | Outcome | Effect type | Point estimate (95%CI) | P-value | Estimator | Heterogeneity P-value | Precision | No. variants |
|---|---|---|---|---|---|---|---|---|
| TG | Glucose (mmol/l) | Conditional on HDL-C & Apo-B | -0.01 (-0.04; 0.01) | 0.338 | IVW | < 0.001 | $4.5\times10^3$ | 187 |
| Apo-B | Glucose (mmol/l) | Conditional on TG & HDL-C | -0.01 (-0.03; 0.02) | 0.625 | IVW | < 0.001 | $7.7\times10^3$ | 187 |
| HDL-C | HbA1c (mmol/mol) | Conditional on Apo-B & TG | -0.05 (-0.14; 0.03) | 0.213 | IVW | < 0.001 | $5.2\times10^2$ | 181 |
| TG | HbA1c (mmol/mol) | Conditional on HDL-C & Apo-B | -0.09 (-0.24; 0.07) | 0.262 | IVW | < 0.001 | $1.7\times10^2$ | 181 |
| Apo-B | HbA1c (mmol/mol) | Conditional on TG & HDL-C | -0.10 (-0.21; 0.02) | 0.097 | IVW | < 0.001 | $2.9\times10^2$ | 181 |
| HDL-C | T2DM (OR) | Conditional on Apo-B & TG | 0.98 (0.95; 1.02) | 0.338 | IVW | < 0.001 | $3.3\times10^3$ | 169 |
| TG | T2DM (OR) | Conditional on HDL-C & Apo-B | 1.15 (1.07; 1.23) | < 0.001 | IVW | < 0.001 | $8.8\times10^2$ | 169 |
| Apo-B | T2DM (OR) | Conditional on TG & HDL-C | 0.88 (0.83; 0.92) | < 0.001 | IVW | < 0.001 | $1.4\times10^3$ | 169 |
| HDL-C | CRP (mg/L) | Conditional on Apo-B & TG | 0.17 (0.11; 0.23) | < 0.001 | IVW | < 0.001 | $1.1\times10^3$ | 174 |
| TG | CRP (mg/L) | Conditional on HDL-C & Apo-B | 0.31 (0.20; 0.42) | < 0.001 | IVW | < 0.001 | $3.1\times10^2$ | 174 |
| Apo-B | CRP (mg/L) | Conditional on TG & HDL-C | -0.14 (-0.23; -0.05) | 0.002 | IVW | < 0.001 | $5.0\times10^2$ | 174 |
| HDL-C | SBP (mmHg) | Conditional on Apo-B & TG | -0.27 (-0.43; -0.10) | < 0.001 | IVW | < 0.001 | $1.4\times10^2$ | 164 |
| TG | SBP (mmHg) | Conditional on HDL-C & Apo-B | 0.25 (-0.06; 0.56) | 0.108 | IVW | < 0.001 | $4.1\times10^1$ | 164 |
| Apo-B | SBP (mmHg) | Conditional on TG & HDL-C | 0.32 (0.08; 0.56) | 0.010 | IVW | < 0.001 | $6.5\times10^1$ | 164 |
| HDL-C | DBP (mmHg) | Conditional on Apo-B & TG | -0.13 (-0.23; -0.04) | 0.006 | IVW | < 0.001 | $4.3\times10^2$ | 160 |
| TG | DBP (mmHg) | Conditional on HDL-C & Apo-B | 0.36 (0.17; 0.54) | < 0.001 | IVW | < 0.001 | $1.1\times10^2$ | 160 |
| Apo-B | DBP (mmHg) | Conditional on TG & HDL-C | -0.23 (-0.39; -0.07) | 0.004 | MR Egger | < 0.001 | $1.5\times10^2$ | 161 |
| HDL-C | PBL (OR) | Conditional on Apo-B & TG | 1.36 (1.06; 1.75) | 0.016 | IVW | 0.003 | $6.2\times10^1$ | 97 |
| TG | PBL (OR) | Conditional on HDL-C & Apo-B | 1.53 (1.02; 2.30) | 0.041 | IVW | 0.003 | $2.3\times10^1$ | 97 |
| Apo-B | PBL (OR) | Conditional on TG & HDL-C | 0.85 (0.59; 1.22) | 0.382 | IVW | 0.003 | $2.9\times10^1$ | 97 |
| HDL-C | BUN (mg/dl) | Conditional on Apo-B & TG | -0.00 (-0.01; -0.00) | 0.032 | IVW | < 0.001 | $1.9\times10^5$ | 190 |
| TG | BUN (mg/dl) | Conditional on HDL-C & Apo-B | -0.01 (-0.01; 0.00) | 0.174 | IVW | < 0.001 | $5.0\times10^4$ | 190 |
| Apo-B | BUN (mg/dl) | Conditional on TG & HDL-C | 0.00 (-0.00; 0.01) | 0.261 | IVW | < 0.001 | $8.0\times10^4$ | 190 |
| HDL-C | eGFR (SD of log(eGFR)) | Conditional on Apo-B & TG | -0.00 (-0.00; 0.00) | 0.269 | IVW | < 0.001 | $8.6\times10^5$ | 180 |
| TG | eGFR (SD of log(eGFR)) | Conditional on HDL-C & Apo-B | 0.00 (-0.00; 0.00) | 0.883 | IVW | < 0.001 | $2.9\times10^5$ | 180 |
| Apo-B | eGFR (SD of log(eGFR)) | Conditional on TG & HDL-C | 0.00 (-0.00; 0.00) | 0.596 | IVW | < 0.001 | $5.1\times10^5$ | 180 |
| HDL-C | CKD (OR) | Conditional on Apo-B & TG | 0.99 (0.94; 1.04) | 0.706 | IVW | < 0.001 | $1.4\times10^3$ | 192 |
| TG | CKD (OR) | Conditional on HDL-C & Apo-B | 1.02 (0.93; 1.11) | 0.749 | IVW | < 0.001 | $4.5\times10^2$ | 192 |
| Apo-B | CKD (OR) | Conditional on TG & HDL-C | 0.96 (0.89; 1.03) | 0.272 | IVW | < 0.001 | $7.4\times10^2$ | 192 |
| HDL-C | IBD (OR) | Conditional on Apo-B & TG | 0.98 (0.89; 1.08) | 0.647 | IVW | < 0.001 | $4.3\times10^2$ | 194 |
| TG | IBD (OR) | Conditional on HDL-C & Apo-B | 0.87 (0.74; 1.02) | 0.081 | IVW | < 0.001 | $1.5\times10^2$ | 194 |

**Table S13:** Multivariable Mendelian randomization effect estimates for one standa
*(continued)*

| Exposure | Outcome | Effect type | Point estimate (95%CI) | P-value | Estimator | Heterogeneity P-value | Precision | No. variants |
|---|---|---|---|---|---|---|---|---|
| Apo-B | IBD (OR) | Conditional on TG & HDL-C | 1.17 (1.03; 1.33) | 0.016 | IVW | < 0.001 | $2.3\times10^2$ | 194 |
| HDL-C | UC (OR) | Conditional on Apo-B & TG | 0.97 (0.86; 1.09) | 0.609 | IVW | 0.025 | $2.8\times10^2$ | 198 |
| TG | UC (OR) | Conditional on HDL-C & Apo-B | 0.80 (0.65; 0.98) | 0.031 | IVW | 0.025 | $9.2\times10^1$ | 198 |
| Apo-B | UC (OR) | Conditional on TG & HDL-C | 1.12 (0.96; 1.32) | 0.152 | IVW | 0.025 | $1.5\times10^2$ | 198 |
| HDL-C | CD (OR) | Conditional on Apo-B & TG | 1.04 (0.91; 1.19) | 0.556 | IVW | 0.014 | $2.2\times10^2$ | 194 |
| TG | CD (OR) | Conditional on HDL-C & Apo-B | 1.18 (0.94; 1.47) | 0.153 | IVW | 0.014 | $7.7\times10^1$ | 194 |
| Apo-B | CD (OR) | Conditional on TG & HDL-C | 1.05 (0.88; 1.25) | 0.614 | IVW | 0.014 | $1.3\times10^2$ | 194 |
| HDL-C | Arthritis (OR) | Conditional on Apo-B & TG | 1.10 (0.98; 1.24) | 0.112 | IVW | 0.321 | $2.7\times10^2$ | 156 |
| TG | Arthritis (OR) | Conditional on HDL-C & Apo-B | 1.16 (0.95; 1.43) | 0.150 | MR Egger | 0.465 | $9.2\times10^1$ | 155 |
| Apo-B | Arthritis (OR) | Conditional on TG & HDL-C | 0.96 (0.81; 1.15) | 0.677 | MR Egger | 0.471 | $1.2\times10^2$ | 156 |
| HDL-C | ALS (OR) | Conditional on Apo-B & TG | 1.09 (0.96; 1.23) | 0.181 | MR Egger | 0.066 | $2.6\times10^2$ | 198 |
| TG | ALS (OR) | Conditional on HDL-C & Apo-B | 1.09 (0.92; 1.28) | 0.326 | IVW | 0.021 | $1.4\times10^2$ | 194 |
| Apo-B | ALS (OR) | Conditional on TG & HDL-C | 1.02 (0.90; 1.15) | 0.768 | IVW | 0.021 | $2.6\times10^2$ | 194 |
| HDL-C | MS (OR) | Conditional on Apo-B & TG | 0.93 (0.74; 1.16) | 0.507 | MR Egger | 0.012 | $7.3\times10^1$ | 38 |
| TG | MS (OR) | Conditional on HDL-C & Apo-B | 0.82 (0.64; 1.04) | 0.100 | IVW | 0.004 | $6.6\times10^1$ | 38 |
| Apo-B | MS (OR) | Conditional on TG & HDL-C | 1.20 (0.99; 1.46) | 0.062 | IVW | 0.004 | $1.0\times10^2$ | 38 |
| HDL-C | Alzheimer (OR) | Conditional on Apo-B & TG | 0.99 (0.97; 1.01) | 0.178 | MR Egger | < 0.001 | $1.3\times10^4$ | 184 |
| TG | Alzheimer (OR) | Conditional on HDL-C & Apo-B | 0.96 (0.94; 0.98) | < 0.001 | IVW | < 0.001 | $6.9\times10^3$ | 181 |
| Apo-B | Alzheimer (OR) | Conditional on TG & HDL-C | 1.05 (1.03; 1.07) | < 0.001 | IVW | < 0.001 | $1.0\times10^4$ | 181 |
| HDL-C | Parkinson (OR) | Conditional on Apo-B & TG | 0.95 (0.86; 1.06) | 0.367 | IVW | < 0.001 | $3.6\times10^2$ | 194 |
| TG | Parkinson (OR) | Conditional on HDL-C & Apo-B | 1.04 (0.86; 1.26) | 0.667 | IVW | < 0.001 | $1.1\times10^2$ | 194 |
| Apo-B | Parkinson (OR) | Conditional on TG & HDL-C | 0.90 (0.77; 1.04) | 0.147 | IVW | < 0.001 | $1.7\times10^2$ | 194 |
| HDL-C | Pancreatic cancer (OR) | Conditional on Apo-B & TG | 1.11 (0.81; 1.51) | 0.506 | IVW | 0.022 | $4.0\times10^1$ | 196 |
| TG | Pancreatic cancer (OR) | Conditional on HDL-C & Apo-B | 0.68 (0.40; 1.15) | 0.149 | IVW | 0.022 | $1.4\times10^1$ | 196 |
| Apo-B | Pancreatic cancer (OR) | Conditional on TG & HDL-C | 1.19 (0.79; 1.79) | 0.405 | IVW | 0.022 | $2.3\times10^1$ | 196 |
| HDL-C | Colon cancer (OR) | Conditional on Apo-B & TG | 1.13 (0.99; 1.28) | 0.060 | IVW | 0.011 | $2.4\times10^2$ | 198 |
| TG | Colon cancer (OR) | Conditional on HDL-C & Apo-B | 1.06 (0.86; 1.32) | 0.575 | IVW | 0.011 | $8.3\times10^1$ | 198 |
| Apo-B | Colon cancer (OR) | Conditional on TG & HDL-C | 1.11 (0.94; 1.31) | 0.227 | IVW | 0.011 | $1.4\times10^2$ | 198 |
| HDL-C | Rectal cancer (OR) | Conditional on Apo-B & TG | 0.85 (0.71; 1.02) | 0.089 | IVW | 0.213 | $1.2\times10^2$ | 196 |
| TG | Rectal cancer (OR) | Conditional on HDL-C & Apo-B | 0.74 (0.55; 1.00) | 0.051 | IVW | 0.213 | $4.2\times10^1$ | 196 |
| Apo-B | Rectal cancer (OR) | Conditional on TG & HDL-C | 1.31 (1.04; 1.66) | 0.024 | IVW | 0.213 | $6.9\times10^1$ | 196 |

**Table S13:** Multivariable Mendelian randomization effect estimates for one standa
*(continued)*

| Exposure | Outcome | Effect type | Point estimate (95%CI) | P-value | Estimator | Heterogeneity P-value | Precision | No. variants |
|---|---|---|---|---|---|---|---|---|
| HDL-C | Melanoma (OR) | Conditional on Apo-B & TG | 1.03 (0.93; 1.14) | 0.554 | IVW | < 0.001 | $4.0\times10^{2}$ | 195 |
| TG | Melanoma (OR) | Conditional on HDL-C & Apo-B | 1.15 (0.98; 1.35) | 0.091 | IVW | < 0.001 | $1.4\times10^{2}$ | 195 |
| Apo-B | Melanoma (OR) | Conditional on TG & HDL-C | 0.98 (0.87; 1.11) | 0.761 | IVW | < 0.001 | $2.5\times10^{2}$ | 195 |
| HDL-C | Lung cancer (OR) | Conditional on Apo-B & TG | 0.89 (0.76; 1.05) | 0.161 | IVW | 0.017 | $1.6\times10^{2}$ | 199 |
| TG | Lung cancer (OR) | Conditional on HDL-C & Apo-B | 0.94 (0.72; 1.22) | 0.630 | IVW | 0.017 | $5.7\times10^{1}$ | 199 |
| Apo-B | Lung cancer (OR) | Conditional on TG & HDL-C | 0.91 (0.75; 1.11) | 0.366 | IVW | 0.017 | $9.6\times10^{1}$ | 199 |

**Table S14:** Genetic variants used in the presented analyses. Note that due to coverage of the GWAS chip the instruments differed across considered outcomes

| Exposure | Chromosome | Base-pair position (Build 37) | Other allele | Effect allele |
|---|---|---|---|---|
| LDL-C | 11 | 116651115 | T | C |
| LDL-C | 11 | 116657561 | T | C |
| LDL-C | 11 | 61565908 | T | C |
| LDL-C | 11 | 61571382 | G | A |
| LDL-C | 11 | 61621194 | G | A |
| LDL-C | 12 | 111844218 | G | A |
| LDL-C | 12 | 111949394 | T | C |
| LDL-C | 12 | 111976776 | T | C |
| LDL-C | 12 | 111993886 | G | A |
| LDL-C | 14 | 94844947 | T | C |
| LDL-C | 14 | 94877868 | T | C |
| LDL-C | 16 | 72052348 | G | A |
| LDL-C | 16 | 72078989 | TG | T |
| LDL-C | 16 | 72108093 | G | A |
| LDL-C | 17 | 45422628 | G | A |
| LDL-C | 17 | 45485982 | T | C |
| LDL-C | 17 | 45498937 | T | G |
| LDL-C | 17 | 45537395 | T | C |
| LDL-C | 17 | 45540743 | T | C |
| LDL-C | 17 | 45735706 | T | C |
| LDL-C | 19 | 10504616 | C | A |
| LDL-C | 19 | 10516198 | T | G |
| LDL-C | 19 | 10633984 | G | A |
| LDL-C | 19 | 10634928 | T | C |
| LDL-C | 19 | 10639533 | T | C |
| LDL-C | 19 | 10643352 | T | G |
| LDL-C | 19 | 10643471 | G | A |
| LDL-C | 19 | 10645495 | G | A |
| LDL-C | 19 | 10646814 | G | A |
| LDL-C | 19 | 10689256 | T | G |
| LDL-C | 19 | 10703280 | G | A |
| LDL-C | 19 | 10734951 | T | C |
| LDL-C | 19 | 10739143 | C | A |
| LDL-C | 19 | 10741907 | G | A |
| LDL-C | 19 | 10742170 | G | A |
| LDL-C | 19 | 10742287 | T | C |
| LDL-C | 19 | 10784858 | G | A |
| LDL-C | 19 | 10798974 | T | C |
| LDL-C | 19 | 10802792 | G | A |
| LDL-C | 19 | 10813364 | T | C |
| LDL-C | 19 | 10901440 | G | A |
| LDL-C | 19 | 10903972 | G | A |
| LDL-C | 19 | 10904700 | T | C |

**Table S14:** Genetic variants used in the presented analyses. Note that due to coverage of the GWAS chip the instruments differed across considered outcomes *(continued)*

| Exposure | Chromosome | Base-pair position (Build 37) | Other allele | Effect allele |
|---|---|---|---|---|
| LDL-C | 19 | 10909953 | T | C |
| LDL-C | 19 | 10916684 | T | C |
| LDL-C | 19 | 10926952 | T | C |
| LDL-C | 19 | 10947522 | T | C |
| LDL-C | 19 | 10948031 | G | A |
| LDL-C | 19 | 10952958 | G | A |
| LDL-C | 19 | 10965270 | T | C |
| LDL-C | 19 | 10967970 | G | A |
| LDL-C | 19 | 10981992 | T | C |
| LDL-C | 19 | 11027626 | C | A |
| LDL-C | 19 | 11054715 | G | A |
| LDL-C | 19 | 11055935 | G | A |
| LDL-C | 19 | 11059389 | G | A |
| LDL-C | 19 | 11065409 | C | A |
| LDL-C | 19 | 11065687 | G | A |
| LDL-C | 19 | 11104747 | G | A |
| LDL-C | 19 | 11105608 | T | C |
| LDL-C | 19 | 11113902 | G | A |
| LDL-C | 19 | 11126160 | T | C |
| LDL-C | 19 | 11128824 | T | C |
| LDL-C | 19 | 11132080 | G | A |
| LDL-C | 19 | 11136315 | T | C |
| LDL-C | 19 | 11139636 | T | G |
| LDL-C | 19 | 11140144 | G | A |
| LDL-C | 19 | 11146499 | T | C |
| LDL-C | 19 | 11159076 | G | A |
| LDL-C | 19 | 11159096 | T | G |
| LDL-C | 19 | 11159525 | T | G |
| LDL-C | 19 | 11163562 | T | C |
| LDL-C | 19 | 11163601 | T | G |
| LDL-C | 19 | 11163897 | T | C |
| LDL-C | 19 | 11164781 | G | A |
| LDL-C | 19 | 11166431 | CT | C |
| LDL-C | 19 | 11169947 | T | C |
| LDL-C | 19 | 11170839 | T | C |
| LDL-C | 19 | 11171281 | C | A |
| LDL-C | 19 | 11174229 | CT | C |
| LDL-C | 19 | 11174625 | G | A |
| LDL-C | 19 | 11174742 | T | C |
| LDL-C | 19 | 11177250 | C | A |
| LDL-C | 19 | 11180047 | T | C |
| LDL-C | 19 | 11183837 | G | A |
| LDL-C | 19 | 11184250 | G | A |
| LDL-C | 19 | 11184293 | T | C |

**Table S14:** Genetic variants used in the presented analyses. Note that due to coverage of the GWAS chip the instruments differed across considered outcomes *(continued)*

| Exposure | Chromosome | Base-pair position (Build 37) | Other allele | Effect allele |
|---|---|---|---|---|
| LDL-C | 19 | 11185014 | G | A |
| LDL-C | 19 | 11185919 | T | C |
| LDL-C | 19 | 11186711 | T | C |
| LDL-C | 19 | 11187611 | G | A |
| LDL-C | 19 | 11188313 | CTTTA | C |
| LDL-C | 19 | 11188899 | T | C |
| LDL-C | 19 | 11189298 | CG | C |
| LDL-C | 19 | 11189764 | T | C |
| LDL-C | 19 | 11190544 | T | C |
| LDL-C | 19 | 11193065 | T | C |
| LDL-C | 19 | 11195030 | G | A |
| LDL-C | 19 | 11202306 | T | G |
| LDL-C | 19 | 11206575 | G | A |
| LDL-C | 19 | 11207102 | T | C |
| LDL-C | 19 | 11207516 | G | A |
| LDL-C | 19 | 11207982 | T | C |
| LDL-C | 19 | 11209757 | CCAAA | C |
| LDL-C | 19 | 11210912 | T | C |
| LDL-C | 19 | 11216617 | G | A |
| LDL-C | 19 | 11221180 | G | A |
| LDL-C | 19 | 11224181 | T | C |
| LDL-C | 19 | 11224265 | G | A |
| LDL-C | 19 | 11227480 | C | A |
| LDL-C | 19 | 11228965 | C | A |
| LDL-C | 19 | 11230881 | T | C |
| LDL-C | 19 | 11236180 | C | A |
| LDL-C | 19 | 11243445 | G | A |
| LDL-C | 19 | 11245071 | GGAGT | G |
| LDL-C | 19 | 11248104 | G | A |
| LDL-C | 19 | 11250139 | G | A |
| LDL-C | 19 | 11257169 | T | C |
| LDL-C | 19 | 11262477 | T | G |
| LDL-C | 19 | 11262969 | T | G |
| LDL-C | 19 | 11263098 | T | C |
| LDL-C | 19 | 11263968 | CA | C |
| LDL-C | 19 | 11266693 | T | G |
| LDL-C | 19 | 11269893 | T | C |
| LDL-C | 19 | 11271714 | G | A |
| LDL-C | 19 | 11274679 | G | A |
| LDL-C | 19 | 11275139 | C | A |
| LDL-C | 19 | 11275258 | G | A |
| LDL-C | 19 | 11276256 | T | C |
| LDL-C | 19 | 19130096 | T | C |
| LDL-C | 19 | 19380513 | G | A |

**Table S14:** Genetic variants used in the presented analyses. Note that due to coverage of the GWAS chip the instruments differed across considered outcomes *(continued)*

| Exposure | Chromosome | Base-pair position (Build 37) | Other allele | Effect allele |
|---|---|---|---|---|
| LDL-C | 19 | 19458388 | G | A |
| LDL-C | 19 | 20217169 | T | C |
| LDL-C | 19 | 41087144 | T | C |
| LDL-C | 19 | 45041317 | T | C |
| LDL-C | 19 | 45121682 | G | A |
| LDL-C | 19 | 45134682 | C | A |
| LDL-C | 19 | 45135841 | T | C |
| LDL-C | 19 | 45138246 | T | C |
| LDL-C | 19 | 45147396 | T | C |
| LDL-C | 19 | 45150614 | G | A |
| LDL-C | 19 | 45156201 | TA | T |
| LDL-C | 19 | 45158411 | T | C |
| LDL-C | 19 | 45159023 | T | C |
| LDL-C | 19 | 45160086 | C | A |
| LDL-C | 19 | 45163671 | T | C |
| LDL-C | 19 | 45167544 | T | C |
| LDL-C | 19 | 45170097 | G | A |
| LDL-C | 19 | 45173951 | C | A |
| LDL-C | 19 | 45176340 | T | C |
| LDL-C | 19 | 45187163 | G | A |
| LDL-C | 19 | 45188471 | T | C |
| LDL-C | 19 | 45189000 | G | A |
| LDL-C | 19 | 45195313 | T | G |
| LDL-C | 19 | 45195744 | CG | C |
| LDL-C | 19 | 45196254 | G | A |
| LDL-C | 19 | 45196581 | G | A |
| LDL-C | 19 | 45196964 | G | A |
| LDL-C | 19 | 45197027 | C | A |
| LDL-C | 19 | 45198936 | T | C |
| LDL-C | 19 | 45220896 | T | C |
| LDL-C | 19 | 45220941 | C | A |
| LDL-C | 19 | 45222982 | T | G |
| LDL-C | 19 | 45223038 | G | A |
| LDL-C | 19 | 45224305 | C | A |
| LDL-C | 19 | 45225345 | T | C |
| LDL-C | 19 | 45232161 | G | A |
| LDL-C | 19 | 45232205 | G | A |
| LDL-C | 19 | 45232374 | G | A |
| LDL-C | 19 | 45233329 | AAATTTATAG | A |
| LDL-C | 19 | 45237812 | T | C |
| LDL-C | 19 | 45237919 | T | C |
| LDL-C | 19 | 45237984 | T | C |
| LDL-C | 19 | 45239185 | G | A |
| LDL-C | 19 | 45239536 | C | A |

**Table S14:** Genetic variants used in the presented analyses. Note that due to coverage of the GWAS chip the instruments differed across considered outcomes *(continued)*

| Exposure | Chromosome | Base-pair position (Build 37) | Other allele | Effect allele |
|---|---|---|---|---|
| LDL-C | 19 | 45241248 | CA | C |
| LDL-C | 19 | 45242107 | G | A |
| LDL-C | 19 | 45242173 | G | A |
| LDL-C | 19 | 45242740 | G | A |
| LDL-C | 19 | 45245884 | G | A |
| LDL-C | 19 | 45247627 | G | A |
| LDL-C | 19 | 45249661 | T | C |
| LDL-C | 19 | 45251156 | C | A |
| LDL-C | 19 | 45253491 | T | C |
| LDL-C | 19 | 45253542 | CTTTG | C |
| LDL-C | 19 | 45253611 | ATTGGC | A |
| LDL-C | 19 | 45276590 | GTTA | G |
| LDL-C | 19 | 45296364 | G | A |
| LDL-C | 19 | 45296806 | T | C |
| LDL-C | 19 | 45301942 | G | A |
| LDL-C | 19 | 45302504 | G | A |
| LDL-C | 19 | 45302778 | G | A |
| LDL-C | 19 | 45302949 | GAA | G |
| LDL-C | 19 | 45303122 | AAAAG | A |
| LDL-C | 19 | 45304061 | G | A |
| LDL-C | 19 | 45305345 | GC | G |
| LDL-C | 19 | 45309431 | T | C |
| LDL-C | 19 | 45313715 | T | C |
| LDL-C | 19 | 45315445 | G | A |
| LDL-C | 19 | 45319631 | G | A |
| LDL-C | 19 | 45323170 | CCATCCCCAACTCATCCT | C |
| LDL-C | 19 | 45324138 | G | A |
| LDL-C | 19 | 45325738 | T | G |
| LDL-C | 19 | 45326664 | T | C |
| LDL-C | 19 | 45327459 | T | C |
| LDL-C | 19 | 45330999 | G | A |
| LDL-C | 19 | 45331103 | G | A |
| LDL-C | 19 | 45333834 | T | C |
| LDL-C | 19 | 45335676 | G | A |
| LDL-C | 19 | 45336035 | T | C |
| LDL-C | 19 | 45337918 | G | A |
| LDL-C | 19 | 45338250 | AT | A |
| LDL-C | 19 | 45342114 | G | A |
| LDL-C | 19 | 45347911 | C | A |
| LDL-C | 19 | 45351891 | G | A |
| LDL-C | 19 | 45356141 | T | C |
| LDL-C | 19 | 45356752 | CT | C |
| LDL-C | 19 | 45357939 | G | A |
| LDL-C | 19 | 45359706 | G | A |

**Table S14:** Genetic variants used in the presented analyses. Note that due to coverage of the GWAS chip the instruments differed across considered outcomes *(continued)*

| Exposure | Chromosome | Base-pair position (Build 37) | Other allele | Effect allele |
|---|---|---|---|---|
| LDL-C | 19 | 45361646 | G | A |
| LDL-C | 19 | 45365447 | G | A |
| LDL-C | 19 | 45369203 | G | A |
| LDL-C | 19 | 45373565 | G | A |
| LDL-C | 19 | 45377334 | G | A |
| LDL-C | 19 | 45379060 | C | A |
| LDL-C | 19 | 45382034 | G | A |
| LDL-C | 19 | 45382675 | G | A |
| LDL-C | 19 | 45383061 | T | G |
| LDL-C | 19 | 45384105 | T | C |
| LDL-C | 19 | 45384405 | T | C |
| LDL-C | 19 | 45385356 | C | A |
| LDL-C | 19 | 45386467 | GTAA | G |
| LDL-C | 19 | 45386634 | T | C |
| LDL-C | 19 | 45387034 | C | A |
| LDL-C | 19 | 45388130 | G | A |
| LDL-C | 19 | 45388241 | T | G |
| LDL-C | 19 | 45388500 | G | A |
| LDL-C | 19 | 45389596 | G | A |
| LDL-C | 19 | 45392254 | T | C |
| LDL-C | 19 | 45393516 | T | C |
| LDL-C | 19 | 45395266 | G | A |
| LDL-C | 19 | 45395330 | G | A |
| LDL-C | 19 | 45395619 | G | A |
| LDL-C | 19 | 45395714 | T | C |
| LDL-C | 19 | 45395844 | G | A |
| LDL-C | 19 | 45396219 | T | C |
| LDL-C | 19 | 45396973 | G | A |
| LDL-C | 19 | 45397952 | T | C |
| LDL-C | 19 | 45400484 | TG | T |
| LDL-C | 19 | 45401868 | T | C |
| LDL-C | 19 | 45402262 | T | G |
| LDL-C | 19 | 45406673 | G | A |
| LDL-C | 19 | 45408628 | T | C |
| LDL-C | 19 | 45408836 | T | G |
| LDL-C | 19 | 45410002 | G | A |
| LDL-C | 19 | 45411941 | T | C |
| LDL-C | 19 | 45412079 | T | C |
| LDL-C | 19 | 45413576 | T | C |
| LDL-C | 19 | 45415640 | G | A |
| LDL-C | 19 | 45416291 | T | C |
| LDL-C | 19 | 45416831 | T | G |
| LDL-C | 19 | 45418486 | G | A |
| LDL-C | 19 | 45421254 | G | A |

**Table S14:** Genetic variants used in the presented analyses. Note that due to coverage of the GWAS chip the instruments differed across considered outcomes *(continued)*

| Exposure | Chromosome | Base-pair position (Build 37) | Other allele | Effect allele |
|---|---|---|---|---|
| LDL-C | 19 | 45421877 | G | A |
| LDL-C | 19 | 45422946 | G | A |
| LDL-C | 19 | 45423944 | T | C |
| LDL-C | 19 | 45425178 | G | A |
| LDL-C | 19 | 45425460 | G | A |
| LDL-C | 19 | 45426792 | G | A |
| LDL-C | 19 | 45428234 | G | A |
| LDL-C | 19 | 45429543 | T | C |
| LDL-C | 19 | 45431897 | G | A |
| LDL-C | 19 | 45435466 | CT | C |
| LDL-C | 19 | 45436216 | G | A |
| LDL-C | 19 | 45436657 | G | A |
| LDL-C | 19 | 45440145 | CTG | C |
| LDL-C | 19 | 45441608 | T | C |
| LDL-C | 19 | 45447178 | T | C |
| LDL-C | 19 | 45447221 | G | A |
| LDL-C | 19 | 45450252 | T | C |
| LDL-C | 19 | 45450328 | G | A |
| LDL-C | 19 | 45451656 | T | G |
| LDL-C | 19 | 45452335 | T | C |
| LDL-C | 19 | 45453763 | G | A |
| LDL-C | 19 | 45455588 | T | C |
| LDL-C | 19 | 45460549 | TTGAC | T |
| LDL-C | 19 | 45464963 | T | C |
| LDL-C | 19 | 45475700 | T | G |
| LDL-C | 19 | 45477111 | T | C |
| LDL-C | 19 | 45478352 | G | A |
| LDL-C | 19 | 45491032 | T | C |
| LDL-C | 19 | 45496303 | T | C |
| LDL-C | 19 | 45514061 | G | A |
| LDL-C | 19 | 45515014 | CTGTGTGTG | C |
| LDL-C | 19 | 45531618 | GT | G |
| LDL-C | 19 | 45541782 | G | A |
| LDL-C | 19 | 45547583 | G | A |
| LDL-C | 19 | 45548255 | G | A |
| LDL-C | 19 | 45552587 | T | C |
| LDL-C | 19 | 45653883 | TAGTC | T |
| LDL-C | 19 | 45672478 | G | A |
| LDL-C | 19 | 45693752 | T | C |
| LDL-C | 19 | 45706865 | G | A |
| LDL-C | 19 | 45723379 | T | C |
| LDL-C | 19 | 45880690 | T | C |
| LDL-C | 19 | 45915038 | T | C |
| LDL-C | 19 | 45987013 | T | C |

**Table S14:** Genetic variants used in the presented analyses. Note that due to coverage of the GWAS chip the instruments differed across considered outcomes *(continued)*

| Exposure | Chromosome | Base-pair position (Build 37) | Other allele | Effect allele |
|---|---|---|---|---|
| LDL-C | 19 | 46018119 | G | A |
| LDL-C | 19 | 46420463 | AAAAAT | A |
| LDL-C | 1 | 109715950 | G | A |
| LDL-C | 1 | 109757295 | G | A |
| LDL-C | 1 | 109776285 | T | C |
| LDL-C | 1 | 109792735 | ACGC | A |
| LDL-C | 1 | 109797763 | T | C |
| LDL-C | 1 | 109798721 | G | A |
| LDL-C | 1 | 109804646 | G | A |
| LDL-C | 1 | 109806442 | C | A |
| LDL-C | 1 | 109810200 | G | A |
| LDL-C | 1 | 109813485 | T | C |
| LDL-C | 1 | 109814880 | T | C |
| LDL-C | 1 | 109815214 | TG | T |
| LDL-C | 1 | 109815252 | G | A |
| LDL-C | 1 | 109818158 | CT | C |
| LDL-C | 1 | 109819615 | T | C |
| LDL-C | 1 | 109819981 | T | C |
| LDL-C | 1 | 109820884 | G | A |
| LDL-C | 1 | 109820919 | T | G |
| LDL-C | 1 | 109822166 | G | A |
| LDL-C | 1 | 109824250 | G | A |
| LDL-C | 1 | 109826760 | TTCCTAACC | T |
| LDL-C | 1 | 109827253 | G | A |
| LDL-C | 1 | 109834938 | T | C |
| LDL-C | 1 | 109839400 | T | C |
| LDL-C | 1 | 109841760 | G | A |
| LDL-C | 1 | 109955569 | T | C |
| LDL-C | 1 | 110019439 | G | A |
| LDL-C | 1 | 110033224 | G | A |
| LDL-C | 1 | 27284913 | T | C |
| LDL-C | 1 | 54598299 | G | A |
| LDL-C | 1 | 54703630 | T | C |
| LDL-C | 1 | 54703702 | C | A |
| LDL-C | 1 | 54708492 | G | A |
| LDL-C | 1 | 54714821 | G | A |
| LDL-C | 1 | 54715751 | T | C |
| LDL-C | 1 | 54717654 | T | C |
| LDL-C | 1 | 54721862 | TC | T |
| LDL-C | 1 | 54722625 | T | C |
| LDL-C | 1 | 54724389 | G | A |
| LDL-C | 1 | 54726703 | G | A |
| LDL-C | 1 | 54728029 | T | C |
| LDL-C | 1 | 54769277 | T | G |

**Table S14:** Genetic variants used in the presented analyses. Note that due to coverage of the GWAS chip the instruments differed across considered outcomes *(continued)*

| Exposure | Chromosome | Base-pair position (Build 37) | Other allele | Effect allele |
|---|---|---|---|---|
| LDL-C | 1 | 54772869 | C | A |
| LDL-C | 1 | 54783595 | T | C |
| LDL-C | 1 | 54834840 | T | C |
| LDL-C | 1 | 54849397 | T | C |
| LDL-C | 1 | 54880705 | T | C |
| LDL-C | 1 | 54889174 | T | C |
| LDL-C | 1 | 54935900 | G | A |
| LDL-C | 1 | 54983266 | T | C |
| LDL-C | 1 | 55083647 | G | A |
| LDL-C | 1 | 55085141 | G | A |
| LDL-C | 1 | 55136356 | T | C |
| LDL-C | 1 | 55138291 | G | A |
| LDL-C | 1 | 55141339 | C | A |
| LDL-C | 1 | 55142111 | T | G |
| LDL-C | 1 | 55160482 | T | G |
| LDL-C | 1 | 55224773 | C | A |
| LDL-C | 1 | 55243587 | G | A |
| LDL-C | 1 | 55334001 | G | A |
| LDL-C | 1 | 55378724 | G | A |
| LDL-C | 1 | 55411580 | G | A |
| LDL-C | 1 | 55417370 | T | C |
| LDL-C | 1 | 55431933 | T | C |
| LDL-C | 1 | 55470153 | T | C |
| LDL-C | 1 | 55489542 | T | C |
| LDL-C | 1 | 55489960 | G | A |
| LDL-C | 1 | 55493584 | C | A |
| LDL-C | 1 | 55494017 | T | C |
| LDL-C | 1 | 55496039 | T | C |
| LDL-C | 1 | 55500025 | C | A |
| LDL-C | 1 | 55500429 | G | A |
| LDL-C | 1 | 55503448 | G | A |
| LDL-C | 1 | 55504650 | G | A |
| LDL-C | 1 | 55505647 | T | G |
| LDL-C | 1 | 55505861 | G | A |
| LDL-C | 1 | 55507649 | T | C |
| LDL-C | 1 | 55511623 | TC | T |
| LDL-C | 1 | 55511995 | G | A |
| LDL-C | 1 | 55518467 | G | A |
| LDL-C | 1 | 55518528 | C | A |
| LDL-C | 1 | 55519174 | G | A |
| LDL-C | 1 | 55519238 | T | C |
| LDL-C | 1 | 55520212 | G | A |
| LDL-C | 1 | 55520265 | T | C |
| LDL-C | 1 | 55520864 | G | A |

**Table S14:** Genetic variants used in the presented analyses. Note that due to coverage of the GWAS chip the instruments differed across considered outcomes *(continued)*

| Exposure | Chromosome | Base-pair position (Build 37) | Other allele | Effect allele |
|---|---|---|---|---|
| LDL-C | 1 | 55521109 | G | A |
| LDL-C | 1 | 55522141 | G | A |
| LDL-C | 1 | 55522425 | G | A |
| LDL-C | 1 | 55523361 | G | A |
| LDL-C | 1 | 55524116 | T | C |
| LDL-C | 1 | 55524237 | G | A |
| LDL-C | 1 | 55524601 | G | A |
| LDL-C | 1 | 55527479 | G | A |
| LDL-C | 1 | 55527918 | T | C |
| LDL-C | 1 | 55538552 | T | G |
| LDL-C | 1 | 55636240 | T | C |
| LDL-C | 1 | 55656075 | T | G |
| LDL-C | 1 | 55711465 | G | A |
| LDL-C | 1 | 55713094 | G | A |
| LDL-C | 1 | 55713628 | G | A |
| LDL-C | 1 | 55716497 | G | A |
| LDL-C | 1 | 55719011 | G | A |
| LDL-C | 1 | 55719166 | T | G |
| LDL-C | 1 | 55719847 | CAT | C |
| LDL-C | 1 | 55720083 | C | A |
| LDL-C | 1 | 55720337 | G | A |
| LDL-C | 1 | 55723756 | C | A |
| LDL-C | 1 | 55726519 | T | C |
| LDL-C | 1 | 55730327 | T | C |
| LDL-C | 1 | 55731772 | TC | T |
| LDL-C | 1 | 55732596 | ATTGT | A |
| LDL-C | 1 | 55732837 | T | C |
| LDL-C | 1 | 55734875 | T | C |
| LDL-C | 1 | 55738663 | T | C |
| LDL-C | 1 | 55739925 | T | C |
| LDL-C | 1 | 55740015 | T | C |
| LDL-C | 1 | 55741377 | G | A |
| LDL-C | 1 | 55742091 | G | A |
| LDL-C | 1 | 55744167 | T | C |
| LDL-C | 1 | 55744966 | G | A |
| LDL-C | 1 | 55746330 | G | A |
| LDL-C | 1 | 55750138 | T | C |
| LDL-C | 1 | 55752440 | T | C |
| LDL-C | 1 | 55752699 | TA | T |
| LDL-C | 1 | 55754727 | G | A |
| LDL-C | 1 | 55767547 | T | C |
| LDL-C | 1 | 55785672 | T | C |
| LDL-C | 1 | 55785815 | T | G |
| LDL-C | 1 | 55786637 | T | C |

**Table S14:** Genetic variants used in the presented analyses. Note that due to coverage of the GWAS chip the instruments differed across considered outcomes *(continued)*

| Exposure | Chromosome | Base-pair position (Build 37) | Other allele | Effect allele |
|---|---|---|---|---|
| LDL-C | 1 | 55793244 | G | A |
| LDL-C | 1 | 55795814 | G | A |
| LDL-C | 1 | 55796086 | T | C |
| LDL-C | 1 | 55808143 | T | C |
| LDL-C | 1 | 55814244 | T | C |
| LDL-C | 1 | 55815584 | G | A |
| LDL-C | 1 | 55820713 | C | A |
| LDL-C | 1 | 55837603 | G | A |
| LDL-C | 1 | 55843810 | T | C |
| LDL-C | 1 | 55847507 | G | A |
| LDL-C | 1 | 55960656 | G | A |
| LDL-C | 1 | 56035239 | T | C |
| LDL-C | 1 | 56258328 | T | C |
| LDL-C | 1 | 56261771 | C | A |
| LDL-C | 1 | 56267033 | G | A |
| LDL-C | 1 | 56321074 | G | A |
| LDL-C | 1 | 56387285 | G | A |
| LDL-C | 1 | 56413519 | T | C |
| LDL-C | 1 | 56414772 | G | A |
| LDL-C | 1 | 56415347 | G | A |
| LDL-C | 1 | 56625395 | G | A |
| LDL-C | 1 | 56763141 | T | C |
| LDL-C | 1 | 56823728 | G | A |
| LDL-C | 1 | 56885874 | G | A |
| LDL-C | 2 | 20705963 | G | A |
| LDL-C | 2 | 20709694 | G | A |
| LDL-C | 2 | 20902748 | C | A |
| LDL-C | 2 | 20903015 | T | C |
| LDL-C | 2 | 20928295 | GCT | G |
| LDL-C | 2 | 20928972 | CT | C |
| LDL-C | 2 | 20955032 | AG | A |
| LDL-C | 2 | 20991528 | T | G |
| LDL-C | 2 | 21050729 | GA | G |
| LDL-C | 2 | 21076264 | TA | T |
| LDL-C | 2 | 21083052 | T | C |
| LDL-C | 2 | 21085410 | T | C |
| LDL-C | 2 | 21087323 | TCTC | T |
| LDL-C | 2 | 21088464 | G | A |
| LDL-C | 2 | 21091409 | C | A |
| LDL-C | 2 | 21093112 | G | A |
| LDL-C | 2 | 21098248 | C | A |
| LDL-C | 2 | 21099311 | T | C |
| LDL-C | 2 | 21100426 | T | C |
| LDL-C | 2 | 21101046 | G | A |

**Table S14:** Genetic variants used in the presented analyses. Note that due to coverage of the GWAS chip the instruments differed across considered outcomes *(continued)*

| Exposure | Chromosome | Base-pair position (Build 37) | Other allele | Effect allele |
|---|---|---|---|---|
| LDL-C | 2 | 21105335 | G | A |
| LDL-C | 2 | 21107494 | T | G |
| LDL-C | 2 | 21111034 | T | C |
| LDL-C | 2 | 21111763 | G | A |
| LDL-C | 2 | 21112689 | G | A |
| LDL-C | 2 | 21121464 | T | C |
| LDL-C | 2 | 21126936 | G | A |
| LDL-C | 2 | 21133883 | G | A |
| LDL-C | 2 | 21134011 | T | C |
| LDL-C | 2 | 21135187 | G | A |
| LDL-C | 2 | 21135577 | G | A |
| LDL-C | 2 | 21135723 | T | G |
| LDL-C | 2 | 21137425 | T | C |
| LDL-C | 2 | 21138066 | T | C |
| LDL-C | 2 | 21140540 | T | C |
| LDL-C | 2 | 21141725 | G | A |
| LDL-C | 2 | 21142994 | T | C |
| LDL-C | 2 | 21143289 | T | C |
| LDL-C | 2 | 21143329 | G | A |
| LDL-C | 2 | 21148274 | C | A |
| LDL-C | 2 | 21150787 | T | C |
| LDL-C | 2 | 21151408 | G | A |
| LDL-C | 2 | 21151917 | T | C |
| LDL-C | 2 | 21152755 | G | A |
| LDL-C | 2 | 21182116 | G | A |
| LDL-C | 2 | 21182710 | G | A |
| LDL-C | 2 | 21187599 | G | A |
| LDL-C | 2 | 21189091 | T | C |
| LDL-C | 2 | 21190024 | T | C |
| LDL-C | 2 | 21190209 | T | C |
| LDL-C | 2 | 21196112 | G | A |
| LDL-C | 2 | 21196346 | T | G |
| LDL-C | 2 | 21197159 | T | C |
| LDL-C | 2 | 21202959 | C | A |
| LDL-C | 2 | 21205920 | T | C |
| LDL-C | 2 | 21215645 | T | C |
| LDL-C | 2 | 21216815 | C | A |
| LDL-C | 2 | 21217490 | T | C |
| LDL-C | 2 | 21219001 | T | G |
| LDL-C | 2 | 21221133 | G | A |
| LDL-C | 2 | 21225753 | T | C |
| LDL-C | 2 | 21231524 | G | A |
| LDL-C | 2 | 21232804 | G | A |
| LDL-C | 2 | 21233972 | T | C |

**Table S14:** Genetic variants used in the presented analyses. Note that due to coverage of the GWAS chip the instruments differed across considered outcomes *(continued)*

| Exposure | Chromosome | Base-pair position (Build 37) | Other allele | Effect allele |
|---|---|---|---|---|
| LDL-C | 2 | 21239667 | T | C |
| LDL-C | 2 | 21241744 | T | C |
| LDL-C | 2 | 21244000 | T | G |
| LDL-C | 2 | 21245367 | G | A |
| LDL-C | 2 | 21247211 | T | C |
| LDL-C | 2 | 21250914 | G | A |
| LDL-C | 2 | 21251040 | T | G |
| LDL-C | 2 | 21259562 | T | C |
| LDL-C | 2 | 21259612 | G | A |
| LDL-C | 2 | 21260601 | G | A |
| LDL-C | 2 | 21262427 | G | A |
| LDL-C | 2 | 21263639 | T | G |
| LDL-C | 2 | 21263900 | G | A |
| LDL-C | 2 | 21266774 | GGCAGCGCCA | G |
| LDL-C | 2 | 21267210 | G | A |
| LDL-C | 2 | 21267461 | G | A |
| LDL-C | 2 | 21270751 | G | A |
| LDL-C | 2 | 21271323 | G | A |
| LDL-C | 2 | 21273490 | T | G |
| LDL-C | 2 | 21277922 | G | A |
| LDL-C | 2 | 21281856 | G | A |
| LDL-C | 2 | 21286057 | T | C |
| LDL-C | 2 | 21288226 | G | A |
| LDL-C | 2 | 21290067 | T | C |
| LDL-C | 2 | 21292377 | C | A |
| LDL-C | 2 | 21294975 | G | A |
| LDL-C | 2 | 21303616 | T | C |
| LDL-C | 2 | 21309335 | T | C |
| LDL-C | 2 | 21311541 | T | G |
| LDL-C | 2 | 21311691 | G | A |
| LDL-C | 2 | 21313577 | G | A |
| LDL-C | 2 | 21313778 | TC | T |
| LDL-C | 2 | 21319986 | GTGATGT | G |
| LDL-C | 2 | 21323868 | CTGT | C |
| LDL-C | 2 | 21327635 | G | A |
| LDL-C | 2 | 21328825 | G | A |
| LDL-C | 2 | 21367696 | TTGAAGAC | T |
| LDL-C | 2 | 21368217 | G | A |
| LDL-C | 2 | 21371068 | T | C |
| LDL-C | 2 | 21378453 | G | A |
| LDL-C | 2 | 21381177 | G | A |
| LDL-C | 2 | 21381214 | C | A |
| LDL-C | 2 | 21381292 | AAAAC | A |
| LDL-C | 2 | 21383353 | T | C |

**Table S14:** Genetic variants used in the presented analyses. Note that due to coverage of the GWAS chip the instruments differed across considered outcomes *(continued)*

| Exposure | Chromosome | Base-pair position (Build 37) | Other allele | Effect allele |
|---|---|---|---|---|
| LDL-C | 2 | 21383514 | G | A |
| LDL-C | 2 | 21389659 | T | G |
| LDL-C | 2 | 21393623 | G | A |
| LDL-C | 2 | 21396092 | GAA | G |
| LDL-C | 2 | 21397182 | G | A |
| LDL-C | 2 | 21401902 | GAAAGAT | G |
| LDL-C | 2 | 21404245 | CTA | C |
| LDL-C | 2 | 21406048 | T | C |
| LDL-C | 2 | 21411649 | G | A |
| LDL-C | 2 | 21413077 | G | A |
| LDL-C | 2 | 21414142 | T | C |
| LDL-C | 2 | 21415728 | T | G |
| LDL-C | 2 | 21419650 | G | A |
| LDL-C | 2 | 21420358 | C | A |
| LDL-C | 2 | 21423218 | AT | A |
| LDL-C | 2 | 21423532 | G | A |
| LDL-C | 2 | 21426683 | AAAAG | A |
| LDL-C | 2 | 21428049 | C | A |
| LDL-C | 2 | 21428185 | G | A |
| LDL-C | 2 | 21434580 | G | A |
| LDL-C | 2 | 21435700 | C | A |
| LDL-C | 2 | 21437321 | T | G |
| LDL-C | 2 | 21438738 | G | A |
| LDL-C | 2 | 21439329 | T | G |
| LDL-C | 2 | 21442944 | TTGCTCCCCTCTGCTTAGCA | T |
| LDL-C | 2 | 21445569 | G | A |
| LDL-C | 2 | 21448511 | G | A |
| LDL-C | 2 | 21449034 | T | C |
| LDL-C | 2 | 21449987 | G | A |
| LDL-C | 2 | 21450714 | G | A |
| LDL-C | 2 | 21451604 | T | C |
| LDL-C | 2 | 21451687 | G | A |
| LDL-C | 2 | 21453733 | G | A |
| LDL-C | 2 | 21457324 | G | A |
| LDL-C | 2 | 21460176 | T | C |
| LDL-C | 2 | 21463922 | T | C |
| LDL-C | 2 | 21467872 | ATT | A |
| LDL-C | 2 | 21469336 | T | C |
| LDL-C | 2 | 21469738 | T | G |
| LDL-C | 2 | 21470596 | G | A |
| LDL-C | 2 | 21470987 | T | C |
| LDL-C | 2 | 21471603 | T | C |
| LDL-C | 2 | 21475944 | T | C |
| LDL-C | 2 | 21476247 | C | A |

**Table S14:** Genetic variants used in the presented analyses. Note that due to coverage of the GWAS chip the instruments differed across considered outcomes *(continued)*

| Exposure | Chromosome | Base-pair position (Build 37) | Other allele | Effect allele |
|---|---|---|---|---|
| LDL-C | 2 | 21476634 | G | A |
| LDL-C | 2 | 21477012 | T | G |
| LDL-C | 2 | 21482889 | TGCACTGAG | T |
| LDL-C | 2 | 21524000 | G | A |
| LDL-C | 2 | 21530659 | G | A |
| LDL-C | 2 | 21534103 | T | C |
| LDL-C | 2 | 21534298 | T | G |
| LDL-C | 2 | 44052743 | GTTA | G |
| LDL-C | 2 | 44057570 | C | A |
| LDL-C | 2 | 44057825 | TCTC | T |
| LDL-C | 2 | 44058201 | G | A |
| LDL-C | 2 | 44073656 | T | C |
| LDL-C | 2 | 44073881 | T | C |
| LDL-C | 2 | 44074000 | G | A |
| LDL-C | 2 | 44074431 | T | C |
| LDL-C | 2 | 44080324 | G | A |
| LDL-C | 2 | 44088039 | G | A |
| LDL-C | 2 | 44088041 | C | A |
| LDL-C | 2 | 44096259 | T | G |
| LDL-C | 2 | 44096402 | G | A |
| LDL-C | 4 | 73240747 | G | A |
| LDL-C | 4 | 73406173 | G | A |
| LDL-C | 4 | 73412023 | T | C |
| LDL-C | 4 | 73542640 | T | G |
| LDL-C | 4 | 74720969 | C | A |
| LDL-C | 4 | 74805105 | G | A |
| LDL-C | 4 | 74970615 | C | A |
| LDL-C | 4 | 75012853 | G | A |
| LDL-C | 4 | 75128036 | G | A |
| LDL-C | 4 | 75242737 | CT | C |
| LDL-C | 4 | 75321980 | T | C |
| LDL-C | 4 | 75370891 | T | C |
| LDL-C | 4 | 75684215 | T | C |
| LDL-C | 5 | 74302658 | G | A |
| LDL-C | 5 | 74340327 | C | A |
| LDL-C | 5 | 74352180 | G | A |
| LDL-C | 5 | 74354875 | T | G |
| LDL-C | 5 | 74360981 | ATCT | A |
| LDL-C | 5 | 74368254 | T | C |
| LDL-C | 5 | 74372141 | T | C |
| LDL-C | 5 | 74384115 | G | A |
| LDL-C | 5 | 74389325 | C | A |
| LDL-C | 5 | 74413713 | T | C |
| LDL-C | 5 | 74420422 | G | A |

**Table S14:** Genetic variants used in the presented analyses. Note that due to coverage of the GWAS chip the instruments differed across considered outcomes *(continued)*

| Exposure | Chromosome | Base-pair position (Build 37) | Other allele | Effect allele |
|---|---|---|---|---|
| LDL-C | 5 | 74443132 | T | C |
| LDL-C | 5 | 74461850 | T | G |
| LDL-C | 5 | 74476396 | G | A |
| LDL-C | 5 | 74477719 | TA | T |
| LDL-C | 5 | 74479238 | T | C |
| LDL-C | 5 | 74480288 | T | C |
| LDL-C | 5 | 74547484 | C | A |
| LDL-C | 5 | 74558893 | G | A |
| LDL-C | 5 | 74560652 | T | C |
| LDL-C | 5 | 74562564 | T | C |
| LDL-C | 5 | 74567359 | G | A |
| LDL-C | 5 | 74574984 | G | A |
| LDL-C | 5 | 74586866 | CT | C |
| LDL-C | 5 | 74587940 | C | A |
| LDL-C | 5 | 74596682 | T | G |
| LDL-C | 5 | 74602699 | C | A |
| LDL-C | 5 | 74604742 | G | A |
| LDL-C | 5 | 74605220 | G | A |
| LDL-C | 5 | 74611232 | TG | T |
| LDL-C | 5 | 74613943 | T | C |
| LDL-C | 5 | 74615021 | T | C |
| LDL-C | 5 | 74616843 | T | C |
| LDL-C | 5 | 74619639 | T | C |
| LDL-C | 5 | 74622121 | TTA | T |
| LDL-C | 5 | 74624234 | G | A |
| LDL-C | 5 | 74625512 | G | A |
| LDL-C | 5 | 74629162 | CAT | C |
| LDL-C | 5 | 74630189 | T | C |
| LDL-C | 5 | 74630277 | T | G |
| LDL-C | 5 | 74631352 | T | C |
| LDL-C | 5 | 74641090 | CT | C |
| LDL-C | 5 | 74643998 | ATTC | A |
| LDL-C | 5 | 74644785 | TA | T |
| LDL-C | 5 | 74651084 | G | A |
| LDL-C | 5 | 74652754 | ATCCATCCGTCATGTCCG | A |
| LDL-C | 5 | 74653462 | T | G |
| LDL-C | 5 | 74656539 | T | C |
| LDL-C | 5 | 74660064 | ATTG | A |
| LDL-C | 5 | 74660356 | T | C |
| LDL-C | 5 | 74673707 | G | A |
| LDL-C | 5 | 74692776 | G | A |
| LDL-C | 5 | 74697050 | T | G |
| LDL-C | 5 | 74717653 | T | C |
| LDL-C | 5 | 74717767 | G | A |

**Table S14:** Genetic variants used in the presented analyses. Note that due to coverage of the GWAS chip the instruments differed across considered outcomes *(continued)*

| Exposure | Chromosome | Base-pair position (Build 37) | Other allele | Effect allele |
|---|---|---|---|---|
| LDL-C | 5 | 74736600 | TA | T |
| LDL-C | 5 | 74745228 | T | G |
| LDL-C | 5 | 74808237 | T | G |
| LDL-C | 5 | 74826735 | G | A |
| LDL-C | 5 | 74832919 | T | C |
| LDL-C | 5 | 74879890 | G | A |
| LDL-C | 5 | 74906963 | C | A |
| LDL-C | 5 | 74952861 | C | A |
| LDL-C | 5 | 74953504 | T | C |
| LDL-C | 5 | 74956517 | T | C |
| LDL-C | 5 | 74965122 | G | A |
| LDL-C | 5 | 74967386 | T | C |
| LDL-C | 5 | 75003056 | G | A |
| LDL-C | 5 | 75003678 | T | C |
| LDL-C | 5 | 75034387 | T | C |
| LDL-C | 5 | 75038267 | G | A |
| LDL-C | 6 | 31901264 | G | A |
| LDL-C | 6 | 32024234 | G | A |
| LDL-C | 6 | 32093029 | T | C |
| LDL-C | 7 | 100897350 | G | A |
| LDL-C | 7 | 100897643 | T | G |
| LDL-C | 7 | 21557854 | G | A |
| LDL-C | 7 | 21563471 | T | C |
| LDL-C | 7 | 21567331 | T | C |
| LDL-C | 7 | 21569089 | G | A |
| LDL-C | 7 | 21576905 | T | C |
| LDL-C | 7 | 21577960 | T | C |
| LDL-C | 7 | 21582917 | T | C |
| LDL-C | 7 | 21586595 | T | C |
| LDL-C | 7 | 21588243 | C | A |
| LDL-C | 7 | 21589030 | T | C |
| LDL-C | 7 | 21589243 | G | A |
| LDL-C | 7 | 21592217 | G | A |
| LDL-C | 7 | 21593289 | T | C |
| LDL-C | 7 | 21596006 | G | A |
| LDL-C | 7 | 21599807 | C | A |
| LDL-C | 7 | 21601659 | GCTCT | G |
| LDL-C | 7 | 21602065 | G | A |
| LDL-C | 7 | 21603601 | G | A |
| LDL-C | 7 | 21603678 | G | A |
| LDL-C | 7 | 21607352 | T | C |
| LDL-C | 7 | 21614216 | T | C |
| LDL-C | 7 | 21627909 | G | A |
| LDL-C | 7 | 21631827 | G | A |

**Table S14:** Genetic variants used in the presented analyses. Note that due to coverage of the GWAS chip the instruments differed across considered outcomes *(continued)*

| Exposure | Chromosome | Base-pair position (Build 37) | Other allele | Effect allele |
|---|---|---|---|---|
| LDL-C | 7 | 21637976 | G | A |
| LDL-C | 7 | 21643513 | T | C |
| LDL-C | 7 | 21647114 | T | G |
| LDL-C | 7 | 21650595 | G | A |
| LDL-C | 7 | 21653877 | G | A |
| LDL-C | 7 | 21670593 | G | A |
| LDL-C | 7 | 21670846 | G | A |
| LDL-C | 7 | 44580876 | T | G |
| LDL-C | 8 | 108336960 | T | G |
| LDL-C | 8 | 108341989 | G | A |
| LDL-C | 8 | 108352779 | T | C |
| LDL-C | 8 | 108354035 | G | A |
| LDL-C | 8 | 126476873 | T | C |
| LDL-C | 8 | 126479362 | T | C |
| LDL-C | 8 | 126486409 | G | A |
| LDL-C | 8 | 126488930 | G | A |
| LDL-C | 8 | 126495818 | T | C |
| LDL-C | 8 | 126506694 | G | A |
| LDL-C | 8 | 144974963 | G | A |
| LDL-C | 8 | 144976572 | G | A |
| LDL-C | 8 | 144976720 | T | C |
| LDL-C | 8 | 144978607 | T | G |
| LDL-C | 8 | 144984283 | T | C |
| LDL-C | 8 | 144992103 | T | C |
| LDL-C | 8 | 144997927 | G | A |
| LDL-C | 8 | 145003777 | G | A |
| LDL-C | 8 | 145004944 | T | C |
| LDL-C | 8 | 145013164 | TC | T |
| LDL-C | 8 | 145031265 | G | A |
| LDL-C | 8 | 145043543 | G | A |
| LDL-C | 8 | 145057868 | G | A |
| LDL-C | 8 | 145058037 | G | A |
| LDL-C | 8 | 145058986 | G | A |
| LDL-C | 8 | 145062020 | T | C |
| LDL-C | 8 | 145068489 | AG | A |
| LDL-C | 8 | 145071742 | TC | T |
| LDL-C | 8 | 145073663 | T | C |
| LDL-C | 8 | 145081248 | G | A |
| LDL-C | 8 | 9173358 | G | A |
| LDL-C | 8 | 9181395 | G | A |
| LDL-C | 8 | 9183596 | G | A |
| LDL-C | 8 | 9183664 | G | A |
| LDL-C | 8 | 9185146 | T | C |
| LDL-C | 8 | 9192423 | G | A |

**Table S14:** Genetic variants used in the presented analyses. Note that due to coverage of the GWAS chip the instruments differed across considered outcomes *(continued)*

| Exposure | Chromosome | Base-pair position (Build 37) | Other allele | Effect allele |
|---|---|---|---|---|
| LDL-C | 8 | 9201787 | T | C |
| LDL-C | 8 | 9206069 | G | A |
| LDL-C | 8 | 9206678 | G | A |
| LDL-C | 8 | 9211723 | G | A |
| LDL-C | 9 | 136139907 | GAAACTGCC | G |
| LDL-C | 9 | 136145425 | C | A |
| LDL-C | 9 | 136146597 | T | C |
| LDL-C | 9 | 136149399 | G | A |
| LDL-C | 9 | 136153875 | T | C |
| LDL-C | 9 | 136154168 | T | C |
| LDL-C | 9 | 136184782 | G | A |
| LDL-C | 9 | 2644315 | T | C |
| HDL-C | 10 | 49910599 | T | C |
| HDL-C | 11 | 116499500 | G | A |
| HDL-C | 11 | 116521316 | G | A |
| HDL-C | 11 | 116522854 | G | A |
| HDL-C | 11 | 116525867 | T | C |
| HDL-C | 11 | 116526322 | C | A |
| HDL-C | 11 | 116586283 | T | C |
| HDL-C | 11 | 116598988 | G | A |
| HDL-C | 11 | 116603724 | C | A |
| HDL-C | 11 | 116606766 | G | A |
| HDL-C | 11 | 116607368 | C | A |
| HDL-C | 11 | 116611985 | G | A |
| HDL-C | 11 | 116614863 | T | C |
| HDL-C | 11 | 116621963 | T | G |
| HDL-C | 11 | 116629949 | T | C |
| HDL-C | 11 | 116631707 | T | C |
| HDL-C | 11 | 116632975 | G | A |
| HDL-C | 11 | 116633862 | G | A |
| HDL-C | 11 | 116639104 | G | A |
| HDL-C | 11 | 116639692 | T | C |
| HDL-C | 11 | 116645275 | T | C |
| HDL-C | 11 | 116651115 | T | C |
| HDL-C | 11 | 116652423 | G | A |
| HDL-C | 11 | 116657561 | T | C |
| HDL-C | 11 | 116660686 | G | A |
| HDL-C | 11 | 116660813 | G | A |
| HDL-C | 11 | 116661826 | T | C |
| HDL-C | 11 | 116662579 | T | C |
| HDL-C | 11 | 116664040 | G | A |
| HDL-C | 11 | 116699395 | C | A |
| HDL-C | 11 | 116705278 | T | C |
| HDL-C | 11 | 116708413 | T | C |

**Table S14:** Genetic variants used in the presented analyses. Note that due to coverage of the GWAS chip the instruments differed across considered outcomes *(continued)*

| Exposure | Chromosome | Base-pair position (Build 37) | Other allele | Effect allele |
|---|---|---|---|---|
| HDL-C | 11 | 116869217 | T | C |
| HDL-C | 11 | 116883665 | T | C |
| HDL-C | 11 | 116933363 | GA | G |
| HDL-C | 11 | 116963312 | T | C |
| HDL-C | 11 | 117027942 | T | C |
| HDL-C | 11 | 117071505 | T | C |
| HDL-C | 11 | 117095283 | T | C |
| HDL-C | 11 | 47246397 | G | A |
| HDL-C | 11 | 47290984 | T | G |
| HDL-C | 11 | 61547237 | T | C |
| HDL-C | 11 | 61564299 | T | G |
| HDL-C | 11 | 61580635 | T | C |
| HDL-C | 11 | 61581656 | G | A |
| HDL-C | 11 | 61581764 | T | C |
| HDL-C | 11 | 61586328 | CT | C |
| HDL-C | 11 | 61589481 | G | A |
| HDL-C | 11 | 61591995 | GAA | G |
| HDL-C | 11 | 61594920 | CT | C |
| HDL-C | 11 | 61597212 | T | C |
| HDL-C | 11 | 61597972 | G | A |
| HDL-C | 11 | 61603237 | T | C |
| HDL-C | 11 | 61605215 | G | A |
| HDL-C | 11 | 61611216 | T | C |
| HDL-C | 11 | 61616012 | G | A |
| HDL-C | 11 | 61618012 | T | C |
| HDL-C | 11 | 61620628 | AAACAAC | A |
| HDL-C | 11 | 61620703 | CTCTT | C |
| HDL-C | 11 | 61621194 | G | A |
| HDL-C | 11 | 61623140 | T | C |
| HDL-C | 11 | 61624181 | T | C |
| HDL-C | 11 | 61624414 | T | C |
| HDL-C | 11 | 61625723 | CA | C |
| HDL-C | 11 | 61626206 | G | A |
| HDL-C | 11 | 61627811 | T | C |
| HDL-C | 11 | 61629122 | G | A |
| HDL-C | 11 | 61629666 | G | A |
| HDL-C | 11 | 61637587 | T | C |
| HDL-C | 11 | 61639488 | T | C |
| HDL-C | 11 | 61641542 | T | G |
| HDL-C | 11 | 61647427 | T | G |
| HDL-C | 11 | 61656117 | G | A |
| HDL-C | 11 | 61656182 | C | A |
| HDL-C | 11 | 61656995 | T | C |
| HDL-C | 11 | 61657110 | T | C |

**Table S14:** Genetic variants used in the presented analyses. Note that due to coverage of the GWAS chip the instruments differed across considered outcomes *(continued)*

| Exposure | Chromosome | Base-pair position (Build 37) | Other allele | Effect allele |
|---|---|---|---|---|
| HDL-C | 12 | 109953417 | G | A |
| HDL-C | 12 | 125264293 | G | A |
| HDL-C | 12 | 125282882 | G | A |
| HDL-C | 12 | 125284748 | G | A |
| HDL-C | 12 | 125286037 | T | C |
| HDL-C | 12 | 125293619 | T | C |
| HDL-C | 12 | 125293686 | G | A |
| HDL-C | 12 | 125295383 | T | C |
| HDL-C | 12 | 125317391 | C | A |
| HDL-C | 12 | 125324197 | G | A |
| HDL-C | 12 | 125324798 | C | A |
| HDL-C | 12 | 125335510 | C | A |
| HDL-C | 12 | 125338336 | T | C |
| HDL-C | 12 | 125339095 | G | A |
| HDL-C | 12 | 125345448 | G | A |
| HDL-C | 12 | 125349480 | G | A |
| HDL-C | 12 | 125351116 | T | C |
| HDL-C | 12 | 125351149 | G | A |
| HDL-C | 12 | 125351772 | T | C |
| HDL-C | 12 | 125352603 | T | C |
| HDL-C | 12 | 125363987 | C | A |
| HDL-C | 12 | 125380232 | G | A |
| HDL-C | 15 | 58557062 | G | A |
| HDL-C | 15 | 58579369 | TATCAC | T |
| HDL-C | 15 | 58580525 | G | A |
| HDL-C | 15 | 58586129 | T | C |
| HDL-C | 15 | 58591415 | T | C |
| HDL-C | 15 | 58611859 | T | G |
| HDL-C | 15 | 58630019 | T | G |
| HDL-C | 15 | 58661083 | T | C |
| HDL-C | 15 | 58670135 | T | C |
| HDL-C | 15 | 58671178 | C | A |
| HDL-C | 15 | 58671721 | G | A |
| HDL-C | 15 | 58671765 | T | G |
| HDL-C | 15 | 58672134 | T | C |
| HDL-C | 15 | 58674051 | G | A |
| HDL-C | 15 | 58674308 | G | A |
| HDL-C | 15 | 58678512 | T | C |
| HDL-C | 15 | 58679668 | G | A |
| HDL-C | 15 | 58679807 | CAGA | C |
| HDL-C | 15 | 58680638 | T | C |
| HDL-C | 15 | 58680639 | T | C |
| HDL-C | 15 | 58680805 | G | A |
| HDL-C | 15 | 58680954 | T | C |

**Table S14:** Genetic variants used in the presented analyses. Note that due to coverage of the GWAS chip the instruments differed across considered outcomes *(continued)*

| Exposure | Chromosome | Base-pair position (Build 37) | Other allele | Effect allele |
|---|---|---|---|---|
| HDL-C | 15 | 58682690 | G | A |
| HDL-C | 15 | 58683366 | G | A |
| HDL-C | 15 | 58683697 | T | C |
| HDL-C | 15 | 58684478 | T | C |
| HDL-C | 15 | 58684967 | T | G |
| HDL-C | 15 | 58686252 | G | A |
| HDL-C | 15 | 58686338 | G | A |
| HDL-C | 15 | 58687530 | G | A |
| HDL-C | 15 | 58687603 | T | C |
| HDL-C | 15 | 58687862 | G | A |
| HDL-C | 15 | 58687880 | T | C |
| HDL-C | 15 | 58688477 | G | A |
| HDL-C | 15 | 58688478 | C | A |
| HDL-C | 15 | 58689187 | T | C |
| HDL-C | 15 | 58690738 | C | A |
| HDL-C | 15 | 58690754 | G | A |
| HDL-C | 15 | 58692202 | C | A |
| HDL-C | 15 | 58692479 | T | C |
| HDL-C | 15 | 58692715 | T | C |
| HDL-C | 15 | 58693860 | T | C |
| HDL-C | 15 | 58694020 | T | G |
| HDL-C | 15 | 58696300 | T | C |
| HDL-C | 15 | 58698113 | T | G |
| HDL-C | 15 | 58698243 | T | G |
| HDL-C | 15 | 58698755 | T | G |
| HDL-C | 15 | 58699010 | T | C |
| HDL-C | 15 | 58699937 | T | C |
| HDL-C | 15 | 58700311 | T | C |
| HDL-C | 15 | 58702325 | G | A |
| HDL-C | 15 | 58703069 | C | A |
| HDL-C | 15 | 58704064 | T | G |
| HDL-C | 15 | 58705029 | T | C |
| HDL-C | 15 | 58705658 | AC | A |
| HDL-C | 15 | 58706797 | G | A |
| HDL-C | 15 | 58709181 | T | C |
| HDL-C | 15 | 58709182 | G | A |
| HDL-C | 15 | 58710627 | C | A |
| HDL-C | 15 | 58710736 | T | C |
| HDL-C | 15 | 58712203 | T | G |
| HDL-C | 15 | 58712945 | C | A |
| HDL-C | 15 | 58712973 | T | C |
| HDL-C | 15 | 58716044 | G | A |
| HDL-C | 15 | 58716257 | G | A |
| HDL-C | 15 | 58716615 | T | G |

**Table S14:** Genetic variants used in the presented analyses. Note that due to coverage of the GWAS chip the instruments differed across considered outcomes *(continued)*

| Exposure | Chromosome | Base-pair position (Build 37) | Other allele | Effect allele |
|---|---|---|---|---|
| HDL-C | 15 | 58717919 | T | C |
| HDL-C | 15 | 58718529 | CA | C |
| HDL-C | 15 | 58719741 | G | A |
| HDL-C | 15 | 58720431 | C | A |
| HDL-C | 15 | 58721302 | G | A |
| HDL-C | 15 | 58721614 | G | A |
| HDL-C | 15 | 58722590 | GC | G |
| HDL-C | 15 | 58723479 | T | C |
| HDL-C | 15 | 58723675 | T | C |
| HDL-C | 15 | 58724371 | T | C |
| HDL-C | 15 | 58724706 | G | A |
| HDL-C | 15 | 58727279 | T | C |
| HDL-C | 15 | 58727325 | G | A |
| HDL-C | 15 | 58728067 | G | A |
| HDL-C | 15 | 58729144 | T | C |
| HDL-C | 15 | 58730498 | T | C |
| HDL-C | 15 | 58731395 | T | C |
| HDL-C | 15 | 58731455 | T | C |
| HDL-C | 15 | 58731567 | G | A |
| HDL-C | 15 | 58735222 | C | A |
| HDL-C | 15 | 58735638 | T | G |
| HDL-C | 15 | 58736778 | T | C |
| HDL-C | 15 | 58737341 | T | G |
| HDL-C | 15 | 58738423 | T | C |
| HDL-C | 15 | 58741891 | G | A |
| HDL-C | 15 | 58742418 | G | A |
| HDL-C | 15 | 58742731 | T | G |
| HDL-C | 15 | 58743022 | T | G |
| HDL-C | 15 | 58743709 | T | C |
| HDL-C | 15 | 58882792 | T | C |
| HDL-C | 15 | 58899609 | T | C |
| HDL-C | 15 | 58947292 | T | C |
| HDL-C | 15 | 59093625 | G | A |
| HDL-C | 15 | 59351989 | G | A |
| HDL-C | 15 | 59566385 | T | C |
| HDL-C | 15 | 59585013 | T | C |
| HDL-C | 15 | 59806496 | T | G |
| HDL-C | 16 | 56742475 | G | A |
| HDL-C | 16 | 56745752 | T | C |
| HDL-C | 16 | 56758596 | G | A |
| HDL-C | 16 | 56759532 | G | A |
| HDL-C | 16 | 56808843 | G | A |
| HDL-C | 16 | 56811400 | G | A |
| HDL-C | 16 | 56827946 | G | A |

**Table S14:** Genetic variants used in the presented analyses. Note that due to coverage of the GWAS chip the instruments differed across considered outcomes *(continued)*

| Exposure | Chromosome | Base-pair position (Build 37) | Other allele | Effect allele |
|---|---|---|---|---|
| HDL-C | 16 | 56866054 | AT | A |
| HDL-C | 16 | 56898198 | G | A |
| HDL-C | 16 | 56899006 | T | C |
| HDL-C | 16 | 56900100 | G | A |
| HDL-C | 16 | 56900931 | T | C |
| HDL-C | 16 | 56909598 | T | C |
| HDL-C | 16 | 56910692 | T | G |
| HDL-C | 16 | 56920104 | G | A |
| HDL-C | 16 | 56920146 | G | A |
| HDL-C | 16 | 56922660 | T | C |
| HDL-C | 16 | 56927720 | T | C |
| HDL-C | 16 | 56929944 | G | A |
| HDL-C | 16 | 56930251 | T | C |
| HDL-C | 16 | 56932531 | G | A |
| HDL-C | 16 | 56933519 | G | A |
| HDL-C | 16 | 56938016 | G | A |
| HDL-C | 16 | 56938522 | T | C |
| HDL-C | 16 | 56938955 | C | A |
| HDL-C | 16 | 56939969 | G | A |
| HDL-C | 16 | 56940513 | T | C |
| HDL-C | 16 | 56940874 | T | C |
| HDL-C | 16 | 56942709 | G | A |
| HDL-C | 16 | 56945049 | C | A |
| HDL-C | 16 | 56952098 | G | A |
| HDL-C | 16 | 56953261 | G | A |
| HDL-C | 16 | 56960147 | G | A |
| HDL-C | 16 | 56972713 | G | A |
| HDL-C | 16 | 56980135 | G | A |
| HDL-C | 16 | 56982549 | T | C |
| HDL-C | 16 | 56984482 | T | C |
| HDL-C | 16 | 56984590 | T | C |
| HDL-C | 16 | 56984763 | G | A |
| HDL-C | 16 | 56985139 | G | A |
| HDL-C | 16 | 56985156 | G | A |
| HDL-C | 16 | 56985514 | G | A |
| HDL-C | 16 | 56985555 | G | A |
| HDL-C | 16 | 56985805 | T | C |
| HDL-C | 16 | 56986762 | T | G |
| HDL-C | 16 | 56988044 | T | C |
| HDL-C | 16 | 56988503 | T | C |
| HDL-C | 16 | 56990716 | C | A |
| HDL-C | 16 | 56991363 | T | C |
| HDL-C | 16 | 56991524 | G | A |
| HDL-C | 16 | 56991741 | T | C |

**Table S14:** Genetic variants used in the presented analyses. Note that due to coverage of the GWAS chip the instruments differed across considered outcomes *(continued)*

| Exposure | Chromosome | Base-pair position (Build 37) | Other allele | Effect allele |
|---|---|---|---|---|
| HDL-C | 16 | 56992017 | G | A |
| HDL-C | 16 | 56993025 | T | G |
| HDL-C | 16 | 56993211 | T | C |
| HDL-C | 16 | 56993324 | C | A |
| HDL-C | 16 | 56993897 | C | A |
| HDL-C | 16 | 56993901 | C | A |
| HDL-C | 16 | 56993909 | C | A |
| HDL-C | 16 | 56994192 | T | C |
| HDL-C | 16 | 56994212 | T | C |
| HDL-C | 16 | 56994244 | TA | T |
| HDL-C | 16 | 56994894 | G | A |
| HDL-C | 16 | 56995038 | T | C |
| HDL-C | 16 | 56995236 | C | A |
| HDL-C | 16 | 56996158 | T | C |
| HDL-C | 16 | 56996211 | G | A |
| HDL-C | 16 | 56996645 | GCC | G |
| HDL-C | 16 | 56997233 | G | A |
| HDL-C | 16 | 56997349 | CA | C |
| HDL-C | 16 | 56998918 | G | A |
| HDL-C | 16 | 56999258 | C | A |
| HDL-C | 16 | 56999328 | T | C |
| HDL-C | 16 | 57000284 | G | A |
| HDL-C | 16 | 57000938 | G | A |
| HDL-C | 16 | 57001254 | TCACA | T |
| HDL-C | 16 | 57001274 | AC | A |
| HDL-C | 16 | 57002118 | T | G |
| HDL-C | 16 | 57002404 | C | A |
| HDL-C | 16 | 57002732 | T | G |
| HDL-C | 16 | 57003146 | T | C |
| HDL-C | 16 | 57004889 | G | A |
| HDL-C | 16 | 57005479 | C | A |
| HDL-C | 16 | 57006590 | T | C |
| HDL-C | 16 | 57007353 | T | C |
| HDL-C | 16 | 57007451 | G | A |
| HDL-C | 16 | 57008048 | G | A |
| HDL-C | 16 | 57009651 | TC | T |
| HDL-C | 16 | 57010232 | T | C |
| HDL-C | 16 | 57010382 | T | G |
| HDL-C | 16 | 57010486 | T | C |
| HDL-C | 16 | 57010948 | T | G |
| HDL-C | 16 | 57011185 | G | A |
| HDL-C | 16 | 57012699 | G | A |
| HDL-C | 16 | 57014610 | T | C |
| HDL-C | 16 | 57014875 | G | A |

**Table S14:** Genetic variants used in the presented analyses. Note that due to coverage of the GWAS chip the instruments differed across considered outcomes *(continued)*

| Exposure | Chromosome | Base-pair position (Build 37) | Other allele | Effect allele |
|---|---|---|---|---|
| HDL-C | 16 | 57016092 | G | A |
| HDL-C | 16 | 57017002 | T | G |
| HDL-C | 16 | 57017319 | G | A |
| HDL-C | 16 | 57019392 | T | C |
| HDL-C | 16 | 57021433 | G | A |
| HDL-C | 16 | 57060097 | T | C |
| HDL-C | 16 | 68090982 | TATATA | T |
| HDL-C | 16 | 68229579 | TGGTCGC | T |
| HDL-C | 17 | 41840849 | T | C |
| HDL-C | 17 | 41926126 | T | C |
| HDL-C | 17 | 41999943 | G | A |
| HDL-C | 18 | 46529282 | T | G |
| HDL-C | 18 | 47049700 | G | A |
| HDL-C | 18 | 47066144 | C | A |
| HDL-C | 18 | 47093864 | T | C |
| HDL-C | 18 | 47096016 | G | A |
| HDL-C | 18 | 47109955 | G | A |
| HDL-C | 18 | 47128176 | T | C |
| HDL-C | 18 | 47129148 | T | C |
| HDL-C | 18 | 47131888 | C | A |
| HDL-C | 18 | 47133828 | G | A |
| HDL-C | 18 | 47134175 | T | C |
| HDL-C | 18 | 47147746 | G | A |
| HDL-C | 18 | 47148299 | T | C |
| HDL-C | 18 | 47148886 | T | C |
| HDL-C | 18 | 47149371 | G | A |
| HDL-C | 18 | 47149635 | C | A |
| HDL-C | 18 | 47156730 | G | A |
| HDL-C | 18 | 47157004 | T | C |
| HDL-C | 18 | 47158186 | T | C |
| HDL-C | 18 | 47158234 | T | C |
| HDL-C | 18 | 47159090 | G | A |
| HDL-C | 18 | 47160814 | G | A |
| HDL-C | 18 | 47161733 | T | C |
| HDL-C | 18 | 47161892 | T | C |
| HDL-C | 18 | 47166694 | G | A |
| HDL-C | 18 | 47168648 | G | A |
| HDL-C | 18 | 47169340 | T | G |
| HDL-C | 18 | 47170139 | ACCTG | A |
| HDL-C | 18 | 47176261 | C | A |
| HDL-C | 18 | 47176793 | G | A |
| HDL-C | 18 | 47179516 | G | A |
| HDL-C | 18 | 47182664 | G | A |
| HDL-C | 18 | 47183012 | T | C |

**Table S14:** Genetic variants used in the presented analyses. Note that due to coverage of the GWAS chip the instruments differed across considered outcomes *(continued)*

| Exposure | Chromosome | Base-pair position (Build 37) | Other allele | Effect allele |
|---|---|---|---|---|
| HDL-C | 18 | 47184006 | G | A |
| HDL-C | 18 | 47243912 | C | A |
| HDL-C | 18 | 47277223 | T | C |
| HDL-C | 19 | 11317744 | T | C |
| HDL-C | 19 | 11346550 | T | C |
| HDL-C | 19 | 11347493 | T | C |
| HDL-C | 19 | 45396665 | T | G |
| HDL-C | 19 | 45413576 | T | C |
| HDL-C | 19 | 8429323 | G | A |
| HDL-C | 1 | 182161829 | T | C |
| HDL-C | 1 | 182168885 | G | A |
| HDL-C | 1 | 182170792 | T | C |
| HDL-C | 1 | 220967683 | G | A |
| HDL-C | 1 | 220970028 | G | A |
| HDL-C | 1 | 220973563 | T | C |
| HDL-C | 1 | 230280696 | T | C |
| HDL-C | 1 | 230293530 | T | C |
| HDL-C | 1 | 230294989 | C | A |
| HDL-C | 1 | 230295245 | T | C |
| HDL-C | 1 | 230295691 | G | A |
| HDL-C | 1 | 230300481 | T | C |
| HDL-C | 1 | 230302521 | T | G |
| HDL-C | 1 | 230303512 | T | C |
| HDL-C | 1 | 230307182 | T | C |
| HDL-C | 1 | 230307222 | T | G |
| HDL-C | 1 | 230309619 | G | A |
| HDL-C | 1 | 230310568 | T | C |
| HDL-C | 1 | 230324067 | T | G |
| HDL-C | 1 | 230324119 | G | A |
| HDL-C | 1 | 230324801 | G | A |
| HDL-C | 20 | 43042364 | T | C |
| HDL-C | 20 | 44534651 | G | A |
| HDL-C | 20 | 44554015 | T | C |
| HDL-C | 20 | 44563007 | T | C |
| HDL-C | 20 | 44576502 | T | C |
| HDL-C | 2 | 136429366 | T | C |
| HDL-C | 2 | 136608646 | G | A |
| HDL-C | 2 | 136636324 | G | A |
| HDL-C | 2 | 136696138 | G | A |
| HDL-C | 2 | 21123352 | C | A |
| HDL-C | 2 | 21134011 | T | C |
| HDL-C | 2 | 21143982 | T | C |
| HDL-C | 2 | 21148274 | C | A |
| HDL-C | 2 | 21198900 | G | A |

**Table S14:** Genetic variants used in the presented analyses. Note that due to coverage of the GWAS chip the instruments differed across considered outcomes *(continued)*

| Exposure | Chromosome | Base-pair position (Build 37) | Other allele | Effect allele |
|---|---|---|---|---|
| HDL-C | 2 | 21208211 | G | A |
| HDL-C | 2 | 21240031 | C | A |
| HDL-C | 2 | 21240364 | G | A |
| HDL-C | 2 | 21245329 | T | G |
| HDL-C | 2 | 21267782 | T | C |
| HDL-C | 2 | 21269809 | G | A |
| HDL-C | 2 | 227105921 | T | C |
| HDL-C | 2 | 227159867 | G | A |
| HDL-C | 4 | 103188709 | T | C |
| HDL-C | 4 | 103198082 | G | A |
| HDL-C | 4 | 69450464 | T | C |
| HDL-C | 8 | 19554301 | T | C |
| HDL-C | 8 | 19662458 | G | A |
| HDL-C | 8 | 19734951 | G | A |
| HDL-C | 8 | 19737157 | G | A |
| HDL-C | 8 | 19755175 | C | A |
| HDL-C | 8 | 19756592 | T | C |
| HDL-C | 8 | 19774890 | C | A |
| HDL-C | 8 | 19776981 | C | A |
| HDL-C | 8 | 19777695 | T | C |
| HDL-C | 8 | 19786891 | T | C |
| HDL-C | 8 | 19791071 | T | C |
| HDL-C | 8 | 19791346 | T | C |
| HDL-C | 8 | 19793581 | T | C |
| HDL-C | 8 | 19801530 | T | G |
| HDL-C | 8 | 19803093 | T | C |
| HDL-C | 8 | 19810787 | C | A |
| HDL-C | 8 | 19811901 | T | C |
| HDL-C | 8 | 19812812 | T | C |
| HDL-C | 8 | 19813180 | G | A |
| HDL-C | 8 | 19813529 | G | A |
| HDL-C | 8 | 19815098 | T | C |
| HDL-C | 8 | 19815189 | T | C |
| HDL-C | 8 | 19815556 | G | A |
| HDL-C | 8 | 19816078 | TA | T |
| HDL-C | 8 | 19816934 | T | C |
| HDL-C | 8 | 19818042 | G | A |
| HDL-C | 8 | 19819439 | G | A |
| HDL-C | 8 | 19820405 | G | A |
| HDL-C | 8 | 19824626 | CTT | C |
| HDL-C | 8 | 19826373 | G | A |
| HDL-C | 8 | 19827848 | C | A |
| HDL-C | 8 | 19830170 | T | G |
| HDL-C | 8 | 19830921 | T | C |

**Table S14:** Genetic variants used in the presented analyses. Note that due to coverage of the GWAS chip the instruments differed across considered outcomes *(continued)*

| Exposure | Chromosome | Base-pair position (Build 37) | Other allele | Effect allele |
|---|---|---|---|---|
| HDL-C | 8 | 19838469 | T | C |
| HDL-C | 8 | 19839709 | T | C |
| HDL-C | 8 | 19843861 | G | A |
| HDL-C | 8 | 19843879 | T | C |
| HDL-C | 8 | 19844222 | G | A |
| HDL-C | 8 | 19849757 | G | A |
| HDL-C | 8 | 19849992 | T | C |
| HDL-C | 8 | 19852156 | T | G |
| HDL-C | 8 | 19852176 | TTC | T |
| HDL-C | 8 | 19855844 | GTATTTT | G |
| HDL-C | 8 | 19855853 | TTTTTA | T |
| HDL-C | 8 | 19856539 | T | C |
| HDL-C | 8 | 19862746 | G | A |
| HDL-C | 8 | 19865175 | G | A |
| HDL-C | 8 | 19865843 | G | A |
| HDL-C | 8 | 19866631 | G | A |
| HDL-C | 8 | 19867853 | G | A |
| HDL-C | 8 | 19868386 | C | A |
| HDL-C | 8 | 19868971 | T | C |
| HDL-C | 8 | 19870271 | C | A |
| HDL-C | 8 | 19870693 | G | A |
| HDL-C | 8 | 19870735 | G | A |
| HDL-C | 8 | 19871080 | G | A |
| HDL-C | 8 | 19871113 | T | C |
| HDL-C | 8 | 19874932 | AAGAC | A |
| HDL-C | 8 | 19875100 | T | C |
| HDL-C | 8 | 19876301 | GA | G |
| HDL-C | 8 | 19878356 | T | C |
| HDL-C | 8 | 19882950 | T | C |
| HDL-C | 8 | 19885726 | T | C |
| HDL-C | 8 | 19888586 | T | C |
| HDL-C | 8 | 19889125 | T | G |
| HDL-C | 8 | 19889399 | T | G |
| HDL-C | 8 | 19890612 | G | A |
| HDL-C | 8 | 19890776 | C | A |
| HDL-C | 8 | 19894504 | T | C |
| HDL-C | 8 | 19899046 | T | C |
| HDL-C | 8 | 19912370 | G | A |
| HDL-C | 8 | 19914598 | C | A |
| HDL-C | 8 | 19922610 | G | A |
| HDL-C | 8 | 19928160 | G | A |
| HDL-C | 8 | 19928582 | G | A |
| HDL-C | 8 | 19931193 | T | C |
| HDL-C | 8 | 19932088 | T | C |

**Table S14:** Genetic variants used in the presented analyses. Note that due to coverage of the GWAS chip the instruments differed across considered outcomes *(continued)*

| Exposure | Chromosome | Base-pair position (Build 37) | Other allele | Effect allele |
|---|---|---|---|---|
| HDL-C | 8 | 19933896 | T | C |
| HDL-C | 8 | 19934339 | T | C |
| HDL-C | 8 | 19936687 | G | A |
| HDL-C | 8 | 19938922 | T | C |
| HDL-C | 8 | 19938946 | T | C |
| HDL-C | 8 | 19939049 | T | C |
| HDL-C | 8 | 19941145 | C | A |
| HDL-C | 8 | 19941448 | T | C |
| HDL-C | 8 | 19942181 | T | G |
| HDL-C | 8 | 19942806 | G | A |
| HDL-C | 8 | 19943027 | G | A |
| HDL-C | 8 | 19943308 | G | A |
| HDL-C | 8 | 9177732 | T | G |
| HDL-C | 8 | 9179298 | T | G |
| HDL-C | 8 | 9181611 | G | A |
| HDL-C | 8 | 9183596 | G | A |
| HDL-C | 8 | 9184691 | T | C |
| HDL-C | 8 | 9187242 | G | A |
| HDL-C | 8 | 9190564 | T | C |
| HDL-C | 8 | 9192423 | G | A |
| HDL-C | 8 | 9201787 | T | C |
| HDL-C | 8 | 9206069 | G | A |
| HDL-C | 8 | 9206678 | G | A |
| HDL-C | 8 | 9219868 | G | A |
| HDL-C | 8 | 9219869 | G | A |
| HDL-C | 9 | 107629336 | T | C |
| HDL-C | 9 | 107635869 | T | C |
| HDL-C | 9 | 107636707 | C | A |
| HDL-C | 9 | 107639180 | G | A |
| HDL-C | 9 | 107642287 | T | C |
| HDL-C | 9 | 107650345 | T | G |
| HDL-C | 9 | 107650355 | T | G |
| HDL-C | 9 | 107651526 | G | A |
| HDL-C | 9 | 107655426 | T | G |
| HDL-C | 9 | 107657070 | G | A |
| HDL-C | 9 | 107658910 | C | A |
| HDL-C | 9 | 107661742 | C | A |
| HDL-C | 9 | 107664301 | T | C |
| HDL-C | 9 | 107665739 | G | A |
| HDL-C | 9 | 107666414 | T | C |
| HDL-C | 9 | 107669073 | T | C |
| HDL-C | 9 | 107669241 | T | C |
| HDL-C | 9 | 107672526 | T | G |
| HDL-C | 9 | 117138278 | T | C |

**Table S14:** Genetic variants used in the presented analyses. Note that due to coverage of the GWAS chip the instruments differed across considered outcomes *(continued)*

| Exposure | Chromosome | Base-pair position (Build 37) | Other allele | Effect allele |
|---|---|---|---|---|
| HDL-C | 9 | 15285329 | T | C |
| HDL-C | 9 | 15289578 | T | C |
| HDL-C | 9 | 15293513 | T | C |
| HDL-C | 9 | 15296034 | C | A |
| HDL-C | 9 | 15298666 | T | C |
| TG | 10 | 65340897 | G | A |
| TG | 11 | 116489979 | T | C |
| TG | 11 | 116493950 | T | G |
| TG | 11 | 116496360 | T | C |
| TG | 11 | 116499500 | G | A |
| TG | 11 | 116511028 | G | A |
| TG | 11 | 116515557 | T | C |
| TG | 11 | 116519655 | T | C |
| TG | 11 | 116519739 | T | C |
| TG | 11 | 116520398 | T | C |
| TG | 11 | 116522223 | CT | C |
| TG | 11 | 116522854 | G | A |
| TG | 11 | 116522961 | T | C |
| TG | 11 | 116522976 | AAGTGG | A |
| TG | 11 | 116524569 | CTT | C |
| TG | 11 | 116525867 | T | C |
| TG | 11 | 116526032 | CCT | C |
| TG | 11 | 116526322 | C | A |
| TG | 11 | 116527728 | G | A |
| TG | 11 | 116528544 | T | C |
| TG | 11 | 116528876 | T | C |
| TG | 11 | 116536418 | G | A |
| TG | 11 | 116552562 | T | C |
| TG | 11 | 116553025 | G | A |
| TG | 11 | 116555033 | TA | T |
| TG | 11 | 116557248 | T | C |
| TG | 11 | 116558427 | G | A |
| TG | 11 | 116559002 | T | C |
| TG | 11 | 116559553 | T | G |
| TG | 11 | 116560447 | T | C |
| TG | 11 | 116560667 | T | G |
| TG | 11 | 116561996 | T | C |
| TG | 11 | 116562580 | C | A |
| TG | 11 | 116567619 | T | C |
| TG | 11 | 116568168 | G | A |
| TG | 11 | 116570942 | G | A |
| TG | 11 | 116571284 | T | C |
| TG | 11 | 116571608 | G | A |
| TG | 11 | 116572089 | T | C |

**Table S14:** Genetic variants used in the presented analyses. Note that due to coverage of the GWAS chip the instruments differed across considered outcomes *(continued)*

| Exposure | Chromosome | Base-pair position (Build 37) | Other allele | Effect allele |
|---|---|---|---|---|
| TG | 11 | 116574488 | G | A |
| TG | 11 | 116576400 | T | C |
| TG | 11 | 116576415 | G | A |
| TG | 11 | 116577441 | G | A |
| TG | 11 | 116578982 | G | A |
| TG | 11 | 116579663 | T | C |
| TG | 11 | 116580816 | T | C |
| TG | 11 | 116581259 | T | C |
| TG | 11 | 116583017 | G | A |
| TG | 11 | 116584987 | T | C |
| TG | 11 | 116585533 | G | A |
| TG | 11 | 116586283 | T | C |
| TG | 11 | 116586423 | T | C |
| TG | 11 | 116590787 | G | A |
| TG | 11 | 116592203 | G | A |
| TG | 11 | 116595756 | T | C |
| TG | 11 | 116596309 | C | A |
| TG | 11 | 116598315 | T | C |
| TG | 11 | 116598988 | G | A |
| TG | 11 | 116599392 | T | C |
| TG | 11 | 116601945 | G | A |
| TG | 11 | 116603677 | G | A |
| TG | 11 | 116603724 | C | A |
| TG | 11 | 116604449 | AGGCTGG | A |
| TG | 11 | 116606766 | G | A |
| TG | 11 | 116607368 | C | A |
| TG | 11 | 116610249 | G | A |
| TG | 11 | 116613660 | T | C |
| TG | 11 | 116613840 | CTAAA | C |
| TG | 11 | 116619605 | G | A |
| TG | 11 | 116620256 | C | A |
| TG | 11 | 116622722 | T | C |
| TG | 11 | 116623659 | T | C |
| TG | 11 | 116624601 | C | A |
| TG | 11 | 116624703 | T | G |
| TG | 11 | 116625875 | T | G |
| TG | 11 | 116631707 | T | C |
| TG | 11 | 116632798 | T | C |
| TG | 11 | 116634173 | T | C |
| TG | 11 | 116639104 | G | A |
| TG | 11 | 116639692 | T | C |
| TG | 11 | 116641028 | T | C |
| TG | 11 | 116645275 | T | C |
| TG | 11 | 116649135 | G | A |

**Table S14:** Genetic variants used in the presented analyses. Note that due to coverage of the GWAS chip the instruments differed across considered outcomes *(continued)*

| Exposure | Chromosome | Base-pair position (Build 37) | Other allele | Effect allele |
|---|---|---|---|---|
| TG | 11 | 116650666 | CTAGT | C |
| TG | 11 | 116651115 | T | C |
| TG | 11 | 116651463 | T | G |
| TG | 11 | 116652301 | T | C |
| TG | 11 | 116654159 | G | A |
| TG | 11 | 116657561 | T | C |
| TG | 11 | 116657590 | CA | C |
| TG | 11 | 116660686 | G | A |
| TG | 11 | 116660813 | G | A |
| TG | 11 | 116662579 | T | C |
| TG | 11 | 116664040 | G | A |
| TG | 11 | 116666099 | T | C |
| TG | 11 | 116667083 | G | A |
| TG | 11 | 116668511 | T | C |
| TG | 11 | 116669828 | G | A |
| TG | 11 | 116670676 | C | A |
| TG | 11 | 116670738 | G | A |
| TG | 11 | 116673149 | T | C |
| TG | 11 | 116673315 | T | G |
| TG | 11 | 116673464 | T | C |
| TG | 11 | 116674818 | G | A |
| TG | 11 | 116684028 | T | C |
| TG | 11 | 116688765 | C | A |
| TG | 11 | 116691480 | G | A |
| TG | 11 | 116692334 | T | C |
| TG | 11 | 116694938 | G | A |
| TG | 11 | 116699389 | G | A |
| TG | 11 | 116700142 | T | C |
| TG | 11 | 116701535 | T | C |
| TG | 11 | 116701850 | T | C |
| TG | 11 | 116703146 | T | C |
| TG | 11 | 116703671 | T | G |
| TG | 11 | 116704178 | C | A |
| TG | 11 | 116705516 | G | A |
| TG | 11 | 116705568 | T | G |
| TG | 11 | 116707338 | G | A |
| TG | 11 | 116707583 | G | A |
| TG | 11 | 116707684 | G | A |
| TG | 11 | 116708020 | G | A |
| TG | 11 | 116710167 | CAT | C |
| TG | 11 | 116710968 | G | A |
| TG | 11 | 116711141 | T | C |
| TG | 11 | 116712615 | G | A |
| TG | 11 | 116718858 | G | A |

**Table S14:** Genetic variants used in the presented analyses. Note that due to coverage of the GWAS chip the instruments differed across considered outcomes *(continued)*

| Exposure | Chromosome | Base-pair position (Build 37) | Other allele | Effect allele |
|---|---|---|---|---|
| TG | 11 | 116720089 | T | C |
| TG | 11 | 116722041 | T | G |
| TG | 11 | 116730638 | G | A |
| TG | 11 | 116732512 | T | G |
| TG | 11 | 116736679 | G | A |
| TG | 11 | 116741572 | C | A |
| TG | 11 | 116758101 | G | A |
| TG | 11 | 116763034 | G | A |
| TG | 11 | 116764021 | T | C |
| TG | 11 | 116772441 | T | C |
| TG | 11 | 116782202 | G | A |
| TG | 11 | 116784824 | C | A |
| TG | 11 | 116807970 | G | A |
| TG | 11 | 116864990 | G | A |
| TG | 11 | 116876621 | T | C |
| TG | 11 | 116896155 | C | A |
| TG | 11 | 116908283 | G | A |
| TG | 11 | 116969390 | T | C |
| TG | 11 | 116972894 | C | A |
| TG | 11 | 116979689 | T | C |
| TG | 11 | 116987046 | G | A |
| TG | 11 | 116990760 | GAAAC | G |
| TG | 11 | 116990851 | G | A |
| TG | 11 | 116991926 | G | A |
| TG | 11 | 116996539 | G | A |
| TG | 11 | 116998879 | T | C |
| TG | 11 | 117004656 | T | C |
| TG | 11 | 117005627 | T | C |
| TG | 11 | 117022352 | C | A |
| TG | 11 | 117026374 | C | A |
| TG | 11 | 117027183 | G | A |
| TG | 11 | 117042377 | G | A |
| TG | 11 | 117045335 | T | C |
| TG | 11 | 117045431 | T | G |
| TG | 11 | 117048367 | T | C |
| TG | 11 | 117049508 | G | A |
| TG | 11 | 117053675 | G | A |
| TG | 11 | 117059347 | T | C |
| TG | 11 | 117062370 | T | C |
| TG | 11 | 117075566 | T | C |
| TG | 11 | 117089628 | G | A |
| TG | 11 | 117141469 | T | C |
| TG | 11 | 117175658 | G | A |
| TG | 12 | 124380047 | G | A |

**Table S14:** Genetic variants used in the presented analyses. Note that due to coverage of the GWAS chip the instruments differed across considered outcomes *(continued)*

| Exposure | Chromosome | Base-pair position (Build 37) | Other allele | Effect allele |
|---|---|---|---|---|
| TG | 15 | 58718998 | T | C |
| TG | 15 | 58719741 | G | A |
| TG | 15 | 58723675 | T | C |
| TG | 15 | 58727325 | G | A |
| TG | 15 | 58730498 | T | C |
| TG | 15 | 58731567 | G | A |
| TG | 15 | 58740094 | T | C |
| TG | 15 | 58742418 | G | A |
| TG | 15 | 58742731 | T | G |
| TG | 16 | 56985514 | G | A |
| TG | 16 | 56986762 | T | G |
| TG | 16 | 56987765 | T | C |
| TG | 16 | 56988044 | T | C |
| TG | 16 | 56990716 | C | A |
| TG | 16 | 56993324 | C | A |
| TG | 16 | 56994244 | TA | T |
| TG | 16 | 56995236 | C | A |
| TG | 16 | 56996211 | G | A |
| TG | 16 | 56997233 | G | A |
| TG | 16 | 56999328 | T | C |
| TG | 16 | 57000885 | G | A |
| TG | 16 | 57001216 | T | C |
| TG | 16 | 57001274 | AC | A |
| TG | 16 | 57001581 | AAAC | A |
| TG | 16 | 57002732 | T | G |
| TG | 16 | 57005479 | C | A |
| TG | 16 | 57006590 | T | C |
| TG | 17 | 41840849 | T | C |
| TG | 17 | 41926126 | T | C |
| TG | 19 | 11185919 | T | C |
| TG | 19 | 11186711 | T | C |
| TG | 19 | 11190544 | T | C |
| TG | 19 | 11195030 | G | A |
| TG | 19 | 11196356 | AC | A |
| TG | 19 | 11196886 | T | C |
| TG | 19 | 11197750 | GA | G |
| TG | 19 | 11198187 | T | C |
| TG | 19 | 11202306 | T | G |
| TG | 19 | 19083483 | G | A |
| TG | 19 | 19104843 | C | A |
| TG | 19 | 19130750 | G | A |
| TG | 19 | 19207229 | G | A |
| TG | 19 | 19262300 | T | C |
| TG | 19 | 19262698 | G | A |

**Table S14:** Genetic variants used in the presented analyses. Note that due to coverage of the GWAS chip the instruments differed across considered outcomes *(continued)*

| Exposure | Chromosome | Base-pair position (Build 37) | Other allele | Effect allele |
|---|---|---|---|---|
| TG | 19 | 19268316 | G | A |
| TG | 19 | 19274869 | G | A |
| TG | 19 | 19283268 | T | C |
| TG | 19 | 19295875 | G | A |
| TG | 19 | 19297355 | G | A |
| TG | 19 | 19325963 | T | G |
| TG | 19 | 19329924 | T | C |
| TG | 19 | 19336608 | T | C |
| TG | 19 | 19366632 | T | C |
| TG | 19 | 19374061 | T | C |
| TG | 19 | 19380513 | G | A |
| TG | 19 | 19393714 | G | A |
| TG | 19 | 19394368 | G | A |
| TG | 19 | 19406869 | T | C |
| TG | 19 | 19407718 | T | C |
| TG | 19 | 19418916 | T | C |
| TG | 19 | 19432290 | AG | A |
| TG | 19 | 19456917 | T | C |
| TG | 19 | 19458388 | G | A |
| TG | 19 | 19461437 | T | C |
| TG | 19 | 19467545 | T | C |
| TG | 19 | 19479497 | T | C |
| TG | 19 | 19494483 | G | A |
| TG | 19 | 19503573 | T | C |
| TG | 19 | 19517169 | G | A |
| TG | 19 | 19525792 | T | C |
| TG | 19 | 19531910 | G | A |
| TG | 19 | 19539891 | T | C |
| TG | 19 | 19544644 | T | C |
| TG | 19 | 19610596 | T | C |
| TG | 19 | 19662220 | T | C |
| TG | 19 | 19685470 | GT | G |
| TG | 19 | 19695228 | T | C |
| TG | 19 | 19721722 | G | A |
| TG | 19 | 19746151 | T | G |
| TG | 19 | 19789528 | G | A |
| TG | 19 | 19819843 | G | A |
| TG | 19 | 19892796 | G | A |
| TG | 19 | 19919422 | G | A |
| TG | 19 | 20100542 | G | A |
| TG | 19 | 20112264 | T | C |
| TG | 19 | 20152519 | T | G |
| TG | 19 | 20162400 | C | A |
| TG | 19 | 20162695 | C | A |

**Table S14:** Genetic variants used in the presented analyses. Note that due to coverage of the GWAS chip the instruments differed across considered outcomes *(continued)*

| Exposure | Chromosome | Base-pair position (Build 37) | Other allele | Effect allele |
|---|---|---|---|---|
| TG | 19 | 20175252 | G | A |
| TG | 19 | 20217169 | T | C |
| TG | 19 | 20495521 | G | A |
| TG | 19 | 23124745 | C | A |
| TG | 19 | 23126236 | T | C |
| TG | 19 | 23198437 | G | A |
| TG | 19 | 23228747 | T | G |
| TG | 19 | 23231399 | G | A |
| TG | 19 | 45386467 | GTAA | G |
| TG | 19 | 45388500 | G | A |
| TG | 19 | 45392254 | T | C |
| TG | 19 | 45394336 | T | C |
| TG | 19 | 45395266 | G | A |
| TG | 19 | 45395619 | G | A |
| TG | 19 | 45395714 | T | C |
| TG | 19 | 45395844 | G | A |
| TG | 19 | 45396219 | T | C |
| TG | 19 | 45404691 | G | A |
| TG | 19 | 45406673 | G | A |
| TG | 19 | 45410002 | G | A |
| TG | 19 | 45411941 | T | C |
| TG | 19 | 45413576 | T | C |
| TG | 19 | 45414451 | T | C |
| TG | 19 | 45416478 | G | A |
| TG | 19 | 45417638 | CTTCG | C |
| TG | 19 | 45421254 | G | A |
| TG | 19 | 45421877 | G | A |
| TG | 19 | 45422946 | G | A |
| TG | 19 | 45425175 | T | G |
| TG | 19 | 45428234 | G | A |
| TG | 19 | 45431658 | G | A |
| TG | 19 | 45443504 | G | A |
| TG | 19 | 45448465 | T | G |
| TG | 19 | 45466299 | CA | C |
| TG | 19 | 45467788 | TAACTC | T |
| TG | 19 | 45483438 | C | A |
| TG | 19 | 45493061 | T | C |
| TG | 19 | 45499388 | T | C |
| TG | 19 | 8429323 | G | A |
| TG | 19 | 8579319 | ATT | A |
| TG | 19 | 8582702 | GC | G |
| TG | 19 | 8596372 | ACAG | A |
| TG | 19 | 8627569 | T | C |
| TG | 1 | 230280696 | T | C |

**Table S14:** Genetic variants used in the presented analyses. Note that due to coverage of the GWAS chip the instruments differed across considered outcomes *(continued)*

| Exposure | Chromosome | Base-pair position (Build 37) | Other allele | Effect allele |
|---|---|---|---|---|
| TG | 1 | 230285056 | G | A |
| TG | 1 | 230290254 | G | A |
| TG | 1 | 230291868 | T | C |
| TG | 1 | 230294989 | C | A |
| TG | 1 | 230295691 | G | A |
| TG | 1 | 230296492 | G | A |
| TG | 1 | 230300481 | T | C |
| TG | 1 | 230302521 | T | G |
| TG | 1 | 230302661 | G | A |
| TG | 1 | 230304914 | G | A |
| TG | 1 | 230304988 | G | A |
| TG | 1 | 230305312 | G | A |
| TG | 1 | 230305477 | T | C |
| TG | 1 | 230309771 | T | C |
| TG | 1 | 230326412 | G | A |
| TG | 1 | 230402624 | G | A |
| TG | 1 | 55505647 | T | G |
| TG | 1 | 55636240 | T | C |
| TG | 1 | 62900811 | G | A |
| TG | 1 | 62901807 | C | A |
| TG | 1 | 62913693 | G | A |
| TG | 1 | 62960250 | T | C |
| TG | 1 | 62960335 | TG | T |
| TG | 1 | 62960575 | T | C |
| TG | 1 | 62988972 | T | C |
| TG | 1 | 62999110 | AT | A |
| TG | 1 | 63018852 | T | C |
| TG | 1 | 63025942 | T | G |
| TG | 1 | 63028198 | T | C |
| TG | 1 | 63034363 | C | A |
| TG | 1 | 63034646 | C | A |
| TG | 1 | 63048276 | C | A |
| TG | 1 | 63085941 | G | A |
| TG | 1 | 63117328 | G | A |
| TG | 1 | 63119256 | T | C |
| TG | 1 | 63135127 | G | A |
| TG | 1 | 63158722 | T | C |
| TG | 1 | 63165761 | G | A |
| TG | 1 | 63167984 | C | A |
| TG | 1 | 63177365 | T | C |
| TG | 1 | 63191577 | C | A |
| TG | 1 | 63191777 | T | C |
| TG | 1 | 63193972 | G | A |
| TG | 1 | 63195692 | T | C |

**Table S14:** Genetic variants used in the presented analyses. Note that due to coverage of the GWAS chip the instruments differed across considered outcomes *(continued)*

| Exposure | Chromosome | Base-pair position (Build 37) | Other allele | Effect allele |
|---|---|---|---|---|
| TG | 1 | 63199347 | TG | T |
| TG | 1 | 63203221 | T | C |
| TG | 1 | 63211706 | T | C |
| TG | 1 | 63248232 | TA | T |
| TG | 1 | 63310444 | G | A |
| TG | 1 | 63316517 | G | A |
| TG | 1 | 63332527 | ATC | A |
| TG | 1 | 63354972 | G | A |
| TG | 20 | 44534651 | G | A |
| TG | 20 | 44551855 | T | C |
| TG | 2 | 21074998 | G | A |
| TG | 2 | 21121464 | T | C |
| TG | 2 | 21123352 | C | A |
| TG | 2 | 21134011 | T | C |
| TG | 2 | 21135577 | G | A |
| TG | 2 | 21138066 | T | C |
| TG | 2 | 21140540 | T | C |
| TG | 2 | 21142994 | T | C |
| TG | 2 | 21145665 | T | C |
| TG | 2 | 21145932 | AC | A |
| TG | 2 | 21148274 | C | A |
| TG | 2 | 21148888 | GA | G |
| TG | 2 | 21150787 | T | C |
| TG | 2 | 21152239 | TG | T |
| TG | 2 | 21184764 | G | A |
| TG | 2 | 21186121 | C | A |
| TG | 2 | 21187641 | T | C |
| TG | 2 | 21188488 | T | C |
| TG | 2 | 21190024 | T | C |
| TG | 2 | 21190107 | T | G |
| TG | 2 | 21190209 | T | C |
| TG | 2 | 21205920 | T | C |
| TG | 2 | 21208211 | G | A |
| TG | 2 | 21216815 | C | A |
| TG | 2 | 21217490 | T | C |
| TG | 2 | 21231524 | G | A |
| TG | 2 | 21232804 | G | A |
| TG | 2 | 21233972 | T | C |
| TG | 2 | 21239667 | T | C |
| TG | 2 | 21240031 | C | A |
| TG | 2 | 21240364 | G | A |
| TG | 2 | 21244000 | T | G |
| TG | 2 | 21245329 | T | G |
| TG | 2 | 21250914 | G | A |

**Table S14:** Genetic variants used in the presented analyses. Note that due to coverage of the GWAS chip the instruments differed across considered outcomes *(continued)*

| Exposure | Chromosome | Base-pair position (Build 37) | Other allele | Effect allele |
|---|---|---|---|---|
| TG | 2 | 21259562 | T | C |
| TG | 2 | 21259612 | G | A |
| TG | 2 | 21262427 | G | A |
| TG | 2 | 21263900 | G | A |
| TG | 2 | 21266774 | GGCAGCGCCA | G |
| TG | 2 | 21267461 | G | A |
| TG | 2 | 21300770 | C | A |
| TG | 2 | 21311541 | T | G |
| TG | 2 | 21345292 | G | A |
| TG | 2 | 21371068 | T | C |
| TG | 2 | 21381177 | G | A |
| TG | 2 | 21383353 | T | C |
| TG | 2 | 21389659 | T | G |
| TG | 2 | 21471603 | T | C |
| TG | 2 | 237756139 | G | A |
| TG | 2 | 23794857 | G | A |
| TG | 2 | 27444546 | T | C |
| TG | 2 | 27450763 | T | G |
| TG | 2 | 27493045 | T | G |
| TG | 2 | 27505758 | T | C |
| TG | 2 | 27525263 | G | A |
| TG | 2 | 27550967 | G | A |
| TG | 2 | 27556721 | G | A |
| TG | 2 | 27558252 | G | A |
| TG | 2 | 27574159 | T | C |
| TG | 2 | 27591804 | T | C |
| TG | 2 | 27598097 | T | C |
| TG | 2 | 27605983 | G | A |
| TG | 2 | 27635463 | G | A |
| TG | 2 | 27642734 | C | A |
| TG | 2 | 27649929 | T | G |
| TG | 2 | 27651693 | T | C |
| TG | 2 | 27685388 | G | A |
| TG | 2 | 27686480 | G | A |
| TG | 2 | 27697953 | CAGA | C |
| TG | 2 | 27702026 | C | A |
| TG | 2 | 27703495 | G | A |
| TG | 2 | 27706640 | T | C |
| TG | 2 | 27711631 | T | C |
| TG | 2 | 27711893 | G | A |
| TG | 2 | 27730940 | T | C |
| TG | 2 | 27731212 | T | C |
| TG | 2 | 27735033 | G | A |
| TG | 2 | 27736389 | T | C |

**Table S14:** Genetic variants used in the presented analyses. Note that due to coverage of the GWAS chip the instruments differed across considered outcomes *(continued)*

| Exposure | Chromosome | Base-pair position (Build 37) | Other allele | Effect allele |
|---|---|---|---|---|
| TG | 2 | 27741105 | G | A |
| TG | 2 | 27742603 | T | C |
| TG | 2 | 27743154 | G | A |
| TG | 2 | 27743215 | G | A |
| TG | 2 | 27748597 | C | A |
| TG | 2 | 27748624 | G | A |
| TG | 2 | 27750545 | G | A |
| TG | 2 | 27751562 | C | A |
| TG | 2 | 27752296 | G | A |
| TG | 2 | 27752463 | G | A |
| TG | 2 | 27752871 | T | C |
| TG | 2 | 27753878 | C | A |
| TG | 2 | 27767488 | GC | G |
| TG | 2 | 27839369 | T | C |
| TG | 2 | 27839539 | T | C |
| TG | 2 | 27851918 | G | A |
| TG | 2 | 27860258 | G | A |
| TG | 2 | 27867573 | GT | G |
| TG | 2 | 27872312 | AATAT | A |
| TG | 2 | 27895073 | T | C |
| TG | 2 | 27928797 | T | C |
| TG | 2 | 27929023 | C | A |
| TG | 2 | 27934731 | G | A |
| TG | 2 | 27987046 | C | A |
| TG | 2 | 27988821 | G | A |
| TG | 2 | 27995781 | C | A |
| TG | 2 | 27995931 | G | A |
| TG | 2 | 28003174 | T | G |
| TG | 2 | 28066122 | T | C |
| TG | 2 | 28113911 | G | A |
| TG | 2 | 28268742 | T | C |
| TG | 2 | 28431037 | T | C |
| TG | 2 | 28443050 | T | C |
| TG | 2 | 28460985 | G | A |
| TG | 2 | 28469456 | G | A |
| TG | 5 | 74562564 | T | C |
| TG | 5 | 74567359 | G | A |
| TG | 5 | 74589385 | T | C |
| TG | 5 | 74596682 | T | G |
| TG | 5 | 74602699 | C | A |
| TG | 5 | 74625487 | C | A |
| TG | 5 | 74656539 | T | C |
| TG | 5 | 74879890 | G | A |
| TG | 6 | 160848743 | G | A |

**Table S14:** Genetic variants used in the presented analyses. Note that due to coverage of the GWAS chip the instruments differed across considered outcomes *(continued)*

| Exposure | Chromosome | Base-pair position (Build 37) | Other allele | Effect allele |
|---|---|---|---|---|
| TG | 6 | 160853746 | C | A |
| TG | 6 | 31847012 | C | A |
| TG | 6 | 31861815 | G | A |
| TG | 6 | 43756169 | T | C |
| TG | 6 | 43758873 | G | A |
| TG | 6 | 52611118 | C | A |
| TG | 7 | 72809618 | T | C |
| TG | 7 | 72822848 | C | A |
| TG | 7 | 72823661 | C | A |
| TG | 7 | 72825125 | G | A |
| TG | 7 | 72829437 | T | G |
| TG | 7 | 72830546 | T | C |
| TG | 7 | 72839952 | T | G |
| TG | 7 | 72842267 | T | C |
| TG | 7 | 72846741 | CTTT | C |
| TG | 7 | 72850178 | T | C |
| TG | 7 | 72854549 | T | C |
| TG | 7 | 72856269 | G | A |
| TG | 7 | 72856430 | T | C |
| TG | 7 | 72857713 | CA | C |
| TG | 7 | 72864869 | G | A |
| TG | 7 | 72870211 | G | A |
| TG | 7 | 72878044 | T | G |
| TG | 7 | 72878645 | G | A |
| TG | 7 | 72881807 | T | C |
| TG | 7 | 72912032 | AAAAAAG | A |
| TG | 7 | 72934510 | G | A |
| TG | 7 | 72987354 | T | C |
| TG | 7 | 72989141 | CA | C |
| TG | 7 | 72993549 | TACTTTTACTG | TCTGGTGGAGAA |
| TG | 7 | 72999043 | G | A |
| TG | 7 | 73003027 | G | A |
| TG | 7 | 73003303 | C | A |
| TG | 7 | 73012042 | G | A |
| TG | 7 | 73018524 | T | C |
| TG | 7 | 73022933 | AAG | A |
| TG | 7 | 73024762 | TC | T |
| TG | 7 | 73026378 | T | C |
| TG | 7 | 73032823 | G | A |
| TG | 7 | 73035857 | T | C |
| TG | 7 | 73042302 | GCTTT | G |
| TG | 7 | 73042443 | T | C |
| TG | 7 | 73046758 | C | A |
| TG | 7 | 73046779 | G | A |

**Table S14:** Genetic variants used in the presented analyses. Note that due to coverage of the GWAS chip the instruments differed across considered outcomes *(continued)*

| Exposure | Chromosome | Base-pair position (Build 37) | Other allele | Effect allele |
|---|---|---|---|---|
| TG | 7 | 73047511 | T | C |
| TG | 7 | 73050599 | G | A |
| TG | 7 | 73050864 | G | A |
| TG | 7 | 73054746 | G | A |
| TG | 7 | 73055461 | C | A |
| TG | 7 | 73056750 | T | C |
| TG | 7 | 73058017 | G | A |
| TG | 7 | 73059019 | T | C |
| TG | 7 | 73059990 | T | C |
| TG | 7 | 73060006 | T | C |
| TG | 8 | 126447308 | T | C |
| TG | 8 | 126467230 | T | G |
| TG | 8 | 126474306 | G | A |
| TG | 8 | 126474356 | G | A |
| TG | 8 | 126474405 | T | G |
| TG | 8 | 126476873 | T | C |
| TG | 8 | 126478644 | G | A |
| TG | 8 | 126479362 | T | C |
| TG | 8 | 126484463 | T | C |
| TG | 8 | 126484526 | T | G |
| TG | 8 | 126486409 | G | A |
| TG | 8 | 126488930 | G | A |
| TG | 8 | 126491304 | T | C |
| TG | 8 | 126491733 | T | G |
| TG | 8 | 126495818 | T | C |
| TG | 8 | 126506694 | G | A |
| TG | 8 | 19550298 | T | C |
| TG | 8 | 19611230 | G | A |
| TG | 8 | 19611556 | T | C |
| TG | 8 | 19648079 | T | C |
| TG | 8 | 19648391 | T | C |
| TG | 8 | 19662458 | G | A |
| TG | 8 | 19664668 | T | C |
| TG | 8 | 19665214 | C | A |
| TG | 8 | 19666645 | G | A |
| TG | 8 | 19672040 | T | G |
| TG | 8 | 19673750 | T | G |
| TG | 8 | 19674033 | T | C |
| TG | 8 | 19678967 | T | C |
| TG | 8 | 19724434 | T | G |
| TG | 8 | 19734951 | G | A |
| TG | 8 | 19748448 | T | C |
| TG | 8 | 19756592 | T | C |
| TG | 8 | 19776981 | C | A |

**Table S14:** Genetic variants used in the presented analyses. Note that due to coverage of the GWAS chip the instruments differed across considered outcomes *(continued)*

| Exposure | Chromosome | Base-pair position (Build 37) | Other allele | Effect allele |
|---|---|---|---|---|
| TG | 8 | 19777695 | T | C |
| TG | 8 | 19785656 | G | A |
| TG | 8 | 19791346 | T | C |
| TG | 8 | 19793581 | T | C |
| TG | 8 | 19803093 | T | C |
| TG | 8 | 19810787 | C | A |
| TG | 8 | 19811901 | T | C |
| TG | 8 | 19812812 | T | C |
| TG | 8 | 19813180 | G | A |
| TG | 8 | 19813529 | G | A |
| TG | 8 | 19815189 | T | C |
| TG | 8 | 19816078 | TA | T |
| TG | 8 | 19816934 | T | C |
| TG | 8 | 19817401 | G | A |
| TG | 8 | 19817476 | T | G |
| TG | 8 | 19818042 | G | A |
| TG | 8 | 19819439 | G | A |
| TG | 8 | 19824626 | CTT | C |
| TG | 8 | 19826373 | G | A |
| TG | 8 | 19826992 | G | A |
| TG | 8 | 19827848 | C | A |
| TG | 8 | 19830170 | T | G |
| TG | 8 | 19830921 | T | C |
| TG | 8 | 19838469 | T | C |
| TG | 8 | 19839709 | T | C |
| TG | 8 | 19843879 | T | C |
| TG | 8 | 19844222 | G | A |
| TG | 8 | 19844447 | CT | C |
| TG | 8 | 19848396 | G | A |
| TG | 8 | 19849757 | G | A |
| TG | 8 | 19849992 | T | C |
| TG | 8 | 19852156 | T | G |
| TG | 8 | 19852176 | TTC | T |
| TG | 8 | 19854773 | T | C |
| TG | 8 | 19855844 | GTATTTT | G |
| TG | 8 | 19855853 | TTTTTA | T |
| TG | 8 | 19856260 | T | G |
| TG | 8 | 19859120 | T | C |
| TG | 8 | 19863024 | T | C |
| TG | 8 | 19865175 | G | A |
| TG | 8 | 19865843 | G | A |
| TG | 8 | 19866631 | G | A |
| TG | 8 | 19867853 | G | A |
| TG | 8 | 19868386 | C | A |

**Table S14:** Genetic variants used in the presented analyses. Note that due to coverage of the GWAS chip the instruments differed across considered outcomes *(continued)*

| Exposure | Chromosome | Base-pair position (Build 37) | Other allele | Effect allele |
|---|---|---|---|---|
| TG | 8 | 19868971 | T | C |
| TG | 8 | 19870693 | G | A |
| TG | 8 | 19870735 | G | A |
| TG | 8 | 19871080 | G | A |
| TG | 8 | 19871113 | T | C |
| TG | 8 | 19871637 | T | C |
| TG | 8 | 19874932 | AAGAC | A |
| TG | 8 | 19875100 | T | C |
| TG | 8 | 19876301 | GA | G |
| TG | 8 | 19878154 | T | C |
| TG | 8 | 19878356 | T | C |
| TG | 8 | 19881801 | G | A |
| TG | 8 | 19882950 | T | C |
| TG | 8 | 19885726 | T | C |
| TG | 8 | 19888586 | T | C |
| TG | 8 | 19889125 | T | G |
| TG | 8 | 19889399 | T | G |
| TG | 8 | 19890612 | G | A |
| TG | 8 | 19890654 | T | C |
| TG | 8 | 19890776 | C | A |
| TG | 8 | 19894504 | T | C |
| TG | 8 | 19899046 | T | C |
| TG | 8 | 19900474 | T | C |
| TG | 8 | 19912370 | G | A |
| TG | 8 | 19914598 | C | A |
| TG | 8 | 19922610 | G | A |
| TG | 8 | 19928160 | G | A |
| TG | 8 | 19928582 | G | A |
| TG | 8 | 19929197 | TC | T |
| TG | 8 | 19931193 | T | C |
| TG | 8 | 19931498 | G | A |
| TG | 8 | 19932863 | T | C |
| TG | 8 | 19933896 | T | C |
| TG | 8 | 19936687 | G | A |
| TG | 8 | 19938922 | T | C |
| TG | 8 | 19938946 | T | C |
| TG | 8 | 19939049 | T | C |
| TG | 8 | 19941145 | C | A |
| TG | 8 | 19941448 | T | C |
| TG | 8 | 19942700 | T | C |
| TG | 8 | 19942908 | C | A |
| TG | 8 | 19943308 | G | A |
| TG | 8 | 19943364 | T | C |
| TG | 8 | 19955382 | T | C |

**Table S14:** Genetic variants used in the presented analyses. Note that due to coverage of the GWAS chip the instruments differed across considered outcomes *(continued)*

| Exposure | Chromosome | Base-pair position (Build 37) | Other allele | Effect allele |
|---|---|---|---|---|
| TG | 8 | 19985025 | G | A |
| TC | 10 | 14426847 | G | A |
| TC | 10 | 14426855 | T | C |
| TC | 10 | 14427327 | C | A |
| TC | 10 | 14428187 | G | A |
| TC | 10 | 14435680 | C | A |
| TC | 11 | 116586283 | T | C |
| TC | 11 | 116607437 | T | C |
| TC | 11 | 116610249 | G | A |
| TC | 11 | 116631707 | T | C |
| TC | 11 | 116639104 | G | A |
| TC | 11 | 116651115 | T | C |
| TC | 11 | 116652423 | G | A |
| TC | 11 | 116652491 | G | A |
| TC | 11 | 116657561 | T | C |
| TC | 11 | 116660686 | G | A |
| TC | 11 | 116662579 | T | C |
| TC | 11 | 116663707 | G | A |
| TC | 11 | 116670676 | C | A |
| TC | 11 | 116671476 | T | C |
| TC | 11 | 116684164 | T | C |
| TC | 11 | 116688765 | C | A |
| TC | 11 | 116692334 | T | C |
| TC | 11 | 116694938 | G | A |
| TC | 11 | 116701535 | T | C |
| TC | 11 | 116702123 | T | C |
| TC | 11 | 116703146 | T | C |
| TC | 11 | 116703671 | T | G |
| TC | 11 | 116704178 | C | A |
| TC | 11 | 116705516 | G | A |
| TC | 11 | 116705568 | T | G |
| TC | 11 | 116707338 | G | A |
| TC | 11 | 116707583 | G | A |
| TC | 11 | 116710968 | G | A |
| TC | 11 | 116711141 | T | C |
| TC | 11 | 116730638 | G | A |
| TC | 11 | 116752163 | T | C |
| TC | 11 | 116772787 | C | A |
| TC | 11 | 116793231 | G | A |
| TC | 11 | 116908283 | G | A |
| TC | 11 | 117045335 | T | C |
| TC | 11 | 117075566 | T | C |
| TC | 11 | 61547237 | T | C |
| TC | 11 | 61580635 | T | C |

**Table S14:** Genetic variants used in the presented analyses. Note that due to coverage of the GWAS chip the instruments differed across considered outcomes *(continued)*

| Exposure | Chromosome | Base-pair position (Build 37) | Other allele | Effect allele |
|---|---|---|---|---|
| TC | 11 | 61581656 | G | A |
| TC | 11 | 61589481 | G | A |
| TC | 11 | 61597212 | T | C |
| TC | 11 | 61597972 | G | A |
| TC | 11 | 61604814 | C | A |
| TC | 11 | 61609750 | T | C |
| TC | 11 | 61621194 | G | A |
| TC | 11 | 61622227 | T | C |
| TC | 11 | 61626206 | G | A |
| TC | 11 | 61639488 | T | C |
| TC | 11 | 61641542 | T | G |
| TC | 11 | 61655305 | G | A |
| TC | 11 | 61656117 | G | A |
| TC | 11 | 61658800 | T | C |
| TC | 11 | 61663691 | G | A |
| TC | 11 | 61674118 | T | C |
| TC | 12 | 111949394 | T | C |
| TC | 15 | 58671178 | C | A |
| TC | 15 | 58671721 | G | A |
| TC | 15 | 58671765 | T | G |
| TC | 15 | 58672134 | T | C |
| TC | 15 | 58674051 | G | A |
| TC | 15 | 58674308 | G | A |
| TC | 15 | 58678512 | T | C |
| TC | 15 | 58679668 | G | A |
| TC | 15 | 58680638 | T | C |
| TC | 15 | 58680639 | T | C |
| TC | 15 | 58680643 | T | G |
| TC | 15 | 58680805 | G | A |
| TC | 15 | 58683366 | G | A |
| TC | 15 | 58683697 | T | C |
| TC | 15 | 58684057 | G | A |
| TC | 15 | 58684967 | T | G |
| TC | 15 | 58686252 | G | A |
| TC | 15 | 58686338 | G | A |
| TC | 15 | 58686939 | T | C |
| TC | 15 | 58687370 | G | A |
| TC | 15 | 58687603 | T | C |
| TC | 15 | 58687880 | T | C |
| TC | 15 | 58688477 | G | A |
| TC | 15 | 58688478 | C | A |
| TC | 15 | 58689187 | T | C |
| TC | 15 | 58690738 | C | A |
| TC | 15 | 58692148 | T | G |

**Table S14:** Genetic variants used in the presented analyses. Note that due to coverage of the GWAS chip the instruments differed across considered outcomes *(continued)*

| Exposure | Chromosome | Base-pair position (Build 37) | Other allele | Effect allele |
|---|---|---|---|---|
| TC | 15 | 58695337 | T | C |
| TC | 15 | 58696300 | T | C |
| TC | 15 | 58698243 | T | G |
| TC | 15 | 58699937 | T | C |
| TC | 15 | 58700311 | T | C |
| TC | 15 | 58702325 | G | A |
| TC | 15 | 58702864 | T | C |
| TC | 15 | 58703069 | C | A |
| TC | 15 | 58705036 | G | A |
| TC | 15 | 58705658 | AC | A |
| TC | 15 | 58706797 | G | A |
| TC | 15 | 58709182 | G | A |
| TC | 15 | 58710627 | C | A |
| TC | 15 | 58712973 | T | C |
| TC | 15 | 58718529 | CA | C |
| TC | 15 | 58719741 | G | A |
| TC | 15 | 58720431 | C | A |
| TC | 15 | 58721614 | G | A |
| TC | 15 | 58722590 | GC | G |
| TC | 15 | 58723675 | T | C |
| TC | 15 | 58724524 | G | A |
| TC | 15 | 58724706 | G | A |
| TC | 15 | 58725839 | G | A |
| TC | 15 | 58727279 | T | C |
| TC | 15 | 58727325 | G | A |
| TC | 15 | 58729144 | T | C |
| TC | 15 | 58730498 | T | C |
| TC | 15 | 58731567 | G | A |
| TC | 15 | 58735005 | G | A |
| TC | 15 | 58737341 | T | G |
| TC | 15 | 58738423 | T | C |
| TC | 15 | 58738760 | G | A |
| TC | 15 | 58741891 | G | A |
| TC | 15 | 58742418 | G | A |
| TC | 15 | 58742731 | T | G |
| TC | 15 | 58743709 | T | C |
| TC | 16 | 72052348 | G | A |
| TC | 16 | 72078989 | TG | T |
| TC | 16 | 72108093 | G | A |
| TC | 17 | 45390472 | T | C |
| TC | 17 | 45402208 | T | C |
| TC | 17 | 45404916 | T | C |
| TC | 17 | 45422628 | G | A |
| TC | 17 | 45441804 | G | A |

**Table S14:** Genetic variants used in the presented analyses. Note that due to coverage of the GWAS chip the instruments differed across considered outcomes *(continued)*

| Exposure | Chromosome | Base-pair position (Build 37) | Other allele | Effect allele |
|---|---|---|---|---|
| TC | 17 | 45443494 | G | A |
| TC | 17 | 45454883 | G | A |
| TC | 17 | 45547185 | T | C |
| TC | 17 | 45554786 | G | A |
| TC | 17 | 45698483 | G | A |
| TC | 17 | 45791123 | T | G |
| TC | 18 | 47106028 | T | C |
| TC | 19 | 10504616 | C | A |
| TC | 19 | 10633984 | G | A |
| TC | 19 | 10636904 | T | C |
| TC | 19 | 10639312 | T | C |
| TC | 19 | 10639533 | T | C |
| TC | 19 | 10640380 | G | A |
| TC | 19 | 10641291 | AG | A |
| TC | 19 | 10646814 | G | A |
| TC | 19 | 10663165 | T | C |
| TC | 19 | 10702970 | T | C |
| TC | 19 | 10703280 | G | A |
| TC | 19 | 10734951 | T | C |
| TC | 19 | 10784858 | G | A |
| TC | 19 | 10802792 | G | A |
| TC | 19 | 10813364 | T | C |
| TC | 19 | 10901440 | G | A |
| TC | 19 | 10903972 | G | A |
| TC | 19 | 10909953 | T | C |
| TC | 19 | 10916684 | T | C |
| TC | 19 | 10926952 | T | C |
| TC | 19 | 10952958 | G | A |
| TC | 19 | 10965270 | T | C |
| TC | 19 | 10967970 | G | A |
| TC | 19 | 11027626 | C | A |
| TC | 19 | 11053375 | T | C |
| TC | 19 | 11054715 | G | A |
| TC | 19 | 11085543 | T | C |
| TC | 19 | 11107133 | T | C |
| TC | 19 | 11113902 | G | A |
| TC | 19 | 11114206 | T | C |
| TC | 19 | 11114591 | T | C |
| TC | 19 | 11122710 | T | G |
| TC | 19 | 11127069 | T | G |
| TC | 19 | 11132080 | G | A |
| TC | 19 | 11136315 | T | C |
| TC | 19 | 11139636 | T | G |
| TC | 19 | 11140144 | G | A |

**Table S14:** Genetic variants used in the presented analyses. Note that due to coverage of the GWAS chip the instruments differed across considered outcomes *(continued)*

| Exposure | Chromosome | Base-pair position (Build 37) | Other allele | Effect allele |
|---|---|---|---|---|
| TC | 19 | 11147694 | T | G |
| TC | 19 | 11159076 | G | A |
| TC | 19 | 11161537 | C | A |
| TC | 19 | 11163562 | T | C |
| TC | 19 | 11163601 | T | G |
| TC | 19 | 11163897 | T | C |
| TC | 19 | 11164460 | G | A |
| TC | 19 | 11164781 | G | A |
| TC | 19 | 11166431 | CT | C |
| TC | 19 | 11169947 | T | C |
| TC | 19 | 11170839 | T | C |
| TC | 19 | 11171281 | C | A |
| TC | 19 | 11172299 | G | A |
| TC | 19 | 11174625 | G | A |
| TC | 19 | 11174742 | T | C |
| TC | 19 | 11177250 | C | A |
| TC | 19 | 11180047 | T | C |
| TC | 19 | 11183837 | G | A |
| TC | 19 | 11184250 | G | A |
| TC | 19 | 11184293 | T | C |
| TC | 19 | 11185014 | G | A |
| TC | 19 | 11185919 | T | C |
| TC | 19 | 11186711 | T | C |
| TC | 19 | 11187611 | G | A |
| TC | 19 | 11188313 | CTTTA | C |
| TC | 19 | 11188899 | T | C |
| TC | 19 | 11189298 | CG | C |
| TC | 19 | 11190544 | T | C |
| TC | 19 | 11190873 | T | C |
| TC | 19 | 11193065 | T | C |
| TC | 19 | 11195030 | G | A |
| TC | 19 | 11202306 | T | G |
| TC | 19 | 11205975 | G | A |
| TC | 19 | 11206575 | G | A |
| TC | 19 | 11207102 | T | C |
| TC | 19 | 11207516 | G | A |
| TC | 19 | 11207982 | T | C |
| TC | 19 | 11209669 | CA | C |
| TC | 19 | 11209757 | CCAAA | C |
| TC | 19 | 11210912 | T | C |
| TC | 19 | 11243445 | G | A |
| TC | 19 | 11245071 | GGAGT | G |
| TC | 19 | 11257169 | T | C |
| TC | 19 | 11262969 | T | G |

**Table S14:** Genetic variants used in the presented analyses. Note that due to coverage of the GWAS chip the instruments differed across considered outcomes *(continued)*

| Exposure | Chromosome | Base-pair position (Build 37) | Other allele | Effect allele |
|---|---|---|---|---|
| TC | 19 | 11277232 | G | A |
| TC | 19 | 11331354 | T | G |
| TC | 19 | 11350488 | T | C |
| TC | 19 | 11517032 | T | C |
| TC | 19 | 19130096 | T | C |
| TC | 19 | 19176612 | G | A |
| TC | 19 | 19329924 | T | C |
| TC | 19 | 19366632 | T | C |
| TC | 19 | 19380513 | G | A |
| TC | 19 | 19393714 | G | A |
| TC | 19 | 19394368 | G | A |
| TC | 19 | 19407718 | T | C |
| TC | 19 | 19432290 | AG | A |
| TC | 19 | 19456917 | T | C |
| TC | 19 | 19458388 | G | A |
| TC | 19 | 19479497 | T | C |
| TC | 19 | 19494483 | G | A |
| TC | 19 | 19544644 | T | C |
| TC | 19 | 19624481 | G | A |
| TC | 19 | 19658472 | T | G |
| TC | 19 | 19721722 | G | A |
| TC | 19 | 19855549 | C | A |
| TC | 19 | 19919422 | G | A |
| TC | 19 | 20217169 | T | C |
| TC | 19 | 45134682 | C | A |
| TC | 19 | 45160086 | C | A |
| TC | 19 | 45167544 | T | C |
| TC | 19 | 45176340 | T | C |
| TC | 19 | 45187163 | G | A |
| TC | 19 | 45190766 | T | C |
| TC | 19 | 45195313 | T | G |
| TC | 19 | 45196964 | G | A |
| TC | 19 | 45197027 | C | A |
| TC | 19 | 45198477 | T | C |
| TC | 19 | 45198936 | T | C |
| TC | 19 | 45220896 | T | C |
| TC | 19 | 45222982 | T | G |
| TC | 19 | 45223038 | G | A |
| TC | 19 | 45227742 | T | C |
| TC | 19 | 45232161 | G | A |
| TC | 19 | 45233329 | AAATTTATAG | A |
| TC | 19 | 45237812 | T | C |
| TC | 19 | 45239536 | C | A |
| TC | 19 | 45242173 | G | A |

**Table S14:** Genetic variants used in the presented analyses. Note that due to coverage of the GWAS chip the instruments differed across considered outcomes *(continued)*

| Exposure | Chromosome | Base-pair position (Build 37) | Other allele | Effect allele |
|---|---|---|---|---|
| TC | 19 | 45247627 | G | A |
| TC | 19 | 45249661 | T | C |
| TC | 19 | 45251156 | C | A |
| TC | 19 | 45253491 | T | C |
| TC | 19 | 45253542 | CTTTG | C |
| TC | 19 | 45253611 | ATTGGC | A |
| TC | 19 | 45276590 | GTTA | G |
| TC | 19 | 45296364 | G | A |
| TC | 19 | 45296806 | T | C |
| TC | 19 | 45302504 | G | A |
| TC | 19 | 45302778 | G | A |
| TC | 19 | 45302949 | GAA | G |
| TC | 19 | 45302951 | G | A |
| TC | 19 | 45303122 | AAAAG | A |
| TC | 19 | 45304061 | G | A |
| TC | 19 | 45315445 | G | A |
| TC | 19 | 45319631 | G | A |
| TC | 19 | 45323170 | CCATCCCCAACTCATCCT | C |
| TC | 19 | 45324138 | G | A |
| TC | 19 | 45325738 | T | G |
| TC | 19 | 45326664 | T | C |
| TC | 19 | 45327459 | T | C |
| TC | 19 | 45328380 | C | A |
| TC | 19 | 45331103 | G | A |
| TC | 19 | 45333834 | T | C |
| TC | 19 | 45335676 | G | A |
| TC | 19 | 45336035 | T | C |
| TC | 19 | 45337918 | G | A |
| TC | 19 | 45347911 | C | A |
| TC | 19 | 45351891 | G | A |
| TC | 19 | 45356141 | T | C |
| TC | 19 | 45357377 | T | C |
| TC | 19 | 45357939 | G | A |
| TC | 19 | 45361646 | G | A |
| TC | 19 | 45365447 | G | A |
| TC | 19 | 45369203 | G | A |
| TC | 19 | 45373565 | G | A |
| TC | 19 | 45377334 | G | A |
| TC | 19 | 45379060 | C | A |
| TC | 19 | 45382034 | G | A |
| TC | 19 | 45382675 | G | A |
| TC | 19 | 45384405 | T | C |
| TC | 19 | 45386467 | GTAA | G |
| TC | 19 | 45386634 | T | C |

**Table S14:** Genetic variants used in the presented analyses. Note that due to coverage of the GWAS chip the instruments differed across considered outcomes *(continued)*

| Exposure | Chromosome | Base-pair position (Build 37) | Other allele | Effect allele |
|---|---|---|---|---|
| TC | 19 | 45387034 | C | A |
| TC | 19 | 45388130 | G | A |
| TC | 19 | 45388241 | T | G |
| TC | 19 | 45388500 | G | A |
| TC | 19 | 45389596 | G | A |
| TC | 19 | 45392254 | T | C |
| TC | 19 | 45395266 | G | A |
| TC | 19 | 45395619 | G | A |
| TC | 19 | 45395714 | T | C |
| TC | 19 | 45395844 | G | A |
| TC | 19 | 45396219 | T | C |
| TC | 19 | 45396973 | G | A |
| TC | 19 | 45397952 | T | C |
| TC | 19 | 45400484 | TG | T |
| TC | 19 | 45401868 | T | C |
| TC | 19 | 45402262 | T | G |
| TC | 19 | 45408628 | T | C |
| TC | 19 | 45408836 | T | G |
| TC | 19 | 45410002 | G | A |
| TC | 19 | 45411941 | T | C |
| TC | 19 | 45412079 | T | C |
| TC | 19 | 45415640 | G | A |
| TC | 19 | 45416831 | T | G |
| TC | 19 | 45418486 | G | A |
| TC | 19 | 45421254 | G | A |
| TC | 19 | 45421650 | G | A |
| TC | 19 | 45421877 | G | A |
| TC | 19 | 45422846 | G | A |
| TC | 19 | 45422946 | G | A |
| TC | 19 | 45423944 | T | C |
| TC | 19 | 45425178 | G | A |
| TC | 19 | 45425460 | G | A |
| TC | 19 | 45426792 | G | A |
| TC | 19 | 45428234 | G | A |
| TC | 19 | 45429543 | T | C |
| TC | 19 | 45431897 | G | A |
| TC | 19 | 45435466 | CT | C |
| TC | 19 | 45436216 | G | A |
| TC | 19 | 45436657 | G | A |
| TC | 19 | 45441608 | T | C |
| TC | 19 | 45447178 | T | C |
| TC | 19 | 45447221 | G | A |
| TC | 19 | 45450252 | T | C |
| TC | 19 | 45453763 | G | A |

**Table S14:** Genetic variants used in the presented analyses. Note that due to coverage of the GWAS chip the instruments differed across considered outcomes *(continued)*

| Exposure | Chromosome | Base-pair position (Build 37) | Other allele | Effect allele |
|---|---|---|---|---|
| TC | 19 | 45478352 | G | A |
| TC | 19 | 45491032 | T | C |
| TC | 19 | 45496303 | T | C |
| TC | 19 | 45531618 | GT | G |
| TC | 19 | 45541782 | G | A |
| TC | 19 | 45653883 | TAGTC | T |
| TC | 19 | 45672478 | G | A |
| TC | 19 | 45723379 | T | C |
| TC | 19 | 45880690 | T | C |
| TC | 1 | 109757295 | G | A |
| TC | 1 | 109792735 | ACGC | A |
| TC | 1 | 109798721 | G | A |
| TC | 1 | 109806442 | C | A |
| TC | 1 | 109813485 | T | C |
| TC | 1 | 109814880 | T | C |
| TC | 1 | 109815133 | G | A |
| TC | 1 | 109815252 | G | A |
| TC | 1 | 109818158 | CT | C |
| TC | 1 | 109819296 | C | A |
| TC | 1 | 109819615 | T | C |
| TC | 1 | 109819981 | T | C |
| TC | 1 | 109820884 | G | A |
| TC | 1 | 109822166 | G | A |
| TC | 1 | 109824250 | G | A |
| TC | 1 | 109826760 | TTCCTAACC | T |
| TC | 1 | 109827253 | G | A |
| TC | 1 | 109834938 | T | C |
| TC | 1 | 109839392 | G | A |
| TC | 1 | 109839400 | T | C |
| TC | 1 | 109955569 | T | C |
| TC | 1 | 110036634 | T | C |
| TC | 1 | 220970028 | G | A |
| TC | 1 | 220970593 | T | G |
| TC | 1 | 25762805 | AT | A |
| TC | 1 | 26987646 | G | A |
| TC | 1 | 54703630 | T | C |
| TC | 1 | 54703702 | C | A |
| TC | 1 | 54708492 | G | A |
| TC | 1 | 54714821 | G | A |
| TC | 1 | 54715751 | T | C |
| TC | 1 | 54717654 | T | C |
| TC | 1 | 54721862 | TC | T |
| TC | 1 | 54722625 | T | C |
| TC | 1 | 54724389 | G | A |

**Table S14:** Genetic variants used in the presented analyses. Note that due to coverage of the GWAS chip the instruments differed across considered outcomes *(continued)*

| Exposure | Chromosome | Base-pair position (Build 37) | Other allele | Effect allele |
|---|---|---|---|---|
| TC | 1 | 54728029 | T | C |
| TC | 1 | 54769277 | T | G |
| TC | 1 | 54772869 | C | A |
| TC | 1 | 54834840 | T | C |
| TC | 1 | 54849397 | T | C |
| TC | 1 | 54880705 | T | C |
| TC | 1 | 54935900 | G | A |
| TC | 1 | 54983266 | T | C |
| TC | 1 | 55082656 | GGT | G |
| TC | 1 | 55138291 | G | A |
| TC | 1 | 55160482 | T | G |
| TC | 1 | 55224773 | C | A |
| TC | 1 | 55243587 | G | A |
| TC | 1 | 55334001 | G | A |
| TC | 1 | 55378724 | G | A |
| TC | 1 | 55411580 | G | A |
| TC | 1 | 55431933 | T | C |
| TC | 1 | 55489542 | T | C |
| TC | 1 | 55493584 | C | A |
| TC | 1 | 55494017 | T | C |
| TC | 1 | 55496039 | T | C |
| TC | 1 | 55503448 | G | A |
| TC | 1 | 55504650 | G | A |
| TC | 1 | 55505647 | T | G |
| TC | 1 | 55505861 | G | A |
| TC | 1 | 55511623 | TC | T |
| TC | 1 | 55518467 | G | A |
| TC | 1 | 55520212 | G | A |
| TC | 1 | 55521109 | G | A |
| TC | 1 | 55524601 | G | A |
| TC | 1 | 55538552 | T | G |
| TC | 1 | 55630151 | T | C |
| TC | 1 | 55636240 | T | C |
| TC | 1 | 55656075 | T | G |
| TC | 1 | 55711465 | G | A |
| TC | 1 | 55713094 | G | A |
| TC | 1 | 55713628 | G | A |
| TC | 1 | 55716497 | G | A |
| TC | 1 | 55719011 | G | A |
| TC | 1 | 55719847 | CAT | C |
| TC | 1 | 55720083 | C | A |
| TC | 1 | 55720337 | G | A |
| TC | 1 | 55723756 | C | A |
| TC | 1 | 55726519 | T | C |

**Table S14:** Genetic variants used in the presented analyses. Note that due to coverage of the GWAS chip the instruments differed across considered outcomes *(continued)*

| Exposure | Chromosome | Base-pair position (Build 37) | Other allele | Effect allele |
|---|---|---|---|---|
| TC | 1 | 55730327 | T | C |
| TC | 1 | 55731772 | TC | T |
| TC | 1 | 55732240 | T | C |
| TC | 1 | 55734875 | T | C |
| TC | 1 | 55738663 | T | C |
| TC | 1 | 55739925 | T | C |
| TC | 1 | 55740015 | T | C |
| TC | 1 | 55741377 | G | A |
| TC | 1 | 55746330 | G | A |
| TC | 1 | 55750138 | T | C |
| TC | 1 | 55752440 | T | C |
| TC | 1 | 55752699 | TA | T |
| TC | 1 | 55767547 | T | C |
| TC | 1 | 55770565 | T | C |
| TC | 1 | 55785672 | T | C |
| TC | 1 | 55785815 | T | G |
| TC | 1 | 55788281 | T | C |
| TC | 1 | 55788609 | T | C |
| TC | 1 | 55793244 | G | A |
| TC | 1 | 55798859 | T | C |
| TC | 1 | 55806465 | T | C |
| TC | 1 | 55808143 | T | C |
| TC | 1 | 55831918 | T | C |
| TC | 1 | 55837603 | G | A |
| TC | 1 | 55960656 | G | A |
| TC | 1 | 56035239 | T | C |
| TC | 1 | 56414772 | G | A |
| TC | 1 | 56608087 | T | C |
| TC | 1 | 56625395 | G | A |
| TC | 1 | 56823728 | G | A |
| TC | 1 | 56885874 | G | A |
| TC | 1 | 62900811 | G | A |
| TC | 1 | 62901807 | C | A |
| TC | 1 | 62917913 | G | A |
| TC | 1 | 62931632 | C | A |
| TC | 1 | 62934002 | G | A |
| TC | 1 | 62936912 | T | C |
| TC | 1 | 62960250 | T | C |
| TC | 1 | 62960575 | T | C |
| TC | 1 | 63048276 | C | A |
| TC | 1 | 63135127 | G | A |
| TC | 1 | 63177539 | T | G |
| TC | 1 | 63179138 | T | C |
| TC | 1 | 63193972 | G | A |

**Table S14:** Genetic variants used in the presented analyses. Note that due to coverage of the GWAS chip the instruments differed across considered outcomes *(continued)*

| Exposure | Chromosome | Base-pair position (Build 37) | Other allele | Effect allele |
|---|---|---|---|---|
| TC | 1 | 63195692 | T | C |
| TC | 1 | 63199936 | T | G |
| TC | 2 | 20896870 | CACGG | C |
| TC | 2 | 20899083 | G | A |
| TC | 2 | 20903015 | T | C |
| TC | 2 | 20928577 | G | A |
| TC | 2 | 20951740 | T | C |
| TC | 2 | 20955032 | AG | A |
| TC | 2 | 20991528 | T | G |
| TC | 2 | 21050729 | GA | G |
| TC | 2 | 21076264 | TA | T |
| TC | 2 | 21083052 | T | C |
| TC | 2 | 21085410 | T | C |
| TC | 2 | 21087323 | TCTC | T |
| TC | 2 | 21091409 | C | A |
| TC | 2 | 21092488 | T | C |
| TC | 2 | 21093112 | G | A |
| TC | 2 | 21100426 | T | C |
| TC | 2 | 21101046 | G | A |
| TC | 2 | 21107494 | T | G |
| TC | 2 | 21112689 | G | A |
| TC | 2 | 21121464 | T | C |
| TC | 2 | 21123352 | C | A |
| TC | 2 | 21126936 | G | A |
| TC | 2 | 21133883 | G | A |
| TC | 2 | 21134011 | T | C |
| TC | 2 | 21135577 | G | A |
| TC | 2 | 21135723 | T | G |
| TC | 2 | 21138066 | T | C |
| TC | 2 | 21140540 | T | C |
| TC | 2 | 21141725 | G | A |
| TC | 2 | 21142994 | T | C |
| TC | 2 | 21143677 | C | A |
| TC | 2 | 21144829 | T | G |
| TC | 2 | 21148274 | C | A |
| TC | 2 | 21150787 | T | C |
| TC | 2 | 21151917 | T | C |
| TC | 2 | 21152755 | G | A |
| TC | 2 | 21182116 | G | A |
| TC | 2 | 21182710 | G | A |
| TC | 2 | 21190024 | T | C |
| TC | 2 | 21190209 | T | C |
| TC | 2 | 21193946 | G | A |
| TC | 2 | 21196112 | G | A |

**Table S14:** Genetic variants used in the presented analyses. Note that due to coverage of the GWAS chip the instruments differed across considered outcomes *(continued)*

| Exposure | Chromosome | Base-pair position (Build 37) | Other allele | Effect allele |
|---|---|---|---|---|
| TC | 2 | 21196346 | T | G |
| TC | 2 | 21197159 | T | C |
| TC | 2 | 21202959 | C | A |
| TC | 2 | 21205920 | T | C |
| TC | 2 | 21216815 | C | A |
| TC | 2 | 21217230 | G | A |
| TC | 2 | 21217490 | T | C |
| TC | 2 | 21231524 | G | A |
| TC | 2 | 21232804 | G | A |
| TC | 2 | 21233972 | T | C |
| TC | 2 | 21239667 | T | C |
| TC | 2 | 21244000 | T | G |
| TC | 2 | 21250914 | G | A |
| TC | 2 | 21251040 | T | G |
| TC | 2 | 21259562 | T | C |
| TC | 2 | 21259612 | G | A |
| TC | 2 | 21260601 | G | A |
| TC | 2 | 21262427 | G | A |
| TC | 2 | 21263900 | G | A |
| TC | 2 | 21266774 | GGCAGCGCCA | G |
| TC | 2 | 21267210 | G | A |
| TC | 2 | 21267461 | G | A |
| TC | 2 | 21270554 | C | A |
| TC | 2 | 21270751 | G | A |
| TC | 2 | 21271323 | G | A |
| TC | 2 | 21286057 | T | C |
| TC | 2 | 21288226 | G | A |
| TC | 2 | 21288321 | G | A |
| TC | 2 | 21299935 | TATCCTGCA | T |
| TC | 2 | 21300770 | C | A |
| TC | 2 | 21303616 | T | C |
| TC | 2 | 21309335 | T | C |
| TC | 2 | 21309856 | G | A |
| TC | 2 | 21311541 | T | G |
| TC | 2 | 21313778 | TC | T |
| TC | 2 | 21323868 | CTGT | C |
| TC | 2 | 21327042 | GCTAGTGCCTA | G |
| TC | 2 | 21327635 | G | A |
| TC | 2 | 21328825 | G | A |
| TC | 2 | 21367696 | TTGAAGAC | T |
| TC | 2 | 21368217 | G | A |
| TC | 2 | 21371068 | T | C |
| TC | 2 | 21372014 | G | A |
| TC | 2 | 21374413 | T | C |

**Table S14:** Genetic variants used in the presented analyses. Note that due to coverage of the GWAS chip the instruments differed across considered outcomes *(continued)*

| Exposure | Chromosome | Base-pair position (Build 37) | Other allele | Effect allele |
|---|---|---|---|---|
| TC | 2 | 21375346 | G | A |
| TC | 2 | 21378453 | G | A |
| TC | 2 | 21381177 | G | A |
| TC | 2 | 21381292 | AAAAC | A |
| TC | 2 | 21383353 | T | C |
| TC | 2 | 21383514 | G | A |
| TC | 2 | 21389659 | T | G |
| TC | 2 | 21396092 | GAA | G |
| TC | 2 | 21397182 | G | A |
| TC | 2 | 21401902 | GAAAGAT | G |
| TC | 2 | 21406048 | T | C |
| TC | 2 | 21410890 | T | C |
| TC | 2 | 21411649 | G | A |
| TC | 2 | 21413077 | G | A |
| TC | 2 | 21414142 | T | C |
| TC | 2 | 21415728 | T | G |
| TC | 2 | 21419650 | G | A |
| TC | 2 | 21420358 | C | A |
| TC | 2 | 21422928 | T | C |
| TC | 2 | 21423532 | G | A |
| TC | 2 | 21426683 | AAAAG | A |
| TC | 2 | 21428185 | G | A |
| TC | 2 | 21434580 | G | A |
| TC | 2 | 21435700 | C | A |
| TC | 2 | 21439329 | T | G |
| TC | 2 | 21442944 | TTGCTCCCCTCTGCTTAGCA | T |
| TC | 2 | 21445569 | G | A |
| TC | 2 | 21448511 | G | A |
| TC | 2 | 21449034 | T | C |
| TC | 2 | 21449987 | G | A |
| TC | 2 | 21450714 | G | A |
| TC | 2 | 21451604 | T | C |
| TC | 2 | 21451687 | G | A |
| TC | 2 | 21453733 | G | A |
| TC | 2 | 21457324 | G | A |
| TC | 2 | 21463922 | T | C |
| TC | 2 | 21463986 | T | C |
| TC | 2 | 21467872 | ATT | A |
| TC | 2 | 21469336 | T | C |
| TC | 2 | 21471603 | T | C |
| TC | 2 | 21471750 | G | A |
| TC | 2 | 21476247 | C | A |
| TC | 2 | 21476634 | G | A |
| TC | 2 | 21524000 | G | A |

**Table S14:** Genetic variants used in the presented analyses. Note that due to coverage of the GWAS chip the instruments differed across considered outcomes *(continued)*

| Exposure | Chromosome | Base-pair position (Build 37) | Other allele | Effect allele |
|---|---|---|---|---|
| TC | 2 | 21530659 | G | A |
| TC | 2 | 21534103 | T | C |
| TC | 2 | 21534298 | T | G |
| TC | 2 | 27598097 | T | C |
| TC | 2 | 27635463 | G | A |
| TC | 2 | 27730940 | T | C |
| TC | 2 | 27742603 | T | C |
| TC | 2 | 27748624 | G | A |
| TC | 2 | 27750545 | G | A |
| TC | 2 | 27752463 | G | A |
| TC | 2 | 27752871 | T | C |
| TC | 2 | 44052743 | GTTA | G |
| TC | 2 | 44057570 | C | A |
| TC | 2 | 44073656 | T | C |
| TC | 2 | 44073881 | T | C |
| TC | 2 | 44074000 | G | A |
| TC | 2 | 44074431 | T | C |
| TC | 2 | 44081627 | T | G |
| TC | 2 | 44084975 | T | C |
| TC | 2 | 44092066 | T | C |
| TC | 2 | 44096259 | T | G |
| TC | 2 | 44096402 | G | A |
| TC | 4 | 73232228 | T | C |
| TC | 4 | 73406173 | G | A |
| TC | 4 | 73412023 | T | C |
| TC | 4 | 73542640 | T | G |
| TC | 4 | 73660066 | T | C |
| TC | 4 | 74720969 | C | A |
| TC | 4 | 74805105 | G | A |
| TC | 4 | 74970615 | C | A |
| TC | 4 | 75128036 | G | A |
| TC | 4 | 75242737 | CT | C |
| TC | 4 | 75370891 | T | C |
| TC | 4 | 75684215 | T | C |
| TC | 5 | 156380067 | T | G |
| TC | 5 | 156390297 | T | C |
| TC | 5 | 156401862 | G | A |
| TC | 5 | 156443066 | G | A |
| TC | 5 | 74302658 | G | A |
| TC | 5 | 74340327 | C | A |
| TC | 5 | 74343769 | G | A |
| TC | 5 | 74352180 | G | A |
| TC | 5 | 74354422 | G | A |
| TC | 5 | 74372141 | T | C |

**Table S14:** Genetic variants used in the presented analyses. Note that due to coverage of the GWAS chip the instruments differed across considered outcomes *(continued)*

| Exposure | Chromosome | Base-pair position (Build 37) | Other allele | Effect allele |
|---|---|---|---|---|
| TC | 5 | 74382081 | G | A |
| TC | 5 | 74384115 | G | A |
| TC | 5 | 74389325 | C | A |
| TC | 5 | 74442964 | T | C |
| TC | 5 | 74443132 | T | C |
| TC | 5 | 74446301 | G | A |
| TC | 5 | 74461850 | T | G |
| TC | 5 | 74461924 | C | A |
| TC | 5 | 74462866 | TC | T |
| TC | 5 | 74476396 | G | A |
| TC | 5 | 74477719 | TA | T |
| TC | 5 | 74491715 | TTCA | T |
| TC | 5 | 74536921 | G | A |
| TC | 5 | 74558893 | G | A |
| TC | 5 | 74562564 | T | C |
| TC | 5 | 74586866 | CT | C |
| TC | 5 | 74587940 | C | A |
| TC | 5 | 74596682 | T | G |
| TC | 5 | 74603479 | G | A |
| TC | 5 | 74604742 | G | A |
| TC | 5 | 74605220 | G | A |
| TC | 5 | 74611232 | TG | T |
| TC | 5 | 74613943 | T | C |
| TC | 5 | 74615021 | T | C |
| TC | 5 | 74622121 | TTA | T |
| TC | 5 | 74625512 | G | A |
| TC | 5 | 74629162 | CAT | C |
| TC | 5 | 74631352 | T | C |
| TC | 5 | 74643998 | ATTC | A |
| TC | 5 | 74644785 | TA | T |
| TC | 5 | 74651084 | G | A |
| TC | 5 | 74652754 | ATCCATCCGTCTGTCCG | A |
| TC | 5 | 74653462 | T | G |
| TC | 5 | 74656539 | T | C |
| TC | 5 | 74660064 | ATTG | A |
| TC | 5 | 74673707 | G | A |
| TC | 5 | 74692776 | G | A |
| TC | 5 | 74736065 | C | A |
| TC | 5 | 74736600 | TA | T |
| TC | 5 | 74745228 | T | G |
| TC | 5 | 74753736 | T | C |
| TC | 5 | 74806104 | T | C |
| TC | 5 | 74808237 | T | G |
| TC | 5 | 74808417 | G | A |

**Table S14:** Genetic variants used in the presented analyses. Note that due to coverage of the GWAS chip the instruments differed across considered outcomes *(continued)*

| Exposure | Chromosome | Base-pair position (Build 37) | Other allele | Effect allele |
|---|---|---|---|---|
| TC | 5 | 74816841 | CT | C |
| TC | 5 | 74829947 | T | C |
| TC | 5 | 74879890 | G | A |
| TC | 5 | 74892002 | T | C |
| TC | 5 | 74906963 | C | A |
| TC | 5 | 74965122 | G | A |
| TC | 5 | 74967386 | T | C |
| TC | 5 | 74971846 | G | A |
| TC | 5 | 74974434 | CT | C |
| TC | 7 | 21567331 | T | C |
| TC | 7 | 21576905 | T | C |
| TC | 7 | 21577960 | T | C |
| TC | 7 | 21582917 | T | C |
| TC | 7 | 21589243 | G | A |
| TC | 7 | 21595043 | C | A |
| TC | 7 | 21595655 | T | C |
| TC | 7 | 21607352 | T | C |
| TC | 7 | 21650595 | G | A |
| TC | 8 | 126478450 | T | C |
| TC | 8 | 126479362 | T | C |
| TC | 8 | 126484526 | T | G |
| TC | 8 | 126485531 | T | C |
| TC | 8 | 126486409 | G | A |
| TC | 8 | 126488930 | G | A |
| TC | 8 | 126493392 | G | A |
| TC | 8 | 126495818 | T | C |
| TC | 8 | 126506694 | G | A |
| TC | 8 | 144985200 | G | A |
| TC | 8 | 144986103 | T | C |
| TC | 8 | 145040108 | T | C |
| TC | 8 | 9173358 | G | A |
| TC | 8 | 9177732 | T | G |
| TC | 8 | 9179298 | T | G |
| TC | 8 | 9181395 | G | A |
| TC | 8 | 9183596 | G | A |
| TC | 8 | 9183664 | G | A |
| TC | 8 | 9184691 | T | C |
| TC | 8 | 9185146 | T | C |
| TC | 8 | 9190564 | T | C |
| TC | 8 | 9192423 | G | A |
| TC | 8 | 9201787 | T | C |
| TC | 8 | 9206069 | G | A |
| TC | 8 | 9206678 | G | A |
| TC | 8 | 9219868 | G | A |

**Table S14:** Genetic variants used in the presented analyses. Note that due to coverage of the GWAS chip the instruments differed across considered outcomes *(continued)*

| Exposure | Chromosome | Base-pair position (Build 37) | Other allele | Effect allele |
|---|---|---|---|---|
| TC | 8 | 9219869 | G | A |
| TC | 8 | 9220139 | G | A |
| TC | 8 | 9224224 | G | A |
| TC | 8 | 9224410 | T | C |
| TC | 8 | 9228403 | C | A |
| TC | 9 | 136145425 | C | A |
| TC | 9 | 136146597 | T | C |
| TC | 9 | 136149399 | G | A |
| TC | 9 | 136153875 | T | C |
| TC | 9 | 136154168 | T | C |
| TC | 9 | 136164593 | T | G |
| TC | 9 | 136184782 | G | A |
| IDL-C | 11 | 116657561 | T | C |
| IDL-C | 11 | 61547237 | T | C |
| IDL-C | 11 | 61580635 | T | C |
| IDL-C | 11 | 61581656 | G | A |
| IDL-C | 11 | 61597212 | T | C |
| IDL-C | 11 | 61597972 | G | A |
| IDL-C | 11 | 61605215 | G | A |
| IDL-C | 11 | 61609750 | T | C |
| IDL-C | 11 | 61618608 | G | A |
| IDL-C | 11 | 61619829 | C | A |
| IDL-C | 11 | 61621194 | G | A |
| IDL-C | 11 | 61622227 | T | C |
| IDL-C | 11 | 61623140 | T | C |
| IDL-C | 11 | 61627881 | T | C |
| IDL-C | 11 | 61629322 | T | C |
| IDL-C | 11 | 61629747 | G | A |
| IDL-C | 11 | 61639488 | T | C |
| IDL-C | 11 | 61641542 | T | G |
| IDL-C | 11 | 61656117 | G | A |
| IDL-C | 11 | 61656995 | T | C |
| IDL-C | 11 | 61657110 | T | C |
| IDL-C | 11 | 61657401 | C | A |
| IDL-C | 11 | 61657926 | G | A |
| IDL-C | 11 | 61663691 | G | A |
| IDL-C | 11 | 61677479 | G | A |
| IDL-C | 11 | 61678576 | T | G |
| IDL-C | 15 | 58671178 | C | A |
| IDL-C | 15 | 58671721 | G | A |
| IDL-C | 15 | 58671765 | T | G |
| IDL-C | 15 | 58672134 | T | C |
| IDL-C | 15 | 58674051 | G | A |
| IDL-C | 15 | 58674308 | G | A |

**Table S14:** Genetic variants used in the presented analyses. Note that due to coverage of the GWAS chip the instruments differed across considered outcomes *(continued)*

| Exposure | Chromosome | Base-pair position (Build 37) | Other allele | Effect allele |
|---|---|---|---|---|
| IDL-C | 15 | 58678512 | T | C |
| IDL-C | 15 | 58679668 | G | A |
| IDL-C | 15 | 58680638 | T | C |
| IDL-C | 15 | 58680639 | T | C |
| IDL-C | 15 | 58680643 | T | G |
| IDL-C | 15 | 58681772 | T | C |
| IDL-C | 15 | 58683366 | G | A |
| IDL-C | 15 | 58683697 | T | C |
| IDL-C | 15 | 58684967 | T | G |
| IDL-C | 15 | 58686252 | G | A |
| IDL-C | 15 | 58686338 | G | A |
| IDL-C | 15 | 58686939 | T | C |
| IDL-C | 15 | 58687603 | T | C |
| IDL-C | 15 | 58687880 | T | C |
| IDL-C | 15 | 58688478 | C | A |
| IDL-C | 15 | 58689187 | T | C |
| IDL-C | 15 | 58693860 | T | C |
| IDL-C | 15 | 58696300 | T | C |
| IDL-C | 15 | 58698243 | T | G |
| IDL-C | 15 | 58699010 | T | C |
| IDL-C | 15 | 58699937 | T | C |
| IDL-C | 15 | 58701931 | G | A |
| IDL-C | 15 | 58702325 | G | A |
| IDL-C | 15 | 58703069 | C | A |
| IDL-C | 15 | 58705658 | AC | A |
| IDL-C | 15 | 58709182 | G | A |
| IDL-C | 15 | 58710627 | C | A |
| IDL-C | 15 | 58712203 | T | G |
| IDL-C | 15 | 58712973 | T | C |
| IDL-C | 15 | 58718529 | CA | C |
| IDL-C | 15 | 58719741 | G | A |
| IDL-C | 15 | 58720431 | C | A |
| IDL-C | 15 | 58721614 | G | A |
| IDL-C | 15 | 58723675 | T | C |
| IDL-C | 15 | 58727279 | T | C |
| IDL-C | 15 | 58727325 | G | A |
| IDL-C | 15 | 58729144 | T | C |
| IDL-C | 15 | 58731567 | G | A |
| IDL-C | 15 | 58736778 | T | C |
| IDL-C | 15 | 58737341 | T | G |
| IDL-C | 15 | 58738423 | T | C |
| IDL-C | 15 | 58738760 | G | A |
| IDL-C | 15 | 58741891 | G | A |
| IDL-C | 15 | 58742731 | T | G |

**Table S14:** Genetic variants used in the presented analyses. Note that due to coverage of the GWAS chip the instruments differed across considered outcomes *(continued)*

| Exposure | Chromosome | Base-pair position (Build 37) | Other allele | Effect allele |
|---|---|---|---|---|
| IDL-C | 15 | 58743709 | T | C |
| IDL-C | 16 | 72052348 | G | A |
| IDL-C | 16 | 72078989 | TG | T |
| IDL-C | 16 | 72108093 | G | A |
| IDL-C | 17 | 45385012 | T | C |
| IDL-C | 17 | 45390472 | T | C |
| IDL-C | 17 | 45404916 | T | C |
| IDL-C | 17 | 45405144 | G | A |
| IDL-C | 17 | 45438952 | T | C |
| IDL-C | 17 | 45441804 | G | A |
| IDL-C | 17 | 45516667 | T | C |
| IDL-C | 17 | 45548958 | C | A |
| IDL-C | 17 | 45556945 | G | A |
| IDL-C | 17 | 45592266 | G | A |
| IDL-C | 17 | 45616332 | T | C |
| IDL-C | 17 | 45732774 | C | A |
| IDL-C | 17 | 45740805 | G | A |
| IDL-C | 17 | 45755810 | G | A |
| IDL-C | 17 | 45763073 | C | A |
| IDL-C | 17 | 45771816 | T | C |
| IDL-C | 17 | 45791123 | T | G |
| IDL-C | 19 | 10437297 | T | G |
| IDL-C | 19 | 10504616 | C | A |
| IDL-C | 19 | 10633984 | G | A |
| IDL-C | 19 | 10634928 | T | C |
| IDL-C | 19 | 10636904 | T | C |
| IDL-C | 19 | 10639312 | T | C |
| IDL-C | 19 | 10639533 | T | C |
| IDL-C | 19 | 10640380 | G | A |
| IDL-C | 19 | 10641291 | AG | A |
| IDL-C | 19 | 10643471 | G | A |
| IDL-C | 19 | 10646814 | G | A |
| IDL-C | 19 | 10654705 | T | C |
| IDL-C | 19 | 10663165 | T | C |
| IDL-C | 19 | 10703280 | G | A |
| IDL-C | 19 | 10715154 | C | A |
| IDL-C | 19 | 10734951 | T | C |
| IDL-C | 19 | 10739265 | T | C |
| IDL-C | 19 | 10784858 | G | A |
| IDL-C | 19 | 10798974 | T | C |
| IDL-C | 19 | 10802792 | G | A |
| IDL-C | 19 | 10813364 | T | C |
| IDL-C | 19 | 10901440 | G | A |
| IDL-C | 19 | 10903972 | G | A |

**Table S14:** Genetic variants used in the presented analyses. Note that due to coverage of the GWAS chip the instruments differed across considered outcomes *(continued)*

| Exposure | Chromosome | Base-pair position (Build 37) | Other allele | Effect allele |
|---|---|---|---|---|
| IDL-C | 19 | 10904700 | T | C |
| IDL-C | 19 | 10909953 | T | C |
| IDL-C | 19 | 10914829 | G | A |
| IDL-C | 19 | 10916684 | T | C |
| IDL-C | 19 | 10926952 | T | C |
| IDL-C | 19 | 10947522 | T | C |
| IDL-C | 19 | 10948031 | G | A |
| IDL-C | 19 | 10952958 | G | A |
| IDL-C | 19 | 10965270 | T | C |
| IDL-C | 19 | 10967970 | G | A |
| IDL-C | 19 | 10981992 | T | C |
| IDL-C | 19 | 11027626 | C | A |
| IDL-C | 19 | 11054715 | G | A |
| IDL-C | 19 | 11055935 | G | A |
| IDL-C | 19 | 11059389 | G | A |
| IDL-C | 19 | 11065409 | C | A |
| IDL-C | 19 | 11065687 | G | A |
| IDL-C | 19 | 11104747 | G | A |
| IDL-C | 19 | 11105608 | T | C |
| IDL-C | 19 | 11113902 | G | A |
| IDL-C | 19 | 11126160 | T | C |
| IDL-C | 19 | 11128824 | T | C |
| IDL-C | 19 | 11132080 | G | A |
| IDL-C | 19 | 11133272 | G | A |
| IDL-C | 19 | 11135654 | T | C |
| IDL-C | 19 | 11139636 | T | G |
| IDL-C | 19 | 11140144 | G | A |
| IDL-C | 19 | 11140703 | T | C |
| IDL-C | 19 | 11146499 | T | C |
| IDL-C | 19 | 11159076 | G | A |
| IDL-C | 19 | 11159525 | T | G |
| IDL-C | 19 | 11163562 | T | C |
| IDL-C | 19 | 11163601 | T | G |
| IDL-C | 19 | 11163897 | T | C |
| IDL-C | 19 | 11164781 | G | A |
| IDL-C | 19 | 11166431 | CT | C |
| IDL-C | 19 | 11169947 | T | C |
| IDL-C | 19 | 11170839 | T | C |
| IDL-C | 19 | 11174229 | CT | C |
| IDL-C | 19 | 11174625 | G | A |
| IDL-C | 19 | 11174742 | T | C |
| IDL-C | 19 | 11177250 | C | A |
| IDL-C | 19 | 11180047 | T | C |
| IDL-C | 19 | 11183837 | G | A |

**Table S14:** Genetic variants used in the presented analyses. Note that due to coverage of the GWAS chip the instruments differed across considered outcomes *(continued)*

| Exposure | Chromosome | Base-pair position (Build 37) | Other allele | Effect allele |
|---|---|---|---|---|
| IDL-C | 19 | 11184250 | G | A |
| IDL-C | 19 | 11184293 | T | C |
| IDL-C | 19 | 11185014 | G | A |
| IDL-C | 19 | 11185919 | T | C |
| IDL-C | 19 | 11186711 | T | C |
| IDL-C | 19 | 11187611 | G | A |
| IDL-C | 19 | 11188313 | CTTTA | C |
| IDL-C | 19 | 11188899 | T | C |
| IDL-C | 19 | 11189298 | CG | C |
| IDL-C | 19 | 11190544 | T | C |
| IDL-C | 19 | 11195030 | G | A |
| IDL-C | 19 | 11202306 | T | G |
| IDL-C | 19 | 11205975 | G | A |
| IDL-C | 19 | 11206575 | G | A |
| IDL-C | 19 | 11207102 | T | C |
| IDL-C | 19 | 11207516 | G | A |
| IDL-C | 19 | 11207982 | T | C |
| IDL-C | 19 | 11209669 | CA | C |
| IDL-C | 19 | 11209757 | CCAAA | C |
| IDL-C | 19 | 11210912 | T | C |
| IDL-C | 19 | 11227070 | T | C |
| IDL-C | 19 | 11227480 | C | A |
| IDL-C | 19 | 11228965 | C | A |
| IDL-C | 19 | 11230881 | T | C |
| IDL-C | 19 | 11243409 | C | A |
| IDL-C | 19 | 11243445 | G | A |
| IDL-C | 19 | 11245071 | GGAGT | G |
| IDL-C | 19 | 11248104 | G | A |
| IDL-C | 19 | 11250139 | G | A |
| IDL-C | 19 | 11250396 | T | C |
| IDL-C | 19 | 11257169 | T | C |
| IDL-C | 19 | 11262477 | T | G |
| IDL-C | 19 | 11262969 | T | G |
| IDL-C | 19 | 11263968 | CA | C |
| IDL-C | 19 | 11269893 | T | C |
| IDL-C | 19 | 11271714 | G | A |
| IDL-C | 19 | 11274679 | G | A |
| IDL-C | 19 | 11275139 | C | A |
| IDL-C | 19 | 11275258 | G | A |
| IDL-C | 19 | 11276256 | T | C |
| IDL-C | 19 | 11330942 | G | A |
| IDL-C | 19 | 11433021 | AAAG | A |
| IDL-C | 19 | 19130096 | T | C |
| IDL-C | 19 | 19176612 | G | A |

**Table S14:** Genetic variants used in the presented analyses. Note that due to coverage of the GWAS chip the instruments differed across considered outcomes *(continued)*

| Exposure | Chromosome | Base-pair position (Build 37) | Other allele | Effect allele |
|---|---|---|---|---|
| IDL-C | 19 | 19380513 | G | A |
| IDL-C | 19 | 19393714 | G | A |
| IDL-C | 19 | 19407718 | T | C |
| IDL-C | 19 | 19432290 | AG | A |
| IDL-C | 19 | 19458388 | G | A |
| IDL-C | 19 | 19479497 | T | C |
| IDL-C | 19 | 19544644 | T | C |
| IDL-C | 19 | 19624481 | G | A |
| IDL-C | 19 | 20217169 | T | C |
| IDL-C | 19 | 45134682 | C | A |
| IDL-C | 19 | 45150614 | G | A |
| IDL-C | 19 | 45158411 | T | C |
| IDL-C | 19 | 45159023 | T | C |
| IDL-C | 19 | 45160086 | C | A |
| IDL-C | 19 | 45167544 | T | C |
| IDL-C | 19 | 45170097 | G | A |
| IDL-C | 19 | 45173951 | C | A |
| IDL-C | 19 | 45176340 | T | C |
| IDL-C | 19 | 45187163 | G | A |
| IDL-C | 19 | 45188620 | GAA | G |
| IDL-C | 19 | 45190766 | T | C |
| IDL-C | 19 | 45190773 | T | G |
| IDL-C | 19 | 45195313 | T | G |
| IDL-C | 19 | 45195744 | CG | C |
| IDL-C | 19 | 45196135 | G | A |
| IDL-C | 19 | 45196254 | G | A |
| IDL-C | 19 | 45196581 | G | A |
| IDL-C | 19 | 45196964 | G | A |
| IDL-C | 19 | 45197027 | C | A |
| IDL-C | 19 | 45198936 | T | C |
| IDL-C | 19 | 45220896 | T | C |
| IDL-C | 19 | 45222982 | T | G |
| IDL-C | 19 | 45223038 | G | A |
| IDL-C | 19 | 45225345 | T | C |
| IDL-C | 19 | 45232161 | G | A |
| IDL-C | 19 | 45232374 | G | A |
| IDL-C | 19 | 45233329 | AAATTTATAG | A |
| IDL-C | 19 | 45237812 | T | C |
| IDL-C | 19 | 45239185 | G | A |
| IDL-C | 19 | 45239536 | C | A |
| IDL-C | 19 | 45242173 | G | A |
| IDL-C | 19 | 45242740 | G | A |
| IDL-C | 19 | 45247627 | G | A |
| IDL-C | 19 | 45249661 | T | C |

**Table S14:** Genetic variants used in the presented analyses. Note that due to coverage of the GWAS chip the instruments differed across considered outcomes *(continued)*

| Exposure | Chromosome | Base-pair position (Build 37) | Other allele | Effect allele |
|---|---|---|---|---|
| IDL-C | 19 | 45251156 | C | A |
| IDL-C | 19 | 45253491 | T | C |
| IDL-C | 19 | 45253542 | CTTTG | C |
| IDL-C | 19 | 45253611 | ATTGGC | A |
| IDL-C | 19 | 45276590 | GTTA | G |
| IDL-C | 19 | 45296364 | G | A |
| IDL-C | 19 | 45296806 | T | C |
| IDL-C | 19 | 45297928 | G | A |
| IDL-C | 19 | 45302504 | G | A |
| IDL-C | 19 | 45302778 | G | A |
| IDL-C | 19 | 45302949 | GAA | G |
| IDL-C | 19 | 45303122 | AAAAG | A |
| IDL-C | 19 | 45304061 | G | A |
| IDL-C | 19 | 45309431 | T | C |
| IDL-C | 19 | 45311305 | TAAC | T |
| IDL-C | 19 | 45313715 | T | C |
| IDL-C | 19 | 45315445 | G | A |
| IDL-C | 19 | 45319631 | G | A |
| IDL-C | 19 | 45323170 | CCATCCCCAAC | CTCATCCT |
| IDL-C | 19 | 45324138 | G | A |
| IDL-C | 19 | 45325738 | T | G |
| IDL-C | 19 | 45327459 | T | C |
| IDL-C | 19 | 45329214 | T | G |
| IDL-C | 19 | 45330999 | G | A |
| IDL-C | 19 | 45331103 | G | A |
| IDL-C | 19 | 45333834 | T | C |
| IDL-C | 19 | 45335676 | G | A |
| IDL-C | 19 | 45336035 | T | C |
| IDL-C | 19 | 45337918 | G | A |
| IDL-C | 19 | 45338250 | AT | A |
| IDL-C | 19 | 45345623 | G | A |
| IDL-C | 19 | 45347911 | C | A |
| IDL-C | 19 | 45351891 | G | A |
| IDL-C | 19 | 45356141 | T | C |
| IDL-C | 19 | 45357939 | G | A |
| IDL-C | 19 | 45359706 | G | A |
| IDL-C | 19 | 45361646 | G | A |
| IDL-C | 19 | 45365447 | G | A |
| IDL-C | 19 | 45369203 | G | A |
| IDL-C | 19 | 45373565 | G | A |
| IDL-C | 19 | 45377334 | G | A |
| IDL-C | 19 | 45379060 | C | A |
| IDL-C | 19 | 45382034 | G | A |
| IDL-C | 19 | 45382675 | G | A |

**Table S14:** Genetic variants used in the presented analyses. Note that due to coverage of the GWAS chip the instruments differed across considered outcomes *(continued)*

| Exposure | Chromosome | Base-pair position (Build 37) | Other allele | Effect allele |
|---|---|---|---|---|
| IDL-C | 19 | 45384105 | T | C |
| IDL-C | 19 | 45384405 | T | C |
| IDL-C | 19 | 45386467 | GTAA | G |
| IDL-C | 19 | 45387034 | C | A |
| IDL-C | 19 | 45388130 | G | A |
| IDL-C | 19 | 45388241 | T | G |
| IDL-C | 19 | 45389596 | G | A |
| IDL-C | 19 | 45390333 | G | A |
| IDL-C | 19 | 45392254 | T | C |
| IDL-C | 19 | 45395266 | G | A |
| IDL-C | 19 | 45395619 | G | A |
| IDL-C | 19 | 45395844 | G | A |
| IDL-C | 19 | 45397952 | T | C |
| IDL-C | 19 | 45398785 | T | C |
| IDL-C | 19 | 45400484 | TG | T |
| IDL-C | 19 | 45401666 | G | A |
| IDL-C | 19 | 45401868 | T | C |
| IDL-C | 19 | 45402262 | T | G |
| IDL-C | 19 | 45408628 | T | C |
| IDL-C | 19 | 45408836 | T | G |
| IDL-C | 19 | 45410002 | G | A |
| IDL-C | 19 | 45410444 | G | A |
| IDL-C | 19 | 45411941 | T | C |
| IDL-C | 19 | 45412079 | T | C |
| IDL-C | 19 | 45413576 | T | C |
| IDL-C | 19 | 45414392 | T | G |
| IDL-C | 19 | 45415640 | G | A |
| IDL-C | 19 | 45415713 | G | A |
| IDL-C | 19 | 45416291 | T | C |
| IDL-C | 19 | 45418486 | G | A |
| IDL-C | 19 | 45418961 | T | C |
| IDL-C | 19 | 45421254 | G | A |
| IDL-C | 19 | 45421744 | T | G |
| IDL-C | 19 | 45422846 | G | A |
| IDL-C | 19 | 45422946 | G | A |
| IDL-C | 19 | 45423636 | G | A |
| IDL-C | 19 | 45423944 | T | C |
| IDL-C | 19 | 45425178 | G | A |
| IDL-C | 19 | 45425460 | G | A |
| IDL-C | 19 | 45426792 | G | A |
| IDL-C | 19 | 45428234 | G | A |
| IDL-C | 19 | 45429543 | T | C |
| IDL-C | 19 | 45431897 | G | A |
| IDL-C | 19 | 45435466 | CT | C |

**Table S14:** Genetic variants used in the presented analyses. Note that due to coverage of the GWAS chip the instruments differed across considered outcomes *(continued)*

| Exposure | Chromosome | Base-pair position (Build 37) | Other allele | Effect allele |
|---|---|---|---|---|
| IDL-C | 19 | 45436216 | G | A |
| IDL-C | 19 | 45447178 | T | C |
| IDL-C | 19 | 45447221 | G | A |
| IDL-C | 19 | 45450252 | T | C |
| IDL-C | 19 | 45453763 | G | A |
| IDL-C | 19 | 45475700 | T | G |
| IDL-C | 19 | 45478352 | G | A |
| IDL-C | 19 | 45491032 | T | C |
| IDL-C | 19 | 45496303 | T | C |
| IDL-C | 19 | 45515014 | CTGTGTGTG | C |
| IDL-C | 19 | 45531618 | GT | G |
| IDL-C | 19 | 45541782 | G | A |
| IDL-C | 19 | 45653883 | TAGTC | T |
| IDL-C | 19 | 45672478 | G | A |
| IDL-C | 19 | 45693752 | T | C |
| IDL-C | 19 | 45706865 | G | A |
| IDL-C | 19 | 45708570 | T | C |
| IDL-C | 19 | 45713488 | GTTTC | G |
| IDL-C | 19 | 45723379 | T | C |
| IDL-C | 19 | 45880690 | T | C |
| IDL-C | 19 | 46018119 | G | A |
| IDL-C | 1 | 109715950 | G | A |
| IDL-C | 1 | 109757295 | G | A |
| IDL-C | 1 | 109776285 | T | C |
| IDL-C | 1 | 109792735 | ACGC | A |
| IDL-C | 1 | 109798721 | G | A |
| IDL-C | 1 | 109800321 | T | C |
| IDL-C | 1 | 109804646 | G | A |
| IDL-C | 1 | 109806442 | C | A |
| IDL-C | 1 | 109810201 | G | A |
| IDL-C | 1 | 109813485 | T | C |
| IDL-C | 1 | 109814880 | T | C |
| IDL-C | 1 | 109815214 | TG | T |
| IDL-C | 1 | 109815252 | G | A |
| IDL-C | 1 | 109818158 | CT | C |
| IDL-C | 1 | 109819615 | T | C |
| IDL-C | 1 | 109819981 | T | C |
| IDL-C | 1 | 109820884 | G | A |
| IDL-C | 1 | 109820919 | T | G |
| IDL-C | 1 | 109821797 | G | A |
| IDL-C | 1 | 109822166 | G | A |
| IDL-C | 1 | 109824250 | G | A |
| IDL-C | 1 | 109826760 | TTCCTAACC | T |
| IDL-C | 1 | 109827253 | G | A |

**Table S14:** Genetic variants used in the presented analyses. Note that due to coverage of the GWAS chip the instruments differed across considered outcomes *(continued)*

| Exposure | Chromosome | Base-pair position (Build 37) | Other allele | Effect allele |
|---|---|---|---|---|
| IDL-C | 1 | 109834938 | T | C |
| IDL-C | 1 | 109839400 | T | C |
| IDL-C | 1 | 109955569 | T | C |
| IDL-C | 1 | 110019439 | G | A |
| IDL-C | 1 | 110033224 | G | A |
| IDL-C | 1 | 222832010 | G | A |
| IDL-C | 1 | 25721154 | G | A |
| IDL-C | 1 | 25752617 | G | A |
| IDL-C | 1 | 25755014 | G | A |
| IDL-C | 1 | 25768937 | G | A |
| IDL-C | 1 | 25780893 | G | A |
| IDL-C | 1 | 25800790 | T | C |
| IDL-C | 1 | 26987646 | G | A |
| IDL-C | 1 | 54380983 | T | C |
| IDL-C | 1 | 54598299 | G | A |
| IDL-C | 1 | 54602648 | T | C |
| IDL-C | 1 | 54703630 | T | C |
| IDL-C | 1 | 54703702 | C | A |
| IDL-C | 1 | 54708492 | G | A |
| IDL-C | 1 | 54714821 | G | A |
| IDL-C | 1 | 54715751 | T | C |
| IDL-C | 1 | 54717654 | T | C |
| IDL-C | 1 | 54721862 | TC | T |
| IDL-C | 1 | 54722625 | T | C |
| IDL-C | 1 | 54723658 | T | C |
| IDL-C | 1 | 54724389 | G | A |
| IDL-C | 1 | 54726703 | G | A |
| IDL-C | 1 | 54728029 | T | C |
| IDL-C | 1 | 54769277 | T | G |
| IDL-C | 1 | 54772869 | C | A |
| IDL-C | 1 | 54783595 | T | C |
| IDL-C | 1 | 54834840 | T | C |
| IDL-C | 1 | 54849397 | T | C |
| IDL-C | 1 | 54880705 | T | C |
| IDL-C | 1 | 54903630 | C | A |
| IDL-C | 1 | 54911365 | T | C |
| IDL-C | 1 | 54935900 | G | A |
| IDL-C | 1 | 54983266 | T | C |
| IDL-C | 1 | 55083647 | G | A |
| IDL-C | 1 | 55085141 | G | A |
| IDL-C | 1 | 55138291 | G | A |
| IDL-C | 1 | 55160482 | T | G |
| IDL-C | 1 | 55224773 | C | A |
| IDL-C | 1 | 55243587 | G | A |

**Table S14:** Genetic variants used in the presented analyses. Note that due to coverage of the GWAS chip the instruments differed across considered outcomes *(continued)*

| Exposure | Chromosome | Base-pair position (Build 37) | Other allele | Effect allele |
|---|---|---|---|---|
| IDL-C | 1 | 55334001 | G | A |
| IDL-C | 1 | 55378724 | G | A |
| IDL-C | 1 | 55411580 | G | A |
| IDL-C | 1 | 55417370 | T | C |
| IDL-C | 1 | 55431933 | T | C |
| IDL-C | 1 | 55470153 | T | C |
| IDL-C | 1 | 55489542 | T | C |
| IDL-C | 1 | 55489960 | G | A |
| IDL-C | 1 | 55493584 | C | A |
| IDL-C | 1 | 55494017 | T | C |
| IDL-C | 1 | 55494735 | T | C |
| IDL-C | 1 | 55496039 | T | C |
| IDL-C | 1 | 55500025 | C | A |
| IDL-C | 1 | 55500429 | G | A |
| IDL-C | 1 | 55503448 | G | A |
| IDL-C | 1 | 55504650 | G | A |
| IDL-C | 1 | 55505647 | T | G |
| IDL-C | 1 | 55505861 | G | A |
| IDL-C | 1 | 55511995 | G | A |
| IDL-C | 1 | 55513061 | T | C |
| IDL-C | 1 | 55515481 | T | C |
| IDL-C | 1 | 55516508 | G | A |
| IDL-C | 1 | 55518166 | G | A |
| IDL-C | 1 | 55518467 | G | A |
| IDL-C | 1 | 55518528 | C | A |
| IDL-C | 1 | 55519174 | G | A |
| IDL-C | 1 | 55519238 | T | C |
| IDL-C | 1 | 55520212 | G | A |
| IDL-C | 1 | 55520265 | T | C |
| IDL-C | 1 | 55520864 | G | A |
| IDL-C | 1 | 55521109 | G | A |
| IDL-C | 1 | 55522425 | G | A |
| IDL-C | 1 | 55523361 | G | A |
| IDL-C | 1 | 55524116 | T | C |
| IDL-C | 1 | 55524237 | G | A |
| IDL-C | 1 | 55524387 | T | C |
| IDL-C | 1 | 55524601 | G | A |
| IDL-C | 1 | 55527479 | G | A |
| IDL-C | 1 | 55527918 | T | C |
| IDL-C | 1 | 55538552 | T | G |
| IDL-C | 1 | 55608005 | T | C |
| IDL-C | 1 | 55630151 | T | C |
| IDL-C | 1 | 55636240 | T | C |
| IDL-C | 1 | 55656075 | T | G |

**Table S14:** Genetic variants used in the presented analyses. Note that due to coverage of the GWAS chip the instruments differed across considered outcomes *(continued)*

| Exposure | Chromosome | Base-pair position (Build 37) | Other allele | Effect allele |
|---|---|---|---|---|
| IDL-C | 1 | 55711465 | G | A |
| IDL-C | 1 | 55713094 | G | A |
| IDL-C | 1 | 55713628 | G | A |
| IDL-C | 1 | 55716497 | G | A |
| IDL-C | 1 | 55719011 | G | A |
| IDL-C | 1 | 55719166 | T | G |
| IDL-C | 1 | 55719847 | CAT | C |
| IDL-C | 1 | 55720083 | C | A |
| IDL-C | 1 | 55720337 | G | A |
| IDL-C | 1 | 55723756 | C | A |
| IDL-C | 1 | 55723817 | G | A |
| IDL-C | 1 | 55726519 | T | C |
| IDL-C | 1 | 55730327 | T | C |
| IDL-C | 1 | 55731772 | TC | T |
| IDL-C | 1 | 55732596 | ATTGT | A |
| IDL-C | 1 | 55732837 | T | C |
| IDL-C | 1 | 55734875 | T | C |
| IDL-C | 1 | 55738663 | T | C |
| IDL-C | 1 | 55739925 | T | C |
| IDL-C | 1 | 55740015 | T | C |
| IDL-C | 1 | 55741377 | G | A |
| IDL-C | 1 | 55750138 | T | C |
| IDL-C | 1 | 55750949 | G | A |
| IDL-C | 1 | 55752440 | T | C |
| IDL-C | 1 | 55752699 | TA | T |
| IDL-C | 1 | 55754727 | G | A |
| IDL-C | 1 | 55767547 | T | C |
| IDL-C | 1 | 55785672 | T | C |
| IDL-C | 1 | 55785815 | T | G |
| IDL-C | 1 | 55788281 | T | C |
| IDL-C | 1 | 55788609 | T | C |
| IDL-C | 1 | 55793244 | G | A |
| IDL-C | 1 | 55795814 | G | A |
| IDL-C | 1 | 55796086 | T | C |
| IDL-C | 1 | 55808143 | T | C |
| IDL-C | 1 | 55814244 | T | C |
| IDL-C | 1 | 55814595 | T | C |
| IDL-C | 1 | 55815284 | T | G |
| IDL-C | 1 | 55831918 | T | C |
| IDL-C | 1 | 55843810 | T | C |
| IDL-C | 1 | 55960656 | G | A |
| IDL-C | 1 | 56034852 | G | A |
| IDL-C | 1 | 56059000 | CAA | C |
| IDL-C | 1 | 56141726 | T | C |

**Table S14:** Genetic variants used in the presented analyses. Note that due to coverage of the GWAS chip the instruments differed across considered outcomes *(continued)*

| Exposure | Chromosome | Base-pair position (Build 37) | Other allele | Effect allele |
|---|---|---|---|---|
| IDL-C | 1 | 56258328 | T | C |
| IDL-C | 1 | 56413519 | T | C |
| IDL-C | 1 | 56414772 | G | A |
| IDL-C | 1 | 56625395 | G | A |
| IDL-C | 1 | 56763141 | T | C |
| IDL-C | 1 | 56823728 | G | A |
| IDL-C | 1 | 56885874 | G | A |
| IDL-C | 1 | 63013134 | G | A |
| IDL-C | 1 | 63049551 | C | A |
| IDL-C | 20 | 39142516 | G | A |
| IDL-C | 2 | 20902748 | C | A |
| IDL-C | 2 | 20903015 | T | C |
| IDL-C | 2 | 20955032 | AG | A |
| IDL-C | 2 | 20991528 | T | G |
| IDL-C | 2 | 21050729 | GA | G |
| IDL-C | 2 | 21074998 | G | A |
| IDL-C | 2 | 21076264 | TA | T |
| IDL-C | 2 | 21085410 | T | C |
| IDL-C | 2 | 21087323 | TCTC | T |
| IDL-C | 2 | 21091409 | C | A |
| IDL-C | 2 | 21092488 | T | C |
| IDL-C | 2 | 21093112 | G | A |
| IDL-C | 2 | 21100426 | T | C |
| IDL-C | 2 | 21105335 | G | A |
| IDL-C | 2 | 21107494 | T | G |
| IDL-C | 2 | 21111034 | T | C |
| IDL-C | 2 | 21111763 | G | A |
| IDL-C | 2 | 21112689 | G | A |
| IDL-C | 2 | 21121464 | T | C |
| IDL-C | 2 | 21123352 | C | A |
| IDL-C | 2 | 21126936 | G | A |
| IDL-C | 2 | 21133883 | G | A |
| IDL-C | 2 | 21134011 | T | C |
| IDL-C | 2 | 21135187 | G | A |
| IDL-C | 2 | 21135577 | G | A |
| IDL-C | 2 | 21138066 | T | C |
| IDL-C | 2 | 21140540 | T | C |
| IDL-C | 2 | 21141725 | G | A |
| IDL-C | 2 | 21142994 | T | C |
| IDL-C | 2 | 21143329 | G | A |
| IDL-C | 2 | 21143677 | C | A |
| IDL-C | 2 | 21144829 | T | G |
| IDL-C | 2 | 21147179 | T | C |
| IDL-C | 2 | 21148274 | C | A |

**Table S14:** Genetic variants used in the presented analyses. Note that due to coverage of the GWAS chip the instruments differed across considered outcomes *(continued)*

| Exposure | Chromosome | Base-pair position (Build 37) | Other allele | Effect allele |
|---|---|---|---|---|
| IDL-C | 2 | 21150787 | T | C |
| IDL-C | 2 | 21151408 | G | A |
| IDL-C | 2 | 21151917 | T | C |
| IDL-C | 2 | 21152755 | G | A |
| IDL-C | 2 | 21182116 | G | A |
| IDL-C | 2 | 21182710 | G | A |
| IDL-C | 2 | 21184847 | G | A |
| IDL-C | 2 | 21187599 | G | A |
| IDL-C | 2 | 21189091 | T | C |
| IDL-C | 2 | 21190024 | T | C |
| IDL-C | 2 | 21190209 | T | C |
| IDL-C | 2 | 21196112 | G | A |
| IDL-C | 2 | 21196346 | T | G |
| IDL-C | 2 | 21197159 | T | C |
| IDL-C | 2 | 21198900 | G | A |
| IDL-C | 2 | 21202959 | C | A |
| IDL-C | 2 | 21205920 | T | C |
| IDL-C | 2 | 21208484 | T | G |
| IDL-C | 2 | 21216815 | C | A |
| IDL-C | 2 | 21217490 | T | C |
| IDL-C | 2 | 21218805 | T | C |
| IDL-C | 2 | 21221133 | G | A |
| IDL-C | 2 | 21221357 | T | G |
| IDL-C | 2 | 21225753 | T | C |
| IDL-C | 2 | 21231524 | G | A |
| IDL-C | 2 | 21232804 | G | A |
| IDL-C | 2 | 21233972 | T | C |
| IDL-C | 2 | 21239667 | T | C |
| IDL-C | 2 | 21241744 | T | C |
| IDL-C | 2 | 21244000 | T | G |
| IDL-C | 2 | 21245367 | G | A |
| IDL-C | 2 | 21247211 | T | C |
| IDL-C | 2 | 21250914 | G | A |
| IDL-C | 2 | 21251040 | T | G |
| IDL-C | 2 | 21259562 | T | C |
| IDL-C | 2 | 21259612 | G | A |
| IDL-C | 2 | 21260601 | G | A |
| IDL-C | 2 | 21262427 | G | A |
| IDL-C | 2 | 21263639 | T | G |
| IDL-C | 2 | 21263900 | G | A |
| IDL-C | 2 | 21266774 | GGCAGCGCCA | G |
| IDL-C | 2 | 21267210 | G | A |
| IDL-C | 2 | 21267461 | G | A |
| IDL-C | 2 | 21270751 | G | A |

**Table S14:** Genetic variants used in the presented analyses. Note that due to coverage of the GWAS chip the instruments differed across considered outcomes *(continued)*

| Exposure | Chromosome | Base-pair position (Build 37) | Other allele | Effect allele |
|---|---|---|---|---|
| IDL-C | 2 | 21271323 | G | A |
| IDL-C | 2 | 21272763 | AG | A |
| IDL-C | 2 | 21277922 | G | A |
| IDL-C | 2 | 21281856 | G | A |
| IDL-C | 2 | 21286057 | T | C |
| IDL-C | 2 | 21288226 | G | A |
| IDL-C | 2 | 21290067 | T | C |
| IDL-C | 2 | 21294975 | G | A |
| IDL-C | 2 | 21300770 | C | A |
| IDL-C | 2 | 21303616 | T | C |
| IDL-C | 2 | 21309335 | T | C |
| IDL-C | 2 | 21311541 | T | G |
| IDL-C | 2 | 21313577 | G | A |
| IDL-C | 2 | 21313778 | TC | T |
| IDL-C | 2 | 21319986 | GTGATGT | G |
| IDL-C | 2 | 21323868 | CTGT | C |
| IDL-C | 2 | 21327635 | G | A |
| IDL-C | 2 | 21328825 | G | A |
| IDL-C | 2 | 21337306 | T | C |
| IDL-C | 2 | 21367696 | TTGAAGAC | T |
| IDL-C | 2 | 21368217 | G | A |
| IDL-C | 2 | 21371068 | T | C |
| IDL-C | 2 | 21372014 | G | A |
| IDL-C | 2 | 21375346 | G | A |
| IDL-C | 2 | 21378453 | G | A |
| IDL-C | 2 | 21378805 | G | A |
| IDL-C | 2 | 21381177 | G | A |
| IDL-C | 2 | 21381292 | AAAAC | A |
| IDL-C | 2 | 21382786 | C | A |
| IDL-C | 2 | 21383353 | T | C |
| IDL-C | 2 | 21383514 | G | A |
| IDL-C | 2 | 21383536 | G | A |
| IDL-C | 2 | 21389659 | T | G |
| IDL-C | 2 | 21396092 | GAA | G |
| IDL-C | 2 | 21397182 | G | A |
| IDL-C | 2 | 21404245 | CTA | C |
| IDL-C | 2 | 21406048 | T | C |
| IDL-C | 2 | 21411649 | G | A |
| IDL-C | 2 | 21413077 | G | A |
| IDL-C | 2 | 21414142 | T | C |
| IDL-C | 2 | 21415728 | T | G |
| IDL-C | 2 | 21419650 | G | A |
| IDL-C | 2 | 21422928 | T | C |
| IDL-C | 2 | 21423218 | AT | A |

**Table S14:** Genetic variants used in the presented analyses. Note that due to coverage of the GWAS chip the instruments differed across considered outcomes *(continued)*

| Exposure | Chromosome | Base-pair position (Build 37) | Other allele | Effect allele |
|---|---|---|---|---|
| IDL-C | 2 | 21423532 | G | A |
| IDL-C | 2 | 21426683 | AAAAG | A |
| IDL-C | 2 | 21434580 | G | A |
| IDL-C | 2 | 21435700 | C | A |
| IDL-C | 2 | 21437321 | T | G |
| IDL-C | 2 | 21438738 | G | A |
| IDL-C | 2 | 21439329 | T | G |
| IDL-C | 2 | 21445569 | G | A |
| IDL-C | 2 | 21448511 | G | A |
| IDL-C | 2 | 21449034 | T | C |
| IDL-C | 2 | 21449278 | TA | T |
| IDL-C | 2 | 21449987 | G | A |
| IDL-C | 2 | 21450714 | G | A |
| IDL-C | 2 | 21451604 | T | C |
| IDL-C | 2 | 21451687 | G | A |
| IDL-C | 2 | 21453733 | G | A |
| IDL-C | 2 | 21457324 | G | A |
| IDL-C | 2 | 21460176 | T | C |
| IDL-C | 2 | 21467872 | ATT | A |
| IDL-C | 2 | 21469336 | T | C |
| IDL-C | 2 | 21470596 | G | A |
| IDL-C | 2 | 21470987 | T | C |
| IDL-C | 2 | 21471603 | T | C |
| IDL-C | 2 | 21471750 | G | A |
| IDL-C | 2 | 21475944 | T | C |
| IDL-C | 2 | 21476247 | C | A |
| IDL-C | 2 | 21476634 | G | A |
| IDL-C | 2 | 21524000 | G | A |
| IDL-C | 2 | 21530659 | G | A |
| IDL-C | 2 | 21534103 | T | C |
| IDL-C | 2 | 21534298 | T | G |
| IDL-C | 2 | 44052743 | GTTA | G |
| IDL-C | 2 | 44055510 | T | C |
| IDL-C | 2 | 44057570 | C | A |
| IDL-C | 2 | 44063357 | G | A |
| IDL-C | 2 | 44065090 | G | A |
| IDL-C | 2 | 44072576 | T | G |
| IDL-C | 2 | 44073656 | T | C |
| IDL-C | 2 | 44074431 | T | C |
| IDL-C | 2 | 44080324 | G | A |
| IDL-C | 2 | 44088039 | G | A |
| IDL-C | 2 | 44088041 | C | A |
| IDL-C | 2 | 44096259 | T | G |
| IDL-C | 2 | 44096402 | G | A |

**Table S14:** Genetic variants used in the presented analyses. Note that due to coverage of the GWAS chip the instruments differed across considered outcomes *(continued)*

| Exposure | Chromosome | Base-pair position (Build 37) | Other allele | Effect allele |
|---|---|---|---|---|
| IDL-C | 4 | 31423582 | G | A |
| IDL-C | 4 | 73406173 | G | A |
| IDL-C | 4 | 73499882 | T | C |
| IDL-C | 4 | 74720969 | C | A |
| IDL-C | 4 | 74805105 | G | A |
| IDL-C | 4 | 75242737 | CT | C |
| IDL-C | 4 | 75370891 | T | C |
| IDL-C | 4 | 75684215 | T | C |
| IDL-C | 5 | 123072033 | T | C |
| IDL-C | 5 | 156382308 | G | A |
| IDL-C | 5 | 156390297 | T | C |
| IDL-C | 5 | 156394093 | T | C |
| IDL-C | 5 | 156394441 | G | A |
| IDL-C | 5 | 156396003 | T | C |
| IDL-C | 5 | 74302658 | G | A |
| IDL-C | 5 | 74340327 | C | A |
| IDL-C | 5 | 74343769 | G | A |
| IDL-C | 5 | 74352180 | G | A |
| IDL-C | 5 | 74354875 | T | G |
| IDL-C | 5 | 74360981 | ATCT | A |
| IDL-C | 5 | 74372141 | T | C |
| IDL-C | 5 | 74380959 | G | A |
| IDL-C | 5 | 74384115 | G | A |
| IDL-C | 5 | 74389325 | C | A |
| IDL-C | 5 | 74443132 | T | C |
| IDL-C | 5 | 74461850 | T | G |
| IDL-C | 5 | 74476396 | G | A |
| IDL-C | 5 | 74477719 | TA | T |
| IDL-C | 5 | 74479238 | T | C |
| IDL-C | 5 | 74547484 | C | A |
| IDL-C | 5 | 74558893 | G | A |
| IDL-C | 5 | 74562564 | T | C |
| IDL-C | 5 | 74574984 | G | A |
| IDL-C | 5 | 74586866 | CT | C |
| IDL-C | 5 | 74587940 | C | A |
| IDL-C | 5 | 74596682 | T | G |
| IDL-C | 5 | 74602699 | C | A |
| IDL-C | 5 | 74604742 | G | A |
| IDL-C | 5 | 74605220 | G | A |
| IDL-C | 5 | 74611232 | TG | T |
| IDL-C | 5 | 74613943 | T | C |
| IDL-C | 5 | 74615021 | T | C |
| IDL-C | 5 | 74616843 | T | C |
| IDL-C | 5 | 74619639 | T | C |

**Table S14:** Genetic variants used in the presented analyses. Note that due to coverage of the GWAS chip the instruments differed across considered outcomes *(continued)*

| Exposure | Chromosome | Base-pair position (Build 37) | Other allele | Effect allele |
|---|---|---|---|---|
| IDL-C | 5 | 74622121 | TTA | T |
| IDL-C | 5 | 74624234 | G | A |
| IDL-C | 5 | 74625512 | G | A |
| IDL-C | 5 | 74629162 | CAT | C |
| IDL-C | 5 | 74630189 | T | C |
| IDL-C | 5 | 74631352 | T | C |
| IDL-C | 5 | 74632133 | C | A |
| IDL-C | 5 | 74643670 | T | C |
| IDL-C | 5 | 74643998 | ATTC | A |
| IDL-C | 5 | 74644785 | TA | T |
| IDL-C | 5 | 74651084 | G | A |
| IDL-C | 5 | 74651909 | G | A |
| IDL-C | 5 | 74652754 | ATCCATCCGTC | ATGTCCG |
| IDL-C | 5 | 74653462 | T | G |
| IDL-C | 5 | 74656539 | T | C |
| IDL-C | 5 | 74660064 | ATTG | A |
| IDL-C | 5 | 74673707 | G | A |
| IDL-C | 5 | 74688688 | G | A |
| IDL-C | 5 | 74692776 | G | A |
| IDL-C | 5 | 74697050 | T | G |
| IDL-C | 5 | 74717653 | T | C |
| IDL-C | 5 | 74725609 | T | C |
| IDL-C | 5 | 74736600 | TA | T |
| IDL-C | 5 | 74745228 | T | G |
| IDL-C | 5 | 74808237 | T | G |
| IDL-C | 5 | 74808331 | G | A |
| IDL-C | 5 | 74816841 | CT | C |
| IDL-C | 5 | 74844362 | G | A |
| IDL-C | 5 | 74875642 | T | C |
| IDL-C | 5 | 74879890 | G | A |
| IDL-C | 5 | 74906963 | C | A |
| IDL-C | 5 | 74965122 | G | A |
| IDL-C | 5 | 74965554 | G | A |
| IDL-C | 5 | 74967386 | T | C |
| IDL-C | 5 | 74971846 | G | A |
| IDL-C | 6 | 32590735 | T | C |
| IDL-C | 6 | 32602483 | T | G |
| IDL-C | 7 | 21567331 | T | C |
| IDL-C | 7 | 21569089 | G | A |
| IDL-C | 7 | 21576905 | T | C |
| IDL-C | 7 | 21577960 | T | C |
| IDL-C | 7 | 21582917 | T | C |
| IDL-C | 7 | 21588243 | C | A |
| IDL-C | 7 | 21589243 | G | A |

**Table S14:** Genetic variants used in the presented analyses. Note that due to coverage of the GWAS chip the instruments differed across considered outcomes *(continued)*

| Exposure | Chromosome | Base-pair position (Build 37) | Other allele | Effect allele |
|---|---|---|---|---|
| IDL-C | 7 | 21592217 | G | A |
| IDL-C | 7 | 21592949 | G | A |
| IDL-C | 7 | 21596006 | G | A |
| IDL-C | 7 | 21598254 | T | C |
| IDL-C | 7 | 21601659 | GCTCT | G |
| IDL-C | 7 | 21603601 | G | A |
| IDL-C | 7 | 21603678 | G | A |
| IDL-C | 7 | 21607352 | T | C |
| IDL-C | 7 | 21614216 | T | C |
| IDL-C | 7 | 21627909 | G | A |
| IDL-C | 7 | 21631827 | G | A |
| IDL-C | 7 | 21650595 | G | A |
| IDL-C | 8 | 126476873 | T | C |
| IDL-C | 8 | 126479362 | T | C |
| IDL-C | 8 | 126486409 | G | A |
| IDL-C | 8 | 126488930 | G | A |
| IDL-C | 8 | 126495818 | T | C |
| IDL-C | 8 | 126506694 | G | A |
| IDL-C | 8 | 144972454 | T | C |
| IDL-C | 8 | 144973902 | T | C |
| IDL-C | 8 | 144974963 | G | A |
| IDL-C | 8 | 144976572 | G | A |
| IDL-C | 8 | 144978607 | T | G |
| IDL-C | 8 | 144984283 | T | C |
| IDL-C | 8 | 144984956 | G | A |
| IDL-C | 8 | 144990528 | G | A |
| IDL-C | 8 | 144992361 | T | C |
| IDL-C | 8 | 145001031 | T | C |
| IDL-C | 8 | 145003777 | G | A |
| IDL-C | 8 | 145004944 | T | C |
| IDL-C | 8 | 145007187 | G | A |
| IDL-C | 8 | 145043543 | G | A |
| IDL-C | 8 | 145044749 | G | A |
| IDL-C | 8 | 145046662 | T | C |
| IDL-C | 8 | 145057868 | G | A |
| IDL-C | 8 | 145058986 | G | A |
| IDL-C | 8 | 145062020 | T | C |
| IDL-C | 8 | 145071742 | TC | T |
| IDL-C | 8 | 145079301 | T | C |
| IDL-C | 8 | 145081248 | G | A |
| IDL-C | 8 | 145082187 | T | C |
| IDL-C | 8 | 59377357 | G | A |
| IDL-C | 8 | 59388565 | T | C |
| IDL-C | 8 | 9173358 | G | A |

**Table S14:** Genetic variants used in the presented analyses. Note that due to coverage of the GWAS chip the instruments differed across considered outcomes *(continued)*

| Exposure | Chromosome | Base-pair position (Build 37) | Other allele | Effect allele |
|---|---|---|---|---|
| IDL-C | 8 | 9181611 | G | A |
| IDL-C | 8 | 9183596 | G | A |
| IDL-C | 8 | 9183664 | G | A |
| IDL-C | 8 | 9184691 | T | C |
| IDL-C | 8 | 9185146 | T | C |
| IDL-C | 8 | 9192423 | G | A |
| IDL-C | 8 | 9194978 | T | C |
| IDL-C | 8 | 9194985 | G | A |
| IDL-C | 8 | 9200251 | T | C |
| IDL-C | 8 | 9201787 | T | C |
| IDL-C | 8 | 9206069 | G | A |
| IDL-C | 9 | 136137065 | G | A |
| IDL-C | 9 | 136138765 | GCGCCCACCACTA | G |
| IDL-C | 9 | 136139265 | C | A |
| IDL-C | 9 | 136139907 | GAAACTGCC | G |
| IDL-C | 9 | 136142217 | T | G |
| IDL-C | 9 | 136143121 | T | C |
| IDL-C | 9 | 136144309 | G | A |
| IDL-C | 9 | 136145425 | C | A |
| IDL-C | 9 | 136145974 | G | A |
| IDL-C | 9 | 136146046 | G | A |
| IDL-C | 9 | 136146597 | T | C |
| IDL-C | 9 | 136149399 | G | A |
| IDL-C | 9 | 136153875 | T | C |
| IDL-C | 9 | 136153981 | T | C |
| IDL-C | 9 | 136154867 | T | G |
| IDL-C | 9 | 136157133 | G | A |
| IDL-C | 9 | 136164593 | T | G |
| IDL-C | 9 | 136184782 | G | A |
| IDL-C | 9 | 136237672 | C | A |
| IDL-C | 9 | 136267149 | T | C |
| VLDL-C | 11 | 116519655 | T | C |
| VLDL-C | 11 | 116519739 | T | C |
| VLDL-C | 11 | 116520398 | T | C |
| VLDL-C | 11 | 116521135 | T | G |
| VLDL-C | 11 | 116521748 | G | A |
| VLDL-C | 11 | 116523931 | T | C |
| VLDL-C | 11 | 116523988 | T | C |
| VLDL-C | 11 | 116525730 | T | C |
| VLDL-C | 11 | 116526322 | C | A |
| VLDL-C | 11 | 116528544 | T | C |
| VLDL-C | 11 | 116529442 | T | C |
| VLDL-C | 11 | 116536418 | G | A |
| VLDL-C | 11 | 116552562 | T | C |

**Table S14:** Genetic variants used in the presented analyses. Note that due to coverage of the GWAS chip the instruments differed across considered outcomes *(continued)*

| Exposure | Chromosome | Base-pair position (Build 37) | Other allele | Effect allele |
|---|---|---|---|---|
| VLDL-C | 11 | 116552815 | T | C |
| VLDL-C | 11 | 116553025 | G | A |
| VLDL-C | 11 | 116558427 | G | A |
| VLDL-C | 11 | 116559553 | T | G |
| VLDL-C | 11 | 116559663 | G | A |
| VLDL-C | 11 | 116572089 | T | C |
| VLDL-C | 11 | 116574488 | G | A |
| VLDL-C | 11 | 116577441 | G | A |
| VLDL-C | 11 | 116578982 | G | A |
| VLDL-C | 11 | 116579663 | T | C |
| VLDL-C | 11 | 116581259 | T | C |
| VLDL-C | 11 | 116583017 | G | A |
| VLDL-C | 11 | 116584504 | T | C |
| VLDL-C | 11 | 116584987 | T | C |
| VLDL-C | 11 | 116586283 | T | C |
| VLDL-C | 11 | 116590787 | G | A |
| VLDL-C | 11 | 116596309 | C | A |
| VLDL-C | 11 | 116598315 | T | C |
| VLDL-C | 11 | 116603724 | C | A |
| VLDL-C | 11 | 116606766 | G | A |
| VLDL-C | 11 | 116607437 | T | C |
| VLDL-C | 11 | 116613660 | T | C |
| VLDL-C | 11 | 116623659 | T | C |
| VLDL-C | 11 | 116624601 | C | A |
| VLDL-C | 11 | 116624703 | T | G |
| VLDL-C | 11 | 116628396 | T | C |
| VLDL-C | 11 | 116631707 | T | C |
| VLDL-C | 11 | 116639104 | G | A |
| VLDL-C | 11 | 116640768 | G | A |
| VLDL-C | 11 | 116645275 | T | C |
| VLDL-C | 11 | 116646107 | G | A |
| VLDL-C | 11 | 116651115 | T | C |
| VLDL-C | 11 | 116652423 | G | A |
| VLDL-C | 11 | 116657561 | T | C |
| VLDL-C | 11 | 116658031 | T | G |
| VLDL-C | 11 | 116660686 | G | A |
| VLDL-C | 11 | 116662579 | T | C |
| VLDL-C | 11 | 116663707 | G | A |
| VLDL-C | 11 | 116664040 | G | A |
| VLDL-C | 11 | 116667083 | G | A |
| VLDL-C | 11 | 116670676 | C | A |
| VLDL-C | 11 | 116673464 | T | C |
| VLDL-C | 11 | 116680418 | T | C |
| VLDL-C | 11 | 116684028 | T | C |

**Table S14:** Genetic variants used in the presented analyses. Note that due to coverage of the GWAS chip the instruments differed across considered outcomes *(continued)*

| Exposure | Chromosome | Base-pair position (Build 37) | Other allele | Effect allele |
|---|---|---|---|---|
| VLDL-C | 11 | 116692334 | T | C |
| VLDL-C | 11 | 116693464 | T | C |
| VLDL-C | 11 | 116695780 | G | A |
| VLDL-C | 11 | 116699389 | G | A |
| VLDL-C | 11 | 116701535 | T | C |
| VLDL-C | 11 | 116703146 | T | C |
| VLDL-C | 11 | 116703671 | T | G |
| VLDL-C | 11 | 116704178 | C | A |
| VLDL-C | 11 | 116705516 | G | A |
| VLDL-C | 11 | 116705568 | T | G |
| VLDL-C | 11 | 116707338 | G | A |
| VLDL-C | 11 | 116707583 | G | A |
| VLDL-C | 11 | 116710968 | G | A |
| VLDL-C | 11 | 116711141 | T | C |
| VLDL-C | 11 | 116722041 | T | G |
| VLDL-C | 11 | 116730638 | G | A |
| VLDL-C | 11 | 116731205 | G | A |
| VLDL-C | 11 | 116762028 | G | A |
| VLDL-C | 11 | 116764021 | T | C |
| VLDL-C | 11 | 116782202 | G | A |
| VLDL-C | 11 | 116807970 | G | A |
| VLDL-C | 11 | 116908283 | G | A |
| VLDL-C | 11 | 116979689 | T | C |
| VLDL-C | 11 | 116990851 | G | A |
| VLDL-C | 11 | 116998879 | T | C |
| VLDL-C | 11 | 117006383 | G | A |
| VLDL-C | 11 | 117037361 | G | A |
| VLDL-C | 11 | 117039325 | T | G |
| VLDL-C | 11 | 117045335 | T | C |
| VLDL-C | 11 | 117062370 | T | C |
| VLDL-C | 11 | 117071970 | T | C |
| VLDL-C | 11 | 117075566 | T | C |
| VLDL-C | 11 | 117085270 | C | A |
| VLDL-C | 11 | 117089628 | G | A |
| VLDL-C | 16 | 56945049 | C | A |
| VLDL-C | 16 | 56985139 | G | A |
| VLDL-C | 16 | 56985514 | G | A |
| VLDL-C | 16 | 56986762 | T | G |
| VLDL-C | 16 | 56988044 | T | C |
| VLDL-C | 16 | 56990716 | C | A |
| VLDL-C | 16 | 56993324 | C | A |
| VLDL-C | 16 | 56993886 | G | A |
| VLDL-C | 16 | 56995236 | C | A |
| VLDL-C | 16 | 56996158 | T | C |

**Table S14:** Genetic variants used in the presented analyses. Note that due to coverage of the GWAS chip the instruments differed across considered outcomes *(continued)*

| Exposure | Chromosome | Base-pair position (Build 37) | Other allele | Effect allele |
|---|---|---|---|---|
| VLDL-C | 16 | 56997233 | G | A |
| VLDL-C | 16 | 56998918 | G | A |
| VLDL-C | 16 | 56999258 | C | A |
| VLDL-C | 16 | 57002732 | T | G |
| VLDL-C | 16 | 57005479 | C | A |
| VLDL-C | 16 | 57006378 | G | A |
| VLDL-C | 16 | 57006590 | T | C |
| VLDL-C | 19 | 11198502 | T | C |
| VLDL-C | 19 | 19329924 | T | C |
| VLDL-C | 19 | 19366632 | T | C |
| VLDL-C | 19 | 19394368 | G | A |
| VLDL-C | 19 | 19407718 | T | C |
| VLDL-C | 19 | 19456917 | T | C |
| VLDL-C | 19 | 19494483 | G | A |
| VLDL-C | 19 | 19544644 | T | C |
| VLDL-C | 19 | 19786099 | G | A |
| VLDL-C | 19 | 19789528 | G | A |
| VLDL-C | 19 | 45388130 | G | A |
| VLDL-C | 19 | 45388500 | G | A |
| VLDL-C | 19 | 45392254 | T | C |
| VLDL-C | 19 | 45395266 | G | A |
| VLDL-C | 19 | 45396219 | T | C |
| VLDL-C | 19 | 45396665 | T | G |
| VLDL-C | 19 | 45411941 | T | C |
| VLDL-C | 19 | 45414451 | T | C |
| VLDL-C | 19 | 45416478 | G | A |
| VLDL-C | 19 | 45418961 | T | C |
| VLDL-C | 19 | 45425175 | T | G |
| VLDL-C | 1 | 62900811 | G | A |
| VLDL-C | 1 | 62960250 | T | C |
| VLDL-C | 1 | 63034363 | C | A |
| VLDL-C | 1 | 63048276 | C | A |
| VLDL-C | 1 | 63049551 | C | A |
| VLDL-C | 1 | 63118196 | C | A |
| VLDL-C | 1 | 63165761 | G | A |
| VLDL-C | 1 | 63167984 | C | A |
| VLDL-C | 1 | 63171063 | T | C |
| VLDL-C | 1 | 63195692 | T | C |
| VLDL-C | 1 | 63199936 | T | G |
| VLDL-C | 2 | 21150787 | T | C |
| VLDL-C | 2 | 21216815 | C | A |
| VLDL-C | 2 | 21217490 | T | C |
| VLDL-C | 2 | 21231524 | G | A |
| VLDL-C | 2 | 21232195 | G | A |

**Table S14:** Genetic variants used in the presented analyses. Note that due to coverage of the GWAS chip the instruments differed across considered outcomes *(continued)*

| Exposure | Chromosome | Base-pair position (Build 37) | Other allele | Effect allele |
|---|---|---|---|---|
| VLDL-C | 2 | 21239667 | T | C |
| VLDL-C | 2 | 21244000 | T | G |
| VLDL-C | 2 | 21259562 | T | C |
| VLDL-C | 2 | 21263900 | G | A |
| VLDL-C | 2 | 21267461 | G | A |
| VLDL-C | 2 | 21273490 | T | G |
| VLDL-C | 2 | 21292377 | C | A |
| VLDL-C | 2 | 21295832 | G | A |
| VLDL-C | 2 | 27598097 | T | C |
| VLDL-C | 2 | 27730940 | T | C |
| VLDL-C | 2 | 27741105 | G | A |
| VLDL-C | 2 | 27742603 | T | C |
| VLDL-C | 7 | 72822848 | C | A |
| VLDL-C | 7 | 72830546 | T | C |
| VLDL-C | 7 | 72850178 | T | C |
| VLDL-C | 7 | 72854549 | T | C |
| VLDL-C | 7 | 72856269 | G | A |
| VLDL-C | 7 | 72856430 | T | C |
| VLDL-C | 7 | 72864869 | G | A |
| VLDL-C | 7 | 72873420 | G | A |
| VLDL-C | 7 | 72878044 | T | G |
| VLDL-C | 7 | 72971231 | T | C |
| VLDL-C | 7 | 72971728 | C | A |
| VLDL-C | 7 | 72987354 | T | C |
| VLDL-C | 7 | 72989390 | G | A |
| VLDL-C | 7 | 72991592 | T | C |
| VLDL-C | 7 | 72995943 | T | C |
| VLDL-C | 7 | 73003027 | G | A |
| VLDL-C | 7 | 73003303 | C | A |
| VLDL-C | 7 | 73007943 | G | A |
| VLDL-C | 7 | 73016862 | T | C |
| VLDL-C | 7 | 73017005 | G | A |
| VLDL-C | 7 | 73018524 | T | C |
| VLDL-C | 7 | 73026378 | T | C |
| VLDL-C | 7 | 73035857 | T | C |
| VLDL-C | 7 | 73040814 | T | G |
| VLDL-C | 7 | 73042085 | T | C |
| VLDL-C | 7 | 73042443 | T | C |
| VLDL-C | 7 | 73046779 | G | A |
| VLDL-C | 7 | 73047551 | T | C |
| VLDL-C | 7 | 73050599 | G | A |
| VLDL-C | 7 | 73054746 | G | A |
| VLDL-C | 7 | 73056750 | T | C |
| VLDL-C | 7 | 73058017 | G | A |

**Table S14:** Genetic variants used in the presented analyses. Note that due to coverage of the GWAS chip the instruments differed across considered outcomes *(continued)*

| Exposure | Chromosome | Base-pair position (Build 37) | Other allele | Effect allele |
|---|---|---|---|---|
| VLDL-C | 8 | 126447308 | T | C |
| VLDL-C | 8 | 126475770 | G | A |
| VLDL-C | 8 | 126478450 | T | C |
| VLDL-C | 8 | 126479315 | G | A |
| VLDL-C | 8 | 126479362 | T | C |
| VLDL-C | 8 | 126480367 | T | C |
| VLDL-C | 8 | 126484526 | T | G |
| VLDL-C | 8 | 126486409 | G | A |
| VLDL-C | 8 | 126488930 | G | A |
| VLDL-C | 8 | 126491304 | T | C |
| VLDL-C | 8 | 126493392 | G | A |
| VLDL-C | 8 | 126506694 | G | A |
| VLDL-C | 8 | 19720622 | T | C |
| VLDL-C | 8 | 19723503 | T | C |
| VLDL-C | 8 | 19734951 | G | A |
| VLDL-C | 8 | 19755977 | T | G |
| VLDL-C | 8 | 19797916 | T | G |
| VLDL-C | 8 | 19810787 | C | A |
| VLDL-C | 8 | 19811901 | T | C |
| VLDL-C | 8 | 19812812 | T | C |
| VLDL-C | 8 | 19813180 | G | A |
| VLDL-C | 8 | 19816371 | T | C |
| VLDL-C | 8 | 19816934 | T | C |
| VLDL-C | 8 | 19818042 | G | A |
| VLDL-C | 8 | 19819439 | G | A |
| VLDL-C | 8 | 19820405 | G | A |
| VLDL-C | 8 | 19824563 | T | C |
| VLDL-C | 8 | 19826373 | G | A |
| VLDL-C | 8 | 19830170 | T | G |
| VLDL-C | 8 | 19830921 | T | C |
| VLDL-C | 8 | 19838469 | T | C |
| VLDL-C | 8 | 19839709 | T | C |
| VLDL-C | 8 | 19843879 | T | C |
| VLDL-C | 8 | 19844222 | G | A |
| VLDL-C | 8 | 19849757 | G | A |
| VLDL-C | 8 | 19849992 | T | C |
| VLDL-C | 8 | 19856539 | T | C |
| VLDL-C | 8 | 19865175 | G | A |
| VLDL-C | 8 | 19865843 | G | A |
| VLDL-C | 8 | 19868971 | T | C |
| VLDL-C | 8 | 19870693 | G | A |
| VLDL-C | 8 | 19873556 | C | A |
| VLDL-C | 8 | 19873566 | G | A |
| VLDL-C | 8 | 19874153 | T | C |

**Table S14:** Genetic variants used in the presented analyses. Note that due to coverage of the GWAS chip the instruments differed across considered outcomes *(continued)*

| Exposure | Chromosome | Base-pair position (Build 37) | Other allele | Effect allele |
|---|---|---|---|---|
| VLDL-C | 8 | 19875004 | G | A |
| VLDL-C | 8 | 19875100 | T | C |
| VLDL-C | 8 | 19882950 | T | C |
| VLDL-C | 8 | 19912060 | G | A |
| VLDL-C | 8 | 19912370 | G | A |
| VLDL-C | 8 | 19928160 | G | A |
| VLDL-C | 8 | 19928582 | G | A |
| VLDL-C | 8 | 19931087 | T | C |
| VLDL-C | 8 | 19931193 | T | C |
| VLDL-C | 8 | 19931498 | G | A |
| VLDL-C | 8 | 19932088 | T | C |
| VLDL-C | 8 | 19932863 | T | C |
| VLDL-C | 8 | 19936687 | G | A |
| VLDL-C | 8 | 19938946 | T | C |
| VLDL-C | 8 | 19939049 | T | C |
| VLDL-C | 8 | 19941448 | T | C |
| Rem-Chol | 11 | 116519739 | T | C |
| Rem-Chol | 11 | 116520398 | T | C |
| Rem-Chol | 11 | 116523931 | T | C |
| Rem-Chol | 11 | 116525730 | T | C |
| Rem-Chol | 11 | 116529442 | T | C |
| Rem-Chol | 11 | 116536418 | G | A |
| Rem-Chol | 11 | 116552815 | T | C |
| Rem-Chol | 11 | 116553025 | G | A |
| Rem-Chol | 11 | 116558427 | G | A |
| Rem-Chol | 11 | 116559553 | T | G |
| Rem-Chol | 11 | 116574488 | G | A |
| Rem-Chol | 11 | 116577441 | G | A |
| Rem-Chol | 11 | 116579663 | T | C |
| Rem-Chol | 11 | 116581259 | T | C |
| Rem-Chol | 11 | 116583017 | G | A |
| Rem-Chol | 11 | 116584504 | T | C |
| Rem-Chol | 11 | 116584987 | T | C |
| Rem-Chol | 11 | 116586283 | T | C |
| Rem-Chol | 11 | 116590787 | G | A |
| Rem-Chol | 11 | 116603724 | C | A |
| Rem-Chol | 11 | 116606766 | G | A |
| Rem-Chol | 11 | 116607437 | T | C |
| Rem-Chol | 11 | 116611457 | T | C |
| Rem-Chol | 11 | 116613660 | T | C |
| Rem-Chol | 11 | 116623659 | T | C |
| Rem-Chol | 11 | 116631707 | T | C |
| Rem-Chol | 11 | 116639104 | G | A |
| Rem-Chol | 11 | 116645275 | T | C |

**Table S14:** Genetic variants used in the presented analyses. Note that due to coverage of the GWAS chip the instruments differed across considered outcomes *(continued)*

| Exposure | Chromosome | Base-pair position (Build 37) | Other allele | Effect allele |
|---|---|---|---|---|
| Rem-Chol | 11 | 116651115 | T | C |
| Rem-Chol | 11 | 116652423 | G | A |
| Rem-Chol | 11 | 116657561 | T | C |
| Rem-Chol | 11 | 116662579 | T | C |
| Rem-Chol | 11 | 116663707 | G | A |
| Rem-Chol | 11 | 116664040 | G | A |
| Rem-Chol | 11 | 116667083 | G | A |
| Rem-Chol | 11 | 116670676 | C | A |
| Rem-Chol | 11 | 116671476 | T | C |
| Rem-Chol | 11 | 116701535 | T | C |
| Rem-Chol | 11 | 116703146 | T | C |
| Rem-Chol | 11 | 116705516 | G | A |
| Rem-Chol | 11 | 116705568 | T | G |
| Rem-Chol | 11 | 116707583 | G | A |
| Rem-Chol | 11 | 116710968 | G | A |
| Rem-Chol | 11 | 116730638 | G | A |
| Rem-Chol | 11 | 116764021 | T | C |
| Rem-Chol | 11 | 116908283 | G | A |
| Rem-Chol | 11 | 116990851 | G | A |
| Rem-Chol | 11 | 117075566 | T | C |
| Rem-Chol | 16 | 56945049 | C | A |
| Rem-Chol | 16 | 56988044 | T | C |
| Rem-Chol | 16 | 56993324 | C | A |
| Rem-Chol | 16 | 56993886 | G | A |
| Rem-Chol | 16 | 57006590 | T | C |
| Rem-Chol | 19 | 11185919 | T | C |
| Rem-Chol | 19 | 11186412 | T | C |
| Rem-Chol | 19 | 11186711 | T | C |
| Rem-Chol | 19 | 11187611 | G | A |
| Rem-Chol | 19 | 11190544 | T | C |
| Rem-Chol | 19 | 11195030 | G | A |
| Rem-Chol | 19 | 11198187 | T | C |
| Rem-Chol | 19 | 11202306 | T | G |
| Rem-Chol | 19 | 11210912 | T | C |
| Rem-Chol | 19 | 11262969 | T | G |
| Rem-Chol | 19 | 11267757 | T | C |
| Rem-Chol | 19 | 11274679 | G | A |
| Rem-Chol | 19 | 11276256 | T | C |
| Rem-Chol | 19 | 45232161 | G | A |
| Rem-Chol | 19 | 45302504 | G | A |
| Rem-Chol | 19 | 45319631 | G | A |
| Rem-Chol | 19 | 45325738 | T | G |
| Rem-Chol | 19 | 45327459 | T | C |
| Rem-Chol | 19 | 45329214 | T | G |

**Table S14:** Genetic variants used in the presented analyses. Note that due to coverage of the GWAS chip the instruments differed across considered outcomes *(continued)*

| Exposure | Chromosome | Base-pair position (Build 37) | Other allele | Effect allele |
|---|---|---|---|---|
| Rem-Chol | 19 | 45335676 | G | A |
| Rem-Chol | 19 | 45336035 | T | C |
| Rem-Chol | 19 | 45338493 | T | C |
| Rem-Chol | 19 | 45342114 | G | A |
| Rem-Chol | 19 | 45342241 | G | A |
| Rem-Chol | 19 | 45359706 | G | A |
| Rem-Chol | 19 | 45361646 | G | A |
| Rem-Chol | 19 | 45382675 | G | A |
| Rem-Chol | 19 | 45384405 | T | C |
| Rem-Chol | 19 | 45388500 | G | A |
| Rem-Chol | 19 | 45392254 | T | C |
| Rem-Chol | 19 | 45395266 | G | A |
| Rem-Chol | 19 | 45395619 | G | A |
| Rem-Chol | 19 | 45395844 | G | A |
| Rem-Chol | 19 | 45396144 | T | C |
| Rem-Chol | 19 | 45396219 | T | C |
| Rem-Chol | 19 | 45396665 | T | G |
| Rem-Chol | 19 | 45397229 | G | A |
| Rem-Chol | 19 | 45410002 | G | A |
| Rem-Chol | 19 | 45411941 | T | C |
| Rem-Chol | 19 | 45412079 | T | C |
| Rem-Chol | 19 | 45415640 | G | A |
| Rem-Chol | 19 | 45415713 | G | A |
| Rem-Chol | 19 | 45415935 | T | G |
| Rem-Chol | 19 | 45421254 | G | A |
| Rem-Chol | 19 | 45421877 | G | A |
| Rem-Chol | 19 | 45422946 | G | A |
| Rem-Chol | 19 | 45425460 | G | A |
| Rem-Chol | 19 | 45426792 | G | A |
| Rem-Chol | 19 | 45428234 | G | A |
| Rem-Chol | 19 | 45447221 | G | A |
| Rem-Chol | 19 | 45541782 | G | A |
| Rem-Chol | 1 | 109817838 | T | C |
| Rem-Chol | 1 | 109821511 | T | G |
| Rem-Chol | 1 | 109822166 | G | A |
| Rem-Chol | 1 | 55505647 | T | G |
| Rem-Chol | 1 | 62900811 | G | A |
| Rem-Chol | 1 | 62960250 | T | C |
| Rem-Chol | 1 | 62960575 | T | C |
| Rem-Chol | 1 | 63034363 | C | A |
| Rem-Chol | 1 | 63048276 | C | A |
| Rem-Chol | 1 | 63049551 | C | A |
| Rem-Chol | 1 | 63118196 | C | A |
| Rem-Chol | 1 | 63165761 | G | A |

**Table S14:** Genetic variants used in the presented analyses. Note that due to coverage of the GWAS chip the instruments differed across considered outcomes *(continued)*

| Exposure | Chromosome | Base-pair position (Build 37) | Other allele | Effect allele |
|---|---|---|---|---|
| Rem-Chol | 1 | 63171063 | T | C |
| Rem-Chol | 1 | 63195692 | T | C |
| Rem-Chol | 1 | 63199936 | T | G |
| Rem-Chol | 2 | 21134656 | T | C |
| Rem-Chol | 2 | 21140540 | T | C |
| Rem-Chol | 2 | 21141725 | G | A |
| Rem-Chol | 2 | 21150787 | T | C |
| Rem-Chol | 2 | 21182116 | G | A |
| Rem-Chol | 2 | 21209542 | T | C |
| Rem-Chol | 2 | 21213544 | T | C |
| Rem-Chol | 2 | 21216815 | C | A |
| Rem-Chol | 2 | 21217490 | T | C |
| Rem-Chol | 2 | 21232195 | G | A |
| Rem-Chol | 2 | 21239667 | T | C |
| Rem-Chol | 2 | 21244000 | T | G |
| Rem-Chol | 2 | 21259562 | T | C |
| Rem-Chol | 2 | 21263900 | G | A |
| Rem-Chol | 2 | 21267461 | G | A |
| Rem-Chol | 2 | 21271323 | G | A |
| Rem-Chol | 2 | 21285711 | C | A |
| Rem-Chol | 2 | 21286057 | T | C |
| Rem-Chol | 2 | 21288226 | G | A |
| Rem-Chol | 2 | 21288321 | G | A |
| Rem-Chol | 2 | 21292377 | C | A |
| Rem-Chol | 2 | 21300770 | C | A |
| Rem-Chol | 2 | 21303616 | T | C |
| Rem-Chol | 2 | 21309335 | T | C |
| Rem-Chol | 2 | 21313577 | G | A |
| Rem-Chol | 2 | 21327635 | G | A |
| Rem-Chol | 2 | 21328825 | G | A |
| Rem-Chol | 2 | 21368217 | G | A |
| Rem-Chol | 2 | 21369926 | T | C |
| Rem-Chol | 2 | 21371068 | T | C |
| Rem-Chol | 2 | 21381177 | G | A |
| Rem-Chol | 2 | 21382786 | C | A |
| Rem-Chol | 2 | 21383353 | T | C |
| Rem-Chol | 2 | 21383524 | G | A |
| Rem-Chol | 2 | 21383536 | G | A |
| Rem-Chol | 2 | 21383742 | G | A |
| Rem-Chol | 2 | 21397182 | G | A |
| Rem-Chol | 2 | 21415728 | T | G |
| Rem-Chol | 2 | 21419650 | G | A |
| Rem-Chol | 2 | 21435700 | C | A |
| Rem-Chol | 2 | 21439329 | T | G |

**Table S14:** Genetic variants used in the presented analyses. Note that due to coverage of the GWAS chip the instruments differed across considered outcomes *(continued)*

| Exposure | Chromosome | Base-pair position (Build 37) | Other allele | Effect allele |
|---|---|---|---|---|
| Rem-Chol | 2 | 21448511 | G | A |
| Rem-Chol | 2 | 21450714 | G | A |
| Rem-Chol | 2 | 21453733 | G | A |
| Rem-Chol | 2 | 21469152 | T | G |
| Rem-Chol | 2 | 21476247 | C | A |
| Rem-Chol | 2 | 21476634 | G | A |
| Rem-Chol | 2 | 44072576 | T | G |
| Rem-Chol | 7 | 72856430 | T | C |
| Rem-Chol | 7 | 72878044 | T | G |
| Rem-Chol | 7 | 72934510 | G | A |
| Rem-Chol | 7 | 72977249 | T | C |
| Rem-Chol | 7 | 72998952 | G | A |
| Rem-Chol | 7 | 72999105 | T | G |
| Rem-Chol | 8 | 126481694 | T | C |
| Rem-Chol | 8 | 126481952 | G | A |
| Rem-Chol | 8 | 126485296 | T | C |
| Rem-Chol | 8 | 126486409 | G | A |
| Rem-Chol | 8 | 126488250 | T | C |
| Rem-Chol | 8 | 126495818 | T | C |
| Rem-Chol | 8 | 126506694 | G | A |
| Rem-Chol | 8 | 126507389 | C | A |
| Apo-B | 11 | 116519655 | T | C |
| Apo-B | 11 | 116520398 | T | C |
| Apo-B | 11 | 116521558 | G | A |
| Apo-B | 11 | 116522233 | TC | T |
| Apo-B | 11 | 116522854 | G | A |
| Apo-B | 11 | 116524569 | CTT | C |
| Apo-B | 11 | 116525867 | T | C |
| Apo-B | 11 | 116526032 | CCT | C |
| Apo-B | 11 | 116526322 | C | A |
| Apo-B | 11 | 116528224 | T | G |
| Apo-B | 11 | 116529313 | G | A |
| Apo-B | 11 | 116536418 | G | A |
| Apo-B | 11 | 116552562 | T | C |
| Apo-B | 11 | 116553025 | G | A |
| Apo-B | 11 | 116555033 | TA | T |
| Apo-B | 11 | 116557248 | T | C |
| Apo-B | 11 | 116559553 | T | G |
| Apo-B | 11 | 116575473 | G | A |
| Apo-B | 11 | 116577441 | G | A |
| Apo-B | 11 | 116578982 | G | A |
| Apo-B | 11 | 116579663 | T | C |
| Apo-B | 11 | 116580816 | T | C |
| Apo-B | 11 | 116581259 | T | C |

**Table S14:** Genetic variants used in the presented analyses. Note that due to coverage of the GWAS chip the instruments differed across considered outcomes *(continued)*

| Exposure | Chromosome | Base-pair position (Build 37) | Other allele | Effect allele |
|---|---|---|---|---|
| Apo-B | 11 | 116581641 | G | A |
| Apo-B | 11 | 116583017 | G | A |
| Apo-B | 11 | 116584987 | T | C |
| Apo-B | 11 | 116585533 | G | A |
| Apo-B | 11 | 116586283 | T | C |
| Apo-B | 11 | 116588629 | T | C |
| Apo-B | 11 | 116590787 | G | A |
| Apo-B | 11 | 116596309 | C | A |
| Apo-B | 11 | 116598988 | G | A |
| Apo-B | 11 | 116600400 | G | A |
| Apo-B | 11 | 116603677 | G | A |
| Apo-B | 11 | 116603724 | C | A |
| Apo-B | 11 | 116606766 | G | A |
| Apo-B | 11 | 116607437 | T | C |
| Apo-B | 11 | 116613660 | T | C |
| Apo-B | 11 | 116623659 | T | C |
| Apo-B | 11 | 116624601 | C | A |
| Apo-B | 11 | 116624703 | T | G |
| Apo-B | 11 | 116628396 | T | C |
| Apo-B | 11 | 116631707 | T | C |
| Apo-B | 11 | 116634173 | T | C |
| Apo-B | 11 | 116639104 | G | A |
| Apo-B | 11 | 116639692 | T | C |
| Apo-B | 11 | 116645275 | T | C |
| Apo-B | 11 | 116649135 | G | A |
| Apo-B | 11 | 116651115 | T | C |
| Apo-B | 11 | 116657561 | T | C |
| Apo-B | 11 | 116657590 | CA | C |
| Apo-B | 11 | 116660686 | G | A |
| Apo-B | 11 | 116662579 | T | C |
| Apo-B | 11 | 116664040 | G | A |
| Apo-B | 11 | 116667083 | G | A |
| Apo-B | 11 | 116670676 | C | A |
| Apo-B | 11 | 116673464 | T | C |
| Apo-B | 11 | 116684028 | T | C |
| Apo-B | 11 | 116688765 | C | A |
| Apo-B | 11 | 116692334 | T | C |
| Apo-B | 11 | 116694938 | G | A |
| Apo-B | 11 | 116699389 | G | A |
| Apo-B | 11 | 116701535 | T | C |
| Apo-B | 11 | 116701850 | T | C |
| Apo-B | 11 | 116703146 | T | C |
| Apo-B | 11 | 116703671 | T | G |
| Apo-B | 11 | 116704178 | C | A |

**Table S14:** Genetic variants used in the presented analyses. Note that due to coverage of the GWAS chip the instruments differed across considered outcomes *(continued)*

| Exposure | Chromosome | Base-pair position (Build 37) | Other allele | Effect allele |
|---|---|---|---|---|
| Apo-B | 11 | 116705516 | G | A |
| Apo-B | 11 | 116705568 | T | G |
| Apo-B | 11 | 116707338 | G | A |
| Apo-B | 11 | 116707583 | G | A |
| Apo-B | 11 | 116707684 | G | A |
| Apo-B | 11 | 116710968 | G | A |
| Apo-B | 11 | 116711141 | T | C |
| Apo-B | 11 | 116722041 | T | G |
| Apo-B | 11 | 116730638 | G | A |
| Apo-B | 11 | 116732512 | T | G |
| Apo-B | 11 | 116736679 | G | A |
| Apo-B | 11 | 116741572 | C | A |
| Apo-B | 11 | 116762028 | G | A |
| Apo-B | 11 | 116764021 | T | C |
| Apo-B | 11 | 116772441 | T | C |
| Apo-B | 11 | 116782202 | G | A |
| Apo-B | 11 | 116807970 | G | A |
| Apo-B | 11 | 116966435 | G | A |
| Apo-B | 11 | 116967666 | T | C |
| Apo-B | 11 | 116969390 | T | C |
| Apo-B | 11 | 116972894 | C | A |
| Apo-B | 11 | 116981580 | G | A |
| Apo-B | 11 | 116990760 | GAAAC | G |
| Apo-B | 11 | 116990851 | G | A |
| Apo-B | 11 | 116993292 | T | C |
| Apo-B | 11 | 116996539 | G | A |
| Apo-B | 11 | 117004656 | T | C |
| Apo-B | 11 | 117027183 | G | A |
| Apo-B | 11 | 117045335 | T | C |
| Apo-B | 11 | 117048367 | T | C |
| Apo-B | 11 | 117049508 | G | A |
| Apo-B | 11 | 117075566 | T | C |
| Apo-B | 15 | 58723479 | T | C |
| Apo-B | 15 | 58723675 | T | C |
| Apo-B | 16 | 56987765 | T | C |
| Apo-B | 16 | 56988044 | T | C |
| Apo-B | 16 | 56989830 | G | A |
| Apo-B | 16 | 56993161 | G | A |
| Apo-B | 16 | 56993324 | C | A |
| Apo-B | 16 | 72052348 | G | A |
| Apo-B | 16 | 72078989 | TG | T |
| Apo-B | 16 | 72108093 | G | A |
| Apo-B | 18 | 3353012 | G | A |
| Apo-B | 19 | 10636323 | G | A |

**Table S14:** Genetic variants used in the presented analyses. Note that due to coverage of the GWAS chip the instruments differed across considered outcomes *(continued)*

| Exposure | Chromosome | Base-pair position (Build 37) | Other allele | Effect allele |
|---|---|---|---|---|
| Apo-B | 19 | 10641608 | G | A |
| Apo-B | 19 | 10643471 | G | A |
| Apo-B | 19 | 10703280 | G | A |
| Apo-B | 19 | 10734951 | T | C |
| Apo-B | 19 | 10901440 | G | A |
| Apo-B | 19 | 10903972 | G | A |
| Apo-B | 19 | 10909953 | T | C |
| Apo-B | 19 | 10914829 | G | A |
| Apo-B | 19 | 10915031 | G | A |
| Apo-B | 19 | 10916684 | T | C |
| Apo-B | 19 | 10926952 | T | C |
| Apo-B | 19 | 10927540 | T | C |
| Apo-B | 19 | 10933022 | T | C |
| Apo-B | 19 | 10948031 | G | A |
| Apo-B | 19 | 10950125 | T | C |
| Apo-B | 19 | 10952958 | G | A |
| Apo-B | 19 | 10954950 | G | A |
| Apo-B | 19 | 10965270 | T | C |
| Apo-B | 19 | 10967970 | G | A |
| Apo-B | 19 | 10981992 | T | C |
| Apo-B | 19 | 11027626 | C | A |
| Apo-B | 19 | 11054715 | G | A |
| Apo-B | 19 | 11055935 | G | A |
| Apo-B | 19 | 11059389 | G | A |
| Apo-B | 19 | 11065687 | G | A |
| Apo-B | 19 | 11075706 | G | A |
| Apo-B | 19 | 11088219 | T | C |
| Apo-B | 19 | 11106790 | T | C |
| Apo-B | 19 | 11112151 | T | C |
| Apo-B | 19 | 11113902 | G | A |
| Apo-B | 19 | 11114206 | T | C |
| Apo-B | 19 | 11126160 | T | C |
| Apo-B | 19 | 11127069 | T | G |
| Apo-B | 19 | 11128824 | T | C |
| Apo-B | 19 | 11132080 | G | A |
| Apo-B | 19 | 11134090 | G | A |
| Apo-B | 19 | 11136315 | T | C |
| Apo-B | 19 | 11139636 | T | G |
| Apo-B | 19 | 11140144 | G | A |
| Apo-B | 19 | 11146499 | T | C |
| Apo-B | 19 | 11159076 | G | A |
| Apo-B | 19 | 11159096 | T | G |
| Apo-B | 19 | 11159525 | T | G |
| Apo-B | 19 | 11163562 | T | C |

**Table S14:** Genetic variants used in the presented analyses. Note that due to coverage of the GWAS chip the instruments differed across considered outcomes *(continued)*

| Exposure | Chromosome | Base-pair position (Build 37) | Other allele | Effect allele |
|---|---|---|---|---|
| Apo-B | 19 | 11163601 | T | G |
| Apo-B | 19 | 11163897 | T | C |
| Apo-B | 19 | 11164781 | G | A |
| Apo-B | 19 | 11166431 | CT | C |
| Apo-B | 19 | 11169514 | T | C |
| Apo-B | 19 | 11169947 | T | C |
| Apo-B | 19 | 11170839 | T | C |
| Apo-B | 19 | 11171281 | C | A |
| Apo-B | 19 | 11172299 | G | A |
| Apo-B | 19 | 11174229 | CT | C |
| Apo-B | 19 | 11174625 | G | A |
| Apo-B | 19 | 11174742 | T | C |
| Apo-B | 19 | 11177250 | C | A |
| Apo-B | 19 | 11177408 | T | C |
| Apo-B | 19 | 11180047 | T | C |
| Apo-B | 19 | 11184250 | G | A |
| Apo-B | 19 | 11184293 | T | C |
| Apo-B | 19 | 11185014 | G | A |
| Apo-B | 19 | 11185919 | T | C |
| Apo-B | 19 | 11186711 | T | C |
| Apo-B | 19 | 11187611 | G | A |
| Apo-B | 19 | 11188313 | CTTTA | C |
| Apo-B | 19 | 11188899 | T | C |
| Apo-B | 19 | 11189298 | CG | C |
| Apo-B | 19 | 11190110 | G | A |
| Apo-B | 19 | 11190544 | T | C |
| Apo-B | 19 | 11195030 | G | A |
| Apo-B | 19 | 11202306 | T | G |
| Apo-B | 19 | 11205975 | G | A |
| Apo-B | 19 | 11206575 | G | A |
| Apo-B | 19 | 11207102 | T | C |
| Apo-B | 19 | 11207516 | G | A |
| Apo-B | 19 | 11207982 | T | C |
| Apo-B | 19 | 11209757 | CCAAA | C |
| Apo-B | 19 | 11210912 | T | C |
| Apo-B | 19 | 11227070 | T | C |
| Apo-B | 19 | 11227480 | C | A |
| Apo-B | 19 | 11228745 | T | C |
| Apo-B | 19 | 11228965 | C | A |
| Apo-B | 19 | 11230881 | T | C |
| Apo-B | 19 | 11236180 | C | A |
| Apo-B | 19 | 11243445 | G | A |
| Apo-B | 19 | 11245071 | GGAGT | G |
| Apo-B | 19 | 11248104 | G | A |

**Table S14:** Genetic variants used in the presented analyses. Note that due to coverage of the GWAS chip the instruments differed across considered outcomes *(continued)*

| Exposure | Chromosome | Base-pair position (Build 37) | Other allele | Effect allele |
|---|---|---|---|---|
| Apo-B | 19 | 11250396 | T | C |
| Apo-B | 19 | 11257169 | T | C |
| Apo-B | 19 | 11262477 | T | G |
| Apo-B | 19 | 11262969 | T | G |
| Apo-B | 19 | 11267757 | T | C |
| Apo-B | 19 | 11271714 | G | A |
| Apo-B | 19 | 11274679 | G | A |
| Apo-B | 19 | 11275139 | C | A |
| Apo-B | 19 | 11275258 | G | A |
| Apo-B | 19 | 11276256 | T | C |
| Apo-B | 19 | 11277232 | G | A |
| Apo-B | 19 | 19130096 | T | C |
| Apo-B | 19 | 19325963 | T | G |
| Apo-B | 19 | 19329924 | T | C |
| Apo-B | 19 | 19366632 | T | C |
| Apo-B | 19 | 19380513 | G | A |
| Apo-B | 19 | 19393714 | G | A |
| Apo-B | 19 | 19394368 | G | A |
| Apo-B | 19 | 19407718 | T | C |
| Apo-B | 19 | 19418866 | T | C |
| Apo-B | 19 | 19432290 | AG | A |
| Apo-B | 19 | 19456917 | T | C |
| Apo-B | 19 | 19458388 | G | A |
| Apo-B | 19 | 19479497 | T | C |
| Apo-B | 19 | 19494483 | G | A |
| Apo-B | 19 | 19578743 | G | A |
| Apo-B | 19 | 19610596 | T | C |
| Apo-B | 19 | 19624481 | G | A |
| Apo-B | 19 | 19658472 | T | G |
| Apo-B | 19 | 19721722 | G | A |
| Apo-B | 19 | 19727152 | G | A |
| Apo-B | 19 | 19746151 | T | G |
| Apo-B | 19 | 19789528 | G | A |
| Apo-B | 19 | 19892796 | G | A |
| Apo-B | 19 | 19919422 | G | A |
| Apo-B | 19 | 20100542 | G | A |
| Apo-B | 19 | 20151617 | GT | G |
| Apo-B | 19 | 20217169 | T | C |
| Apo-B | 19 | 45122043 | C | A |
| Apo-B | 19 | 45134682 | C | A |
| Apo-B | 19 | 45135841 | T | C |
| Apo-B | 19 | 45156201 | TA | T |
| Apo-B | 19 | 45160086 | C | A |
| Apo-B | 19 | 45176340 | T | C |

**Table S14:** Genetic variants used in the presented analyses. Note that due to coverage of the GWAS chip the instruments differed across considered outcomes *(continued)*

| Exposure | Chromosome | Base-pair position (Build 37) | Other allele | Effect allele |
|---|---|---|---|---|
| Apo-B | 19 | 45187163 | G | A |
| Apo-B | 19 | 45197027 | C | A |
| Apo-B | 19 | 45220896 | T | C |
| Apo-B | 19 | 45222982 | T | G |
| Apo-B | 19 | 45223038 | G | A |
| Apo-B | 19 | 45224236 | G | A |
| Apo-B | 19 | 45225345 | T | C |
| Apo-B | 19 | 45231074 | CCCTT | C |
| Apo-B | 19 | 45232161 | G | A |
| Apo-B | 19 | 45232205 | G | A |
| Apo-B | 19 | 45233271 | TA | T |
| Apo-B | 19 | 45233329 | AAATTTATAG | A |
| Apo-B | 19 | 45234124 | G | A |
| Apo-B | 19 | 45237812 | T | C |
| Apo-B | 19 | 45237919 | T | C |
| Apo-B | 19 | 45239185 | G | A |
| Apo-B | 19 | 45239536 | C | A |
| Apo-B | 19 | 45242173 | G | A |
| Apo-B | 19 | 45242740 | G | A |
| Apo-B | 19 | 45247627 | G | A |
| Apo-B | 19 | 45251156 | C | A |
| Apo-B | 19 | 45253491 | T | C |
| Apo-B | 19 | 45253542 | CTTTG | C |
| Apo-B | 19 | 45253611 | ATTGGC | A |
| Apo-B | 19 | 45276590 | GTTA | G |
| Apo-B | 19 | 45296364 | G | A |
| Apo-B | 19 | 45296806 | T | C |
| Apo-B | 19 | 45302504 | G | A |
| Apo-B | 19 | 45302778 | G | A |
| Apo-B | 19 | 45302949 | GAA | G |
| Apo-B | 19 | 45302951 | G | A |
| Apo-B | 19 | 45303122 | AAAAG | A |
| Apo-B | 19 | 45315445 | G | A |
| Apo-B | 19 | 45319631 | G | A |
| Apo-B | 19 | 45324138 | G | A |
| Apo-B | 19 | 45325738 | T | G |
| Apo-B | 19 | 45326664 | T | C |
| Apo-B | 19 | 45327459 | T | C |
| Apo-B | 19 | 45331103 | G | A |
| Apo-B | 19 | 45333834 | T | C |
| Apo-B | 19 | 45335676 | G | A |
| Apo-B | 19 | 45336035 | T | C |
| Apo-B | 19 | 45337249 | C | A |
| Apo-B | 19 | 45337918 | G | A |

**Table S14:** Genetic variants used in the presented analyses. Note that due to coverage of the GWAS chip the instruments differed across considered outcomes *(continued)*

| Exposure | Chromosome | Base-pair position (Build 37) | Other allele | Effect allele |
|---|---|---|---|---|
| Apo-B | 19 | 45338250 | AT | A |
| Apo-B | 19 | 45345708 | G | A |
| Apo-B | 19 | 45347911 | C | A |
| Apo-B | 19 | 45351937 | T | C |
| Apo-B | 19 | 45356141 | T | C |
| Apo-B | 19 | 45357377 | T | C |
| Apo-B | 19 | 45358026 | T | C |
| Apo-B | 19 | 45358353 | T | C |
| Apo-B | 19 | 45361646 | G | A |
| Apo-B | 19 | 45365447 | G | A |
| Apo-B | 19 | 45369203 | G | A |
| Apo-B | 19 | 45373565 | G | A |
| Apo-B | 19 | 45377334 | G | A |
| Apo-B | 19 | 45379060 | C | A |
| Apo-B | 19 | 45382034 | G | A |
| Apo-B | 19 | 45382675 | G | A |
| Apo-B | 19 | 45384405 | T | C |
| Apo-B | 19 | 45385356 | C | A |
| Apo-B | 19 | 45386467 | GTAA | G |
| Apo-B | 19 | 45387034 | C | A |
| Apo-B | 19 | 45387596 | G | A |
| Apo-B | 19 | 45388500 | G | A |
| Apo-B | 19 | 45389596 | G | A |
| Apo-B | 19 | 45392254 | T | C |
| Apo-B | 19 | 45393516 | T | C |
| Apo-B | 19 | 45395266 | G | A |
| Apo-B | 19 | 45395619 | G | A |
| Apo-B | 19 | 45395714 | T | C |
| Apo-B | 19 | 45395844 | G | A |
| Apo-B | 19 | 45396219 | T | C |
| Apo-B | 19 | 45399896 | G | A |
| Apo-B | 19 | 45400484 | TG | T |
| Apo-B | 19 | 45401868 | T | C |
| Apo-B | 19 | 45402262 | T | G |
| Apo-B | 19 | 45404691 | G | A |
| Apo-B | 19 | 45406673 | G | A |
| Apo-B | 19 | 45408628 | T | C |
| Apo-B | 19 | 45410002 | G | A |
| Apo-B | 19 | 45411941 | T | C |
| Apo-B | 19 | 45412079 | T | C |
| Apo-B | 19 | 45414451 | T | C |
| Apo-B | 19 | 45415640 | G | A |
| Apo-B | 19 | 45416478 | G | A |
| Apo-B | 19 | 45416831 | T | G |

**Table S14:** Genetic variants used in the presented analyses. Note that due to coverage of the GWAS chip the instruments differed across considered outcomes *(continued)*

| Exposure | Chromosome | Base-pair position (Build 37) | Other allele | Effect allele |
|---|---|---|---|---|
| Apo-B | 19 | 45418486 | G | A |
| Apo-B | 19 | 45418961 | T | C |
| Apo-B | 19 | 45421204 | T | C |
| Apo-B | 19 | 45421254 | G | A |
| Apo-B | 19 | 45422946 | G | A |
| Apo-B | 19 | 45423944 | T | C |
| Apo-B | 19 | 45425178 | G | A |
| Apo-B | 19 | 45426792 | G | A |
| Apo-B | 19 | 45428234 | G | A |
| Apo-B | 19 | 45429543 | T | C |
| Apo-B | 19 | 45436216 | G | A |
| Apo-B | 19 | 45436657 | G | A |
| Apo-B | 19 | 45441608 | T | C |
| Apo-B | 19 | 45447221 | G | A |
| Apo-B | 19 | 45478352 | G | A |
| Apo-B | 19 | 45491032 | T | C |
| Apo-B | 19 | 45531618 | GT | G |
| Apo-B | 19 | 45541782 | G | A |
| Apo-B | 19 | 45653883 | TAGTC | T |
| Apo-B | 19 | 45672478 | G | A |
| Apo-B | 19 | 45723379 | T | C |
| Apo-B | 19 | 45880690 | T | C |
| Apo-B | 1 | 109814880 | T | C |
| Apo-B | 1 | 109815252 | G | A |
| Apo-B | 1 | 109817245 | T | C |
| Apo-B | 1 | 109818158 | CT | C |
| Apo-B | 1 | 109819296 | C | A |
| Apo-B | 1 | 109820884 | G | A |
| Apo-B | 1 | 109821511 | T | G |
| Apo-B | 1 | 109822166 | G | A |
| Apo-B | 1 | 27284913 | T | C |
| Apo-B | 1 | 3704452 | T | G |
| Apo-B | 1 | 3738018 | T | C |
| Apo-B | 1 | 54708492 | G | A |
| Apo-B | 1 | 54715326 | C | A |
| Apo-B | 1 | 54721862 | TC | T |
| Apo-B | 1 | 54728029 | T | C |
| Apo-B | 1 | 54834840 | T | C |
| Apo-B | 1 | 54849397 | T | C |
| Apo-B | 1 | 54880705 | T | C |
| Apo-B | 1 | 54935900 | G | A |
| Apo-B | 1 | 54983266 | T | C |
| Apo-B | 1 | 55083647 | G | A |
| Apo-B | 1 | 55085141 | G | A |

**Table S14:** Genetic variants used in the presented analyses. Note that due to coverage of the GWAS chip the instruments differed across considered outcomes *(continued)*

| Exposure | Chromosome | Base-pair position (Build 37) | Other allele | Effect allele |
|---|---|---|---|---|
| Apo-B | 1 | 55138291 | G | A |
| Apo-B | 1 | 55160482 | T | G |
| Apo-B | 1 | 55224773 | C | A |
| Apo-B | 1 | 55243587 | G | A |
| Apo-B | 1 | 55334001 | G | A |
| Apo-B | 1 | 55378724 | G | A |
| Apo-B | 1 | 55411580 | G | A |
| Apo-B | 1 | 55431933 | T | C |
| Apo-B | 1 | 55489542 | T | C |
| Apo-B | 1 | 55493584 | C | A |
| Apo-B | 1 | 55494017 | T | C |
| Apo-B | 1 | 55496039 | T | C |
| Apo-B | 1 | 55503448 | G | A |
| Apo-B | 1 | 55505647 | T | G |
| Apo-B | 1 | 55505861 | G | A |
| Apo-B | 1 | 55518467 | G | A |
| Apo-B | 1 | 55519015 | T | G |
| Apo-B | 1 | 55520212 | G | A |
| Apo-B | 1 | 55521109 | G | A |
| Apo-B | 1 | 55523361 | G | A |
| Apo-B | 1 | 55524116 | T | C |
| Apo-B | 1 | 55524237 | G | A |
| Apo-B | 1 | 55524601 | G | A |
| Apo-B | 1 | 55527918 | T | C |
| Apo-B | 1 | 55538552 | T | G |
| Apo-B | 1 | 55636240 | T | C |
| Apo-B | 1 | 55656075 | T | G |
| Apo-B | 1 | 55711465 | G | A |
| Apo-B | 1 | 55713094 | G | A |
| Apo-B | 1 | 55713628 | G | A |
| Apo-B | 1 | 55716497 | G | A |
| Apo-B | 1 | 55719011 | G | A |
| Apo-B | 1 | 55720083 | C | A |
| Apo-B | 1 | 55720337 | G | A |
| Apo-B | 1 | 55726519 | T | C |
| Apo-B | 1 | 55730327 | T | C |
| Apo-B | 1 | 55731772 | TC | T |
| Apo-B | 1 | 55732596 | ATTGT | A |
| Apo-B | 1 | 55732837 | T | C |
| Apo-B | 1 | 55734875 | T | C |
| Apo-B | 1 | 55738663 | T | C |
| Apo-B | 1 | 55739925 | T | C |
| Apo-B | 1 | 55744966 | G | A |
| Apo-B | 1 | 55746330 | G | A |

**Table S14:** Genetic variants used in the presented analyses. Note that due to coverage of the GWAS chip the instruments differed across considered outcomes *(continued)*

| Exposure | Chromosome | Base-pair position (Build 37) | Other allele | Effect allele |
|---|---|---|---|---|
| Apo-B | 1 | 55750138 | T | C |
| Apo-B | 1 | 55767547 | T | C |
| Apo-B | 1 | 55779387 | T | G |
| Apo-B | 1 | 55785672 | T | C |
| Apo-B | 1 | 55785815 | T | G |
| Apo-B | 1 | 55798859 | T | C |
| Apo-B | 1 | 55806465 | T | C |
| Apo-B | 1 | 56034852 | G | A |
| Apo-B | 1 | 56549124 | T | C |
| Apo-B | 1 | 56608087 | T | C |
| Apo-B | 1 | 56625395 | G | A |
| Apo-B | 1 | 62900811 | G | A |
| Apo-B | 1 | 62901807 | C | A |
| Apo-B | 1 | 62934002 | G | A |
| Apo-B | 1 | 62936912 | T | C |
| Apo-B | 1 | 62944947 | CA | C |
| Apo-B | 1 | 62960250 | T | C |
| Apo-B | 1 | 62960575 | T | C |
| Apo-B | 1 | 63025942 | T | G |
| Apo-B | 1 | 63048276 | C | A |
| Apo-B | 1 | 63119256 | T | C |
| Apo-B | 1 | 63121101 | TC | T |
| Apo-B | 1 | 63135127 | G | A |
| Apo-B | 1 | 63177365 | T | C |
| Apo-B | 1 | 63177539 | T | G |
| Apo-B | 1 | 63191577 | C | A |
| Apo-B | 1 | 63191777 | T | C |
| Apo-B | 1 | 63193972 | G | A |
| Apo-B | 1 | 63195692 | T | C |
| Apo-B | 1 | 63203221 | T | C |
| Apo-B | 20 | 39142516 | G | A |
| Apo-B | 20 | 39183949 | T | C |
| Apo-B | 20 | 39193406 | T | C |
| Apo-B | 2 | 20900832 | T | C |
| Apo-B | 2 | 20901660 | T | G |
| Apo-B | 2 | 20903015 | T | C |
| Apo-B | 2 | 20918951 | TA | T |
| Apo-B | 2 | 20928577 | G | A |
| Apo-B | 2 | 20967410 | G | A |
| Apo-B | 2 | 21050729 | GA | G |
| Apo-B | 2 | 21074998 | G | A |
| Apo-B | 2 | 21076264 | TA | T |
| Apo-B | 2 | 21085410 | T | C |
| Apo-B | 2 | 21087323 | TCTC | T |

**Table S14:** Genetic variants used in the presented analyses. Note that due to coverage of the GWAS chip the instruments differed across considered outcomes *(continued)*

| Exposure | Chromosome | Base-pair position (Build 37) | Other allele | Effect allele |
|---|---|---|---|---|
| Apo-B | 2 | 21091409 | C | A |
| Apo-B | 2 | 21092488 | T | C |
| Apo-B | 2 | 21093112 | G | A |
| Apo-B | 2 | 21100426 | T | C |
| Apo-B | 2 | 21101046 | G | A |
| Apo-B | 2 | 21108159 | G | A |
| Apo-B | 2 | 21121464 | T | C |
| Apo-B | 2 | 21123352 | C | A |
| Apo-B | 2 | 21126936 | G | A |
| Apo-B | 2 | 21133883 | G | A |
| Apo-B | 2 | 21134011 | T | C |
| Apo-B | 2 | 21135577 | G | A |
| Apo-B | 2 | 21138066 | T | C |
| Apo-B | 2 | 21140540 | T | C |
| Apo-B | 2 | 21142994 | T | C |
| Apo-B | 2 | 21148274 | C | A |
| Apo-B | 2 | 21149424 | G | A |
| Apo-B | 2 | 21150787 | T | C |
| Apo-B | 2 | 21151917 | T | C |
| Apo-B | 2 | 21152755 | G | A |
| Apo-B | 2 | 21182116 | G | A |
| Apo-B | 2 | 21182710 | G | A |
| Apo-B | 2 | 21190024 | T | C |
| Apo-B | 2 | 21190209 | T | C |
| Apo-B | 2 | 21196112 | G | A |
| Apo-B | 2 | 21196346 | T | G |
| Apo-B | 2 | 21197159 | T | C |
| Apo-B | 2 | 21202959 | C | A |
| Apo-B | 2 | 21205920 | T | C |
| Apo-B | 2 | 21208211 | G | A |
| Apo-B | 2 | 21216815 | C | A |
| Apo-B | 2 | 21217490 | T | C |
| Apo-B | 2 | 21231524 | G | A |
| Apo-B | 2 | 21232804 | G | A |
| Apo-B | 2 | 21233972 | T | C |
| Apo-B | 2 | 21239661 | T | C |
| Apo-B | 2 | 21239667 | T | C |
| Apo-B | 2 | 21240031 | C | A |
| Apo-B | 2 | 21240364 | G | A |
| Apo-B | 2 | 21244000 | T | G |
| Apo-B | 2 | 21245329 | T | G |
| Apo-B | 2 | 21250914 | G | A |
| Apo-B | 2 | 21259562 | T | C |
| Apo-B | 2 | 21259612 | G | A |

**Table S14:** Genetic variants used in the presented analyses. Note that due to coverage of the GWAS chip the instruments differed across considered outcomes *(continued)*

| Exposure | Chromosome | Base-pair position (Build 37) | Other allele | Effect allele |
|---|---|---|---|---|
| Apo-B | 2 | 21262427 | G | A |
| Apo-B | 2 | 21263900 | G | A |
| Apo-B | 2 | 21266774 | GGCAGCGCCA | G |
| Apo-B | 2 | 21267210 | G | A |
| Apo-B | 2 | 21267461 | G | A |
| Apo-B | 2 | 21270554 | C | A |
| Apo-B | 2 | 21271323 | G | A |
| Apo-B | 2 | 21286057 | T | C |
| Apo-B | 2 | 21288226 | G | A |
| Apo-B | 2 | 21290067 | T | C |
| Apo-B | 2 | 21294975 | G | A |
| Apo-B | 2 | 21300770 | C | A |
| Apo-B | 2 | 21303616 | T | C |
| Apo-B | 2 | 21309335 | T | C |
| Apo-B | 2 | 21309856 | G | A |
| Apo-B | 2 | 21311541 | T | G |
| Apo-B | 2 | 21313577 | G | A |
| Apo-B | 2 | 21313778 | TC | T |
| Apo-B | 2 | 21323868 | CTGT | C |
| Apo-B | 2 | 21327042 | GCTAGTGCCTA | G |
| Apo-B | 2 | 21327635 | G | A |
| Apo-B | 2 | 21328825 | G | A |
| Apo-B | 2 | 21333932 | C | A |
| Apo-B | 2 | 21337306 | T | C |
| Apo-B | 2 | 21364562 | T | C |
| Apo-B | 2 | 21367696 | TTGAAGAC | T |
| Apo-B | 2 | 21368217 | G | A |
| Apo-B | 2 | 21371068 | T | C |
| Apo-B | 2 | 21372014 | G | A |
| Apo-B | 2 | 21378013 | CA | C |
| Apo-B | 2 | 21378453 | G | A |
| Apo-B | 2 | 21378805 | G | A |
| Apo-B | 2 | 21381177 | G | A |
| Apo-B | 2 | 21383353 | T | C |
| Apo-B | 2 | 21383514 | G | A |
| Apo-B | 2 | 21388401 | GA | G |
| Apo-B | 2 | 21389659 | T | G |
| Apo-B | 2 | 21393623 | G | A |
| Apo-B | 2 | 21399216 | T | G |
| Apo-B | 2 | 21406048 | T | C |
| Apo-B | 2 | 21410890 | T | C |
| Apo-B | 2 | 21413077 | G | A |
| Apo-B | 2 | 21414142 | T | C |
| Apo-B | 2 | 21415728 | T | G |

**Table S14:** Genetic variants used in the presented analyses. Note that due to coverage of the GWAS chip the instruments differed across considered outcomes *(continued)*

| Exposure | Chromosome | Base-pair position (Build 37) | Other allele | Effect allele |
|---|---|---|---|---|
| Apo-B | 2 | 21419650 | G | A |
| Apo-B | 2 | 21422928 | T | C |
| Apo-B | 2 | 21428049 | C | A |
| Apo-B | 2 | 21428185 | G | A |
| Apo-B | 2 | 21428289 | G | A |
| Apo-B | 2 | 21434580 | G | A |
| Apo-B | 2 | 21435700 | C | A |
| Apo-B | 2 | 21437321 | T | G |
| Apo-B | 2 | 21438738 | G | A |
| Apo-B | 2 | 21439329 | T | G |
| Apo-B | 2 | 21442944 | TTGCTCCCCTCTGCTTAGCA | T |
| Apo-B | 2 | 21445569 | G | A |
| Apo-B | 2 | 21448511 | G | A |
| Apo-B | 2 | 21449034 | T | C |
| Apo-B | 2 | 21449987 | G | A |
| Apo-B | 2 | 21450714 | G | A |
| Apo-B | 2 | 21451604 | T | C |
| Apo-B | 2 | 21451687 | G | A |
| Apo-B | 2 | 21453733 | G | A |
| Apo-B | 2 | 21457324 | G | A |
| Apo-B | 2 | 21463922 | T | C |
| Apo-B | 2 | 21467872 | ATT | A |
| Apo-B | 2 | 21469336 | T | C |
| Apo-B | 2 | 21471603 | T | C |
| Apo-B | 2 | 21471750 | G | A |
| Apo-B | 2 | 21476247 | C | A |
| Apo-B | 2 | 21476634 | G | A |
| Apo-B | 2 | 21524000 | G | A |
| Apo-B | 2 | 21530659 | G | A |
| Apo-B | 2 | 27574159 | T | C |
| Apo-B | 2 | 27591804 | T | C |
| Apo-B | 2 | 27598097 | T | C |
| Apo-B | 2 | 27635463 | G | A |
| Apo-B | 2 | 27648726 | T | C |
| Apo-B | 2 | 27678471 | G | A |
| Apo-B | 2 | 27685388 | G | A |
| Apo-B | 2 | 27730940 | T | C |
| Apo-B | 2 | 27742603 | T | C |
| Apo-B | 2 | 27748597 | C | A |
| Apo-B | 2 | 27748624 | G | A |
| Apo-B | 2 | 27750545 | G | A |
| Apo-B | 2 | 27752463 | G | A |
| Apo-B | 2 | 27752871 | T | C |
| Apo-B | 2 | 27988821 | G | A |

**Table S14:** Genetic variants used in the presented analyses. Note that due to coverage of the GWAS chip the instruments differed across considered outcomes *(continued)*

| Exposure | Chromosome | Base-pair position (Build 37) | Other allele | Effect allele |
|---|---|---|---|---|
| Apo-B | 2 | 44057570 | C | A |
| Apo-B | 2 | 44057825 | TCTC | T |
| Apo-B | 2 | 44058201 | G | A |
| Apo-B | 2 | 44073656 | T | C |
| Apo-B | 2 | 44073881 | T | C |
| Apo-B | 2 | 44074000 | G | A |
| Apo-B | 2 | 44088041 | C | A |
| Apo-B | 2 | 44092066 | T | C |
| Apo-B | 2 | 44096259 | T | G |
| Apo-B | 2 | 44096402 | G | A |
| Apo-B | 4 | 63875850 | T | C |
| Apo-B | 4 | 73406173 | G | A |
| Apo-B | 4 | 73499882 | T | C |
| Apo-B | 5 | 156367092 | T | C |
| Apo-B | 5 | 156381370 | G | A |
| Apo-B | 5 | 156390297 | T | C |
| Apo-B | 5 | 156443066 | G | A |
| Apo-B | 5 | 156482099 | T | C |
| Apo-B | 5 | 74302658 | G | A |
| Apo-B | 5 | 74340327 | C | A |
| Apo-B | 5 | 74343769 | G | A |
| Apo-B | 5 | 74352180 | G | A |
| Apo-B | 5 | 74354422 | G | A |
| Apo-B | 5 | 74372141 | T | C |
| Apo-B | 5 | 74379827 | G | A |
| Apo-B | 5 | 74382081 | G | A |
| Apo-B | 5 | 74384115 | G | A |
| Apo-B | 5 | 74389325 | C | A |
| Apo-B | 5 | 74443132 | T | C |
| Apo-B | 5 | 74461850 | T | G |
| Apo-B | 5 | 74461924 | C | A |
| Apo-B | 5 | 74480288 | T | C |
| Apo-B | 5 | 74536921 | G | A |
| Apo-B | 5 | 74537513 | G | A |
| Apo-B | 5 | 74547484 | C | A |
| Apo-B | 5 | 74558893 | G | A |
| Apo-B | 5 | 74562564 | T | C |
| Apo-B | 5 | 74565199 | GTCTCCGTCTGCC | |
| Apo-B | 5 | 74567359 | G | A |
| Apo-B | 5 | 74586866 | CT | C |
| Apo-B | 5 | 74587940 | C | A |
| Apo-B | 5 | 74596682 | T | G |
| Apo-B | 5 | 74603479 | G | A |
| Apo-B | 5 | 74604742 | G | A |

**Table S14:** Genetic variants used in the presented analyses. Note that due to coverage of the GWAS chip the instruments differed across considered outcomes *(continued)*

| Exposure | Chromosome | Base-pair position (Build 37) | Other allele | Effect allele |
|---|---|---|---|---|
| Apo-B | 5 | 74608732 | T | C |
| Apo-B | 5 | 74611232 | TG | T |
| Apo-B | 5 | 74613943 | T | C |
| Apo-B | 5 | 74615021 | T | C |
| Apo-B | 5 | 74616843 | T | C |
| Apo-B | 5 | 74617262 | G | A |
| Apo-B | 5 | 74619132 | T | C |
| Apo-B | 5 | 74622121 | TTA | T |
| Apo-B | 5 | 74625512 | G | A |
| Apo-B | 5 | 74629162 | CAT | C |
| Apo-B | 5 | 74630189 | T | C |
| Apo-B | 5 | 74630277 | T | G |
| Apo-B | 5 | 74631352 | T | C |
| Apo-B | 5 | 74644785 | TA | T |
| Apo-B | 5 | 74651084 | G | A |
| Apo-B | 5 | 74652754 | ATCCATCCGTCTGTCCG | A |
| Apo-B | 5 | 74653462 | T | G |
| Apo-B | 5 | 74656539 | T | C |
| Apo-B | 5 | 74660064 | ATTG | A |
| Apo-B | 5 | 74660356 | T | C |
| Apo-B | 5 | 74666492 | T | C |
| Apo-B | 5 | 74673707 | G | A |
| Apo-B | 5 | 74675076 | T | C |
| Apo-B | 5 | 74692776 | G | A |
| Apo-B | 5 | 74717653 | T | C |
| Apo-B | 5 | 74736600 | TA | T |
| Apo-B | 5 | 74745228 | T | G |
| Apo-B | 5 | 74753736 | T | C |
| Apo-B | 5 | 74808237 | T | G |
| Apo-B | 5 | 74808417 | G | A |
| Apo-B | 5 | 74810384 | G | A |
| Apo-B | 5 | 74816841 | CT | C |
| Apo-B | 5 | 74879890 | G | A |
| Apo-B | 5 | 74906963 | C | A |
| Apo-B | 5 | 74965122 | G | A |
| Apo-B | 5 | 74967386 | T | C |
| Apo-B | 7 | 21563471 | T | C |
| Apo-B | 7 | 21569089 | G | A |
| Apo-B | 7 | 21576905 | T | C |
| Apo-B | 7 | 21577960 | T | C |
| Apo-B | 7 | 21581561 | G | A |
| Apo-B | 7 | 21582917 | T | C |
| Apo-B | 7 | 21582963 | T | G |
| Apo-B | 7 | 21586595 | T | C |

**Table S14:** Genetic variants used in the presented analyses. Note that due to coverage of the GWAS chip the instruments differed across considered outcomes *(continued)*

| Exposure | Chromosome | Base-pair position (Build 37) | Other allele | Effect allele |
|---|---|---|---|---|
| Apo-B | 7 | 21588243 | C | A |
| Apo-B | 7 | 21589030 | T | C |
| Apo-B | 7 | 21589243 | G | A |
| Apo-B | 7 | 21589580 | G | A |
| Apo-B | 7 | 21592949 | G | A |
| Apo-B | 7 | 21595655 | T | C |
| Apo-B | 7 | 21596006 | G | A |
| Apo-B | 7 | 21597487 | C | A |
| Apo-B | 7 | 21597631 | T | G |
| Apo-B | 7 | 21599233 | T | C |
| Apo-B | 7 | 21601444 | T | C |
| Apo-B | 7 | 21601659 | GCTCT | G |
| Apo-B | 7 | 21603601 | G | A |
| Apo-B | 7 | 21603678 | G | A |
| Apo-B | 7 | 21607352 | T | C |
| Apo-B | 7 | 21610379 | G | A |
| Apo-B | 7 | 21627909 | G | A |
| Apo-B | 7 | 21629476 | G | A |
| Apo-B | 7 | 21631827 | G | A |
| Apo-B | 7 | 21637976 | G | A |
| Apo-B | 7 | 21647114 | T | G |
| Apo-B | 7 | 21650595 | G | A |
| Apo-B | 7 | 21670593 | G | A |
| Apo-B | 7 | 21670846 | G | A |
| Apo-B | 7 | 21686001 | T | C |
| Apo-B | 7 | 21691610 | T | G |
| Apo-B | 7 | 21693503 | G | A |
| Apo-B | 7 | 21693716 | G | A |
| Apo-B | 7 | 21693996 | T | C |
| Apo-B | 7 | 21702980 | G | A |
| Apo-B | 7 | 21712184 | T | C |
| Apo-B | 7 | 72856269 | G | A |
| Apo-B | 7 | 72971231 | T | C |
| Apo-B | 7 | 73029437 | T | C |
| Apo-B | 7 | 73046758 | C | A |
| Apo-B | 7 | 73046779 | G | A |
| Apo-B | 7 | 73047520 | T | C |
| Apo-B | 7 | 73050864 | G | A |
| Apo-B | 7 | 73051306 | T | C |
| Apo-B | 8 | 126447308 | T | C |
| Apo-B | 8 | 126475770 | G | A |
| Apo-B | 8 | 126476873 | T | C |
| Apo-B | 8 | 126479362 | T | C |
| Apo-B | 8 | 126480367 | T | C |

**Table S14:** Genetic variants used in the presented analyses. Note that due to coverage of the GWAS chip the instruments differed across considered outcomes *(continued)*

| Exposure | Chromosome | Base-pair position (Build 37) | Other allele | Effect allele |
|---|---|---|---|---|
| Apo-B | 8 | 126484526 | T | G |
| Apo-B | 8 | 126485531 | T | C |
| Apo-B | 8 | 126486409 | G | A |
| Apo-B | 8 | 126488930 | G | A |
| Apo-B | 8 | 126491304 | T | C |
| Apo-B | 8 | 126495818 | T | C |
| Apo-B | 8 | 126499878 | G | A |
| Apo-B | 8 | 126501177 | T | C |
| Apo-B | 8 | 126506694 | G | A |
| Apo-B | 8 | 145028505 | CG | C |
| Apo-B | 8 | 19816934 | T | C |
| Apo-B | 8 | 19817401 | G | A |
| Apo-B | 8 | 19818042 | G | A |
| Apo-B | 8 | 19819439 | G | A |
| Apo-B | 8 | 19820405 | G | A |
| Apo-B | 8 | 19824563 | T | C |
| Apo-B | 8 | 19824626 | CTT | C |
| Apo-B | 8 | 19830170 | T | G |
| Apo-B | 8 | 19830921 | T | C |
| Apo-B | 8 | 19838469 | T | C |
| Apo-B | 8 | 19839709 | T | C |
| Apo-B | 8 | 19843879 | T | C |
| Apo-B | 8 | 19844222 | G | A |
| Apo-B | 8 | 19849757 | G | A |
| Apo-B | 8 | 19849992 | T | C |
| Apo-B | 8 | 19852156 | T | G |
| Apo-B | 8 | 19856260 | T | G |
| Apo-B | 8 | 19868386 | C | A |
| Apo-B | 8 | 19868971 | T | C |
| Apo-B | 8 | 19874932 | AAGAC | A |
| Apo-B | 8 | 19882950 | T | C |
| Apo-B | 8 | 19912370 | G | A |
| Apo-B | 8 | 19928582 | G | A |
| Apo-B | 8 | 19929778 | T | G |
| Apo-B | 8 | 19931498 | G | A |
| Apo-B | 8 | 19932863 | T | C |
| Apo-B | 8 | 19936687 | G | A |
| Apo-B | 8 | 19938946 | T | C |
| Apo-B | 8 | 19939049 | T | C |
| Apo-B | 8 | 59395980 | CA | C |
| Apo-B | 9 | 136146597 | T | C |
| Apo-B | 9 | 136149399 | G | A |
| Apo-B | 9 | 136151806 | T | C |
| Apo-B | 9 | 136153981 | T | C |

**Table S14:** Genetic variants used in the presented analyses. Note that due to coverage of the GWAS chip the instruments differed across considered outcomes *(continued)*

| Exposure | Chromosome | Base-pair position (Build 37) | Other allele | Effect allele |
|---|---|---|---|---|
| Apo-B | 9 | 136154168 | T | C |
| Apo-B | 9 | 136154304 | T | C |
| Apo-B | 9 | 136184782 | G | A |
| Apo-B | 9 | 136267149 | T | C |
| Apo-A1 | 11 | 116701535 | T | C |
| Apo-A1 | 11 | 116705100 | T | G |
| Apo-A1 | 11 | 116705278 | T | C |
| Apo-A1 | 11 | 116705516 | G | A |
| Apo-A1 | 11 | 116711141 | T | C |
| Apo-A1 | 11 | 116728277 | G | A |
| Apo-A1 | 11 | 116752163 | T | C |
| Apo-A1 | 11 | 116772787 | C | A |
| Apo-A1 | 11 | 116972935 | T | C |
| Apo-A1 | 11 | 116975704 | T | G |
| Apo-A1 | 11 | 116981335 | G | A |
| Apo-A1 | 11 | 61547237 | T | C |
| Apo-A1 | 11 | 61564299 | T | G |
| Apo-A1 | 11 | 61580635 | T | C |
| Apo-A1 | 11 | 61581656 | G | A |
| Apo-A1 | 11 | 61582708 | T | C |
| Apo-A1 | 11 | 61586328 | CT | C |
| Apo-A1 | 11 | 61589481 | G | A |
| Apo-A1 | 11 | 61591995 | GAA | G |
| Apo-A1 | 11 | 61594920 | CT | C |
| Apo-A1 | 11 | 61597212 | T | C |
| Apo-A1 | 11 | 61597972 | G | A |
| Apo-A1 | 11 | 61601872 | T | C |
| Apo-A1 | 11 | 61601908 | G | A |
| Apo-A1 | 11 | 61603237 | T | C |
| Apo-A1 | 11 | 61603510 | C | A |
| Apo-A1 | 11 | 61605215 | G | A |
| Apo-A1 | 11 | 61609750 | T | C |
| Apo-A1 | 11 | 61611216 | T | C |
| Apo-A1 | 11 | 61616012 | G | A |
| Apo-A1 | 11 | 61618012 | T | C |
| Apo-A1 | 11 | 61618608 | G | A |
| Apo-A1 | 11 | 61620628 | AAACAAC | A |
| Apo-A1 | 11 | 61620703 | CTCTT | C |
| Apo-A1 | 11 | 61621194 | G | A |
| Apo-A1 | 11 | 61623793 | G | A |
| Apo-A1 | 11 | 61624181 | T | C |
| Apo-A1 | 11 | 61624414 | T | C |
| Apo-A1 | 11 | 61625723 | CA | C |
| Apo-A1 | 11 | 61626206 | G | A |

**Table S14:** Genetic variants used in the presented analyses. Note that due to coverage of the GWAS chip the instruments differed across considered outcomes *(continued)*

| Exposure | Chromosome | Base-pair position (Build 37) | Other allele | Effect allele |
|---|---|---|---|---|
| Apo-A1 | 11 | 61629122 | G | A |
| Apo-A1 | 11 | 61629666 | G | A |
| Apo-A1 | 11 | 61637587 | T | C |
| Apo-A1 | 11 | 61639488 | T | C |
| Apo-A1 | 11 | 61641542 | T | G |
| Apo-A1 | 11 | 61647427 | T | G |
| Apo-A1 | 11 | 61656117 | G | A |
| Apo-A1 | 11 | 61656182 | C | A |
| Apo-A1 | 11 | 61656995 | T | C |
| Apo-A1 | 11 | 61657110 | T | C |
| Apo-A1 | 11 | 61659296 | G | A |
| Apo-A1 | 11 | 61803540 | G | A |
| Apo-A1 | 12 | 109953417 | G | A |
| Apo-A1 | 12 | 125265201 | G | A |
| Apo-A1 | 12 | 125286650 | T | C |
| Apo-A1 | 12 | 125287216 | C | A |
| Apo-A1 | 12 | 125317391 | C | A |
| Apo-A1 | 12 | 125324798 | C | A |
| Apo-A1 | 12 | 125335510 | C | A |
| Apo-A1 | 12 | 125338336 | T | C |
| Apo-A1 | 12 | 125345448 | G | A |
| Apo-A1 | 12 | 125349480 | G | A |
| Apo-A1 | 12 | 125350849 | T | C |
| Apo-A1 | 12 | 125351116 | T | C |
| Apo-A1 | 12 | 125351149 | G | A |
| Apo-A1 | 12 | 125351772 | T | C |
| Apo-A1 | 12 | 125352603 | T | C |
| Apo-A1 | 12 | 125363987 | C | A |
| Apo-A1 | 12 | 125380232 | G | A |
| Apo-A1 | 12 | 47223857 | CAA | C |
| Apo-A1 | 12 | 79276836 | G | A |
| Apo-A1 | 13 | 66479744 | G | A |
| Apo-A1 | 15 | 58478950 | T | C |
| Apo-A1 | 15 | 58489359 | C | A |
| Apo-A1 | 15 | 58501481 | T | G |
| Apo-A1 | 15 | 58520259 | T | C |
| Apo-A1 | 15 | 58561006 | G | A |
| Apo-A1 | 15 | 58579956 | G | A |
| Apo-A1 | 15 | 58580525 | G | A |
| Apo-A1 | 15 | 58582294 | T | C |
| Apo-A1 | 15 | 58586129 | T | C |
| Apo-A1 | 15 | 58607320 | G | A |
| Apo-A1 | 15 | 58612816 | G | A |
| Apo-A1 | 15 | 58623632 | T | G |

**Table S14:** Genetic variants used in the presented analyses. Note that due to coverage of the GWAS chip the instruments differed across considered outcomes *(continued)*

| Exposure | Chromosome | Base-pair position (Build 37) | Other allele | Effect allele |
|---|---|---|---|---|
| Apo-A1 | 15 | 58624396 | T | C |
| Apo-A1 | 15 | 58630019 | T | G |
| Apo-A1 | 15 | 58641140 | G | A |
| Apo-A1 | 15 | 58661083 | T | C |
| Apo-A1 | 15 | 58669703 | C | A |
| Apo-A1 | 15 | 58671178 | C | A |
| Apo-A1 | 15 | 58671765 | T | G |
| Apo-A1 | 15 | 58672134 | T | C |
| Apo-A1 | 15 | 58674051 | G | A |
| Apo-A1 | 15 | 58674308 | G | A |
| Apo-A1 | 15 | 58678512 | T | C |
| Apo-A1 | 15 | 58679668 | G | A |
| Apo-A1 | 15 | 58679807 | CAGA | C |
| Apo-A1 | 15 | 58680638 | T | C |
| Apo-A1 | 15 | 58680805 | G | A |
| Apo-A1 | 15 | 58680954 | T | C |
| Apo-A1 | 15 | 58682029 | TGAA | T |
| Apo-A1 | 15 | 58682690 | G | A |
| Apo-A1 | 15 | 58683366 | G | A |
| Apo-A1 | 15 | 58683697 | T | C |
| Apo-A1 | 15 | 58684478 | T | C |
| Apo-A1 | 15 | 58684967 | T | G |
| Apo-A1 | 15 | 58685970 | T | C |
| Apo-A1 | 15 | 58686252 | G | A |
| Apo-A1 | 15 | 58686338 | G | A |
| Apo-A1 | 15 | 58687252 | GA | G |
| Apo-A1 | 15 | 58687530 | G | A |
| Apo-A1 | 15 | 58687603 | T | C |
| Apo-A1 | 15 | 58687862 | G | A |
| Apo-A1 | 15 | 58687880 | T | C |
| Apo-A1 | 15 | 58688477 | G | A |
| Apo-A1 | 15 | 58688478 | C | A |
| Apo-A1 | 15 | 58689187 | T | C |
| Apo-A1 | 15 | 58690754 | G | A |
| Apo-A1 | 15 | 58691202 | G | A |
| Apo-A1 | 15 | 58692007 | G | A |
| Apo-A1 | 15 | 58692202 | C | A |
| Apo-A1 | 15 | 58692479 | T | C |
| Apo-A1 | 15 | 58692936 | T | C |
| Apo-A1 | 15 | 58693860 | T | C |
| Apo-A1 | 15 | 58694020 | T | G |
| Apo-A1 | 15 | 58696300 | T | C |
| Apo-A1 | 15 | 58697072 | T | C |
| Apo-A1 | 15 | 58698113 | T | G |

**Table S14:** Genetic variants used in the presented analyses. Note that due to coverage of the GWAS chip the instruments differed across considered outcomes *(continued)*

| Exposure | Chromosome | Base-pair position (Build 37) | Other allele | Effect allele |
|---|---|---|---|---|
| Apo-A1 | 15 | 58698243 | T | G |
| Apo-A1 | 15 | 58698755 | T | G |
| Apo-A1 | 15 | 58699010 | T | C |
| Apo-A1 | 15 | 58699937 | T | C |
| Apo-A1 | 15 | 58702325 | G | A |
| Apo-A1 | 15 | 58703069 | C | A |
| Apo-A1 | 15 | 58703477 | G | A |
| Apo-A1 | 15 | 58703895 | G | A |
| Apo-A1 | 15 | 58705029 | T | C |
| Apo-A1 | 15 | 58705658 | AC | A |
| Apo-A1 | 15 | 58706797 | G | A |
| Apo-A1 | 15 | 58709181 | T | C |
| Apo-A1 | 15 | 58709182 | G | A |
| Apo-A1 | 15 | 58709725 | T | G |
| Apo-A1 | 15 | 58709750 | G | A |
| Apo-A1 | 15 | 58710627 | C | A |
| Apo-A1 | 15 | 58710736 | T | C |
| Apo-A1 | 15 | 58712203 | T | G |
| Apo-A1 | 15 | 58712945 | C | A |
| Apo-A1 | 15 | 58712973 | T | C |
| Apo-A1 | 15 | 58716044 | G | A |
| Apo-A1 | 15 | 58716257 | G | A |
| Apo-A1 | 15 | 58716615 | T | G |
| Apo-A1 | 15 | 58718529 | CA | C |
| Apo-A1 | 15 | 58719741 | G | A |
| Apo-A1 | 15 | 58720262 | T | C |
| Apo-A1 | 15 | 58720405 | G | A |
| Apo-A1 | 15 | 58720431 | C | A |
| Apo-A1 | 15 | 58721447 | G | A |
| Apo-A1 | 15 | 58721614 | G | A |
| Apo-A1 | 15 | 58722590 | GC | G |
| Apo-A1 | 15 | 58723479 | T | C |
| Apo-A1 | 15 | 58723675 | T | C |
| Apo-A1 | 15 | 58724706 | G | A |
| Apo-A1 | 15 | 58727279 | T | C |
| Apo-A1 | 15 | 58727325 | G | A |
| Apo-A1 | 15 | 58727687 | T | C |
| Apo-A1 | 15 | 58728067 | G | A |
| Apo-A1 | 15 | 58729144 | T | C |
| Apo-A1 | 15 | 58730498 | T | C |
| Apo-A1 | 15 | 58731455 | T | C |
| Apo-A1 | 15 | 58731567 | G | A |
| Apo-A1 | 15 | 58737341 | T | G |
| Apo-A1 | 15 | 58738423 | T | C |

**Table S14:** Genetic variants used in the presented analyses. Note that due to coverage of the GWAS chip the instruments differed across considered outcomes *(continued)*

| Exposure | Chromosome | Base-pair position (Build 37) | Other allele | Effect allele |
|---|---|---|---|---|
| Apo-A1 | 15 | 58738760 | G | A |
| Apo-A1 | 15 | 58741891 | G | A |
| Apo-A1 | 15 | 58742418 | G | A |
| Apo-A1 | 15 | 58742731 | T | G |
| Apo-A1 | 15 | 58882792 | T | C |
| Apo-A1 | 15 | 58899609 | T | C |
| Apo-A1 | 15 | 58931340 | G | A |
| Apo-A1 | 15 | 59080862 | C | A |
| Apo-A1 | 15 | 59531818 | T | C |
| Apo-A1 | 15 | 59566385 | T | C |
| Apo-A1 | 15 | 59806496 | T | G |
| Apo-A1 | 16 | 56745752 | T | C |
| Apo-A1 | 16 | 56759532 | G | A |
| Apo-A1 | 16 | 56811400 | G | A |
| Apo-A1 | 16 | 56819919 | T | C |
| Apo-A1 | 16 | 56827946 | G | A |
| Apo-A1 | 16 | 56846804 | T | C |
| Apo-A1 | 16 | 56899006 | T | C |
| Apo-A1 | 16 | 56900100 | G | A |
| Apo-A1 | 16 | 56909598 | T | C |
| Apo-A1 | 16 | 56910692 | T | G |
| Apo-A1 | 16 | 56927296 | T | C |
| Apo-A1 | 16 | 56929944 | G | A |
| Apo-A1 | 16 | 56938955 | C | A |
| Apo-A1 | 16 | 56945049 | C | A |
| Apo-A1 | 16 | 56984590 | T | C |
| Apo-A1 | 16 | 56985139 | G | A |
| Apo-A1 | 16 | 56985156 | G | A |
| Apo-A1 | 16 | 56985514 | G | A |
| Apo-A1 | 16 | 56985555 | G | A |
| Apo-A1 | 16 | 56985805 | T | C |
| Apo-A1 | 16 | 56986762 | T | G |
| Apo-A1 | 16 | 56988044 | T | C |
| Apo-A1 | 16 | 56988502 | G | A |
| Apo-A1 | 16 | 56990716 | C | A |
| Apo-A1 | 16 | 56991363 | T | C |
| Apo-A1 | 16 | 56991524 | G | A |
| Apo-A1 | 16 | 56991741 | T | C |
| Apo-A1 | 16 | 56992017 | G | A |
| Apo-A1 | 16 | 56993025 | T | G |
| Apo-A1 | 16 | 56993211 | T | C |
| Apo-A1 | 16 | 56993324 | C | A |
| Apo-A1 | 16 | 56993897 | C | A |
| Apo-A1 | 16 | 56993901 | C | A |

**Table S14:** Genetic variants used in the presented analyses. Note that due to coverage of the GWAS chip the instruments differed across considered outcomes *(continued)*

| Exposure | Chromosome | Base-pair position (Build 37) | Other allele | Effect allele |
|---|---|---|---|---|
| Apo-A1 | 16 | 56993909 | C | A |
| Apo-A1 | 16 | 56994192 | T | C |
| Apo-A1 | 16 | 56994244 | TA | T |
| Apo-A1 | 16 | 56994894 | G | A |
| Apo-A1 | 16 | 56995038 | T | C |
| Apo-A1 | 16 | 56995236 | C | A |
| Apo-A1 | 16 | 56996158 | T | C |
| Apo-A1 | 16 | 56996211 | G | A |
| Apo-A1 | 16 | 56996288 | G | A |
| Apo-A1 | 16 | 56996645 | GCC | G |
| Apo-A1 | 16 | 56997233 | G | A |
| Apo-A1 | 16 | 56997349 | CA | C |
| Apo-A1 | 16 | 56998918 | G | A |
| Apo-A1 | 16 | 56999258 | C | A |
| Apo-A1 | 16 | 56999328 | T | C |
| Apo-A1 | 16 | 57000284 | G | A |
| Apo-A1 | 16 | 57000938 | G | A |
| Apo-A1 | 16 | 57001254 | TCACA | T |
| Apo-A1 | 16 | 57001274 | AC | A |
| Apo-A1 | 16 | 57001985 | C | A |
| Apo-A1 | 16 | 57002404 | C | A |
| Apo-A1 | 16 | 57002732 | T | G |
| Apo-A1 | 16 | 57003146 | T | C |
| Apo-A1 | 16 | 57005479 | C | A |
| Apo-A1 | 16 | 57006590 | T | C |
| Apo-A1 | 16 | 57007353 | T | C |
| Apo-A1 | 16 | 57007451 | G | A |
| Apo-A1 | 16 | 57008227 | G | A |
| Apo-A1 | 16 | 57009651 | TC | T |
| Apo-A1 | 16 | 57010232 | T | C |
| Apo-A1 | 16 | 57010486 | T | C |
| Apo-A1 | 16 | 57011185 | G | A |
| Apo-A1 | 16 | 57012699 | G | A |
| Apo-A1 | 16 | 57014875 | G | A |
| Apo-A1 | 16 | 57016092 | G | A |
| Apo-A1 | 16 | 57017002 | T | G |
| Apo-A1 | 16 | 57017319 | G | A |
| Apo-A1 | 16 | 57019392 | T | C |
| Apo-A1 | 16 | 67675409 | TA | T |
| Apo-A1 | 16 | 68021125 | C | A |
| Apo-A1 | 16 | 68073644 | CAAGG | C |
| Apo-A1 | 16 | 68127222 | TAG | T |
| Apo-A1 | 16 | 68305702 | TTTTTC | T |
| Apo-A1 | 16 | 68305708 | TTTTC | T |

**Table S14:** Genetic variants used in the presented analyses. Note that due to coverage of the GWAS chip the instruments differed across considered outcomes *(continued)*

| Exposure | Chromosome | Base-pair position (Build 37) | Other allele | Effect allele |
|---|---|---|---|---|
| Apo-A1 | 17 | 45531884 | T | G |
| Apo-A1 | 17 | 45542260 | T | G |
| Apo-A1 | 18 | 46529282 | T | G |
| Apo-A1 | 18 | 46704724 | ACAT | A |
| Apo-A1 | 18 | 46723931 | CT | C |
| Apo-A1 | 18 | 47049700 | G | A |
| Apo-A1 | 18 | 47062054 | T | G |
| Apo-A1 | 18 | 47066144 | C | A |
| Apo-A1 | 18 | 47068878 | T | G |
| Apo-A1 | 18 | 47085440 | T | C |
| Apo-A1 | 18 | 47093864 | T | C |
| Apo-A1 | 18 | 47096016 | G | A |
| Apo-A1 | 18 | 47109955 | G | A |
| Apo-A1 | 18 | 47118219 | T | C |
| Apo-A1 | 18 | 47122158 | C | A |
| Apo-A1 | 18 | 47126133 | T | C |
| Apo-A1 | 18 | 47126999 | G | A |
| Apo-A1 | 18 | 47128176 | T | C |
| Apo-A1 | 18 | 47129148 | T | C |
| Apo-A1 | 18 | 47131781 | G | A |
| Apo-A1 | 18 | 47132464 | T | C |
| Apo-A1 | 18 | 47133828 | G | A |
| Apo-A1 | 18 | 47134066 | T | C |
| Apo-A1 | 18 | 47134175 | T | C |
| Apo-A1 | 18 | 47142243 | G | A |
| Apo-A1 | 18 | 47143458 | T | C |
| Apo-A1 | 18 | 47145202 | G | A |
| Apo-A1 | 18 | 47146125 | G | A |
| Apo-A1 | 18 | 47147746 | G | A |
| Apo-A1 | 18 | 47148299 | T | C |
| Apo-A1 | 18 | 47148886 | T | C |
| Apo-A1 | 18 | 47149371 | G | A |
| Apo-A1 | 18 | 47151264 | G | A |
| Apo-A1 | 18 | 47157093 | T | C |
| Apo-A1 | 18 | 47157439 | T | C |
| Apo-A1 | 18 | 47159090 | G | A |
| Apo-A1 | 18 | 47159917 | C | A |
| Apo-A1 | 18 | 47160814 | G | A |
| Apo-A1 | 18 | 47160953 | T | G |
| Apo-A1 | 18 | 47161733 | T | C |
| Apo-A1 | 18 | 47161892 | T | C |
| Apo-A1 | 18 | 47166694 | G | A |
| Apo-A1 | 18 | 47168648 | G | A |
| Apo-A1 | 18 | 47169340 | T | G |

**Table S14:** Genetic variants used in the presented analyses. Note that due to coverage of the GWAS chip the instruments differed across considered outcomes *(continued)*

| Exposure | Chromosome | Base-pair position (Build 37) | Other allele | Effect allele |
|---|---|---|---|---|
| Apo-A1 | 18 | 47170139 | ACCTG | A |
| Apo-A1 | 18 | 47175670 | C | A |
| Apo-A1 | 18 | 47176261 | C | A |
| Apo-A1 | 18 | 47176793 | G | A |
| Apo-A1 | 18 | 47176911 | G | A |
| Apo-A1 | 18 | 47176917 | T | C |
| Apo-A1 | 18 | 47179516 | G | A |
| Apo-A1 | 18 | 47182664 | G | A |
| Apo-A1 | 18 | 47183012 | T | C |
| Apo-A1 | 18 | 47196870 | G | A |
| Apo-A1 | 18 | 47227334 | G | A |
| Apo-A1 | 18 | 47243912 | C | A |
| Apo-A1 | 18 | 47385799 | G | A |
| Apo-A1 | 19 | 11190549 | G | A |
| Apo-A1 | 19 | 11190652 | G | A |
| Apo-A1 | 19 | 11202306 | T | G |
| Apo-A1 | 19 | 11206729 | T | C |
| Apo-A1 | 19 | 11206969 | G | A |
| Apo-A1 | 19 | 11210912 | T | C |
| Apo-A1 | 19 | 11317744 | T | C |
| Apo-A1 | 19 | 11330942 | G | A |
| Apo-A1 | 19 | 11331354 | T | G |
| Apo-A1 | 19 | 11341029 | T | C |
| Apo-A1 | 19 | 11346550 | T | C |
| Apo-A1 | 19 | 11347493 | T | C |
| Apo-A1 | 19 | 11348521 | G | A |
| Apo-A1 | 19 | 45296806 | T | C |
| Apo-A1 | 19 | 45302778 | G | A |
| Apo-A1 | 19 | 45318351 | G | A |
| Apo-A1 | 19 | 45369203 | G | A |
| Apo-A1 | 19 | 45389596 | G | A |
| Apo-A1 | 19 | 45408628 | T | C |
| Apo-A1 | 19 | 45412079 | T | C |
| Apo-A1 | 19 | 45415640 | G | A |
| Apo-A1 | 19 | 45416831 | T | G |
| Apo-A1 | 19 | 45426792 | G | A |
| Apo-A1 | 1 | 220967683 | G | A |
| Apo-A1 | 1 | 220970028 | G | A |
| Apo-A1 | 1 | 220970499 | G | A |
| Apo-A1 | 1 | 220973563 | T | C |
| Apo-A1 | 1 | 220974545 | T | C |
| Apo-A1 | 1 | 226064353 | TTTGG | T |
| Apo-A1 | 1 | 62901807 | C | A |
| Apo-A1 | 1 | 62902874 | G | A |

**Table S14:** Genetic variants used in the presented analyses. Note that due to coverage of the GWAS chip the instruments differed across considered outcomes *(continued)*

| Exposure | Chromosome | Base-pair position (Build 37) | Other allele | Effect allele |
|---|---|---|---|---|
| Apo-A1 | 1 | 62931632 | C | A |
| Apo-A1 | 1 | 62934002 | G | A |
| Apo-A1 | 1 | 62960250 | T | C |
| Apo-A1 | 1 | 62960575 | T | C |
| Apo-A1 | 1 | 63048276 | C | A |
| Apo-A1 | 1 | 63083289 | C | A |
| Apo-A1 | 1 | 63124542 | T | C |
| Apo-A1 | 1 | 63171063 | T | C |
| Apo-A1 | 1 | 63171199 | T | C |
| Apo-A1 | 1 | 63177539 | T | G |
| Apo-A1 | 20 | 43042364 | T | C |
| Apo-A1 | 22 | 21916166 | T | C |
| Apo-A1 | 22 | 21966442 | G | A |
| Apo-A1 | 22 | 21971010 | G | A |
| Apo-A1 | 22 | 21982892 | T | C |
| Apo-A1 | 22 | 21984379 | G | A |
| Apo-A1 | 2 | 136381348 | G | A |
| Apo-A1 | 2 | 136479302 | T | C |
| Apo-A1 | 2 | 136555659 | T | C |
| Apo-A1 | 2 | 136583192 | G | A |
| Apo-A1 | 2 | 136698098 | T | C |
| Apo-A1 | 2 | 136707982 | T | C |
| Apo-A1 | 2 | 136765951 | T | C |
| Apo-A1 | 2 | 136783169 | G | A |
| Apo-A1 | 2 | 136796019 | G | A |
| Apo-A1 | 2 | 136800195 | T | C |
| Apo-A1 | 2 | 213190035 | T | C |
| Apo-A1 | 4 | 103146888 | C | A |
| Apo-A1 | 4 | 103188709 | T | C |
| Apo-A1 | 4 | 69310287 | T | C |
| Apo-A1 | 4 | 69331331 | GT | G |
| Apo-A1 | 4 | 69339257 | G | A |
| Apo-A1 | 4 | 69374720 | T | C |
| Apo-A1 | 4 | 69374819 | T | C |
| Apo-A1 | 4 | 69375985 | T | C |
| Apo-A1 | 4 | 69384338 | AT | A |
| Apo-A1 | 4 | 69388595 | G | A |
| Apo-A1 | 4 | 69393588 | C | A |
| Apo-A1 | 4 | 69403830 | G | A |
| Apo-A1 | 4 | 69420232 | G | A |
| Apo-A1 | 4 | 69433714 | T | C |
| Apo-A1 | 4 | 69457410 | T | C |
| Apo-A1 | 4 | 69459490 | T | C |
| Apo-A1 | 4 | 69477030 | TAA | T |

**Table S14:** Genetic variants used in the presented analyses. Note that due to coverage of the GWAS chip the instruments differed across considered outcomes *(continued)*

| Exposure | Chromosome | Base-pair position (Build 37) | Other allele | Effect allele |
|---|---|---|---|---|
| Apo-A1 | 4 | 73406173 | G | A |
| Apo-A1 | 7 | 1067403 | G | A |
| Apo-A1 | 7 | 1091625 | G | A |
| Apo-A1 | 7 | 1096846 | G | A |
| Apo-A1 | 7 | 1097183 | G | A |
| Apo-A1 | 8 | 19734951 | G | A |
| Apo-A1 | 8 | 19748448 | T | C |
| Apo-A1 | 8 | 19755175 | C | A |
| Apo-A1 | 8 | 19755626 | C | A |
| Apo-A1 | 8 | 19755977 | T | G |
| Apo-A1 | 8 | 19756592 | T | C |
| Apo-A1 | 8 | 19767203 | T | C |
| Apo-A1 | 8 | 19774890 | C | A |
| Apo-A1 | 8 | 19776981 | C | A |
| Apo-A1 | 8 | 19786891 | T | C |
| Apo-A1 | 8 | 19791071 | T | C |
| Apo-A1 | 8 | 19791346 | T | C |
| Apo-A1 | 8 | 19793581 | T | C |
| Apo-A1 | 8 | 19803093 | T | C |
| Apo-A1 | 8 | 19812812 | T | C |
| Apo-A1 | 8 | 19813180 | G | A |
| Apo-A1 | 8 | 19813529 | G | A |
| Apo-A1 | 8 | 19813702 | G | A |
| Apo-A1 | 8 | 19815619 | T | C |
| Apo-A1 | 8 | 19816934 | T | C |
| Apo-A1 | 8 | 19818042 | G | A |
| Apo-A1 | 8 | 19819077 | T | G |
| Apo-A1 | 8 | 19819439 | G | A |
| Apo-A1 | 8 | 19819536 | T | G |
| Apo-A1 | 8 | 19820405 | G | A |
| Apo-A1 | 8 | 19824563 | T | C |
| Apo-A1 | 8 | 19826318 | G | A |
| Apo-A1 | 8 | 19830170 | T | G |
| Apo-A1 | 8 | 19830921 | T | C |
| Apo-A1 | 8 | 19838469 | T | C |
| Apo-A1 | 8 | 19839709 | T | C |
| Apo-A1 | 8 | 19843861 | G | A |
| Apo-A1 | 8 | 19843879 | T | C |
| Apo-A1 | 8 | 19844222 | G | A |
| Apo-A1 | 8 | 19846580 | C | A |
| Apo-A1 | 8 | 19849757 | G | A |
| Apo-A1 | 8 | 19849992 | T | C |
| Apo-A1 | 8 | 19856260 | T | G |
| Apo-A1 | 8 | 19865843 | G | A |

**Table S14:** Genetic variants used in the presented analyses. Note that due to coverage of the GWAS chip the instruments differed across considered outcomes *(continued)*

| Exposure | Chromosome | Base-pair position (Build 37) | Other allele | Effect allele |
|---|---|---|---|---|
| Apo-A1 | 8 | 19868386 | C | A |
| Apo-A1 | 8 | 19868971 | T | C |
| Apo-A1 | 8 | 19870271 | C | A |
| Apo-A1 | 8 | 19870693 | G | A |
| Apo-A1 | 8 | 19875004 | G | A |
| Apo-A1 | 8 | 19875100 | T | C |
| Apo-A1 | 8 | 19875408 | G | A |
| Apo-A1 | 8 | 19889399 | T | G |
| Apo-A1 | 8 | 19912370 | G | A |
| Apo-A1 | 8 | 19916120 | G | A |
| Apo-A1 | 8 | 19919655 | T | C |
| Apo-A1 | 8 | 19931087 | T | C |
| Apo-A1 | 8 | 19934339 | T | C |
| Apo-A1 | 8 | 19939049 | T | C |
| Apo-A1 | 8 | 9177732 | T | G |
| Apo-A1 | 8 | 9179298 | T | G |
| Apo-A1 | 8 | 9181611 | G | A |
| Apo-A1 | 8 | 9183596 | G | A |
| Apo-A1 | 8 | 9184691 | T | C |
| Apo-A1 | 8 | 9185146 | T | C |
| Apo-A1 | 8 | 9190564 | T | C |
| Apo-A1 | 8 | 9192423 | G | A |
| Apo-A1 | 8 | 9201787 | T | C |
| Apo-A1 | 8 | 9206069 | G | A |
| Apo-A1 | 8 | 9206678 | G | A |
| Apo-A1 | 8 | 9219868 | G | A |
| Apo-A1 | 8 | 9219869 | G | A |
| Apo-A1 | 8 | 9231661 | G | A |
| Apo-A1 | 9 | 107629058 | T | C |
| Apo-A1 | 9 | 107629336 | T | C |
| Apo-A1 | 9 | 107631134 | G | A |
| Apo-A1 | 9 | 107632644 | G | A |
| Apo-A1 | 9 | 107635869 | T | C |
| Apo-A1 | 9 | 107637306 | G | A |
| Apo-A1 | 9 | 107638969 | AG | A |
| Apo-A1 | 9 | 107639180 | G | A |
| Apo-A1 | 9 | 107645080 | T | C |
| Apo-A1 | 9 | 107645922 | T | C |
| Apo-A1 | 9 | 107649557 | AAAAC | A |
| Apo-A1 | 9 | 107650345 | T | G |
| Apo-A1 | 9 | 107650355 | T | G |
| Apo-A1 | 9 | 107650820 | T | C |
| Apo-A1 | 9 | 107651526 | G | A |
| Apo-A1 | 9 | 107656122 | C | A |

**Table S14:** Genetic variants used in the presented analyses. Note that due to coverage of the GWAS chip the instruments differed across considered outcomes *(continued)*

| Exposure | Chromosome | Base-pair position (Build 37) | Other allele | Effect allele |
|---|---|---|---|---|
| Apo-A1 | 9 | 107656409 | GC | G |
| Apo-A1 | 9 | 107657070 | G | A |
| Apo-A1 | 9 | 107658910 | C | A |
| Apo-A1 | 9 | 107661742 | C | A |
| Apo-A1 | 9 | 107664301 | T | C |
| Apo-A1 | 9 | 107665739 | G | A |
| Apo-A1 | 9 | 107669073 | T | C |
| Apo-A1 | 9 | 107669241 | T | C |
| Apo-A1 | 9 | 107672526 | T | G |
| Apo-A1 | 9 | 15285448 | T | G |
| Apo-A1 | 9 | 15285670 | G | A |
| Apo-A1 | 9 | 15286386 | T | C |
| Apo-A1 | 9 | 15287389 | G | A |
| Apo-A1 | 9 | 15289578 | T | C |
| Apo-A1 | 9 | 15293427 | G | A |
| Apo-A1 | 9 | 15294596 | G | A |
| Apo-A1 | 9 | 15296034 | C | A |
| Apo-A1 | 9 | 15297629 | G | A |
| Apo-A1 | 9 | 15298666 | T | C |
| Apo-A1 | 9 | 15301428 | C | A |
| Apo-B (cis) | 2 | 21182116 | G | A |
| Apo-B (cis) | 2 | 21182710 | G | A |
| Apo-B (cis) | 2 | 21184764 | G | A |
| Apo-B (cis) | 2 | 21187641 | T | C |
| Apo-B (cis) | 2 | 21190024 | T | C |
| Apo-B (cis) | 2 | 21190209 | T | C |
| Apo-B (cis) | 2 | 21190288 | CGCCT | C |
| Apo-B (cis) | 2 | 21193946 | G | A |
| Apo-B (cis) | 2 | 21196112 | G | A |
| Apo-B (cis) | 2 | 21196346 | T | G |
| Apo-B (cis) | 2 | 21197159 | T | C |
| Apo-B (cis) | 2 | 21202959 | C | A |
| Apo-B (cis) | 2 | 21205563 | G | A |
| Apo-B (cis) | 2 | 21205920 | T | C |
| Apo-B (cis) | 2 | 21208211 | G | A |
| Apo-B (cis) | 2 | 21216815 | C | A |
| Apo-B (cis) | 2 | 21217490 | T | C |
| Apo-B (cis) | 2 | 21231524 | G | A |
| Apo-B (cis) | 2 | 21232804 | G | A |
| Apo-B (cis) | 2 | 21233972 | T | C |
| Apo-B (cis) | 2 | 21239667 | T | C |
| Apo-B (cis) | 2 | 21240031 | C | A |
| Apo-B (cis) | 2 | 21240364 | G | A |
| Apo-B (cis) | 2 | 21244000 | T | G |

**Table S14:** Genetic variants used in the presented analyses. Note that due to coverage of the GWAS chip the instruments differed across considered outcomes *(continued)*

| Exposure | Chromosome | Base-pair position (Build 37) | Other allele | Effect allele |
|---|---|---|---|---|
| Apo-B (cis) | 2 | 21245329 | T | G |
| Apo-B (cis) | 2 | 21245367 | G | A |
| Apo-B (cis) | 2 | 21250914 | G | A |
| Apo-B (cis) | 2 | 21251040 | T | G |
| Apo-B (cis) | 2 | 21259562 | T | C |
| Apo-B (cis) | 2 | 21259612 | G | A |
| Apo-B (cis) | 2 | 21260601 | G | A |
| Apo-B (cis) | 2 | 21262427 | G | A |
| Apo-B (cis) | 2 | 21263900 | G | A |
| Apo-B (cis) | 2 | 21266774 | GGCAGCGCCA | G |
| Apo-B (cis) | 2 | 21267210 | G | A |
| Apo-B (cis) | 2 | 21267461 | G | A |
| Apo-B (cis) | 2 | 21269809 | G | A |
| Apo-B (cis) | 2 | 21270057 | G | A |
| Apo-B (cis) | 2 | 21270554 | C | A |
| Apo-B (cis) | 2 | 21270751 | G | A |
| Apo-B (cis) | 2 | 21271323 | G | A |
| Apo-B (cis) | 2 | 21272763 | AG | A |
| Apo-B (cis) | 2 | 21286057 | T | C |
| Apo-B (cis) | 2 | 21288226 | G | A |
| Apo-B (cis) | 2 | 21288970 | C | A |
| Apo-B (cis) | 2 | 21294975 | G | A |
| Apo-B (cis) | 2 | 21295065 | T | C |
| Apo-B (cis) | 2 | 21299935 | TATCCTGCA | T |
| Apo-B (cis) | 2 | 21300770 | C | A |
| Apo-B (cis) | 2 | 21301892 | T | C |
| Apo-B (cis) | 2 | 21302020 | T | C |
| Apo-B (cis) | 2 | 21303470 | T | C |
| Apo-B (cis) | 2 | 21303616 | T | C |
| Apo-B (cis) | 2 | 21309335 | T | C |
| Apo-B (cis) | 2 | 21311541 | T | G |
| Apo-B (cis) | 2 | 21312210 | T | C |
| Apo-B (cis) | 2 | 21313577 | G | A |
| Apo-B (cis) | 2 | 21313778 | TC | T |
| Apo-B (cis) | 2 | 21314851 | G | A |
| Apo-B (cis) | 2 | 21315664 | T | C |
| Apo-A1 (cis) | 11 | 116686288 | T | C |
| Apo-A1 (cis) | 11 | 116692334 | T | C |
| Apo-A1 (cis) | 11 | 116700142 | T | C |
| Apo-A1 (cis) | 11 | 116701535 | T | C |
| Apo-A1 (cis) | 11 | 116703671 | T | G |
| Apo-A1 (cis) | 11 | 116705100 | T | G |
| Apo-A1 (cis) | 11 | 116705278 | T | C |
| Apo-A1 (cis) | 11 | 116705516 | G | A |

**Table S14:** Genetic variants used in the presented analyses. Note that due to coverage of the GWAS chip the instruments differed across considered outcomes *(continued)*

| Exposure | Chromosome | Base-pair position (Build 37) | Other allele | Effect allele |
|---|---|---|---|---|
| Apo-A1 (cis) | 11 | 116708020 | G | A |
| Apo-A1 (cis) | 11 | 116710167 | CAT | C |
| Apo-A1 (cis) | 11 | 116710968 | G | A |
| Apo-A1 (cis) | 11 | 116711141 | T | C |
| Apo-A1 (cis) | 11 | 116711180 | G | A |
| Apo-A1 (cis) | 11 | 116711974 | T | C |
| Apo-A1 (cis) | 11 | 116720089 | T | C |
| Apo-A1 (cis) | 11 | 116728277 | G | A |
| Apo-A1 (cis) | 11 | 116733565 | G | A |
| Apo-A1 (cis) | 11 | 116734545 | G | A |
| Apo-A1 (cis) | 11 | 116741553 | G | A |
| Apo-A1 (cis) | 11 | 116741572 | C | A |
| Apo-A1 (cis) | 11 | 116744018 | T | C |
| Apo-A1 (cis) | 11 | 116750874 | C | A |
| Apo-A1 (cis) | 11 | 116752163 | T | C |
| Apo-A1 (cis) | 11 | 116755584 | G | A |
| LDL-C (GLGC) | 10 | 113902418 | C | A |
| LDL-C (GLGC) | 10 | 113913222 | T | C |
| LDL-C (GLGC) | 10 | 17260290 | G | A |
| LDL-C (GLGC) | 10 | 94762340 | G | A |
| LDL-C (GLGC) | 11 | 116519739 | T | C |
| LDL-C (GLGC) | 11 | 116522330 | T | C |
| LDL-C (GLGC) | 11 | 116525730 | T | C |
| LDL-C (GLGC) | 11 | 116531739 | T | C |
| LDL-C (GLGC) | 11 | 116574488 | G | A |
| LDL-C (GLGC) | 11 | 116584987 | T | C |
| LDL-C (GLGC) | 11 | 116586283 | T | C |
| LDL-C (GLGC) | 11 | 116596309 | C | A |
| LDL-C (GLGC) | 11 | 116603724 | C | A |
| LDL-C (GLGC) | 11 | 116604514 | G | A |
| LDL-C (GLGC) | 11 | 116607368 | C | A |
| LDL-C (GLGC) | 11 | 116610249 | G | A |
| LDL-C (GLGC) | 11 | 116612128 | G | A |
| LDL-C (GLGC) | 11 | 116613660 | T | C |
| LDL-C (GLGC) | 11 | 116617240 | C | A |
| LDL-C (GLGC) | 11 | 116623659 | T | C |
| LDL-C (GLGC) | 11 | 116624703 | T | G |
| LDL-C (GLGC) | 11 | 116645275 | T | C |
| LDL-C (GLGC) | 11 | 116646858 | T | C |
| LDL-C (GLGC) | 11 | 116649135 | G | A |
| LDL-C (GLGC) | 11 | 116652301 | T | C |
| LDL-C (GLGC) | 11 | 116652351 | G | A |
| LDL-C (GLGC) | 11 | 116652423 | G | A |
| LDL-C (GLGC) | 11 | 116655600 | G | A |

**Table S14:** Genetic variants used in the presented analyses. Note that due to coverage of the GWAS chip the instruments differed across considered outcomes *(continued)*

| Exposure | Chromosome | Base-pair position (Build 37) | Other allele | Effect allele |
|---|---|---|---|---|
| LDL-C (GLGC) | 11 | 116657561 | T | C |
| LDL-C (GLGC) | 11 | 116660686 | G | A |
| LDL-C (GLGC) | 11 | 116661826 | T | C |
| LDL-C (GLGC) | 11 | 116662579 | T | C |
| LDL-C (GLGC) | 11 | 116663707 | G | A |
| LDL-C (GLGC) | 11 | 116665079 | T | C |
| LDL-C (GLGC) | 11 | 116670182 | T | C |
| LDL-C (GLGC) | 11 | 116670676 | C | A |
| LDL-C (GLGC) | 11 | 116722041 | T | G |
| LDL-C (GLGC) | 11 | 116730638 | G | A |
| LDL-C (GLGC) | 11 | 116732512 | T | G |
| LDL-C (GLGC) | 11 | 116916529 | T | C |
| LDL-C (GLGC) | 11 | 116943354 | C | A |
| LDL-C (GLGC) | 11 | 117075566 | T | C |
| LDL-C (GLGC) | 11 | 122534504 | T | C |
| LDL-C (GLGC) | 11 | 126060666 | G | A |
| LDL-C (GLGC) | 11 | 126084834 | G | A |
| LDL-C (GLGC) | 11 | 126110978 | C | A |
| LDL-C (GLGC) | 11 | 126152647 | G | A |
| LDL-C (GLGC) | 11 | 126180370 | T | G |
| LDL-C (GLGC) | 11 | 126183756 | T | C |
| LDL-C (GLGC) | 11 | 126217870 | T | C |
| LDL-C (GLGC) | 11 | 126218541 | G | A |
| LDL-C (GLGC) | 11 | 126227723 | T | G |
| LDL-C (GLGC) | 11 | 126229617 | T | C |
| LDL-C (GLGC) | 11 | 126232385 | T | C |
| LDL-C (GLGC) | 11 | 126241696 | G | A |
| LDL-C (GLGC) | 11 | 126242912 | G | A |
| LDL-C (GLGC) | 11 | 126243952 | G | A |
| LDL-C (GLGC) | 11 | 126244955 | G | A |
| LDL-C (GLGC) | 11 | 126250680 | G | A |
| LDL-C (GLGC) | 11 | 126257779 | G | A |
| LDL-C (GLGC) | 11 | 126258062 | G | A |
| LDL-C (GLGC) | 11 | 126260580 | G | A |
| LDL-C (GLGC) | 11 | 126270374 | G | A |
| LDL-C (GLGC) | 11 | 18626834 | C | A |
| LDL-C (GLGC) | 11 | 18632984 | T | C |
| LDL-C (GLGC) | 11 | 18656271 | T | C |
| LDL-C (GLGC) | 11 | 61537529 | T | G |
| LDL-C (GLGC) | 11 | 61543499 | T | C |
| LDL-C (GLGC) | 11 | 61548874 | G | A |
| LDL-C (GLGC) | 11 | 61549458 | G | A |
| LDL-C (GLGC) | 11 | 61551927 | C | A |
| LDL-C (GLGC) | 11 | 61564299 | T | G |

**Table S14:** Genetic variants used in the presented analyses. Note that due to coverage of the GWAS chip the instruments differed across considered outcomes *(continued)*

| Exposure | Chromosome | Base-pair position (Build 37) | Other allele | Effect allele |
|---|---|---|---|---|
| LDL-C (GLGC) | 11 | 61565908 | T | C |
| LDL-C (GLGC) | 11 | 61570783 | T | C |
| LDL-C (GLGC) | 11 | 61580635 | T | C |
| LDL-C (GLGC) | 11 | 61582708 | T | C |
| LDL-C (GLGC) | 11 | 61593816 | T | C |
| LDL-C (GLGC) | 11 | 61595564 | T | C |
| LDL-C (GLGC) | 11 | 61597212 | T | C |
| LDL-C (GLGC) | 11 | 61597972 | G | A |
| LDL-C (GLGC) | 11 | 61600327 | G | A |
| LDL-C (GLGC) | 11 | 61601378 | T | C |
| LDL-C (GLGC) | 11 | 61601908 | G | A |
| LDL-C (GLGC) | 11 | 61603237 | T | C |
| LDL-C (GLGC) | 11 | 61605215 | G | A |
| LDL-C (GLGC) | 11 | 61605613 | T | C |
| LDL-C (GLGC) | 11 | 61611694 | G | A |
| LDL-C (GLGC) | 11 | 61616012 | G | A |
| LDL-C (GLGC) | 11 | 61618608 | G | A |
| LDL-C (GLGC) | 11 | 61618831 | T | C |
| LDL-C (GLGC) | 11 | 61621194 | G | A |
| LDL-C (GLGC) | 11 | 61623793 | G | A |
| LDL-C (GLGC) | 11 | 61624181 | T | C |
| LDL-C (GLGC) | 11 | 61624414 | T | C |
| LDL-C (GLGC) | 11 | 61627484 | G | A |
| LDL-C (GLGC) | 11 | 61627881 | T | C |
| LDL-C (GLGC) | 11 | 61629122 | G | A |
| LDL-C (GLGC) | 11 | 61635294 | G | A |
| LDL-C (GLGC) | 11 | 61637057 | G | A |
| LDL-C (GLGC) | 11 | 61638044 | G | A |
| LDL-C (GLGC) | 11 | 61639488 | T | C |
| LDL-C (GLGC) | 11 | 61639573 | G | A |
| LDL-C (GLGC) | 11 | 61650747 | G | A |
| LDL-C (GLGC) | 11 | 61656117 | G | A |
| LDL-C (GLGC) | 11 | 61657110 | T | C |
| LDL-C (GLGC) | 11 | 61657401 | C | A |
| LDL-C (GLGC) | 11 | 61657593 | T | C |
| LDL-C (GLGC) | 11 | 61663691 | G | A |
| LDL-C (GLGC) | 11 | 61677479 | G | A |
| LDL-C (GLGC) | 11 | 61678576 | T | G |
| LDL-C (GLGC) | 11 | 61713590 | G | A |
| LDL-C (GLGC) | 12 | 109095137 | G | A |
| LDL-C (GLGC) | 12 | 111818701 | G | A |
| LDL-C (GLGC) | 12 | 111849271 | G | A |
| LDL-C (GLGC) | 12 | 111893537 | G | A |
| LDL-C (GLGC) | 12 | 111910219 | G | A |

**Table S14:** Genetic variants used in the presented analyses. Note that due to coverage of the GWAS chip the instruments differed across considered outcomes *(continued)*

| Exposure | Chromosome | Base-pair position (Build 37) | Other allele | Effect allele |
|---|---|---|---|---|
| LDL-C (GLGC) | 12 | 111941120 | T | C |
| LDL-C (GLGC) | 12 | 111976776 | T | C |
| LDL-C (GLGC) | 12 | 111993886 | G | A |
| LDL-C (GLGC) | 12 | 112007756 | T | C |
| LDL-C (GLGC) | 12 | 112113658 | T | C |
| LDL-C (GLGC) | 12 | 112123284 | G | A |
| LDL-C (GLGC) | 12 | 112206895 | G | A |
| LDL-C (GLGC) | 12 | 112211833 | T | C |
| LDL-C (GLGC) | 12 | 112338539 | T | C |
| LDL-C (GLGC) | 12 | 112375990 | C | A |
| LDL-C (GLGC) | 12 | 112463296 | G | A |
| LDL-C (GLGC) | 12 | 112521448 | G | A |
| LDL-C (GLGC) | 12 | 112871372 | G | A |
| LDL-C (GLGC) | 12 | 112922529 | G | A |
| LDL-C (GLGC) | 12 | 113024793 | G | A |
| LDL-C (GLGC) | 12 | 121359586 | G | A |
| LDL-C (GLGC) | 12 | 121376416 | T | C |
| LDL-C (GLGC) | 12 | 121380544 | T | C |
| LDL-C (GLGC) | 12 | 121388962 | C | A |
| LDL-C (GLGC) | 12 | 121400205 | T | C |
| LDL-C (GLGC) | 12 | 121403724 | G | A |
| LDL-C (GLGC) | 12 | 121405210 | T | C |
| LDL-C (GLGC) | 12 | 121419056 | T | C |
| LDL-C (GLGC) | 12 | 121420260 | G | A |
| LDL-C (GLGC) | 12 | 121420807 | G | A |
| LDL-C (GLGC) | 12 | 121423386 | G | A |
| LDL-C (GLGC) | 12 | 121432302 | T | G |
| LDL-C (GLGC) | 12 | 121435427 | G | A |
| LDL-C (GLGC) | 12 | 121435475 | G | A |
| LDL-C (GLGC) | 12 | 121436376 | C | A |
| LDL-C (GLGC) | 12 | 121438844 | T | C |
| LDL-C (GLGC) | 12 | 121439192 | T | G |
| LDL-C (GLGC) | 12 | 121439433 | G | A |
| LDL-C (GLGC) | 12 | 121443116 | G | A |
| LDL-C (GLGC) | 12 | 121450165 | T | C |
| LDL-C (GLGC) | 12 | 121454622 | T | C |
| LDL-C (GLGC) | 12 | 121457995 | G | A |
| LDL-C (GLGC) | 12 | 121470256 | G | A |
| LDL-C (GLGC) | 12 | 121471337 | G | A |
| LDL-C (GLGC) | 12 | 121487632 | T | C |
| LDL-C (GLGC) | 13 | 32896846 | G | A |
| LDL-C (GLGC) | 13 | 32936646 | T | C |
| LDL-C (GLGC) | 13 | 32940345 | C | A |
| LDL-C (GLGC) | 13 | 32953388 | T | C |

**Table S14:** Genetic variants used in the presented analyses. Note that due to coverage of the GWAS chip the instruments differed across considered outcomes *(continued)*

| Exposure | Chromosome | Base-pair position (Build 37) | Other allele | Effect allele |
|---|---|---|---|---|
| LDL-C (GLGC) | 14 | 24872209 | T | C |
| LDL-C (GLGC) | 14 | 24883058 | T | C |
| LDL-C (GLGC) | 14 | 24883630 | G | A |
| LDL-C (GLGC) | 14 | 24883887 | G | A |
| LDL-C (GLGC) | 14 | 71096344 | T | G |
| LDL-C (GLGC) | 14 | 94795492 | G | A |
| LDL-C (GLGC) | 16 | 56985139 | G | A |
| LDL-C (GLGC) | 16 | 56985156 | G | A |
| LDL-C (GLGC) | 16 | 56985514 | G | A |
| LDL-C (GLGC) | 16 | 56985555 | G | A |
| LDL-C (GLGC) | 16 | 56987369 | T | C |
| LDL-C (GLGC) | 16 | 56988044 | T | C |
| LDL-C (GLGC) | 16 | 56990716 | C | A |
| LDL-C (GLGC) | 16 | 56993025 | T | G |
| LDL-C (GLGC) | 16 | 56993161 | G | A |
| LDL-C (GLGC) | 16 | 56993211 | T | C |
| LDL-C (GLGC) | 16 | 56993324 | C | A |
| LDL-C (GLGC) | 16 | 56994894 | G | A |
| LDL-C (GLGC) | 16 | 56995236 | C | A |
| LDL-C (GLGC) | 16 | 56996158 | T | C |
| LDL-C (GLGC) | 16 | 56997233 | G | A |
| LDL-C (GLGC) | 16 | 56999258 | C | A |
| LDL-C (GLGC) | 16 | 56999328 | T | C |
| LDL-C (GLGC) | 16 | 57000938 | G | A |
| LDL-C (GLGC) | 16 | 57002732 | T | G |
| LDL-C (GLGC) | 16 | 57004889 | G | A |
| LDL-C (GLGC) | 16 | 57005479 | C | A |
| LDL-C (GLGC) | 16 | 57006590 | T | C |
| LDL-C (GLGC) | 16 | 57007451 | G | A |
| LDL-C (GLGC) | 16 | 57009941 | T | C |
| LDL-C (GLGC) | 16 | 71906597 | T | C |
| LDL-C (GLGC) | 16 | 71951845 | C | A |
| LDL-C (GLGC) | 16 | 71983664 | T | G |
| LDL-C (GLGC) | 16 | 72001110 | G | A |
| LDL-C (GLGC) | 16 | 72007232 | G | A |
| LDL-C (GLGC) | 16 | 72008225 | G | A |
| LDL-C (GLGC) | 16 | 72009676 | T | G |
| LDL-C (GLGC) | 16 | 72011181 | T | G |
| LDL-C (GLGC) | 16 | 72018417 | T | C |
| LDL-C (GLGC) | 16 | 72018771 | C | A |
| LDL-C (GLGC) | 16 | 72022065 | G | A |
| LDL-C (GLGC) | 16 | 72022821 | G | A |
| LDL-C (GLGC) | 16 | 72028930 | T | G |
| LDL-C (GLGC) | 16 | 72031860 | G | A |

**Table S14:** Genetic variants used in the presented analyses. Note that due to coverage of the GWAS chip the instruments differed across considered outcomes *(continued)*

| Exposure | Chromosome | Base-pair position (Build 37) | Other allele | Effect allele |
|---|---|---|---|---|
| LDL-C (GLGC) | 16 | 72035359 | T | C |
| LDL-C (GLGC) | 16 | 72036577 | G | A |
| LDL-C (GLGC) | 16 | 72037206 | C | A |
| LDL-C (GLGC) | 16 | 72042682 | C | A |
| LDL-C (GLGC) | 16 | 72043039 | T | C |
| LDL-C (GLGC) | 16 | 72048080 | T | G |
| LDL-C (GLGC) | 16 | 72048332 | T | C |
| LDL-C (GLGC) | 16 | 72052348 | G | A |
| LDL-C (GLGC) | 16 | 72059269 | T | C |
| LDL-C (GLGC) | 16 | 72067263 | G | A |
| LDL-C (GLGC) | 16 | 72073122 | G | A |
| LDL-C (GLGC) | 16 | 72078485 | T | C |
| LDL-C (GLGC) | 16 | 72108093 | G | A |
| LDL-C (GLGC) | 16 | 72114002 | T | C |
| LDL-C (GLGC) | 16 | 72116467 | G | A |
| LDL-C (GLGC) | 16 | 72137561 | G | A |
| LDL-C (GLGC) | 16 | 72144174 | T | C |
| LDL-C (GLGC) | 16 | 72183959 | G | A |
| LDL-C (GLGC) | 16 | 72193036 | G | A |
| LDL-C (GLGC) | 16 | 72248088 | T | C |
| LDL-C (GLGC) | 16 | 72252752 | G | A |
| LDL-C (GLGC) | 16 | 72256903 | T | C |
| LDL-C (GLGC) | 16 | 72312727 | G | A |
| LDL-C (GLGC) | 16 | 72409698 | T | C |
| LDL-C (GLGC) | 16 | 72469463 | T | C |
| LDL-C (GLGC) | 16 | 72653326 | G | A |
| LDL-C (GLGC) | 16 | 72800567 | G | A |
| LDL-C (GLGC) | 17 | 45312813 | T | C |
| LDL-C (GLGC) | 17 | 45313178 | G | A |
| LDL-C (GLGC) | 17 | 45393665 | T | C |
| LDL-C (GLGC) | 17 | 45393739 | C | A |
| LDL-C (GLGC) | 17 | 45422628 | G | A |
| LDL-C (GLGC) | 17 | 45425115 | G | A |
| LDL-C (GLGC) | 17 | 45498937 | T | G |
| LDL-C (GLGC) | 17 | 45750596 | G | A |
| LDL-C (GLGC) | 17 | 45768836 | T | C |
| LDL-C (GLGC) | 17 | 64210580 | C | A |
| LDL-C (GLGC) | 17 | 67150176 | T | C |
| LDL-C (GLGC) | 17 | 67345501 | T | C |
| LDL-C (GLGC) | 17 | 7088923 | G | A |
| LDL-C (GLGC) | 17 | 7091650 | T | C |
| LDL-C (GLGC) | 17 | 73782191 | T | C |
| LDL-C (GLGC) | 17 | 8129160 | G | A |
| LDL-C (GLGC) | 18 | 47157004 | T | C |

**Table S14:** Genetic variants used in the presented analyses. Note that due to coverage of the GWAS chip the instruments differed across considered outcomes *(continued)*

| Exposure | Chromosome | Base-pair position (Build 37) | Other allele | Effect allele |
|---|---|---|---|---|
| LDL-C (GLGC) | 19 | 10680241 | G | A |
| LDL-C (GLGC) | 19 | 10686123 | G | A |
| LDL-C (GLGC) | 19 | 10728320 | G | A |
| LDL-C (GLGC) | 19 | 10742170 | G | A |
| LDL-C (GLGC) | 19 | 10742287 | T | C |
| LDL-C (GLGC) | 19 | 10793749 | G | A |
| LDL-C (GLGC) | 19 | 10801185 | G | A |
| LDL-C (GLGC) | 19 | 10802792 | G | A |
| LDL-C (GLGC) | 19 | 10813364 | T | C |
| LDL-C (GLGC) | 19 | 10824915 | G | A |
| LDL-C (GLGC) | 19 | 10830281 | T | G |
| LDL-C (GLGC) | 19 | 10894728 | G | A |
| LDL-C (GLGC) | 19 | 10903972 | G | A |
| LDL-C (GLGC) | 19 | 10904700 | T | C |
| LDL-C (GLGC) | 19 | 10909953 | T | C |
| LDL-C (GLGC) | 19 | 10916684 | T | C |
| LDL-C (GLGC) | 19 | 10927540 | T | C |
| LDL-C (GLGC) | 19 | 10947522 | T | C |
| LDL-C (GLGC) | 19 | 10950125 | T | C |
| LDL-C (GLGC) | 19 | 10961273 | G | A |
| LDL-C (GLGC) | 19 | 10981352 | G | A |
| LDL-C (GLGC) | 19 | 10987013 | G | A |
| LDL-C (GLGC) | 19 | 11014937 | T | C |
| LDL-C (GLGC) | 19 | 11019994 | T | C |
| LDL-C (GLGC) | 19 | 11022159 | T | C |
| LDL-C (GLGC) | 19 | 11023434 | T | C |
| LDL-C (GLGC) | 19 | 11042743 | G | A |
| LDL-C (GLGC) | 19 | 11052985 | G | A |
| LDL-C (GLGC) | 19 | 11065021 | G | A |
| LDL-C (GLGC) | 19 | 11065409 | C | A |
| LDL-C (GLGC) | 19 | 11083210 | T | G |
| LDL-C (GLGC) | 19 | 11086746 | T | C |
| LDL-C (GLGC) | 19 | 11092139 | G | A |
| LDL-C (GLGC) | 19 | 11105608 | T | C |
| LDL-C (GLGC) | 19 | 11114352 | G | A |
| LDL-C (GLGC) | 19 | 11128596 | G | A |
| LDL-C (GLGC) | 19 | 11132080 | G | A |
| LDL-C (GLGC) | 19 | 11136315 | T | C |
| LDL-C (GLGC) | 19 | 11139636 | T | G |
| LDL-C (GLGC) | 19 | 11146499 | T | C |
| LDL-C (GLGC) | 19 | 11159525 | T | G |
| LDL-C (GLGC) | 19 | 11163562 | T | C |
| LDL-C (GLGC) | 19 | 11163601 | T | G |
| LDL-C (GLGC) | 19 | 11164781 | G | A |

**Table S14:** Genetic variants used in the presented analyses. Note that due to coverage of the GWAS chip the instruments differed across considered outcomes *(continued)*

| Exposure | Chromosome | Base-pair position (Build 37) | Other allele | Effect allele |
|---|---|---|---|---|
| LDL-C (GLGC) | 19 | 11166827 | G | A |
| LDL-C (GLGC) | 19 | 11169947 | T | C |
| LDL-C (GLGC) | 19 | 11171281 | C | A |
| LDL-C (GLGC) | 19 | 11174625 | G | A |
| LDL-C (GLGC) | 19 | 11180047 | T | C |
| LDL-C (GLGC) | 19 | 11183837 | G | A |
| LDL-C (GLGC) | 19 | 11186711 | T | C |
| LDL-C (GLGC) | 19 | 11188899 | T | C |
| LDL-C (GLGC) | 19 | 11190764 | G | A |
| LDL-C (GLGC) | 19 | 11192226 | T | C |
| LDL-C (GLGC) | 19 | 11195030 | G | A |
| LDL-C (GLGC) | 19 | 11202306 | T | G |
| LDL-C (GLGC) | 19 | 11205975 | G | A |
| LDL-C (GLGC) | 19 | 11206575 | G | A |
| LDL-C (GLGC) | 19 | 11207516 | G | A |
| LDL-C (GLGC) | 19 | 11210912 | T | C |
| LDL-C (GLGC) | 19 | 11224265 | G | A |
| LDL-C (GLGC) | 19 | 11227480 | C | A |
| LDL-C (GLGC) | 19 | 11227554 | T | C |
| LDL-C (GLGC) | 19 | 11230881 | T | C |
| LDL-C (GLGC) | 19 | 11233941 | G | A |
| LDL-C (GLGC) | 19 | 11236804 | T | C |
| LDL-C (GLGC) | 19 | 11238473 | C | A |
| LDL-C (GLGC) | 19 | 11238492 | G | A |
| LDL-C (GLGC) | 19 | 11240053 | G | A |
| LDL-C (GLGC) | 19 | 11241780 | G | A |
| LDL-C (GLGC) | 19 | 11241915 | G | A |
| LDL-C (GLGC) | 19 | 11242658 | T | C |
| LDL-C (GLGC) | 19 | 11243445 | G | A |
| LDL-C (GLGC) | 19 | 11243735 | T | C |
| LDL-C (GLGC) | 19 | 11256285 | G | A |
| LDL-C (GLGC) | 19 | 11257018 | T | C |
| LDL-C (GLGC) | 19 | 11263650 | T | G |
| LDL-C (GLGC) | 19 | 11264514 | T | C |
| LDL-C (GLGC) | 19 | 11266584 | G | A |
| LDL-C (GLGC) | 19 | 11266693 | T | G |
| LDL-C (GLGC) | 19 | 11275139 | C | A |
| LDL-C (GLGC) | 19 | 11284302 | T | C |
| LDL-C (GLGC) | 19 | 11315817 | T | G |
| LDL-C (GLGC) | 19 | 11327608 | T | C |
| LDL-C (GLGC) | 19 | 11380212 | T | C |
| LDL-C (GLGC) | 19 | 11425200 | T | G |
| LDL-C (GLGC) | 19 | 19275735 | T | C |
| LDL-C (GLGC) | 19 | 19295875 | G | A |

**Table S14:** Genetic variants used in the presented analyses. Note that due to coverage of the GWAS chip the instruments differed across considered outcomes *(continued)*

| Exposure | Chromosome | Base-pair position (Build 37) | Other allele | Effect allele |
|---|---|---|---|---|
| LDL-C (GLGC) | 19 | 19325963 | T | G |
| LDL-C (GLGC) | 19 | 19329924 | T | C |
| LDL-C (GLGC) | 19 | 19336608 | T | C |
| LDL-C (GLGC) | 19 | 19374068 | T | C |
| LDL-C (GLGC) | 19 | 19379549 | T | C |
| LDL-C (GLGC) | 19 | 19381715 | G | A |
| LDL-C (GLGC) | 19 | 19392401 | T | C |
| LDL-C (GLGC) | 19 | 19406869 | T | C |
| LDL-C (GLGC) | 19 | 19407718 | T | C |
| LDL-C (GLGC) | 19 | 19453521 | T | G |
| LDL-C (GLGC) | 19 | 19456917 | T | C |
| LDL-C (GLGC) | 19 | 19467545 | T | C |
| LDL-C (GLGC) | 19 | 19478022 | T | G |
| LDL-C (GLGC) | 19 | 19488718 | T | C |
| LDL-C (GLGC) | 19 | 19508013 | G | A |
| LDL-C (GLGC) | 19 | 19525792 | T | C |
| LDL-C (GLGC) | 19 | 19531910 | G | A |
| LDL-C (GLGC) | 19 | 19539891 | T | C |
| LDL-C (GLGC) | 19 | 19588546 | C | A |
| LDL-C (GLGC) | 19 | 19646272 | T | C |
| LDL-C (GLGC) | 19 | 19650096 | G | A |
| LDL-C (GLGC) | 19 | 19657500 | G | A |
| LDL-C (GLGC) | 19 | 19662220 | T | C |
| LDL-C (GLGC) | 19 | 19664077 | T | C |
| LDL-C (GLGC) | 19 | 19665581 | G | A |
| LDL-C (GLGC) | 19 | 19666574 | T | C |
| LDL-C (GLGC) | 19 | 19668916 | G | A |
| LDL-C (GLGC) | 19 | 19692579 | T | C |
| LDL-C (GLGC) | 19 | 19717056 | G | A |
| LDL-C (GLGC) | 19 | 19746151 | T | G |
| LDL-C (GLGC) | 19 | 19789528 | G | A |
| LDL-C (GLGC) | 19 | 19819843 | G | A |
| LDL-C (GLGC) | 19 | 44675801 | T | C |
| LDL-C (GLGC) | 19 | 44830892 | C | A |
| LDL-C (GLGC) | 19 | 44855757 | G | A |
| LDL-C (GLGC) | 19 | 44880214 | T | C |
| LDL-C (GLGC) | 19 | 44880360 | T | C |
| LDL-C (GLGC) | 19 | 44952066 | T | G |
| LDL-C (GLGC) | 19 | 44957508 | G | A |
| LDL-C (GLGC) | 19 | 44988716 | G | A |
| LDL-C (GLGC) | 19 | 45008634 | C | A |
| LDL-C (GLGC) | 19 | 45015020 | T | C |
| LDL-C (GLGC) | 19 | 45017064 | T | C |
| LDL-C (GLGC) | 19 | 45130550 | T | C |

**Table S14:** Genetic variants used in the presented analyses. Note that due to coverage of the GWAS chip the instruments differed across considered outcomes *(continued)*

| Exposure | Chromosome | Base-pair position (Build 37) | Other allele | Effect allele |
|---|---|---|---|---|
| LDL-C (GLGC) | 19 | 45133475 | G | A |
| LDL-C (GLGC) | 19 | 45138246 | T | C |
| LDL-C (GLGC) | 19 | 45146103 | T | C |
| LDL-C (GLGC) | 19 | 45150433 | G | A |
| LDL-C (GLGC) | 19 | 45150614 | G | A |
| LDL-C (GLGC) | 19 | 45163671 | T | C |
| LDL-C (GLGC) | 19 | 45165912 | C | A |
| LDL-C (GLGC) | 19 | 45176340 | T | C |
| LDL-C (GLGC) | 19 | 45189332 | T | C |
| LDL-C (GLGC) | 19 | 45195313 | T | G |
| LDL-C (GLGC) | 19 | 45202027 | G | A |
| LDL-C (GLGC) | 19 | 45215884 | T | C |
| LDL-C (GLGC) | 19 | 45227742 | T | C |
| LDL-C (GLGC) | 19 | 45232205 | G | A |
| LDL-C (GLGC) | 19 | 45232374 | G | A |
| LDL-C (GLGC) | 19 | 45237812 | T | C |
| LDL-C (GLGC) | 19 | 45242107 | G | A |
| LDL-C (GLGC) | 19 | 45242173 | G | A |
| LDL-C (GLGC) | 19 | 45245015 | G | A |
| LDL-C (GLGC) | 19 | 45247627 | G | A |
| LDL-C (GLGC) | 19 | 45254168 | C | A |
| LDL-C (GLGC) | 19 | 45296806 | T | C |
| LDL-C (GLGC) | 19 | 45298069 | G | A |
| LDL-C (GLGC) | 19 | 45313715 | T | C |
| LDL-C (GLGC) | 19 | 45326768 | T | C |
| LDL-C (GLGC) | 19 | 45327459 | T | C |
| LDL-C (GLGC) | 19 | 45328476 | T | C |
| LDL-C (GLGC) | 19 | 45329214 | T | G |
| LDL-C (GLGC) | 19 | 45333834 | T | C |
| LDL-C (GLGC) | 19 | 45336035 | T | C |
| LDL-C (GLGC) | 19 | 45344458 | G | A |
| LDL-C (GLGC) | 19 | 45351746 | G | A |
| LDL-C (GLGC) | 19 | 45351937 | T | C |
| LDL-C (GLGC) | 19 | 45361574 | G | A |
| LDL-C (GLGC) | 19 | 45368320 | T | C |
| LDL-C (GLGC) | 19 | 45371168 | G | A |
| LDL-C (GLGC) | 19 | 45372354 | T | G |
| LDL-C (GLGC) | 19 | 45372959 | T | C |
| LDL-C (GLGC) | 19 | 45373276 | G | A |
| LDL-C (GLGC) | 19 | 45373565 | G | A |
| LDL-C (GLGC) | 19 | 45377467 | G | A |
| LDL-C (GLGC) | 19 | 45379060 | C | A |
| LDL-C (GLGC) | 19 | 45382034 | G | A |
| LDL-C (GLGC) | 19 | 45383061 | T | G |

**Table S14:** Genetic variants used in the presented analyses. Note that due to coverage of the GWAS chip the instruments differed across considered outcomes *(continued)*

| Exposure | Chromosome | Base-pair position (Build 37) | Other allele | Effect allele |
|---|---|---|---|---|
| LDL-C (GLGC) | 19 | 45389596 | G | A |
| LDL-C (GLGC) | 19 | 45392254 | T | C |
| LDL-C (GLGC) | 19 | 45395266 | G | A |
| LDL-C (GLGC) | 19 | 45395619 | G | A |
| LDL-C (GLGC) | 19 | 45399896 | G | A |
| LDL-C (GLGC) | 19 | 45401666 | G | A |
| LDL-C (GLGC) | 19 | 45408836 | T | G |
| LDL-C (GLGC) | 19 | 45410002 | G | A |
| LDL-C (GLGC) | 19 | 45410444 | G | A |
| LDL-C (GLGC) | 19 | 45412079 | T | C |
| LDL-C (GLGC) | 19 | 45413576 | T | C |
| LDL-C (GLGC) | 19 | 45414451 | T | C |
| LDL-C (GLGC) | 19 | 45415640 | G | A |
| LDL-C (GLGC) | 19 | 45416291 | T | C |
| LDL-C (GLGC) | 19 | 45422846 | G | A |
| LDL-C (GLGC) | 19 | 45422946 | G | A |
| LDL-C (GLGC) | 19 | 45423636 | G | A |
| LDL-C (GLGC) | 19 | 45429543 | T | C |
| LDL-C (GLGC) | 19 | 45447221 | G | A |
| LDL-C (GLGC) | 19 | 45449284 | T | G |
| LDL-C (GLGC) | 19 | 45455588 | T | C |
| LDL-C (GLGC) | 19 | 45493133 | T | G |
| LDL-C (GLGC) | 19 | 45496303 | T | C |
| LDL-C (GLGC) | 19 | 45523369 | T | G |
| LDL-C (GLGC) | 19 | 45618902 | T | C |
| LDL-C (GLGC) | 19 | 45619946 | G | A |
| LDL-C (GLGC) | 19 | 45636319 | C | A |
| LDL-C (GLGC) | 19 | 46018119 | G | A |
| LDL-C (GLGC) | 19 | 46180184 | T | C |
| LDL-C (GLGC) | 19 | 46202172 | T | C |
| LDL-C (GLGC) | 19 | 46207810 | C | A |
| LDL-C (GLGC) | 19 | 46295223 | G | A |
| LDL-C (GLGC) | 19 | 49208543 | T | C |
| LDL-C (GLGC) | 19 | 49208978 | G | A |
| LDL-C (GLGC) | 19 | 49214274 | G | A |
| LDL-C (GLGC) | 19 | 49228272 | T | C |
| LDL-C (GLGC) | 19 | 49246866 | G | A |
| LDL-C (GLGC) | 1 | 109703023 | C | A |
| LDL-C (GLGC) | 1 | 109703430 | T | C |
| LDL-C (GLGC) | 1 | 109705022 | G | A |
| LDL-C (GLGC) | 1 | 109709796 | C | A |
| LDL-C (GLGC) | 1 | 109713271 | G | A |
| LDL-C (GLGC) | 1 | 109715950 | G | A |
| LDL-C (GLGC) | 1 | 109723090 | T | C |

**Table S14:** Genetic variants used in the presented analyses. Note that due to coverage of the GWAS chip the instruments differed across considered outcomes *(continued)*

| Exposure | Chromosome | Base-pair position (Build 37) | Other allele | Effect allele |
|---|---|---|---|---|
| LDL-C (GLGC) | 1 | 109723188 | T | C |
| LDL-C (GLGC) | 1 | 109727832 | G | A |
| LDL-C (GLGC) | 1 | 109740364 | T | G |
| LDL-C (GLGC) | 1 | 109747635 | T | C |
| LDL-C (GLGC) | 1 | 109750671 | T | C |
| LDL-C (GLGC) | 1 | 109760555 | T | C |
| LDL-C (GLGC) | 1 | 109768161 | T | C |
| LDL-C (GLGC) | 1 | 109772918 | C | A |
| LDL-C (GLGC) | 1 | 109776285 | T | C |
| LDL-C (GLGC) | 1 | 109778237 | T | C |
| LDL-C (GLGC) | 1 | 109782037 | G | A |
| LDL-C (GLGC) | 1 | 109782190 | T | C |
| LDL-C (GLGC) | 1 | 109783205 | T | G |
| LDL-C (GLGC) | 1 | 109783261 | T | C |
| LDL-C (GLGC) | 1 | 109783648 | T | C |
| LDL-C (GLGC) | 1 | 109784781 | G | A |
| LDL-C (GLGC) | 1 | 109787493 | C | A |
| LDL-C (GLGC) | 1 | 109789795 | T | C |
| LDL-C (GLGC) | 1 | 109795608 | T | C |
| LDL-C (GLGC) | 1 | 109797763 | T | C |
| LDL-C (GLGC) | 1 | 109798721 | G | A |
| LDL-C (GLGC) | 1 | 109800321 | T | C |
| LDL-C (GLGC) | 1 | 109801121 | T | C |
| LDL-C (GLGC) | 1 | 109806442 | C | A |
| LDL-C (GLGC) | 1 | 109806834 | T | C |
| LDL-C (GLGC) | 1 | 109808117 | T | C |
| LDL-C (GLGC) | 1 | 109810200 | G | A |
| LDL-C (GLGC) | 1 | 109810544 | C | A |
| LDL-C (GLGC) | 1 | 109810981 | G | A |
| LDL-C (GLGC) | 1 | 109813020 | G | A |
| LDL-C (GLGC) | 1 | 109813485 | T | C |
| LDL-C (GLGC) | 1 | 109813719 | T | C |
| LDL-C (GLGC) | 1 | 109814880 | T | C |
| LDL-C (GLGC) | 1 | 109815074 | G | A |
| LDL-C (GLGC) | 1 | 109815252 | G | A |
| LDL-C (GLGC) | 1 | 109817192 | G | A |
| LDL-C (GLGC) | 1 | 109817245 | T | C |
| LDL-C (GLGC) | 1 | 109818306 | T | G |
| LDL-C (GLGC) | 1 | 109820110 | T | C |
| LDL-C (GLGC) | 1 | 109820884 | G | A |
| LDL-C (GLGC) | 1 | 109820919 | T | G |
| LDL-C (GLGC) | 1 | 109821307 | T | G |
| LDL-C (GLGC) | 1 | 109822166 | G | A |
| LDL-C (GLGC) | 1 | 109822509 | T | C |

**Table S14:** Genetic variants used in the presented analyses. Note that due to coverage of the GWAS chip the instruments differed across considered outcomes *(continued)*

| Exposure | Chromosome | Base-pair position (Build 37) | Other allele | Effect allele |
|---|---|---|---|---|
| LDL-C (GLGC) | 1 | 109823962 | T | C |
| LDL-C (GLGC) | 1 | 109824250 | G | A |
| LDL-C (GLGC) | 1 | 109826136 | T | C |
| LDL-C (GLGC) | 1 | 109827253 | G | A |
| LDL-C (GLGC) | 1 | 109832283 | G | A |
| LDL-C (GLGC) | 1 | 109834938 | T | C |
| LDL-C (GLGC) | 1 | 109835163 | T | C |
| LDL-C (GLGC) | 1 | 109835387 | T | G |
| LDL-C (GLGC) | 1 | 109837423 | C | A |
| LDL-C (GLGC) | 1 | 109838215 | G | A |
| LDL-C (GLGC) | 1 | 109838918 | T | C |
| LDL-C (GLGC) | 1 | 109839896 | T | C |
| LDL-C (GLGC) | 1 | 109840169 | T | C |
| LDL-C (GLGC) | 1 | 109840629 | G | A |
| LDL-C (GLGC) | 1 | 109845100 | T | G |
| LDL-C (GLGC) | 1 | 109887191 | T | C |
| LDL-C (GLGC) | 1 | 109891133 | T | C |
| LDL-C (GLGC) | 1 | 109909149 | T | C |
| LDL-C (GLGC) | 1 | 109912051 | G | A |
| LDL-C (GLGC) | 1 | 109919525 | G | A |
| LDL-C (GLGC) | 1 | 109922377 | G | A |
| LDL-C (GLGC) | 1 | 109950607 | G | A |
| LDL-C (GLGC) | 1 | 109955569 | T | C |
| LDL-C (GLGC) | 1 | 109967104 | C | A |
| LDL-C (GLGC) | 1 | 109968410 | C | A |
| LDL-C (GLGC) | 1 | 109999838 | C | A |
| LDL-C (GLGC) | 1 | 110009802 | G | A |
| LDL-C (GLGC) | 1 | 110019439 | G | A |
| LDL-C (GLGC) | 1 | 110023610 | G | A |
| LDL-C (GLGC) | 1 | 110040680 | T | C |
| LDL-C (GLGC) | 1 | 110065906 | G | A |
| LDL-C (GLGC) | 1 | 110082473 | T | G |
| LDL-C (GLGC) | 1 | 110109039 | G | A |
| LDL-C (GLGC) | 1 | 110173775 | T | G |
| LDL-C (GLGC) | 1 | 110186563 | T | C |
| LDL-C (GLGC) | 1 | 110201580 | T | C |
| LDL-C (GLGC) | 1 | 110296289 | T | C |
| LDL-C (GLGC) | 1 | 110366083 | T | C |
| LDL-C (GLGC) | 1 | 150958836 | G | A |
| LDL-C (GLGC) | 1 | 220970028 | G | A |
| LDL-C (GLGC) | 1 | 220972343 | G | A |
| LDL-C (GLGC) | 1 | 220973563 | T | C |
| LDL-C (GLGC) | 1 | 220974545 | T | C |
| LDL-C (GLGC) | 1 | 220977650 | C | A |

**Table S14:** Genetic variants used in the presented analyses. Note that due to coverage of the GWAS chip the instruments differed across considered outcomes *(continued)*

| Exposure | Chromosome | Base-pair position (Build 37) | Other allele | Effect allele |
|---|---|---|---|---|
| LDL-C (GLGC) | 1 | 220983402 | T | C |
| LDL-C (GLGC) | 1 | 220988260 | T | C |
| LDL-C (GLGC) | 1 | 221004168 | T | C |
| LDL-C (GLGC) | 1 | 221010205 | T | C |
| LDL-C (GLGC) | 1 | 221010513 | T | G |
| LDL-C (GLGC) | 1 | 221040401 | T | C |
| LDL-C (GLGC) | 1 | 221055463 | G | A |
| LDL-C (GLGC) | 1 | 221057662 | G | A |
| LDL-C (GLGC) | 1 | 221106024 | T | C |
| LDL-C (GLGC) | 1 | 221107232 | G | A |
| LDL-C (GLGC) | 1 | 234840803 | G | A |
| LDL-C (GLGC) | 1 | 234845257 | T | C |
| LDL-C (GLGC) | 1 | 234850374 | T | C |
| LDL-C (GLGC) | 1 | 234854779 | T | C |
| LDL-C (GLGC) | 1 | 234872431 | G | A |
| LDL-C (GLGC) | 1 | 25674785 | T | C |
| LDL-C (GLGC) | 1 | 25684012 | T | C |
| LDL-C (GLGC) | 1 | 25687901 | T | C |
| LDL-C (GLGC) | 1 | 25688276 | G | A |
| LDL-C (GLGC) | 1 | 25758419 | T | C |
| LDL-C (GLGC) | 1 | 25768937 | G | A |
| LDL-C (GLGC) | 1 | 25780893 | G | A |
| LDL-C (GLGC) | 1 | 25802056 | G | A |
| LDL-C (GLGC) | 1 | 25816847 | T | C |
| LDL-C (GLGC) | 1 | 25844714 | G | A |
| LDL-C (GLGC) | 1 | 25851977 | G | A |
| LDL-C (GLGC) | 1 | 25863179 | G | A |
| LDL-C (GLGC) | 1 | 27001893 | G | A |
| LDL-C (GLGC) | 1 | 27138393 | T | C |
| LDL-C (GLGC) | 1 | 27285195 | T | C |
| LDL-C (GLGC) | 1 | 27397604 | T | C |
| LDL-C (GLGC) | 1 | 55485796 | T | C |
| LDL-C (GLGC) | 1 | 55486064 | G | A |
| LDL-C (GLGC) | 1 | 55486117 | G | A |
| LDL-C (GLGC) | 1 | 55487648 | G | A |
| LDL-C (GLGC) | 1 | 55488369 | G | A |
| LDL-C (GLGC) | 1 | 55488989 | G | A |
| LDL-C (GLGC) | 1 | 55496039 | T | C |
| LDL-C (GLGC) | 1 | 55496556 | T | C |
| LDL-C (GLGC) | 1 | 55498745 | T | C |
| LDL-C (GLGC) | 1 | 55499156 | T | C |
| LDL-C (GLGC) | 1 | 55500206 | T | C |
| LDL-C (GLGC) | 1 | 55500429 | G | A |
| LDL-C (GLGC) | 1 | 55502672 | C | A |

**Table S14:** Genetic variants used in the presented analyses. Note that due to coverage of the GWAS chip the instruments differed across considered outcomes *(continued)*

| Exposure | Chromosome | Base-pair position (Build 37) | Other allele | Effect allele |
|---|---|---|---|---|
| LDL-C (GLGC) | 1 | 55503448 | G | A |
| LDL-C (GLGC) | 1 | 55504650 | G | A |
| LDL-C (GLGC) | 1 | 55505647 | T | G |
| LDL-C (GLGC) | 1 | 55505668 | T | C |
| LDL-C (GLGC) | 1 | 55505861 | G | A |
| LDL-C (GLGC) | 1 | 55509872 | T | C |
| LDL-C (GLGC) | 1 | 55513061 | T | C |
| LDL-C (GLGC) | 1 | 55516004 | C | A |
| LDL-C (GLGC) | 1 | 55517344 | T | C |
| LDL-C (GLGC) | 1 | 55518166 | G | A |
| LDL-C (GLGC) | 1 | 55518316 | T | C |
| LDL-C (GLGC) | 1 | 55518752 | T | C |
| LDL-C (GLGC) | 1 | 55520864 | G | A |
| LDL-C (GLGC) | 1 | 55524237 | G | A |
| LDL-C (GLGC) | 1 | 55524842 | G | A |
| LDL-C (GLGC) | 1 | 55525726 | T | G |
| LDL-C (GLGC) | 1 | 55526296 | G | A |
| LDL-C (GLGC) | 1 | 55527323 | G | A |
| LDL-C (GLGC) | 1 | 55528629 | G | A |
| LDL-C (GLGC) | 1 | 55529187 | G | A |
| LDL-C (GLGC) | 1 | 55538552 | T | G |
| LDL-C (GLGC) | 1 | 55551718 | G | A |
| LDL-C (GLGC) | 1 | 55590465 | T | C |
| LDL-C (GLGC) | 1 | 55602644 | C | A |
| LDL-C (GLGC) | 1 | 55652983 | T | G |
| LDL-C (GLGC) | 1 | 55656075 | T | G |
| LDL-C (GLGC) | 1 | 55695535 | T | C |
| LDL-C (GLGC) | 1 | 55711465 | G | A |
| LDL-C (GLGC) | 1 | 55713094 | G | A |
| LDL-C (GLGC) | 1 | 55713628 | G | A |
| LDL-C (GLGC) | 1 | 55713961 | T | C |
| LDL-C (GLGC) | 1 | 55715412 | T | C |
| LDL-C (GLGC) | 1 | 55716497 | G | A |
| LDL-C (GLGC) | 1 | 55719166 | T | G |
| LDL-C (GLGC) | 1 | 55724704 | G | A |
| LDL-C (GLGC) | 1 | 55730327 | T | C |
| LDL-C (GLGC) | 1 | 55738663 | T | C |
| LDL-C (GLGC) | 1 | 55739925 | T | C |
| LDL-C (GLGC) | 1 | 55741377 | G | A |
| LDL-C (GLGC) | 1 | 55742322 | T | C |
| LDL-C (GLGC) | 1 | 55767547 | T | C |
| LDL-C (GLGC) | 1 | 55770565 | T | C |
| LDL-C (GLGC) | 1 | 55785672 | T | C |
| LDL-C (GLGC) | 1 | 55806465 | T | C |

**Table S14:** Genetic variants used in the presented analyses. Note that due to coverage of the GWAS chip the instruments differed across considered outcomes *(continued)*

| Exposure | Chromosome | Base-pair position (Build 37) | Other allele | Effect allele |
|---|---|---|---|---|
| LDL-C (GLGC) | 1 | 55808143 | T | C |
| LDL-C (GLGC) | 1 | 55814164 | T | C |
| LDL-C (GLGC) | 1 | 55815584 | G | A |
| LDL-C (GLGC) | 1 | 55820713 | C | A |
| LDL-C (GLGC) | 1 | 56240278 | T | C |
| LDL-C (GLGC) | 1 | 62902874 | G | A |
| LDL-C (GLGC) | 1 | 62917913 | G | A |
| LDL-C (GLGC) | 1 | 62931632 | C | A |
| LDL-C (GLGC) | 1 | 62960250 | T | C |
| LDL-C (GLGC) | 1 | 62991905 | T | C |
| LDL-C (GLGC) | 1 | 62995263 | T | C |
| LDL-C (GLGC) | 1 | 62996373 | T | C |
| LDL-C (GLGC) | 1 | 63049551 | C | A |
| LDL-C (GLGC) | 1 | 63083289 | C | A |
| LDL-C (GLGC) | 1 | 63135127 | G | A |
| LDL-C (GLGC) | 1 | 63171063 | T | C |
| LDL-C (GLGC) | 1 | 63191777 | T | C |
| LDL-C (GLGC) | 1 | 63193972 | G | A |
| LDL-C (GLGC) | 1 | 63194009 | T | C |
| LDL-C (GLGC) | 1 | 63195692 | T | C |
| LDL-C (GLGC) | 1 | 63199348 | T | G |
| LDL-C (GLGC) | 1 | 63199936 | T | G |
| LDL-C (GLGC) | 1 | 63203221 | T | C |
| LDL-C (GLGC) | 1 | 63211706 | T | C |
| LDL-C (GLGC) | 1 | 63252766 | T | C |
| LDL-C (GLGC) | 1 | 63310444 | G | A |
| LDL-C (GLGC) | 1 | 63358816 | T | C |
| LDL-C (GLGC) | 1 | 92680371 | G | A |
| LDL-C (GLGC) | 1 | 92761654 | T | C |
| LDL-C (GLGC) | 1 | 92846681 | G | A |
| LDL-C (GLGC) | 1 | 92904381 | T | C |
| LDL-C (GLGC) | 1 | 92979354 | G | A |
| LDL-C (GLGC) | 1 | 92987575 | G | A |
| LDL-C (GLGC) | 1 | 93009438 | C | A |
| LDL-C (GLGC) | 1 | 93108402 | G | A |
| LDL-C (GLGC) | 1 | 93160902 | T | C |
| LDL-C (GLGC) | 1 | 93167854 | G | A |
| LDL-C (GLGC) | 1 | 93269824 | T | C |
| LDL-C (GLGC) | 1 | 93348423 | T | C |
| LDL-C (GLGC) | 20 | 12969400 | G | A |
| LDL-C (GLGC) | 20 | 17845921 | C | A |
| LDL-C (GLGC) | 20 | 17847735 | T | C |
| LDL-C (GLGC) | 20 | 17850340 | G | A |
| LDL-C (GLGC) | 20 | 34046501 | G | A |

**Table S14:** Genetic variants used in the presented analyses. Note that due to coverage of the GWAS chip the instruments differed across considered outcomes *(continued)*

| Exposure | Chromosome | Base-pair position (Build 37) | Other allele | Effect allele |
|---|---|---|---|---|
| LDL-C (GLGC) | 20 | 34154371 | C | A |
| LDL-C (GLGC) | 20 | 39086995 | T | G |
| LDL-C (GLGC) | 20 | 39091514 | G | A |
| LDL-C (GLGC) | 20 | 39091892 | G | A |
| LDL-C (GLGC) | 20 | 39092895 | G | A |
| LDL-C (GLGC) | 20 | 39147287 | G | A |
| LDL-C (GLGC) | 20 | 39154095 | G | A |
| LDL-C (GLGC) | 20 | 39159119 | T | C |
| LDL-C (GLGC) | 20 | 39163431 | T | C |
| LDL-C (GLGC) | 20 | 39167592 | G | A |
| LDL-C (GLGC) | 20 | 39193406 | T | C |
| LDL-C (GLGC) | 20 | 39200914 | G | A |
| LDL-C (GLGC) | 20 | 39216286 | T | G |
| LDL-C (GLGC) | 20 | 39228784 | T | C |
| LDL-C (GLGC) | 20 | 39234223 | G | A |
| LDL-C (GLGC) | 20 | 39237558 | G | A |
| LDL-C (GLGC) | 20 | 39275470 | G | A |
| LDL-C (GLGC) | 20 | 39282720 | C | A |
| LDL-C (GLGC) | 20 | 39291784 | G | A |
| LDL-C (GLGC) | 20 | 39293397 | T | C |
| LDL-C (GLGC) | 20 | 39308345 | T | C |
| LDL-C (GLGC) | 20 | 39617398 | G | A |
| LDL-C (GLGC) | 20 | 39636663 | T | C |
| LDL-C (GLGC) | 20 | 39662842 | G | A |
| LDL-C (GLGC) | 20 | 39678289 | T | C |
| LDL-C (GLGC) | 20 | 39690342 | T | C |
| LDL-C (GLGC) | 20 | 39709255 | T | C |
| LDL-C (GLGC) | 20 | 39710496 | G | A |
| LDL-C (GLGC) | 20 | 39720830 | G | A |
| LDL-C (GLGC) | 20 | 39721373 | G | A |
| LDL-C (GLGC) | 20 | 39769975 | T | C |
| LDL-C (GLGC) | 20 | 39780932 | G | A |
| LDL-C (GLGC) | 20 | 39794489 | G | A |
| LDL-C (GLGC) | 20 | 39797465 | T | C |
| LDL-C (GLGC) | 20 | 39821898 | T | C |
| LDL-C (GLGC) | 20 | 39870578 | C | A |
| LDL-C (GLGC) | 20 | 39882406 | T | G |
| LDL-C (GLGC) | 20 | 39897007 | T | C |
| LDL-C (GLGC) | 20 | 39907749 | C | A |
| LDL-C (GLGC) | 20 | 39915515 | G | A |
| LDL-C (GLGC) | 20 | 39916437 | T | C |
| LDL-C (GLGC) | 20 | 39938122 | T | C |
| LDL-C (GLGC) | 20 | 39960342 | G | A |
| LDL-C (GLGC) | 20 | 39964330 | T | C |

**Table S14:** Genetic variants used in the presented analyses. Note that due to coverage of the GWAS chip the instruments differed across considered outcomes *(continued)*

| Exposure | Chromosome | Base-pair position (Build 37) | Other allele | Effect allele |
|---|---|---|---|---|
| LDL-C (GLGC) | 20 | 39969513 | C | A |
| LDL-C (GLGC) | 20 | 39974316 | G | A |
| LDL-C (GLGC) | 20 | 40021751 | C | A |
| LDL-C (GLGC) | 20 | 40054210 | G | A |
| LDL-C (GLGC) | 20 | 40066770 | G | A |
| LDL-C (GLGC) | 20 | 40078085 | T | C |
| LDL-C (GLGC) | 20 | 40237810 | T | G |
| LDL-C (GLGC) | 20 | 40267858 | G | A |
| LDL-C (GLGC) | 20 | 43042364 | T | C |
| LDL-C (GLGC) | 22 | 30198025 | T | C |
| LDL-C (GLGC) | 22 | 30241061 | G | A |
| LDL-C (GLGC) | 22 | 30378703 | T | C |
| LDL-C (GLGC) | 22 | 30413877 | T | C |
| LDL-C (GLGC) | 22 | 46627603 | T | C |
| LDL-C (GLGC) | 2 | 118680551 | T | G |
| LDL-C (GLGC) | 2 | 118690820 | T | C |
| LDL-C (GLGC) | 2 | 118706990 | G | A |
| LDL-C (GLGC) | 2 | 118815958 | G | A |
| LDL-C (GLGC) | 2 | 118849850 | G | A |
| LDL-C (GLGC) | 2 | 118879253 | T | C |
| LDL-C (GLGC) | 2 | 121309488 | T | C |
| LDL-C (GLGC) | 2 | 121322038 | T | C |
| LDL-C (GLGC) | 2 | 135755629 | G | A |
| LDL-C (GLGC) | 2 | 135777504 | G | A |
| LDL-C (GLGC) | 2 | 135837906 | C | A |
| LDL-C (GLGC) | 2 | 135893372 | G | A |
| LDL-C (GLGC) | 2 | 135952405 | T | C |
| LDL-C (GLGC) | 2 | 136092061 | C | A |
| LDL-C (GLGC) | 2 | 136393658 | G | A |
| LDL-C (GLGC) | 2 | 136459227 | G | A |
| LDL-C (GLGC) | 2 | 136608646 | G | A |
| LDL-C (GLGC) | 2 | 136714178 | G | A |
| LDL-C (GLGC) | 2 | 136787402 | T | C |
| LDL-C (GLGC) | 2 | 136808949 | T | C |
| LDL-C (GLGC) | 2 | 165528876 | T | C |
| LDL-C (GLGC) | 2 | 165539661 | T | C |
| LDL-C (GLGC) | 2 | 169827018 | T | C |
| LDL-C (GLGC) | 2 | 169830051 | T | C |
| LDL-C (GLGC) | 2 | 169830155 | G | A |
| LDL-C (GLGC) | 2 | 169830328 | G | A |
| LDL-C (GLGC) | 2 | 203708307 | G | A |
| LDL-C (GLGC) | 2 | 20887226 | T | G |
| LDL-C (GLGC) | 2 | 20985414 | T | C |
| LDL-C (GLGC) | 2 | 21004251 | T | C |

**Table S14:** Genetic variants used in the presented analyses. Note that due to coverage of the GWAS chip the instruments differed across considered outcomes *(continued)*

| Exposure | Chromosome | Base-pair position (Build 37) | Other allele | Effect allele |
|---|---|---|---|---|
| LDL-C (GLGC) | 2 | 21097962 | T | C |
| LDL-C (GLGC) | 2 | 21100438 | T | G |
| LDL-C (GLGC) | 2 | 21111763 | G | A |
| LDL-C (GLGC) | 2 | 21112689 | G | A |
| LDL-C (GLGC) | 2 | 21118983 | C | A |
| LDL-C (GLGC) | 2 | 21121130 | T | G |
| LDL-C (GLGC) | 2 | 21121464 | T | C |
| LDL-C (GLGC) | 2 | 21123352 | C | A |
| LDL-C (GLGC) | 2 | 21125742 | T | C |
| LDL-C (GLGC) | 2 | 21133883 | G | A |
| LDL-C (GLGC) | 2 | 21134064 | T | C |
| LDL-C (GLGC) | 2 | 21135577 | G | A |
| LDL-C (GLGC) | 2 | 21135628 | T | C |
| LDL-C (GLGC) | 2 | 21137986 | G | A |
| LDL-C (GLGC) | 2 | 21139625 | T | G |
| LDL-C (GLGC) | 2 | 21140540 | T | C |
| LDL-C (GLGC) | 2 | 21141725 | G | A |
| LDL-C (GLGC) | 2 | 21141832 | C | A |
| LDL-C (GLGC) | 2 | 21142994 | T | C |
| LDL-C (GLGC) | 2 | 21143329 | G | A |
| LDL-C (GLGC) | 2 | 21145499 | G | A |
| LDL-C (GLGC) | 2 | 21147179 | T | C |
| LDL-C (GLGC) | 2 | 21148274 | C | A |
| LDL-C (GLGC) | 2 | 21187599 | G | A |
| LDL-C (GLGC) | 2 | 21189091 | T | C |
| LDL-C (GLGC) | 2 | 21190024 | T | C |
| LDL-C (GLGC) | 2 | 21190209 | T | C |
| LDL-C (GLGC) | 2 | 21192112 | C | A |
| LDL-C (GLGC) | 2 | 21194868 | T | C |
| LDL-C (GLGC) | 2 | 21196112 | G | A |
| LDL-C (GLGC) | 2 | 21196346 | T | G |
| LDL-C (GLGC) | 2 | 21196684 | T | G |
| LDL-C (GLGC) | 2 | 21197159 | T | C |
| LDL-C (GLGC) | 2 | 21198426 | T | C |
| LDL-C (GLGC) | 2 | 21205457 | G | A |
| LDL-C (GLGC) | 2 | 21206052 | G | A |
| LDL-C (GLGC) | 2 | 21206906 | T | C |
| LDL-C (GLGC) | 2 | 21208211 | G | A |
| LDL-C (GLGC) | 2 | 21213193 | T | C |
| LDL-C (GLGC) | 2 | 21214832 | G | A |
| LDL-C (GLGC) | 2 | 21215645 | T | C |
| LDL-C (GLGC) | 2 | 21216815 | C | A |
| LDL-C (GLGC) | 2 | 21217230 | G | A |
| LDL-C (GLGC) | 2 | 21217490 | T | C |

**Table S14:** Genetic variants used in the presented analyses. Note that due to coverage of the GWAS chip the instruments differed across considered outcomes *(continued)*

| Exposure | Chromosome | Base-pair position (Build 37) | Other allele | Effect allele |
|---|---|---|---|---|
| LDL-C (GLGC) | 2 | 21218805 | T | C |
| LDL-C (GLGC) | 2 | 21221133 | G | A |
| LDL-C (GLGC) | 2 | 21225753 | T | C |
| LDL-C (GLGC) | 2 | 21228827 | T | C |
| LDL-C (GLGC) | 2 | 21231592 | G | A |
| LDL-C (GLGC) | 2 | 21232195 | G | A |
| LDL-C (GLGC) | 2 | 21233972 | T | C |
| LDL-C (GLGC) | 2 | 21237544 | G | A |
| LDL-C (GLGC) | 2 | 21237786 | T | G |
| LDL-C (GLGC) | 2 | 21239918 | T | G |
| LDL-C (GLGC) | 2 | 21240364 | G | A |
| LDL-C (GLGC) | 2 | 21241505 | G | A |
| LDL-C (GLGC) | 2 | 21244000 | T | G |
| LDL-C (GLGC) | 2 | 21247211 | T | C |
| LDL-C (GLGC) | 2 | 21249716 | T | C |
| LDL-C (GLGC) | 2 | 21250914 | G | A |
| LDL-C (GLGC) | 2 | 21257927 | T | C |
| LDL-C (GLGC) | 2 | 21259562 | T | C |
| LDL-C (GLGC) | 2 | 21259612 | G | A |
| LDL-C (GLGC) | 2 | 21260601 | G | A |
| LDL-C (GLGC) | 2 | 21261507 | T | C |
| LDL-C (GLGC) | 2 | 21261998 | C | A |
| LDL-C (GLGC) | 2 | 21262427 | G | A |
| LDL-C (GLGC) | 2 | 21263639 | T | G |
| LDL-C (GLGC) | 2 | 21263900 | G | A |
| LDL-C (GLGC) | 2 | 21267247 | T | G |
| LDL-C (GLGC) | 2 | 21267461 | G | A |
| LDL-C (GLGC) | 2 | 21269809 | G | A |
| LDL-C (GLGC) | 2 | 21270057 | G | A |
| LDL-C (GLGC) | 2 | 21270751 | G | A |
| LDL-C (GLGC) | 2 | 21271270 | C | A |
| LDL-C (GLGC) | 2 | 21271323 | G | A |
| LDL-C (GLGC) | 2 | 21276962 | G | A |
| LDL-C (GLGC) | 2 | 21288321 | G | A |
| LDL-C (GLGC) | 2 | 21290067 | T | C |
| LDL-C (GLGC) | 2 | 21293135 | T | C |
| LDL-C (GLGC) | 2 | 21295227 | G | A |
| LDL-C (GLGC) | 2 | 21297149 | G | A |
| LDL-C (GLGC) | 2 | 21300770 | C | A |
| LDL-C (GLGC) | 2 | 21301892 | T | C |
| LDL-C (GLGC) | 2 | 21303470 | T | C |
| LDL-C (GLGC) | 2 | 21309335 | T | C |
| LDL-C (GLGC) | 2 | 21309856 | G | A |
| LDL-C (GLGC) | 2 | 21311541 | T | G |

**Table S14:** Genetic variants used in the presented analyses. Note that due to coverage of the GWAS chip the instruments differed across considered outcomes *(continued)*

| Exposure | Chromosome | Base-pair position (Build 37) | Other allele | Effect allele |
|---|---|---|---|---|
| LDL-C (GLGC) | 2 | 21311691 | G | A |
| LDL-C (GLGC) | 2 | 21312210 | T | C |
| LDL-C (GLGC) | 2 | 21315664 | T | C |
| LDL-C (GLGC) | 2 | 21318633 | T | C |
| LDL-C (GLGC) | 2 | 21319066 | T | G |
| LDL-C (GLGC) | 2 | 21327635 | G | A |
| LDL-C (GLGC) | 2 | 21328825 | G | A |
| LDL-C (GLGC) | 2 | 21337306 | T | C |
| LDL-C (GLGC) | 2 | 21348574 | G | A |
| LDL-C (GLGC) | 2 | 21349199 | T | C |
| LDL-C (GLGC) | 2 | 21364562 | T | C |
| LDL-C (GLGC) | 2 | 21366682 | T | C |
| LDL-C (GLGC) | 2 | 21368217 | G | A |
| LDL-C (GLGC) | 2 | 21377935 | G | A |
| LDL-C (GLGC) | 2 | 21377938 | G | A |
| LDL-C (GLGC) | 2 | 21378453 | G | A |
| LDL-C (GLGC) | 2 | 21381177 | G | A |
| LDL-C (GLGC) | 2 | 21385541 | G | A |
| LDL-C (GLGC) | 2 | 21386078 | C | A |
| LDL-C (GLGC) | 2 | 21389019 | G | A |
| LDL-C (GLGC) | 2 | 21389659 | T | G |
| LDL-C (GLGC) | 2 | 21396001 | C | A |
| LDL-C (GLGC) | 2 | 21399216 | T | G |
| LDL-C (GLGC) | 2 | 21400661 | G | A |
| LDL-C (GLGC) | 2 | 21402981 | T | C |
| LDL-C (GLGC) | 2 | 21403965 | G | A |
| LDL-C (GLGC) | 2 | 21415466 | G | A |
| LDL-C (GLGC) | 2 | 21428049 | C | A |
| LDL-C (GLGC) | 2 | 21430401 | T | C |
| LDL-C (GLGC) | 2 | 21438738 | G | A |
| LDL-C (GLGC) | 2 | 21439329 | T | G |
| LDL-C (GLGC) | 2 | 21445569 | G | A |
| LDL-C (GLGC) | 2 | 21448511 | G | A |
| LDL-C (GLGC) | 2 | 21448887 | T | C |
| LDL-C (GLGC) | 2 | 21449408 | T | C |
| LDL-C (GLGC) | 2 | 21449987 | G | A |
| LDL-C (GLGC) | 2 | 21450714 | G | A |
| LDL-C (GLGC) | 2 | 21451604 | T | C |
| LDL-C (GLGC) | 2 | 21451687 | G | A |
| LDL-C (GLGC) | 2 | 21451827 | G | A |
| LDL-C (GLGC) | 2 | 21453211 | G | A |
| LDL-C (GLGC) | 2 | 21453733 | G | A |
| LDL-C (GLGC) | 2 | 21460176 | T | C |
| LDL-C (GLGC) | 2 | 21460827 | T | C |

**Table S14:** Genetic variants used in the presented analyses. Note that due to coverage of the GWAS chip the instruments differed across considered outcomes *(continued)*

| Exposure | Chromosome | Base-pair position (Build 37) | Other allele | Effect allele |
|---|---|---|---|---|
| LDL-C (GLGC) | 2 | 21463790 | G | A |
| LDL-C (GLGC) | 2 | 21464498 | G | A |
| LDL-C (GLGC) | 2 | 21469152 | T | G |
| LDL-C (GLGC) | 2 | 21470987 | T | C |
| LDL-C (GLGC) | 2 | 21476634 | G | A |
| LDL-C (GLGC) | 2 | 21493269 | T | C |
| LDL-C (GLGC) | 2 | 21502727 | G | A |
| LDL-C (GLGC) | 2 | 21507500 | G | A |
| LDL-C (GLGC) | 2 | 21514481 | C | A |
| LDL-C (GLGC) | 2 | 21527036 | G | A |
| LDL-C (GLGC) | 2 | 21535673 | G | A |
| LDL-C (GLGC) | 2 | 21538345 | G | A |
| LDL-C (GLGC) | 2 | 21538810 | C | A |
| LDL-C (GLGC) | 2 | 21538826 | T | C |
| LDL-C (GLGC) | 2 | 216287093 | G | A |
| LDL-C (GLGC) | 2 | 216304384 | T | C |
| LDL-C (GLGC) | 2 | 21769639 | G | A |
| LDL-C (GLGC) | 2 | 234605835 | T | C |
| LDL-C (GLGC) | 2 | 234679384 | T | C |
| LDL-C (GLGC) | 2 | 27730940 | T | C |
| LDL-C (GLGC) | 2 | 27731212 | T | C |
| LDL-C (GLGC) | 2 | 43990829 | G | A |
| LDL-C (GLGC) | 2 | 43998726 | G | A |
| LDL-C (GLGC) | 2 | 44004099 | T | G |
| LDL-C (GLGC) | 2 | 44022970 | T | C |
| LDL-C (GLGC) | 2 | 44036689 | G | A |
| LDL-C (GLGC) | 2 | 44037488 | G | A |
| LDL-C (GLGC) | 2 | 44042317 | G | A |
| LDL-C (GLGC) | 2 | 44043393 | G | A |
| LDL-C (GLGC) | 2 | 44046585 | T | C |
| LDL-C (GLGC) | 2 | 44048346 | T | C |
| LDL-C (GLGC) | 2 | 44057102 | G | A |
| LDL-C (GLGC) | 2 | 44057570 | C | A |
| LDL-C (GLGC) | 2 | 44058119 | G | A |
| LDL-C (GLGC) | 2 | 44065090 | G | A |
| LDL-C (GLGC) | 2 | 44072576 | T | G |
| LDL-C (GLGC) | 2 | 44073656 | T | C |
| LDL-C (GLGC) | 2 | 44079004 | T | C |
| LDL-C (GLGC) | 2 | 44079310 | C | A |
| LDL-C (GLGC) | 2 | 44080324 | G | A |
| LDL-C (GLGC) | 2 | 44081042 | T | C |
| LDL-C (GLGC) | 2 | 44085035 | C | A |
| LDL-C (GLGC) | 2 | 44092066 | T | C |
| LDL-C (GLGC) | 2 | 44094845 | G | A |

**Table S14:** Genetic variants used in the presented analyses. Note that due to coverage of the GWAS chip the instruments differed across considered outcomes *(continued)*

| Exposure | Chromosome | Base-pair position (Build 37) | Other allele | Effect allele |
|---|---|---|---|---|
| LDL-C (GLGC) | 2 | 44096402 | G | A |
| LDL-C (GLGC) | 2 | 44097114 | G | A |
| LDL-C (GLGC) | 2 | 44099433 | C | A |
| LDL-C (GLGC) | 2 | 44260914 | T | C |
| LDL-C (GLGC) | 2 | 62871225 | T | C |
| LDL-C (GLGC) | 2 | 62884053 | G | A |
| LDL-C (GLGC) | 2 | 63069112 | T | G |
| LDL-C (GLGC) | 2 | 63149557 | G | A |
| LDL-C (GLGC) | 2 | 63259881 | T | C |
| LDL-C (GLGC) | 2 | 63280690 | T | C |
| LDL-C (GLGC) | 2 | 63362468 | G | A |
| LDL-C (GLGC) | 2 | 63562115 | G | A |
| LDL-C (GLGC) | 3 | 12267648 | G | A |
| LDL-C (GLGC) | 3 | 12272199 | G | A |
| LDL-C (GLGC) | 3 | 12288569 | C | A |
| LDL-C (GLGC) | 3 | 12296469 | G | A |
| LDL-C (GLGC) | 3 | 12323413 | G | A |
| LDL-C (GLGC) | 3 | 12327431 | T | G |
| LDL-C (GLGC) | 3 | 12344441 | G | A |
| LDL-C (GLGC) | 3 | 12388563 | C | A |
| LDL-C (GLGC) | 3 | 12397392 | G | A |
| LDL-C (GLGC) | 3 | 12402601 | G | A |
| LDL-C (GLGC) | 3 | 12403119 | T | G |
| LDL-C (GLGC) | 3 | 12430075 | T | C |
| LDL-C (GLGC) | 3 | 12449528 | T | G |
| LDL-C (GLGC) | 3 | 12461615 | G | A |
| LDL-C (GLGC) | 3 | 12466490 | G | A |
| LDL-C (GLGC) | 3 | 12467044 | T | C |
| LDL-C (GLGC) | 3 | 12470239 | C | A |
| LDL-C (GLGC) | 3 | 12489342 | T | G |
| LDL-C (GLGC) | 3 | 12502955 | T | G |
| LDL-C (GLGC) | 3 | 12674199 | T | C |
| LDL-C (GLGC) | 3 | 12709167 | G | A |
| LDL-C (GLGC) | 3 | 132163200 | T | G |
| LDL-C (GLGC) | 3 | 132257961 | T | G |
| LDL-C (GLGC) | 3 | 32533010 | T | C |
| LDL-C (GLGC) | 3 | 32538247 | T | C |
| LDL-C (GLGC) | 3 | 58293867 | G | A |
| LDL-C (GLGC) | 3 | 58381287 | G | A |
| LDL-C (GLGC) | 4 | 151303318 | G | A |
| LDL-C (GLGC) | 4 | 3434885 | T | G |
| LDL-C (GLGC) | 4 | 3473139 | G | A |
| LDL-C (GLGC) | 5 | 122855416 | G | A |
| LDL-C (GLGC) | 5 | 156111450 | T | C |

**Table S14:** Genetic variants used in the presented analyses. Note that due to coverage of the GWAS chip the instruments differed across considered outcomes *(continued)*

| Exposure | Chromosome | Base-pair position (Build 37) | Other allele | Effect allele |
|---|---|---|---|---|
| LDL-C (GLGC) | 5 | 156273481 | G | A |
| LDL-C (GLGC) | 5 | 156354477 | G | A |
| LDL-C (GLGC) | 5 | 156369171 | T | C |
| LDL-C (GLGC) | 5 | 156376703 | G | A |
| LDL-C (GLGC) | 5 | 156381370 | G | A |
| LDL-C (GLGC) | 5 | 156382308 | G | A |
| LDL-C (GLGC) | 5 | 156390297 | T | C |
| LDL-C (GLGC) | 5 | 156394441 | G | A |
| LDL-C (GLGC) | 5 | 156396003 | T | C |
| LDL-C (GLGC) | 5 | 156398253 | G | A |
| LDL-C (GLGC) | 5 | 156405946 | T | C |
| LDL-C (GLGC) | 5 | 156432215 | G | A |
| LDL-C (GLGC) | 5 | 156442657 | G | A |
| LDL-C (GLGC) | 5 | 156449748 | G | A |
| LDL-C (GLGC) | 5 | 156466179 | T | G |
| LDL-C (GLGC) | 5 | 156468539 | G | A |
| LDL-C (GLGC) | 5 | 156469783 | G | A |
| LDL-C (GLGC) | 5 | 156472930 | G | A |
| LDL-C (GLGC) | 5 | 156475398 | T | G |
| LDL-C (GLGC) | 5 | 156479509 | G | A |
| LDL-C (GLGC) | 5 | 74259550 | G | A |
| LDL-C (GLGC) | 5 | 74263455 | T | C |
| LDL-C (GLGC) | 5 | 74264281 | T | C |
| LDL-C (GLGC) | 5 | 74266343 | G | A |
| LDL-C (GLGC) | 5 | 74276566 | C | A |
| LDL-C (GLGC) | 5 | 74280980 | G | A |
| LDL-C (GLGC) | 5 | 74291869 | G | A |
| LDL-C (GLGC) | 5 | 74300613 | T | G |
| LDL-C (GLGC) | 5 | 74302658 | G | A |
| LDL-C (GLGC) | 5 | 74340327 | C | A |
| LDL-C (GLGC) | 5 | 74343769 | G | A |
| LDL-C (GLGC) | 5 | 74345369 | C | A |
| LDL-C (GLGC) | 5 | 74352180 | G | A |
| LDL-C (GLGC) | 5 | 74361140 | T | C |
| LDL-C (GLGC) | 5 | 74365757 | C | A |
| LDL-C (GLGC) | 5 | 74382081 | G | A |
| LDL-C (GLGC) | 5 | 74382382 | T | G |
| LDL-C (GLGC) | 5 | 74384115 | G | A |
| LDL-C (GLGC) | 5 | 74442964 | T | C |
| LDL-C (GLGC) | 5 | 74443132 | T | C |
| LDL-C (GLGC) | 5 | 74461924 | C | A |
| LDL-C (GLGC) | 5 | 74466833 | G | A |
| LDL-C (GLGC) | 5 | 74490547 | T | C |
| LDL-C (GLGC) | 5 | 74497885 | T | C |

**Table S14:** Genetic variants used in the presented analyses. Note that due to coverage of the GWAS chip the instruments differed across considered outcomes *(continued)*

| Exposure | Chromosome | Base-pair position (Build 37) | Other allele | Effect allele |
|---|---|---|---|---|
| LDL-C (GLGC) | 5 | 74534372 | G | A |
| LDL-C (GLGC) | 5 | 74562564 | T | C |
| LDL-C (GLGC) | 5 | 74570531 | G | A |
| LDL-C (GLGC) | 5 | 74574984 | G | A |
| LDL-C (GLGC) | 5 | 74582162 | G | A |
| LDL-C (GLGC) | 5 | 74596682 | T | G |
| LDL-C (GLGC) | 5 | 74602699 | C | A |
| LDL-C (GLGC) | 5 | 74604742 | G | A |
| LDL-C (GLGC) | 5 | 74605220 | G | A |
| LDL-C (GLGC) | 5 | 74611574 | T | G |
| LDL-C (GLGC) | 5 | 74616843 | T | C |
| LDL-C (GLGC) | 5 | 74620225 | T | C |
| LDL-C (GLGC) | 5 | 74620377 | T | C |
| LDL-C (GLGC) | 5 | 74624234 | G | A |
| LDL-C (GLGC) | 5 | 74625512 | G | A |
| LDL-C (GLGC) | 5 | 74631262 | G | A |
| LDL-C (GLGC) | 5 | 74631352 | T | C |
| LDL-C (GLGC) | 5 | 74636484 | C | A |
| LDL-C (GLGC) | 5 | 74637213 | T | C |
| LDL-C (GLGC) | 5 | 74641707 | G | A |
| LDL-C (GLGC) | 5 | 74648496 | T | G |
| LDL-C (GLGC) | 5 | 74651084 | G | A |
| LDL-C (GLGC) | 5 | 74656539 | T | C |
| LDL-C (GLGC) | 5 | 74668589 | T | C |
| LDL-C (GLGC) | 5 | 74670454 | T | C |
| LDL-C (GLGC) | 5 | 74673707 | G | A |
| LDL-C (GLGC) | 5 | 74690076 | T | C |
| LDL-C (GLGC) | 5 | 74692776 | G | A |
| LDL-C (GLGC) | 5 | 74700126 | C | A |
| LDL-C (GLGC) | 5 | 74707762 | G | A |
| LDL-C (GLGC) | 5 | 74735038 | G | A |
| LDL-C (GLGC) | 5 | 74786083 | G | A |
| LDL-C (GLGC) | 5 | 74808237 | T | G |
| LDL-C (GLGC) | 5 | 74808417 | G | A |
| LDL-C (GLGC) | 5 | 74876613 | T | C |
| LDL-C (GLGC) | 5 | 74879890 | G | A |
| LDL-C (GLGC) | 5 | 74898606 | T | C |
| LDL-C (GLGC) | 5 | 74910870 | G | A |
| LDL-C (GLGC) | 5 | 74926778 | G | A |
| LDL-C (GLGC) | 5 | 74929312 | G | A |
| LDL-C (GLGC) | 5 | 74929647 | G | A |
| LDL-C (GLGC) | 5 | 74946518 | G | A |
| LDL-C (GLGC) | 5 | 74951901 | G | A |
| LDL-C (GLGC) | 5 | 74952861 | C | A |

**Table S14:** Genetic variants used in the presented analyses. Note that due to coverage of the GWAS chip the instruments differed across considered outcomes *(continued)*

| Exposure | Chromosome | Base-pair position (Build 37) | Other allele | Effect allele |
|---|---|---|---|---|
| LDL-C (GLGC) | 5 | 74959970 | G | A |
| LDL-C (GLGC) | 5 | 74961796 | T | G |
| LDL-C (GLGC) | 5 | 74964128 | T | C |
| LDL-C (GLGC) | 5 | 74965122 | G | A |
| LDL-C (GLGC) | 5 | 74965554 | G | A |
| LDL-C (GLGC) | 5 | 74967386 | T | C |
| LDL-C (GLGC) | 5 | 74970556 | T | G |
| LDL-C (GLGC) | 5 | 74970850 | G | A |
| LDL-C (GLGC) | 5 | 74971846 | G | A |
| LDL-C (GLGC) | 5 | 74972507 | T | G |
| LDL-C (GLGC) | 5 | 74975030 | T | C |
| LDL-C (GLGC) | 5 | 74984269 | T | C |
| LDL-C (GLGC) | 5 | 74984619 | G | A |
| LDL-C (GLGC) | 5 | 74986481 | T | C |
| LDL-C (GLGC) | 5 | 74988369 | G | A |
| LDL-C (GLGC) | 5 | 74997736 | T | C |
| LDL-C (GLGC) | 5 | 75003678 | T | C |
| LDL-C (GLGC) | 5 | 75003878 | T | C |
| LDL-C (GLGC) | 5 | 75012317 | T | C |
| LDL-C (GLGC) | 5 | 75015698 | G | A |
| LDL-C (GLGC) | 5 | 75024380 | T | G |
| LDL-C (GLGC) | 5 | 75030034 | G | A |
| LDL-C (GLGC) | 5 | 75034387 | T | C |
| LDL-C (GLGC) | 5 | 75037086 | G | A |
| LDL-C (GLGC) | 5 | 75042913 | G | A |
| LDL-C (GLGC) | 5 | 75132546 | T | G |
| LDL-C (GLGC) | 6 | 100600097 | T | C |
| LDL-C (GLGC) | 6 | 116296970 | T | G |
| LDL-C (GLGC) | 6 | 116305064 | T | C |
| LDL-C (GLGC) | 6 | 116309649 | T | C |
| LDL-C (GLGC) | 6 | 116373616 | T | C |
| LDL-C (GLGC) | 6 | 116381994 | T | C |
| LDL-C (GLGC) | 6 | 160402705 | G | A |
| LDL-C (GLGC) | 6 | 160415554 | T | C |
| LDL-C (GLGC) | 6 | 160443390 | G | A |
| LDL-C (GLGC) | 6 | 160468278 | G | A |
| LDL-C (GLGC) | 6 | 160494521 | T | C |
| LDL-C (GLGC) | 6 | 160496817 | T | C |
| LDL-C (GLGC) | 6 | 160508172 | T | C |
| LDL-C (GLGC) | 6 | 160514283 | G | A |
| LDL-C (GLGC) | 6 | 160537308 | T | C |
| LDL-C (GLGC) | 6 | 160538921 | G | A |
| LDL-C (GLGC) | 6 | 160543148 | T | C |
| LDL-C (GLGC) | 6 | 160543562 | G | A |

**Table S14:** Genetic variants used in the presented analyses. Note that due to coverage of the GWAS chip the instruments differed across considered outcomes *(continued)*

| Exposure | Chromosome | Base-pair position (Build 37) | Other allele | Effect allele |
|---|---|---|---|---|
| LDL-C (GLGC) | 6 | 160551486 | T | C |
| LDL-C (GLGC) | 6 | 160560933 | T | C |
| LDL-C (GLGC) | 6 | 160567928 | T | G |
| LDL-C (GLGC) | 6 | 160571499 | G | A |
| LDL-C (GLGC) | 6 | 160572866 | C | A |
| LDL-C (GLGC) | 6 | 160574037 | T | C |
| LDL-C (GLGC) | 6 | 160575366 | G | A |
| LDL-C (GLGC) | 6 | 160578860 | T | C |
| LDL-C (GLGC) | 6 | 160608804 | C | A |
| LDL-C (GLGC) | 6 | 160668389 | G | A |
| LDL-C (GLGC) | 6 | 160735281 | T | C |
| LDL-C (GLGC) | 6 | 160737950 | T | C |
| LDL-C (GLGC) | 6 | 160754712 | G | A |
| LDL-C (GLGC) | 6 | 160770360 | G | A |
| LDL-C (GLGC) | 6 | 160771192 | G | A |
| LDL-C (GLGC) | 6 | 160816409 | T | C |
| LDL-C (GLGC) | 6 | 160894878 | T | C |
| LDL-C (GLGC) | 6 | 160903622 | G | A |
| LDL-C (GLGC) | 6 | 160906086 | G | A |
| LDL-C (GLGC) | 6 | 160914460 | G | A |
| LDL-C (GLGC) | 6 | 160917559 | T | C |
| LDL-C (GLGC) | 6 | 160922870 | G | A |
| LDL-C (GLGC) | 6 | 160961137 | T | C |
| LDL-C (GLGC) | 6 | 160963230 | T | C |
| LDL-C (GLGC) | 6 | 160980330 | T | C |
| LDL-C (GLGC) | 6 | 160981736 | C | A |
| LDL-C (GLGC) | 6 | 160998148 | T | G |
| LDL-C (GLGC) | 6 | 161010118 | G | A |
| LDL-C (GLGC) | 6 | 161018174 | T | C |
| LDL-C (GLGC) | 6 | 161029728 | T | C |
| LDL-C (GLGC) | 6 | 161069941 | G | A |
| LDL-C (GLGC) | 6 | 16107867 | C | A |
| LDL-C (GLGC) | 6 | 161082695 | T | C |
| LDL-C (GLGC) | 6 | 161082799 | T | C |
| LDL-C (GLGC) | 6 | 16109163 | C | A |
| LDL-C (GLGC) | 6 | 16110186 | T | C |
| LDL-C (GLGC) | 6 | 16110445 | G | A |
| LDL-C (GLGC) | 6 | 161108144 | C | A |
| LDL-C (GLGC) | 6 | 161108536 | T | C |
| LDL-C (GLGC) | 6 | 16113321 | G | A |
| LDL-C (GLGC) | 6 | 16113733 | T | C |
| LDL-C (GLGC) | 6 | 16120819 | T | C |
| LDL-C (GLGC) | 6 | 16126934 | G | A |
| LDL-C (GLGC) | 6 | 16127407 | T | C |

**Table S14:** Genetic variants used in the presented analyses. Note that due to coverage of the GWAS chip the instruments differed across considered outcomes *(continued)*

| Exposure | Chromosome | Base-pair position (Build 37) | Other allele | Effect allele |
|---|---|---|---|---|
| LDL-C (GLGC) | 6 | 16134247 | T | C |
| LDL-C (GLGC) | 6 | 161385757 | G | A |
| LDL-C (GLGC) | 6 | 16160569 | T | G |
| LDL-C (GLGC) | 6 | 16162508 | T | C |
| LDL-C (GLGC) | 6 | 16175022 | G | A |
| LDL-C (GLGC) | 6 | 16175295 | G | A |
| LDL-C (GLGC) | 6 | 16193416 | T | G |
| LDL-C (GLGC) | 6 | 16196759 | G | A |
| LDL-C (GLGC) | 6 | 25488583 | T | C |
| LDL-C (GLGC) | 6 | 25821770 | T | C |
| LDL-C (GLGC) | 6 | 25842951 | T | G |
| LDL-C (GLGC) | 6 | 26093141 | G | A |
| LDL-C (GLGC) | 6 | 31011845 | T | C |
| LDL-C (GLGC) | 6 | 31019460 | G | A |
| LDL-C (GLGC) | 6 | 31129707 | T | C |
| LDL-C (GLGC) | 6 | 31237592 | C | A |
| LDL-C (GLGC) | 6 | 31238318 | G | A |
| LDL-C (GLGC) | 6 | 31240692 | G | A |
| LDL-C (GLGC) | 6 | 31243785 | T | G |
| LDL-C (GLGC) | 6 | 31253444 | T | C |
| LDL-C (GLGC) | 6 | 31265490 | T | C |
| LDL-C (GLGC) | 6 | 31272261 | T | C |
| LDL-C (GLGC) | 6 | 31336100 | C | A |
| LDL-C (GLGC) | 6 | 31341156 | G | A |
| LDL-C (GLGC) | 6 | 31352631 | G | A |
| LDL-C (GLGC) | 6 | 31353593 | G | A |
| LDL-C (GLGC) | 6 | 31353950 | C | A |
| LDL-C (GLGC) | 6 | 31354182 | T | C |
| LDL-C (GLGC) | 6 | 31355119 | T | C |
| LDL-C (GLGC) | 6 | 31360341 | T | C |
| LDL-C (GLGC) | 6 | 31362930 | T | C |
| LDL-C (GLGC) | 6 | 31377047 | T | C |
| LDL-C (GLGC) | 6 | 31378977 | G | A |
| LDL-C (GLGC) | 6 | 31380449 | T | C |
| LDL-C (GLGC) | 6 | 31380732 | T | C |
| LDL-C (GLGC) | 6 | 31384578 | G | A |
| LDL-C (GLGC) | 6 | 31384792 | T | C |
| LDL-C (GLGC) | 6 | 31407643 | T | C |
| LDL-C (GLGC) | 6 | 31407828 | T | C |
| LDL-C (GLGC) | 6 | 31409528 | T | C |
| LDL-C (GLGC) | 6 | 31410649 | T | C |
| LDL-C (GLGC) | 6 | 31440552 | T | G |
| LDL-C (GLGC) | 6 | 31441349 | G | A |
| LDL-C (GLGC) | 6 | 31507447 | T | C |

**Table S14:** Genetic variants used in the presented analyses. Note that due to coverage of the GWAS chip the instruments differed across considered outcomes *(continued)*

| Exposure | Chromosome | Base-pair position (Build 37) | Other allele | Effect allele |
|---|---|---|---|---|
| LDL-C (GLGC) | 6 | 31512799 | G | A |
| LDL-C (GLGC) | 6 | 31847012 | C | A |
| LDL-C (GLGC) | 6 | 31883957 | G | A |
| LDL-C (GLGC) | 6 | 31929014 | C | A |
| LDL-C (GLGC) | 6 | 31930441 | T | G |
| LDL-C (GLGC) | 6 | 31947460 | T | G |
| LDL-C (GLGC) | 6 | 32046275 | C | A |
| LDL-C (GLGC) | 6 | 32265528 | T | C |
| LDL-C (GLGC) | 6 | 32341398 | G | A |
| LDL-C (GLGC) | 6 | 32345086 | G | A |
| LDL-C (GLGC) | 6 | 32351566 | T | C |
| LDL-C (GLGC) | 6 | 32358533 | G | A |
| LDL-C (GLGC) | 6 | 32360341 | C | A |
| LDL-C (GLGC) | 6 | 32361388 | T | C |
| LDL-C (GLGC) | 6 | 32366421 | T | C |
| LDL-C (GLGC) | 6 | 32367604 | T | C |
| LDL-C (GLGC) | 6 | 32376471 | C | A |
| LDL-C (GLGC) | 6 | 32411362 | C | A |
| LDL-C (GLGC) | 6 | 32412435 | G | A |
| LDL-C (GLGC) | 6 | 32415858 | G | A |
| LDL-C (GLGC) | 6 | 32415975 | G | A |
| LDL-C (GLGC) | 6 | 32581889 | T | C |
| LDL-C (GLGC) | 6 | 32626272 | C | A |
| LDL-C (GLGC) | 6 | 32652317 | G | A |
| LDL-C (GLGC) | 6 | 32658310 | G | A |
| LDL-C (GLGC) | 6 | 32663644 | G | A |
| LDL-C (GLGC) | 6 | 32665420 | T | G |
| LDL-C (GLGC) | 6 | 32667364 | T | C |
| LDL-C (GLGC) | 6 | 32668804 | T | C |
| LDL-C (GLGC) | 6 | 32670576 | G | A |
| LDL-C (GLGC) | 6 | 32670874 | C | A |
| LDL-C (GLGC) | 6 | 32671103 | T | C |
| LDL-C (GLGC) | 6 | 32681277 | G | A |
| LDL-C (GLGC) | 6 | 32782582 | T | C |
| LDL-C (GLGC) | 6 | 32784620 | T | C |
| LDL-C (GLGC) | 6 | 32787570 | T | C |
| LDL-C (GLGC) | 6 | 32796967 | G | A |
| LDL-C (GLGC) | 6 | 34618893 | G | A |
| LDL-C (GLGC) | 6 | 34638847 | G | A |
| LDL-C (GLGC) | 6 | 35133074 | G | A |
| LDL-C (GLGC) | 6 | 39250837 | G | A |
| LDL-C (GLGC) | 6 | 52452585 | G | A |
| LDL-C (GLGC) | 6 | 52454299 | G | A |
| LDL-C (GLGC) | 6 | 52529666 | C | A |

**Table S14:** Genetic variants used in the presented analyses. Note that due to coverage of the GWAS chip the instruments differed across considered outcomes *(continued)*

| Exposure | Chromosome | Base-pair position (Build 37) | Other allele | Effect allele |
|---|---|---|---|---|
| LDL-C (GLGC) | 7 | 21488929 | G | A |
| LDL-C (GLGC) | 7 | 21496496 | T | C |
| LDL-C (GLGC) | 7 | 21563471 | T | C |
| LDL-C (GLGC) | 7 | 21582917 | T | C |
| LDL-C (GLGC) | 7 | 21589243 | G | A |
| LDL-C (GLGC) | 7 | 21601444 | T | C |
| LDL-C (GLGC) | 7 | 21603678 | G | A |
| LDL-C (GLGC) | 7 | 21607352 | T | C |
| LDL-C (GLGC) | 7 | 21612705 | T | C |
| LDL-C (GLGC) | 7 | 21615629 | T | G |
| LDL-C (GLGC) | 7 | 21627909 | G | A |
| LDL-C (GLGC) | 7 | 21647114 | T | G |
| LDL-C (GLGC) | 7 | 21654602 | T | C |
| LDL-C (GLGC) | 7 | 25991826 | T | C |
| LDL-C (GLGC) | 7 | 25997536 | G | A |
| LDL-C (GLGC) | 7 | 44570067 | T | C |
| LDL-C (GLGC) | 7 | 44570717 | C | A |
| LDL-C (GLGC) | 7 | 44577622 | G | A |
| LDL-C (GLGC) | 7 | 44580876 | T | G |
| LDL-C (GLGC) | 7 | 44580991 | T | C |
| LDL-C (GLGC) | 7 | 44581986 | T | C |
| LDL-C (GLGC) | 7 | 44585052 | T | C |
| LDL-C (GLGC) | 7 | 44602187 | T | G |
| LDL-C (GLGC) | 7 | 44615161 | G | A |
| LDL-C (GLGC) | 7 | 44617793 | G | A |
| LDL-C (GLGC) | 7 | 44643802 | G | A |
| LDL-C (GLGC) | 7 | 44657475 | T | C |
| LDL-C (GLGC) | 7 | 44709372 | T | C |
| LDL-C (GLGC) | 8 | 116648089 | G | A |
| LDL-C (GLGC) | 8 | 116648565 | C | A |
| LDL-C (GLGC) | 8 | 116650219 | G | A |
| LDL-C (GLGC) | 8 | 116650491 | T | C |
| LDL-C (GLGC) | 8 | 116659534 | G | A |
| LDL-C (GLGC) | 8 | 116663922 | G | A |
| LDL-C (GLGC) | 8 | 116670347 | T | C |
| LDL-C (GLGC) | 8 | 126447308 | T | C |
| LDL-C (GLGC) | 8 | 126449406 | T | C |
| LDL-C (GLGC) | 8 | 126456376 | G | A |
| LDL-C (GLGC) | 8 | 126459962 | G | A |
| LDL-C (GLGC) | 8 | 126459990 | G | A |
| LDL-C (GLGC) | 8 | 126460535 | G | A |
| LDL-C (GLGC) | 8 | 126463365 | G | A |
| LDL-C (GLGC) | 8 | 126468621 | T | G |
| LDL-C (GLGC) | 8 | 126470817 | G | A |

**Table S14:** Genetic variants used in the presented analyses. Note that due to coverage of the GWAS chip the instruments differed across considered outcomes *(continued)*

| Exposure | Chromosome | Base-pair position (Build 37) | Other allele | Effect allele |
|---|---|---|---|---|
| LDL-C (GLGC) | 8 | 126472487 | G | A |
| LDL-C (GLGC) | 8 | 126473441 | G | A |
| LDL-C (GLGC) | 8 | 126474306 | G | A |
| LDL-C (GLGC) | 8 | 126474356 | G | A |
| LDL-C (GLGC) | 8 | 126478644 | G | A |
| LDL-C (GLGC) | 8 | 126478758 | G | A |
| LDL-C (GLGC) | 8 | 126480367 | T | C |
| LDL-C (GLGC) | 8 | 126480526 | T | C |
| LDL-C (GLGC) | 8 | 126482077 | G | A |
| LDL-C (GLGC) | 8 | 126485531 | T | C |
| LDL-C (GLGC) | 8 | 126486287 | T | G |
| LDL-C (GLGC) | 8 | 126486409 | G | A |
| LDL-C (GLGC) | 8 | 126486906 | G | A |
| LDL-C (GLGC) | 8 | 126488235 | T | C |
| LDL-C (GLGC) | 8 | 126488930 | G | A |
| LDL-C (GLGC) | 8 | 126489818 | G | A |
| LDL-C (GLGC) | 8 | 126491304 | T | C |
| LDL-C (GLGC) | 8 | 126493392 | G | A |
| LDL-C (GLGC) | 8 | 126493746 | G | A |
| LDL-C (GLGC) | 8 | 126494562 | T | C |
| LDL-C (GLGC) | 8 | 126494764 | G | A |
| LDL-C (GLGC) | 8 | 126495818 | T | C |
| LDL-C (GLGC) | 8 | 126501177 | T | C |
| LDL-C (GLGC) | 8 | 126501528 | G | A |
| LDL-C (GLGC) | 8 | 126502526 | G | A |
| LDL-C (GLGC) | 8 | 126504726 | T | C |
| LDL-C (GLGC) | 8 | 126523523 | T | C |
| LDL-C (GLGC) | 8 | 126526063 | T | G |
| LDL-C (GLGC) | 8 | 144986793 | T | C |
| LDL-C (GLGC) | 8 | 144992103 | T | C |
| LDL-C (GLGC) | 8 | 145005561 | T | C |
| LDL-C (GLGC) | 8 | 145009610 | G | A |
| LDL-C (GLGC) | 8 | 145031265 | G | A |
| LDL-C (GLGC) | 8 | 145043543 | G | A |
| LDL-C (GLGC) | 8 | 145058986 | G | A |
| LDL-C (GLGC) | 8 | 145059425 | T | C |
| LDL-C (GLGC) | 8 | 145074452 | T | C |
| LDL-C (GLGC) | 8 | 18255709 | G | A |
| LDL-C (GLGC) | 8 | 18272377 | T | C |
| LDL-C (GLGC) | 8 | 18272438 | T | C |
| LDL-C (GLGC) | 8 | 55422424 | T | C |
| LDL-C (GLGC) | 8 | 55422440 | T | C |
| LDL-C (GLGC) | 8 | 55437524 | G | A |
| LDL-C (GLGC) | 8 | 59320389 | G | A |

**Table S14:** Genetic variants used in the presented analyses. Note that due to coverage of the GWAS chip the instruments differed across considered outcomes *(continued)*

| Exposure | Chromosome | Base-pair position (Build 37) | Other allele | Effect allele |
|---|---|---|---|---|
| LDL-C (GLGC) | 8 | 59332694 | G | A |
| LDL-C (GLGC) | 8 | 59339279 | T | C |
| LDL-C (GLGC) | 8 | 59388565 | T | C |
| LDL-C (GLGC) | 8 | 59412066 | T | G |
| LDL-C (GLGC) | 8 | 59544413 | G | A |
| LDL-C (GLGC) | 8 | 9168146 | T | C |
| LDL-C (GLGC) | 8 | 9171735 | T | C |
| LDL-C (GLGC) | 8 | 9172718 | G | A |
| LDL-C (GLGC) | 8 | 9175433 | T | C |
| LDL-C (GLGC) | 8 | 9177732 | T | G |
| LDL-C (GLGC) | 8 | 9178135 | T | C |
| LDL-C (GLGC) | 8 | 9178921 | G | A |
| LDL-C (GLGC) | 8 | 9183596 | G | A |
| LDL-C (GLGC) | 8 | 9184231 | G | A |
| LDL-C (GLGC) | 8 | 9184691 | T | C |
| LDL-C (GLGC) | 8 | 9187242 | G | A |
| LDL-C (GLGC) | 8 | 9192423 | G | A |
| LDL-C (GLGC) | 8 | 9197250 | G | A |
| LDL-C (GLGC) | 8 | 9201787 | T | C |
| LDL-C (GLGC) | 8 | 9206069 | G | A |
| LDL-C (GLGC) | 8 | 9206678 | G | A |
| LDL-C (GLGC) | 8 | 9219622 | T | G |
| LDL-C (GLGC) | 8 | 9228403 | C | A |
| LDL-C (GLGC) | 9 | 107653426 | G | A |
| LDL-C (GLGC) | 9 | 107664301 | T | C |
| LDL-C (GLGC) | 9 | 107665739 | G | A |
| LDL-C (GLGC) | 9 | 107667147 | T | C |
| LDL-C (GLGC) | 9 | 107672365 | G | A |
| LDL-C (GLGC) | 9 | 136042782 | G | A |
| LDL-C (GLGC) | 9 | 136049780 | G | A |
| LDL-C (GLGC) | 9 | 136057106 | G | A |
| LDL-C (GLGC) | 9 | 136064766 | G | A |
| LDL-C (GLGC) | 9 | 136075679 | C | A |
| LDL-C (GLGC) | 9 | 136084941 | T | C |
| LDL-C (GLGC) | 9 | 136087047 | T | C |
| LDL-C (GLGC) | 9 | 136087931 | T | G |
| LDL-C (GLGC) | 9 | 136093199 | G | A |
| LDL-C (GLGC) | 9 | 136102199 | T | C |
| LDL-C (GLGC) | 9 | 136102946 | G | A |
| LDL-C (GLGC) | 9 | 136103153 | T | C |
| LDL-C (GLGC) | 9 | 136109279 | G | A |
| LDL-C (GLGC) | 9 | 136112570 | T | C |
| LDL-C (GLGC) | 9 | 136115176 | G | A |
| LDL-C (GLGC) | 9 | 136120091 | G | A |

**Table S14:** Genetic variants used in the presented analyses. Note that due to coverage of the GWAS chip the instruments differed across considered outcomes *(continued)*

| Exposure | Chromosome | Base-pair position (Build 37) | Other allele | Effect allele |
|---|---|---|---|---|
| LDL-C (GLGC) | 9 | 136121652 | G | A |
| LDL-C (GLGC) | 9 | 136128558 | T | C |
| LDL-C (GLGC) | 9 | 136131322 | T | G |
| LDL-C (GLGC) | 9 | 136132350 | T | C |
| LDL-C (GLGC) | 9 | 136132617 | G | A |
| LDL-C (GLGC) | 9 | 136132633 | G | A |
| LDL-C (GLGC) | 9 | 136132754 | C | A |
| LDL-C (GLGC) | 9 | 136132873 | T | C |
| LDL-C (GLGC) | 9 | 136133714 | G | A |
| LDL-C (GLGC) | 9 | 136137065 | G | A |
| LDL-C (GLGC) | 9 | 136139265 | C | A |
| LDL-C (GLGC) | 9 | 136139297 | T | C |
| LDL-C (GLGC) | 9 | 136139754 | T | C |
| LDL-C (GLGC) | 9 | 136142185 | T | C |
| LDL-C (GLGC) | 9 | 136145471 | G | A |
| LDL-C (GLGC) | 9 | 136149399 | G | A |
| LDL-C (GLGC) | 9 | 136149722 | G | A |
| LDL-C (GLGC) | 9 | 136153875 | T | C |
| LDL-C (GLGC) | 9 | 136153981 | T | C |
| LDL-C (GLGC) | 9 | 136154867 | T | G |
| LDL-C (GLGC) | 9 | 136155359 | G | A |
| LDL-C (GLGC) | 9 | 136155444 | T | C |
| LDL-C (GLGC) | 9 | 136157133 | G | A |
| LDL-C (GLGC) | 9 | 136182475 | G | A |
| LDL-C (GLGC) | 9 | 136191010 | T | C |
| LDL-C (GLGC) | 9 | 136194595 | T | C |
| LDL-C (GLGC) | 9 | 136237672 | C | A |
| LDL-C (GLGC) | 9 | 136246666 | T | C |
| LDL-C (GLGC) | 9 | 136267149 | T | C |
| LDL-C (GLGC) | 9 | 136268084 | G | A |
| LDL-C (GLGC) | 9 | 136269576 | C | A |
| LDL-C (GLGC) | 9 | 136323754 | G | A |
| LDL-C (GLGC) | 9 | 136329954 | T | C |
| LDL-C (GLGC) | 9 | 136330428 | G | A |
| LDL-C (GLGC) | 9 | 136334910 | T | C |
| LDL-C (GLGC) | 9 | 136348194 | T | C |
| LDL-C (GLGC) | 9 | 136359599 | T | C |
| LDL-C (GLGC) | 9 | 2640759 | G | A |
| HDL-C (GLGC) | 10 | 113921825 | G | A |
| HDL-C (GLGC) | 10 | 113940329 | T | C |
| HDL-C (GLGC) | 10 | 113991397 | T | C |
| HDL-C (GLGC) | 10 | 113992861 | T | G |
| HDL-C (GLGC) | 10 | 114045333 | T | C |
| HDL-C (GLGC) | 10 | 114048792 | T | C |

**Table S14:** Genetic variants used in the presented analyses. Note that due to coverage of the GWAS chip the instruments differed across considered outcomes *(continued)*

| Exposure | Chromosome | Base-pair position (Build 37) | Other allele | Effect allele |
|---|---|---|---|---|
| HDL-C (GLGC) | 10 | 115789375 | T | C |
| HDL-C (GLGC) | 10 | 115792787 | G | A |
| HDL-C (GLGC) | 10 | 45932264 | T | C |
| HDL-C (GLGC) | 10 | 45938746 | T | C |
| HDL-C (GLGC) | 10 | 45958881 | C | A |
| HDL-C (GLGC) | 10 | 45988597 | G | A |
| HDL-C (GLGC) | 10 | 46031747 | G | A |
| HDL-C (GLGC) | 10 | 46076235 | G | A |
| HDL-C (GLGC) | 10 | 46088061 | T | C |
| HDL-C (GLGC) | 10 | 46094214 | T | G |
| HDL-C (GLGC) | 10 | 5799613 | G | A |
| HDL-C (GLGC) | 10 | 65050659 | G | A |
| HDL-C (GLGC) | 10 | 65066186 | T | G |
| HDL-C (GLGC) | 10 | 65350383 | G | A |
| HDL-C (GLGC) | 10 | 94805665 | G | A |
| HDL-C (GLGC) | 11 | 10388782 | G | A |
| HDL-C (GLGC) | 11 | 109979945 | T | C |
| HDL-C (GLGC) | 11 | 116505440 | G | A |
| HDL-C (GLGC) | 11 | 116520398 | T | C |
| HDL-C (GLGC) | 11 | 116522854 | G | A |
| HDL-C (GLGC) | 11 | 116522961 | T | C |
| HDL-C (GLGC) | 11 | 116523844 | G | A |
| HDL-C (GLGC) | 11 | 116525730 | T | C |
| HDL-C (GLGC) | 11 | 116526322 | C | A |
| HDL-C (GLGC) | 11 | 116527282 | T | C |
| HDL-C (GLGC) | 11 | 116529188 | T | G |
| HDL-C (GLGC) | 11 | 116529442 | T | C |
| HDL-C (GLGC) | 11 | 116531739 | T | C |
| HDL-C (GLGC) | 11 | 116532221 | T | C |
| HDL-C (GLGC) | 11 | 116536418 | G | A |
| HDL-C (GLGC) | 11 | 116536476 | T | C |
| HDL-C (GLGC) | 11 | 116552562 | T | C |
| HDL-C (GLGC) | 11 | 116553025 | G | A |
| HDL-C (GLGC) | 11 | 116554228 | C | A |
| HDL-C (GLGC) | 11 | 116562580 | C | A |
| HDL-C (GLGC) | 11 | 116566117 | T | C |
| HDL-C (GLGC) | 11 | 116571608 | G | A |
| HDL-C (GLGC) | 11 | 116572089 | T | C |
| HDL-C (GLGC) | 11 | 116574336 | G | A |
| HDL-C (GLGC) | 11 | 116574463 | G | A |
| HDL-C (GLGC) | 11 | 116574488 | G | A |
| HDL-C (GLGC) | 11 | 116576400 | T | C |
| HDL-C (GLGC) | 11 | 116576415 | G | A |
| HDL-C (GLGC) | 11 | 116578223 | T | C |

**Table S14:** Genetic variants used in the presented analyses. Note that due to coverage of the GWAS chip the instruments differed across considered outcomes *(continued)*

| Exposure | Chromosome | Base-pair position (Build 37) | Other allele | Effect allele |
|---|---|---|---|---|
| HDL-C (GLGC) | 11 | 116579663 | T | C |
| HDL-C (GLGC) | 11 | 116581259 | T | C |
| HDL-C (GLGC) | 11 | 116584987 | T | C |
| HDL-C (GLGC) | 11 | 116586283 | T | C |
| HDL-C (GLGC) | 11 | 116590787 | G | A |
| HDL-C (GLGC) | 11 | 116595756 | T | C |
| HDL-C (GLGC) | 11 | 116596309 | C | A |
| HDL-C (GLGC) | 11 | 116598988 | G | A |
| HDL-C (GLGC) | 11 | 116599392 | T | C |
| HDL-C (GLGC) | 11 | 116603677 | G | A |
| HDL-C (GLGC) | 11 | 116603724 | C | A |
| HDL-C (GLGC) | 11 | 116604070 | T | C |
| HDL-C (GLGC) | 11 | 116607368 | C | A |
| HDL-C (GLGC) | 11 | 116610249 | G | A |
| HDL-C (GLGC) | 11 | 116611457 | T | C |
| HDL-C (GLGC) | 11 | 116612128 | G | A |
| HDL-C (GLGC) | 11 | 116615244 | G | A |
| HDL-C (GLGC) | 11 | 116619867 | C | A |
| HDL-C (GLGC) | 11 | 116621770 | G | A |
| HDL-C (GLGC) | 11 | 116624601 | C | A |
| HDL-C (GLGC) | 11 | 116624703 | T | G |
| HDL-C (GLGC) | 11 | 116629949 | T | C |
| HDL-C (GLGC) | 11 | 116633825 | T | C |
| HDL-C (GLGC) | 11 | 116633947 | G | A |
| HDL-C (GLGC) | 11 | 116639941 | G | A |
| HDL-C (GLGC) | 11 | 116640237 | G | A |
| HDL-C (GLGC) | 11 | 116645275 | T | C |
| HDL-C (GLGC) | 11 | 116648776 | C | A |
| HDL-C (GLGC) | 11 | 116649135 | G | A |
| HDL-C (GLGC) | 11 | 116649538 | G | A |
| HDL-C (GLGC) | 11 | 116652351 | G | A |
| HDL-C (GLGC) | 11 | 116652423 | G | A |
| HDL-C (GLGC) | 11 | 116653762 | T | C |
| HDL-C (GLGC) | 11 | 116654159 | G | A |
| HDL-C (GLGC) | 11 | 116655600 | G | A |
| HDL-C (GLGC) | 11 | 116657561 | T | C |
| HDL-C (GLGC) | 11 | 116658031 | T | G |
| HDL-C (GLGC) | 11 | 116660686 | G | A |
| HDL-C (GLGC) | 11 | 116660813 | G | A |
| HDL-C (GLGC) | 11 | 116661826 | T | C |
| HDL-C (GLGC) | 11 | 116662579 | T | C |
| HDL-C (GLGC) | 11 | 116667545 | T | C |
| HDL-C (GLGC) | 11 | 116670676 | C | A |
| HDL-C (GLGC) | 11 | 116673768 | G | A |

**Table S14:** Genetic variants used in the presented analyses. Note that due to coverage of the GWAS chip the instruments differed across considered outcomes *(continued)*

| Exposure | Chromosome | Base-pair position (Build 37) | Other allele | Effect allele |
|---|---|---|---|---|
| HDL-C (GLGC) | 11 | 116721826 | T | C |
| HDL-C (GLGC) | 11 | 116723171 | T | C |
| HDL-C (GLGC) | 11 | 116734545 | G | A |
| HDL-C (GLGC) | 11 | 116758454 | G | A |
| HDL-C (GLGC) | 11 | 116826870 | T | C |
| HDL-C (GLGC) | 11 | 116831912 | T | C |
| HDL-C (GLGC) | 11 | 116871223 | G | A |
| HDL-C (GLGC) | 11 | 116907449 | T | C |
| HDL-C (GLGC) | 11 | 116912730 | T | C |
| HDL-C (GLGC) | 11 | 116929995 | G | A |
| HDL-C (GLGC) | 11 | 116942753 | T | C |
| HDL-C (GLGC) | 11 | 116959039 | G | A |
| HDL-C (GLGC) | 11 | 116980641 | T | G |
| HDL-C (GLGC) | 11 | 117027942 | T | C |
| HDL-C (GLGC) | 11 | 117046197 | G | A |
| HDL-C (GLGC) | 11 | 117065476 | G | A |
| HDL-C (GLGC) | 11 | 117075566 | T | C |
| HDL-C (GLGC) | 11 | 117094491 | T | C |
| HDL-C (GLGC) | 11 | 117183650 | T | C |
| HDL-C (GLGC) | 11 | 117267884 | G | A |
| HDL-C (GLGC) | 11 | 122504251 | T | G |
| HDL-C (GLGC) | 11 | 122508163 | T | C |
| HDL-C (GLGC) | 11 | 122509237 | G | A |
| HDL-C (GLGC) | 11 | 122522375 | T | C |
| HDL-C (GLGC) | 11 | 122563338 | G | A |
| HDL-C (GLGC) | 11 | 126228000 | T | C |
| HDL-C (GLGC) | 11 | 126228659 | T | G |
| HDL-C (GLGC) | 11 | 13355770 | T | C |
| HDL-C (GLGC) | 11 | 13357183 | G | A |
| HDL-C (GLGC) | 11 | 14276999 | G | A |
| HDL-C (GLGC) | 11 | 14504463 | C | A |
| HDL-C (GLGC) | 11 | 45723298 | G | A |
| HDL-C (GLGC) | 11 | 45734387 | T | C |
| HDL-C (GLGC) | 11 | 45737527 | T | C |
| HDL-C (GLGC) | 11 | 45739162 | T | C |
| HDL-C (GLGC) | 11 | 45740020 | T | C |
| HDL-C (GLGC) | 11 | 45740173 | G | A |
| HDL-C (GLGC) | 11 | 45747590 | T | G |
| HDL-C (GLGC) | 11 | 45755505 | T | C |
| HDL-C (GLGC) | 11 | 45756960 | C | A |
| HDL-C (GLGC) | 11 | 45757401 | T | C |
| HDL-C (GLGC) | 11 | 45762123 | G | A |
| HDL-C (GLGC) | 11 | 45839709 | G | A |
| HDL-C (GLGC) | 11 | 46308549 | T | C |

**Table S14:** Genetic variants used in the presented analyses. Note that due to coverage of the GWAS chip the instruments differed across considered outcomes *(continued)*

| Exposure | Chromosome | Base-pair position (Build 37) | Other allele | Effect allele |
|---|---|---|---|---|
| HDL-C (GLGC) | 11 | 46309968 | G | A |
| HDL-C (GLGC) | 11 | 46702160 | T | C |
| HDL-C (GLGC) | 11 | 46722221 | T | C |
| HDL-C (GLGC) | 11 | 46725196 | T | C |
| HDL-C (GLGC) | 11 | 46745003 | T | C |
| HDL-C (GLGC) | 11 | 46749648 | G | A |
| HDL-C (GLGC) | 11 | 46768041 | G | A |
| HDL-C (GLGC) | 11 | 46769788 | C | A |
| HDL-C (GLGC) | 11 | 46798631 | G | A |
| HDL-C (GLGC) | 11 | 46805575 | G | A |
| HDL-C (GLGC) | 11 | 46810916 | C | A |
| HDL-C (GLGC) | 11 | 46857279 | T | C |
| HDL-C (GLGC) | 11 | 46872873 | G | A |
| HDL-C (GLGC) | 11 | 46890165 | T | C |
| HDL-C (GLGC) | 11 | 46890197 | G | A |
| HDL-C (GLGC) | 11 | 46897446 | G | A |
| HDL-C (GLGC) | 11 | 46898771 | T | C |
| HDL-C (GLGC) | 11 | 46911704 | T | C |
| HDL-C (GLGC) | 11 | 46917983 | G | A |
| HDL-C (GLGC) | 11 | 46939027 | G | A |
| HDL-C (GLGC) | 11 | 46975125 | T | C |
| HDL-C (GLGC) | 11 | 46981646 | G | A |
| HDL-C (GLGC) | 11 | 47127153 | G | A |
| HDL-C (GLGC) | 11 | 47160623 | T | C |
| HDL-C (GLGC) | 11 | 47179861 | C | A |
| HDL-C (GLGC) | 11 | 47182370 | G | A |
| HDL-C (GLGC) | 11 | 47201924 | G | A |
| HDL-C (GLGC) | 11 | 47245803 | G | A |
| HDL-C (GLGC) | 11 | 47246397 | G | A |
| HDL-C (GLGC) | 11 | 47247621 | G | A |
| HDL-C (GLGC) | 11 | 47249294 | G | A |
| HDL-C (GLGC) | 11 | 47255598 | T | C |
| HDL-C (GLGC) | 11 | 47266808 | T | G |
| HDL-C (GLGC) | 11 | 47269159 | T | C |
| HDL-C (GLGC) | 11 | 47270255 | T | C |
| HDL-C (GLGC) | 11 | 47275064 | G | A |
| HDL-C (GLGC) | 11 | 47277090 | T | C |
| HDL-C (GLGC) | 11 | 47300155 | G | A |
| HDL-C (GLGC) | 11 | 47303275 | T | C |
| HDL-C (GLGC) | 11 | 47306585 | G | A |
| HDL-C (GLGC) | 11 | 47312892 | T | G |
| HDL-C (GLGC) | 11 | 47316691 | C | A |
| HDL-C (GLGC) | 11 | 47346145 | G | A |
| HDL-C (GLGC) | 11 | 47346940 | G | A |

**Table S14:** Genetic variants used in the presented analyses. Note that due to coverage of the GWAS chip the instruments differed across considered outcomes *(continued)*

| Exposure | Chromosome | Base-pair position (Build 37) | Other allele | Effect allele |
|---|---|---|---|---|
| HDL-C (GLGC) | 11 | 47349969 | C | A |
| HDL-C (GLGC) | 11 | 47350287 | G | A |
| HDL-C (GLGC) | 11 | 47359706 | C | A |
| HDL-C (GLGC) | 11 | 47365199 | G | A |
| HDL-C (GLGC) | 11 | 47370607 | T | C |
| HDL-C (GLGC) | 11 | 47370957 | C | A |
| HDL-C (GLGC) | 11 | 47378399 | G | A |
| HDL-C (GLGC) | 11 | 47380593 | G | A |
| HDL-C (GLGC) | 11 | 47380995 | T | C |
| HDL-C (GLGC) | 11 | 47401171 | C | A |
| HDL-C (GLGC) | 11 | 47411736 | G | A |
| HDL-C (GLGC) | 11 | 47419129 | G | A |
| HDL-C (GLGC) | 11 | 47430458 | T | C |
| HDL-C (GLGC) | 11 | 47441513 | G | A |
| HDL-C (GLGC) | 11 | 47454271 | C | A |
| HDL-C (GLGC) | 11 | 47454701 | G | A |
| HDL-C (GLGC) | 11 | 47456763 | G | A |
| HDL-C (GLGC) | 11 | 47465955 | G | A |
| HDL-C (GLGC) | 11 | 47468791 | T | C |
| HDL-C (GLGC) | 11 | 47470345 | G | A |
| HDL-C (GLGC) | 11 | 47471053 | T | C |
| HDL-C (GLGC) | 11 | 47558220 | G | A |
| HDL-C (GLGC) | 11 | 47566352 | T | C |
| HDL-C (GLGC) | 11 | 47607912 | T | C |
| HDL-C (GLGC) | 11 | 47611152 | T | C |
| HDL-C (GLGC) | 11 | 47629793 | T | C |
| HDL-C (GLGC) | 11 | 47650993 | T | C |
| HDL-C (GLGC) | 11 | 47658314 | T | C |
| HDL-C (GLGC) | 11 | 47663049 | G | A |
| HDL-C (GLGC) | 11 | 47702395 | G | A |
| HDL-C (GLGC) | 11 | 47773226 | T | C |
| HDL-C (GLGC) | 11 | 47803907 | T | C |
| HDL-C (GLGC) | 11 | 47858755 | T | C |
| HDL-C (GLGC) | 11 | 47868474 | T | C |
| HDL-C (GLGC) | 11 | 47877493 | T | C |
| HDL-C (GLGC) | 11 | 47888629 | G | A |
| HDL-C (GLGC) | 11 | 47901269 | T | C |
| HDL-C (GLGC) | 11 | 47910823 | T | G |
| HDL-C (GLGC) | 11 | 47969152 | T | C |
| HDL-C (GLGC) | 11 | 47983477 | G | A |
| HDL-C (GLGC) | 11 | 48018355 | G | A |
| HDL-C (GLGC) | 11 | 48024893 | G | A |
| HDL-C (GLGC) | 11 | 48028343 | T | G |
| HDL-C (GLGC) | 11 | 48050391 | G | A |

**Table S14:** Genetic variants used in the presented analyses. Note that due to coverage of the GWAS chip the instruments differed across considered outcomes *(continued)*

| Exposure | Chromosome | Base-pair position (Build 37) | Other allele | Effect allele |
|---|---|---|---|---|
| HDL-C (GLGC) | 11 | 48050995 | G | A |
| HDL-C (GLGC) | 11 | 48051442 | C | A |
| HDL-C (GLGC) | 11 | 48059091 | T | C |
| HDL-C (GLGC) | 11 | 48094879 | T | C |
| HDL-C (GLGC) | 11 | 48097480 | T | C |
| HDL-C (GLGC) | 11 | 48115089 | T | C |
| HDL-C (GLGC) | 11 | 48120936 | G | A |
| HDL-C (GLGC) | 11 | 48150576 | G | A |
| HDL-C (GLGC) | 11 | 48157869 | T | C |
| HDL-C (GLGC) | 11 | 48207259 | G | A |
| HDL-C (GLGC) | 11 | 48234680 | G | A |
| HDL-C (GLGC) | 11 | 48246457 | G | A |
| HDL-C (GLGC) | 11 | 48293931 | T | C |
| HDL-C (GLGC) | 11 | 48382471 | G | A |
| HDL-C (GLGC) | 11 | 48518893 | G | A |
| HDL-C (GLGC) | 11 | 48590678 | T | G |
| HDL-C (GLGC) | 11 | 48616694 | G | A |
| HDL-C (GLGC) | 11 | 48626363 | C | A |
| HDL-C (GLGC) | 11 | 48688784 | T | C |
| HDL-C (GLGC) | 11 | 48807911 | T | C |
| HDL-C (GLGC) | 11 | 49011009 | T | C |
| HDL-C (GLGC) | 11 | 49012509 | G | A |
| HDL-C (GLGC) | 11 | 49381198 | G | A |
| HDL-C (GLGC) | 11 | 49505175 | T | C |
| HDL-C (GLGC) | 11 | 49610094 | T | G |
| HDL-C (GLGC) | 11 | 49644966 | T | C |
| HDL-C (GLGC) | 11 | 50096095 | T | G |
| HDL-C (GLGC) | 11 | 50367359 | T | C |
| HDL-C (GLGC) | 11 | 51526861 | G | A |
| HDL-C (GLGC) | 11 | 56180753 | G | A |
| HDL-C (GLGC) | 11 | 56436140 | G | A |
| HDL-C (GLGC) | 11 | 61546592 | G | A |
| HDL-C (GLGC) | 11 | 61548559 | G | A |
| HDL-C (GLGC) | 11 | 61548874 | G | A |
| HDL-C (GLGC) | 11 | 61549458 | G | A |
| HDL-C (GLGC) | 11 | 61560081 | G | A |
| HDL-C (GLGC) | 11 | 61564299 | T | G |
| HDL-C (GLGC) | 11 | 61565908 | T | C |
| HDL-C (GLGC) | 11 | 61580635 | T | C |
| HDL-C (GLGC) | 11 | 61593816 | T | C |
| HDL-C (GLGC) | 11 | 61597212 | T | C |
| HDL-C (GLGC) | 11 | 61597972 | G | A |
| HDL-C (GLGC) | 11 | 61600342 | C | A |
| HDL-C (GLGC) | 11 | 61601908 | G | A |

**Table S14:** Genetic variants used in the presented analyses. Note that due to coverage of the GWAS chip the instruments differed across considered outcomes *(continued)*

| Exposure | Chromosome | Base-pair position (Build 37) | Other allele | Effect allele |
|---|---|---|---|---|
| HDL-C (GLGC) | 11 | 61603237 | T | C |
| HDL-C (GLGC) | 11 | 61605215 | G | A |
| HDL-C (GLGC) | 11 | 61605613 | T | C |
| HDL-C (GLGC) | 11 | 61609750 | T | C |
| HDL-C (GLGC) | 11 | 61611694 | G | A |
| HDL-C (GLGC) | 11 | 61616012 | G | A |
| HDL-C (GLGC) | 11 | 61618608 | G | A |
| HDL-C (GLGC) | 11 | 61618831 | T | C |
| HDL-C (GLGC) | 11 | 61623793 | G | A |
| HDL-C (GLGC) | 11 | 61624181 | T | C |
| HDL-C (GLGC) | 11 | 61624414 | T | C |
| HDL-C (GLGC) | 11 | 61626921 | T | G |
| HDL-C (GLGC) | 11 | 61629122 | G | A |
| HDL-C (GLGC) | 11 | 61639488 | T | C |
| HDL-C (GLGC) | 11 | 61641542 | T | G |
| HDL-C (GLGC) | 11 | 61647288 | G | A |
| HDL-C (GLGC) | 11 | 61655305 | G | A |
| HDL-C (GLGC) | 11 | 61656117 | G | A |
| HDL-C (GLGC) | 11 | 65391317 | G | A |
| HDL-C (GLGC) | 11 | 75455021 | C | A |
| HDL-C (GLGC) | 12 | 107174646 | C | A |
| HDL-C (GLGC) | 12 | 107255986 | T | G |
| HDL-C (GLGC) | 12 | 109779616 | T | C |
| HDL-C (GLGC) | 12 | 109783690 | T | C |
| HDL-C (GLGC) | 12 | 109795166 | T | G |
| HDL-C (GLGC) | 12 | 109800531 | T | C |
| HDL-C (GLGC) | 12 | 109832545 | T | G |
| HDL-C (GLGC) | 12 | 109840940 | G | A |
| HDL-C (GLGC) | 12 | 109866528 | C | A |
| HDL-C (GLGC) | 12 | 109874230 | G | A |
| HDL-C (GLGC) | 12 | 109875981 | G | A |
| HDL-C (GLGC) | 12 | 109880279 | C | A |
| HDL-C (GLGC) | 12 | 109883374 | T | C |
| HDL-C (GLGC) | 12 | 109890339 | G | A |
| HDL-C (GLGC) | 12 | 109890510 | T | C |
| HDL-C (GLGC) | 12 | 109898732 | G | A |
| HDL-C (GLGC) | 12 | 109969686 | T | C |
| HDL-C (GLGC) | 12 | 109977464 | T | G |
| HDL-C (GLGC) | 12 | 109980516 | T | C |
| HDL-C (GLGC) | 12 | 109980619 | G | A |
| HDL-C (GLGC) | 12 | 109986935 | G | A |
| HDL-C (GLGC) | 12 | 109998097 | G | A |
| HDL-C (GLGC) | 12 | 110000193 | T | C |
| HDL-C (GLGC) | 12 | 110002777 | T | C |

**Table S14:** Genetic variants used in the presented analyses. Note that due to coverage of the GWAS chip the instruments differed across considered outcomes *(continued)*

| Exposure | Chromosome | Base-pair position (Build 37) | Other allele | Effect allele |
|---|---|---|---|---|
| HDL-C (GLGC) | 12 | 110004040 | T | C |
| HDL-C (GLGC) | 12 | 110010866 | G | A |
| HDL-C (GLGC) | 12 | 110027795 | G | A |
| HDL-C (GLGC) | 12 | 110046051 | T | C |
| HDL-C (GLGC) | 12 | 110048015 | T | C |
| HDL-C (GLGC) | 12 | 110055044 | T | C |
| HDL-C (GLGC) | 12 | 110059431 | G | A |
| HDL-C (GLGC) | 12 | 111910219 | G | A |
| HDL-C (GLGC) | 12 | 112083162 | T | C |
| HDL-C (GLGC) | 12 | 112871372 | G | A |
| HDL-C (GLGC) | 12 | 122781897 | G | A |
| HDL-C (GLGC) | 12 | 123171218 | G | A |
| HDL-C (GLGC) | 12 | 123200768 | T | G |
| HDL-C (GLGC) | 12 | 123311981 | T | C |
| HDL-C (GLGC) | 12 | 123332032 | T | C |
| HDL-C (GLGC) | 12 | 123344021 | G | A |
| HDL-C (GLGC) | 12 | 123387922 | T | G |
| HDL-C (GLGC) | 12 | 123727443 | G | A |
| HDL-C (GLGC) | 12 | 123736084 | T | G |
| HDL-C (GLGC) | 12 | 123775127 | G | A |
| HDL-C (GLGC) | 12 | 123791227 | T | C |
| HDL-C (GLGC) | 12 | 123796238 | T | C |
| HDL-C (GLGC) | 12 | 123798074 | G | A |
| HDL-C (GLGC) | 12 | 123800376 | G | A |
| HDL-C (GLGC) | 12 | 124317416 | G | A |
| HDL-C (GLGC) | 12 | 124393697 | T | G |
| HDL-C (GLGC) | 12 | 124399550 | T | C |
| HDL-C (GLGC) | 12 | 124417889 | T | C |
| HDL-C (GLGC) | 12 | 124446728 | G | A |
| HDL-C (GLGC) | 12 | 124460167 | T | G |
| HDL-C (GLGC) | 12 | 124494540 | T | C |
| HDL-C (GLGC) | 12 | 124505444 | T | C |
| HDL-C (GLGC) | 12 | 124506631 | T | C |
| HDL-C (GLGC) | 12 | 124520331 | G | A |
| HDL-C (GLGC) | 12 | 124537575 | G | A |
| HDL-C (GLGC) | 12 | 124561803 | T | C |
| HDL-C (GLGC) | 12 | 124566777 | T | C |
| HDL-C (GLGC) | 12 | 124583446 | G | A |
| HDL-C (GLGC) | 12 | 125259888 | G | A |
| HDL-C (GLGC) | 12 | 125260645 | G | A |
| HDL-C (GLGC) | 12 | 125261593 | T | C |
| HDL-C (GLGC) | 12 | 125261839 | T | C |
| HDL-C (GLGC) | 12 | 125262104 | G | A |
| HDL-C (GLGC) | 12 | 125264293 | G | A |

**Table S14:** Genetic variants used in the presented analyses. Note that due to coverage of the GWAS chip the instruments differed across considered outcomes *(continued)*

| Exposure | Chromosome | Base-pair position (Build 37) | Other allele | Effect allele |
|---|---|---|---|---|
| HDL-C (GLGC) | 12 | 125265636 | G | A |
| HDL-C (GLGC) | 12 | 125289265 | G | A |
| HDL-C (GLGC) | 12 | 125320155 | T | G |
| HDL-C (GLGC) | 12 | 125327352 | G | A |
| HDL-C (GLGC) | 12 | 125327404 | T | C |
| HDL-C (GLGC) | 12 | 125328375 | G | A |
| HDL-C (GLGC) | 12 | 125335253 | C | A |
| HDL-C (GLGC) | 12 | 125351116 | T | C |
| HDL-C (GLGC) | 12 | 125380232 | G | A |
| HDL-C (GLGC) | 12 | 125439806 | T | C |
| HDL-C (GLGC) | 12 | 20463526 | G | A |
| HDL-C (GLGC) | 12 | 20473758 | C | A |
| HDL-C (GLGC) | 12 | 57696677 | G | A |
| HDL-C (GLGC) | 12 | 57809456 | T | C |
| HDL-C (GLGC) | 12 | 57848639 | G | A |
| HDL-C (GLGC) | 12 | 57851182 | G | A |
| HDL-C (GLGC) | 12 | 58012233 | G | A |
| HDL-C (GLGC) | 14 | 105277209 | G | A |
| HDL-C (GLGC) | 15 | 41827655 | T | C |
| HDL-C (GLGC) | 15 | 42683787 | G | A |
| HDL-C (GLGC) | 15 | 44244523 | T | C |
| HDL-C (GLGC) | 15 | 44269775 | C | A |
| HDL-C (GLGC) | 15 | 58448287 | G | A |
| HDL-C (GLGC) | 15 | 58450274 | G | A |
| HDL-C (GLGC) | 15 | 58525552 | T | G |
| HDL-C (GLGC) | 15 | 58542197 | T | C |
| HDL-C (GLGC) | 15 | 58544607 | G | A |
| HDL-C (GLGC) | 15 | 58549619 | T | G |
| HDL-C (GLGC) | 15 | 58552606 | T | C |
| HDL-C (GLGC) | 15 | 58555270 | C | A |
| HDL-C (GLGC) | 15 | 58557062 | G | A |
| HDL-C (GLGC) | 15 | 58558746 | T | C |
| HDL-C (GLGC) | 15 | 58576226 | T | C |
| HDL-C (GLGC) | 15 | 58577163 | G | A |
| HDL-C (GLGC) | 15 | 58579956 | G | A |
| HDL-C (GLGC) | 15 | 58580525 | G | A |
| HDL-C (GLGC) | 15 | 58580781 | T | C |
| HDL-C (GLGC) | 15 | 58582540 | T | C |
| HDL-C (GLGC) | 15 | 58591415 | T | C |
| HDL-C (GLGC) | 15 | 58593730 | T | C |
| HDL-C (GLGC) | 15 | 58604998 | G | A |
| HDL-C (GLGC) | 15 | 58610852 | T | C |
| HDL-C (GLGC) | 15 | 58611859 | T | G |
| HDL-C (GLGC) | 15 | 58613666 | G | A |

**Table S14:** Genetic variants used in the presented analyses. Note that due to coverage of the GWAS chip the instruments differed across considered outcomes *(continued)*

| Exposure | Chromosome | Base-pair position (Build 37) | Other allele | Effect allele |
|---|---|---|---|---|
| HDL-C (GLGC) | 15 | 58619349 | C | A |
| HDL-C (GLGC) | 15 | 58619457 | G | A |
| HDL-C (GLGC) | 15 | 58623328 | T | G |
| HDL-C (GLGC) | 15 | 58623424 | G | A |
| HDL-C (GLGC) | 15 | 58623465 | C | A |
| HDL-C (GLGC) | 15 | 58623525 | T | C |
| HDL-C (GLGC) | 15 | 58630019 | T | G |
| HDL-C (GLGC) | 15 | 58635583 | C | A |
| HDL-C (GLGC) | 15 | 58635891 | G | A |
| HDL-C (GLGC) | 15 | 58643839 | T | C |
| HDL-C (GLGC) | 15 | 58649477 | T | C |
| HDL-C (GLGC) | 15 | 58654491 | T | C |
| HDL-C (GLGC) | 15 | 58666341 | T | C |
| HDL-C (GLGC) | 15 | 58666679 | T | C |
| HDL-C (GLGC) | 15 | 58671178 | C | A |
| HDL-C (GLGC) | 15 | 58672134 | T | C |
| HDL-C (GLGC) | 15 | 58674051 | G | A |
| HDL-C (GLGC) | 15 | 58674308 | G | A |
| HDL-C (GLGC) | 15 | 58676119 | G | A |
| HDL-C (GLGC) | 15 | 58678512 | T | C |
| HDL-C (GLGC) | 15 | 58678869 | T | C |
| HDL-C (GLGC) | 15 | 58680638 | T | C |
| HDL-C (GLGC) | 15 | 58680805 | G | A |
| HDL-C (GLGC) | 15 | 58680954 | T | C |
| HDL-C (GLGC) | 15 | 58682690 | G | A |
| HDL-C (GLGC) | 15 | 58682783 | C | A |
| HDL-C (GLGC) | 15 | 58683366 | G | A |
| HDL-C (GLGC) | 15 | 58683410 | T | C |
| HDL-C (GLGC) | 15 | 58683585 | G | A |
| HDL-C (GLGC) | 15 | 58684381 | G | A |
| HDL-C (GLGC) | 15 | 58684478 | T | C |
| HDL-C (GLGC) | 15 | 58685560 | T | C |
| HDL-C (GLGC) | 15 | 58685970 | T | C |
| HDL-C (GLGC) | 15 | 58686338 | G | A |
| HDL-C (GLGC) | 15 | 58686754 | T | C |
| HDL-C (GLGC) | 15 | 58686809 | T | C |
| HDL-C (GLGC) | 15 | 58686939 | T | C |
| HDL-C (GLGC) | 15 | 58687040 | T | C |
| HDL-C (GLGC) | 15 | 58687609 | T | G |
| HDL-C (GLGC) | 15 | 58687880 | T | C |
| HDL-C (GLGC) | 15 | 58688186 | C | A |
| HDL-C (GLGC) | 15 | 58690754 | G | A |
| HDL-C (GLGC) | 15 | 58691208 | T | C |
| HDL-C (GLGC) | 15 | 58692007 | G | A |

**Table S14:** Genetic variants used in the presented analyses. Note that due to coverage of the GWAS chip the instruments differed across considered outcomes *(continued)*

| Exposure | Chromosome | Base-pair position (Build 37) | Other allele | Effect allele |
|---|---|---|---|---|
| HDL-C (GLGC) | 15 | 58692202 | C | A |
| HDL-C (GLGC) | 15 | 58692936 | T | C |
| HDL-C (GLGC) | 15 | 58694020 | T | G |
| HDL-C (GLGC) | 15 | 58695592 | G | A |
| HDL-C (GLGC) | 15 | 58697072 | T | C |
| HDL-C (GLGC) | 15 | 58699010 | T | C |
| HDL-C (GLGC) | 15 | 58699937 | T | C |
| HDL-C (GLGC) | 15 | 58703069 | C | A |
| HDL-C (GLGC) | 15 | 58703895 | G | A |
| HDL-C (GLGC) | 15 | 58710627 | C | A |
| HDL-C (GLGC) | 15 | 58711654 | T | C |
| HDL-C (GLGC) | 15 | 58712203 | T | G |
| HDL-C (GLGC) | 15 | 58712973 | T | C |
| HDL-C (GLGC) | 15 | 58716044 | G | A |
| HDL-C (GLGC) | 15 | 58716615 | T | G |
| HDL-C (GLGC) | 15 | 58717428 | T | C |
| HDL-C (GLGC) | 15 | 58718998 | T | C |
| HDL-C (GLGC) | 15 | 58719741 | G | A |
| HDL-C (GLGC) | 15 | 58720431 | C | A |
| HDL-C (GLGC) | 15 | 58721447 | G | A |
| HDL-C (GLGC) | 15 | 58721614 | G | A |
| HDL-C (GLGC) | 15 | 58723479 | T | C |
| HDL-C (GLGC) | 15 | 58723675 | T | C |
| HDL-C (GLGC) | 15 | 58723939 | G | A |
| HDL-C (GLGC) | 15 | 58724524 | G | A |
| HDL-C (GLGC) | 15 | 58724706 | G | A |
| HDL-C (GLGC) | 15 | 58727279 | T | C |
| HDL-C (GLGC) | 15 | 58727325 | G | A |
| HDL-C (GLGC) | 15 | 58729144 | T | C |
| HDL-C (GLGC) | 15 | 58731455 | T | C |
| HDL-C (GLGC) | 15 | 58731567 | G | A |
| HDL-C (GLGC) | 15 | 58734645 | T | C |
| HDL-C (GLGC) | 15 | 58738760 | G | A |
| HDL-C (GLGC) | 15 | 58740094 | T | C |
| HDL-C (GLGC) | 15 | 58742731 | T | G |
| HDL-C (GLGC) | 15 | 58743709 | T | C |
| HDL-C (GLGC) | 15 | 58751877 | T | G |
| HDL-C (GLGC) | 15 | 58853109 | G | A |
| HDL-C (GLGC) | 15 | 59010962 | T | C |
| HDL-C (GLGC) | 15 | 63348087 | G | A |
| HDL-C (GLGC) | 15 | 63366282 | G | A |
| HDL-C (GLGC) | 15 | 63367032 | T | C |
| HDL-C (GLGC) | 15 | 63379643 | T | G |
| HDL-C (GLGC) | 15 | 63396867 | G | A |

**Table S14:** Genetic variants used in the presented analyses. Note that due to coverage of the GWAS chip the instruments differed across considered outcomes *(continued)*

| Exposure | Chromosome | Base-pair position (Build 37) | Other allele | Effect allele |
|---|---|---|---|---|
| HDL-C (GLGC) | 15 | 63400584 | G | A |
| HDL-C (GLGC) | 15 | 63402574 | G | A |
| HDL-C (GLGC) | 15 | 63413084 | T | C |
| HDL-C (GLGC) | 15 | 63425337 | G | A |
| HDL-C (GLGC) | 15 | 63426912 | T | G |
| HDL-C (GLGC) | 15 | 63433766 | G | A |
| HDL-C (GLGC) | 15 | 63434763 | G | A |
| HDL-C (GLGC) | 16 | 53800954 | T | C |
| HDL-C (GLGC) | 16 | 53804340 | T | C |
| HDL-C (GLGC) | 16 | 53812614 | G | A |
| HDL-C (GLGC) | 16 | 53815161 | T | G |
| HDL-C (GLGC) | 16 | 53818708 | T | G |
| HDL-C (GLGC) | 16 | 53828752 | G | A |
| HDL-C (GLGC) | 16 | 53830465 | G | A |
| HDL-C (GLGC) | 16 | 56387000 | T | C |
| HDL-C (GLGC) | 16 | 56490549 | G | A |
| HDL-C (GLGC) | 16 | 56626769 | G | A |
| HDL-C (GLGC) | 16 | 56671140 | T | C |
| HDL-C (GLGC) | 16 | 56690858 | T | C |
| HDL-C (GLGC) | 16 | 56722572 | T | C |
| HDL-C (GLGC) | 16 | 56735782 | G | A |
| HDL-C (GLGC) | 16 | 56742746 | T | C |
| HDL-C (GLGC) | 16 | 56765226 | T | C |
| HDL-C (GLGC) | 16 | 56793346 | C | A |
| HDL-C (GLGC) | 16 | 56811692 | T | C |
| HDL-C (GLGC) | 16 | 56815620 | G | A |
| HDL-C (GLGC) | 16 | 56824183 | T | G |
| HDL-C (GLGC) | 16 | 56846804 | T | C |
| HDL-C (GLGC) | 16 | 56868700 | G | A |
| HDL-C (GLGC) | 16 | 56873789 | T | C |
| HDL-C (GLGC) | 16 | 56873840 | G | A |
| HDL-C (GLGC) | 16 | 56880842 | T | C |
| HDL-C (GLGC) | 16 | 56883924 | G | A |
| HDL-C (GLGC) | 16 | 56900931 | T | C |
| HDL-C (GLGC) | 16 | 56905432 | T | C |
| HDL-C (GLGC) | 16 | 56912903 | T | C |
| HDL-C (GLGC) | 16 | 56926195 | G | A |
| HDL-C (GLGC) | 16 | 56928216 | T | C |
| HDL-C (GLGC) | 16 | 56933519 | G | A |
| HDL-C (GLGC) | 16 | 56936563 | G | A |
| HDL-C (GLGC) | 16 | 56938016 | G | A |
| HDL-C (GLGC) | 16 | 56938522 | T | C |
| HDL-C (GLGC) | 16 | 56940128 | T | C |
| HDL-C (GLGC) | 16 | 56942709 | G | A |

**Table S14:** Genetic variants used in the presented analyses. Note that due to coverage of the GWAS chip the instruments differed across considered outcomes *(continued)*

| Exposure | Chromosome | Base-pair position (Build 37) | Other allele | Effect allele |
|---|---|---|---|---|
| HDL-C (GLGC) | 16 | 56945539 | G | A |
| HDL-C (GLGC) | 16 | 56945619 | T | G |
| HDL-C (GLGC) | 16 | 56948618 | C | A |
| HDL-C (GLGC) | 16 | 56964719 | G | A |
| HDL-C (GLGC) | 16 | 56965707 | G | A |
| HDL-C (GLGC) | 16 | 56968039 | G | A |
| HDL-C (GLGC) | 16 | 56969148 | G | A |
| HDL-C (GLGC) | 16 | 56984482 | T | C |
| HDL-C (GLGC) | 16 | 56984590 | T | C |
| HDL-C (GLGC) | 16 | 56984763 | G | A |
| HDL-C (GLGC) | 16 | 56985139 | G | A |
| HDL-C (GLGC) | 16 | 56985156 | G | A |
| HDL-C (GLGC) | 16 | 56985514 | G | A |
| HDL-C (GLGC) | 16 | 56985555 | G | A |
| HDL-C (GLGC) | 16 | 56987015 | T | C |
| HDL-C (GLGC) | 16 | 56987369 | T | C |
| HDL-C (GLGC) | 16 | 56988044 | T | C |
| HDL-C (GLGC) | 16 | 56989590 | T | C |
| HDL-C (GLGC) | 16 | 56990716 | C | A |
| HDL-C (GLGC) | 16 | 56991363 | T | C |
| HDL-C (GLGC) | 16 | 56991524 | G | A |
| HDL-C (GLGC) | 16 | 56992017 | G | A |
| HDL-C (GLGC) | 16 | 56993025 | T | G |
| HDL-C (GLGC) | 16 | 56993161 | G | A |
| HDL-C (GLGC) | 16 | 56993211 | T | C |
| HDL-C (GLGC) | 16 | 56993324 | C | A |
| HDL-C (GLGC) | 16 | 56993909 | C | A |
| HDL-C (GLGC) | 16 | 56993935 | T | C |
| HDL-C (GLGC) | 16 | 56994192 | T | C |
| HDL-C (GLGC) | 16 | 56994528 | T | C |
| HDL-C (GLGC) | 16 | 56994894 | G | A |
| HDL-C (GLGC) | 16 | 56995236 | C | A |
| HDL-C (GLGC) | 16 | 56996158 | T | C |
| HDL-C (GLGC) | 16 | 56997233 | G | A |
| HDL-C (GLGC) | 16 | 56999258 | C | A |
| HDL-C (GLGC) | 16 | 56999328 | T | C |
| HDL-C (GLGC) | 16 | 57000284 | G | A |
| HDL-C (GLGC) | 16 | 57000885 | G | A |
| HDL-C (GLGC) | 16 | 57000938 | G | A |
| HDL-C (GLGC) | 16 | 57002404 | C | A |
| HDL-C (GLGC) | 16 | 57002732 | T | G |
| HDL-C (GLGC) | 16 | 57003146 | T | C |
| HDL-C (GLGC) | 16 | 57004889 | G | A |
| HDL-C (GLGC) | 16 | 57005479 | C | A |

**Table S14:** Genetic variants used in the presented analyses. Note that due to coverage of the GWAS chip the instruments differed across considered outcomes *(continued)*

| Exposure | Chromosome | Base-pair position (Build 37) | Other allele | Effect allele |
|---|---|---|---|---|
| HDL-C (GLGC) | 16 | 57005762 | T | C |
| HDL-C (GLGC) | 16 | 57006072 | G | A |
| HDL-C (GLGC) | 16 | 57006590 | T | C |
| HDL-C (GLGC) | 16 | 57007192 | T | C |
| HDL-C (GLGC) | 16 | 57007451 | G | A |
| HDL-C (GLGC) | 16 | 57011185 | G | A |
| HDL-C (GLGC) | 16 | 57014875 | G | A |
| HDL-C (GLGC) | 16 | 57016092 | G | A |
| HDL-C (GLGC) | 16 | 57017002 | T | G |
| HDL-C (GLGC) | 16 | 57017662 | G | A |
| HDL-C (GLGC) | 16 | 57019392 | T | C |
| HDL-C (GLGC) | 16 | 57019532 | C | A |
| HDL-C (GLGC) | 16 | 57021433 | G | A |
| HDL-C (GLGC) | 16 | 57024662 | T | C |
| HDL-C (GLGC) | 16 | 57025063 | G | A |
| HDL-C (GLGC) | 16 | 57026775 | G | A |
| HDL-C (GLGC) | 16 | 57031811 | G | A |
| HDL-C (GLGC) | 16 | 57032461 | T | G |
| HDL-C (GLGC) | 16 | 57034115 | T | C |
| HDL-C (GLGC) | 16 | 57034645 | T | C |
| HDL-C (GLGC) | 16 | 57042912 | G | A |
| HDL-C (GLGC) | 16 | 57048118 | T | C |
| HDL-C (GLGC) | 16 | 57048707 | G | A |
| HDL-C (GLGC) | 16 | 57050348 | T | G |
| HDL-C (GLGC) | 16 | 57052509 | C | A |
| HDL-C (GLGC) | 16 | 57052901 | T | G |
| HDL-C (GLGC) | 16 | 57060724 | G | A |
| HDL-C (GLGC) | 16 | 57061189 | T | C |
| HDL-C (GLGC) | 16 | 57065121 | C | A |
| HDL-C (GLGC) | 16 | 57065556 | T | C |
| HDL-C (GLGC) | 16 | 57072004 | G | A |
| HDL-C (GLGC) | 16 | 57074451 | T | C |
| HDL-C (GLGC) | 16 | 57075253 | C | A |
| HDL-C (GLGC) | 16 | 57080528 | C | A |
| HDL-C (GLGC) | 16 | 57083232 | G | A |
| HDL-C (GLGC) | 16 | 57116036 | C | A |
| HDL-C (GLGC) | 16 | 57141666 | C | A |
| HDL-C (GLGC) | 16 | 57154547 | C | A |
| HDL-C (GLGC) | 16 | 57353934 | G | A |
| HDL-C (GLGC) | 16 | 67081372 | G | A |
| HDL-C (GLGC) | 16 | 67144295 | T | C |
| HDL-C (GLGC) | 16 | 67144367 | G | A |
| HDL-C (GLGC) | 16 | 67187211 | T | G |
| HDL-C (GLGC) | 16 | 67207933 | T | C |

**Table S14:** Genetic variants used in the presented analyses. Note that due to coverage of the GWAS chip the instruments differed across considered outcomes *(continued)*

| Exposure | Chromosome | Base-pair position (Build 37) | Other allele | Effect allele |
|---|---|---|---|---|
| HDL-C (GLGC) | 16 | 67241282 | T | C |
| HDL-C (GLGC) | 16 | 67305513 | C | A |
| HDL-C (GLGC) | 16 | 67358816 | G | A |
| HDL-C (GLGC) | 16 | 67390448 | G | A |
| HDL-C (GLGC) | 16 | 67398369 | T | C |
| HDL-C (GLGC) | 16 | 67419152 | G | A |
| HDL-C (GLGC) | 16 | 67555339 | C | A |
| HDL-C (GLGC) | 16 | 67605794 | T | C |
| HDL-C (GLGC) | 16 | 67627635 | T | G |
| HDL-C (GLGC) | 16 | 67635751 | T | C |
| HDL-C (GLGC) | 16 | 67642935 | T | C |
| HDL-C (GLGC) | 16 | 67651879 | G | A |
| HDL-C (GLGC) | 16 | 67671804 | T | G |
| HDL-C (GLGC) | 16 | 67677001 | T | C |
| HDL-C (GLGC) | 16 | 67682234 | G | A |
| HDL-C (GLGC) | 16 | 67699948 | T | C |
| HDL-C (GLGC) | 16 | 67708130 | T | C |
| HDL-C (GLGC) | 16 | 67708897 | G | A |
| HDL-C (GLGC) | 16 | 67758778 | T | C |
| HDL-C (GLGC) | 16 | 67796315 | C | A |
| HDL-C (GLGC) | 16 | 67801391 | G | A |
| HDL-C (GLGC) | 16 | 67827247 | G | A |
| HDL-C (GLGC) | 16 | 67833171 | T | C |
| HDL-C (GLGC) | 16 | 67863451 | T | C |
| HDL-C (GLGC) | 16 | 67898492 | G | A |
| HDL-C (GLGC) | 16 | 67902070 | G | A |
| HDL-C (GLGC) | 16 | 67925435 | G | A |
| HDL-C (GLGC) | 16 | 68012849 | T | C |
| HDL-C (GLGC) | 16 | 68013471 | T | C |
| HDL-C (GLGC) | 16 | 68021125 | C | A |
| HDL-C (GLGC) | 16 | 68024995 | G | A |
| HDL-C (GLGC) | 16 | 68057880 | G | A |
| HDL-C (GLGC) | 16 | 68068627 | G | A |
| HDL-C (GLGC) | 16 | 68074857 | G | A |
| HDL-C (GLGC) | 16 | 68104422 | T | C |
| HDL-C (GLGC) | 16 | 68106846 | T | C |
| HDL-C (GLGC) | 16 | 68128104 | G | A |
| HDL-C (GLGC) | 16 | 68181177 | C | A |
| HDL-C (GLGC) | 16 | 68190239 | C | A |
| HDL-C (GLGC) | 16 | 68206844 | G | A |
| HDL-C (GLGC) | 16 | 68218336 | G | A |
| HDL-C (GLGC) | 16 | 68225515 | G | A |
| HDL-C (GLGC) | 16 | 68270229 | T | C |
| HDL-C (GLGC) | 16 | 68280893 | T | C |

**Table S14:** Genetic variants used in the presented analyses. Note that due to coverage of the GWAS chip the instruments differed across considered outcomes *(continued)*

| Exposure | Chromosome | Base-pair position (Build 37) | Other allele | Effect allele |
|---|---|---|---|---|
| HDL-C (GLGC) | 16 | 68286427 | C | A |
| HDL-C (GLGC) | 16 | 68289313 | C | A |
| HDL-C (GLGC) | 16 | 68290200 | G | A |
| HDL-C (GLGC) | 16 | 68295598 | G | A |
| HDL-C (GLGC) | 16 | 68297228 | G | A |
| HDL-C (GLGC) | 16 | 68299771 | G | A |
| HDL-C (GLGC) | 16 | 68301410 | G | A |
| HDL-C (GLGC) | 16 | 68307447 | T | C |
| HDL-C (GLGC) | 16 | 68310312 | T | C |
| HDL-C (GLGC) | 16 | 68321878 | C | A |
| HDL-C (GLGC) | 16 | 68323654 | G | A |
| HDL-C (GLGC) | 16 | 68344363 | C | A |
| HDL-C (GLGC) | 16 | 68369213 | T | C |
| HDL-C (GLGC) | 16 | 68393405 | T | C |
| HDL-C (GLGC) | 16 | 68409435 | C | A |
| HDL-C (GLGC) | 16 | 68423506 | T | C |
| HDL-C (GLGC) | 16 | 68428326 | G | A |
| HDL-C (GLGC) | 16 | 81519766 | G | A |
| HDL-C (GLGC) | 16 | 81524274 | C | A |
| HDL-C (GLGC) | 16 | 81534790 | T | C |
| HDL-C (GLGC) | 16 | 81549708 | T | C |
| HDL-C (GLGC) | 17 | 37389409 | G | A |
| HDL-C (GLGC) | 17 | 37409801 | G | A |
| HDL-C (GLGC) | 17 | 37425238 | T | G |
| HDL-C (GLGC) | 17 | 37430519 | T | C |
| HDL-C (GLGC) | 17 | 37447218 | T | C |
| HDL-C (GLGC) | 17 | 37499949 | T | C |
| HDL-C (GLGC) | 17 | 37561613 | G | A |
| HDL-C (GLGC) | 17 | 37573065 | C | A |
| HDL-C (GLGC) | 17 | 37609120 | G | A |
| HDL-C (GLGC) | 17 | 37631924 | G | A |
| HDL-C (GLGC) | 17 | 37682657 | G | A |
| HDL-C (GLGC) | 17 | 37709782 | G | A |
| HDL-C (GLGC) | 17 | 37715551 | T | G |
| HDL-C (GLGC) | 17 | 37722515 | G | A |
| HDL-C (GLGC) | 17 | 37727950 | T | C |
| HDL-C (GLGC) | 17 | 37745979 | G | A |
| HDL-C (GLGC) | 17 | 37767453 | G | A |
| HDL-C (GLGC) | 17 | 37770005 | T | C |
| HDL-C (GLGC) | 17 | 37781849 | G | A |
| HDL-C (GLGC) | 17 | 37784990 | G | A |
| HDL-C (GLGC) | 17 | 37790939 | T | C |
| HDL-C (GLGC) | 17 | 37807698 | C | A |
| HDL-C (GLGC) | 17 | 37814080 | G | A |

**Table S14:** Genetic variants used in the presented analyses. Note that due to coverage of the GWAS chip the instruments differed across considered outcomes *(continued)*

| Exposure | Chromosome | Base-pair position (Build 37) | Other allele | Effect allele |
|---|---|---|---|---|
| HDL-C (GLGC) | 17 | 37824545 | G | A |
| HDL-C (GLGC) | 17 | 37828745 | G | A |
| HDL-C (GLGC) | 17 | 37831613 | G | A |
| HDL-C (GLGC) | 17 | 37832279 | T | C |
| HDL-C (GLGC) | 17 | 37834357 | C | A |
| HDL-C (GLGC) | 17 | 37836581 | G | A |
| HDL-C (GLGC) | 17 | 37858678 | G | A |
| HDL-C (GLGC) | 17 | 37863708 | T | C |
| HDL-C (GLGC) | 17 | 37866005 | T | C |
| HDL-C (GLGC) | 17 | 37922259 | G | A |
| HDL-C (GLGC) | 17 | 37949924 | G | A |
| HDL-C (GLGC) | 17 | 37970149 | G | A |
| HDL-C (GLGC) | 17 | 37985801 | G | A |
| HDL-C (GLGC) | 17 | 38035116 | G | A |
| HDL-C (GLGC) | 17 | 38066240 | T | C |
| HDL-C (GLGC) | 17 | 38069949 | T | C |
| HDL-C (GLGC) | 17 | 38080865 | G | A |
| HDL-C (GLGC) | 17 | 38121993 | G | A |
| HDL-C (GLGC) | 17 | 38122680 | T | G |
| HDL-C (GLGC) | 17 | 41930569 | G | A |
| HDL-C (GLGC) | 17 | 41978756 | T | G |
| HDL-C (GLGC) | 17 | 45010721 | G | A |
| HDL-C (GLGC) | 17 | 66833954 | G | A |
| HDL-C (GLGC) | 17 | 66857775 | G | A |
| HDL-C (GLGC) | 17 | 66869321 | G | A |
| HDL-C (GLGC) | 17 | 66895188 | T | C |
| HDL-C (GLGC) | 17 | 66895479 | T | G |
| HDL-C (GLGC) | 17 | 66898257 | C | A |
| HDL-C (GLGC) | 17 | 66899290 | G | A |
| HDL-C (GLGC) | 17 | 66901215 | T | G |
| HDL-C (GLGC) | 17 | 66903293 | G | A |
| HDL-C (GLGC) | 17 | 76391454 | T | G |
| HDL-C (GLGC) | 17 | 76392144 | G | A |
| HDL-C (GLGC) | 17 | 76395430 | T | C |
| HDL-C (GLGC) | 17 | 76403984 | G | A |
| HDL-C (GLGC) | 17 | 76409809 | G | A |
| HDL-C (GLGC) | 18 | 46972438 | G | A |
| HDL-C (GLGC) | 18 | 47029576 | G | A |
| HDL-C (GLGC) | 18 | 47033207 | T | C |
| HDL-C (GLGC) | 18 | 47045445 | C | A |
| HDL-C (GLGC) | 18 | 47049032 | G | A |
| HDL-C (GLGC) | 18 | 47062054 | T | G |
| HDL-C (GLGC) | 18 | 47074869 | G | A |
| HDL-C (GLGC) | 18 | 47075308 | G | A |

**Table S14:** Genetic variants used in the presented analyses. Note that due to coverage of the GWAS chip the instruments differed across considered outcomes *(continued)*

| Exposure | Chromosome | Base-pair position (Build 37) | Other allele | Effect allele |
|---|---|---|---|---|
| HDL-C (GLGC) | 18 | 47076063 | C | A |
| HDL-C (GLGC) | 18 | 47086932 | T | C |
| HDL-C (GLGC) | 18 | 47093790 | T | C |
| HDL-C (GLGC) | 18 | 47093864 | T | C |
| HDL-C (GLGC) | 18 | 47097398 | G | A |
| HDL-C (GLGC) | 18 | 47099186 | G | A |
| HDL-C (GLGC) | 18 | 47102102 | G | A |
| HDL-C (GLGC) | 18 | 47118398 | T | C |
| HDL-C (GLGC) | 18 | 47123763 | T | C |
| HDL-C (GLGC) | 18 | 47126133 | T | C |
| HDL-C (GLGC) | 18 | 47128779 | T | C |
| HDL-C (GLGC) | 18 | 47131684 | T | C |
| HDL-C (GLGC) | 18 | 47131888 | C | A |
| HDL-C (GLGC) | 18 | 47132464 | T | C |
| HDL-C (GLGC) | 18 | 47133828 | G | A |
| HDL-C (GLGC) | 18 | 47135834 | T | C |
| HDL-C (GLGC) | 18 | 47137027 | T | C |
| HDL-C (GLGC) | 18 | 47142243 | G | A |
| HDL-C (GLGC) | 18 | 47145202 | G | A |
| HDL-C (GLGC) | 18 | 47145717 | G | A |
| HDL-C (GLGC) | 18 | 47146125 | G | A |
| HDL-C (GLGC) | 18 | 47146166 | T | C |
| HDL-C (GLGC) | 18 | 47147746 | G | A |
| HDL-C (GLGC) | 18 | 47148299 | T | C |
| HDL-C (GLGC) | 18 | 47148886 | T | C |
| HDL-C (GLGC) | 18 | 47149023 | T | C |
| HDL-C (GLGC) | 18 | 47149371 | G | A |
| HDL-C (GLGC) | 18 | 47151264 | G | A |
| HDL-C (GLGC) | 18 | 47153264 | C | A |
| HDL-C (GLGC) | 18 | 47154779 | T | C |
| HDL-C (GLGC) | 18 | 47156433 | T | C |
| HDL-C (GLGC) | 18 | 47156730 | G | A |
| HDL-C (GLGC) | 18 | 47157439 | T | C |
| HDL-C (GLGC) | 18 | 47158186 | T | C |
| HDL-C (GLGC) | 18 | 47159090 | G | A |
| HDL-C (GLGC) | 18 | 47159857 | G | A |
| HDL-C (GLGC) | 18 | 47160814 | G | A |
| HDL-C (GLGC) | 18 | 47160953 | T | G |
| HDL-C (GLGC) | 18 | 47161225 | T | C |
| HDL-C (GLGC) | 18 | 47161628 | G | A |
| HDL-C (GLGC) | 18 | 47161733 | T | C |
| HDL-C (GLGC) | 18 | 47162207 | G | A |
| HDL-C (GLGC) | 18 | 47164926 | T | C |
| HDL-C (GLGC) | 18 | 47166694 | G | A |

**Table S14:** Genetic variants used in the presented analyses. Note that due to coverage of the GWAS chip the instruments differed across considered outcomes *(continued)*

| Exposure | Chromosome | Base-pair position (Build 37) | Other allele | Effect allele |
|---|---|---|---|---|
| HDL-C (GLGC) | 18 | 47167506 | T | G |
| HDL-C (GLGC) | 18 | 47169333 | G | A |
| HDL-C (GLGC) | 18 | 47172730 | T | C |
| HDL-C (GLGC) | 18 | 47175467 | T | C |
| HDL-C (GLGC) | 18 | 47175670 | C | A |
| HDL-C (GLGC) | 18 | 47176261 | C | A |
| HDL-C (GLGC) | 18 | 47176911 | G | A |
| HDL-C (GLGC) | 18 | 47176917 | T | C |
| HDL-C (GLGC) | 18 | 47177608 | C | A |
| HDL-C (GLGC) | 18 | 47179516 | G | A |
| HDL-C (GLGC) | 18 | 47181568 | G | A |
| HDL-C (GLGC) | 18 | 47182569 | G | A |
| HDL-C (GLGC) | 18 | 47182838 | G | A |
| HDL-C (GLGC) | 18 | 47183794 | G | A |
| HDL-C (GLGC) | 18 | 47184684 | G | A |
| HDL-C (GLGC) | 18 | 47189138 | G | A |
| HDL-C (GLGC) | 18 | 47192294 | C | A |
| HDL-C (GLGC) | 18 | 47203955 | G | A |
| HDL-C (GLGC) | 18 | 47205432 | G | A |
| HDL-C (GLGC) | 18 | 47206583 | T | C |
| HDL-C (GLGC) | 18 | 47209971 | G | A |
| HDL-C (GLGC) | 18 | 47213682 | G | A |
| HDL-C (GLGC) | 18 | 47243912 | C | A |
| HDL-C (GLGC) | 18 | 47249297 | G | A |
| HDL-C (GLGC) | 18 | 47261744 | C | A |
| HDL-C (GLGC) | 18 | 47266045 | C | A |
| HDL-C (GLGC) | 18 | 47275339 | T | C |
| HDL-C (GLGC) | 18 | 47278147 | T | C |
| HDL-C (GLGC) | 18 | 47280410 | T | C |
| HDL-C (GLGC) | 18 | 47288632 | C | A |
| HDL-C (GLGC) | 18 | 47295988 | T | G |
| HDL-C (GLGC) | 18 | 47385799 | G | A |
| HDL-C (GLGC) | 18 | 47394395 | C | A |
| HDL-C (GLGC) | 18 | 47394538 | T | C |
| HDL-C (GLGC) | 18 | 47400116 | T | C |
| HDL-C (GLGC) | 18 | 47404743 | T | C |
| HDL-C (GLGC) | 18 | 47430194 | T | C |
| HDL-C (GLGC) | 18 | 47457559 | T | C |
| HDL-C (GLGC) | 18 | 47464423 | T | C |
| HDL-C (GLGC) | 18 | 47482530 | T | C |
| HDL-C (GLGC) | 18 | 57739179 | T | G |
| HDL-C (GLGC) | 18 | 57787559 | T | C |
| HDL-C (GLGC) | 18 | 57811982 | G | A |
| HDL-C (GLGC) | 18 | 57851097 | T | C |

**Table S14:** Genetic variants used in the presented analyses. Note that due to coverage of the GWAS chip the instruments differed across considered outcomes *(continued)*

| Exposure | Chromosome | Base-pair position (Build 37) | Other allele | Effect allele |
|---|---|---|---|---|
| HDL-C (GLGC) | 18 | 57897803 | T | C |
| HDL-C (GLGC) | 18 | 57922526 | C | A |
| HDL-C (GLGC) | 19 | 11334179 | T | C |
| HDL-C (GLGC) | 19 | 11346550 | T | C |
| HDL-C (GLGC) | 19 | 11347493 | T | C |
| HDL-C (GLGC) | 19 | 11347657 | T | C |
| HDL-C (GLGC) | 19 | 33899065 | G | A |
| HDL-C (GLGC) | 19 | 33906123 | T | C |
| HDL-C (GLGC) | 19 | 33909710 | T | G |
| HDL-C (GLGC) | 19 | 45242173 | G | A |
| HDL-C (GLGC) | 19 | 45247627 | G | A |
| HDL-C (GLGC) | 19 | 45329214 | T | G |
| HDL-C (GLGC) | 19 | 45333834 | T | C |
| HDL-C (GLGC) | 19 | 45392254 | T | C |
| HDL-C (GLGC) | 19 | 45395266 | G | A |
| HDL-C (GLGC) | 19 | 45395619 | G | A |
| HDL-C (GLGC) | 19 | 45403216 | T | C |
| HDL-C (GLGC) | 19 | 45410002 | G | A |
| HDL-C (GLGC) | 19 | 45412079 | T | C |
| HDL-C (GLGC) | 19 | 45414451 | T | C |
| HDL-C (GLGC) | 19 | 45415640 | G | A |
| HDL-C (GLGC) | 19 | 45422846 | G | A |
| HDL-C (GLGC) | 19 | 45422946 | G | A |
| HDL-C (GLGC) | 19 | 45443504 | G | A |
| HDL-C (GLGC) | 19 | 45448036 | T | C |
| HDL-C (GLGC) | 19 | 45448465 | T | G |
| HDL-C (GLGC) | 19 | 45453144 | T | C |
| HDL-C (GLGC) | 19 | 45453239 | G | A |
| HDL-C (GLGC) | 19 | 45458146 | C | A |
| HDL-C (GLGC) | 19 | 47589895 | T | C |
| HDL-C (GLGC) | 19 | 52323088 | G | A |
| HDL-C (GLGC) | 19 | 52324216 | G | A |
| HDL-C (GLGC) | 19 | 52327784 | T | C |
| HDL-C (GLGC) | 19 | 52550401 | T | G |
| HDL-C (GLGC) | 19 | 54762408 | T | C |
| HDL-C (GLGC) | 19 | 54779101 | T | C |
| HDL-C (GLGC) | 19 | 54779455 | T | C |
| HDL-C (GLGC) | 19 | 54784130 | T | C |
| HDL-C (GLGC) | 19 | 54785593 | G | A |
| HDL-C (GLGC) | 19 | 54793048 | C | A |
| HDL-C (GLGC) | 19 | 54793188 | G | A |
| HDL-C (GLGC) | 19 | 54795299 | T | C |
| HDL-C (GLGC) | 19 | 54796630 | G | A |
| HDL-C (GLGC) | 19 | 54800225 | G | A |

**Table S14:** Genetic variants used in the presented analyses. Note that due to coverage of the GWAS chip the instruments differed across considered outcomes *(continued)*

| Exposure | Chromosome | Base-pair position (Build 37) | Other allele | Effect allele |
|---|---|---|---|---|
| HDL-C (GLGC) | 19 | 54800371 | T | C |
| HDL-C (GLGC) | 19 | 54810370 | G | A |
| HDL-C (GLGC) | 19 | 54813762 | T | C |
| HDL-C (GLGC) | 19 | 54827040 | C | A |
| HDL-C (GLGC) | 19 | 54827480 | T | G |
| HDL-C (GLGC) | 19 | 54835771 | T | C |
| HDL-C (GLGC) | 19 | 54837165 | G | A |
| HDL-C (GLGC) | 19 | 7970635 | G | A |
| HDL-C (GLGC) | 19 | 8433196 | C | A |
| HDL-C (GLGC) | 19 | 8441779 | T | C |
| HDL-C (GLGC) | 19 | 8453822 | T | C |
| HDL-C (GLGC) | 19 | 8468337 | T | C |
| HDL-C (GLGC) | 19 | 8469738 | T | C |
| HDL-C (GLGC) | 19 | 8503025 | G | A |
| HDL-C (GLGC) | 19 | 8586086 | C | A |
| HDL-C (GLGC) | 19 | 8610313 | G | A |
| HDL-C (GLGC) | 19 | 8615589 | G | A |
| HDL-C (GLGC) | 1 | 109814880 | T | C |
| HDL-C (GLGC) | 1 | 109815252 | G | A |
| HDL-C (GLGC) | 1 | 109817192 | G | A |
| HDL-C (GLGC) | 1 | 109818306 | T | G |
| HDL-C (GLGC) | 1 | 109821307 | T | G |
| HDL-C (GLGC) | 1 | 109822166 | G | A |
| HDL-C (GLGC) | 1 | 110470764 | G | A |
| HDL-C (GLGC) | 1 | 110478911 | G | A |
| HDL-C (GLGC) | 1 | 110499669 | G | A |
| HDL-C (GLGC) | 1 | 110500175 | T | C |
| HDL-C (GLGC) | 1 | 156700651 | T | G |
| HDL-C (GLGC) | 1 | 178515312 | G | A |
| HDL-C (GLGC) | 1 | 182054977 | T | C |
| HDL-C (GLGC) | 1 | 182058556 | G | A |
| HDL-C (GLGC) | 1 | 182117324 | G | A |
| HDL-C (GLGC) | 1 | 182119332 | G | A |
| HDL-C (GLGC) | 1 | 182121598 | G | A |
| HDL-C (GLGC) | 1 | 182161829 | T | C |
| HDL-C (GLGC) | 1 | 182168885 | G | A |
| HDL-C (GLGC) | 1 | 182200659 | T | C |
| HDL-C (GLGC) | 1 | 220970028 | G | A |
| HDL-C (GLGC) | 1 | 220970593 | T | G |
| HDL-C (GLGC) | 1 | 230278826 | G | A |
| HDL-C (GLGC) | 1 | 230281068 | G | A |
| HDL-C (GLGC) | 1 | 230281698 | G | A |
| HDL-C (GLGC) | 1 | 230283764 | G | A |
| HDL-C (GLGC) | 1 | 230285056 | G | A |

**Table S14:** Genetic variants used in the presented analyses. Note that due to coverage of the GWAS chip the instruments differed across considered outcomes *(continued)*

| Exposure | Chromosome | Base-pair position (Build 37) | Other allele | Effect allele |
|---|---|---|---|---|
| HDL-C (GLGC) | 1 | 230287128 | T | G |
| HDL-C (GLGC) | 1 | 230290254 | G | A |
| HDL-C (GLGC) | 1 | 230292499 | T | C |
| HDL-C (GLGC) | 1 | 230295691 | G | A |
| HDL-C (GLGC) | 1 | 230300586 | T | C |
| HDL-C (GLGC) | 1 | 230301811 | T | G |
| HDL-C (GLGC) | 1 | 230304988 | G | A |
| HDL-C (GLGC) | 1 | 230307222 | T | G |
| HDL-C (GLGC) | 1 | 230309619 | G | A |
| HDL-C (GLGC) | 1 | 230321019 | T | C |
| HDL-C (GLGC) | 1 | 230324067 | T | G |
| HDL-C (GLGC) | 1 | 230324364 | T | C |
| HDL-C (GLGC) | 1 | 230324801 | G | A |
| HDL-C (GLGC) | 1 | 230389838 | T | G |
| HDL-C (GLGC) | 1 | 230398102 | G | A |
| HDL-C (GLGC) | 1 | 230419344 | G | A |
| HDL-C (GLGC) | 1 | 230427840 | G | A |
| HDL-C (GLGC) | 1 | 26900708 | G | A |
| HDL-C (GLGC) | 1 | 26902388 | T | G |
| HDL-C (GLGC) | 1 | 27013133 | G | A |
| HDL-C (GLGC) | 1 | 27138393 | T | C |
| HDL-C (GLGC) | 1 | 27230033 | G | A |
| HDL-C (GLGC) | 1 | 27236212 | T | C |
| HDL-C (GLGC) | 1 | 27248645 | G | A |
| HDL-C (GLGC) | 1 | 39550621 | T | C |
| HDL-C (GLGC) | 1 | 39569571 | T | C |
| HDL-C (GLGC) | 1 | 39583686 | T | C |
| HDL-C (GLGC) | 1 | 39719519 | T | C |
| HDL-C (GLGC) | 1 | 39730172 | G | A |
| HDL-C (GLGC) | 1 | 39797055 | G | A |
| HDL-C (GLGC) | 1 | 39826005 | G | A |
| HDL-C (GLGC) | 1 | 39908506 | G | A |
| HDL-C (GLGC) | 1 | 39945297 | T | C |
| HDL-C (GLGC) | 1 | 39950080 | T | C |
| HDL-C (GLGC) | 1 | 39961999 | G | A |
| HDL-C (GLGC) | 1 | 40018509 | T | C |
| HDL-C (GLGC) | 1 | 40028180 | G | A |
| HDL-C (GLGC) | 1 | 40037902 | G | A |
| HDL-C (GLGC) | 1 | 40064961 | G | A |
| HDL-C (GLGC) | 1 | 40069939 | G | A |
| HDL-C (GLGC) | 1 | 93620393 | G | A |
| HDL-C (GLGC) | 1 | 93724924 | G | A |
| HDL-C (GLGC) | 1 | 93761896 | G | A |
| HDL-C (GLGC) | 1 | 93816400 | G | A |

**Table S14:** Genetic variants used in the presented analyses. Note that due to coverage of the GWAS chip the instruments differed across considered outcomes *(continued)*

| Exposure | Chromosome | Base-pair position (Build 37) | Other allele | Effect allele |
|---|---|---|---|---|
| HDL-C (GLGC) | 1 | 93817946 | G | A |
| HDL-C (GLGC) | 20 | 33175466 | T | C |
| HDL-C (GLGC) | 20 | 33409350 | G | A |
| HDL-C (GLGC) | 20 | 33529766 | T | G |
| HDL-C (GLGC) | 20 | 33777983 | G | A |
| HDL-C (GLGC) | 20 | 43038249 | T | C |
| HDL-C (GLGC) | 20 | 43042364 | T | C |
| HDL-C (GLGC) | 20 | 44427418 | G | A |
| HDL-C (GLGC) | 20 | 44496029 | C | A |
| HDL-C (GLGC) | 20 | 44532255 | G | A |
| HDL-C (GLGC) | 20 | 44534651 | G | A |
| HDL-C (GLGC) | 20 | 44536932 | G | A |
| HDL-C (GLGC) | 20 | 44545048 | T | C |
| HDL-C (GLGC) | 20 | 44547068 | G | A |
| HDL-C (GLGC) | 20 | 44564682 | G | A |
| HDL-C (GLGC) | 20 | 44576502 | T | C |
| HDL-C (GLGC) | 20 | 44586036 | G | A |
| HDL-C (GLGC) | 20 | 44592364 | T | C |
| HDL-C (GLGC) | 20 | 44594228 | G | A |
| HDL-C (GLGC) | 20 | 44604822 | G | A |
| HDL-C (GLGC) | 20 | 44607601 | G | A |
| HDL-C (GLGC) | 20 | 44615001 | T | C |
| HDL-C (GLGC) | 20 | 44630804 | G | A |
| HDL-C (GLGC) | 20 | 44633111 | T | C |
| HDL-C (GLGC) | 20 | 44643111 | G | A |
| HDL-C (GLGC) | 20 | 44643592 | G | A |
| HDL-C (GLGC) | 20 | 44644965 | G | A |
| HDL-C (GLGC) | 20 | 44714510 | G | A |
| HDL-C (GLGC) | 20 | 44734371 | G | A |
| HDL-C (GLGC) | 22 | 21910280 | T | C |
| HDL-C (GLGC) | 22 | 21968221 | G | A |
| HDL-C (GLGC) | 22 | 21979096 | T | C |
| HDL-C (GLGC) | 22 | 21982892 | T | C |
| HDL-C (GLGC) | 22 | 21985094 | G | A |
| HDL-C (GLGC) | 2 | 165508300 | T | C |
| HDL-C (GLGC) | 2 | 165508389 | T | C |
| HDL-C (GLGC) | 2 | 165509579 | G | A |
| HDL-C (GLGC) | 2 | 165513091 | T | C |
| HDL-C (GLGC) | 2 | 165528876 | T | C |
| HDL-C (GLGC) | 2 | 165539661 | T | C |
| HDL-C (GLGC) | 2 | 165540800 | T | C |
| HDL-C (GLGC) | 2 | 165543199 | G | A |
| HDL-C (GLGC) | 2 | 165545615 | T | C |
| HDL-C (GLGC) | 2 | 165548569 | G | A |

**Table S14:** Genetic variants used in the presented analyses. Note that due to coverage of the GWAS chip the instruments differed across considered outcomes *(continued)*

| Exposure | Chromosome | Base-pair position (Build 37) | Other allele | Effect allele |
|---|---|---|---|---|
| HDL-C (GLGC) | 2 | 165557318 | T | C |
| HDL-C (GLGC) | 2 | 165558252 | T | C |
| HDL-C (GLGC) | 2 | 165602181 | G | A |
| HDL-C (GLGC) | 2 | 165611362 | G | A |
| HDL-C (GLGC) | 2 | 165623322 | T | G |
| HDL-C (GLGC) | 2 | 165625830 | T | C |
| HDL-C (GLGC) | 2 | 165630201 | C | A |
| HDL-C (GLGC) | 2 | 165634551 | G | A |
| HDL-C (GLGC) | 2 | 165638070 | T | C |
| HDL-C (GLGC) | 2 | 165642448 | G | A |
| HDL-C (GLGC) | 2 | 165644545 | T | C |
| HDL-C (GLGC) | 2 | 21112689 | G | A |
| HDL-C (GLGC) | 2 | 21118983 | C | A |
| HDL-C (GLGC) | 2 | 21121464 | T | C |
| HDL-C (GLGC) | 2 | 21123352 | C | A |
| HDL-C (GLGC) | 2 | 21134040 | C | A |
| HDL-C (GLGC) | 2 | 21135577 | G | A |
| HDL-C (GLGC) | 2 | 21135628 | T | C |
| HDL-C (GLGC) | 2 | 21136327 | T | C |
| HDL-C (GLGC) | 2 | 21138066 | T | C |
| HDL-C (GLGC) | 2 | 21138740 | T | G |
| HDL-C (GLGC) | 2 | 21140540 | T | C |
| HDL-C (GLGC) | 2 | 21141832 | C | A |
| HDL-C (GLGC) | 2 | 21142994 | T | C |
| HDL-C (GLGC) | 2 | 21144829 | T | G |
| HDL-C (GLGC) | 2 | 21145499 | G | A |
| HDL-C (GLGC) | 2 | 21148274 | C | A |
| HDL-C (GLGC) | 2 | 211540507 | C | A |
| HDL-C (GLGC) | 2 | 21184764 | G | A |
| HDL-C (GLGC) | 2 | 21186121 | C | A |
| HDL-C (GLGC) | 2 | 21187641 | T | C |
| HDL-C (GLGC) | 2 | 21188488 | T | C |
| HDL-C (GLGC) | 2 | 21190024 | T | C |
| HDL-C (GLGC) | 2 | 21190209 | T | C |
| HDL-C (GLGC) | 2 | 21193946 | G | A |
| HDL-C (GLGC) | 2 | 21198900 | G | A |
| HDL-C (GLGC) | 2 | 21205563 | G | A |
| HDL-C (GLGC) | 2 | 21208211 | G | A |
| HDL-C (GLGC) | 2 | 21212571 | T | G |
| HDL-C (GLGC) | 2 | 21217490 | T | C |
| HDL-C (GLGC) | 2 | 21224853 | T | C |
| HDL-C (GLGC) | 2 | 21232195 | G | A |
| HDL-C (GLGC) | 2 | 21233972 | T | C |
| HDL-C (GLGC) | 2 | 21240031 | C | A |

**Table S14:** Genetic variants used in the presented analyses. Note that due to coverage of the GWAS chip the instruments differed across considered outcomes *(continued)*

| Exposure | Chromosome | Base-pair position (Build 37) | Other allele | Effect allele |
|---|---|---|---|---|
| HDL-C (GLGC) | 2 | 21240364 | G | A |
| HDL-C (GLGC) | 2 | 21244000 | T | G |
| HDL-C (GLGC) | 2 | 21244358 | T | G |
| HDL-C (GLGC) | 2 | 21245329 | T | G |
| HDL-C (GLGC) | 2 | 21245813 | G | A |
| HDL-C (GLGC) | 2 | 21250914 | G | A |
| HDL-C (GLGC) | 2 | 21259562 | T | C |
| HDL-C (GLGC) | 2 | 21259612 | G | A |
| HDL-C (GLGC) | 2 | 21262427 | G | A |
| HDL-C (GLGC) | 2 | 21263900 | G | A |
| HDL-C (GLGC) | 2 | 21278773 | T | G |
| HDL-C (GLGC) | 2 | 21311691 | G | A |
| HDL-C (GLGC) | 2 | 21321158 | G | A |
| HDL-C (GLGC) | 2 | 21323369 | G | A |
| HDL-C (GLGC) | 2 | 21371068 | T | C |
| HDL-C (GLGC) | 2 | 21389659 | T | G |
| HDL-C (GLGC) | 2 | 227019461 | T | C |
| HDL-C (GLGC) | 2 | 227020653 | G | A |
| HDL-C (GLGC) | 2 | 227020853 | G | A |
| HDL-C (GLGC) | 2 | 227034499 | G | A |
| HDL-C (GLGC) | 2 | 227046342 | T | C |
| HDL-C (GLGC) | 2 | 227052396 | C | A |
| HDL-C (GLGC) | 2 | 227066471 | T | C |
| HDL-C (GLGC) | 2 | 227066574 | C | A |
| HDL-C (GLGC) | 2 | 227068080 | C | A |
| HDL-C (GLGC) | 2 | 227069522 | T | C |
| HDL-C (GLGC) | 2 | 227088577 | C | A |
| HDL-C (GLGC) | 2 | 227098387 | G | A |
| HDL-C (GLGC) | 2 | 227105921 | T | C |
| HDL-C (GLGC) | 2 | 227113202 | T | C |
| HDL-C (GLGC) | 2 | 227123439 | T | C |
| HDL-C (GLGC) | 2 | 227156662 | T | C |
| HDL-C (GLGC) | 2 | 227156730 | T | C |
| HDL-C (GLGC) | 2 | 227177236 | C | A |
| HDL-C (GLGC) | 2 | 85546932 | C | A |
| HDL-C (GLGC) | 2 | 85549527 | T | C |
| HDL-C (GLGC) | 2 | 85552186 | C | A |
| HDL-C (GLGC) | 3 | 11400249 | T | C |
| HDL-C (GLGC) | 3 | 119786887 | C | A |
| HDL-C (GLGC) | 3 | 12448994 | T | C |
| HDL-C (GLGC) | 3 | 131751775 | C | A |
| HDL-C (GLGC) | 3 | 135926622 | T | G |
| HDL-C (GLGC) | 3 | 135932359 | T | C |
| HDL-C (GLGC) | 3 | 136069472 | T | G |

**Table S14:** Genetic variants used in the presented analyses. Note that due to coverage of the GWAS chip the instruments differed across considered outcomes *(continued)*

| Exposure | Chromosome | Base-pair position (Build 37) | Other allele | Effect allele |
|---|---|---|---|---|
| HDL-C (GLGC) | 3 | 156797648 | T | C |
| HDL-C (GLGC) | 3 | 156798732 | G | A |
| HDL-C (GLGC) | 3 | 47061183 | G | A |
| HDL-C (GLGC) | 3 | 49898273 | G | A |
| HDL-C (GLGC) | 3 | 49971514 | C | A |
| HDL-C (GLGC) | 3 | 50002245 | T | C |
| HDL-C (GLGC) | 3 | 50172397 | T | C |
| HDL-C (GLGC) | 3 | 50174197 | T | G |
| HDL-C (GLGC) | 3 | 50240338 | T | C |
| HDL-C (GLGC) | 3 | 52431671 | G | A |
| HDL-C (GLGC) | 3 | 52491447 | C | A |
| HDL-C (GLGC) | 3 | 52492707 | T | C |
| HDL-C (GLGC) | 3 | 52506426 | T | C |
| HDL-C (GLGC) | 3 | 52532118 | G | A |
| HDL-C (GLGC) | 3 | 52677478 | G | A |
| HDL-C (GLGC) | 3 | 52844534 | T | C |
| HDL-C (GLGC) | 3 | 52877270 | G | A |
| HDL-C (GLGC) | 3 | 53102389 | G | A |
| HDL-C (GLGC) | 3 | 53118739 | T | C |
| HDL-C (GLGC) | 3 | 53125922 | T | C |
| HDL-C (GLGC) | 4 | 100014805 | G | A |
| HDL-C (GLGC) | 4 | 102707791 | G | A |
| HDL-C (GLGC) | 4 | 103144890 | G | A |
| HDL-C (GLGC) | 4 | 103183510 | T | C |
| HDL-C (GLGC) | 4 | 103188709 | T | C |
| HDL-C (GLGC) | 4 | 103212512 | T | C |
| HDL-C (GLGC) | 4 | 103363860 | G | A |
| HDL-C (GLGC) | 4 | 103373286 | G | A |
| HDL-C (GLGC) | 4 | 157670537 | T | C |
| HDL-C (GLGC) | 4 | 157734675 | G | A |
| HDL-C (GLGC) | 4 | 26027797 | T | C |
| HDL-C (GLGC) | 4 | 26062990 | G | A |
| HDL-C (GLGC) | 4 | 87769929 | T | C |
| HDL-C (GLGC) | 4 | 87976387 | G | A |
| HDL-C (GLGC) | 4 | 87985166 | T | C |
| HDL-C (GLGC) | 4 | 87990334 | T | G |
| HDL-C (GLGC) | 4 | 87999483 | G | A |
| HDL-C (GLGC) | 4 | 88008782 | G | A |
| HDL-C (GLGC) | 4 | 88053085 | T | C |
| HDL-C (GLGC) | 4 | 88057353 | G | A |
| HDL-C (GLGC) | 4 | 89726283 | C | A |
| HDL-C (GLGC) | 4 | 89740128 | G | A |
| HDL-C (GLGC) | 4 | 89742179 | T | C |
| HDL-C (GLGC) | 5 | 53299435 | T | C |

**Table S14:** Genetic variants used in the presented analyses. Note that due to coverage of the GWAS chip the instruments differed across considered outcomes *(continued)*

| Exposure | Chromosome | Base-pair position (Build 37) | Other allele | Effect allele |
|---|---|---|---|---|
| HDL-C (GLGC) | 5 | 53300662 | G | A |
| HDL-C (GLGC) | 5 | 53303595 | T | C |
| HDL-C (GLGC) | 5 | 53304620 | T | G |
| HDL-C (GLGC) | 5 | 55796319 | T | C |
| HDL-C (GLGC) | 5 | 55803533 | G | A |
| HDL-C (GLGC) | 5 | 55806751 | G | A |
| HDL-C (GLGC) | 5 | 55816888 | T | C |
| HDL-C (GLGC) | 5 | 55856375 | T | G |
| HDL-C (GLGC) | 5 | 55860866 | T | G |
| HDL-C (GLGC) | 5 | 55861786 | T | C |
| HDL-C (GLGC) | 5 | 67714246 | G | A |
| HDL-C (GLGC) | 6 | 109574095 | G | A |
| HDL-C (GLGC) | 6 | 127398595 | C | A |
| HDL-C (GLGC) | 6 | 127409882 | T | C |
| HDL-C (GLGC) | 6 | 127414801 | T | C |
| HDL-C (GLGC) | 6 | 127421733 | T | C |
| HDL-C (GLGC) | 6 | 127431488 | G | A |
| HDL-C (GLGC) | 6 | 127432657 | G | A |
| HDL-C (GLGC) | 6 | 127436064 | T | C |
| HDL-C (GLGC) | 6 | 127436313 | G | A |
| HDL-C (GLGC) | 6 | 127451665 | T | C |
| HDL-C (GLGC) | 6 | 127455138 | T | C |
| HDL-C (GLGC) | 6 | 127457260 | G | A |
| HDL-C (GLGC) | 6 | 127471533 | T | C |
| HDL-C (GLGC) | 6 | 127476516 | G | A |
| HDL-C (GLGC) | 6 | 127481154 | C | A |
| HDL-C (GLGC) | 6 | 127496512 | G | A |
| HDL-C (GLGC) | 6 | 139829666 | T | C |
| HDL-C (GLGC) | 6 | 139831180 | G | A |
| HDL-C (GLGC) | 6 | 139838419 | C | A |
| HDL-C (GLGC) | 6 | 139839423 | C | A |
| HDL-C (GLGC) | 6 | 139840693 | C | A |
| HDL-C (GLGC) | 6 | 139843583 | T | C |
| HDL-C (GLGC) | 6 | 139843787 | G | A |
| HDL-C (GLGC) | 6 | 139844429 | G | A |
| HDL-C (GLGC) | 6 | 152039964 | T | C |
| HDL-C (GLGC) | 6 | 152040291 | T | C |
| HDL-C (GLGC) | 6 | 152042260 | C | A |
| HDL-C (GLGC) | 6 | 160737122 | G | A |
| HDL-C (GLGC) | 6 | 160929904 | G | A |
| HDL-C (GLGC) | 6 | 161082461 | T | C |
| HDL-C (GLGC) | 6 | 25905111 | G | A |
| HDL-C (GLGC) | 6 | 25918855 | G | A |
| HDL-C (GLGC) | 6 | 31081434 | G | A |

**Table S14:** Genetic variants used in the presented analyses. Note that due to coverage of the GWAS chip the instruments differed across considered outcomes *(continued)*

| Exposure | Chromosome | Base-pair position (Build 37) | Other allele | Effect allele |
|---|---|---|---|---|
| HDL-C (GLGC) | 6 | 31100869 | G | A |
| HDL-C (GLGC) | 6 | 31100942 | G | A |
| HDL-C (GLGC) | 6 | 31105413 | G | A |
| HDL-C (GLGC) | 6 | 31331829 | T | C |
| HDL-C (GLGC) | 6 | 31352631 | G | A |
| HDL-C (GLGC) | 6 | 31353593 | G | A |
| HDL-C (GLGC) | 6 | 31362930 | T | C |
| HDL-C (GLGC) | 6 | 31380449 | T | C |
| HDL-C (GLGC) | 6 | 31388214 | G | A |
| HDL-C (GLGC) | 6 | 31601843 | T | C |
| HDL-C (GLGC) | 6 | 31603770 | G | A |
| HDL-C (GLGC) | 6 | 32189841 | T | G |
| HDL-C (GLGC) | 6 | 32363215 | T | C |
| HDL-C (GLGC) | 6 | 32380262 | G | A |
| HDL-C (GLGC) | 6 | 32389648 | G | A |
| HDL-C (GLGC) | 6 | 32408842 | G | A |
| HDL-C (GLGC) | 6 | 32433167 | T | G |
| HDL-C (GLGC) | 6 | 32444198 | T | G |
| HDL-C (GLGC) | 6 | 32575983 | G | A |
| HDL-C (GLGC) | 6 | 32664458 | T | C |
| HDL-C (GLGC) | 6 | 32669373 | T | C |
| HDL-C (GLGC) | 6 | 32678999 | G | A |
| HDL-C (GLGC) | 6 | 32681631 | T | C |
| HDL-C (GLGC) | 6 | 34482498 | G | A |
| HDL-C (GLGC) | 6 | 34503338 | G | A |
| HDL-C (GLGC) | 6 | 34546560 | T | C |
| HDL-C (GLGC) | 6 | 34618893 | G | A |
| HDL-C (GLGC) | 6 | 34622676 | G | A |
| HDL-C (GLGC) | 6 | 34715341 | T | C |
| HDL-C (GLGC) | 6 | 34726838 | G | A |
| HDL-C (GLGC) | 6 | 34727031 | G | A |
| HDL-C (GLGC) | 6 | 34739726 | T | G |
| HDL-C (GLGC) | 6 | 34739932 | G | A |
| HDL-C (GLGC) | 6 | 34764443 | G | A |
| HDL-C (GLGC) | 6 | 34807894 | T | G |
| HDL-C (GLGC) | 6 | 34824636 | G | A |
| HDL-C (GLGC) | 6 | 35057331 | G | A |
| HDL-C (GLGC) | 6 | 35133074 | G | A |
| HDL-C (GLGC) | 6 | 35162141 | T | C |
| HDL-C (GLGC) | 6 | 43757896 | C | A |
| HDL-C (GLGC) | 6 | 43758873 | G | A |
| HDL-C (GLGC) | 6 | 43764551 | T | C |
| HDL-C (GLGC) | 7 | 1083777 | G | A |
| HDL-C (GLGC) | 7 | 130433944 | C | A |

**Table S14:** Genetic variants used in the presented analyses. Note that due to coverage of the GWAS chip the instruments differed across considered outcomes *(continued)*

| Exposure | Chromosome | Base-pair position (Build 37) | Other allele | Effect allele |
|---|---|---|---|---|
| HDL-C (GLGC) | 7 | 130437476 | G | A |
| HDL-C (GLGC) | 7 | 130437689 | G | A |
| HDL-C (GLGC) | 7 | 130445981 | T | C |
| HDL-C (GLGC) | 7 | 130453382 | G | A |
| HDL-C (GLGC) | 7 | 130466091 | G | A |
| HDL-C (GLGC) | 7 | 130466854 | G | A |
| HDL-C (GLGC) | 7 | 130468190 | T | C |
| HDL-C (GLGC) | 7 | 150307167 | G | A |
| HDL-C (GLGC) | 7 | 150529449 | T | C |
| HDL-C (GLGC) | 7 | 17823624 | T | C |
| HDL-C (GLGC) | 7 | 17908546 | T | C |
| HDL-C (GLGC) | 7 | 17911038 | T | C |
| HDL-C (GLGC) | 7 | 18016544 | T | G |
| HDL-C (GLGC) | 7 | 50305863 | T | G |
| HDL-C (GLGC) | 7 | 6396719 | G | A |
| HDL-C (GLGC) | 7 | 6434904 | G | A |
| HDL-C (GLGC) | 7 | 6474073 | T | C |
| HDL-C (GLGC) | 7 | 72856269 | G | A |
| HDL-C (GLGC) | 7 | 72856430 | T | C |
| HDL-C (GLGC) | 7 | 72878645 | G | A |
| HDL-C (GLGC) | 7 | 72885810 | G | A |
| HDL-C (GLGC) | 7 | 72911843 | T | C |
| HDL-C (GLGC) | 7 | 73017005 | G | A |
| HDL-C (GLGC) | 7 | 73026378 | T | C |
| HDL-C (GLGC) | 7 | 73035857 | T | C |
| HDL-C (GLGC) | 7 | 73037366 | G | A |
| HDL-C (GLGC) | 7 | 73042085 | T | C |
| HDL-C (GLGC) | 7 | 73058017 | G | A |
| HDL-C (GLGC) | 8 | 10641965 | G | A |
| HDL-C (GLGC) | 8 | 10643164 | T | C |
| HDL-C (GLGC) | 8 | 10671260 | G | A |
| HDL-C (GLGC) | 8 | 10678089 | C | A |
| HDL-C (GLGC) | 8 | 10683325 | C | A |
| HDL-C (GLGC) | 8 | 10685586 | T | G |
| HDL-C (GLGC) | 8 | 116452340 | T | C |
| HDL-C (GLGC) | 8 | 116464988 | T | C |
| HDL-C (GLGC) | 8 | 116480753 | G | A |
| HDL-C (GLGC) | 8 | 116575459 | G | A |
| HDL-C (GLGC) | 8 | 116599199 | T | G |
| HDL-C (GLGC) | 8 | 116628403 | T | C |
| HDL-C (GLGC) | 8 | 116645056 | T | C |
| HDL-C (GLGC) | 8 | 121868551 | T | G |
| HDL-C (GLGC) | 8 | 121875083 | G | A |
| HDL-C (GLGC) | 8 | 126474306 | G | A |

**Table S14:** Genetic variants used in the presented analyses. Note that due to coverage of the GWAS chip the instruments differed across considered outcomes *(continued)*

| Exposure | Chromosome | Base-pair position (Build 37) | Other allele | Effect allele |
|---|---|---|---|---|
| HDL-C (GLGC) | 8 | 126474356 | G | A |
| HDL-C (GLGC) | 8 | 126474405 | T | G |
| HDL-C (GLGC) | 8 | 126479315 | G | A |
| HDL-C (GLGC) | 8 | 126480367 | T | C |
| HDL-C (GLGC) | 8 | 126480526 | T | C |
| HDL-C (GLGC) | 8 | 126481952 | G | A |
| HDL-C (GLGC) | 8 | 126486409 | G | A |
| HDL-C (GLGC) | 8 | 126488235 | T | C |
| HDL-C (GLGC) | 8 | 126491304 | T | C |
| HDL-C (GLGC) | 8 | 126493392 | G | A |
| HDL-C (GLGC) | 8 | 126493746 | G | A |
| HDL-C (GLGC) | 8 | 126501177 | T | C |
| HDL-C (GLGC) | 8 | 126514676 | T | C |
| HDL-C (GLGC) | 8 | 126516988 | T | C |
| HDL-C (GLGC) | 8 | 126523523 | T | C |
| HDL-C (GLGC) | 8 | 126644200 | T | G |
| HDL-C (GLGC) | 8 | 144245875 | G | A |
| HDL-C (GLGC) | 8 | 144284709 | T | C |
| HDL-C (GLGC) | 8 | 144303418 | G | A |
| HDL-C (GLGC) | 8 | 19514934 | T | C |
| HDL-C (GLGC) | 8 | 19555762 | T | C |
| HDL-C (GLGC) | 8 | 19576448 | T | C |
| HDL-C (GLGC) | 8 | 19577267 | G | A |
| HDL-C (GLGC) | 8 | 19605966 | G | A |
| HDL-C (GLGC) | 8 | 19626120 | T | C |
| HDL-C (GLGC) | 8 | 19632090 | T | G |
| HDL-C (GLGC) | 8 | 19639509 | T | C |
| HDL-C (GLGC) | 8 | 19650333 | T | C |
| HDL-C (GLGC) | 8 | 19662458 | G | A |
| HDL-C (GLGC) | 8 | 19710468 | T | C |
| HDL-C (GLGC) | 8 | 19712564 | T | C |
| HDL-C (GLGC) | 8 | 19715236 | G | A |
| HDL-C (GLGC) | 8 | 19717091 | G | A |
| HDL-C (GLGC) | 8 | 19720833 | T | C |
| HDL-C (GLGC) | 8 | 19726130 | G | A |
| HDL-C (GLGC) | 8 | 19727047 | T | C |
| HDL-C (GLGC) | 8 | 19728815 | T | G |
| HDL-C (GLGC) | 8 | 19734951 | G | A |
| HDL-C (GLGC) | 8 | 19738260 | G | A |
| HDL-C (GLGC) | 8 | 19738812 | T | G |
| HDL-C (GLGC) | 8 | 19739243 | T | G |
| HDL-C (GLGC) | 8 | 19748448 | T | C |
| HDL-C (GLGC) | 8 | 19748856 | T | C |
| HDL-C (GLGC) | 8 | 19749287 | G | A |

**Table S14:** Genetic variants used in the presented analyses. Note that due to coverage of the GWAS chip the instruments differed across considered outcomes *(continued)*

| Exposure | Chromosome | Base-pair position (Build 37) | Other allele | Effect allele |
|---|---|---|---|---|
| HDL-C (GLGC) | 8 | 19749390 | T | C |
| HDL-C (GLGC) | 8 | 19751939 | T | C |
| HDL-C (GLGC) | 8 | 19755626 | C | A |
| HDL-C (GLGC) | 8 | 19755977 | T | G |
| HDL-C (GLGC) | 8 | 19756813 | T | C |
| HDL-C (GLGC) | 8 | 19757300 | T | C |
| HDL-C (GLGC) | 8 | 19757319 | G | A |
| HDL-C (GLGC) | 8 | 19762054 | T | G |
| HDL-C (GLGC) | 8 | 19762816 | C | A |
| HDL-C (GLGC) | 8 | 19774890 | C | A |
| HDL-C (GLGC) | 8 | 19782715 | G | A |
| HDL-C (GLGC) | 8 | 19783362 | C | A |
| HDL-C (GLGC) | 8 | 19786891 | T | C |
| HDL-C (GLGC) | 8 | 19797916 | T | G |
| HDL-C (GLGC) | 8 | 19799641 | T | G |
| HDL-C (GLGC) | 8 | 19805708 | G | A |
| HDL-C (GLGC) | 8 | 19808030 | G | A |
| HDL-C (GLGC) | 8 | 19810787 | C | A |
| HDL-C (GLGC) | 8 | 19811417 | T | C |
| HDL-C (GLGC) | 8 | 19811901 | T | C |
| HDL-C (GLGC) | 8 | 19812812 | T | C |
| HDL-C (GLGC) | 8 | 19813180 | G | A |
| HDL-C (GLGC) | 8 | 19813667 | T | G |
| HDL-C (GLGC) | 8 | 19813702 | G | A |
| HDL-C (GLGC) | 8 | 19814289 | T | C |
| HDL-C (GLGC) | 8 | 19815098 | T | C |
| HDL-C (GLGC) | 8 | 19815189 | T | C |
| HDL-C (GLGC) | 8 | 19816371 | T | C |
| HDL-C (GLGC) | 8 | 19816934 | T | C |
| HDL-C (GLGC) | 8 | 19818042 | G | A |
| HDL-C (GLGC) | 8 | 19818436 | C | A |
| HDL-C (GLGC) | 8 | 19819439 | G | A |
| HDL-C (GLGC) | 8 | 19820396 | G | A |
| HDL-C (GLGC) | 8 | 19823192 | T | C |
| HDL-C (GLGC) | 8 | 19824669 | C | A |
| HDL-C (GLGC) | 8 | 19824896 | T | G |
| HDL-C (GLGC) | 8 | 19826373 | G | A |
| HDL-C (GLGC) | 8 | 19830170 | T | G |
| HDL-C (GLGC) | 8 | 19830766 | T | C |
| HDL-C (GLGC) | 8 | 19830921 | T | C |
| HDL-C (GLGC) | 8 | 19838469 | T | C |
| HDL-C (GLGC) | 8 | 19843137 | C | A |
| HDL-C (GLGC) | 8 | 19843462 | C | A |
| HDL-C (GLGC) | 8 | 19847060 | G | A |

**Table S14:** Genetic variants used in the presented analyses. Note that due to coverage of the GWAS chip the instruments differed across considered outcomes *(continued)*

| Exposure | Chromosome | Base-pair position (Build 37) | Other allele | Effect allele |
|---|---|---|---|---|
| HDL-C (GLGC) | 8 | 19847259 | T | C |
| HDL-C (GLGC) | 8 | 19847690 | C | A |
| HDL-C (GLGC) | 8 | 19848080 | G | A |
| HDL-C (GLGC) | 8 | 19848396 | G | A |
| HDL-C (GLGC) | 8 | 19851897 | T | C |
| HDL-C (GLGC) | 8 | 19852156 | T | G |
| HDL-C (GLGC) | 8 | 19853146 | T | C |
| HDL-C (GLGC) | 8 | 19854773 | T | C |
| HDL-C (GLGC) | 8 | 19856539 | T | C |
| HDL-C (GLGC) | 8 | 19859120 | T | C |
| HDL-C (GLGC) | 8 | 19860197 | C | A |
| HDL-C (GLGC) | 8 | 19862746 | G | A |
| HDL-C (GLGC) | 8 | 19863024 | T | C |
| HDL-C (GLGC) | 8 | 19863114 | G | A |
| HDL-C (GLGC) | 8 | 19863301 | G | A |
| HDL-C (GLGC) | 8 | 19863873 | T | C |
| HDL-C (GLGC) | 8 | 19865509 | C | A |
| HDL-C (GLGC) | 8 | 19865843 | G | A |
| HDL-C (GLGC) | 8 | 19868971 | T | C |
| HDL-C (GLGC) | 8 | 19869366 | T | G |
| HDL-C (GLGC) | 8 | 19870693 | G | A |
| HDL-C (GLGC) | 8 | 19871637 | T | C |
| HDL-C (GLGC) | 8 | 19875365 | G | A |
| HDL-C (GLGC) | 8 | 19878154 | T | C |
| HDL-C (GLGC) | 8 | 19878356 | T | C |
| HDL-C (GLGC) | 8 | 19879809 | G | A |
| HDL-C (GLGC) | 8 | 19881186 | T | C |
| HDL-C (GLGC) | 8 | 19889125 | T | G |
| HDL-C (GLGC) | 8 | 19890612 | G | A |
| HDL-C (GLGC) | 8 | 19890654 | T | C |
| HDL-C (GLGC) | 8 | 19890776 | C | A |
| HDL-C (GLGC) | 8 | 19899046 | T | C |
| HDL-C (GLGC) | 8 | 19914598 | C | A |
| HDL-C (GLGC) | 8 | 19915284 | T | C |
| HDL-C (GLGC) | 8 | 19916120 | G | A |
| HDL-C (GLGC) | 8 | 19919655 | T | C |
| HDL-C (GLGC) | 8 | 19922610 | G | A |
| HDL-C (GLGC) | 8 | 19928160 | G | A |
| HDL-C (GLGC) | 8 | 19931855 | T | C |
| HDL-C (GLGC) | 8 | 19932088 | T | C |
| HDL-C (GLGC) | 8 | 19933896 | T | C |
| HDL-C (GLGC) | 8 | 19936687 | G | A |
| HDL-C (GLGC) | 8 | 19939049 | T | C |
| HDL-C (GLGC) | 8 | 19941027 | T | C |

**Table S14:** Genetic variants used in the presented analyses. Note that due to coverage of the GWAS chip the instruments differed across considered outcomes *(continued)*

| Exposure | Chromosome | Base-pair position (Build 37) | Other allele | Effect allele |
|---|---|---|---|---|
| HDL-C (GLGC) | 8 | 19941078 | T | G |
| HDL-C (GLGC) | 8 | 19941145 | C | A |
| HDL-C (GLGC) | 8 | 19941448 | T | C |
| HDL-C (GLGC) | 8 | 19942908 | C | A |
| HDL-C (GLGC) | 8 | 19943027 | G | A |
| HDL-C (GLGC) | 8 | 19943308 | G | A |
| HDL-C (GLGC) | 8 | 19943364 | T | C |
| HDL-C (GLGC) | 8 | 19944078 | T | C |
| HDL-C (GLGC) | 8 | 19949300 | T | C |
| HDL-C (GLGC) | 8 | 19951752 | T | C |
| HDL-C (GLGC) | 8 | 19952028 | T | G |
| HDL-C (GLGC) | 8 | 19955606 | G | A |
| HDL-C (GLGC) | 8 | 19968929 | T | C |
| HDL-C (GLGC) | 8 | 19970889 | G | A |
| HDL-C (GLGC) | 8 | 9168146 | T | C |
| HDL-C (GLGC) | 8 | 9170178 | T | C |
| HDL-C (GLGC) | 8 | 9171735 | T | C |
| HDL-C (GLGC) | 8 | 9172718 | G | A |
| HDL-C (GLGC) | 8 | 9175433 | T | C |
| HDL-C (GLGC) | 8 | 9177732 | T | G |
| HDL-C (GLGC) | 8 | 9178135 | T | C |
| HDL-C (GLGC) | 8 | 9178921 | G | A |
| HDL-C (GLGC) | 8 | 9183207 | G | A |
| HDL-C (GLGC) | 8 | 9183358 | G | A |
| HDL-C (GLGC) | 8 | 9183596 | G | A |
| HDL-C (GLGC) | 8 | 9184691 | T | C |
| HDL-C (GLGC) | 8 | 9187242 | G | A |
| HDL-C (GLGC) | 8 | 9187595 | G | A |
| HDL-C (GLGC) | 8 | 9190564 | T | C |
| HDL-C (GLGC) | 8 | 9192423 | G | A |
| HDL-C (GLGC) | 8 | 9192927 | T | C |
| HDL-C (GLGC) | 8 | 9194172 | G | A |
| HDL-C (GLGC) | 8 | 9196365 | G | A |
| HDL-C (GLGC) | 8 | 9197250 | G | A |
| HDL-C (GLGC) | 8 | 9198185 | G | A |
| HDL-C (GLGC) | 8 | 9200042 | G | A |
| HDL-C (GLGC) | 8 | 9200787 | T | C |
| HDL-C (GLGC) | 8 | 9201787 | T | C |
| HDL-C (GLGC) | 8 | 9204425 | G | A |
| HDL-C (GLGC) | 8 | 9205531 | G | A |
| HDL-C (GLGC) | 8 | 9206069 | G | A |
| HDL-C (GLGC) | 8 | 9206678 | G | A |
| HDL-C (GLGC) | 8 | 9219868 | G | A |
| HDL-C (GLGC) | 8 | 9223532 | T | G |

**Table S14:** Genetic variants used in the presented analyses. Note that due to coverage of the GWAS chip the instruments differed across considered outcomes *(continued)*

| Exposure | Chromosome | Base-pair position (Build 37) | Other allele | Effect allele |
|---|---|---|---|---|
| HDL-C (GLGC) | 8 | 9228403 | C | A |
| HDL-C (GLGC) | 8 | 9231661 | G | A |
| HDL-C (GLGC) | 9 | 107548375 | C | A |
| HDL-C (GLGC) | 9 | 107557315 | G | A |
| HDL-C (GLGC) | 9 | 107559059 | G | A |
| HDL-C (GLGC) | 9 | 107562804 | T | C |
| HDL-C (GLGC) | 9 | 107563269 | T | C |
| HDL-C (GLGC) | 9 | 107569337 | T | C |
| HDL-C (GLGC) | 9 | 107582780 | G | A |
| HDL-C (GLGC) | 9 | 107586753 | T | C |
| HDL-C (GLGC) | 9 | 107588777 | T | C |
| HDL-C (GLGC) | 9 | 107592189 | C | A |
| HDL-C (GLGC) | 9 | 107604197 | T | G |
| HDL-C (GLGC) | 9 | 107604722 | C | A |
| HDL-C (GLGC) | 9 | 107617337 | T | C |
| HDL-C (GLGC) | 9 | 107617433 | T | C |
| HDL-C (GLGC) | 9 | 107624242 | G | A |
| HDL-C (GLGC) | 9 | 107626389 | T | C |
| HDL-C (GLGC) | 9 | 107629104 | T | C |
| HDL-C (GLGC) | 9 | 107635869 | T | C |
| HDL-C (GLGC) | 9 | 107639414 | G | A |
| HDL-C (GLGC) | 9 | 107642287 | T | C |
| HDL-C (GLGC) | 9 | 107645080 | T | C |
| HDL-C (GLGC) | 9 | 107645674 | T | C |
| HDL-C (GLGC) | 9 | 107647019 | T | C |
| HDL-C (GLGC) | 9 | 107647077 | G | A |
| HDL-C (GLGC) | 9 | 107647220 | T | C |
| HDL-C (GLGC) | 9 | 107648458 | G | A |
| HDL-C (GLGC) | 9 | 107651526 | G | A |
| HDL-C (GLGC) | 9 | 107653426 | G | A |
| HDL-C (GLGC) | 9 | 107656122 | C | A |
| HDL-C (GLGC) | 9 | 107657070 | G | A |
| HDL-C (GLGC) | 9 | 107658910 | C | A |
| HDL-C (GLGC) | 9 | 107661122 | G | A |
| HDL-C (GLGC) | 9 | 107661150 | C | A |
| HDL-C (GLGC) | 9 | 107662708 | T | C |
| HDL-C (GLGC) | 9 | 107664301 | T | C |
| HDL-C (GLGC) | 9 | 107664961 | G | A |
| HDL-C (GLGC) | 9 | 107665511 | T | C |
| HDL-C (GLGC) | 9 | 107665739 | G | A |
| HDL-C (GLGC) | 9 | 107665751 | T | C |
| HDL-C (GLGC) | 9 | 107667147 | T | C |
| HDL-C (GLGC) | 9 | 107669073 | T | C |
| HDL-C (GLGC) | 9 | 107669241 | T | C |

**Table S14:** Genetic variants used in the presented analyses. Note that due to coverage of the GWAS chip the instruments differed across considered outcomes *(continued)*

| Exposure | Chromosome | Base-pair position (Build 37) | Other allele | Effect allele |
|---|---|---|---|---|
| HDL-C (GLGC) | 9 | 107672365 | G | A |
| HDL-C (GLGC) | 9 | 107684230 | T | G |
| HDL-C (GLGC) | 9 | 107690450 | G | A |
| HDL-C (GLGC) | 9 | 15284100 | T | C |
| HDL-C (GLGC) | 9 | 15285448 | T | G |
| HDL-C (GLGC) | 9 | 15288045 | G | A |
| HDL-C (GLGC) | 9 | 15289578 | T | C |
| HDL-C (GLGC) | 9 | 15290012 | G | A |
| HDL-C (GLGC) | 9 | 15291239 | T | C |
| HDL-C (GLGC) | 9 | 15293513 | T | C |
| HDL-C (GLGC) | 9 | 15296034 | C | A |
| HDL-C (GLGC) | 9 | 15298666 | T | C |
| HDL-C (GLGC) | 9 | 15301428 | C | A |
| TG (GLGC) | 10 | 113910721 | G | A |
| TG (GLGC) | 10 | 113940329 | T | C |
| TG (GLGC) | 10 | 113975832 | T | C |
| TG (GLGC) | 10 | 114045333 | T | C |
| TG (GLGC) | 10 | 114154815 | T | C |
| TG (GLGC) | 10 | 5196273 | G | A |
| TG (GLGC) | 10 | 5202925 | G | A |
| TG (GLGC) | 10 | 5254146 | G | A |
| TG (GLGC) | 10 | 5254188 | G | A |
| TG (GLGC) | 10 | 5254847 | G | A |
| TG (GLGC) | 10 | 5272499 | T | C |
| TG (GLGC) | 10 | 5286162 | G | A |
| TG (GLGC) | 10 | 64914467 | G | A |
| TG (GLGC) | 10 | 64948684 | T | C |
| TG (GLGC) | 10 | 65048306 | C | A |
| TG (GLGC) | 10 | 65066186 | T | G |
| TG (GLGC) | 10 | 65121565 | C | A |
| TG (GLGC) | 10 | 65133156 | C | A |
| TG (GLGC) | 10 | 65179414 | G | A |
| TG (GLGC) | 10 | 65261184 | T | C |
| TG (GLGC) | 10 | 65337153 | T | C |
| TG (GLGC) | 10 | 65384567 | G | A |
| TG (GLGC) | 10 | 94819053 | T | C |
| TG (GLGC) | 10 | 94839642 | G | A |
| TG (GLGC) | 10 | 94839724 | T | G |
| TG (GLGC) | 10 | 94845036 | G | A |
| TG (GLGC) | 11 | 116352622 | T | C |
| TG (GLGC) | 11 | 116383064 | G | A |
| TG (GLGC) | 11 | 116449283 | T | C |
| TG (GLGC) | 11 | 116496360 | T | C |
| TG (GLGC) | 11 | 116511522 | T | C |

**Table S14:** Genetic variants used in the presented analyses. Note that due to coverage of the GWAS chip the instruments differed across considered outcomes *(continued)*

| Exposure | Chromosome | Base-pair position (Build 37) | Other allele | Effect allele |
|---|---|---|---|---|
| TG (GLGC) | 11 | 116514417 | T | C |
| TG (GLGC) | 11 | 116519655 | T | C |
| TG (GLGC) | 11 | 116519739 | T | C |
| TG (GLGC) | 11 | 116520398 | T | C |
| TG (GLGC) | 11 | 116521748 | G | A |
| TG (GLGC) | 11 | 116522854 | G | A |
| TG (GLGC) | 11 | 116523063 | T | G |
| TG (GLGC) | 11 | 116523844 | G | A |
| TG (GLGC) | 11 | 116525867 | T | C |
| TG (GLGC) | 11 | 116525995 | T | C |
| TG (GLGC) | 11 | 116526322 | C | A |
| TG (GLGC) | 11 | 116527282 | T | C |
| TG (GLGC) | 11 | 116528663 | G | A |
| TG (GLGC) | 11 | 116528876 | T | C |
| TG (GLGC) | 11 | 116529024 | T | C |
| TG (GLGC) | 11 | 116529188 | T | G |
| TG (GLGC) | 11 | 116529442 | T | C |
| TG (GLGC) | 11 | 116529468 | T | C |
| TG (GLGC) | 11 | 116529524 | G | A |
| TG (GLGC) | 11 | 116529641 | T | G |
| TG (GLGC) | 11 | 116530061 | G | A |
| TG (GLGC) | 11 | 116530251 | G | A |
| TG (GLGC) | 11 | 116531739 | T | C |
| TG (GLGC) | 11 | 116532066 | T | G |
| TG (GLGC) | 11 | 116532221 | T | C |
| TG (GLGC) | 11 | 116532314 | G | A |
| TG (GLGC) | 11 | 116535126 | T | C |
| TG (GLGC) | 11 | 116535435 | T | C |
| TG (GLGC) | 11 | 116536418 | G | A |
| TG (GLGC) | 11 | 116550115 | G | A |
| TG (GLGC) | 11 | 116551538 | T | C |
| TG (GLGC) | 11 | 116552562 | T | C |
| TG (GLGC) | 11 | 116553025 | G | A |
| TG (GLGC) | 11 | 116554228 | C | A |
| TG (GLGC) | 11 | 116555415 | T | C |
| TG (GLGC) | 11 | 116556865 | G | A |
| TG (GLGC) | 11 | 116557216 | C | A |
| TG (GLGC) | 11 | 116557314 | T | G |
| TG (GLGC) | 11 | 116559002 | T | C |
| TG (GLGC) | 11 | 116560447 | T | C |
| TG (GLGC) | 11 | 116560667 | T | G |
| TG (GLGC) | 11 | 116561996 | T | C |
| TG (GLGC) | 11 | 116562580 | C | A |
| TG (GLGC) | 11 | 116564408 | T | C |

**Table S14:** Genetic variants used in the presented analyses. Note that due to coverage of the GWAS chip the instruments differed across considered outcomes *(continued)*

| Exposure | Chromosome | Base-pair position (Build 37) | Other allele | Effect allele |
|---|---|---|---|---|
| TG (GLGC) | 11 | 116566117 | T | C |
| TG (GLGC) | 11 | 116571284 | T | C |
| TG (GLGC) | 11 | 116571608 | G | A |
| TG (GLGC) | 11 | 116572089 | T | C |
| TG (GLGC) | 11 | 116572205 | T | C |
| TG (GLGC) | 11 | 116573120 | G | A |
| TG (GLGC) | 11 | 116574336 | G | A |
| TG (GLGC) | 11 | 116574488 | G | A |
| TG (GLGC) | 11 | 116574584 | G | A |
| TG (GLGC) | 11 | 116575473 | G | A |
| TG (GLGC) | 11 | 116575555 | G | A |
| TG (GLGC) | 11 | 116576400 | T | C |
| TG (GLGC) | 11 | 116576415 | G | A |
| TG (GLGC) | 11 | 116577043 | G | A |
| TG (GLGC) | 11 | 116578223 | T | C |
| TG (GLGC) | 11 | 116579663 | T | C |
| TG (GLGC) | 11 | 116581259 | T | C |
| TG (GLGC) | 11 | 116581641 | G | A |
| TG (GLGC) | 11 | 116581800 | T | C |
| TG (GLGC) | 11 | 116584317 | T | C |
| TG (GLGC) | 11 | 116584987 | T | C |
| TG (GLGC) | 11 | 116585533 | G | A |
| TG (GLGC) | 11 | 116586283 | T | C |
| TG (GLGC) | 11 | 116587849 | T | C |
| TG (GLGC) | 11 | 116590787 | G | A |
| TG (GLGC) | 11 | 116595756 | T | C |
| TG (GLGC) | 11 | 116596309 | C | A |
| TG (GLGC) | 11 | 116598988 | G | A |
| TG (GLGC) | 11 | 116599392 | T | C |
| TG (GLGC) | 11 | 116599504 | T | C |
| TG (GLGC) | 11 | 116600021 | T | C |
| TG (GLGC) | 11 | 116601945 | G | A |
| TG (GLGC) | 11 | 116603677 | G | A |
| TG (GLGC) | 11 | 116603724 | C | A |
| TG (GLGC) | 11 | 116607641 | T | C |
| TG (GLGC) | 11 | 116610048 | T | C |
| TG (GLGC) | 11 | 116610249 | G | A |
| TG (GLGC) | 11 | 116611457 | T | C |
| TG (GLGC) | 11 | 116612128 | G | A |
| TG (GLGC) | 11 | 116613184 | T | C |
| TG (GLGC) | 11 | 116613660 | T | C |
| TG (GLGC) | 11 | 116617240 | C | A |
| TG (GLGC) | 11 | 116619867 | C | A |
| TG (GLGC) | 11 | 116621770 | G | A |

**Table S14:** Genetic variants used in the presented analyses. Note that due to coverage of the GWAS chip the instruments differed across considered outcomes *(continued)*

| Exposure | Chromosome | Base-pair position (Build 37) | Other allele | Effect allele |
|---|---|---|---|---|
| TG (GLGC) | 11 | 116623659 | T | C |
| TG (GLGC) | 11 | 116624601 | C | A |
| TG (GLGC) | 11 | 116624703 | T | G |
| TG (GLGC) | 11 | 116625875 | T | G |
| TG (GLGC) | 11 | 116628401 | T | C |
| TG (GLGC) | 11 | 116632798 | T | C |
| TG (GLGC) | 11 | 116633947 | G | A |
| TG (GLGC) | 11 | 116634173 | T | C |
| TG (GLGC) | 11 | 116640237 | G | A |
| TG (GLGC) | 11 | 116641028 | T | C |
| TG (GLGC) | 11 | 116642137 | T | C |
| TG (GLGC) | 11 | 116643436 | T | G |
| TG (GLGC) | 11 | 116645275 | T | C |
| TG (GLGC) | 11 | 116646858 | T | C |
| TG (GLGC) | 11 | 116647120 | T | G |
| TG (GLGC) | 11 | 116648776 | C | A |
| TG (GLGC) | 11 | 116649135 | G | A |
| TG (GLGC) | 11 | 116649538 | G | A |
| TG (GLGC) | 11 | 116651115 | T | C |
| TG (GLGC) | 11 | 116652301 | T | C |
| TG (GLGC) | 11 | 116652351 | G | A |
| TG (GLGC) | 11 | 116652423 | G | A |
| TG (GLGC) | 11 | 116654159 | G | A |
| TG (GLGC) | 11 | 116655600 | G | A |
| TG (GLGC) | 11 | 116657561 | T | C |
| TG (GLGC) | 11 | 116660686 | G | A |
| TG (GLGC) | 11 | 116660813 | G | A |
| TG (GLGC) | 11 | 116661488 | T | C |
| TG (GLGC) | 11 | 116662579 | T | C |
| TG (GLGC) | 11 | 116667545 | T | C |
| TG (GLGC) | 11 | 116668511 | T | C |
| TG (GLGC) | 11 | 116669428 | T | C |
| TG (GLGC) | 11 | 116670182 | T | C |
| TG (GLGC) | 11 | 116670676 | C | A |
| TG (GLGC) | 11 | 116670738 | G | A |
| TG (GLGC) | 11 | 116677723 | G | A |
| TG (GLGC) | 11 | 116681008 | G | A |
| TG (GLGC) | 11 | 116691634 | C | A |
| TG (GLGC) | 11 | 116721826 | T | C |
| TG (GLGC) | 11 | 116722041 | T | G |
| TG (GLGC) | 11 | 116728277 | G | A |
| TG (GLGC) | 11 | 116730638 | G | A |
| TG (GLGC) | 11 | 116733316 | T | C |
| TG (GLGC) | 11 | 116734545 | G | A |

**Table S14:** Genetic variants used in the presented analyses. Note that due to coverage of the GWAS chip the instruments differed across considered outcomes *(continued)*

| Exposure | Chromosome | Base-pair position (Build 37) | Other allele | Effect allele |
|---|---|---|---|---|
| TG (GLGC) | 11 | 116736679 | G | A |
| TG (GLGC) | 11 | 116738046 | G | A |
| TG (GLGC) | 11 | 116741572 | C | A |
| TG (GLGC) | 11 | 116744018 | T | C |
| TG (GLGC) | 11 | 116763034 | G | A |
| TG (GLGC) | 11 | 116772441 | T | C |
| TG (GLGC) | 11 | 116772787 | C | A |
| TG (GLGC) | 11 | 116807970 | G | A |
| TG (GLGC) | 11 | 116829653 | T | G |
| TG (GLGC) | 11 | 116857003 | T | C |
| TG (GLGC) | 11 | 116880456 | T | C |
| TG (GLGC) | 11 | 116896155 | C | A |
| TG (GLGC) | 11 | 116908283 | G | A |
| TG (GLGC) | 11 | 116915742 | T | C |
| TG (GLGC) | 11 | 116919511 | T | C |
| TG (GLGC) | 11 | 116929384 | C | A |
| TG (GLGC) | 11 | 116932561 | T | C |
| TG (GLGC) | 11 | 116942753 | T | C |
| TG (GLGC) | 11 | 116944846 | T | C |
| TG (GLGC) | 11 | 116972894 | C | A |
| TG (GLGC) | 11 | 116972935 | T | C |
| TG (GLGC) | 11 | 116980641 | T | G |
| TG (GLGC) | 11 | 116985641 | T | G |
| TG (GLGC) | 11 | 116988936 | G | A |
| TG (GLGC) | 11 | 116991926 | G | A |
| TG (GLGC) | 11 | 116996539 | G | A |
| TG (GLGC) | 11 | 116997798 | T | C |
| TG (GLGC) | 11 | 117034729 | T | C |
| TG (GLGC) | 11 | 117037065 | T | C |
| TG (GLGC) | 11 | 117037361 | G | A |
| TG (GLGC) | 11 | 117046197 | G | A |
| TG (GLGC) | 11 | 117048417 | G | A |
| TG (GLGC) | 11 | 117059347 | T | C |
| TG (GLGC) | 11 | 117062370 | T | C |
| TG (GLGC) | 11 | 117065476 | G | A |
| TG (GLGC) | 11 | 117068534 | C | A |
| TG (GLGC) | 11 | 117070857 | G | A |
| TG (GLGC) | 11 | 117074299 | T | C |
| TG (GLGC) | 11 | 117075566 | T | C |
| TG (GLGC) | 11 | 117085270 | C | A |
| TG (GLGC) | 11 | 117088450 | G | A |
| TG (GLGC) | 11 | 117089628 | G | A |
| TG (GLGC) | 11 | 117091821 | G | A |
| TG (GLGC) | 11 | 117095361 | T | C |

**Table S14:** Genetic variants used in the presented analyses. Note that due to coverage of the GWAS chip the instruments differed across considered outcomes *(continued)*

| Exposure | Chromosome | Base-pair position (Build 37) | Other allele | Effect allele |
|---|---|---|---|---|
| TG (GLGC) | 11 | 117097952 | T | C |
| TG (GLGC) | 11 | 117100594 | G | A |
| TG (GLGC) | 11 | 117151412 | G | A |
| TG (GLGC) | 11 | 117182708 | T | G |
| TG (GLGC) | 11 | 117199870 | T | C |
| TG (GLGC) | 11 | 27526105 | T | C |
| TG (GLGC) | 11 | 46722221 | T | C |
| TG (GLGC) | 11 | 46798631 | G | A |
| TG (GLGC) | 11 | 46898771 | T | C |
| TG (GLGC) | 11 | 47179829 | G | A |
| TG (GLGC) | 11 | 47259668 | T | C |
| TG (GLGC) | 11 | 47275064 | G | A |
| TG (GLGC) | 11 | 47298360 | G | A |
| TG (GLGC) | 11 | 61543961 | T | C |
| TG (GLGC) | 11 | 61547237 | T | C |
| TG (GLGC) | 11 | 61548559 | G | A |
| TG (GLGC) | 11 | 61560081 | G | A |
| TG (GLGC) | 11 | 61565908 | T | C |
| TG (GLGC) | 11 | 61580635 | T | C |
| TG (GLGC) | 11 | 61581764 | T | C |
| TG (GLGC) | 11 | 61592362 | G | A |
| TG (GLGC) | 11 | 61595564 | T | C |
| TG (GLGC) | 11 | 61597212 | T | C |
| TG (GLGC) | 11 | 61597972 | G | A |
| TG (GLGC) | 11 | 61600327 | G | A |
| TG (GLGC) | 11 | 61600342 | C | A |
| TG (GLGC) | 11 | 61601908 | G | A |
| TG (GLGC) | 11 | 61603237 | T | C |
| TG (GLGC) | 11 | 61605215 | G | A |
| TG (GLGC) | 11 | 61605613 | T | C |
| TG (GLGC) | 11 | 61609750 | T | C |
| TG (GLGC) | 11 | 61611694 | G | A |
| TG (GLGC) | 11 | 61616012 | G | A |
| TG (GLGC) | 11 | 61618608 | G | A |
| TG (GLGC) | 11 | 61618831 | T | C |
| TG (GLGC) | 11 | 61621194 | G | A |
| TG (GLGC) | 11 | 61623793 | G | A |
| TG (GLGC) | 11 | 61624181 | T | C |
| TG (GLGC) | 11 | 61624414 | T | C |
| TG (GLGC) | 11 | 61626921 | T | G |
| TG (GLGC) | 11 | 61627881 | T | C |
| TG (GLGC) | 11 | 61629122 | G | A |
| TG (GLGC) | 11 | 61633182 | T | C |
| TG (GLGC) | 11 | 61639488 | T | C |

**Table S14:** Genetic variants used in the presented analyses. Note that due to coverage of the GWAS chip the instruments differed across considered outcomes *(continued)*

| Exposure | Chromosome | Base-pair position (Build 37) | Other allele | Effect allele |
|---|---|---|---|---|
| TG (GLGC) | 11 | 61647288 | G | A |
| TG (GLGC) | 11 | 61655305 | G | A |
| TG (GLGC) | 11 | 61656117 | G | A |
| TG (GLGC) | 11 | 61745694 | T | C |
| TG (GLGC) | 12 | 107174646 | C | A |
| TG (GLGC) | 12 | 122249091 | G | A |
| TG (GLGC) | 12 | 124393697 | T | G |
| TG (GLGC) | 12 | 124405871 | G | A |
| TG (GLGC) | 12 | 124460167 | T | G |
| TG (GLGC) | 12 | 124475940 | G | A |
| TG (GLGC) | 12 | 133069894 | G | A |
| TG (GLGC) | 12 | 21331549 | T | C |
| TG (GLGC) | 12 | 56951219 | G | A |
| TG (GLGC) | 12 | 57735045 | G | A |
| TG (GLGC) | 12 | 57809456 | T | C |
| TG (GLGC) | 13 | 51221618 | C | A |
| TG (GLGC) | 13 | 95284980 | C | A |
| TG (GLGC) | 15 | 42683787 | G | A |
| TG (GLGC) | 15 | 44016417 | T | C |
| TG (GLGC) | 15 | 44106624 | G | A |
| TG (GLGC) | 15 | 44224668 | G | A |
| TG (GLGC) | 15 | 44234776 | C | A |
| TG (GLGC) | 15 | 44399628 | G | A |
| TG (GLGC) | 15 | 44406021 | C | A |
| TG (GLGC) | 15 | 44515864 | T | C |
| TG (GLGC) | 15 | 44526684 | G | A |
| TG (GLGC) | 15 | 58674051 | G | A |
| TG (GLGC) | 15 | 58674308 | G | A |
| TG (GLGC) | 15 | 58678512 | T | C |
| TG (GLGC) | 15 | 58680638 | T | C |
| TG (GLGC) | 15 | 58683366 | G | A |
| TG (GLGC) | 15 | 58684057 | G | A |
| TG (GLGC) | 15 | 58684478 | T | C |
| TG (GLGC) | 15 | 58687880 | T | C |
| TG (GLGC) | 15 | 58689187 | T | C |
| TG (GLGC) | 15 | 58690754 | G | A |
| TG (GLGC) | 15 | 58692936 | T | C |
| TG (GLGC) | 15 | 58699010 | T | C |
| TG (GLGC) | 15 | 58699937 | T | C |
| TG (GLGC) | 15 | 58702864 | T | C |
| TG (GLGC) | 15 | 58703069 | C | A |
| TG (GLGC) | 15 | 58710627 | C | A |
| TG (GLGC) | 15 | 58712973 | T | C |
| TG (GLGC) | 15 | 58718998 | T | C |

**Table S14:** Genetic variants used in the presented analyses. Note that due to coverage of the GWAS chip the instruments differed across considered outcomes *(continued)*

| Exposure | Chromosome | Base-pair position (Build 37) | Other allele | Effect allele |
|---|---|---|---|---|
| TG (GLGC) | 15 | 58719741 | G | A |
| TG (GLGC) | 15 | 58723675 | T | C |
| TG (GLGC) | 15 | 58723939 | G | A |
| TG (GLGC) | 15 | 58725839 | G | A |
| TG (GLGC) | 15 | 58727325 | G | A |
| TG (GLGC) | 15 | 58731567 | G | A |
| TG (GLGC) | 15 | 58735005 | G | A |
| TG (GLGC) | 15 | 58741177 | T | C |
| TG (GLGC) | 15 | 63396088 | G | A |
| TG (GLGC) | 15 | 63396867 | G | A |
| TG (GLGC) | 15 | 63413084 | T | C |
| TG (GLGC) | 15 | 63413501 | G | A |
| TG (GLGC) | 15 | 63425337 | G | A |
| TG (GLGC) | 15 | 63433766 | G | A |
| TG (GLGC) | 15 | 63434763 | G | A |
| TG (GLGC) | 15 | 72287856 | G | A |
| TG (GLGC) | 15 | 73109629 | G | A |
| TG (GLGC) | 16 | 15129459 | G | A |
| TG (GLGC) | 16 | 15129940 | T | C |
| TG (GLGC) | 16 | 15129970 | G | A |
| TG (GLGC) | 16 | 30823047 | T | G |
| TG (GLGC) | 16 | 30982225 | G | A |
| TG (GLGC) | 16 | 31011183 | G | A |
| TG (GLGC) | 16 | 31048079 | T | C |
| TG (GLGC) | 16 | 31088625 | G | A |
| TG (GLGC) | 16 | 31121793 | G | A |
| TG (GLGC) | 16 | 53804340 | T | C |
| TG (GLGC) | 16 | 53810686 | T | C |
| TG (GLGC) | 16 | 53812614 | G | A |
| TG (GLGC) | 16 | 53813367 | T | G |
| TG (GLGC) | 16 | 53818708 | T | G |
| TG (GLGC) | 16 | 56926195 | G | A |
| TG (GLGC) | 16 | 56985139 | G | A |
| TG (GLGC) | 16 | 56985514 | G | A |
| TG (GLGC) | 16 | 56985555 | G | A |
| TG (GLGC) | 16 | 56987369 | T | C |
| TG (GLGC) | 16 | 56988044 | T | C |
| TG (GLGC) | 16 | 56990716 | C | A |
| TG (GLGC) | 16 | 56993211 | T | C |
| TG (GLGC) | 16 | 56993324 | C | A |
| TG (GLGC) | 16 | 56994894 | G | A |
| TG (GLGC) | 16 | 56995236 | C | A |
| TG (GLGC) | 16 | 56996158 | T | C |
| TG (GLGC) | 16 | 56996211 | G | A |

**Table S14:** Genetic variants used in the presented analyses. Note that due to coverage of the GWAS chip the instruments differed across considered outcomes *(continued)*

| Exposure | Chromosome | Base-pair position (Build 37) | Other allele | Effect allele |
|---|---|---|---|---|
| TG (GLGC) | 16 | 56997233 | G | A |
| TG (GLGC) | 16 | 56999258 | C | A |
| TG (GLGC) | 16 | 56999328 | T | C |
| TG (GLGC) | 16 | 57000938 | G | A |
| TG (GLGC) | 16 | 57002732 | T | G |
| TG (GLGC) | 16 | 57005479 | C | A |
| TG (GLGC) | 16 | 57006590 | T | C |
| TG (GLGC) | 16 | 57007451 | G | A |
| TG (GLGC) | 16 | 72108093 | G | A |
| TG (GLGC) | 16 | 81534790 | T | C |
| TG (GLGC) | 17 | 41865627 | T | G |
| TG (GLGC) | 17 | 41878166 | C | A |
| TG (GLGC) | 17 | 41930569 | G | A |
| TG (GLGC) | 17 | 45750596 | G | A |
| TG (GLGC) | 17 | 47440466 | G | A |
| TG (GLGC) | 17 | 65870073 | T | C |
| TG (GLGC) | 17 | 65947640 | G | A |
| TG (GLGC) | 19 | 19267990 | T | C |
| TG (GLGC) | 19 | 19315180 | G | A |
| TG (GLGC) | 19 | 19325963 | T | G |
| TG (GLGC) | 19 | 19329924 | T | C |
| TG (GLGC) | 19 | 19336608 | T | C |
| TG (GLGC) | 19 | 19378416 | T | C |
| TG (GLGC) | 19 | 19379549 | T | C |
| TG (GLGC) | 19 | 19392401 | T | C |
| TG (GLGC) | 19 | 19406869 | T | C |
| TG (GLGC) | 19 | 19407718 | T | C |
| TG (GLGC) | 19 | 19431423 | T | G |
| TG (GLGC) | 19 | 19456917 | T | C |
| TG (GLGC) | 19 | 19467545 | T | C |
| TG (GLGC) | 19 | 19478022 | T | G |
| TG (GLGC) | 19 | 19508013 | G | A |
| TG (GLGC) | 19 | 19525792 | T | C |
| TG (GLGC) | 19 | 19531910 | G | A |
| TG (GLGC) | 19 | 19532682 | G | A |
| TG (GLGC) | 19 | 19539891 | T | C |
| TG (GLGC) | 19 | 19579046 | T | G |
| TG (GLGC) | 19 | 19591066 | G | A |
| TG (GLGC) | 19 | 19599389 | G | A |
| TG (GLGC) | 19 | 19616953 | T | G |
| TG (GLGC) | 19 | 19650096 | G | A |
| TG (GLGC) | 19 | 19658472 | T | G |
| TG (GLGC) | 19 | 19664077 | T | C |
| TG (GLGC) | 19 | 19666574 | T | C |

**Table S14:** Genetic variants used in the presented analyses. Note that due to coverage of the GWAS chip the instruments differed across considered outcomes *(continued)*

| Exposure | Chromosome | Base-pair position (Build 37) | Other allele | Effect allele |
|---|---|---|---|---|
| TG (GLGC) | 19 | 19668916 | G | A |
| TG (GLGC) | 19 | 19692579 | T | C |
| TG (GLGC) | 19 | 19717056 | G | A |
| TG (GLGC) | 19 | 19738554 | G | A |
| TG (GLGC) | 19 | 19746151 | T | G |
| TG (GLGC) | 19 | 19789528 | G | A |
| TG (GLGC) | 19 | 19795560 | C | A |
| TG (GLGC) | 19 | 19819843 | G | A |
| TG (GLGC) | 19 | 19841752 | T | C |
| TG (GLGC) | 19 | 19920967 | C | A |
| TG (GLGC) | 19 | 33899065 | G | A |
| TG (GLGC) | 19 | 35556744 | T | C |
| TG (GLGC) | 19 | 45242173 | G | A |
| TG (GLGC) | 19 | 45247627 | G | A |
| TG (GLGC) | 19 | 45322744 | G | A |
| TG (GLGC) | 19 | 45370838 | G | A |
| TG (GLGC) | 19 | 45373739 | G | A |
| TG (GLGC) | 19 | 45377098 | C | A |
| TG (GLGC) | 19 | 45379791 | T | C |
| TG (GLGC) | 19 | 45382984 | T | G |
| TG (GLGC) | 19 | 45389596 | G | A |
| TG (GLGC) | 19 | 45392254 | T | C |
| TG (GLGC) | 19 | 45395266 | G | A |
| TG (GLGC) | 19 | 45395619 | G | A |
| TG (GLGC) | 19 | 45398785 | T | C |
| TG (GLGC) | 19 | 45399896 | G | A |
| TG (GLGC) | 19 | 45404691 | G | A |
| TG (GLGC) | 19 | 45406673 | G | A |
| TG (GLGC) | 19 | 45408836 | T | G |
| TG (GLGC) | 19 | 45410002 | G | A |
| TG (GLGC) | 19 | 45412079 | T | C |
| TG (GLGC) | 19 | 45413576 | T | C |
| TG (GLGC) | 19 | 45414451 | T | C |
| TG (GLGC) | 19 | 45415640 | G | A |
| TG (GLGC) | 19 | 45416478 | G | A |
| TG (GLGC) | 19 | 45422846 | G | A |
| TG (GLGC) | 19 | 45422946 | G | A |
| TG (GLGC) | 19 | 45453165 | T | C |
| TG (GLGC) | 19 | 45458146 | C | A |
| TG (GLGC) | 19 | 45490428 | G | A |
| TG (GLGC) | 19 | 47589895 | T | C |
| TG (GLGC) | 19 | 50028163 | G | A |
| TG (GLGC) | 19 | 7184801 | T | C |
| TG (GLGC) | 19 | 7205240 | T | C |

**Table S14:** Genetic variants used in the presented analyses. Note that due to coverage of the GWAS chip the instruments differed across considered outcomes *(continued)*

| Exposure | Chromosome | Base-pair position (Build 37) | Other allele | Effect allele |
|---|---|---|---|---|
| TG (GLGC) | 19 | 7222832 | T | C |
| TG (GLGC) | 19 | 7224431 | G | A |
| TG (GLGC) | 19 | 8586086 | C | A |
| TG (GLGC) | 19 | 8610313 | G | A |
| TG (GLGC) | 19 | 8615589 | G | A |
| TG (GLGC) | 1 | 108656403 | T | G |
| TG (GLGC) | 1 | 154245142 | T | G |
| TG (GLGC) | 1 | 230238821 | T | C |
| TG (GLGC) | 1 | 230264607 | G | A |
| TG (GLGC) | 1 | 230281068 | G | A |
| TG (GLGC) | 1 | 230283764 | G | A |
| TG (GLGC) | 1 | 230287128 | T | G |
| TG (GLGC) | 1 | 230290254 | G | A |
| TG (GLGC) | 1 | 230291441 | C | A |
| TG (GLGC) | 1 | 230295245 | T | C |
| TG (GLGC) | 1 | 230295691 | G | A |
| TG (GLGC) | 1 | 230296470 | C | A |
| TG (GLGC) | 1 | 230300586 | T | C |
| TG (GLGC) | 1 | 230301451 | G | A |
| TG (GLGC) | 1 | 230304988 | G | A |
| TG (GLGC) | 1 | 230307222 | T | G |
| TG (GLGC) | 1 | 230309619 | G | A |
| TG (GLGC) | 1 | 230311176 | G | A |
| TG (GLGC) | 1 | 230321019 | T | C |
| TG (GLGC) | 1 | 230322747 | T | C |
| TG (GLGC) | 1 | 230324801 | G | A |
| TG (GLGC) | 1 | 230326412 | G | A |
| TG (GLGC) | 1 | 230398102 | G | A |
| TG (GLGC) | 1 | 27138393 | T | C |
| TG (GLGC) | 1 | 27236212 | T | C |
| TG (GLGC) | 1 | 27285195 | T | C |
| TG (GLGC) | 1 | 39703847 | G | A |
| TG (GLGC) | 1 | 39797055 | G | A |
| TG (GLGC) | 1 | 39908506 | G | A |
| TG (GLGC) | 1 | 39950080 | T | C |
| TG (GLGC) | 1 | 40018509 | T | C |
| TG (GLGC) | 1 | 40028180 | G | A |
| TG (GLGC) | 1 | 40064961 | G | A |
| TG (GLGC) | 1 | 62833553 | G | A |
| TG (GLGC) | 1 | 62836976 | T | C |
| TG (GLGC) | 1 | 62856695 | G | A |
| TG (GLGC) | 1 | 62860310 | G | A |
| TG (GLGC) | 1 | 62861261 | G | A |
| TG (GLGC) | 1 | 62869139 | G | A |

**Table S14:** Genetic variants used in the presented analyses. Note that due to coverage of the GWAS chip the instruments differed across considered outcomes *(continued)*

| Exposure | Chromosome | Base-pair position (Build 37) | Other allele | Effect allele |
|---|---|---|---|---|
| TG (GLGC) | 1 | 62902874 | G | A |
| TG (GLGC) | 1 | 62917913 | G | A |
| TG (GLGC) | 1 | 62931632 | C | A |
| TG (GLGC) | 1 | 62932120 | T | C |
| TG (GLGC) | 1 | 62953373 | G | A |
| TG (GLGC) | 1 | 62960250 | T | C |
| TG (GLGC) | 1 | 62960575 | T | C |
| TG (GLGC) | 1 | 62991905 | T | C |
| TG (GLGC) | 1 | 62996373 | T | C |
| TG (GLGC) | 1 | 62997573 | C | A |
| TG (GLGC) | 1 | 63022004 | G | A |
| TG (GLGC) | 1 | 63028198 | T | C |
| TG (GLGC) | 1 | 63049551 | C | A |
| TG (GLGC) | 1 | 63055280 | G | A |
| TG (GLGC) | 1 | 63083289 | C | A |
| TG (GLGC) | 1 | 63108374 | G | A |
| TG (GLGC) | 1 | 63117328 | G | A |
| TG (GLGC) | 1 | 63135127 | G | A |
| TG (GLGC) | 1 | 63164029 | T | C |
| TG (GLGC) | 1 | 63177539 | T | G |
| TG (GLGC) | 1 | 63181548 | G | A |
| TG (GLGC) | 1 | 63182135 | T | C |
| TG (GLGC) | 1 | 63191565 | G | A |
| TG (GLGC) | 1 | 63191777 | T | C |
| TG (GLGC) | 1 | 63193972 | G | A |
| TG (GLGC) | 1 | 63194009 | T | C |
| TG (GLGC) | 1 | 63195692 | T | C |
| TG (GLGC) | 1 | 63199348 | T | G |
| TG (GLGC) | 1 | 63199936 | T | G |
| TG (GLGC) | 1 | 63203221 | T | C |
| TG (GLGC) | 1 | 63203906 | G | A |
| TG (GLGC) | 1 | 63248080 | G | A |
| TG (GLGC) | 1 | 63252215 | C | A |
| TG (GLGC) | 1 | 63261652 | G | A |
| TG (GLGC) | 1 | 63268786 | T | C |
| TG (GLGC) | 1 | 63310444 | G | A |
| TG (GLGC) | 1 | 63316576 | T | C |
| TG (GLGC) | 1 | 63342206 | G | A |
| TG (GLGC) | 1 | 63346976 | G | A |
| TG (GLGC) | 1 | 63354972 | G | A |
| TG (GLGC) | 1 | 63358816 | T | C |
| TG (GLGC) | 1 | 63375116 | T | C |
| TG (GLGC) | 20 | 33340313 | T | C |
| TG (GLGC) | 20 | 33525407 | G | A |

**Table S14:** Genetic variants used in the presented analyses. Note that due to coverage of the GWAS chip the instruments differed across considered outcomes *(continued)*

| Exposure | Chromosome | Base-pair position (Build 37) | Other allele | Effect allele |
|---|---|---|---|---|
| TG (GLGC) | 20 | 39110599 | G | A |
| TG (GLGC) | 20 | 39118662 | T | C |
| TG (GLGC) | 20 | 44427418 | G | A |
| TG (GLGC) | 20 | 44496029 | C | A |
| TG (GLGC) | 20 | 44534651 | G | A |
| TG (GLGC) | 20 | 44536932 | G | A |
| TG (GLGC) | 20 | 44545048 | T | C |
| TG (GLGC) | 20 | 44564682 | G | A |
| TG (GLGC) | 20 | 44574533 | T | C |
| TG (GLGC) | 20 | 44576502 | T | C |
| TG (GLGC) | 20 | 44588241 | T | C |
| TG (GLGC) | 20 | 44592364 | T | C |
| TG (GLGC) | 20 | 44594228 | G | A |
| TG (GLGC) | 20 | 44604822 | G | A |
| TG (GLGC) | 20 | 44607601 | G | A |
| TG (GLGC) | 20 | 44610737 | T | G |
| TG (GLGC) | 20 | 44615001 | T | C |
| TG (GLGC) | 20 | 44623967 | T | C |
| TG (GLGC) | 20 | 44628668 | C | A |
| TG (GLGC) | 20 | 44630804 | G | A |
| TG (GLGC) | 20 | 44634550 | T | C |
| TG (GLGC) | 20 | 44639511 | T | C |
| TG (GLGC) | 20 | 44640225 | G | A |
| TG (GLGC) | 20 | 44643111 | G | A |
| TG (GLGC) | 20 | 44643592 | G | A |
| TG (GLGC) | 20 | 44644965 | G | A |
| TG (GLGC) | 20 | 44645010 | T | C |
| TG (GLGC) | 20 | 45592842 | T | C |
| TG (GLGC) | 20 | 45598564 | G | A |
| TG (GLGC) | 22 | 38518622 | G | A |
| TG (GLGC) | 22 | 38544298 | G | A |
| TG (GLGC) | 22 | 38546033 | T | C |
| TG (GLGC) | 22 | 38546118 | T | C |
| TG (GLGC) | 22 | 38546700 | G | A |
| TG (GLGC) | 22 | 38562056 | T | C |
| TG (GLGC) | 22 | 38595411 | G | A |
| TG (GLGC) | 22 | 38898051 | T | C |
| TG (GLGC) | 2 | 165504565 | T | G |
| TG (GLGC) | 2 | 165507843 | T | C |
| TG (GLGC) | 2 | 165509579 | G | A |
| TG (GLGC) | 2 | 165513091 | T | C |
| TG (GLGC) | 2 | 165528876 | T | C |
| TG (GLGC) | 2 | 165539661 | T | C |
| TG (GLGC) | 2 | 165551201 | T | C |

**Table S14:** Genetic variants used in the presented analyses. Note that due to coverage of the GWAS chip the instruments differed across considered outcomes *(continued)*

| Exposure | Chromosome | Base-pair position (Build 37) | Other allele | Effect allele |
|---|---|---|---|---|
| TG (GLGC) | 2 | 165557318 | T | C |
| TG (GLGC) | 2 | 165558252 | T | C |
| TG (GLGC) | 2 | 165562893 | T | C |
| TG (GLGC) | 2 | 165602181 | G | A |
| TG (GLGC) | 2 | 165606954 | T | C |
| TG (GLGC) | 2 | 21118983 | C | A |
| TG (GLGC) | 2 | 21121130 | T | G |
| TG (GLGC) | 2 | 21121464 | T | C |
| TG (GLGC) | 2 | 21123352 | C | A |
| TG (GLGC) | 2 | 21134656 | T | C |
| TG (GLGC) | 2 | 21135577 | G | A |
| TG (GLGC) | 2 | 21135628 | T | C |
| TG (GLGC) | 2 | 21136327 | T | C |
| TG (GLGC) | 2 | 21136564 | T | C |
| TG (GLGC) | 2 | 21138740 | T | G |
| TG (GLGC) | 2 | 21140540 | T | C |
| TG (GLGC) | 2 | 21141832 | C | A |
| TG (GLGC) | 2 | 21142994 | T | C |
| TG (GLGC) | 2 | 21143329 | G | A |
| TG (GLGC) | 2 | 21144829 | T | G |
| TG (GLGC) | 2 | 21145499 | G | A |
| TG (GLGC) | 2 | 21148274 | C | A |
| TG (GLGC) | 2 | 21184764 | G | A |
| TG (GLGC) | 2 | 21187641 | T | C |
| TG (GLGC) | 2 | 21188488 | T | C |
| TG (GLGC) | 2 | 21189068 | T | C |
| TG (GLGC) | 2 | 21190024 | T | C |
| TG (GLGC) | 2 | 21190209 | T | C |
| TG (GLGC) | 2 | 21192523 | G | A |
| TG (GLGC) | 2 | 21196112 | G | A |
| TG (GLGC) | 2 | 21196346 | T | G |
| TG (GLGC) | 2 | 21197159 | T | C |
| TG (GLGC) | 2 | 21205563 | G | A |
| TG (GLGC) | 2 | 21206183 | T | G |
| TG (GLGC) | 2 | 21208211 | G | A |
| TG (GLGC) | 2 | 21214209 | T | C |
| TG (GLGC) | 2 | 21217490 | T | C |
| TG (GLGC) | 2 | 21224853 | T | C |
| TG (GLGC) | 2 | 21232195 | G | A |
| TG (GLGC) | 2 | 21233972 | T | C |
| TG (GLGC) | 2 | 21240031 | C | A |
| TG (GLGC) | 2 | 21240364 | G | A |
| TG (GLGC) | 2 | 21244000 | T | G |
| TG (GLGC) | 2 | 21245329 | T | G |

**Table S14:** Genetic variants used in the presented analyses. Note that due to coverage of the GWAS chip the instruments differed across considered outcomes *(continued)*

| Exposure | Chromosome | Base-pair position (Build 37) | Other allele | Effect allele |
|---|---|---|---|---|
| TG (GLGC) | 2 | 21250914 | G | A |
| TG (GLGC) | 2 | 21259562 | T | C |
| TG (GLGC) | 2 | 21259612 | G | A |
| TG (GLGC) | 2 | 21263900 | G | A |
| TG (GLGC) | 2 | 21300770 | C | A |
| TG (GLGC) | 2 | 21389659 | T | G |
| TG (GLGC) | 2 | 21539609 | C | A |
| TG (GLGC) | 2 | 227020399 | G | A |
| TG (GLGC) | 2 | 227020653 | G | A |
| TG (GLGC) | 2 | 227020853 | G | A |
| TG (GLGC) | 2 | 227034499 | G | A |
| TG (GLGC) | 2 | 227036603 | C | A |
| TG (GLGC) | 2 | 227052396 | C | A |
| TG (GLGC) | 2 | 227068080 | C | A |
| TG (GLGC) | 2 | 227087284 | G | A |
| TG (GLGC) | 2 | 227105921 | T | C |
| TG (GLGC) | 2 | 227113202 | T | C |
| TG (GLGC) | 2 | 227176093 | G | A |
| TG (GLGC) | 2 | 27342894 | T | C |
| TG (GLGC) | 2 | 27361947 | G | A |
| TG (GLGC) | 2 | 27386726 | C | A |
| TG (GLGC) | 2 | 27386799 | T | C |
| TG (GLGC) | 2 | 27388447 | G | A |
| TG (GLGC) | 2 | 27389195 | G | A |
| TG (GLGC) | 2 | 27404846 | C | A |
| TG (GLGC) | 2 | 27405497 | T | C |
| TG (GLGC) | 2 | 27414134 | T | C |
| TG (GLGC) | 2 | 27420605 | G | A |
| TG (GLGC) | 2 | 27424636 | G | A |
| TG (GLGC) | 2 | 27430781 | G | A |
| TG (GLGC) | 2 | 27444546 | T | C |
| TG (GLGC) | 2 | 27445294 | G | A |
| TG (GLGC) | 2 | 27451000 | T | C |
| TG (GLGC) | 2 | 27463351 | T | C |
| TG (GLGC) | 2 | 27466368 | T | C |
| TG (GLGC) | 2 | 27474869 | G | A |
| TG (GLGC) | 2 | 27479543 | T | C |
| TG (GLGC) | 2 | 27487805 | G | A |
| TG (GLGC) | 2 | 27504312 | G | A |
| TG (GLGC) | 2 | 27505758 | T | C |
| TG (GLGC) | 2 | 27509032 | T | C |
| TG (GLGC) | 2 | 27519153 | T | G |
| TG (GLGC) | 2 | 27525661 | G | A |
| TG (GLGC) | 2 | 27533619 | T | C |

**Table S14:** Genetic variants used in the presented analyses. Note that due to coverage of the GWAS chip the instruments differed across considered outcomes *(continued)*

| Exposure | Chromosome | Base-pair position (Build 37) | Other allele | Effect allele |
|---|---|---|---|---|
| TG (GLGC) | 2 | 27542041 | T | C |
| TG (GLGC) | 2 | 27550967 | G | A |
| TG (GLGC) | 2 | 27556721 | G | A |
| TG (GLGC) | 2 | 27558252 | G | A |
| TG (GLGC) | 2 | 27584666 | G | A |
| TG (GLGC) | 2 | 27590666 | T | C |
| TG (GLGC) | 2 | 27591708 | G | A |
| TG (GLGC) | 2 | 27591804 | T | C |
| TG (GLGC) | 2 | 27606098 | G | A |
| TG (GLGC) | 2 | 27609212 | T | C |
| TG (GLGC) | 2 | 27620827 | G | A |
| TG (GLGC) | 2 | 27625005 | T | G |
| TG (GLGC) | 2 | 27635463 | G | A |
| TG (GLGC) | 2 | 27636458 | T | C |
| TG (GLGC) | 2 | 27639153 | G | A |
| TG (GLGC) | 2 | 27642734 | C | A |
| TG (GLGC) | 2 | 27677763 | T | C |
| TG (GLGC) | 2 | 27686911 | T | C |
| TG (GLGC) | 2 | 27703495 | G | A |
| TG (GLGC) | 2 | 27714526 | T | C |
| TG (GLGC) | 2 | 27718474 | T | C |
| TG (GLGC) | 2 | 27730817 | G | A |
| TG (GLGC) | 2 | 27730940 | T | C |
| TG (GLGC) | 2 | 27731212 | T | C |
| TG (GLGC) | 2 | 27737637 | G | A |
| TG (GLGC) | 2 | 27742603 | T | C |
| TG (GLGC) | 2 | 27743154 | G | A |
| TG (GLGC) | 2 | 27743215 | G | A |
| TG (GLGC) | 2 | 27747949 | G | A |
| TG (GLGC) | 2 | 27748597 | C | A |
| TG (GLGC) | 2 | 27748624 | G | A |
| TG (GLGC) | 2 | 27750139 | T | C |
| TG (GLGC) | 2 | 27755168 | G | A |
| TG (GLGC) | 2 | 27778091 | G | A |
| TG (GLGC) | 2 | 27778167 | C | A |
| TG (GLGC) | 2 | 27796159 | T | G |
| TG (GLGC) | 2 | 27801418 | C | A |
| TG (GLGC) | 2 | 27811115 | T | G |
| TG (GLGC) | 2 | 27811722 | G | A |
| TG (GLGC) | 2 | 27815510 | C | A |
| TG (GLGC) | 2 | 27831607 | T | C |
| TG (GLGC) | 2 | 27839369 | T | C |
| TG (GLGC) | 2 | 27860258 | G | A |
| TG (GLGC) | 2 | 27876666 | T | G |

**Table S14:** Genetic variants used in the presented analyses. Note that due to coverage of the GWAS chip the instruments differed across considered outcomes *(continued)*

| Exposure | Chromosome | Base-pair position (Build 37) | Other allele | Effect allele |
|---|---|---|---|---|
| TG (GLGC) | 2 | 27902987 | T | C |
| TG (GLGC) | 2 | 27907444 | T | C |
| TG (GLGC) | 2 | 27928797 | T | C |
| TG (GLGC) | 2 | 27929023 | C | A |
| TG (GLGC) | 2 | 27930837 | T | C |
| TG (GLGC) | 2 | 27934524 | T | C |
| TG (GLGC) | 2 | 27935352 | C | A |
| TG (GLGC) | 2 | 27939071 | T | C |
| TG (GLGC) | 2 | 27949052 | G | A |
| TG (GLGC) | 2 | 27960776 | T | C |
| TG (GLGC) | 2 | 27968454 | C | A |
| TG (GLGC) | 2 | 27972445 | G | A |
| TG (GLGC) | 2 | 27972833 | G | A |
| TG (GLGC) | 2 | 27974971 | G | A |
| TG (GLGC) | 2 | 27980944 | T | C |
| TG (GLGC) | 2 | 27987046 | C | A |
| TG (GLGC) | 2 | 27988821 | G | A |
| TG (GLGC) | 2 | 27993142 | T | G |
| TG (GLGC) | 2 | 27993843 | T | C |
| TG (GLGC) | 2 | 27995931 | G | A |
| TG (GLGC) | 2 | 28002002 | T | C |
| TG (GLGC) | 2 | 28003174 | T | G |
| TG (GLGC) | 2 | 28024331 | T | C |
| TG (GLGC) | 2 | 28033538 | C | A |
| TG (GLGC) | 2 | 28041273 | G | A |
| TG (GLGC) | 2 | 28062396 | T | C |
| TG (GLGC) | 2 | 28073894 | T | C |
| TG (GLGC) | 2 | 28079775 | G | A |
| TG (GLGC) | 2 | 28085544 | C | A |
| TG (GLGC) | 2 | 28113911 | G | A |
| TG (GLGC) | 2 | 28123374 | T | G |
| TG (GLGC) | 2 | 28147541 | T | C |
| TG (GLGC) | 2 | 28151354 | G | A |
| TG (GLGC) | 2 | 28153101 | T | C |
| TG (GLGC) | 2 | 28203294 | T | C |
| TG (GLGC) | 2 | 28217327 | T | C |
| TG (GLGC) | 2 | 28225861 | T | G |
| TG (GLGC) | 2 | 28261674 | T | C |
| TG (GLGC) | 2 | 28268742 | T | C |
| TG (GLGC) | 2 | 28281545 | G | A |
| TG (GLGC) | 2 | 28299267 | G | A |
| TG (GLGC) | 2 | 28301540 | T | C |
| TG (GLGC) | 2 | 28311121 | G | A |
| TG (GLGC) | 2 | 28315770 | T | C |

**Table S14:** Genetic variants used in the presented analyses. Note that due to coverage of the GWAS chip the instruments differed across considered outcomes *(continued)*

| Exposure | Chromosome | Base-pair position (Build 37) | Other allele | Effect allele |
|---|---|---|---|---|
| TG (GLGC) | 2 | 28317086 | G | A |
| TG (GLGC) | 2 | 28319961 | T | G |
| TG (GLGC) | 2 | 28320033 | T | C |
| TG (GLGC) | 2 | 28321056 | G | A |
| TG (GLGC) | 2 | 28323869 | T | C |
| TG (GLGC) | 2 | 28326514 | G | A |
| TG (GLGC) | 2 | 28344285 | G | A |
| TG (GLGC) | 2 | 28350611 | G | A |
| TG (GLGC) | 2 | 28358334 | T | C |
| TG (GLGC) | 2 | 28359016 | T | C |
| TG (GLGC) | 2 | 28364272 | T | C |
| TG (GLGC) | 2 | 28381001 | G | A |
| TG (GLGC) | 2 | 28387200 | G | A |
| TG (GLGC) | 2 | 28398241 | G | A |
| TG (GLGC) | 2 | 28408662 | T | C |
| TG (GLGC) | 2 | 28412558 | G | A |
| TG (GLGC) | 2 | 28414302 | C | A |
| TG (GLGC) | 2 | 28418189 | G | A |
| TG (GLGC) | 2 | 28420462 | T | C |
| TG (GLGC) | 2 | 28421381 | T | C |
| TG (GLGC) | 2 | 28421461 | G | A |
| TG (GLGC) | 2 | 28421473 | T | C |
| TG (GLGC) | 2 | 28424472 | T | C |
| TG (GLGC) | 2 | 28426725 | G | A |
| TG (GLGC) | 2 | 28432237 | G | A |
| TG (GLGC) | 2 | 28443050 | T | C |
| TG (GLGC) | 2 | 28469456 | G | A |
| TG (GLGC) | 2 | 28493709 | C | A |
| TG (GLGC) | 2 | 28502878 | T | C |
| TG (GLGC) | 2 | 28503941 | G | A |
| TG (GLGC) | 2 | 28517861 | G | A |
| TG (GLGC) | 2 | 28521057 | T | C |
| TG (GLGC) | 2 | 28523834 | T | C |
| TG (GLGC) | 2 | 28523995 | T | C |
| TG (GLGC) | 2 | 28542934 | G | A |
| TG (GLGC) | 2 | 28556276 | G | A |
| TG (GLGC) | 2 | 28557080 | T | C |
| TG (GLGC) | 2 | 28560427 | T | C |
| TG (GLGC) | 2 | 28602911 | C | A |
| TG (GLGC) | 2 | 28603637 | C | A |
| TG (GLGC) | 2 | 28604879 | G | A |
| TG (GLGC) | 2 | 28635740 | T | C |
| TG (GLGC) | 2 | 28640327 | T | C |
| TG (GLGC) | 2 | 28645120 | C | A |

**Table S14:** Genetic variants used in the presented analyses. Note that due to coverage of the GWAS chip the instruments differed across considered outcomes *(continued)*

| Exposure | Chromosome | Base-pair position (Build 37) | Other allele | Effect allele |
|---|---|---|---|---|
| TG (GLGC) | 2 | 28645802 | T | C |
| TG (GLGC) | 2 | 28652529 | G | A |
| TG (GLGC) | 2 | 28655788 | T | C |
| TG (GLGC) | 2 | 28661371 | G | A |
| TG (GLGC) | 3 | 12486964 | C | A |
| TG (GLGC) | 3 | 12487612 | T | G |
| TG (GLGC) | 3 | 12489342 | T | G |
| TG (GLGC) | 3 | 135926622 | T | G |
| TG (GLGC) | 3 | 135932359 | T | C |
| TG (GLGC) | 3 | 136069472 | T | G |
| TG (GLGC) | 3 | 170724955 | T | C |
| TG (GLGC) | 3 | 170727408 | G | A |
| TG (GLGC) | 4 | 3438643 | G | A |
| TG (GLGC) | 4 | 3473139 | G | A |
| TG (GLGC) | 4 | 3480136 | T | C |
| TG (GLGC) | 4 | 87279096 | T | C |
| TG (GLGC) | 4 | 87473776 | G | A |
| TG (GLGC) | 4 | 87769929 | T | C |
| TG (GLGC) | 4 | 87863770 | T | C |
| TG (GLGC) | 4 | 87873164 | G | A |
| TG (GLGC) | 4 | 87976387 | G | A |
| TG (GLGC) | 4 | 87984331 | G | A |
| TG (GLGC) | 4 | 87985115 | T | G |
| TG (GLGC) | 4 | 87985166 | T | C |
| TG (GLGC) | 4 | 87985530 | T | G |
| TG (GLGC) | 4 | 87996623 | G | A |
| TG (GLGC) | 4 | 87996745 | G | A |
| TG (GLGC) | 4 | 87999483 | G | A |
| TG (GLGC) | 4 | 88007227 | T | C |
| TG (GLGC) | 4 | 88008782 | G | A |
| TG (GLGC) | 4 | 88057353 | G | A |
| TG (GLGC) | 4 | 88066259 | G | A |
| TG (GLGC) | 4 | 88094650 | T | C |
| TG (GLGC) | 4 | 88102967 | T | C |
| TG (GLGC) | 4 | 88124762 | C | A |
| TG (GLGC) | 4 | 88148812 | C | A |
| TG (GLGC) | 4 | 88151993 | G | A |
| TG (GLGC) | 4 | 88160018 | G | A |
| TG (GLGC) | 4 | 88178919 | T | C |
| TG (GLGC) | 4 | 88180631 | T | G |
| TG (GLGC) | 4 | 88186509 | G | A |
| TG (GLGC) | 4 | 88197499 | T | C |
| TG (GLGC) | 4 | 88199484 | T | G |
| TG (GLGC) | 4 | 88211056 | G | A |

**Table S14:** Genetic variants used in the presented analyses. Note that due to coverage of the GWAS chip the instruments differed across considered outcomes *(continued)*

| Exposure | Chromosome | Base-pair position (Build 37) | Other allele | Effect allele |
|---|---|---|---|---|
| TG (GLGC) | 4 | 89740128 | G | A |
| TG (GLGC) | 4 | 89741269 | G | A |
| TG (GLGC) | 5 | 156369171 | T | C |
| TG (GLGC) | 5 | 156381370 | G | A |
| TG (GLGC) | 5 | 156382308 | G | A |
| TG (GLGC) | 5 | 156390297 | T | C |
| TG (GLGC) | 5 | 156396003 | T | C |
| TG (GLGC) | 5 | 156432215 | G | A |
| TG (GLGC) | 5 | 156460095 | T | C |
| TG (GLGC) | 5 | 156472930 | G | A |
| TG (GLGC) | 5 | 156473185 | G | A |
| TG (GLGC) | 5 | 156479509 | G | A |
| TG (GLGC) | 5 | 173372625 | G | A |
| TG (GLGC) | 5 | 173388311 | T | G |
| TG (GLGC) | 5 | 53297304 | T | C |
| TG (GLGC) | 5 | 53298025 | G | A |
| TG (GLGC) | 5 | 55841824 | G | A |
| TG (GLGC) | 5 | 55854153 | T | C |
| TG (GLGC) | 5 | 55856375 | T | G |
| TG (GLGC) | 5 | 55859146 | T | C |
| TG (GLGC) | 5 | 55859952 | G | A |
| TG (GLGC) | 5 | 55860866 | T | G |
| TG (GLGC) | 5 | 55861786 | T | C |
| TG (GLGC) | 5 | 55868097 | T | C |
| TG (GLGC) | 5 | 55876283 | G | A |
| TG (GLGC) | 6 | 127281547 | T | C |
| TG (GLGC) | 6 | 127379919 | T | C |
| TG (GLGC) | 6 | 127380482 | T | C |
| TG (GLGC) | 6 | 127388186 | T | C |
| TG (GLGC) | 6 | 127392977 | G | A |
| TG (GLGC) | 6 | 127398595 | C | A |
| TG (GLGC) | 6 | 127409882 | T | C |
| TG (GLGC) | 6 | 127414801 | T | C |
| TG (GLGC) | 6 | 127436064 | T | C |
| TG (GLGC) | 6 | 127436313 | G | A |
| TG (GLGC) | 6 | 127454893 | G | A |
| TG (GLGC) | 6 | 127455138 | T | C |
| TG (GLGC) | 6 | 127461682 | C | A |
| TG (GLGC) | 6 | 130349475 | T | G |
| TG (GLGC) | 6 | 139828916 | G | A |
| TG (GLGC) | 6 | 139835689 | T | C |
| TG (GLGC) | 6 | 139838419 | C | A |
| TG (GLGC) | 6 | 139840693 | C | A |
| TG (GLGC) | 6 | 160770360 | G | A |

**Table S14:** Genetic variants used in the presented analyses. Note that due to coverage of the GWAS chip the instruments differed across considered outcomes *(continued)*

| Exposure | Chromosome | Base-pair position (Build 37) | Other allele | Effect allele |
|---|---|---|---|---|
| TG (GLGC) | 6 | 160797625 | T | G |
| TG (GLGC) | 6 | 160838361 | G | A |
| TG (GLGC) | 6 | 160838648 | G | A |
| TG (GLGC) | 6 | 160851766 | T | C |
| TG (GLGC) | 6 | 160906086 | G | A |
| TG (GLGC) | 6 | 160917559 | T | C |
| TG (GLGC) | 6 | 29442701 | T | C |
| TG (GLGC) | 6 | 29821692 | G | A |
| TG (GLGC) | 6 | 29821982 | T | C |
| TG (GLGC) | 6 | 30231224 | G | A |
| TG (GLGC) | 6 | 30296253 | G | A |
| TG (GLGC) | 6 | 30335621 | G | A |
| TG (GLGC) | 6 | 30618906 | G | A |
| TG (GLGC) | 6 | 30688427 | T | G |
| TG (GLGC) | 6 | 30765579 | C | A |
| TG (GLGC) | 6 | 30798697 | T | C |
| TG (GLGC) | 6 | 30800577 | T | C |
| TG (GLGC) | 6 | 30864279 | T | C |
| TG (GLGC) | 6 | 30867527 | G | A |
| TG (GLGC) | 6 | 30932068 | G | A |
| TG (GLGC) | 6 | 30993590 | G | A |
| TG (GLGC) | 6 | 30998742 | T | G |
| TG (GLGC) | 6 | 30999997 | T | C |
| TG (GLGC) | 6 | 31001496 | T | G |
| TG (GLGC) | 6 | 31002013 | G | A |
| TG (GLGC) | 6 | 31002616 | T | C |
| TG (GLGC) | 6 | 31007081 | C | A |
| TG (GLGC) | 6 | 31008598 | C | A |
| TG (GLGC) | 6 | 31009055 | T | C |
| TG (GLGC) | 6 | 31010200 | G | A |
| TG (GLGC) | 6 | 31011247 | T | C |
| TG (GLGC) | 6 | 31015182 | G | A |
| TG (GLGC) | 6 | 31016978 | C | A |
| TG (GLGC) | 6 | 31018546 | T | C |
| TG (GLGC) | 6 | 31081205 | T | C |
| TG (GLGC) | 6 | 31081409 | T | C |
| TG (GLGC) | 6 | 31081434 | G | A |
| TG (GLGC) | 6 | 31082404 | T | C |
| TG (GLGC) | 6 | 31082987 | G | A |
| TG (GLGC) | 6 | 31084048 | G | A |
| TG (GLGC) | 6 | 31084435 | G | A |
| TG (GLGC) | 6 | 31085382 | C | A |
| TG (GLGC) | 6 | 31085770 | G | A |
| TG (GLGC) | 6 | 31087133 | T | C |

**Table S14:** Genetic variants used in the presented analyses. Note that due to coverage of the GWAS chip the instruments differed across considered outcomes *(continued)*

| Exposure | Chromosome | Base-pair position (Build 37) | Other allele | Effect allele |
|---|---|---|---|---|
| TG (GLGC) | 6 | 31087354 | T | C |
| TG (GLGC) | 6 | 31088202 | G | A |
| TG (GLGC) | 6 | 31088922 | G | A |
| TG (GLGC) | 6 | 31091992 | G | A |
| TG (GLGC) | 6 | 31093066 | G | A |
| TG (GLGC) | 6 | 31097301 | T | C |
| TG (GLGC) | 6 | 31106177 | T | C |
| TG (GLGC) | 6 | 31107187 | G | A |
| TG (GLGC) | 6 | 31108607 | G | A |
| TG (GLGC) | 6 | 31117075 | C | A |
| TG (GLGC) | 6 | 31118511 | T | G |
| TG (GLGC) | 6 | 31119422 | T | C |
| TG (GLGC) | 6 | 31119589 | T | G |
| TG (GLGC) | 6 | 31145839 | T | C |
| TG (GLGC) | 6 | 31145991 | T | C |
| TG (GLGC) | 6 | 31157593 | G | A |
| TG (GLGC) | 6 | 31162963 | G | A |
| TG (GLGC) | 6 | 31167927 | T | C |
| TG (GLGC) | 6 | 31170528 | G | A |
| TG (GLGC) | 6 | 31170713 | T | C |
| TG (GLGC) | 6 | 31170914 | G | A |
| TG (GLGC) | 6 | 31171257 | C | A |
| TG (GLGC) | 6 | 31174527 | G | A |
| TG (GLGC) | 6 | 31176335 | T | C |
| TG (GLGC) | 6 | 31184196 | G | A |
| TG (GLGC) | 6 | 31186245 | G | A |
| TG (GLGC) | 6 | 31195218 | T | C |
| TG (GLGC) | 6 | 31198709 | T | C |
| TG (GLGC) | 6 | 31199766 | G | A |
| TG (GLGC) | 6 | 31202665 | G | A |
| TG (GLGC) | 6 | 31204008 | G | A |
| TG (GLGC) | 6 | 31228410 | T | C |
| TG (GLGC) | 6 | 31229404 | T | C |
| TG (GLGC) | 6 | 31229701 | T | C |
| TG (GLGC) | 6 | 31230800 | G | A |
| TG (GLGC) | 6 | 31237449 | T | C |
| TG (GLGC) | 6 | 31240041 | T | C |
| TG (GLGC) | 6 | 31240128 | T | C |
| TG (GLGC) | 6 | 31240293 | T | C |
| TG (GLGC) | 6 | 31240431 | T | G |
| TG (GLGC) | 6 | 31242191 | G | A |
| TG (GLGC) | 6 | 31243347 | T | C |
| TG (GLGC) | 6 | 31243615 | G | A |
| TG (GLGC) | 6 | 31243884 | G | A |

**Table S14:** Genetic variants used in the presented analyses. Note that due to coverage of the GWAS chip the instruments differed across considered outcomes *(continued)*

| Exposure | Chromosome | Base-pair position (Build 37) | Other allele | Effect allele |
|---|---|---|---|---|
| TG (GLGC) | 6 | 31244021 | G | A |
| TG (GLGC) | 6 | 31245821 | G | A |
| TG (GLGC) | 6 | 31248591 | C | A |
| TG (GLGC) | 6 | 31250642 | T | C |
| TG (GLGC) | 6 | 31252469 | T | G |
| TG (GLGC) | 6 | 31254088 | T | C |
| TG (GLGC) | 6 | 31255286 | G | A |
| TG (GLGC) | 6 | 31257511 | C | A |
| TG (GLGC) | 6 | 31258837 | G | A |
| TG (GLGC) | 6 | 31260218 | T | C |
| TG (GLGC) | 6 | 31260397 | G | A |
| TG (GLGC) | 6 | 31265539 | G | A |
| TG (GLGC) | 6 | 31265737 | G | A |
| TG (GLGC) | 6 | 31266117 | C | A |
| TG (GLGC) | 6 | 31267422 | C | A |
| TG (GLGC) | 6 | 31268832 | T | C |
| TG (GLGC) | 6 | 31269173 | G | A |
| TG (GLGC) | 6 | 31269553 | T | C |
| TG (GLGC) | 6 | 31270118 | C | A |
| TG (GLGC) | 6 | 31271285 | G | A |
| TG (GLGC) | 6 | 31271716 | G | A |
| TG (GLGC) | 6 | 31272261 | T | C |
| TG (GLGC) | 6 | 31272612 | G | A |
| TG (GLGC) | 6 | 31273745 | T | C |
| TG (GLGC) | 6 | 31308630 | G | A |
| TG (GLGC) | 6 | 31317182 | T | G |
| TG (GLGC) | 6 | 31318177 | G | A |
| TG (GLGC) | 6 | 31320538 | T | C |
| TG (GLGC) | 6 | 31327334 | T | G |
| TG (GLGC) | 6 | 31327660 | G | A |
| TG (GLGC) | 6 | 31334945 | T | C |
| TG (GLGC) | 6 | 31335901 | G | A |
| TG (GLGC) | 6 | 31347057 | T | C |
| TG (GLGC) | 6 | 31348077 | G | A |
| TG (GLGC) | 6 | 31353329 | G | A |
| TG (GLGC) | 6 | 31386131 | G | A |
| TG (GLGC) | 6 | 31417191 | G | A |
| TG (GLGC) | 6 | 31417742 | T | C |
| TG (GLGC) | 6 | 31420500 | T | G |
| TG (GLGC) | 6 | 31433558 | G | A |
| TG (GLGC) | 6 | 31448862 | C | A |
| TG (GLGC) | 6 | 31448976 | C | A |
| TG (GLGC) | 6 | 31489644 | G | A |
| TG (GLGC) | 6 | 31496915 | G | A |

**Table S14:** Genetic variants used in the presented analyses. Note that due to coverage of the GWAS chip the instruments differed across considered outcomes *(continued)*

| Exposure | Chromosome | Base-pair position (Build 37) | Other allele | Effect allele |
|---|---|---|---|---|
| TG (GLGC) | 6 | 31509967 | T | G |
| TG (GLGC) | 6 | 31533606 | T | C |
| TG (GLGC) | 6 | 31543031 | G | A |
| TG (GLGC) | 6 | 31568469 | G | A |
| TG (GLGC) | 6 | 31629953 | G | A |
| TG (GLGC) | 6 | 31639129 | G | A |
| TG (GLGC) | 6 | 31655438 | G | A |
| TG (GLGC) | 6 | 31675501 | G | A |
| TG (GLGC) | 6 | 31679307 | G | A |
| TG (GLGC) | 6 | 31686497 | G | A |
| TG (GLGC) | 6 | 31704294 | T | G |
| TG (GLGC) | 6 | 31725285 | G | A |
| TG (GLGC) | 6 | 31726253 | T | C |
| TG (GLGC) | 6 | 31764899 | T | C |
| TG (GLGC) | 6 | 31837250 | G | A |
| TG (GLGC) | 6 | 31839309 | T | C |
| TG (GLGC) | 6 | 31839331 | T | C |
| TG (GLGC) | 6 | 31847012 | C | A |
| TG (GLGC) | 6 | 31851234 | G | A |
| TG (GLGC) | 6 | 31851354 | G | A |
| TG (GLGC) | 6 | 31856070 | G | A |
| TG (GLGC) | 6 | 31883957 | G | A |
| TG (GLGC) | 6 | 31916400 | G | A |
| TG (GLGC) | 6 | 31927342 | G | A |
| TG (GLGC) | 6 | 31930351 | G | A |
| TG (GLGC) | 6 | 31940897 | G | A |
| TG (GLGC) | 6 | 31947460 | T | G |
| TG (GLGC) | 6 | 32050758 | T | C |
| TG (GLGC) | 6 | 32138545 | C | A |
| TG (GLGC) | 6 | 32165444 | G | A |
| TG (GLGC) | 6 | 32172993 | T | C |
| TG (GLGC) | 6 | 32184345 | T | C |
| TG (GLGC) | 6 | 32186872 | G | A |
| TG (GLGC) | 6 | 32190620 | T | C |
| TG (GLGC) | 6 | 32192617 | T | C |
| TG (GLGC) | 6 | 32194339 | T | C |
| TG (GLGC) | 6 | 32194609 | G | A |
| TG (GLGC) | 6 | 32205942 | T | C |
| TG (GLGC) | 6 | 32209027 | T | C |
| TG (GLGC) | 6 | 32210799 | G | A |
| TG (GLGC) | 6 | 32213150 | G | A |
| TG (GLGC) | 6 | 32213210 | G | A |
| TG (GLGC) | 6 | 32215188 | T | C |
| TG (GLGC) | 6 | 32215796 | T | G |

**Table S14:** Genetic variants used in the presented analyses. Note that due to coverage of the GWAS chip the instruments differed across considered outcomes *(continued)*

| Exposure | Chromosome | Base-pair position (Build 37) | Other allele | Effect allele |
|---|---|---|---|---|
| TG (GLGC) | 6 | 32217092 | T | C |
| TG (GLGC) | 6 | 32217232 | G | A |
| TG (GLGC) | 6 | 32218843 | G | A |
| TG (GLGC) | 6 | 32221228 | T | C |
| TG (GLGC) | 6 | 32234953 | T | G |
| TG (GLGC) | 6 | 32289318 | T | C |
| TG (GLGC) | 6 | 32290954 | G | A |
| TG (GLGC) | 6 | 32335204 | G | A |
| TG (GLGC) | 6 | 32338695 | G | A |
| TG (GLGC) | 6 | 32357133 | T | C |
| TG (GLGC) | 6 | 32357715 | T | C |
| TG (GLGC) | 6 | 32399337 | T | C |
| TG (GLGC) | 6 | 32401079 | G | A |
| TG (GLGC) | 6 | 32570994 | T | C |
| TG (GLGC) | 6 | 32575983 | G | A |
| TG (GLGC) | 6 | 32578082 | T | C |
| TG (GLGC) | 6 | 32590925 | G | A |
| TG (GLGC) | 6 | 32602269 | T | G |
| TG (GLGC) | 6 | 32605884 | T | C |
| TG (GLGC) | 6 | 32614917 | T | C |
| TG (GLGC) | 6 | 32626272 | C | A |
| TG (GLGC) | 6 | 32626537 | T | G |
| TG (GLGC) | 6 | 32626750 | G | A |
| TG (GLGC) | 6 | 32634467 | T | C |
| TG (GLGC) | 6 | 32659645 | G | A |
| TG (GLGC) | 6 | 32667910 | G | A |
| TG (GLGC) | 6 | 32669373 | T | C |
| TG (GLGC) | 6 | 32682812 | C | A |
| TG (GLGC) | 6 | 32682862 | T | C |
| TG (GLGC) | 6 | 32684257 | G | A |
| TG (GLGC) | 6 | 32751962 | T | C |
| TG (GLGC) | 6 | 43757896 | C | A |
| TG (GLGC) | 6 | 43758873 | G | A |
| TG (GLGC) | 6 | 43764551 | T | C |
| TG (GLGC) | 7 | 116358044 | G | A |
| TG (GLGC) | 7 | 130432481 | T | C |
| TG (GLGC) | 7 | 130437476 | G | A |
| TG (GLGC) | 7 | 130437689 | G | A |
| TG (GLGC) | 7 | 130453382 | G | A |
| TG (GLGC) | 7 | 130466091 | G | A |
| TG (GLGC) | 7 | 130466854 | G | A |
| TG (GLGC) | 7 | 25939161 | T | C |
| TG (GLGC) | 7 | 25939723 | T | G |
| TG (GLGC) | 7 | 25955754 | G | A |

**Table S14:** Genetic variants used in the presented analyses. Note that due to coverage of the GWAS chip the instruments differed across considered outcomes *(continued)*

| Exposure | Chromosome | Base-pair position (Build 37) | Other allele | Effect allele |
|---|---|---|---|---|
| TG (GLGC) | 7 | 25971153 | G | A |
| TG (GLGC) | 7 | 25991826 | T | C |
| TG (GLGC) | 7 | 25997536 | G | A |
| TG (GLGC) | 7 | 26397239 | C | A |
| TG (GLGC) | 7 | 71979623 | G | A |
| TG (GLGC) | 7 | 72313144 | T | C |
| TG (GLGC) | 7 | 72722981 | G | A |
| TG (GLGC) | 7 | 72825125 | G | A |
| TG (GLGC) | 7 | 72850178 | T | C |
| TG (GLGC) | 7 | 72856269 | G | A |
| TG (GLGC) | 7 | 72870211 | G | A |
| TG (GLGC) | 7 | 72878044 | T | G |
| TG (GLGC) | 7 | 72878645 | G | A |
| TG (GLGC) | 7 | 72891754 | T | C |
| TG (GLGC) | 7 | 72904810 | T | C |
| TG (GLGC) | 7 | 72911843 | T | C |
| TG (GLGC) | 7 | 72921609 | T | C |
| TG (GLGC) | 7 | 72934510 | G | A |
| TG (GLGC) | 7 | 72974869 | T | C |
| TG (GLGC) | 7 | 72977898 | G | A |
| TG (GLGC) | 7 | 72981883 | C | A |
| TG (GLGC) | 7 | 72992350 | T | C |
| TG (GLGC) | 7 | 73012042 | G | A |
| TG (GLGC) | 7 | 73020301 | T | C |
| TG (GLGC) | 7 | 73026378 | T | C |
| TG (GLGC) | 7 | 73042085 | T | C |
| TG (GLGC) | 7 | 73042443 | T | C |
| TG (GLGC) | 7 | 73042585 | T | C |
| TG (GLGC) | 7 | 73043101 | G | A |
| TG (GLGC) | 7 | 73043665 | T | C |
| TG (GLGC) | 7 | 73045204 | G | A |
| TG (GLGC) | 7 | 73046758 | C | A |
| TG (GLGC) | 7 | 73046779 | G | A |
| TG (GLGC) | 7 | 73050864 | G | A |
| TG (GLGC) | 7 | 73057550 | G | A |
| TG (GLGC) | 7 | 73058017 | G | A |
| TG (GLGC) | 7 | 73059019 | T | C |
| TG (GLGC) | 7 | 73059990 | T | C |
| TG (GLGC) | 7 | 73060006 | T | C |
| TG (GLGC) | 7 | 73063515 | T | C |
| TG (GLGC) | 7 | 73087884 | T | C |
| TG (GLGC) | 7 | 73096993 | T | C |
| TG (GLGC) | 7 | 73097654 | T | C |
| TG (GLGC) | 7 | 73107003 | C | A |

**Table S14:** Genetic variants used in the presented analyses. Note that due to coverage of the GWAS chip the instruments differed across considered outcomes *(continued)*

| Exposure | Chromosome | Base-pair position (Build 37) | Other allele | Effect allele |
|---|---|---|---|---|
| TG (GLGC) | 7 | 73113483 | T | C |
| TG (GLGC) | 7 | 73121347 | G | A |
| TG (GLGC) | 7 | 73122923 | G | A |
| TG (GLGC) | 7 | 73123473 | G | A |
| TG (GLGC) | 8 | 10190040 | T | C |
| TG (GLGC) | 8 | 10585194 | T | C |
| TG (GLGC) | 8 | 10600254 | G | A |
| TG (GLGC) | 8 | 10606672 | T | C |
| TG (GLGC) | 8 | 10611882 | G | A |
| TG (GLGC) | 8 | 10614275 | T | G |
| TG (GLGC) | 8 | 10620026 | T | C |
| TG (GLGC) | 8 | 10635222 | T | G |
| TG (GLGC) | 8 | 10641965 | G | A |
| TG (GLGC) | 8 | 10643164 | T | C |
| TG (GLGC) | 8 | 10649086 | C | A |
| TG (GLGC) | 8 | 10661734 | G | A |
| TG (GLGC) | 8 | 10668047 | G | A |
| TG (GLGC) | 8 | 10671260 | G | A |
| TG (GLGC) | 8 | 10674467 | T | C |
| TG (GLGC) | 8 | 10675188 | T | C |
| TG (GLGC) | 8 | 10677981 | T | C |
| TG (GLGC) | 8 | 10682777 | G | A |
| TG (GLGC) | 8 | 10683127 | T | C |
| TG (GLGC) | 8 | 10683623 | G | A |
| TG (GLGC) | 8 | 10689050 | T | C |
| TG (GLGC) | 8 | 10690319 | G | A |
| TG (GLGC) | 8 | 10701959 | T | C |
| TG (GLGC) | 8 | 10702534 | G | A |
| TG (GLGC) | 8 | 10705896 | T | C |
| TG (GLGC) | 8 | 10706162 | G | A |
| TG (GLGC) | 8 | 10716028 | G | A |
| TG (GLGC) | 8 | 10797831 | G | A |
| TG (GLGC) | 8 | 10836024 | T | C |
| TG (GLGC) | 8 | 10982051 | G | A |
| TG (GLGC) | 8 | 11012977 | T | C |
| TG (GLGC) | 8 | 11037034 | G | A |
| TG (GLGC) | 8 | 11045161 | G | A |
| TG (GLGC) | 8 | 11069960 | T | C |
| TG (GLGC) | 8 | 11087475 | G | A |
| TG (GLGC) | 8 | 11376266 | T | C |
| TG (GLGC) | 8 | 11396856 | C | A |
| TG (GLGC) | 8 | 11411823 | G | A |
| TG (GLGC) | 8 | 11433909 | T | C |
| TG (GLGC) | 8 | 11444516 | G | A |

**Table S14:** Genetic variants used in the presented analyses. Note that due to coverage of the GWAS chip the instruments differed across considered outcomes *(continued)*

| Exposure | Chromosome | Base-pair position (Build 37) | Other allele | Effect allele |
|---|---|---|---|---|
| TG (GLGC) | 8 | 11450587 | T | G |
| TG (GLGC) | 8 | 11508731 | T | C |
| TG (GLGC) | 8 | 11616410 | T | C |
| TG (GLGC) | 8 | 11622293 | T | C |
| TG (GLGC) | 8 | 11654796 | G | A |
| TG (GLGC) | 8 | 11689228 | T | G |
| TG (GLGC) | 8 | 11829308 | T | C |
| TG (GLGC) | 8 | 11843758 | G | A |
| TG (GLGC) | 8 | 126444788 | G | A |
| TG (GLGC) | 8 | 126447308 | T | C |
| TG (GLGC) | 8 | 126447473 | G | A |
| TG (GLGC) | 8 | 126448789 | G | A |
| TG (GLGC) | 8 | 126449406 | T | C |
| TG (GLGC) | 8 | 126452509 | T | C |
| TG (GLGC) | 8 | 126456376 | G | A |
| TG (GLGC) | 8 | 126457553 | G | A |
| TG (GLGC) | 8 | 126459962 | G | A |
| TG (GLGC) | 8 | 126459990 | G | A |
| TG (GLGC) | 8 | 126462070 | G | A |
| TG (GLGC) | 8 | 126463365 | G | A |
| TG (GLGC) | 8 | 126468621 | T | G |
| TG (GLGC) | 8 | 126470817 | G | A |
| TG (GLGC) | 8 | 126472487 | G | A |
| TG (GLGC) | 8 | 126473441 | G | A |
| TG (GLGC) | 8 | 126474306 | G | A |
| TG (GLGC) | 8 | 126474356 | G | A |
| TG (GLGC) | 8 | 126474405 | T | G |
| TG (GLGC) | 8 | 126480367 | T | C |
| TG (GLGC) | 8 | 126480526 | T | C |
| TG (GLGC) | 8 | 126481694 | T | C |
| TG (GLGC) | 8 | 126482077 | G | A |
| TG (GLGC) | 8 | 126485531 | T | C |
| TG (GLGC) | 8 | 126486287 | T | G |
| TG (GLGC) | 8 | 126486409 | G | A |
| TG (GLGC) | 8 | 126486906 | G | A |
| TG (GLGC) | 8 | 126487857 | G | A |
| TG (GLGC) | 8 | 126488930 | G | A |
| TG (GLGC) | 8 | 126489818 | G | A |
| TG (GLGC) | 8 | 126491304 | T | C |
| TG (GLGC) | 8 | 126493392 | G | A |
| TG (GLGC) | 8 | 126493746 | G | A |
| TG (GLGC) | 8 | 126494562 | T | C |
| TG (GLGC) | 8 | 126494764 | G | A |
| TG (GLGC) | 8 | 126495818 | T | C |

**Table S14:** Genetic variants used in the presented analyses. Note that due to coverage of the GWAS chip the instruments differed across considered outcomes *(continued)*

| Exposure | Chromosome | Base-pair position (Build 37) | Other allele | Effect allele |
|---|---|---|---|---|
| TG (GLGC) | 8 | 126501177 | T | C |
| TG (GLGC) | 8 | 126504726 | T | C |
| TG (GLGC) | 8 | 126504762 | G | A |
| TG (GLGC) | 8 | 126514676 | T | C |
| TG (GLGC) | 8 | 126516197 | G | A |
| TG (GLGC) | 8 | 126516988 | T | C |
| TG (GLGC) | 8 | 126520544 | G | A |
| TG (GLGC) | 8 | 126521465 | G | A |
| TG (GLGC) | 8 | 126523523 | T | C |
| TG (GLGC) | 8 | 126525035 | G | A |
| TG (GLGC) | 8 | 126525814 | G | A |
| TG (GLGC) | 8 | 126526063 | T | G |
| TG (GLGC) | 8 | 126528722 | G | A |
| TG (GLGC) | 8 | 126541659 | G | A |
| TG (GLGC) | 8 | 126546197 | T | G |
| TG (GLGC) | 8 | 126559887 | T | C |
| TG (GLGC) | 8 | 126630159 | T | C |
| TG (GLGC) | 8 | 126645018 | T | C |
| TG (GLGC) | 8 | 126648134 | T | C |
| TG (GLGC) | 8 | 18211158 | T | C |
| TG (GLGC) | 8 | 18228116 | G | A |
| TG (GLGC) | 8 | 18255709 | G | A |
| TG (GLGC) | 8 | 18260598 | G | A |
| TG (GLGC) | 8 | 18262546 | T | C |
| TG (GLGC) | 8 | 18266774 | G | A |
| TG (GLGC) | 8 | 18266953 | T | C |
| TG (GLGC) | 8 | 18267338 | G | A |
| TG (GLGC) | 8 | 18267878 | G | A |
| TG (GLGC) | 8 | 18271120 | T | C |
| TG (GLGC) | 8 | 18272881 | G | A |
| TG (GLGC) | 8 | 18286997 | T | C |
| TG (GLGC) | 8 | 19507270 | T | G |
| TG (GLGC) | 8 | 19514934 | T | C |
| TG (GLGC) | 8 | 19515570 | T | G |
| TG (GLGC) | 8 | 19564149 | G | A |
| TG (GLGC) | 8 | 19568696 | G | A |
| TG (GLGC) | 8 | 19576448 | T | C |
| TG (GLGC) | 8 | 19576471 | T | G |
| TG (GLGC) | 8 | 19576883 | T | C |
| TG (GLGC) | 8 | 19577267 | G | A |
| TG (GLGC) | 8 | 19590044 | G | A |
| TG (GLGC) | 8 | 19605966 | G | A |
| TG (GLGC) | 8 | 19614339 | T | C |
| TG (GLGC) | 8 | 19621333 | T | C |

**Table S14:** Genetic variants used in the presented analyses. Note that due to coverage of the GWAS chip the instruments differed across considered outcomes *(continued)*

| Exposure | Chromosome | Base-pair position (Build 37) | Other allele | Effect allele |
|---|---|---|---|---|
| TG (GLGC) | 8 | 19622349 | T | C |
| TG (GLGC) | 8 | 19622968 | G | A |
| TG (GLGC) | 8 | 19623129 | C | A |
| TG (GLGC) | 8 | 19625912 | G | A |
| TG (GLGC) | 8 | 19632090 | T | G |
| TG (GLGC) | 8 | 19634829 | T | C |
| TG (GLGC) | 8 | 19635117 | T | C |
| TG (GLGC) | 8 | 19639509 | T | C |
| TG (GLGC) | 8 | 19650333 | T | C |
| TG (GLGC) | 8 | 19662458 | G | A |
| TG (GLGC) | 8 | 19709575 | T | C |
| TG (GLGC) | 8 | 19712564 | T | C |
| TG (GLGC) | 8 | 19715236 | G | A |
| TG (GLGC) | 8 | 19717091 | G | A |
| TG (GLGC) | 8 | 19720833 | T | C |
| TG (GLGC) | 8 | 19726130 | G | A |
| TG (GLGC) | 8 | 19727047 | T | C |
| TG (GLGC) | 8 | 19728815 | T | G |
| TG (GLGC) | 8 | 19734951 | G | A |
| TG (GLGC) | 8 | 19738260 | G | A |
| TG (GLGC) | 8 | 19738812 | T | G |
| TG (GLGC) | 8 | 19739243 | T | G |
| TG (GLGC) | 8 | 19748856 | T | C |
| TG (GLGC) | 8 | 19749390 | T | C |
| TG (GLGC) | 8 | 19751939 | T | C |
| TG (GLGC) | 8 | 19752207 | G | A |
| TG (GLGC) | 8 | 19755626 | C | A |
| TG (GLGC) | 8 | 19755977 | T | G |
| TG (GLGC) | 8 | 19757319 | G | A |
| TG (GLGC) | 8 | 19762816 | C | A |
| TG (GLGC) | 8 | 19774890 | C | A |
| TG (GLGC) | 8 | 19775521 | T | C |
| TG (GLGC) | 8 | 19782715 | G | A |
| TG (GLGC) | 8 | 19786891 | T | C |
| TG (GLGC) | 8 | 19797916 | T | G |
| TG (GLGC) | 8 | 19799641 | T | G |
| TG (GLGC) | 8 | 19800005 | G | A |
| TG (GLGC) | 8 | 19802402 | T | C |
| TG (GLGC) | 8 | 19805708 | G | A |
| TG (GLGC) | 8 | 19810787 | C | A |
| TG (GLGC) | 8 | 19811417 | T | C |
| TG (GLGC) | 8 | 19812812 | T | C |
| TG (GLGC) | 8 | 19813180 | G | A |
| TG (GLGC) | 8 | 19813667 | T | G |

**Table S14:** Genetic variants used in the presented analyses. Note that due to coverage of the GWAS chip the instruments differed across considered outcomes *(continued)*

| Exposure | Chromosome | Base-pair position (Build 37) | Other allele | Effect allele |
|---|---|---|---|---|
| TG (GLGC) | 8 | 19813702 | G | A |
| TG (GLGC) | 8 | 19814289 | T | C |
| TG (GLGC) | 8 | 19815098 | T | C |
| TG (GLGC) | 8 | 19815189 | T | C |
| TG (GLGC) | 8 | 19816371 | T | C |
| TG (GLGC) | 8 | 19816934 | T | C |
| TG (GLGC) | 8 | 19818042 | G | A |
| TG (GLGC) | 8 | 19818436 | C | A |
| TG (GLGC) | 8 | 19818976 | C | A |
| TG (GLGC) | 8 | 19819439 | G | A |
| TG (GLGC) | 8 | 19823192 | T | C |
| TG (GLGC) | 8 | 19824669 | C | A |
| TG (GLGC) | 8 | 19824896 | T | G |
| TG (GLGC) | 8 | 19826373 | G | A |
| TG (GLGC) | 8 | 19830170 | T | G |
| TG (GLGC) | 8 | 19830921 | T | C |
| TG (GLGC) | 8 | 19831977 | G | A |
| TG (GLGC) | 8 | 19833729 | T | C |
| TG (GLGC) | 8 | 19833966 | T | C |
| TG (GLGC) | 8 | 19838469 | T | C |
| TG (GLGC) | 8 | 19843137 | C | A |
| TG (GLGC) | 8 | 19843462 | C | A |
| TG (GLGC) | 8 | 19847060 | G | A |
| TG (GLGC) | 8 | 19847690 | C | A |
| TG (GLGC) | 8 | 19848080 | G | A |
| TG (GLGC) | 8 | 19848396 | G | A |
| TG (GLGC) | 8 | 19852156 | T | G |
| TG (GLGC) | 8 | 19852948 | C | A |
| TG (GLGC) | 8 | 19853146 | T | C |
| TG (GLGC) | 8 | 19854361 | T | G |
| TG (GLGC) | 8 | 19854773 | T | C |
| TG (GLGC) | 8 | 19855030 | T | C |
| TG (GLGC) | 8 | 19856539 | T | C |
| TG (GLGC) | 8 | 19858330 | T | C |
| TG (GLGC) | 8 | 19859120 | T | C |
| TG (GLGC) | 8 | 19860197 | C | A |
| TG (GLGC) | 8 | 19862746 | G | A |
| TG (GLGC) | 8 | 19863024 | T | C |
| TG (GLGC) | 8 | 19863301 | G | A |
| TG (GLGC) | 8 | 19863873 | T | C |
| TG (GLGC) | 8 | 19865843 | G | A |
| TG (GLGC) | 8 | 19868971 | T | C |
| TG (GLGC) | 8 | 19869366 | T | G |
| TG (GLGC) | 8 | 19870693 | G | A |

**Table S14:** Genetic variants used in the presented analyses. Note that due to coverage of the GWAS chip the instruments differed across considered outcomes *(continued)*

| Exposure | Chromosome | Base-pair position (Build 37) | Other allele | Effect allele |
|---|---|---|---|---|
| TG (GLGC) | 8 | 19871629 | T | C |
| TG (GLGC) | 8 | 19871637 | T | C |
| TG (GLGC) | 8 | 19875365 | G | A |
| TG (GLGC) | 8 | 19878154 | T | C |
| TG (GLGC) | 8 | 19878356 | T | C |
| TG (GLGC) | 8 | 19879809 | G | A |
| TG (GLGC) | 8 | 19881186 | T | C |
| TG (GLGC) | 8 | 19889125 | T | G |
| TG (GLGC) | 8 | 19890612 | G | A |
| TG (GLGC) | 8 | 19890654 | T | C |
| TG (GLGC) | 8 | 19890776 | C | A |
| TG (GLGC) | 8 | 19899046 | T | C |
| TG (GLGC) | 8 | 19914598 | C | A |
| TG (GLGC) | 8 | 19916120 | G | A |
| TG (GLGC) | 8 | 19919655 | T | C |
| TG (GLGC) | 8 | 19922610 | G | A |
| TG (GLGC) | 8 | 19928160 | G | A |
| TG (GLGC) | 8 | 19932088 | T | C |
| TG (GLGC) | 8 | 19933896 | T | C |
| TG (GLGC) | 8 | 19936687 | G | A |
| TG (GLGC) | 8 | 19939049 | T | C |
| TG (GLGC) | 8 | 19941027 | T | C |
| TG (GLGC) | 8 | 19941078 | T | G |
| TG (GLGC) | 8 | 19941145 | C | A |
| TG (GLGC) | 8 | 19941448 | T | C |
| TG (GLGC) | 8 | 19941854 | G | A |
| TG (GLGC) | 8 | 19943027 | G | A |
| TG (GLGC) | 8 | 19943308 | G | A |
| TG (GLGC) | 8 | 19943364 | T | C |
| TG (GLGC) | 8 | 19944078 | T | C |
| TG (GLGC) | 8 | 19949300 | T | C |
| TG (GLGC) | 8 | 19961274 | T | C |
| TG (GLGC) | 8 | 19968929 | T | C |
| TG (GLGC) | 8 | 59353534 | T | C |
| TG (GLGC) | 8 | 8263945 | G | A |
| TG (GLGC) | 8 | 8269691 | G | A |
| TG (GLGC) | 8 | 8273043 | T | G |
| TG (GLGC) | 8 | 8279462 | T | G |
| TG (GLGC) | 8 | 8692477 | T | C |
| TG (GLGC) | 8 | 9184930 | T | C |
| TG (GLGC) | 8 | 9192927 | T | C |
| TG (GLGC) | 8 | 9239458 | T | G |
| TG (GLGC) | 8 | 9265464 | C | A |
| TG (GLGC) | 8 | 9267117 | T | C |

**Table S14:** Genetic variants used in the presented analyses. Note that due to coverage of the GWAS chip the instruments differed across considered outcomes *(continued)*

| Exposure | Chromosome | Base-pair position (Build 37) | Other allele | Effect allele |
|---|---|---|---|---|
| TG (GLGC) | 8 | 9267251 | T | G |
| TG (GLGC) | 8 | 9370559 | T | C |
| TG (GLGC) | 8 | 9395532 | G | A |
| TG (GLGC) | 8 | 9438928 | T | C |
| TG (GLGC) | 8 | 9511654 | G | A |
| TG (GLGC) | 8 | 9571445 | T | C |
| TG (GLGC) | 8 | 9669775 | G | A |
| TG (GLGC) | 8 | 9794369 | T | C |
| TG (GLGC) | 9 | 107664301 | T | C |
| TG (GLGC) | 9 | 15304782 | C | A |
| TG (GLGC) | 9 | 16904080 | G | A |

**Table S15:** Conditional F-statistics for the considered multivariable Mendelian randomiation models

| Exposure | Median | Quantile: 0.15 | Quantile: 0.85 | Conditional model |
|---|---|---|---|---|
| Apo-A1 | 15.75 | 15.62 | 16.05 | LDL-C |
| HDL-C | 18.36 | 17.84 | 19.28 | LDL-C |
| Apo-B | 11.02 | 9.11 | 11.54 | LDL-C |
| VLDL-C | 7.88 | 7.80 | 8.02 | LDL-C |
| IDL-C | 6.41 | 5.98 | 6.93 | LDL-C |
| Rem-chol | 6.15 | 6.05 | 6.46 | LDL-C |
| TG | 13.34 | 12.88 | 13.79 | LDL-C |
| TC | 14.17 | 12.57 | 14.82 | LDL-C |
| Apo-A1 | 15.60 | 15.52 | 15.81 | HDL-C |
| Apo-B | 18.24 | 17.40 | 19.00 | HDL-C |
| LDL-C | 25.85 | 23.58 | 26.17 | HDL-C |
| VLDL-C | 7.49 | 7.45 | 7.60 | HDL-C |
| IDL-C | 24.86 | 24.20 | 25.51 | HDL-C |
| Rem-chol | 7.40 | 7.40 | 7.65 | HDL-C |
| TG | 12.27 | 11.90 | 13.42 | HDL-C |
| TC | 29.57 | 28.92 | 29.95 | HDL-C |
| Apo-A1 | 16.45 | 16.26 | 16.78 | TG |
| HDL-C | 14.54 | 14.37 | 14.86 | TG |
| Apo-B | 18.11 | 16.85 | 19.30 | TG |
| LDL-C | 19.85 | 16.94 | 20.80 | TG |
| VLDL-C | 3.13 | 3.07 | 3.13 | TG |
| IDL-C | 18.96 | 16.92 | 19.54 | TG |
| Rem-chol | 6.61 | 6.58 | 6.75 | TG |
| TC | 18.12 | 16.67 | 18.64 | TG |
| Apo-A1 | 3.16 | 3.01 | 3.24 | LDL-C, HDL-C & TG |
| HDL-C | 11.11 | 10.94 | 11.31 | LDL-C & TG |
| Apo-B | 2.65 | 2.61 | 2.75 | LDL-C, HDL-C & TG |
| LDL-C | 16.71 | 14.95 | 17.83 | HDL-C & TG |
| VLDL-C | 2.74 | 2.72 | 2.83 | LDL-C, HDL-C & TG |
| IDL-C | 5.49 | 5.42 | 5.74 | LDL-C, HDL-C & TG |
| Rem-chol | 3.15 | 3.13 | 3.29 | LDL-C, HDL-C & TG |
| TG | 8.61 | 8.46 | 9.37 | LDL-C & HDL-C |
| TC | 2.81 | 2.79 | 2.93 | LDL-C, HDL-C & TG |

### *Supplementary results*

#### *NMR GWAS participants characteristics*

Genetic instruments for cholesterol-containing lipoprotein and apolipoprotein concentrations were sourced from a meta-analysis of Kettunen et al, and the UCLEB consortium. The former included data from Finnish, Estonian, Dutch and German cohorts, with a median age of 46.9 years, median BMI of 26.35 kg/$m^2$ and 54% women enrolled, see [2]. UCLEB consisted of UK-based cohorts with a mean age of 58.6 years, mean BMI of 26.4 kg/$m^2$, and 44% women included [1]. Please see the source publications referred to in the main manuscript for the subject characteristics of the outcome data.

#### *Validation of the LDL-C, HDL-C and TG MR effects on disease incidence and biomarkers*

The univariable MR effects of LDL-C, HDL-C, and TG were validated using independent replication data from GLGC based on clinical chemistry measurements (Figure S2), showing strong agreement both in terms of effect direction and significance.

#### *Univariable MR of Apo-B and Apo-A1 concentration*

The MR analyses described for lipid fractions used instruments selected from across the genome (genome-wide MR). In these analyses, horizontal pleiotropy was addressed analytically (see methods). As described by Schmidt *et al.* 2020 [4] MR analyses of protein concentration, such as Apo-B and Apo-A1, may be protected further against horizontal pleiotropy by selecting instrument from a cis window around the protein encoding gene. Here we compared results from genome-wide MR to that of cis-MR and explored agreement (Figure S2, and Tables S11 -12).

Higher Apo-B concentration was positively associated with the risk of CHD, (ischemic) stroke, CD, Alzheimer's disease, and furthermore increased cIMT, carotid plaque and SBP. Conversely, higher genetically instrumented Apo-B concentration was associated with lower HbA1c concentration as well as with pancreatic cancer risk. Aside from a directionally discordant effect on HF and arthritis, results from both cis and genome-wide approached agreed in both effect direction and magnitude, providing empirical support for analytical correction of horizontal pleiotropy. For HF and arthritis, we made further comparisons against the LDL-C, IDL-C and VLDL-C (the carriers Apo-B) effects, providing support for the observed genome-wide Apo-B effect where higher concentrations increased HF risk and lowered arthritis risk (Figure S2). Due to the modest number of cis variants for Apo-A1 (2 APOA1 variants compared to 8 for APOB; Tables S11 -12) agreement between both analytical methods was limited. Instead, we focussed on the genome-wide MR results, where we expect the larger number of variant leads to better control of horizontal pleiotropy. The genome-wide MR results suggest that higher ApoA-1 concentration decreases the risk of CHD, and T2DM, decreases carotid plaque size, DBP, while increasing CRP concentrations (Figure S2).

### Acknowledgements

#### UCLEB consortium

We gratefully acknowledge the following UCLEB contributors: Dale C, Dudbridge F, Engmann J, Jefferis B, Kuh D, Mclachlan S, Menon U, Price A, Talmud P, Tillin T, Walker A, Ben-Shlomo Y, Cooper Y, Hypponen E, Jefferis B, Kumari M, Shah T, Talmud P, Whincup P.
